# Population genomic insights into the evolution of the SARS-CoV-2 Omicron variant

**DOI:** 10.1101/2022.06.27.22276933

**Authors:** Kritika M. Garg, Vinita Lamba, Balaji Chattopadhyay

## Abstract

A thorough understanding of the patterns of population subdivision of a pathogen can prevent disease spread. For SARS-CoV-2, the availability of millions of genomes makes this task analytically challenging. Our study used population genomic methods and identified subtle subdivisions within the Omicron variant, in addition to that captured by the Pango lineage. Further, some of the identified clusters of the Omicron variant revealed statistically significant signatures of selection or expansion revealing the role of microevolutionary processes in the spread of the virus. These are crucial information for policy makers as preventive measures can be designed to mitigate further spread based on a holistic understanding of the variability of the virus and evolutionary processes aiding its spread.

## MAIN TEXT

The exponential increase in the number of SARS-CoV-2 genomes has brought with it a unique set of challenges for data analysis. With over ten million genome sequences already available, new algorithms are being designed to tackle the deluge of data. Most available analytical tools are designed to provide the overall evolutionary relationship between various lineages. However, obtaining a finer level understanding of the diversity and subdivision within a lineage can provide important insights into pathogen evolution, particularly during ongoing pandemics [1]. In this study, we utilized population genomic methods to understand the subdivision of the Omicron lineage of SARS-CoV-2 across the globe in an attempt to elucidate the evolutionary history of the variant.

We downloaded all SARS-CoV-2 genome sequences belonging to the Omicron lineage available up to 31 January 2022 from the GISAID repository (https://www.gisaid.org/, see appendix Table S1) and cleaned them using Nextclade CLI [2] (supporting information). We retained 14,002 good quality sequences, most of which were from Denmark (n=11,272). We aligned the filtered SARS-CoV-2 genomes to the Wuhan reference genome (accession ID: MN908947.1) in Nextalign CLI [2] using default parameters. Further, we assigned the lineage for each sequence using the pangolin web server version 3.1.20 and 4.0.6 accessed on 3^rd^ March and 6^th^ May 2022 respectively [1].

We used two different approaches to understand the population subdivision within our SARS-CoV-2 dataset. For the first approach, we constructed a haplotype network using the program VENAS (Viral genome Evolution Network Analysis System) to generate the SARS-CoV-2 evolution and transmission network [3]. VENAS can analyze thousands of genomes collected in a short span of time (in few minutes) and is a useful tool for tracking changes across a transmission chain. It identifies mutations across alignments and constructs a network based on hamming distances. In VENAS we first estimated the effective parsimony-informative sites (ePIS) and minor allele frequencies (MAF) using default settings and retained 5,253 genomes. These were then used to construct an evolutionary network which was viewed in Cytoscape 3.9.1 using perfuse-force directed method [4]. Default filtering settings in VENAS removed all sequences belonging to BA.1 lineage and hence, we analyzed sequences from this lineage separately (n = 260 genomes for subdivision analysis).

For the second approach we used the discriminant analysis of principal components (DAPC) method to understand the fine-scale subdivision patterns observed in each lineage (based on Pango lineage definitions mentioned above). DAPC is a useful method to detect subdivision as it maximizes between-group differences while minimizing within-group variability [5]. It is a relatively fast method to detect complex subdivision patterns form genomic data. We used the filtered genomes obtained from VENAS and performed DAPC analysis for both lineages using R adegenet package [5,6]. We first identified the optimal number of clusters within each dataset using K-means algorithm and then performed DAPC analysis. We further identified the unique mutations for each of the DAPC clusters and only considered mutation which were present in at least 70% of the sequences belonging to the cluster.

To estimate the level of genomic diversity within our dataset, we characterized all substitutions in reference to the Wuhan genome using VENUS. We considered all 14,003 good quality sequences and identified the mutations for the 12 functional open reading frames (ORF). We further estimated Tajima’s D for the spike protein sequences for all the clusters identified in DAPC using MEGA 10.2.6 [7]. Tajima’s D test is widely used to identify signatures of microevolutionary forces like population fluctuations and selection acting upon populations. We estimated Tajima’s D for each genetic cluster identified by DAPC to avoid confounding effects of population subdivision.

We observed similar pattern of subdivision for each Omicron lineage using both VENUS and DAPC. While VENAS produced numerous nodes/groups (111 for BA.1 and 1,046 BA.2), these were nested within fewer broader subdivisions retrieved by DAPC (Fig. 1). We identified five clusters for the BA.1 lineage and ten clusters for BA.2 lineage. However, when we visualized our results, we observed that two clusters for the BA.1 lineage were clubbed together and four clusters for the BA.2 lineage were clubbed together (Fig. 1B and 1D). Further, no sequences were assigned to clusters 1 and 3 for the BA. 2 lineage. Thus, effectively only four clusters for both BA.1 and BA.2 lineages were identified by the DAPC method. Most of these clusters shared mutations and only a few private mutations were identified (Fig. 1B and 1D). Although, both the methods use different approaches, together they provide a robust understanding of the finer subdivision patterns within fast evolving lineages.

**Figure 1:**
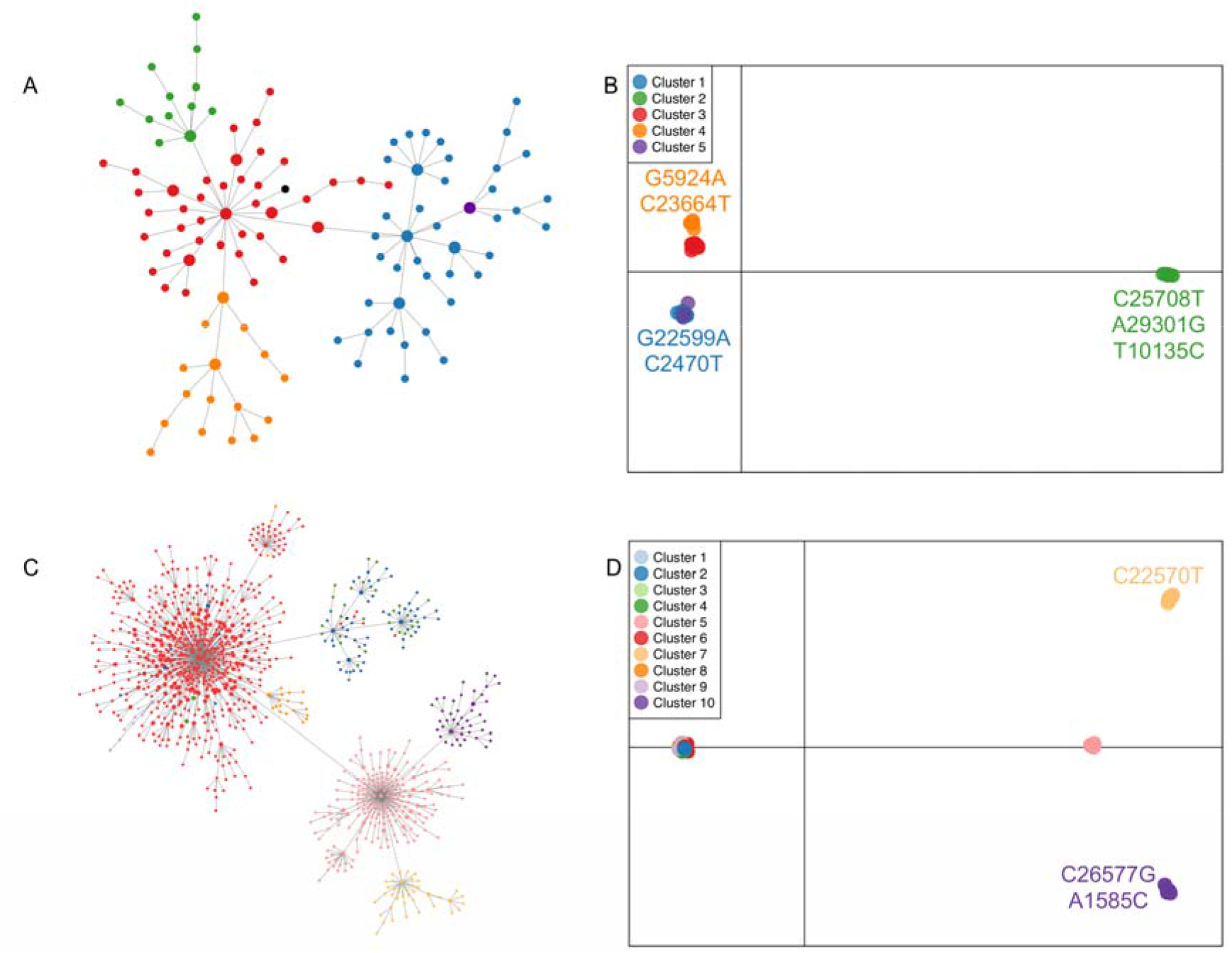
Genetic subdivision observed in Omicron lineage of SARS-CoV-2 based on haplotype network A) BA.1 lineage and C) BA.2 lineage. Panels B and D depict observed genetic subdivision based on DAPC analysis for BA.1 and BA.2 lineages respectively. The Wuhan sequence is denoted in black colored node in panels A and C. Private mutation if any are depicted in the DAPC plot. A private mutation must be present in at least 70% of sequences within the cluster.

At the start of the study, Pango lineage definition 3.1.20 was available which divided the sequences into BA.1, BA.1.1, and BA.2 lineages and our population genomic based clustering identified fine-scale subdivision within these lineages. With the updated Pango lineages version 4.0.6 which is available now, there is broad agreement with the lineage definitions between our methodology (DAPC+ VENAS) and Pango lineage. Using our population genomics pipeline, we could identify mutations used for Pango lineage definitions. For example, G22599A and G5924A are defining mutations for BA.1.1 (cluster 1 and 5) and BA.1.17 (cluster 4) respectively (Fig. 1B). Our method also identified fine-scale subdivision within the Pango definitions, for example, DAPC clusters 5, 7, and 10 all correspond to BA.2.9 lineage from Denmark (Fig. 1D) and were segregated based on three unique mutations (Fig. 1D). One of the mutations unique to cluster 7, C22570T is also used to define the BA.2.9 lineage. However, within our dataset this mutation was only present in cluster 7, highlighting finer subdivision within this lineage (Fig. 1D).

A detailed inspection of the pattern of substitution among the study genomes revealed that as expected, the spike protein, ORF1a, and ORF1b harbored the maximum number of variations (Fig. S1). The most frequent mutations observed were C to T transition and G to T transversion (Fig. S2). Tajima’s D values for the spike protein sequences were negative for all the DAPC clusters, with significant values observed only for four clusters (Table S2). Significant negative Tajima’s D suggests that these DAPC clusters (BA.1 lineage: cluster 1; BA.2 lineage: cluster 2, 5, and 6) are undergoing rapid expansions from a small population size and/or have experienced recent selective sweeps, making them potential target for surveillance and monitoring programs. Interestingly, cluster 2 of BA.1 lineage which did not exhibit a signature of expansion and/or selection, also harbors three unique mutations (C25708T, A29301G, T10135C), which have been identified as suppressors of the spread of this variant by Yang et al. [8] (Fig. 1A and 1B; Table S2). In conjunction, these results identified evolutionary processes affecting virus transmission, highlighting the importance of our approach in identifying sub-lineages of concern and hotspots of spread of SARS-CoV-2. Using a combination of population genomic methods, we could recover subtle variations (within established lineage definitions), some of which can spread more than others. This provides an easy analytical framework which can be used by policy makers to identify variants of potential concern thereby facilitating disease mitigation.

## Data Availability

All data produced are available online at GISAID. We gratefully acknowledge the following Authors from the Originating laboratories responsible for obtaining the specimens and the Submitting laboratories where genetic sequence data were generated and shared via GISAID Initiative, on which this research is based. A full acknowledgement table can be found in Appendix Table S1.

## ACKNOWLEDGEMENTS

B.C. acknowledges the startup funding from Trivedi School of Biosciences (TSB), Ashoka University, India and K.M.G. acknowledges the support from the DBT-Ramalingaswami Fellowship (No. BT/HRD/35/02/2006). V.L. was supported by TSB fellowship. We gratefully acknowledge the following Authors from the Originating laboratories responsible for obtaining the specimens and the Submitting laboratories where genetic sequence data were generated and shared via GISAID Initiative, on which this research is based. A full acknowledgement table can be found in Appendix Table S1.

## DECLARATION OF INTEREST STATEMENT

The authors declare no conflict of interest

## SUPPLEMENTARY MATERIAL

### TABLES

Table S1: Details of the SARS-CoV-2 genome sequences obtained from GISAID.

Table S2: Tajima’s D estimate for the various clusters identified using DAPC. Values in bold indicate significant demographic expansion or selection.

### FIGURES

Figure S1: Distribution of the number of mutations for each open reading frame of SARS-CoV-2 sequences belonging to omicron lineage analyzed in this study. We characterized the mutation in reference to the Wuhan genome.

Figure S2: Frequency distribution of the number of A) transitions, and B) transversion for each open reading frame of SARS-CoV-2 sequences belonging to omicron lineage analyzed in this study. We characterized the mutation in reference to the Wuhan genome.

## Notes

### Competing Interest Statement

The authors have declared no competing interest.

## REFERENCES

1. O’Toole Á, Scher E, Underwood A, Jackson B, Hill V, McCrone JT, et al. Assignment of epidemiological lineages in an emerging pandemic using the pangolin tool. Virus Evol. 2021;7:veab064.

2. Aksamentov I, Roemer C, Hodcroft EB, Neher RA. Nextclade: clade assignment, mutation calling and quality control for viral genomes. J Open Source Softw. 2021;6:3773.

3. Ling Y, Cao R, Qian J, Li J, Zhou H, Yuan L, et al. An interactive viral genome evolution network analysis system enabling rapid large-scale molecular tracing of SARS-CoV-2. Sci Bull (Beijing). 2022;67:665–9.

4. Ideker T. Cytoscape: a software environment for integrated models of biomolecular interaction networks. Genome Res. 2003;13:2498–504.

5. Jombart T. adegenet: a R package for the multivariate analysis of genetic markers. Bioinformatics. 2008;24:1403–5.

6. R Core Team. R: A language and environment for statistical computing. R Foundation for Statistical Computing, Vienna, Austria. 2019. URL https://www.R-project.org/.

7. Kumar S, Tamura K, Nei M. MEGA: molecular evolutionary genetics analysis software for microcomputers. Bioinformatics. 1994;10:189–91.

8. Yang HC, Wang JH, Yang CT, Lin YC, Hsieh HN, Chen PW, Liao HC, Chen CH, Liao JC. Subtyping the major SARS-CoV-2 variants reveals different transmission dynamics. BioRxiv 2022. 10.1101/2022.04.10.486823

