## Supplementary material for "Population genomic insights into the evolution of the SARS-CoV-2 Omicron variant"

**Data cleanup**

We selected only full-length SARS-CoV-2 virus genomes available up to 31 January 2022 from the GISAID repository (https://www.gisaid.org/). The genomes were filtered to remove low-quality sequences (>5% NNNs), genomes without the collection date, and genomes with >0.005% unique amino acid mutations. Only sequences obtained from humans were used for all analysis. We retained 20,067 genomes, which were further filtered for quality using Nextclade CLI [1]. Nextclade examines each query sequence for flaws that could suggest sequencing or assembly errors and assigns a score for each sequence based on the number of N’s, ambiguous sites, private mutation, and stop codons. We considered only good quality sequences for further analysis and retained 14,002 sequences.

**REFERENCES**

1. Aksamentov I, Roemer C, Hodcroft EB, Neher RA. Nextclade: clade assignment, mutation calling and quality control for viral genomes. J Open Source Softw. 2021;6:3773.

**TABLES**

Table S1: Details of the SARS-CoV-2 genome sequences obtained from GISAID. (separate file)

Table S2: Tajima’s D estimate for the various clusters identified using DAPC. Values in bold indicate significant demographic expansion or selection.

| Cluster ID | Number of sequences | Number of segregating sites | Theta | Nucleotide diversity | Tajima’s D |
| --- | --- | --- | --- | --- | --- |
| BA.1 lineage | | | | | |
| Cluster 1 | 41 | 16 | 0.00293 | 0.00071 | **-2.425328** |
| Cluster 2 | 12 | 7 | 0.00182 | 0.00095 | -1.866946 |
| Cluster 3 | 40 | 11 | 0.00203 | 0.00073 | -1.948315 |
| Cluster 4 | 16 | 8 | 0.00189 | 0.00105 | -1.598913 |
| Cluster 5 | 1 | NA | NA | NA | NA |
| BA.2 lineage | | | | | |
| Cluster 1 | 0 | NA | NA | NA | NA |
| Cluster 2 | 61 | 8 | 0.001343 | 0.00028 | **-2.08083** |
| Cluster 3 | 0 | NA | NA | NA | NA |
| Cluster 4 | 31 | 2 | 0.000393 | 0.00010 | -1.50558 |
| Cluster 5 | 207 | 19 | 0.002527 | 0.00032 | **-2.31247** |
| Cluster 6 | 632 | 50 | 0.005591 | 0.00027 | **-2.59423** |
| Cluster 7 | 40 | 3 | 0.000554 | 0.00019 | -1.4309 |
| Cluster 8 | 22 | 1 | 0.000215 | 0.00007 | -1.16240 |
| Cluster 9 | 8 | 1 | 0.000303 | 0.00020 | -1.05482 |
| Cluster 10 | 44 | 3 | 0.000542 | 0.00027 | -1.07839 |

**FIGURES**

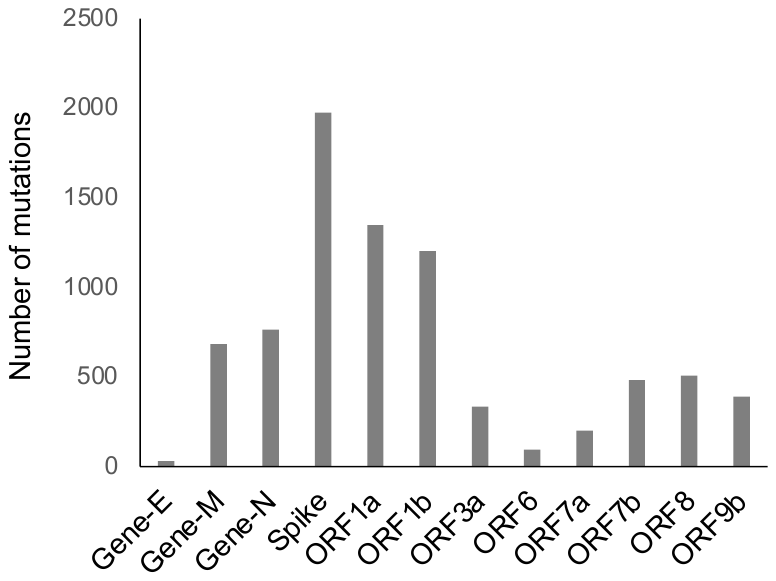

Figure S1: Distribution of the number of mutations for each open reading frame of SARS-CoV-2 sequences belonging to omicron lineage analyzed in this study. We characterized the mutation in reference to the Wuhan genome.

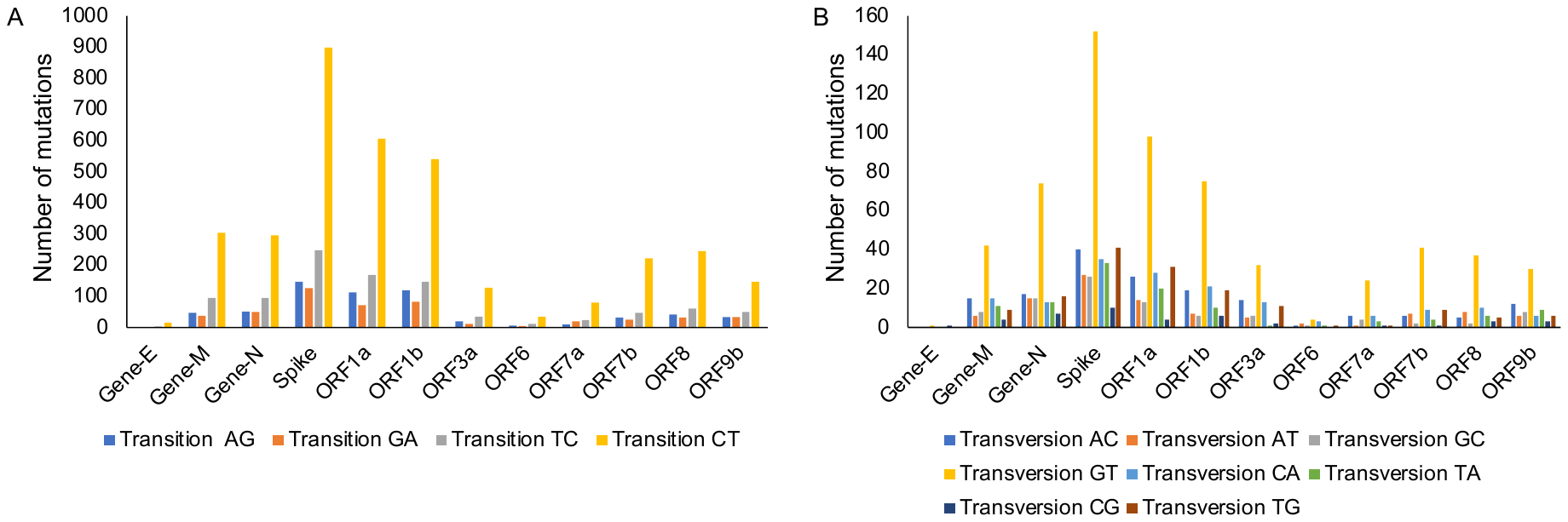

Figure S2: Frequency distribution of the number of A) transitions, and B) transversion for each open reading frame of SARS-CoV-2 sequences belonging to omicron lineage analyzed in this study. We characterized the mutation in reference to the Wuhan genome.
