## Supplementary Table S1 for "Population genomic insights into the evolution of the SARS-CoV-2 Omicron variant"

We gratefully acknowledge the following Authors from the Originating laboratories responsible for obtaining the specimens, as well as the Submitting laboratories where the genome data were generated and shared via GISAID, on which this research is based.

All Submitters of data may be contacted directly via www.gisaid.org

Authors are sorted alphabetically.

| Accession ID | Originating Laboratory | Submitting Laboratory | Authors |
|---|---|---|---|
| EPI_ISL_7543686 | 2 Military Hospital wc MAA | NHLS/UCT | Arash Iranzadeh; Bruna Galvao; Carolyn Williamson; Deelan Doolabh; Diana Hardie; Gert Marais; Innocent Mudau; Luicer Olubayo; Lynn Tyers; Marvin Hsiao; Nokuzola Mbhele; Rageema Joseph; Stephen Korsman |
| EPI_ISL_7987418 | 4Cyte Pathology | NSW Health Pathology - Institute of Clinical Pathology and Medical Research; Westmead Hospital; University of Sydney | Arnott A.; Draper J.; Gall M.; Martinez E.; Rockett R.; Sintchenko V.; on behalf of ICPMR |
| EPI_ISL_7929179 | A. Krumbholz, Labor Dr. Krause und Kollegen MVZ GmbH, Kiel | Charité Universitätsmedizin Berlin, Institut für Virologie | Andi Krumbholz; Barbara Mühlemann; Christian Drosten; Julia Schneider; Julia Tesch; Jörn Beheim-Schwarzbach; Talitha Veith; Terry Jones; Tobias Bleicker; Victor M Corman |
| EPI_ISL_8077636, EPI_ISL_8078464, EPI_ISL_8149646, EPI_ISL_8149727, EPI_ISL_8150203 | Aegis Sciences Corporation | Centers for Disease Control and Prevention Division of Viral Diseases, Pathogen Discovery | Alec Vest; Benjamin Rambo-Martin; Christopher Gulvick; Clinton Paden; Cyndi Clark; Dakota Howard; Dhwani Batra; Dillon Nall; Duncan MacCannell; Erisa Sula; Ethan Sanders; Holly Houdeshell; Jason Caravas; Kristine Lacek; Matthew Hardison; Matthew Schmerer; Ola Kvalvaag; Patrick Campbell; Peter Cook; Rob Case; Scott Sammons; Shatavia Morrison; Shaun Westlund; Tymeckia Kendall; Victoria Caban Figueroa; Vikramsinha Ghorpade; Yvette Unoarumhi |
| EPI_ISL_8056916 | Alfa Health Care Diagnostic Center | National Public Health Laboratory | National Public Health Laboratory Team/ Alfa Health Care Diagnosyic Center |
| EPI_ISL_8015285 | Analytica | Institute of Medical Virology, University of Zurich | Alexandra Trkola; Annette Audigé; Catharine Aquino; Cyril Shah; Daniel Ehrsam; Gabriela Ziltener; Guido Bloemberg; Hubert Rehrauer; Isabel Stürmer; Joel Wirz; Jon Huder; Jürg Böni; Kevin Steiner; Maria Grünberg; Maryam Zaheri; Michael Huber; Riccarda Capaul; Stefan Schmutz; Verena Kufner; Weihong Qi |
| EPI_ISL_8132322 | Arcispedale Santa Maria Nuova Autoimmunita Allergologia e Biotecnologie Innovative"" | Istituto Zooprofilattico Sperimentale della Lombardia e dell'Emilia Romagna (IZSLER), Risk Analysis and Genomic Epidemiology Unit | Erika Scaltriti; Ilaria Menozzi; Marina Morganti; Stefano Pongolini |
| EPI_ISL_7616000 | Ayass Bioscience LLC | Ayass Bioscience LLC | Kevin Zhu; Lina Abi Mosleh; Mohamad Ammar Ayass; Natalya Griko; Nazanin Taheri |
| EPI_ISL_8132350, EPI_ISL_8132365 | Azienda Ospedaliero - Universitaria di Modena Policlinico - Virologia e Microbiologia Molecolare"" | Istituto Zooprofilattico Sperimentale della Lombardia e dell'Emilia Romagna (IZSLER), Risk Analysis and Genomic Epidemiology Unit | Erika Scaltriti; Ilaria Menozzi; Marina Morganti; Stefano Pongolini |
| EPI_ISL_8132317, EPI_ISL_8134455 | Azienda Sanitaria Locale di Piacenza - Presidio Ospedaliero - Laboratorio di Microbiologia"" | Istituto Zooprofilattico Sperimentale della Lombardia e dell'Emilia Romagna (IZSLER), Risk Analysis and Genomic Epidemiology Unit | Erika Scaltriti; Ilaria Menozzi; Marina Morganti; Stefano Pongolini |
| EPI_ISL_7196120, EPI_ISL_7196121 | Bidh Lab | National Public Health Laboratory | Bidh Laboartory; National Public Health Laboratory Team |
| EPI_ISL_7566330, EPI_ISL_7566360 | Botswana Harvard HIV Reference Laboratory | Botswana Harvard AIDS Institute Partnership, Plot 1836 North Ring Road, Princess Marina Hospital, Gaborone | Boitumelo Zuze; Botshelo Radibe; Dorcas Maruapula; Doreen Ditshwanelo; Joseph Makhema; Keoratile Ntshambiwa; Kgomotso Moruisi; Legodile Kooepile; Mosepele Mosepele; Mphaphi B. Mbulawa; Ontlametse T. Bareng; Pamela Smith-Lawrence; Roger Shapiro; Sefetogi Ramaologa; Shahin Lockman; Sikhulile Moyo; Simani Gaseitsiwe; Thongbotho Mphoyakgosi; Wonderful T. Choga |
| EPI_ISL_7834399, EPI_ISL_7834407, EPI_ISL_7834409, EPI_ISL_7834416, EPI_ISL_7834419, EPI_ISL_7834422, EPI_ISL_7834424, EPI_ISL_7834425, EPI_ISL_7834426, EPI_ISL_7834432, EPI_ISL_7834433, EPI_ISL_7834434, EPI_ISL_7834437, EPI_ISL_7834438, EPI_ISL_7834439, EPI_ISL_7834440, EPI_ISL_7834441, EPI_ISL_7834442, EPI_ISL_7834443, EPI_ISL_7834444, EPI_ISL_7834446, EPI_ISL_7834447, EPI_ISL_7834449, EPI_ISL_7834451, EPI_ISL_7834452, EPI_ISL_7834453, EPI_ISL_7834455, EPI_ISL_7834456, EPI_ISL_7834457, EPI_ISL_7834458, EPI_ISL_7834459, EPI_ISL_7834461, EPI_ISL_7834462, EPI_ISL_7834463, EPI_ISL_7834465, EPI_ISL_7834466, EPI_ISL_7834467, EPI_ISL_7834468, EPI_ISL_7834472, EPI_ISL_7834473, EPI_ISL_7834475, EPI_ISL_7834476, EPI_ISL_7834477, EPI_ISL_7834480, EPI_ISL_7834481, EPI_ISL_7834482, EPI_ISL_7834493, EPI_ISL_7834502, EPI_ISL_7834514, EPI_ISL_7834555, EPI_ISL_7834590, EPI_ISL_7834591, EPI_ISL_7834600 | | | |
| see above | Botswana Harvard HIV Reference Laboratory | Botswana Harvard HIV Reference Laboratory | Boitumelo Zuze; Botshelo Radibe; Dorcas Maruapula; Joseph Makhema; Keoratile Ntshambiwa; Kgomotso Moruisi; Legodile Kooepile; Mosepele Mosepele; Mphaphi B. Mbulawa; Ontlametse T. Bareng; Pamela Smith-Lawrence; Roger Shapiro; Sefetogi Ramaologa; Shahin Lockman; Sikhulile Moyo; Simani Gaseitsiwe; Thongbotho Mphoyakgosi; Wonderful T. Choga |
| EPI_ISL_7747509 | CAPRISA_HILLCREST | CERI, Centre for Epidemic Response and Innvoation, Stellenbosch University and KRISP, KZN Research Innovation and Sequencing Platform, UKZN. | Aida Sivro; Arisha Maharaj; Giandhari J; Naidoo Y; Natasha Samsunder; Pillay S; Ramphal U; Ramphal Y; San JE; Tegally H; Tshiabuila D; Wilkinson E; de Oliveira T |
| EPI_ISL_8109017 | CHC Dakor | Gujarat Biotechnology Research Centre | Akhilesh Modi; Apurvasinh Puvar; Bhadreshsinh Gohil; Chaitanya Joshi; Disha Vora; Janvi Raval; Jaykumar Rangani; Madhvi Joshi; Nimesh Patel; Nitin Savaliya; Nitin Shukla; Priyank Chavda; Ramesh Pandit; Roshani Mishra; Sonal Sharma; Tasnim Trivedi; Zarna Patel |
| EPI_ISL_7337481, EPI_ISL_7337497 | Charlotte Maxeke Johannesburg Academic Hospital | National Institute for Communicable Diseases of the National Health Laboratory Service | Amoako DG; Bhiman JN; Everatt J; Ismail A; Mahlangu B; Mnguni A; Mohale T; Ntuli N; Scheepers C |
| EPI_ISL_7988984, EPI_ISL_7988985, EPI_ISL_8099510 | Clinical Microbiology Laboratory, Tel Aviv Sourasky Medical Center | Clinical Microbiology Laboratory, Tel Aviv Sourasky Medical Center | Alon Ziv; Amos Adler; Katya Levytskyi; Lior Handler; Ora Halutz |
| EPI_ISL_7829627 | Clinique Saint-Pierre Ottignies | UCLouvain/IREC/MBLG-CTMA | Benoit Kabamba Mukadi; Bertrand Bearzatto; Jean-Luc Gala; Nicolas Pinte; Paul Blanpain; Simon Ophélie; Valentin Coste |
| EPI_ISL_7829625, EPI_ISL_7829626, EPI_ISL_7829631 | Cliniques universitaires Saint-Luc | UCLouvain/IREC/MBLG-CTMA | Benoit Kabamba Mukadi; Bertrand Bearzatto; Jean-Luc Gala; Nicolas Pinte; Paul Blanpain; Simon Ophélie; Valentin Coste |
| EPI_ISL_8122388 | Colorado Department of Public Health and Environment | Colorado Department of Public Health and Environment | Alexandria Rossheim; Diana Ir; Emily A. Travanty; Laura Bankers; Mandy Waters; Michael A. Martin; Molly C. Hetherington-Rauth; Sarah Elizabeth Totten; Shannon R. Matzinger |
| EPI_ISL_7906576, EPI_ISL_8002167, EPI_ISL_8002210, EPI_ISL_8053581, EPI_ISL_8053663, EPI_ISL_8053917, EPI_ISL_8134681, EPI_ISL_8135005, EPI_ISL_8135069, EPI_ISL_8135113, EPI_ISL_8135280, EPI_ISL_8135299, EPI_ISL_8135305, EPI_ISL_8135486, EPI_ISL_8135495, EPI_ISL_8135593, EPI_ISL_8135685, EPI_ISL_8135710, EPI_ISL_8135886, EPI_ISL_8136001, EPI_ISL_8136019, EPI_ISL_8136050, EPI_ISL_8136091, EPI_ISL_8136635, EPI_ISL_8136749, EPI_ISL_8136811, EPI_ISL_8136950, EPI_ISL_8137224, EPI_ISL_8137308, EPI_ISL_8137566, EPI_ISL_8137955, EPI_ISL_8138431, EPI_ISL_8138663, EPI_ISL_8138852, EPI_ISL_8139086, EPI_ISL_8139268 | | | |
| see above | Department of Bacteria, Parasites and Fungi, Statens Serum Institut, Copenhagen, Denmark | Statens Serum Institut Bioinformatics and Microbial Genomics | Danish Covid-19 Genome Consortium |
| EPI_ISL_8020345 | Department of Clinical Microbiology | GIGA Medical Genomics | Bouchra Boujemla; Claire Gourzonès; Cécile Meex; Keith Durkin; Laurent Gillet; Maria Artesi; Marie-Pierre Hayette; Nadine Cambisano; Nathalie Renotte; Olivier Ek; Sébastien Bontems; Vincent Bours |
| EPI_ISL_7910635 | Department of Clinical Microbiology and Center for Genomic Medicine, Rigshospitalet, Copenhagen, Denmark | Statens Serum Institut Bioinformatics and Microbial Genomics | Danish Covid-19 Genome Consortium |
| EPI_ISL_7192734 | Department of Health Technology and Informatics, The Hong Kong Polytechnic University | Department of Health Technology and Informatics, The Hong Kong Polytechnic University | Alan Ka-Lun Wu; Alex Yat-Man Ho; Barry Kin-Chung Wong; Chloe Toi-Mei Chan; David Ho-Keung Shum; Denise Sze-Hang Wong; Gilman Kit-Hang Siu; Hiu-Yin Lao; Hoi-Ching Jim; Ivan Tak-Fai Wong; Jake Siu-Lun Leung; Kam-Tong Yip; Kenneth Siu-Sing Leung; Kingsley King-Gee Tam; Kitty Sau-Chun Fung; Kristine Luk; Lam-Kwong Lee; Miranda Chong-Yee Yau; Sandy Ka-Yee Chau; Shea Ping Yip; Tak-Lun Que; Timothy Ting-Leung Ng; Wing Cheong Yam; Wing-Hei Lo; Wing-Kin To; Yvette Wai-Man Lai |
| EPI_ISL_7623676 | Dept of Microbilogy, Maulana Azad Medical College | J6094_MAMC_LNH | Avinash Lomash; Mohammad Faruq; Oves Siddiqui; SCOG_MAMC_LNH; Seema Kapoor. Sunil Kumar Polipalli; Somesh Kumar; Suresh Kumar |
| EPI_ISL_8063678 | Fondazione IRCCS Ca' Granda Ospedale Maggiore Policlinico | Fondazione IRCCS Ca' Granda Ospedale Maggiore Policlinico | Ferruccio Ceriotti; Sara Uceda Renteria |
| EPI_ISL_8060345, EPI_ISL_8060974, EPI_ISL_8080063, EPI_ISL_8091398, EPI_ISL_8157487 | Fulgent Genetics | Centers for Disease Control and Prevention Division of Viral Diseases, Pathogen Discovery | Becky Tsai; Benafsh Sapra; Benjamin Rambo-Martin; Christopher Gulvick; Clinton Paden; Dakota Howard; Dhwani Batra; Doreen Ng; Duncan MacCannell; Erisa Sula; Harry Gao; James Xie; Jason Caravas; John Gao; Joseph Fierro; Kristine Lacek; Matthew Schmerer; Mickey Li; Peter Cook; Scott Sammons; Shatavia Morrison; Tymeckia Kendall; Victoria Caban Figueroa; Yan Meng; Yvette Unoarumhi |
| EPI_ISL_8020172 | Gesundheitsdirektion Zürich | Institute of Medical Virology, University of Zurich | Alexandra Trkola; Annette Audigé; Catharine Aquino; Cyril Shah; Daniel Ehrsam; Gabriela Ziltener; Guido Bloemberg; Hubert Rehrauer; Isabel Stürmer; Joel Wirz; Jon Huder; Jürg Böni; Kevin Steiner; Maria Grünberg; Maryam Zaheri; Michael Huber; Riccarda Capaul; Stefan Schmutz; Verena Kufner; Weihong Qi |
| EPI_ISL_7740761 | HELEN JOSEPH LABORATORY | National Institute for Communicable Diseases of the National Health Laboratory Service | Amoako DG; Bhiman JN; Everatt J; Ismail A; Mahlangu B; Mnguni A; Mohale T; Ntuli N; Scheepers C; Wolter N |
| EPI_ISL_7337441 | Helen Joseph Laboratory | National Institute for Communicable Diseases of the National Health Laboratory Service | Amoako DG; Bhiman JN; Everatt J; Ismail A; Mahlangu B; Mnguni A; Mohale T; Ntuli N; Scheepers C |
| EPI_ISL_7265104, EPI_ISL_7749559 | Histopath | NSW Health Pathology - Institute of Clinical Pathology and Medical Research; Westmead Hospital; University of Sydney | Arnott A.; Draper J.; Gall M.; Martinez E.; Rockett R.; Sintchenko V.; on behalf of ICPMR |
| EPI_ISL_8002519 | Hospital General Universitario Gregorio Marañón | Hospital General Universitario Gregorio Marañón | Cristina Rodriguez-Grande; Darío García de Viedma; Jorge Rodríguez-Grande; Julia Suárez; Laura Pérez-Lago; Marta Herranz Martin; Patricia Muñoz; Pedro Sola Campoy; Pilar Catalán; Sergio Buenestado Serrano |
| EPI_ISL_7734081, EPI_ISL_7977800, EPI_ISL_7978212, EPI_ISL_7978248, EPI_ISL_7978362, EPI_ISL_7978363, EPI_ISL_7978635, EPI_ISL_7978638, EPI_ISL_7979184, EPI_ISL_7979535, EPI_ISL_8030372 | | | |
| see above | Houston Methodist Hospital | Houston Methodist Hospital | Akanksha Batajoo; Ilya J. Finkelstein; James J. Davis; Jessica Cambric; Jimmy Gollihar; Kristina Reppond; Layne Pruitt; Madison N. Shyer; Marcus Nguyen; Matthew Ojeda Saavedra; Paul A. Christensen; Prasanti Yerramilli; Randall J. Olsen; Rashi Thakur; Regan Mangham; Richard Snehall; Robert Olson; Ryan Gadd; S. Wesley Long; Sindy Pena; Sishir Subedi; and James M. Musser |
| EPI_ISL_8119611 | IZSM-Virologia | Istituto Zooprofilattico Sperimentale del Mezzogiorno - Unità Operativa Complessa di Virologia | Antonio Limone; Claudio de Martinis; Ester De Carlo; Giovanna Fusco; Loredana Cozzolino; Lorena Cardillo; Maurizio Viscardi |
| EPI_ISL_7496734 | Idaho Bureau of Laboratories | Idaho Bureau of Laboratories | Aimee Ceniseros; Christian Loera; Christopher Ball; Matthew Charles Burns; R. Beukelman; Robert L. Voermans |
| EPI_ISL_7381102 | Indian Council of Medical Research-National Institute of Virology, Microbial Containment Complex | Indian Council of Medical Research-National Institute of Virology, Microbial Containment Complex | Pragya D. Yadav |
| EPI_ISL_8100211, EPI_ISL_8100260 | Institute of Microbiology and Immunology, | Institute of Microbiology and Immunology, Faculty of | Alen Suljič; Andraž Celar; Domen Lazar; Doroteja Vlaj; Mario Poljak; Miša Korva; Patricija Pozvek; Samo Zakotnik; Tatjana Avšič – Županc; Tina Gabrovšek; Tina Živič; Tomaž Mark Zorec; Špela Pleh |

| | | | |
|---|---|---|---|
| | Faculty of Medicine, University of Ljubljana | Medicine, University of Ljubljana | |
| EPI_ISL_7955281 | Integrated Covid Hub North East | Wellcome Sanger Institute for the COVID-19 Genomics UK (COG-UK) Consortium | Cordelia Langford; David K. Jackson; Dominic Kwiatkowski; Ewan Harrison; Ian Johnston; Integrated Covid Hub North East and Alex Alderton; Jeffrey Barrett; John Sillitoe on behalf of the Wellcome Sanger Institute COVID-19 Surveillance Team; Roberto Amato; Sonia Goncalves |
| EPI_ISL_7632052, EPI_ISL_8015288, EPI_ISL_8120111 | Invenimus AG | Institute of Medical Virology, University of Zurich | Alexandra Trkola; Annette Audigé; Catharine Aquino; Cyril Shah; Daniel Ehrsam; Gabriela Ziltener; Guido Bloemberg; Hubert Rehrauer; Isabel Stürmer; Joel Wirz; Jon Huder; Jürg Böni; Kevin Steiner; Maria Grünberg; Maryam Zaheri; Michael Huber; Riccarda Capaul; Stefan Schmutz; Verena Kufner; Weihong Qi |
| EPI_ISL_7789703, EPI_ISL_7790175, EPI_ISL_7982412 | KU Leuven, Rega Institute, Clinical and Epidemiological Virology | KU Leuven, Rega Institute, Clinical and Epidemiological Virology | Bert Vanmechelen; Joan Marti-Carerras; Piet Maes; Tony Wawina-Bokalanga |
| EPI_ISL_8015322, EPI_ISL_8015323, EPI_ISL_8119804 | Kantonsspital Aarau | Institute of Medical Virology, University of Zurich | Alexandra Trkola; Annette Audigé; Catharine Aquino; Cyril Shah; Daniel Ehrsam; Gabriela Ziltener; Guido Bloemberg; Hubert Rehrauer; Isabel Stürmer; Joel Wirz; Jon Huder; Jürg Böni; Kevin Steiner; Maria Grünberg; Maryam Zaheri; Michael Huber; Riccarda Capaul; Stefan Schmutz; Verena Kufner; Weihong Qi |
| EPI_ISL_8015340, EPI_ISL_8015409, EPI_ISL_8120125 | Kantonsspital Winterthur | Institute of Medical Virology, University of Zurich | Alexandra Trkola; Annette Audigé; Catharine Aquino; Cyril Shah; Daniel Ehrsam; Gabriela Ziltener; Guido Bloemberg; Hubert Rehrauer; Isabel Stürmer; Joel Wirz; Jon Huder; Jürg Böni; Kevin Steiner; Maria Grünberg; Maryam Zaheri; Michael Huber; Riccarda Capaul; Stefan Schmutz; Verena Kufner; Weihong Qi |
| EPI_ISL_8097223, EPI_ISL_8097234, EPI_ISL_8097249, EPI_ISL_8097255, EPI_ISL_8097263, EPI_ISL_8097265, EPI_ISL_8097268, EPI_ISL_8097272, EPI_ISL_8097273, EPI_ISL_8097276, EPI_ISL_8097279, EPI_ISL_8097282 | | | |
| see above | Kasane Primary Hospital Laboratory | Botswana Harvard HIV Reference Laboratory | Agnes Karutwaeng; Boitumelo Zuze; Botshelo Radibe; Dorcas Maruapula; Joseph Makhema; Keoratile Ntshambiwa; Legodile Kooepile; Mosepele Mosepele; Mphapi B. Mbulawa; Ontlametse T. Bareng; Roger Shapiro; Rose Munyere; Sefetogi Ramaologa; Shahin Lockman; Sikhulile Moyo; Simani Gaseitsiwe; Thela Tefelo; Thongbotho Mphoyakgosi; Wonderful T. Choga |
| EPI_ISL_7986436, EPI_ISL_7986439 | Kasturba Hospital Molecular Lab - INSACOG | Kasturba hospital WGS | Jayanthi Shastri; Vidushi Chitalia |
| EPI_ISL_7701129 | LANCET LABORATORY | National Institute for Communicable Diseases of the National Health Laboratory Service | Amoako DG; Bhiman JN; Everatt J; Ismail A; Mahlangu B; Mnguni A; Mohale T; Ntuli N; Scheepers C; Wolter N |
| EPI_ISL_7651289 | Laboratory of Molecular Biology and Cancer Immunology, Faculty of Sciences, Lebanese University | Microbial Pathogenomics Lab - LAU | Alissar Zaghlout; Bassam Badran; Fadi Abdel Sater; Georgi Merhi; Jad Koweyes; Nada Ghosn; Rawan Makki; Sima tokajian |
| EPI_ISL_7834554 | Lancet Laboratories Gaborone | Botswana Harvard HIV Reference Laboratory | Boitumelo Zuze; Botshelo Radibe; Dorcas Maruapula; Ellen Nkitse; Joseph Makhema; Keoratile Ntshambiwa; Kgomotso Moruisi; Legodile Kooepile; Leyi Ncube; Mosana Moyo; Mosepele Mosepele; Mphaphi B. Mbulawa; Ontlametse T. Bareng; Pamela Smith-Lawrence; Roger Shapiro; Sefetogi Ramaologa; Shahin Lockman; Sikhulile Moyo; Simani Gaseitsiwe; Thongbotho Mphoyakgosi; Wonderful T. Choga |
| EPI_ISL_7682986, EPI_ISL_7682990 | Lighthouse Lab in Glasgow | Wellcome Sanger Institute for the COVID-19 Genomics UK (COG-UK) Consortium | Anna Dominiczak and Alex Alderton; Carol Clugston; Cordelia Langford; David Gray; David K. Jackson; Dominic Kwiatkowski; Ewan Harrison; Harper VanSteenhouse; Ian Johnston; Jeffrey Barrett; John Sillitoe on behalf of the Wellcome Sanger Institute COVID-19 Surveillance Team; Roberto Amato; Sonia Goncalves; Yumi Kasai |
| EPI_ISL_7864703 | Lok Kayak Hospital | Lok Kayak Hospital | Avinash Lomash; Mohammed FARUQ; Oves Siddiqui; Prashanth Suravajhala SURESH Kumar; Somesh Kumar; Sunil k Polipalli; seema Kapoor and SCOG_MAMC_LNH Lab |
| EPI_ISL_7877006, EPI_ISL_7877026, EPI_ISL_7877093, EPI_ISL_7877115, EPI_ISL_7877191, EPI_ISL_7877201, EPI_ISL_7877202, EPI_ISL_7877203, EPI_ISL_7877297, EPI_ISL_7889640, EPI_ISL_7889641 | | | |
| see above | Lok Nayak Hospital | Lok Nayak Hospital | ; Avinash Lomash; Lok Nayak Hospital; Mohammed FARUQ; Oves Siddiqui; Prashanth Suravajhala; SURESH Kumar; Somesh Kumar; Sunil k Polipalli; seema Kapoor and Genome Sequencing Lab |
| EPI_ISL_7876997 | Lok Nayak Hospital, Delhi | Lok Nayak Hospital, metro Station Central, Jawaharlal Nehru Marg, Near Delhi Gate, New Delhi, Delhi 110002 | Avinash Lomash; Lok Nayak Hospital; Mohammed FARUQ; Oves Siddiqui; Prashanth Suravajhala; SURESH Kumar; Somesh Kumar; Sunil k Polipalli; seema Kapoor and Genome Sequencing Lab |
| EPI_ISL_8123968 | MS PHL | MS PHL | Beata Karowlewicz; Daphne Ware |
| EPI_ISL_7433660 | Medizinisch-Diagnostisches Labor Kempten allgäulab | Robert Koch Institute | |
| EPI_ISL_7620911 | Medlab Pathology | NSW Health Pathology - Institute of Clinical Pathology and Medical Research; Westmead Hospital; University of Sydney | Arnott A.; Draper J.; Gall M.; Martinez E.; Rockett R.; Sintchenko V.; on behalf of ICPMR |
| EPI_ISL_6854346, EPI_ISL_6854347, EPI_ISL_6854348 | Microbiologia e Virologia Cotugno | Microbiologia e Virologia Cotugno | Antonio Canonico; Antonio Fascione; Claudia Tiberio; Enza Mallardo; Francesco Nappo; Giovanni D'Auria; Giuseppe di Gennaro; Ilaria Cavallaro; Luigi Atripaldi |
| EPI_ISL_7552702, EPI_ISL_7552703, EPI_ISL_7552707 | National Health Laboratory | Botswana Harvard AIDS Institute Partnership, Plot 1836 North Ring Road, Princess Marina Hospital, Gaborone | Boitumelo Zuze; Botshelo Radibe; Dorcas Maruapula; Doreen Ditshwanelo; Joseph Makhema; Keoratile Ntshambiwa; Kgomotso Moruisi; Legodile Kooepile; Mosepele Mosepele; Mphaphi B. Mbulawa; Ontlametse T. Bareng; Pamela Smith-Lawrence; Roger Shapiro; Sefetogi Ramaologa; Shahin Lockman; Sikhulile Moyo; Simani Gaseitsiwe; Thongbotho Mphoyakgosi; Wonderful T. Choga |
| EPI_ISL_7545782, EPI_ISL_7747500 | National Health Laboratory Service, Kwazulu-Natal, South Africa | CERI, Centre for Epidemic Response and Innvoation, Stellenbosch University and KRISP, KZN Research Innovation and Sequencing Platform, UKZN. | Arisha Maharaj; Giandhari J; Naidoo Y; Nokukhanya Mdlalose; Pillay S; Ramphal U; Ramphal Y; San JE; Tegally H; Tshiabuila D; Wilkinson E; de Oliveira T |
| EPI_ISL_7969370 | National Platform bis UMONS/Jolimont | National Platform bis UMONS/Jolimont | Eric Tarantino; Florian Juszczak; Gautier Detry; Guillaume Bayon-Vicente; Laetitia Gheysen; Ruddy Wattiez |
| EPI_ISL_8092780, EPI_ISL_8092783, EPI_ISL_8112317, EPI_ISL_8112348 | National Public Health Laboratory, National Centre for Infectious Diseases | National Public Health Laboratory, National Centre for Infectious Diseases | Benny Yeo; Katherine Ching; Lin Cui; Raymond Tzer Pin Lin; Royce Ang; Samuel Loo; Zhenyang Zhou |
| EPI_ISL_8065975 | Noguchi Memorial Institute for Medical Research, University of Ghana, Legon, Ghana | Noguchi Memorial Institute for Medical Research, University of Ghana, Legon, Ghana | Bright Adu; Hilda Opoku Frempong; John K. Odoom; Joseph K. H. Bonney; Joyce Appiah-Kubi; Keren O. Attiku; Mohktar; Peace Uche; Quaneeta; Seyram B. Avonyo |
| EPI_ISL_8097205 | Nyanyangbwe HIV Reference Laboratory | Botswana Harvard HIV Reference Laboratory | Boitumelo Zuze; Botshelo Radibe; David Matema; Dorcas Maruapula; Joseph Makhema; Keoratile Ntshambiwa; Legodile Kooepile; Lindani Bochena; Mosepele Mosepele; Mphapi B. Mbulawa; Ontlametse T. Bareng; Roger Shapiro; Sefetogi Ramaologa; Shahin Lockman; Sikhulile Moyo; Simani Gaseitsiwe; Thela Tefelo; Thongbotho Mphoyakgosi; Wonderful T. Choga |
| EPI_ISL_7552440, EPI_ISL_7799914, EPI_ISL_7799945, EPI_ISL_8083804, EPI_ISL_8083805, EPI_ISL_8083815, EPI_ISL_8083816, EPI_ISL_8083822, EPI_ISL_8083823, EPI_ISL_8083825, EPI_ISL_8083826, EPI_ISL_8083830, EPI_ISL_8083832, EPI_ISL_8083833, EPI_ISL_8083835, EPI_ISL_8083836, EPI_ISL_8083839, EPI_ISL_8083843, EPI_ISL_8083844, EPI_ISL_8083846, EPI_ISL_8083850, EPI_ISL_8083852, EPI_ISL_8083853, EPI_ISL_8083854, EPI_ISL_8083855, EPI_ISL_8083860, EPI_ISL_8083864, EPI_ISL_8083867, EPI_ISL_8083868, EPI_ISL_8083870, EPI_ISL_8083872, EPI_ISL_8083873, EPI_ISL_8083874, EPI_ISL_8083875, EPI_ISL_8083876, EPI_ISL_8083878, EPI_ISL_8083879, EPI_ISL_8083880, EPI_ISL_8083882, EPI_ISL_8083884, EPI_ISL_8083888, EPI_ISL_8083889, EPI_ISL_8083890, EPI_ISL_8083894, EPI_ISL_8083896, EPI_ISL_8083900, EPI_ISL_8083901, EPI_ISL_8083902, EPI_ISL_8083905, EPI_ISL_8083906, EPI_ISL_8083907, EPI_ISL_8083908, EPI_ISL_8083909 | | | |
| see above | Ochsner Health | BioInfoExperts | Amy Feehan; Ben Lain; Chris Huston; David J. Nolan; Judy Crabtree; Julia-Garcia-Diaz; Lucio Miele; Rebecca Rose; Samual Moot; Susanna L. Lamers; Tessa LaFleur |
| EPI_ISL_8142222 | P.D.U Medical College Rajkot | Gujarat Biotechnology Research Centre | Akhilesh Modi; Apurvasinh Puvar; Bhadreshsinh Gohil; Chaitanya Joshi; Disha Vora; Janvi Raval; Jaykumar Rangani; Madhvi Joshi; Madhvlika Mistry; Nimesh Patel; Nitin Savaliya; Nitin Shukla; Priyank Chavda; Ramesh Pandit; Roshani Mishra; Sonal Sharma; Tasnim Trivedi; Zarna Patel |
| EPI_ISL_7190366, EPI_ISL_8132552, EPI_ISL_8132660, EPI_ISL_8132661, EPI_ISL_8132662, EPI_ISL_8132663, EPI_ISL_8132665, EPI_ISL_8132667, EPI_ISL_8132668, EPI_ISL_8132669, EPI_ISL_8132674, EPI_ISL_8132677, EPI_ISL_8132678, EPI_ISL_8132681, EPI_ISL_8132683, EPI_ISL_8132686, EPI_ISL_8132687, EPI_ISL_8132689, EPI_ISL_8132690, EPI_ISL_8132691, EPI_ISL_8132692, EPI_ISL_8132693, EPI_ISL_8132694, EPI_ISL_8132696, EPI_ISL_8132697, EPI_ISL_8132698, EPI_ISL_8132700, EPI_ISL_8132701, EPI_ISL_8132734, EPI_ISL_8132736, EPI_ISL_8132737, EPI_ISL_8132740, EPI_ISL_8132741, EPI_ISL_8132743, EPI_ISL_8132747, EPI_ISL_8132748, EPI_ISL_8132752, EPI_ISL_8132755, EPI_ISL_8132756, EPI_ISL_8132758, EPI_ISL_8132764, EPI_ISL_8132765, EPI_ISL_8132767, EPI_ISL_8132770, EPI_ISL_8132773, EPI_ISL_8132777, EPI_ISL_8132778, EPI_ISL_8132782, EPI_ISL_8132783, EPI_ISL_8132787, EPI_ISL_8132788, EPI_ISL_8132789, EPI_ISL_8132792, EPI_ISL_8132793, EPI_ISL_8132794, EPI_ISL_8132795, EPI_ISL_8132797, EPI_ISL_8132800, EPI_ISL_8132805, EPI_ISL_8132806, EPI_ISL_8132808, EPI_ISL_8132810, EPI_ISL_8132811, EPI_ISL_8132813, EPI_ISL_8132814, EPI_ISL_8132817, EPI_ISL_8132819, EPI_ISL_8132821, EPI_ISL_8132822, EPI_ISL_8132823, EPI_ISL_8132825, EPI_ISL_8132826, EPI_ISL_8132827, EPI_ISL_8132829, EPI_ISL_8132830, EPI_ISL_8132831, EPI_ISL_8132832, EPI_ISL_8132834, EPI_ISL_8132835, EPI_ISL_8132836, EPI_ISL_8132837, EPI_ISL_8132885 | | | |
| see above | PHV-FSS | PHV-FSS | Chenwei Wang on behalf of Q-PHIRE Genomics |
| EPI_ISL_7834572 | Palapye Primary Hospital Laboratory | Botswana Harvard HIV Reference Laboratory | Boitumelo Zuze; Botshelo Radibe; Dorcas Maruapula; Joseph Makhema; Keoratile Ntshambiwa; Kgomotso Moruisi; Legodile Kooepile; Mosepele Mosepele; Mphaphi B. Mbulawa; Ontlametse T. Bareng; Pamela Smith-Lawrence; Roger Shapiro; Sefetogi Ramaologa; Shahin Lockman; Sikhulile Moyo; Simani Gaseitsiwe; Thongbotho Mphoyakgosi; Wonderful T. Choga |
| EPI_ISL_7987968 | Pathology North - Hunter - NSW Health Pathology | NSW Health Pathology - Institute of Clinical Pathology and Medical Research; Westmead Hospital; University of Sydney | Arnott A.; Draper J.; Gall M.; Martinez E.; Rockett R.; Sintchenko V.; on behalf of ICPMR |
| EPI_ISL_8124579 | Plateforme de testing Namuroise | Plateforme de testing Namuroise | Degosserie Jonathan; Demars Aurore; Denis Olivier; Gilliard Nicolas; Maschietto Céline; Mullier François; Nobis Chloé; Otto Gaetan |
| EPI_ISL_8144416 | Premier Laboratory Services | BioInfoExperts | Amy Feehan; Ben Lain; Chris Huston; David J. Nolan; Judy Crabtree; Julia-Garcia-Diaz; Lucio Miele; Rebecca Rose; Samual Moot; Susanna L. Lamers; Tessa LaFleur |
| EPI_ISL_7703322 | Premier Laboratory Services | BioInfoExperts LLC | Cheryl Metzler; David Nolan; Rebecca Rose; Susanna Lamers; Valerie Murray |
| EPI_ISL_7345322, EPI_ISL_7464049, EPI_ISL_7464059, EPI_ISL_7509826, EPI_ISL_7509828, EPI_ISL_7509830, EPI_ISL_7563604, EPI_ISL_7583354, EPI_ISL_7583375, EPI_ISL_7583411, EPI_ISL_7685061, EPI_ISL_7725851, EPI_ISL_7725891, EPI_ISL_7725913, EPI_ISL_7725915, EPI_ISL_7725927, EPI_ISL_7843337, EPI_ISL_7843349, EPI_ISL_7843355, EPI_ISL_7992659, EPI_ISL_8090517 | | | |
| see above | Respiratory Virus Unit, Microbiology Services Colindale, Public Health England | COVID-19 Genomics UK (COG-UK) Consortium | PHE Covid Sequencing Team |
| EPI_ISL_7862534 | Rosalind Franklin Laboratory | Wellcome Sanger Institute for the COVID-19 Genomics UK (COG-UK) Consortium | Cordelia Langford; David K. Jackson; Dominic Kwiatkowski; Donald Fraser; Ewan Harrison; Ian Johnston; Jeffrey Barrett; John Sillitoe on behalf of the Wellcome Sanger Institute COVID-19 Surveillance Team; Rob Howes; Roberto Amato; Sonia Goncalves; Suki Lee; The Rosalind Franklin Laboratory and Alex Alderton |
| EPI_ISL_8112531, EPI_ISL_8112532, EPI_ISL_8112533, EPI_ISL_8112534, EPI_ISL_8112535, EPI_ISL_8112536, EPI_ISL_8112537, EPI_ISL_8112538, EPI_ISL_8112539, EPI_ISL_8112540, EPI_ISL_8112541, EPI_ISL_8112542, EPI_ISL_8112543, EPI_ISL_8112544, EPI_ISL_8112545, EPI_ISL_8112546, EPI_ISL_8112547, EPI_ISL_8112548, EPI_ISL_8112549, EPI_ISL_8112563 | | | |
| see above | Servicio Microbiología Hospital La Paz | Servicio Microbiología Hospital La Paz | Elie Dahdouh; Fernando Lázaro; Iván Bloise; Jesús Mingorance Cruz; Rubén Cáceres |
| EPI_ISL_7625809, EPI_ISL_7896816, EPI_ISL_7896817, EPI_ISL_7896818, EPI_ISL_7896819, EPI_ISL_7896820, EPI_ISL_7896822, EPI_ISL_7896831 | | | |
| see above | Servicio de Microbiologia Hospital Ramon y Cajal | Servicio de Microbiologia Hospital Ramon y Cajal | Galan JC; Martinez L. Abreu M; Ponce M; y Gonzalez-Alba JM |
| EPI_ISL_7632035, EPI_ISL_7632045, EPI_ISL_8020218, EPI_ISL_8020223, EPI_ISL_8020226, EPI_ISL_8119786, EPI_ISL_8119794, EPI_ISL_8119796, EPI_ISL_8119797, EPI_ISL_8119798, EPI_ISL_8119799, EPI_ISL_8119800 | | | |
| see above | Stadtspital Triemli | Institute of Medical Virology, University of Zurich | Alexandra Trkola; Annette Audigé; Catharine Aquino; Cyril Shah; Daniel Ehrsam; Gabriela Ziltener; Guido Bloemberg; Hubert Rehrauer; Isabel Stürmer; Joel Wirz; Jon Huder; Jürg Böni; Kevin Steiner; Maria Grünberg; Maryam Zaheri; Michael Huber; Riccarda Capaul; Stefan Schmutz; Verena Kufner; Weihong Qi |
| EPI_ISL_7445855 | State Hygienic Laboratory at the University of Iowa | State Hygienic Laboratory at the University of Iowa | Alankar Kampoowale; Anna Yakos; Cindy Toll; Davis Rieckenberg; Erik Twait; Jeff Benfer; Kris Eveland; Krishnaveni Sompallae; Kristen Zanon; Mariah Knutson; Mohammed Allam; Valerie Reeb; Wes Hottel |
| EPI_ISL_7621663, EPI_ISL_7621668 | Sydney South West Pathology Service (SSWPS) - Liverpool Hospital - NSW Health Pathology | NSW Health Pathology - Institute of Clinical Pathology and Medical Research; Westmead Hospital; University of Sydney | Arnott A.; Draper J.; Gall M.; Martinez E.; Rockett R.; Sintchenko V.; on behalf of ICPMR |
| EPI_ISL_7830897, EPI_ISL_8128055, EPI_ISL_8131707, EPI_ISL_8131768, EPI_ISL_8131770, EPI_ISL_8131794, EPI_ISL_8131832, EPI_ISL_8131883 | | | |
| see above | UW Virology Lab | UW Virology Lab | Alexander Greninger; Hong Xie; Isabel Arnould; Keith R Jerome; Meei-Li Huang; Nathan Breit; Patrick Mathias; Pavitra Roychoudhury; Pooneh Hajian; Robert J. Livingston; Sean Ellis; Seffir T. Wendm; Shah Mohamed Bakhash |
| EPI_ISL_7696303, EPI_ISL_7983388, EPI_ISL_7983537 | University of Liège COVID-19 testing center | GIGA Medical Genomics | Bouchra Boujemla; Claire Gourzonès; Cécile Meex; Keith Durkin; Laurent Gillet; Maria Artesi; Marie-Pierre Hayette; Nadine Cambisano; Nathalie Renotte; Olivier Ek; Sébastien Bontems; Vincent Bours |

| | | | |
|---|---|---|---|
| EPI_ISL_8092797, EPI_ISL_8092799, EPI_ISL_8092800, EPI_ISL_8092807 | University of Oregon COVID-19 MAP Laboratory | University of Oregon Genomics and Cell Charactericzation Core Facility (GC3F) | Ariana White; Demi Glidden; Douglas Turnbull; Jason Carriere; Jason Sydes; Jeff Bishop; Megan Criss; Peter Batzel; Tina Arredondo |
| EPI_ISL_8015470, EPI_ISL_8015502, EPI_ISL_8020107, EPI_ISL_8119785, EPI_ISL_8119790, EPI_ISL_8119793 | Universität Zürich | Institute of Medical Virology, University of Zurich | Alexandra Trkola; Annette Audigé; Catharine Aquino; Cyril Shah; Daniel Ehrsam; Gabriela Ziltener; Guido Bloemberg; Hubert Rehrauer; Isabel Stürmer; Joel Wirz; Jon Huder; Jürg Böni; Kevin Steiner; Maria Grünberg; Maryam Zaheri; Michael Huber; Riccarda Capaul; Stefan Schmutz; Verena Kufner; Weihong Qi |
| EPI_ISL_8119773, EPI_ISL_8120214 | Universitäts-Kinderspital Zürich | Institute of Medical Virology, University of Zurich | Alexandra Trkola; Annette Audigé; Catharine Aquino; Cyril Shah; Daniel Ehrsam; Gabriela Ziltener; Guido Bloemberg; Hubert Rehrauer; Isabel Stürmer; Joel Wirz; Jon Huder; Jürg Böni; Kevin Steiner; Maria Grünberg; Maryam Zaheri; Michael Huber; Riccarda Capaul; Stefan Schmutz; Verena Kufner; Weihong Qi |
| EPI_ISL_8015421, EPI_ISL_8015430, EPI_ISL_8015435, EPI_ISL_8015487, EPI_ISL_8015488, EPI_ISL_8015517, EPI_ISL_8020132, EPI_ISL_8020177, EPI_ISL_8020180, EPI_ISL_8020186, EPI_ISL_8020187, EPI_ISL_8020196, EPI_ISL_8120107, EPI_ISL_8120108, EPI_ISL_8120109, EPI_ISL_8120112, EPI_ISL_8120113, EPI_ISL_8120198, EPI_ISL_8120199, EPI_ISL_8120200, EPI_ISL_8120201, EPI_ISL_8120203, EPI_ISL_8120204, EPI_ISL_8120205, EPI_ISL_8120206, EPI_ISL_8120207, EPI_ISL_8120211, EPI_ISL_8120221, EPI_ISL_8120222, EPI_ISL_8120224, EPI_ISL_8120226 | | | |
| see above | UniversitätsSpital Zürich | Institute of Medical Virology, University of Zurich | Alexandra Trkola; Annette Audigé; Catharine Aquino; Cyril Shah; Daniel Ehrsam; Gabriela Ziltener; Guido Bloemberg; Hubert Rehrauer; Isabel Stürmer; Joel Wirz; Jon Huder; Jürg Böni; Kevin Steiner; Maria Grünberg; Maryam Zaheri; Michael Huber; Riccarda Capaul; Stefan Schmutz; Verena Kufner; Weihong Qi |
| EPI_ISL_7566875 | Viollier AG | Department of Biosystems Science and Engineering, ETH Zürich | Andrea Patrignani; Andrea Patrizia Salzmann; Andreia Cabral de Gouvea; Catharine Aquino Fournier; Chaoran Chen; Christian Urban; Christiane Beckmann; Christoph Noppen; Daniel Ehrsam; David Dreifuss; Doris Popovic; Franziska Singer; Griffin White; Hai Bui; Henriette Kurth; Isabel Stürmer; Ivan Topolsky; Jay Tracy; Kim Philipp Jablonski; Lara Fuhrmann; Laura Neff; Lennart Opitz; Louis du Plessis; Maria Domenica Moccia; Matteo Carrara; Maurice Redondo; Niko Beerenwinkel; Olivier Kobel; Pelin Icer; Ralph Schlapbach; Sarah Nadeau; Shuqing Yu; Simon Grüter; Tanja Stadler; Timothy Sykes |
| EPI_ISL_7285845, EPI_ISL_7285846 | Virology Laboratory, Scientific Department, Army Medical Center | Virology Laboratory, Scientific Department, Army Medical Center | Anella Monte; Anna Anselmo; Antonella Fortunato; Filippo Molinari; Florigio Lista; Francesco Giordani; Giancarlo Petralito; Giandomenico Cerreto; Giulia Campoli; Lucia Nicosia; Marzia Cavalli; Pietro Marco D'Angelo; Riccardo De Sanctis; Rossella Brandi; Silvia Fillo; Vanessa Vera Fain |
| EPI_ISL_8128502 | ZARV/NHLS, Department Medical Virology, University of Pretoria | CERI, Centre for Epidemic Response and Innvoation, Stellenbosch University and KRISP, KZN Research Innovation and Sequencing Platform, UKZN. | Adriano Mendes; Amy Strydom; Arisha Maharaj; Giandhari J; Micheala Davids; Naidoo Y; Pillay S; Ramphal U; Ramphal Y; San JE; Sim Mayaphi and Marietjie Venter; Tegally H; Tshiabuila D; Wilkinson E; de Oliveira T |
| EPI_ISL_8020197 | Zentrallabor Zürich | Institute of Medical Virology, University of Zurich | Alexandra Trkola; Annette Audigé; Catharine Aquino; Cyril Shah; Daniel Ehrsam; Gabriela Ziltener; Guido Bloemberg; Hubert Rehrauer; Isabel Stürmer; Joel Wirz; Jon Huder; Jürg Böni; Kevin Steiner; Maria Grünberg; Maryam Zaheri; Michael Huber; Riccarda Capaul; Stefan Schmutz; Verena Kufner; Weihong Qi |

We gratefully acknowledge the following Authors from the Originating laboratories responsible for obtaining the specimens, as well as the Submitting laboratories where the genome data were generated and shared via GISAID, on which this research is based.

All Submitters of data may be contacted directly via www.gisaid.org

Authors are sorted alphabetically.

| Accession ID | Originating Laboratory | Submitting Laboratory | Authors |
|---|---|---|---|
| EPI_ISL_8285616 | ADUGODI DISPENSARY | National Center for Biological Sciences, TIFR - Rockefeller Foundation | Awadhesh Pandit; Bhagyashree Madhav Shelar; Darshan Sreenivas; Dimple Notani; Lakshminarayanan CP; Manisha Bharadwaj; Satyajit Mayor; Uma Ramakrishnan |
| EPI_ISL_8307826, EPI_ISL_8308252, EPI_ISL_8311241, EPI_ISL_8311243 | Aegis Sciences Corporation | Centers for Disease Control and Prevention Division of Viral Diseases, Pathogen Discovery | Alec Vest; Benjamin Rambo-Martin; Christopher Gulvick; Clinton Paden; Cyndi Clark; Dakota Howard; Dhwani Batra; Dillon Nall; Duncan MacCannell; Erisa Sula; Ethan Sanders; Holly Houdeshell; Jason Caravas; Kristine Lacek; Matthew Hardison; Matthew Schmerer; Ola Kvalvaag; Patrick Campbell; Peter Cook; Rob Case; Scott Sammons; Shatavia Morrison; Shaun Westlund; Tymeckia Kendall; Victoria Caban Figueroa; Vikramsinha Ghorpade; Yvette Unoarumhi |
| EPI_ISL_8281485, EPI_ISL_8281542, EPI_ISL_8281566 | Allergy, Immunology and Cell Biology Unit (AICBU) | Allergy, Immunology and Cell Biology Unit (AICBU) | Ayesha Wijesinghe; Chandima Jeewandara; Deshni Jayathilaka; Dinuka Ariyaratne; Diyanath Ranasinghe; Dumni Guasinghe; Farha Bary; Gathsaurie Neelika Malavige; Heshan Kuruppu; Tibutius Thanesh |
| EPI_ISL_8274562 | Berkshire and Surrey Pathology Services Lighthouse Laboratory | Wellcome Sanger Institute for the COVID-19 Genomics UK (COG-UK) Consortium | Berkshire and Surrey Pathology Services Lighthouse Laboratory and Alex Alderton; Cordelia Langford; David K. Jackson; Dominic Kwiatkowski; Ewan Harrison; Ian Johnston; Jeffrey Barrett; John Sillitoe on behalf of the Wellcome Sanger Institute COVID-19 Surveillance Team; Roberto Amato; Sonia Goncalves |
| EPI_ISL_8312616 | BioneXt Lab | Laboratoire national de sante, Microbiology, Microbial Genomics Platform | Anke Wienecke-Baldacchino; Catherine Ragimbeau; Elodie Solarino; Fatu Djabi; Jessica Tapp; Lise Pignon; Raoul Salmon; Sibel Berger; Tamir Abdelrahman; Thibault Ferrandon; Virginie Jover |
| EPI_ISL_8294239 | CHTMAD | Instituto Nacional de Saude (INSA) | Borges et al |
| EPI_ISL_8314702 | CHU Purpan - Laboratoire de Virologie - Institut Federatif de Biologie | CHU Purpan - Laboratoire de Virologie - Institut Fédératif de Biologie | Boyer P.; Carcenac R.; Ferrer V.; Harter A.; Izopet J.; Jeanne N.; Latour J.; Ranger N.; Tremeaux P. |
| EPI_ISL_8206925, EPI_ISL_8206972 | Clinical Microbiology Laboratory, Tel Aviv Sourasky Medical Center | Clinical Microbiology Laboratory, Tel Aviv Sourasky Medical Center | Alon Ziv; Amos Adler; Katya Levytskyi; Lior Handler; Ora Halutz |
| EPI_ISL_8317093 | Cliniques universitaires Saint-Luc | UCLouvain/IREC/MBLG-CTMA | Benoit Kabamba Mukadi; Bertrand Bearzatto; Jean-Luc Gala; Nicolas Pinte; Paul Blanpain; Simon Ophélie; Valentin Coste |
| EPI_ISL_8158100, EPI_ISL_8158748, EPI_ISL_8159764, EPI_ISL_8186098, EPI_ISL_8186108 | Cytocheck Laboratory LLC | MAWD Pathology Group, PA | Kelli Fayard; Matison Lewis; Pradip Manna PhD; Shelby Herner; Stephanie Romanowicz |
| EPI_ISL_8170859, EPI_ISL_8170887, EPI_ISL_8170916, EPI_ISL_8170947, EPI_ISL_8171081, EPI_ISL_8171085, EPI_ISL_8171165, EPI_ISL_8171189, EPI_ISL_8171210, EPI_ISL_8171306, EPI_ISL_8171336, EPI_ISL_8171472, EPI_ISL_8171474, EPI_ISL_8171498, EPI_ISL_8171700, EPI_ISL_8171712, EPI_ISL_8171770, EPI_ISL_8172004, EPI_ISL_8172098, EPI_ISL_8172115, EPI_ISL_8172152, EPI_ISL_8172167, EPI_ISL_8172173, EPI_ISL_8172318, EPI_ISL_8200536, EPI_ISL_8200559, EPI_ISL_8200570, EPI_ISL_8200585, EPI_ISL_8200606, EPI_ISL_8200608, EPI_ISL_8200613, EPI_ISL_8200650, EPI_ISL_8200658, EPI_ISL_8200697, EPI_ISL_8200709, EPI_ISL_8200746, EPI_ISL_8200758, EPI_ISL_8200789, EPI_ISL_8200795, EPI_ISL_8200885, EPI_ISL_8200913, EPI_ISL_8200918, EPI_ISL_8200964, EPI_ISL_8201020, EPI_ISL_8201041, EPI_ISL_8201176, EPI_ISL_8201193, EPI_ISL_8201235, EPI_ISL_8201238, EPI_ISL_8201246, EPI_ISL_8201249, EPI_ISL_8201252, EPI_ISL_8201264, EPI_ISL_8201269, EPI_ISL_8201288, EPI_ISL_8201357, EPI_ISL_8201360, EPI_ISL_8201379, EPI_ISL_8201398, EPI_ISL_8201408, EPI_ISL_8201447, EPI_ISL_8201501, EPI_ISL_8201529, EPI_ISL_8201543, EPI_ISL_8201573, EPI_ISL_8201574, EPI_ISL_8201590, EPI_ISL_8201598, EPI_ISL_8201724, EPI_ISL_8201727, EPI_ISL_8201760, EPI_ISL_8201763, EPI_ISL_8201766, EPI_ISL_8201769, EPI_ISL_8201791, EPI_ISL_8201803, EPI_ISL_8201812, EPI_ISL_8201825, EPI_ISL_8201833, EPI_ISL_8201850, EPI_ISL_8201869, EPI_ISL_8201946, EPI_ISL_8201960, EPI_ISL_8202006, EPI_ISL_8202023, EPI_ISL_8202121, EPI_ISL_8202132, EPI_ISL_8202141, EPI_ISL_8202166, EPI_ISL_8202174, EPI_ISL_8202227, EPI_ISL_8202240, EPI_ISL_8202287, EPI_ISL_8202349, EPI_ISL_8202351, EPI_ISL_8202357, EPI_ISL_8202422, EPI_ISL_8202430, EPI_ISL_8202434, EPI_ISL_8202435, EPI_ISL_8202451, EPI_ISL_8202478, EPI_ISL_8202510, EPI_ISL_8202512, EPI_ISL_8202549, EPI_ISL_8202567, EPI_ISL_8202574, EPI_ISL_8202626, EPI_ISL_8202846, EPI_ISL_8202860, EPI_ISL_8202872, EPI_ISL_8202885, EPI_ISL_8202912, EPI_ISL_8202953, EPI_ISL_8249423, EPI_ISL_8249427, EPI_ISL_8249534, EPI_ISL_8249540, EPI_ISL_8249544, EPI_ISL_8249572, EPI_ISL_8249603, EPI_ISL_8249605, EPI_ISL_8249624, EPI_ISL_8249716, EPI_ISL_8249719, EPI_ISL_8249725, EPI_ISL_8249734, EPI_ISL_8249752, EPI_ISL_8249798, EPI_ISL_8249805, EPI_ISL_8249888, EPI_ISL_8249938, EPI_ISL_8249939, EPI_ISL_8249969, EPI_ISL_8249990, EPI_ISL_8249998, EPI_ISL_8250001, EPI_ISL_8250093, EPI_ISL_8250094, EPI_ISL_8250108, EPI_ISL_8250147, EPI_ISL_8250159, EPI_ISL_8250179, EPI_ISL_8250181, EPI_ISL_8250225, EPI_ISL_8250226, EPI_ISL_8250268, EPI_ISL_8250293, EPI_ISL_8250311, EPI_ISL_8250321, EPI_ISL_8250357, EPI_ISL_8250510, EPI_ISL_8250664, EPI_ISL_8250687, EPI_ISL_8250703, EPI_ISL_8250795, EPI_ISL_8250798, EPI_ISL_8250799, EPI_ISL_8250824, EPI_ISL_8250834, EPI_ISL_8250837, EPI_ISL_8250909, EPI_ISL_8250982, EPI_ISL_8250999, EPI_ISL_8251013, EPI_ISL_8251017, EPI_ISL_8251029, EPI_ISL_8251592, EPI_ISL_8251603, EPI_ISL_8251634, EPI_ISL_8251644, EPI_ISL_8251679 | | | |
| see above | Department of Bacteria, Parasites and Fungi, Statens Serum Institut, Copenhagen, Denmark | Statens Serum Institut Bioinformatics and Microbial Genomics | Danish Covid-19 Genome Consortium |
| EPI_ISL_8185870 | Department of Clinical Microbiology | GIGA Medical Genomics | Bouchra Boujemla; Claire Gourzonès; Cécile Meex; Keith Durkin; Laurent Gillet; Maria Artesi; Marie-Pierre Hayette; Nadine Cambisano; Nathalie Renotte; Olivier Ek; Sébastien Bontems; Vincent Bours |
| EPI_ISL_8183263, EPI_ISL_8290748 | Fulgent Genetics | Centers for Disease Control and Prevention Division of Viral Diseases, Pathogen Discovery | Becky Tsai; Benafsh Sapra; Benjamin Rambo-Martin; Christopher Gulvick; Clinton Paden; Dakota Howard; Dhwani Batra; Doreen Ng; Duncan MacCannell; Erisa Sula; Harry Gao; James Xie; Jason Caravas; John Gao; Joseph Fierro; Kristine Lacek; Matthew Schmerer; Mickey Li; Peter Cook; Scott Sammons; Shatavia Morrison; Tymeckia Kendall; Victoria Caban Figueroa; Yan Meng; Yvette Unoarumhi |
| EPI_ISL_8260969, EPI_ISL_8261006, EPI_ISL_8261017, EPI_ISL_8261025, EPI_ISL_8261030, EPI_ISL_8261144, EPI_ISL_8261145, EPI_ISL_8261157, EPI_ISL_8261159, EPI_ISL_8261164, EPI_ISL_8261170, EPI_ISL_8261175, EPI_ISL_8261183, EPI_ISL_8261188, EPI_ISL_8261206, EPI_ISL_8261210, EPI_ISL_8261294, EPI_ISL_8261300 | | | |
| see above | Infinity Biologix | Centers for Disease Control and Prevention Division of Viral Diseases, Pathogen Discovery | Benjamin Rambo-Martin; Chirayu Goswami; Christian Bixby; Christopher Gulvick; Clinton Paden; Dakota Howard; Dhwani Batra; Duncan MacCannell; Erisa Sula; Jason Caravas; Jonathan Schultz; Kristine Lacek; Matthew Schmerer; Peter Cook; Robin Grimwood; Russ Hager; Scott Sammons; Shatavia Morrison; Tymeckia Kendall; Victoria Caban Figueroa; Yihe Wang; Yvette Unoarumhi |
| EPI_ISL_8159888 | Johns Hopkins Hospital Department of Pathology | Johns Hopkins Hospital Department of Pathology | Amary Fall; C. Paul Morris; David Gaston; Heba H. Mostafa; Julie M. Norton; Matthew Schwartz; Michael Forman; Raghda Eldesouki |
| EPI_ISL_8183503, EPI_ISL_8184726, EPI_ISL_8184728, EPI_ISL_8317248, EPI_ISL_8317271 | LABOPAT | LABOPAT | Cynthia Penaloza; Luis Mendoza; Silvia Montilla |
| EPI_ISL_8222049 | Labor Dr. Wisplinghoff - Köln | Robert Koch Institute | |
| EPI_ISL_8182911 | Laboratory for Molecular Diagnostics,IPHMN | Laboratory for Molecular Diagnostics,IPHMN | Danijela Vujošević; Marija Govedarica; Rejhan Hot |
| EPI_ISL_8274098, EPI_ISL_8275396 | Lighthouse Lab in Alderley Park | Wellcome Sanger Institute for the COVID-19 Genomics UK (COG-UK) Consortium | Cordelia Langford; David K. Jackson; Dominic Kwiatkowski; Ewan Harrison; Ian Johnston; Jacquelyn Wynn; Jeffrey Barrett; John Sillitoe on behalf of the Wellcome Sanger Institute COVID-19 Surveillance Team; Mairead Hyland; Roberto Amato; Sonia Goncalves; The Lighthouse Lab in Alderley Park and Alex Alderton |
| EPI_ISL_8276469, EPI_ISL_8280437, EPI_ISL_8280529 | Lighthouse Lab in Glasgow | Wellcome Sanger Institute for the COVID-19 Genomics UK (COG-UK) Consortium | Anna Dominiczak and Alex Alderton; Carol Clugston; Cordelia Langford; David Gray; David K. Jackson; Dominic Kwiatkowski; Ewan Harrison; Harper VanSteenhouse; Ian Johnston; Jeffrey Barrett; John Sillitoe on behalf of the Wellcome Sanger Institute COVID-19 Surveillance Team; Roberto Amato; Sonia Goncalves; Yumi Kasai |
| EPI_ISL_8272463, EPI_ISL_8273411, EPI_ISL_8281329 | Lighthouse Lab in Milton Keynes | Wellcome Sanger Institute for the COVID-19 Genomics UK (COG-UK) Consortium | Cordelia Langford; David K. Jackson; Dominic Kwiatkowski; Ewan Harrison; Ian Johnston; Jeffrey Barrett; John Sillitoe on behalf of the Wellcome Sanger Institute COVID-19 Surveillance Team; Roberto Amato; Sonia Goncalves; The Lighthouse Lab in Milton Keynes and Alex Alderton |
| EPI_ISL_8280527 | Lighthouse Laboratory Plymouth | Wellcome Sanger Institute for the COVID-19 Genomics UK (COG-UK) Consortium | Cordelia Langford; David K. Jackson; Dominic Kwiatkowski; Ewan Harrison; Ian Johnston; Jeffrey Barrett; John Sillitoe on behalf of the Wellcome Sanger Institute COVID-19 Surveillance Team; Lighthouse Laboratory Plymouth and Alex Alderton; Roberto Amato; Sonia Goncalves |
| EPI_ISL_8182744, EPI_ISL_8186596, EPI_ISL_8186689 | MAWD Pathology Group, PA | MAWD Pathology Group, PA | Kelli Fayard; Kelsi Eastman; Matison Lewis; Pradip Manna PhD; Shelby Herner; Stephanie Romanowicz |
| EPI_ISL_8285588 | MEDALL CLUMAX DIAGNOSTICS/BBMP | National Center for Biological Sciences, TIFR - Rockefeller Foundation | Awadhesh Pandit; Bhagyashree Madhav Shelar; Darshan Sreenivas; Dimple Notani; Lakshminarayanan CP; Manisha Bharadwaj; Satyajit Mayor; Uma Ramakrishnan |
| EPI_ISL_8307323, EPI_ISL_8307325, EPI_ISL_8307332 | MRC/UVRI & LSHTM Uganda Research Unit, Uganda Virus Research Institute | MRC/UVRI & LSHTM Uganda Research Unit, Uganda Virus Research Institute | Alfred Ssekagiri; Deogratius Ssemwanga; Hamidah Namagembe; Henry Kyobe-Bbosa; Isaac Ssewanyana; Jocelyn Kiconco; John Kayiwa; Julius Lutwama; Nicholas Bbosa; Pontiano Kaleebu; Ronald Kiiza; Susan Nabadda; Tom Lutalo |
| EPI_ISL_8309860, EPI_ISL_8309978 | Mako Medical | Centers for Disease Control and Prevention Division of Viral Diseases, Pathogen Discovery | Benjamin Rambo-Martin; Christopher Gulvick; Clinton Paden; Dakota Howard; Dhwani Batra; Duncan MacCannell; Erisa Sula; Jason Caravas; Kristine Lacek; Lauren Moon; Matthew Schmerer; Matthew Tugwell; Peter Cook; Scott Sammons; Shatavia Morrison; Tymeckia Kendall; Victoria Caban Figueroa; Yvette Unoarumhi |
| EPI_ISL_8229937, EPI_ISL_8229951, EPI_ISL_8229963, EPI_ISL_8229982, EPI_ISL_8229990, EPI_ISL_8229996, EPI_ISL_8249273, EPI_ISL_8249298 | | | |
| see above | Medizinisch-Diagnostisches Labor Kempten allgäulab | Robert Koch Institute | |
| EPI_ISL_8308168, EPI_ISL_8308189, EPI_ISL_8308194 | Michigan Department of Health and Human Services, Bureau of Laboratories | Michigan Department of Health and Human Services, Bureau of Laboratories | Blankenship HM; Riner D; Soehnlen MK |
| EPI_ISL_8188943 | Middlemore Hospital | Institute of Environmental Science and Research (ESR) | Anja Werno; Antje van der Linden; Arlo Upton; Chris Mansell; Clare Gebbie; David Hammer; Dhanisha Patel; Dragana Drinkovic; Erasmus Smit; Gary McAuliffe; Hana Sofia Andersson; Hermes Perez; James Ussher; Jill Sherwood; Jing Wang; Joep de Ligt; Josh Freeman; Julia Howard; Juliet Elvy; Lauren Jelly; Mary DeAlmeida; Matt Blakiston; Matt Storey; Matthew Rogers; Max Bloomfield; Michael Addidle; Michelle Balm; Muhammad Faisal; Nikki Freed; Olin Silander; Olivia Stroeven; Rachel Boyle; Sally Roberts; SallyAnn Harbison; Sarah Cockerton; Sarah Jefferies; Sharmini Muttaiyah; Susan Morpeth; Susan Taylor; Timothy Blackmore; Vani Sathyendran; Veronica Playle; Virginia Hope; Xiaoyun Ren |
| EPI_ISL_8164601, EPI_ISL_8164656, | Ministry of Health Turkey | Ministry of Health Turkey | Fatma Bayrakdar; Gülay Korukluoğlu; Süleyman Yalcin; Yasemin Coşgun |

| | | | |
|---|---|---|---|
| EPI_ISL_8164657 | | | |
| EPI_ISL_8172051, EPI_ISL_8288172 | Originating lab: Wales Specialist Virology Centre Sequencing lab: Pathogen Genomics Unit | Public Health Wales Microbiology Cardiff Wales Specialist Virology Centre | Alec Birchley; Alexander Adams; Amy Gaskin; Angela Marchbank; Bree Gatica-Wilcox; Catherine Moore; Jason Coombes; Joanne Watkins; Joel Southgate; Johnathan Evans; Laura Gifford; Lauren Gilbert; Lee Graham; Malorie Perry; Matthew Bull; Nicole Pacchiarini; Sally Corden; Sara Kumziene-Summerhayes; Sara Rey; Sarah Taylor; Simon Cottrell; Sophie Jones; Tom Connor |
| EPI_ISL_8242901, EPI_ISL_8242906, EPI_ISL_8287185, EPI_ISL_8287224 | Respiratory Virus Unit, Microbiology Services Colindale, Public Health England | COVID-19 Genomics UK (COG-UK) Consortium | PHE Covid Sequencing Team |
| EPI_ISL_8185869 | SYNLAB | GIGA Medical Genomics | Bouchra Boujemla; Claire Gourzonès; Cécile Meex; Keith Durkin; Laurent Gillet; Maria Artesi; Marie-Pierre Hayette; Nadine Cambisano; Nathalie Renotte; Olivier Ek; Sébastien Bontems; Vincent Bours |
| EPI_ISL_8185077 | Sema4 | Sema4 | Angela Yannes; Bisram Deocharan; Feras Hantash; Matthew Capozziello; Rui Gu; Victoria Parkington; Zdenek Markovic |
| EPI_ISL_8294887, EPI_ISL_8318769 | Tulane University School of Medicine | Tulane University School of Medicine | Di Tian |
| EPI_ISL_8163266, EPI_ISL_8163275, EPI_ISL_8163276, EPI_ISL_8163281, EPI_ISL_8163282, EPI_ISL_8163300, EPI_ISL_8163304, EPI_ISL_8163321, EPI_ISL_8163341, EPI_ISL_8163366 | | | |
| see above | UW Virology Lab | UW Virology Lab | Alexander Greninger; Hong Xie; Isabel Arnould; Keith R Jerome; Meei-Li Huang; Nathan Breit; Patrick Mathias; Pavitra Roychoudhury; Pooneh Hajian; Robert J. Livingston; Sean Ellis; Seffir T. Wendm; Shah Mohamed Bakhash |

We gratefully acknowledge the following Authors from the Originating laboratories responsible for obtaining the specimens, as well as the Submitting laboratories where the genome data were generated and shared via GISAID, on which this research is based.

All Submitters of data may be contacted directly via www.gisaid.org

Authors are sorted alphabetically.

| Accession ID | Originating Laboratory | Submitting Laboratory | Authors |
| --- | --- | --- | --- |
| EPI_ISL_8352208, EPI_ISL_8356546, EPI_ISL_8356663, EPI_ISL_8358068, EPI_ISL_8370307, EPI_ISL_8394586, EPI_ISL_8395229 | | | |
| see above | Aegis Sciences Corporation | Centers for Disease Control and Prevention Division of Viral Diseases, Pathogen Discovery | Alec Vest; Benjamin Rambo-Martin; Christopher Gulvick; Clinton Paden; Cyndi Clark; Dakota Howard; Dhwani Batra; Dillon Nall; Duncan MacCannell; Erisa Sula; Ethan Sanders; Holly Houdeshell; Jason Caravas; Kristine Lacek; Matthew Hardison; Matthew Schmerer; Ola Kvalvaag; Patrick Campbell; Peter Cook; Rob Case; Scott Sammons; Shatavia Morrison; Shaun Westlund; Tymeckia Kendall; Victoria Caban Figueroa; Vikramsinha Ghorpade; Yvette Unoarumhi |
| EPI_ISL_8338494 | Azienda Ospedaliero - Universitaria di Modena Policlinico - Virologia e Microbiologia Molecolare"" | Istituto Zooprofilattico Sperimentale della Lombardia e dell'Emilia Romagna (IZSLER), Risk Analysis and Genomic Epidemiology Unit | Erika Scaltriti; Ilaria Menozzi; Marina Morganti; Stefano Pongolini |
| EPI_ISL_8379784 | Bayerisches Landesamt fuer Gesundheit und Lebensmittelsicherheit (LGL) | Bayerisches Landesamt fuer Gesundheit und Lebensmittelsicherheit (LGL) | Alexandra Dangel; Andreas Sing; Annika Sprenger; Carola Berger; Laura Weise; Nikolaus Ackermann; Sabrina Hepner |
| EPI_ISL_8354549 | CDPH VBL | California Department of Public Health | CDPH COVIDNet |
| EPI_ISL_8401203 | CHU UCL Namur, Avenue G. Thérasse 1, 5530 Yvoir, Belgium | Plateforme de testing Namuroise | Degosserie Jonathan; Demars Aurore; Denis Olivier; Laurent Hélène; Maschietto Céline; Mullier François; Nobis Chloé; Otto Gaetan |
| EPI_ISL_8378658 | Clinical Virology | Clinical Bacteriology, University Hospital Basel | Adrian Egli; Alfredo Mari; Fanny Wegner; Hans Hirsch; Helena MB Seth-Smith; Julia Bielicki; Karoline Leuzinger; Manuel Battegay; Tim Roloff |
| EPI_ISL_8378837 | Colorado Department of Public Health and Environment | Colorado Department of Public Health and Environment | Alexandria Rossheim; Arianna Smith; Diana Ir; Emily A. Travanty; Laura Bankers; Mandy Waters; Michael Martin; Molly C. Hetherington-Rauth; Shannon R. Matzinger |
| EPI_ISL_8344062, EPI_ISL_8344068, EPI_ISL_8344077, EPI_ISL_8344098, EPI_ISL_8344151, EPI_ISL_8344154, EPI_ISL_8344171, EPI_ISL_8344172, EPI_ISL_8344182, EPI_ISL_8344200, EPI_ISL_8344204, EPI_ISL_8344207, EPI_ISL_8344226, EPI_ISL_8344242, EPI_ISL_8344272, EPI_ISL_8344294, EPI_ISL_8344329, EPI_ISL_8344341, EPI_ISL_8344346, EPI_ISL_8344368, EPI_ISL_8344380, EPI_ISL_8344397, EPI_ISL_8344436, EPI_ISL_8344440, EPI_ISL_8344449, EPI_ISL_8344458, EPI_ISL_8344480, EPI_ISL_8344489, EPI_ISL_8344493, EPI_ISL_8344523, EPI_ISL_8344526, EPI_ISL_8344527, EPI_ISL_8344534, EPI_ISL_8344562, EPI_ISL_8344564, EPI_ISL_8344660, EPI_ISL_8344692, EPI_ISL_8344697, EPI_ISL_8344700, EPI_ISL_8344739, EPI_ISL_8344744, EPI_ISL_8344745, EPI_ISL_8344755, EPI_ISL_8344769, EPI_ISL_8344781, EPI_ISL_8344897, EPI_ISL_8344903, EPI_ISL_8344936, EPI_ISL_8344957, EPI_ISL_8344963, EPI_ISL_8344992, EPI_ISL_8345013, EPI_ISL_8345024, EPI_ISL_8345047, EPI_ISL_8345186, EPI_ISL_8345207, EPI_ISL_8345214, EPI_ISL_8345232, EPI_ISL_8345234, EPI_ISL_8345237, EPI_ISL_8345254, EPI_ISL_8345258, EPI_ISL_8345311, EPI_ISL_8345319, EPI_ISL_8345328, EPI_ISL_8345347, EPI_ISL_8345432, EPI_ISL_8345514, EPI_ISL_8345533, EPI_ISL_8345538, EPI_ISL_8345568, EPI_ISL_8345599, EPI_ISL_8345627, EPI_ISL_8345637, EPI_ISL_8345653, EPI_ISL_8345772, EPI_ISL_8345775, EPI_ISL_8345802, EPI_ISL_8345854, EPI_ISL_8345914, EPI_ISL_8345921, EPI_ISL_8345923, EPI_ISL_8345928, EPI_ISL_8345930, EPI_ISL_8345934, EPI_ISL_8345977, EPI_ISL_8345990, EPI_ISL_8345999, EPI_ISL_8346020, EPI_ISL_8346023, EPI_ISL_8346030, EPI_ISL_8346041, EPI_ISL_8346097, EPI_ISL_8346102, EPI_ISL_8346126, EPI_ISL_8346204, EPI_ISL_8346206, EPI_ISL_8346273, EPI_ISL_8346283, EPI_ISL_8346317, EPI_ISL_8346348, EPI_ISL_8346360, EPI_ISL_8346370, EPI_ISL_8346381, EPI_ISL_8346416, EPI_ISL_8346462, EPI_ISL_8346477, EPI_ISL_8346478, EPI_ISL_8346511, EPI_ISL_8346598, EPI_ISL_8346604, EPI_ISL_8346607, EPI_ISL_8346620, EPI_ISL_8346627, EPI_ISL_8346645, EPI_ISL_8346646, EPI_ISL_8346685, EPI_ISL_8346687, EPI_ISL_8346719, EPI_ISL_8346749, EPI_ISL_8346768, EPI_ISL_8346770, EPI_ISL_8346780, EPI_ISL_8346782, EPI_ISL_8346786, EPI_ISL_8346812, EPI_ISL_8346821, EPI_ISL_8346824, EPI_ISL_8346856, EPI_ISL_8346862, EPI_ISL_8346866, EPI_ISL_8346885, EPI_ISL_8346890, EPI_ISL_8346899, EPI_ISL_8346912, EPI_ISL_8346921, EPI_ISL_8346927, EPI_ISL_8346937, EPI_ISL_8346951, EPI_ISL_8347027, EPI_ISL_8347055, EPI_ISL_8347064, EPI_ISL_8347066, EPI_ISL_8347087, EPI_ISL_8347106, EPI_ISL_8347137, EPI_ISL_8347147, EPI_ISL_8347179, EPI_ISL_8347187, EPI_ISL_8347204, EPI_ISL_8347241, EPI_ISL_8347351, EPI_ISL_8347371, EPI_ISL_8347392, EPI_ISL_8347396, EPI_ISL_8347398, EPI_ISL_8347427, EPI_ISL_8347442, EPI_ISL_8347446, EPI_ISL_8347448, EPI_ISL_8347467, EPI_ISL_8347476, EPI_ISL_8347521, EPI_ISL_8347538, EPI_ISL_8347706, EPI_ISL_8347713, EPI_ISL_8381420, EPI_ISL_8381436, EPI_ISL_8381456, EPI_ISL_8381463, EPI_ISL_8381467, EPI_ISL_8381480, EPI_ISL_8381494, EPI_ISL_8381504, EPI_ISL_8381506, EPI_ISL_8381509, EPI_ISL_8381574, EPI_ISL_8381581, EPI_ISL_8381589, EPI_ISL_8381621, EPI_ISL_8381642, EPI_ISL_8381643, EPI_ISL_8381652, EPI_ISL_8381670, EPI_ISL_8381677, EPI_ISL_8381685, EPI_ISL_8381690, EPI_ISL_8381703, EPI_ISL_8381713, EPI_ISL_8381722, EPI_ISL_8382103, EPI_ISL_8382106, EPI_ISL_8382114, EPI_ISL_8382128, EPI_ISL_8382131, EPI_ISL_8382132, EPI_ISL_8382143, EPI_ISL_8382148, EPI_ISL_8382173, EPI_ISL_8382180, EPI_ISL_8382192, EPI_ISL_8382196, EPI_ISL_8382215, EPI_ISL_8382247, EPI_ISL_8382252, EPI_ISL_8382253, EPI_ISL_8382261, EPI_ISL_8382277, EPI_ISL_8382285, EPI_ISL_8382314, EPI_ISL_8382329, EPI_ISL_8382350, EPI_ISL_8382354, EPI_ISL_8382359, EPI_ISL_8382383, EPI_ISL_8382394, EPI_ISL_8382415, EPI_ISL_8382434, EPI_ISL_8382441, EPI_ISL_8382451, EPI_ISL_8382463, EPI_ISL_8382464, EPI_ISL_8382467, EPI_ISL_8382490, EPI_ISL_8382504, EPI_ISL_8382509, EPI_ISL_8382523, EPI_ISL_8382555, EPI_ISL_8382565, EPI_ISL_8382577, EPI_ISL_8382589, EPI_ISL_8382600, EPI_ISL_8382604, EPI_ISL_8382635, EPI_ISL_8382639, EPI_ISL_8382649, EPI_ISL_8382650, EPI_ISL_8382661, EPI_ISL_8382716, EPI_ISL_8382745, EPI_ISL_8382757, EPI_ISL_8382784, EPI_ISL_8382791, EPI_ISL_8382794, EPI_ISL_8382800, EPI_ISL_8382802, EPI_ISL_8382811, EPI_ISL_8382820, EPI_ISL_8382823, EPI_ISL_8382829, EPI_ISL_8382834, EPI_ISL_8382840 | | | |
| see above | Department of Bacteria, Parasites and Fungi, Statens Serum Institut, Copenhagen, Denmark | Statens Serum Institut Bioinformatics and Microbial Genomics | Danish Covid-19 Genome Consortium |
| EPI_ISL_8380408, EPI_ISL_8380410 | Gandhi Medical College and Hospital (GMCH), Secunderabad | NIV Influenza | D.R.Manisha Rani; Dr.G.Sushma Rajya Lakshmi; Dr.K.Nagamani; Dr.Sunitha Pakalapaty |
| EPI_ISL_8329270, EPI_ISL_8329272, EPI_ISL_8329274, EPI_ISL_8329277, EPI_ISL_8329279, EPI_ISL_8329280, EPI_ISL_8329282, EPI_ISL_8329286, EPI_ISL_8329292, EPI_ISL_8329293, EPI_ISL_8329294, EPI_ISL_8329298, EPI_ISL_8329300, EPI_ISL_8329303, EPI_ISL_8329304, EPI_ISL_8329305, EPI_ISL_8329306, EPI_ISL_8329307, EPI_ISL_8329308, EPI_ISL_8329310, EPI_ISL_8329313, EPI_ISL_8329316, EPI_ISL_8329319, EPI_ISL_8329323, EPI_ISL_8329325, EPI_ISL_8329328, EPI_ISL_8329332, EPI_ISL_8329336, EPI_ISL_8329342, EPI_ISL_8329343, EPI_ISL_8329345, EPI_ISL_8329348, EPI_ISL_8329349, EPI_ISL_8329350, EPI_ISL_8329351, EPI_ISL_8329352, EPI_ISL_8329353, EPI_ISL_8329354, EPI_ISL_8329357 | | | |
| see above | Idaho Bureau of Laboratories | Idaho Bureau of Laboratories | Aimee Ceniseros; Christian Loera; Christopher Ball; Matthew Charles Burns; R. Beukelman; Robert L. Voermans |
| EPI_ISL_8364636, EPI_ISL_8365040 | Laboratory Corporation of America | Centers for Disease Control and Prevention Division of Viral Diseases, Pathogen Discovery | Amanda Douglas; Amanda Suchanek; Andrea Throop; Ayla Burns; Benjamin Rambo-Martin; Bobbi Croy; Brian Krueger; Brian Norvell; Christopher Gulvick; Christos Petropoulos; Clinton Paden; Craig Lukasik; Dakota Howard; Debbie Boles; Dhwani Batra; Duncan MacCannell; Eyad Almasri; Goran Stevovic; Howard Engler; Hrushikesh Deshmukh; Jake Humphrey; Jana Schroth; Jason Caravas; Joe Voshell; John Pruitt; Jonathan Meltzer; Jonathan Williams; Kimberly Wagner; Kristine Lacek; Lax Iyer; Lisa Pfefferle; Lyndon Tilson; Manoj Jain; Marcia Eisenberg; Mary Cristobal; Mary Williamson; Matthew Robinson; Matthew Schmerer; Michael Levandoski; Mike Sapeta; Mindy Nye; Minoo Agarwal; Mohan Kolli; Nuthawin Charoensri; Oren Cohen; Peter Cook; Prashant Gupta; Qian Zeng; Rama Ghatti; Scott Parker; Scott Ryan; Scott Sammons; Shatavia Morrison; Stanley Letovsky; Steven Ragan; Suresh Selvaraju; Susan Countryman; Susan Hicks; Suzanne Dale; Thomas Urban; Tim Kuphal; Tricia Zwiefelhofer; Tymeckia Kendall; Victoria Caban Figueroa; Vincent Drouillon; Yvette Unoarumhi |
| EPI_ISL_8371879, EPI_ISL_8372975 | MAWD Pathology Group, PA | MAWD Pathology Group, PA | Kelli Fayard; Matison Lewis; Pradip Manna PhD; Shelby Herner; Stephanie Romanowicz |
| EPI_ISL_8336261, EPI_ISL_8336431, EPI_ISL_8336432, EPI_ISL_8336433, EPI_ISL_8336434, EPI_ISL_8336435, EPI_ISL_8336439, EPI_ISL_8336441 | | | |
| see above | Ministry of Health Turkey | Ministry of Health Turkey | Fatma Bayrakdar; Gülay Korukluoğlu; Gültekin Ünal; Süleyman Yalcin; Yasemin Coşgun |
| EPI_ISL_8384065, EPI_ISL_8384066, EPI_ISL_8384067, EPI_ISL_8384068, EPI_ISL_8384069, EPI_ISL_8384070, EPI_ISL_8384071, EPI_ISL_8384072 | | | |
| see above | National Institute of Public Health | National Institute of Public Health | Alexander Nagy; Helena Jirincova; Jan Moskalyk; Jaromira Vecerova; Timotej Suri |
| EPI_ISL_8380869 | Servicio Microbiología Hospital La Paz | Servicio Microbiología Hospital La Paz | Elie Dahdouh; Fernando Lázaro; Iván Bloise; Jesús Mingorance Cruz; Rubén Cáceres |
| EPI_ISL_8388410 | StarMed Healthcare | UNC Charlotte Environmental Monitoring Laboratory | Cynthia Gibas; Jannatul Ferdous; Jessica Schlueter; Juan Bolanos; Kevin Lambirth; Samuel Kunkleman; Torri Weathers |
| EPI_ISL_8360501, EPI_ISL_8360516, EPI_ISL_8360598, EPI_ISL_8360650 | State Laboratories Division, Hawaii State Department of Health | State Laboratories Division, Hawaii State Department of Health | Ayana Garnet; Briana Ofilas; Cheryl-lynn Daquip; Cheyenne Barela; Daniel Strange; Drew Kuwazaki; Edward Desmond; Jeffrey Au; Mark Nagata; Pamela O'Brien; Razvan Sultana; Remedios Gose; Samantha Cotter; Samantha Sruba |
| EPI_ISL_8382062 | Sweden, Vastragotaland, Unilabs | Unilabs, Eskilstuna, Sweden | Emma Arvidsson |
| EPI_ISL_8354884, EPI_ISL_8355008 | UW Virology Lab | UW Virology Lab | Alexander Greninger; Hong Xie; Isabel Arnould; Keith R Jerome; Meei-Li Huang; Nathan Breit; Patrick Mathias; Pavitra Roychoudhury; Pooneh Hajian; Ricardo Perez; Robert J. Livingston; Sean Ellis; Seffir T. Wendm; Shah Mohamed Bakhash |
| EPI_ISL_8360079 | University of New Mexico Hospital | Center for Global Health, University of New Mexico Health Sciences Center | Darrell Dinwiddie; Daryl Domman; Jesse Young; Jon Femling; Justin Bacca; Kurt Schwalm; Valerie Morley |
| EPI_ISL_8379873 | WHO National Influenza Centre Russian Federation | WHO National Influenza Centre Russian Federation | Andrey Komissarov; Artem Fadeev; Daria Danilenko; Dmitry Lioznov; Elena Nabieva; Georgii Bazykin; Kirill Varchenko; Ksenia Safina; Kseniya Komissarova; Maria Pisareva; Mikhail Bakaev; Nikita Yolshin; Oula Mansour; Tamila Musaeva; Veronika Eder |

We gratefully acknowledge the following Authors from the Originating laboratories responsible for obtaining the specimens, as well as the Submitting laboratories where the genome data were generated and shared via GISAID, on which this research is based.

All Submitters of data may be contacted directly via www.gisaid.org

Authors are sorted alphabetically.

Acknowledgement EPI_SET Identifier: EPI_SET_20220527xa

| Accession ID | Originating Laboratory | Submitting Laboratory | Authors |
|---|---|---|---|
| EPI_ISL_8424966, EPI_ISL_8425144, EPI_ISL_8426200 | Aegis Sciences Corporation | Centers for Disease Control and Prevention Division of Viral Diseases, Pathogen Discovery | Alec Vest; Benjamin Rambo-Martin; Christopher Gulvick; Clinton Paden; Cyndi Clark; Dakota Howard; Dhwani Batra; Dillon Nall; Duncan MacCannell; Erisa Sula; Ethan Sanders; Holly Houdeshell; Jason Caravas; Kristine Lacek; Matthew Hardison; Matthew Schmerer; Ola Kvalvaag; Patrick Campbell; Peter Cook; Rob Case; Scott Sammons; Shatavia Morrison; Shaun Westlund; Tymeckia Kendall; Victoria Caban Figueroa; Vikramsinha Ghorpade; Yvette Unoarumhi |
| EPI_ISL_8417976, EPI_ISL_8417980, EPI_ISL_8417981, EPI_ISL_8417983, EPI_ISL_8417990, EPI_ISL_8417994, EPI_ISL_8417999, EPI_ISL_8418107, EPI_ISL_8418122, EPI_ISL_8418124, EPI_ISL_8418127, EPI_ISL_8418135, EPI_ISL_8418142, EPI_ISL_8418144, EPI_ISL_8418160, EPI_ISL_8418161, EPI_ISL_8418172, EPI_ISL_8418173, EPI_ISL_8418175, EPI_ISL_8418180, EPI_ISL_8418182, EPI_ISL_8418185, EPI_ISL_8418186, EPI_ISL_8418187, EPI_ISL_8418208, EPI_ISL_8418216, EPI_ISL_8418217, EPI_ISL_8418223, EPI_ISL_8418231, EPI_ISL_8418234, EPI_ISL_8418237, EPI_ISL_8418241, EPI_ISL_8418244, EPI_ISL_8418246, EPI_ISL_8418251, EPI_ISL_8418258, EPI_ISL_8418260, EPI_ISL_8418270, EPI_ISL_8418271, EPI_ISL_8418281, EPI_ISL_8418282, EPI_ISL_8418329, EPI_ISL_8418421, EPI_ISL_8418498, EPI_ISL_8418502, EPI_ISL_8418513, EPI_ISL_8418522, EPI_ISL_8418527, EPI_ISL_8418528, EPI_ISL_8418537, EPI_ISL_8418538, EPI_ISL_8418546, EPI_ISL_8418557, EPI_ISL_8418562, EPI_ISL_8418566, EPI_ISL_8418577, EPI_ISL_8418578, EPI_ISL_8418587, EPI_ISL_8418588, EPI_ISL_8418593, EPI_ISL_8418598, EPI_ISL_8418600, EPI_ISL_8418603, EPI_ISL_8418614, EPI_ISL_8418619, EPI_ISL_8418628, EPI_ISL_8418638, EPI_ISL_8418639, EPI_ISL_8418648, EPI_ISL_8418656, EPI_ISL_8418661, EPI_ISL_8418662, EPI_ISL_8418676, EPI_ISL_8418678, EPI_ISL_8418679, EPI_ISL_8418693, EPI_ISL_8418704, EPI_ISL_8418705, EPI_ISL_8418706, EPI_ISL_8418718, EPI_ISL_8418731, EPI_ISL_8418762, EPI_ISL_8418764, EPI_ISL_8418767, EPI_ISL_8418773, EPI_ISL_8418787, EPI_ISL_8418827, EPI_ISL_8418830, EPI_ISL_8418843, EPI_ISL_8418853, EPI_ISL_8418856, EPI_ISL_8418859, EPI_ISL_8418875, EPI_ISL_8418900, EPI_ISL_8418917, EPI_ISL_8418922, EPI_ISL_8418936, EPI_ISL_8418938, EPI_ISL_8418945, EPI_ISL_8418948, EPI_ISL_8418953, EPI_ISL_8418961, EPI_ISL_8418968, EPI_ISL_8418969, EPI_ISL_8418972, EPI_ISL_8418975, EPI_ISL_8418986, EPI_ISL_8418987, EPI_ISL_8418989, EPI_ISL_8418990, EPI_ISL_8418996, EPI_ISL_8418999, EPI_ISL_8419005, EPI_ISL_8419027, EPI_ISL_8419032, EPI_ISL_8419052, EPI_ISL_8419062, EPI_ISL_8419075, EPI_ISL_8419077, EPI_ISL_8419078, EPI_ISL_8419094, EPI_ISL_8419095, EPI_ISL_8419099, EPI_ISL_8419115, EPI_ISL_8419117, EPI_ISL_8419126, EPI_ISL_8419142, EPI_ISL_8419145, EPI_ISL_8419152, EPI_ISL_8419153, EPI_ISL_8419161, EPI_ISL_8419180, EPI_ISL_8419181, EPI_ISL_8419182, EPI_ISL_8419188, EPI_ISL_8419192, EPI_ISL_8419211, EPI_ISL_8419213, EPI_ISL_8419224, EPI_ISL_8419226, EPI_ISL_8419237, EPI_ISL_8419241, EPI_ISL_8419243, EPI_ISL_8419252, EPI_ISL_8419269, EPI_ISL_8419278, EPI_ISL_8419285, EPI_ISL_8419289, EPI_ISL_8419290, EPI_ISL_8419292, EPI_ISL_8419302, EPI_ISL_8419303, EPI_ISL_8419305, EPI_ISL_8419309, EPI_ISL_8419314, EPI_ISL_8419324, EPI_ISL_8419329, EPI_ISL_8419332, EPI_ISL_8419336, EPI_ISL_8419342, EPI_ISL_8419355, EPI_ISL_8419356, EPI_ISL_8419360, EPI_ISL_8419367, EPI_ISL_8419368, EPI_ISL_8419378, EPI_ISL_8419379, EPI_ISL_8419383, EPI_ISL_8419386, EPI_ISL_8419388, EPI_ISL_8419395, EPI_ISL_8419396, EPI_ISL_8419406, EPI_ISL_8419411, EPI_ISL_8419415, EPI_ISL_8419426, EPI_ISL_8419429, EPI_ISL_8419442, EPI_ISL_8419447, EPI_ISL_8419454, EPI_ISL_8419459, EPI_ISL_8419465, EPI_ISL_8419468, EPI_ISL_8419475, EPI_ISL_8419477, EPI_ISL_8419481, EPI_ISL_8419499, EPI_ISL_8419501, EPI_ISL_8419505, EPI_ISL_8419516, EPI_ISL_8419518, EPI_ISL_8419558, EPI_ISL_8419589, EPI_ISL_8419667, EPI_ISL_8419673, EPI_ISL_8419682, EPI_ISL_8419697, EPI_ISL_8419699, EPI_ISL_8419702, EPI_ISL_8419703, EPI_ISL_8419704, EPI_ISL_8419716, EPI_ISL_8419720, EPI_ISL_8419727, EPI_ISL_8419733, EPI_ISL_8419745, EPI_ISL_8419752, EPI_ISL_8419766, EPI_ISL_8419771, EPI_ISL_8419774, EPI_ISL_8419778, EPI_ISL_8419779, EPI_ISL_8419786, EPI_ISL_8419787, EPI_ISL_8419790, EPI_ISL_8419806, EPI_ISL_8419841, EPI_ISL_8419847, EPI_ISL_8419850, EPI_ISL_8419857, EPI_ISL_8419872, EPI_ISL_8419873, EPI_ISL_8419875, EPI_ISL_8419882, EPI_ISL_8419884, EPI_ISL_8419888, EPI_ISL_8419905, EPI_ISL_8419910, EPI_ISL_8419915, EPI_ISL_8419932, EPI_ISL_8419933, EPI_ISL_8419936, EPI_ISL_8419937, EPI_ISL_8419938, EPI_ISL_8419947, EPI_ISL_8419948, EPI_ISL_8420649, EPI_ISL_8420658, EPI_ISL_8420669, EPI_ISL_8420681, EPI_ISL_8420684, EPI_ISL_8420685, EPI_ISL_8420691, EPI_ISL_8420712 | | | |
| see above | Department of Bacteria, Parasites and Fungi, Statens Serum Institut, Copenhagen, Denmark | Statens Serum Institut Bioinformatics and Microbial Genomics | Danish Covid-19 Genome Consortium |
| EPI_ISL_8414381 | MVZ Labor Dr. Fenner und Kollegen (Standort Hamburg) | Robert Koch Institute | |
| EPI_ISL_8421377 | Medical Virology and BSL3 Laboratory | Centre de Séquençage Génomique | Abbad Anas; Amalou Ghita; Anga Latifa; Barakat Abdelhamid; Bouzidi Aymane; Charoute Hicham; Chgouri Fatima; Dersi Noureddine; EL Hamouchi Adil; El Oualid Abdelmjid; Faouzi Abdellah; Harmak Houda; Maaroufi Abderrahmane; Nadifiyine Saloua; Nourlil Jalal; Omondi Francis Carey; Somda Soro Georgina Charlene; Zemmouri Faouzia; Zouheir Yassine |
| EPI_ISL_8414362, EPI_ISL_8414363 | National Institute of Public Health | National Institute of Public Health | Alexander Nagy; Helena Jirincova; Jan Moskalyk; Jaromira Vecerova; Timotej Suri |
| EPI_ISL_8410970 | SYNLAB MVZ Leverkusen | Robert Koch Institute | |
| EPI_ISL_8427960, EPI_ISL_8428054 | The New York Center for Travel and Tropical Medicine | Biotia | Christopher Mason; David Danko; Dorottya Nagy-Szakal; Mara Couto-Rodriguez; Marilyne Debieu; Niamh O'Hara; Xavier Jirau Serrano |

We gratefully acknowledge the following Authors from the Originating laboratories responsible for obtaining the specimens, as well as the Submitting laboratories where the genome data were generated and shared via GISAID, on which this research is based.

All Submitters of data may be contacted directly via www.gisaid.org

Authors are sorted alphabetically.

Acknowledgement EPI_SET Identifier: EPI_SET_20220527ng

| Accession ID | Originating Laboratory | Submitting Laboratory | Authors |
|---|---|---|---|
| EPI_ISL_8552074, EPI_ISL_8552075, EPI_ISL_8552076, EPI_ISL_8552084, EPI_ISL_8552086, EPI_ISL_8552090, EPI_ISL_8552098, EPI_ISL_8554348, EPI_ISL_8555368, EPI_ISL_8555369, EPI_ISL_8555536, EPI_ISL_8555537 | | | |
| see above | Aegis Sciences Corporation | Centers for Disease Control and Prevention Division of Viral Diseases, Pathogen Discovery | Alec Vest; Benjamin Rambo-Martin; Christopher Gulvick; Clinton Paden; Cyndi Clark; Dakota Howard; Dhwani Batra; Dillon Nall; Duncan MacCannell; Erisa Sula; Ethan Sanders; Holly Houdeshell; Jason Caravas; Kristine Lacek; Matthew Hardison; Matthew Schmerer; Ola Kvalvaag; Patrick Campbell; Peter Cook; Rob Case; Scott Sammons; Shatavia Morrison; Shaun Westlund; Tymeckia Kendall; Victoria Caban Figueroa; Vikramsinha Ghorpade; Yvette Unoarumhi |
| EPI_ISL_8543197, EPI_ISL_8543200, EPI_ISL_8543202, EPI_ISL_8543208 | Arcispedale Santa Maria Nuova Autoimmunita Allergologia e Biotecnologie Innovative"" | Istituto Zooprofilattico Sperimentale della Lombardia e dell'Emilia Romagna (IZSLER), Risk Analysis and Genomic Epidemiology Unit | Erika Scaltriti; Ilaria Menozzi; Marina Morganti; Stefano Pongolini |
| EPI_ISL_8509194 | Arizona State University | Arizona State University | Efrem S. Lim; James C. Hu; Joshua LaBaer; LaRinda A. Holland; Matthew F. Smith; Nathaniel Johnson; Regan A. Sullins; Steven C. Holland; Vel Murugan |
| EPI_ISL_8543243, EPI_ISL_8543245 | Azienda Ospedaliero - Universitaria di Modena Policlinico - Virologia e Microbiologia Molecolare"" | Istituto Zooprofilattico Sperimentale della Lombardia e dell'Emilia Romagna (IZSLER), Risk Analysis and Genomic Epidemiology Unit | Erika Scaltriti; Ilaria Menozzi; Marina Morganti; Stefano Pongolini |
| EPI_ISL_8519792, EPI_ISL_8519793 | Bidh Lab | National Public Health Laboratory | National Public Health Labaoratory Team/Bidh Lab Team |
| EPI_ISL_8547008 | Clinical Microbiology Laboratory, Tel Aviv Sourasky Medical Center | Clinical Microbiology Laboratory, Tel Aviv Sourasky Medical Center | Alon Ziv; Amos Adler; Goel Morad; Katya Levytskyi; Lior Handler; Ora Halutz |
| EPI_ISL_8515049, EPI_ISL_8555439, EPI_ISL_8555440, EPI_ISL_8555441 | Colorado Department of Public Health and Environment | Colorado Department of Public Health and Environment | Alexandria Rossheim; Arianna Smith; Diana Ir; Emily A. Travanty; Laura Bankers; Mandy Waters; Michael Martin; Molly C. Hetherington-Rauth; Shannon R. Matzinger |
| EPI_ISL_8528767, EPI_ISL_8528771, EPI_ISL_8528816, EPI_ISL_8528823, EPI_ISL_8528827, EPI_ISL_8528832, EPI_ISL_8528836, EPI_ISL_8528838, EPI_ISL_8528839, EPI_ISL_8528842, EPI_ISL_8528845, EPI_ISL_8528853, EPI_ISL_8528854, EPI_ISL_8528870, EPI_ISL_8528876, EPI_ISL_8528879, EPI_ISL_8528880, EPI_ISL_8528883, EPI_ISL_8528885, EPI_ISL_8528886, EPI_ISL_8528888, EPI_ISL_8528896, EPI_ISL_8528897, EPI_ISL_8528912, EPI_ISL_8528926, EPI_ISL_8528929, EPI_ISL_8528933, EPI_ISL_8528937, EPI_ISL_8528938, EPI_ISL_8528939, EPI_ISL_8528940, EPI_ISL_8528944, EPI_ISL_8528954, EPI_ISL_8528961, EPI_ISL_8528973, EPI_ISL_8528974, EPI_ISL_8528976, EPI_ISL_8528979, EPI_ISL_8528989, EPI_ISL_8528998, EPI_ISL_8529001, EPI_ISL_8529022, EPI_ISL_8529031, EPI_ISL_8529032, EPI_ISL_8529037, EPI_ISL_8529041, EPI_ISL_8529060, EPI_ISL_8529069, EPI_ISL_8529074, EPI_ISL_8529075, EPI_ISL_8529076, EPI_ISL_8529099, EPI_ISL_8529105, EPI_ISL_8529108, EPI_ISL_8529112, EPI_ISL_8529121, EPI_ISL_8529134, EPI_ISL_8529136, EPI_ISL_8529139, EPI_ISL_8529142, EPI_ISL_8529144, EPI_ISL_8529147, EPI_ISL_8529148, EPI_ISL_8529165, EPI_ISL_8529166, EPI_ISL_8529172, EPI_ISL_8529176, EPI_ISL_8529181, EPI_ISL_8529187, EPI_ISL_8529191, EPI_ISL_8529194, EPI_ISL_8529204, EPI_ISL_8529210, EPI_ISL_8529212, EPI_ISL_8529228, EPI_ISL_8529233, EPI_ISL_8529244, EPI_ISL_8529268, EPI_ISL_8529269, EPI_ISL_8529279, EPI_ISL_8529301, EPI_ISL_8529309, EPI_ISL_8529320, EPI_ISL_8529321, EPI_ISL_8529322, EPI_ISL_8529324, EPI_ISL_8529325, EPI_ISL_8529326, EPI_ISL_8529339, EPI_ISL_8529345, EPI_ISL_8529349, EPI_ISL_8529357, EPI_ISL_8529362, EPI_ISL_8529367, EPI_ISL_8529368, EPI_ISL_8529369, EPI_ISL_8529377, EPI_ISL_8529398, EPI_ISL_8529402, EPI_ISL_8529404, EPI_ISL_8529408, EPI_ISL_8529415, EPI_ISL_8529427, EPI_ISL_8529428, EPI_ISL_8529430, EPI_ISL_8529435, EPI_ISL_8529443, EPI_ISL_8529444, EPI_ISL_8529451, EPI_ISL_8529458, EPI_ISL_8529461, EPI_ISL_8529462, EPI_ISL_8529466, EPI_ISL_8529471, EPI_ISL_8529477, EPI_ISL_8529479, EPI_ISL_8529501, EPI_ISL_8529506, EPI_ISL_8529512, EPI_ISL_8529518, EPI_ISL_8529519, EPI_ISL_8529523, EPI_ISL_8529524, EPI_ISL_8529525, EPI_ISL_8529527, EPI_ISL_8529528, EPI_ISL_8529529, EPI_ISL_8529540, EPI_ISL_8529543, EPI_ISL_8529562, EPI_ISL_8529563, EPI_ISL_8529567, EPI_ISL_8529569, EPI_ISL_8529579, EPI_ISL_8529580, EPI_ISL_8529584, EPI_ISL_8529587, EPI_ISL_8529591, EPI_ISL_8529598, EPI_ISL_8529620, EPI_ISL_8529622, EPI_ISL_8529634, EPI_ISL_8529635, EPI_ISL_8529647, EPI_ISL_8529648, EPI_ISL_8529659, EPI_ISL_8529665, EPI_ISL_8529673, EPI_ISL_8529675, EPI_ISL_8529680, EPI_ISL_8529688, EPI_ISL_8529694, EPI_ISL_8529702, EPI_ISL_8529708, EPI_ISL_8529709, EPI_ISL_8529711, EPI_ISL_8529718, EPI_ISL_8529726, EPI_ISL_8529727, EPI_ISL_8529733, EPI_ISL_8529735, EPI_ISL_8529736, EPI_ISL_8529749, EPI_ISL_8529754, EPI_ISL_8529784, EPI_ISL_8529806, EPI_ISL_8529815, EPI_ISL_8529817, EPI_ISL_8529820, EPI_ISL_8529846, EPI_ISL_8529875, EPI_ISL_8529878, EPI_ISL_8529889, EPI_ISL_8529891, EPI_ISL_8529893, EPI_ISL_8529897, EPI_ISL_8529904, EPI_ISL_8529905, EPI_ISL_8529907, EPI_ISL_8529911, EPI_ISL_8529918, EPI_ISL_8529922, EPI_ISL_8529930, EPI_ISL_8529943, EPI_ISL_8529955, EPI_ISL_8529989, EPI_ISL_8529991, EPI_ISL_8529998, EPI_ISL_8530000, EPI_ISL_8530002, EPI_ISL_8530017, EPI_ISL_8530020, EPI_ISL_8530022, EPI_ISL_8530025, EPI_ISL_8530035, EPI_ISL_8530041, EPI_ISL_8530042, EPI_ISL_8530045, EPI_ISL_8530063, EPI_ISL_8530064, EPI_ISL_8530069 | | | |
| see above | Department of Bacteria, Parasites and Fungi, Statens Serum Institut, Copenhagen, Denmark | Statens Serum Institut Bioinformatics and Microbial Genomics | Danish Covid-19 Genome Consortium |
| EPI_ISL_8508635 | Dianalabs SA | Genesupport | Geraldine Jost; Katia Jaton; Nadia Liassine; Tanguy ARAUD |
| EPI_ISL_8504750, EPI_ISL_8504754 | Fulgent Genetics | Centers for Disease Control and Prevention Division of Viral Diseases, Pathogen Discovery | Becky Tsai; Benafsh Sapra; Benjamin Rambo-Martin; Christopher Gulvick; Clinton Paden; Dakota Howard; Dhwani Batra; Doreen Ng; Duncan MacCannell; Erisa Sula; Harry Gao; James Xie; Jason Caravas; John Gao; Joseph Fierro; Kristine Lacek; Matthew Schmerer; Mickey Li; Peter Cook; Scott Sammons; Shatavia Morrison; Tymeckia Kendall; Victoria Caban Figueroa; Yan Meng; Yvette Unoarumhi |
| EPI_ISL_8527502, EPI_ISL_8527509, EPI_ISL_8527512, EPI_ISL_8527524, EPI_ISL_8527529, EPI_ISL_8527533, EPI_ISL_8527536, EPI_ISL_8527537, EPI_ISL_8527542, EPI_ISL_8527543, EPI_ISL_8527548, EPI_ISL_8527550, EPI_ISL_8527552, EPI_ISL_8527554, EPI_ISL_8527564, EPI_ISL_8527568, EPI_ISL_8527571, EPI_ISL_8527572, EPI_ISL_8527573, EPI_ISL_8527574, EPI_ISL_8527575, EPI_ISL_8527578, EPI_ISL_8527579, EPI_ISL_8527580, EPI_ISL_8527585, EPI_ISL_8527586, EPI_ISL_8527589, EPI_ISL_8527591 | | | |
| see above | Gandhi Medical College and Hospital (GMCH), Secunderabad | NIV Influenza | D.R.Manisha Rani; Dr.G.Sushma Rajya Lakshmi; Dr.K.Nagamani; Dr.Sunitha Pakalapaty |
| EPI_ISL_8547703, EPI_ISL_8547714, EPI_ISL_8547723, EPI_ISL_8547724, EPI_ISL_8547736, EPI_ISL_8547746, EPI_ISL_8547858, EPI_ISL_8547987, EPI_ISL_8548249, EPI_ISL_8548251, EPI_ISL_8548252, EPI_ISL_8548253, EPI_ISL_8548261, EPI_ISL_8548264 | | | |
| see above | Helix | Centers for Disease Control and Prevention Division of Viral Diseases, Pathogen Discovery | Benjamin Rambo-Martin; Christopher Gulvick; Clinton Paden; Dakota Howard; Dhwani Batra; Duncan MacCannell; Erisa Sula; Helix CA; Jason Caravas; Kristine Lacek; Matthew Schmerer; Peter Cook; Scott Sammons; Shatavia Morrison; Tymeckia Kendall; Victoria Caban Figueroa; Yvette Unoarumhi |
| EPI_ISL_8553407 | Hospital Universitari Vall d'Hebron - Vall d'Hebron Institut de Recerca | Hospital Universitari Vall d'Hebron - Vall d'Hebron Institut de Recerca | Alejandra González-Sánchez; Andrés Antón; Ariadna Rando; Carla Castillo; Cristina Andrés; Damir Garcia-Cehic; Josep Quer; Juliana Esperalba; Karen García; Maria Carmen Martin; Maria Gema Codina; Maria Piñana; Rodrigo Vásquez; Tomàs Pumarola |
| EPI_ISL_8525201, EPI_ISL_8525219, EPI_ISL_8525234 | Indira Gandhi Memorial Hospital | Indira Gandhi Memorial Hospital | D. Fathmath Nazla Rafeeq; Dr. Ibrahim Afzal; Mr. Ibrahim Nishan Ahmed; Ms. Aishath Shuhudha; Ms. Aminath Shazleena Abdul Rahman; Ms. Soafy Mohamed |
| EPI_ISL_8505377 | Infinity Biologix | Centers for Disease Control and Prevention Division of Viral Diseases, Pathogen Discovery | Benjamin Rambo-Martin; Chirayu Goswami; Christian Bixby; Christopher Gulvick; Clinton Paden; Dakota Howard; Dhwani Batra; Duncan MacCannell; Erisa Sula; Jason Caravas; Jonathan Schultz; Kristine Lacek; Matthew Schmerer; Peter Cook; Robin Grimwood; Russ Hager; Scott Sammons; Shatavia Morrison; Tymeckia Kendall; Victoria Caban Figueroa; Yihe Wang; Yvette Unoarumhi |
| EPI_ISL_8538550 | Institute for Infectious Diseases | Institute for Infectious Diseases, University of Bern | Alban Ramette; Christian Baumann; Cora Sägesser; Franziska Suter-Riniker; Loïc Borcard; Miguel A Terrazos Miani; Nicole Liechti; Pascal Bittel; Peter Keller; Sonja Gempeler; Stefan Neuenschwander; Stephen L Leib |
| EPI_ISL_8519789 | Interpid Nepal | National Public Health Laboratory | National Public Health Labaoratory Team/Intrepid Lab Team |
| EPI_ISL_8519794 | Kathmandu Medical College | National Public Health Laboratory | National Public Health Labaortory Team/Kathmandu Medical College Team |
| EPI_ISL_8515274, EPI_ISL_8515275, EPI_ISL_8515276, EPI_ISL_8515279, EPI_ISL_8515286, EPI_ISL_8515287, EPI_ISL_8515289, EPI_ISL_8515293, EPI_ISL_8515302, EPI_ISL_8515324, EPI_ISL_8515325, EPI_ISL_8515332, EPI_ISL_8515342, EPI_ISL_8515348, EPI_ISL_8515349, EPI_ISL_8515377, EPI_ISL_8515381, EPI_ISL_8515382, EPI_ISL_8515383, EPI_ISL_8515398, EPI_ISL_8515399, EPI_ISL_8515401, EPI_ISL_8515402, EPI_ISL_8515410, EPI_ISL_8515411, EPI_ISL_8515412, EPI_ISL_8515414, EPI_ISL_8515415, EPI_ISL_8515422, EPI_ISL_8515423, EPI_ISL_8515428, EPI_ISL_8515432, EPI_ISL_8515433, EPI_ISL_8515435, EPI_ISL_8515436, EPI_ISL_8515438, EPI_ISL_8515439, EPI_ISL_8515441, EPI_ISL_8515443, EPI_ISL_8515445, EPI_ISL_8515448, EPI_ISL_8515451, EPI_ISL_8515452, EPI_ISL_8515455, EPI_ISL_8515457, EPI_ISL_8515464, EPI_ISL_8515467, EPI_ISL_8515468, EPI_ISL_8515470, EPI_ISL_8515472, EPI_ISL_8515473, EPI_ISL_8515475, EPI_ISL_8515477, EPI_ISL_8544443, EPI_ISL_8544444, EPI_ISL_8544445, EPI_ISL_8544446, EPI_ISL_8544447, EPI_ISL_8544448, EPI_ISL_8544449, EPI_ISL_8544450, EPI_ISL_8544451, EPI_ISL_8544452, EPI_ISL_8544453, EPI_ISL_8544454, EPI_ISL_8544455, EPI_ISL_8544456, EPI_ISL_8544457, EPI_ISL_8544458, EPI_ISL_8544463, EPI_ISL_8544464, EPI_ISL_8544465, EPI_ISL_8544466, EPI_ISL_8544467, EPI_ISL_8544468, EPI_ISL_8544469, EPI_ISL_8544470, EPI_ISL_8544471, EPI_ISL_8544472, EPI_ISL_8544473, EPI_ISL_8544474, EPI_ISL_8544475 | | | |
| see above | Lab. Microbiologia e Virologia Cotugno A.O. dei Colli | TIGEM | Antonio Grimaldi Patrizia Annunziata Francesco Panariello Claudia Tiberio Teresa Giuliano Valentina Bouche Chiara Colantuono Michela Daniele Lucio Di Filippo Anna Manfredi Marcello Salvi Antonio Limone Luigi Atripaldi Andrea Ballabio Davide Cacchiarelli |
| EPI_ISL_8546646, EPI_ISL_8546647 | Labo Analyses Med | National Reference Center for Viruses of Respiratory Infections, Institut Pasteur, Paris | Angela Brisebarre; Camille Capel; Christophe Malabat; Corinne Maufrais; Etienne Simon-Lorière; Fabienne HUET; Frédéric Lemoine; Julien Fumey; Laurence PROTS; Louise Lefrançois; Marion Barbet; Maud Vanpeene; Méline Bizard; Slim El Khiari; Sylvie Behillil; Sylvie Van der Werf; Vincent Enouf |
| EPI_ISL_8547156 | Laboratorio Analisi Osp. Città di Castello - Azienda USL Umbria1 | Istituto Zooprofilattico Sperimentale dell'Abruzzo e Molise "G. Caporale" | Ancora M; Biagetti M; Calistri P; Cammà C; Curini V; Delli Compagni E; Di Domenico M; Di Pasquale A; Giammarioli M; Lorusso A; Malagigi V; Mangone I; Marcacci M; Puglia I; Rinaldi A; Savini G; Scialabba S; Tacconi P |
| EPI_ISL_8533328, EPI_ISL_8533337, EPI_ISL_8533338, EPI_ISL_8533359 | Lighthouse Lab in Glasgow | Wellcome Sanger Institute for the COVID-19 Genomics UK (COG-UK) Consortium | Anna Dominiczak and Alex Alderton; Carol Clugston; Cordelia Langford; David Gray; David K. Jackson; Dominic Kwiatkowski; Ewan Harrison; Harper VanSteenhouse; Ian Johnston; Jeffrey Barrett; John Sillitoe on behalf of the Wellcome Sanger Institute COVID-19 Surveillance Team; Roberto Amato; Sonia Goncalves; Yumi Kasai |
| EPI_ISL_8523907 | Medical Virology and BSL3 Laboratory | Centre de Séquençage Génomique | Abbad Anas; Amalou Ghita; Anga Latifa; Barakat Abdelhamid; Bouzidi Aymane; Charoute Hicham; Chgouri Fatima; Dersi Noureddine; EL Hamouchi Adil; El Oualid Abdelmjid; Faouzi Abdellah; Harmak Houda; Maaroufi Abderrahmane; Nadifiyine Saloua; Nourlil Jalal; Omondi Francis Carey; Somda Soro Georgina Charlene; Zemmouri Faouzia; Zouheir Yassine |
| EPI_ISL_8556454, EPI_ISL_8556456, EPI_ISL_8556458, EPI_ISL_8556473, EPI_ISL_8556502 | Michigan Department of Health and Human Services, Bureau of Laboratories | Michigan Department of Health and Human Services, Bureau of Laboratories | Blankenship HM; Riner D; Soehnlen MK |
| EPI_ISL_8513840, EPI_ISL_8513858, EPI_ISL_8513930, EPI_ISL_8514024, EPI_ISL_8514025, EPI_ISL_8514026, EPI_ISL_8514105 | | | |
| see above | Ministry of Health Turkey | Ministry of Health Turkey | Fatma Bayrakdar; Gulay Korukluoglu; Suleyman Yalcin; Yasemin Cosgun |
| EPI_ISL_8509383, EPI_ISL_8509390, EPI_ISL_8509403, EPI_ISL_8509405, EPI_ISL_8509416, EPI_ISL_8509420 | Molecular Biology Laboratory, Vidant Medical Center | Brody Integrative Genomics Core, East Carolina University | Changhong Yin; Heather Duncan; John T. Fallon; Kimberly P. Briley; Weihua Huang |
| EPI_ISL_8540327, EPI_ISL_8540328 | National Hematology and Transfusiology Center, Department of Medical Genetics | National Hematology and Transfusiology Center, Department of Medical Genetics | Agha Rza Aghayev |
| EPI_ISL_8519771 | National Public Health Laboratory | National Public Health Laboratory | National Public Health Labaoratory Team |
| EPI_ISL_8519790 | Nepal Korea Friendship Hospital | National Public Health Laboratory | National Public Health Labaoratory Team/Nepal Korea Friendship Hospital Team |
| EPI_ISL_8519772 | PPHL-5 | National Public Health Laboratory | National Public Health Labaoratory Team//Provincial Public Health Laboratory -5 COVID Lab Team |
| EPI_ISL_8519773 | Patan Hospital | National Public Health Laboratory | National Public Health Labaoratory Team /PATAN Hospital |
| EPI_ISL_8557231, EPI_ISL_8557382 | Quest Diagnostics Incorporated | Centers for Disease Control and Prevention Division of Viral Diseases, Pathogen Discovery | A. Gerasimova; A. Perez; B. Anderson; Benjamin Rambo-Martin; Christopher Gulvick; Clinton Paden; Dakota Howard; Dhwani Batra; Duncan MacCannell; Erisa Sula; F. Lacbawan; I. Shlyakhter; Jason Caravas; K. Livingston; Kristine Lacek; L. Bernstein; M. Hua; Matthew Schmerer; P. Tanpaiboon; Peter Cook; R. Kagan; R. Owen; R. Rolando; S. Rosenthal; Scott Sammons; Shatavia Morrison; Tymeckia Kendall; Victoria Caban Figueroa; Y. Liu; Yvette Unoarumhi |
| EPI_ISL_8488281, EPI_ISL_8488283, EPI_ISL_8488289, EPI_ISL_8488292, EPI_ISL_8517222, EPI_ISL_8517384, EPI_ISL_8517424, EPI_ISL_8539753, EPI_ISL_8539761, EPI_ISL_8539773, EPI_ISL_8539784, EPI_ISL_8539787, EPI_ISL_8539864, EPI_ISL_8539916 | | | |
| see above | Respiratory Virus Unit, Microbiology Services Colindale, Public Health England | COVID-19 Genomics UK (COG-UK) Consortium | PHE Covid Sequencing Team |
| EPI_ISL_8519791 | Shahid Gangalal Heart Center | National Public Health Laboratory | National Public Health Labaoratory Team/Shahid Gangalal Heart center Team |
| EPI_ISL_8519774, EPI_ISL_8519776, EPI_ISL_8519777, EPI_ISL_8519779, EPI_ISL_8519780, EPI_ISL_8519781, EPI_ISL_8519782, EPI_ISL_8519783, EPI_ISL_8519784, EPI_ISL_8519785, EPI_ISL_8519786, EPI_ISL_8519787, EPI_ISL_8519788 | | | |

| | | | |
|---|---|---|---|
| see above | Shree Birendra Hospital | National Public Health Laboratory | National Public Health Labaoratory Team/Shree Birendra Hospital COVID -19 Lab team |
| EPI_ISL_8494337 | Supratech Micropath Laboratory Research Institute Pvt Ltd, Ahmedabad | Gujarat Biotechnology Research Centre | Akhilesh Modi; Apurvasinh Puvar; Bhadreshsinh Gohil; Chaitanya Joshi; Disha Vora; Janvi Raval; Jaykumar Rangani; Madhvi Joshi; Nimesh Patel; Nitin Savaliya; Nitin Shukla; Priyank Chavda; Ramesh Pandit; Roshani Mishra; Shiva; Sonal Sharma; Tasnim Trivedi; Zarna Patel |
| EPI_ISL_8487263, EPI_ISL_8487371 | The Jackson Laboratory | The Jackson Laboratory | Adams M; Kelly K; Li L; Long E; Omerza G; Renzette N |
| EPI_ISL_8519196, EPI_ISL_8519205, EPI_ISL_8519227 | UW Virology Lab | UW Virology Lab | Alexander Greninger; Hong Xie; Isabel Arnould; Keith R Jerome; Meei-Li Huang; Nathan Breit; Patrick Mathias; Pavitra Roychoudhury; Pooneh Hajian; Ricardo Perez; Robert J. Livingston; Sean Ellis; Seffir T. Wendm; Shah Mohamed Bakhash |
| EPI_ISL_8519054 | University of New Mexico Hospital | Center for Global Health, University of New Mexico Health Sciences Center | Darrell Dinwiddie; Daryl Domman; Jesse Young; Jon Femling; Justin Bacca; Kurt Schwalm; Valerie Morley |
| EPI_ISL_8543599 | University of Wisconsin-Madison AIDS Vaccine Research Laboratories | University of Wisconsin-Madison AIDS Vaccine Research Laboratories | Gage Moreno; Katarina Braun; et al. AIDS Vaccine Research Laboratories |

We gratefully acknowledge the following Authors from the Originating laboratories responsible for obtaining the specimens, as well as the Submitting laboratories where the genome data were generated and shared via GISAID, on which this research is based.

All Submitters of data may be contacted directly via www.gisaid.org

Authors are sorted alphabetically.

Acknowledgement EPI_SET Identifier: EPI_SET_20220527ws

| Accession ID | Originating Laboratory | Submitting Laboratory | Authors |
|---|---|---|---|
| EPI_ISL_8568509 | Arcispedale Santa Maria Nuova Autoimmunita Allergologia e Biotecnologie Innovative"" | Istituto Zooprofilattico Sperimentale della Lombardia e dell'Emilia Romagna (IZSLER), Risk Analysis and Genomic Epidemiology Unit | Erika Scaltriti; Ilaria Menozzi; Marina Morganti; Stefano Pongolini |
| EPI_ISL_8568510 | Azienda Sanitaria Locale di Piacenza - Presidio Ospedaliero - Laboratorio di Microbiologia"" | Istituto Zooprofilattico Sperimentale della Lombardia e dell'Emilia Romagna (IZSLER), Risk Analysis and Genomic Epidemiology Unit | Erika Scaltriti; Ilaria Menozzi; Marina Morganti; Stefano Pongolini |
| EPI_ISL_8564603, EPI_ISL_8564609, EPI_ISL_8564615, EPI_ISL_8564665, EPI_ISL_8564674, EPI_ISL_8564687, EPI_ISL_8564697, EPI_ISL_8564713, EPI_ISL_8564730, EPI_ISL_8564732, EPI_ISL_8564744, EPI_ISL_8564752, EPI_ISL_8564763, EPI_ISL_8564766, EPI_ISL_8564773, EPI_ISL_8564778, EPI_ISL_8564782, EPI_ISL_8564817, EPI_ISL_8564830, EPI_ISL_8564842, EPI_ISL_8564879, EPI_ISL_8564904, EPI_ISL_8564911, EPI_ISL_8564917, EPI_ISL_8564928, EPI_ISL_8564932, EPI_ISL_8564933, EPI_ISL_8564935, EPI_ISL_8565526, EPI_ISL_8565533, EPI_ISL_8565535, EPI_ISL_8565576, EPI_ISL_8565588, EPI_ISL_8565589, EPI_ISL_8565591 | | | |
| see above | Curative Labs | Curative Labs | Elias L. Salfati; Eugenia Khorosheva; George Way; J.Cesar Ignacio-Espinoza; Janet Chen; Mikhail Hanewich-Hollatz; Nabjot Sandhu; Sophia Quasem; Vladimir Slepnev; Zhiyi Xie |
| EPI_ISL_8578158, EPI_ISL_8578167, EPI_ISL_8578168, EPI_ISL_8578177, EPI_ISL_8578182, EPI_ISL_8578189, EPI_ISL_8578190, EPI_ISL_8578202, EPI_ISL_8578203, EPI_ISL_8578205, EPI_ISL_8578209, EPI_ISL_8578213, EPI_ISL_8578218, EPI_ISL_8578254, EPI_ISL_8578258, EPI_ISL_8578259, EPI_ISL_8578264, EPI_ISL_8578270, EPI_ISL_8578373, EPI_ISL_8578374, EPI_ISL_8578383, EPI_ISL_8578386, EPI_ISL_8578393, EPI_ISL_8578396, EPI_ISL_8578406, EPI_ISL_8578409, EPI_ISL_8578410, EPI_ISL_8578415, EPI_ISL_8578424, EPI_ISL_8578425, EPI_ISL_8578426, EPI_ISL_8578428, EPI_ISL_8578446, EPI_ISL_8578454, EPI_ISL_8578459, EPI_ISL_8578461, EPI_ISL_8578472, EPI_ISL_8578474, EPI_ISL_8578484, EPI_ISL_8578485, EPI_ISL_8578488, EPI_ISL_8578499, EPI_ISL_8578507, EPI_ISL_8578517, EPI_ISL_8578521, EPI_ISL_8578522, EPI_ISL_8578528, EPI_ISL_8578529, EPI_ISL_8578535, EPI_ISL_8578536, EPI_ISL_8578545, EPI_ISL_8578547, EPI_ISL_8578552, EPI_ISL_8578558, EPI_ISL_8578570, EPI_ISL_8578576, EPI_ISL_8578580, EPI_ISL_8578590, EPI_ISL_8578606, EPI_ISL_8578615, EPI_ISL_8578618, EPI_ISL_8578622, EPI_ISL_8578629, EPI_ISL_8578631, EPI_ISL_8578654, EPI_ISL_8578660, EPI_ISL_8578672, EPI_ISL_8578675, EPI_ISL_8578677, EPI_ISL_8578678, EPI_ISL_8578680, EPI_ISL_8578686, EPI_ISL_8578689, EPI_ISL_8578695, EPI_ISL_8578699, EPI_ISL_8578706, EPI_ISL_8578721, EPI_ISL_8578723, EPI_ISL_8578735, EPI_ISL_8578742, EPI_ISL_8578747, EPI_ISL_8578748, EPI_ISL_8578753, EPI_ISL_8578761, EPI_ISL_8578769, EPI_ISL_8578770, EPI_ISL_8578807, EPI_ISL_8578811, EPI_ISL_8578814, EPI_ISL_8578815, EPI_ISL_8578823, EPI_ISL_8578832, EPI_ISL_8578843, EPI_ISL_8578844, EPI_ISL_8578849, EPI_ISL_8578866, EPI_ISL_8578868, EPI_ISL_8578870, EPI_ISL_8578874, EPI_ISL_8578885, EPI_ISL_8578892, EPI_ISL_8578896, EPI_ISL_8578908, EPI_ISL_8578912, EPI_ISL_8578933, EPI_ISL_8578937, EPI_ISL_8578938, EPI_ISL_8578943, EPI_ISL_8578963, EPI_ISL_8578964, EPI_ISL_8578969, EPI_ISL_8578982, EPI_ISL_8578989, EPI_ISL_8578998, EPI_ISL_8579013, EPI_ISL_8579015, EPI_ISL_8579017, EPI_ISL_8579019, EPI_ISL_8579028, EPI_ISL_8579030, EPI_ISL_8579031, EPI_ISL_8579033, EPI_ISL_8579034, EPI_ISL_8579039, EPI_ISL_8579045, EPI_ISL_8579046, EPI_ISL_8579047, EPI_ISL_8579056, EPI_ISL_8579058, EPI_ISL_8579065, EPI_ISL_8579078, EPI_ISL_8579083, EPI_ISL_8579103, EPI_ISL_8579112, EPI_ISL_8579116, EPI_ISL_8579125, EPI_ISL_8579130, EPI_ISL_8579145, EPI_ISL_8579151, EPI_ISL_8579152, EPI_ISL_8579154, EPI_ISL_8579158, EPI_ISL_8579161, EPI_ISL_8579164, EPI_ISL_8579167, EPI_ISL_8579172, EPI_ISL_8579180, EPI_ISL_8579181, EPI_ISL_8579186, EPI_ISL_8579187, EPI_ISL_8579190, EPI_ISL_8579211, EPI_ISL_8579222, EPI_ISL_8579228, EPI_ISL_8579242, EPI_ISL_8579244, EPI_ISL_8579246, EPI_ISL_8579248, EPI_ISL_8579253, EPI_ISL_8579260, EPI_ISL_8579263, EPI_ISL_8579280, EPI_ISL_8579281, EPI_ISL_8579289, EPI_ISL_8579298, EPI_ISL_8579303, EPI_ISL_8579304, EPI_ISL_8579310, EPI_ISL_8579327, EPI_ISL_8579333, EPI_ISL_8579338, EPI_ISL_8579339, EPI_ISL_8579349, EPI_ISL_8579351, EPI_ISL_8579353, EPI_ISL_8579357, EPI_ISL_8579359, EPI_ISL_8579363, EPI_ISL_8579367, EPI_ISL_8579372, EPI_ISL_8579376, EPI_ISL_8579382, EPI_ISL_8579392, EPI_ISL_8579399, EPI_ISL_8579403, EPI_ISL_8579406, EPI_ISL_8579407, EPI_ISL_8579411, EPI_ISL_8579414, EPI_ISL_8579416, EPI_ISL_8579419, EPI_ISL_8579426, EPI_ISL_8579429, EPI_ISL_8579440, EPI_ISL_8579446, EPI_ISL_8579447, EPI_ISL_8579451, EPI_ISL_8579459, EPI_ISL_8579460, EPI_ISL_8579462, EPI_ISL_8579474, EPI_ISL_8579476, EPI_ISL_8579479, EPI_ISL_8579490, EPI_ISL_8579500, EPI_ISL_8579501, EPI_ISL_8579514, EPI_ISL_8579519, EPI_ISL_8579521, EPI_ISL_8579540, EPI_ISL_8579551, EPI_ISL_8579556, EPI_ISL_8579557, EPI_ISL_8579564, EPI_ISL_8579566, EPI_ISL_8579576, EPI_ISL_8579586, EPI_ISL_8579587, EPI_ISL_8579588, EPI_ISL_8579599, EPI_ISL_8579600, EPI_ISL_8579602, EPI_ISL_8579605, EPI_ISL_8579606, EPI_ISL_8579610, EPI_ISL_8579614, EPI_ISL_8579619, EPI_ISL_8579634, EPI_ISL_8579641, EPI_ISL_8579644, EPI_ISL_8579645, EPI_ISL_8579649, EPI_ISL_8579657, EPI_ISL_8579944, EPI_ISL_8580301, EPI_ISL_8580507, EPI_ISL_8580513, EPI_ISL_8580529, EPI_ISL_8580557, EPI_ISL_8580558, EPI_ISL_8580559, EPI_ISL_8580560, EPI_ISL_8580561, EPI_ISL_8580566, EPI_ISL_8580570, EPI_ISL_8580584, EPI_ISL_8580606, EPI_ISL_8580610, EPI_ISL_8580617, EPI_ISL_8580639, EPI_ISL_8580641, EPI_ISL_8580653, EPI_ISL_8580671, EPI_ISL_8580677, EPI_ISL_8580691, EPI_ISL_8580699, EPI_ISL_8580709, EPI_ISL_8580715, EPI_ISL_8580717, EPI_ISL_8580728, EPI_ISL_8580749, EPI_ISL_8580753, EPI_ISL_8580760, EPI_ISL_8580762, EPI_ISL_8580770, EPI_ISL_8580775, EPI_ISL_8580795, EPI_ISL_8580801, EPI_ISL_8580806, EPI_ISL_8580810, EPI_ISL_8580822, EPI_ISL_8580826, EPI_ISL_8580841, EPI_ISL_8580845, EPI_ISL_8580846, EPI_ISL_8580847, EPI_ISL_8580853, EPI_ISL_8580866, EPI_ISL_8580871, EPI_ISL_8580880, EPI_ISL_8580881, EPI_ISL_8580886, EPI_ISL_8580892, EPI_ISL_8580894, EPI_ISL_8580898, EPI_ISL_8580900, EPI_ISL_8580904, EPI_ISL_8580908, EPI_ISL_8580912, EPI_ISL_8580917, EPI_ISL_8580919, EPI_ISL_8580929, EPI_ISL_8580933, EPI_ISL_8580944, EPI_ISL_8580948, EPI_ISL_8580950, EPI_ISL_8580958, EPI_ISL_8580961, EPI_ISL_8580970, EPI_ISL_8580994, EPI_ISL_8581005, EPI_ISL_8581009, EPI_ISL_8581019, EPI_ISL_8581045, EPI_ISL_8581050, EPI_ISL_8581070, EPI_ISL_8581079, EPI_ISL_8581081, EPI_ISL_8581091, EPI_ISL_8581092, EPI_ISL_8581097, EPI_ISL_8581100, EPI_ISL_8581103, EPI_ISL_8581142, EPI_ISL_8581143, EPI_ISL_8581145, EPI_ISL_8581150, EPI_ISL_8581152, EPI_ISL_8581153, EPI_ISL_8581154, EPI_ISL_8581161, EPI_ISL_8581167, EPI_ISL_8581183, EPI_ISL_8581186, EPI_ISL_8581192, EPI_ISL_8581195, EPI_ISL_8581199, EPI_ISL_8581201, EPI_ISL_8581206, EPI_ISL_8581208, EPI_ISL_8581210, EPI_ISL_8581212, EPI_ISL_8581222, EPI_ISL_8581227, EPI_ISL_8581229, EPI_ISL_8581232, EPI_ISL_8581245, EPI_ISL_8581254, EPI_ISL_8581256, EPI_ISL_8581260, EPI_ISL_8581263, EPI_ISL_8581265, EPI_ISL_8581272, EPI_ISL_8581275, EPI_ISL_8581277, EPI_ISL_8581286, EPI_ISL_8581291, EPI_ISL_8581302, EPI_ISL_8581317, EPI_ISL_8581323, EPI_ISL_8581331, EPI_ISL_8581334, EPI_ISL_8581339, EPI_ISL_8581341, EPI_ISL_8581344, EPI_ISL_8581347, EPI_ISL_8581348, EPI_ISL_8581351, EPI_ISL_8581353, EPI_ISL_8581358, EPI_ISL_8581361, EPI_ISL_8581373, EPI_ISL_8581376, EPI_ISL_8581386, EPI_ISL_8581388, EPI_ISL_8581421, EPI_ISL_8581426 | | | |
| see above | Department of Bacteria, Parasites and Fungi, Statens Serum Institut, Copenhagen, Denmark | Statens Serum Institut Bioinformatics and Microbial Genomics | Danish Covid-19 Genome Consortium |
| EPI_ISL_8578469 | Department of Clinical Microbiology and Center for Genomic Medicine, Rigshospitalet, Copenhagen, Denmark | Statens Serum Institut Bioinformatics and Microbial Genomics | Danish Covid-19 Genome Consortium |
| EPI_ISL_8565890, EPI_ISL_8565986, EPI_ISL_8566049, EPI_ISL_8566068, EPI_ISL_8566070, EPI_ISL_8566074, EPI_ISL_8566093, EPI_ISL_8566124, EPI_ISL_8566163, EPI_ISL_8566164, EPI_ISL_8566165, EPI_ISL_8566166, EPI_ISL_8566185, EPI_ISL_8566201, EPI_ISL_8566202, EPI_ISL_8566204, EPI_ISL_8566205, EPI_ISL_8566206, EPI_ISL_8566208, EPI_ISL_8566219, EPI_ISL_8566246, EPI_ISL_8566247, EPI_ISL_8566248, EPI_ISL_8566249, EPI_ISL_8568575, EPI_ISL_8568637, EPI_ISL_8568644, EPI_ISL_8568648, EPI_ISL_8568685, EPI_ISL_8568711, EPI_ISL_8568716, EPI_ISL_8568719, EPI_ISL_8568737, EPI_ISL_8568757, EPI_ISL_8568842, EPI_ISL_8568853, EPI_ISL_8568868, EPI_ISL_8568887, EPI_ISL_8568910, EPI_ISL_8568927, EPI_ISL_8568932, EPI_ISL_8568939, EPI_ISL_8568951, EPI_ISL_8568961, EPI_ISL_8568962, EPI_ISL_8568972 | | | |
| see above | Institute of Microbiology and Immunology, Faculty of Medicine, University of Ljubljana | Institute of Microbiology and Immunology, Faculty of Medicine, University of Ljubljana | Alen Suljič; Andraž Celar; Domen Lazar; Doroteja Vlaj; Mario Poljak; Miša Korva; Patricija Pozvek; Samo Zakotnik; Tatjana Avšič – Županc; Tina Gabrovšek; Tina Živič; Tomaž Mark Zorec; Špela Pleh |
| EPI_ISL_8576550, EPI_ISL_8576583, EPI_ISL_8576592, EPI_ISL_8576739, EPI_ISL_8576806, EPI_ISL_8576817, EPI_ISL_8576841, EPI_ISL_8576860, EPI_ISL_8576897, EPI_ISL_8576928, EPI_ISL_8576955, EPI_ISL_8576982, EPI_ISL_8577004, EPI_ISL_8577008, EPI_ISL_8577051, EPI_ISL_8577066 | | | |
| see above | Molecular Biology Laboratory, Vidant Medical Center | Brody Integrative Genomics Core, East Carolina University | Changhong Yin; Heather Duncan; John T. Fallon; Kimberly P. Briley; Weihua Huang |
| EPI_ISL_8562554, EPI_ISL_8562558, EPI_ISL_8562560, EPI_ISL_8562561, EPI_ISL_8562562, EPI_ISL_8562566, EPI_ISL_8562592, EPI_ISL_8562593, EPI_ISL_8562594, EPI_ISL_8562638, EPI_ISL_8562640, EPI_ISL_8562661, EPI_ISL_8562665, EPI_ISL_8566891 | | | |
| see above | National Public Health Laboratory, National Centre for Infectious Diseases | National Public Health Laboratory, National Centre for Infectious Diseases | Benny Yeo; Lin Cui; Raymond Tzer Pin Lin; Royce Ang; Samuel Loo; Zhenyang Zhou |
| EPI_ISL_8559646, EPI_ISL_8559670, EPI_ISL_8559900, EPI_ISL_8560013, EPI_ISL_8560180, EPI_ISL_8560294, EPI_ISL_8560301, EPI_ISL_8560339, EPI_ISL_8560340, EPI_ISL_8560342, EPI_ISL_8560357, EPI_ISL_8560423, EPI_ISL_8560425, EPI_ISL_8560428, EPI_ISL_8560490, EPI_ISL_8560498, EPI_ISL_8560500 | | | |
| see above | SARS-CoV-2 testing team, National Institute of Infectious Diseases | Pathogen Genomics Center, National Institute of Infectious Diseases | Hazuka Y Furihata; Kentaro Itokawa; Makoto Kuroda; Masanori Hashino; Masumichi Saito; Naomi Nojiri; Nozomu Hanaoka; Rina Tanaka; Tsuguto Fujimoto; Tsuyoshi Sekizuka |
| EPI_ISL_8565215 | University of New Mexico Hospital | Center for Global Health, University of New Mexico Health Sciences Center | Darrell Dinwiddie; Daryl Domman; Jesse Young; Jon Femling; Justin Bacca; Kurt Schwalm; Valerie Morley |

We gratefully acknowledge the following Authors from the Originating laboratories responsible for obtaining the specimens, as well as the Submitting laboratories where the genome data were generated and shared via GISAID, on which this research is based.

All Submitters of data may be contacted directly via www.gisaid.org

Authors are sorted alphabetically.

| Accession ID | Originating Laboratory | Submitting Laboratory | Authors |
|---|---|---|---|
| EPI_ISL_8593661 | City of Milwaukee Health Department Laboratory | City of Milwaukee Health Department Laboratory | Amy Bauer; Joshua Weiner; Manjeet Khubbar; Nandhakumar Balakrishnan; Sanjib Bhattacharyya |
| EPI_ISL_8594433, EPI_ISL_8594435, EPI_ISL_8594436, EPI_ISL_8594437 | Cliniques universitaires Saint-Luc | UCLouvain/IREC/MBLG-CTMA | Benoit Kabamba Mukadi; Bertrand Bearzatto; Jean-Luc Gala; Nicolas Pinte; Paul Blanpain; Simon Ophélie; Valentin Coste |
| EPI_ISL_8583616, EPI_ISL_8583617, EPI_ISL_8583622, EPI_ISL_8583623, EPI_ISL_8583628, EPI_ISL_8583629, EPI_ISL_8583631, EPI_ISL_8583633, EPI_ISL_8583638, EPI_ISL_8583640, EPI_ISL_8583641, EPI_ISL_8583642, EPI_ISL_8583646, EPI_ISL_8583654, EPI_ISL_8583670, EPI_ISL_8583675, EPI_ISL_8583683, EPI_ISL_8583686, EPI_ISL_8583690, EPI_ISL_8583701, EPI_ISL_8583704, EPI_ISL_8583707, EPI_ISL_8583709, EPI_ISL_8583710, EPI_ISL_8583714, EPI_ISL_8583718, EPI_ISL_8583719, EPI_ISL_8583720, EPI_ISL_8583729, EPI_ISL_8583733, EPI_ISL_8583739, EPI_ISL_8583744 | | | |
| see above | Department of Bacteria, Parasites and Fungi, Statens Serum Institut, Copenhagen, Denmark | Statens Serum Institut Bioinformatics and Microbial Genomics | Danish Covid-19 Genome Consortium |
| EPI_ISL_8596354 | Edmonton Provincial Lab | Public Health Agency of Canada (PHAC) National Microbiology Laboratory | Buss; Croxen M; Deo A; Dieu P; E; Ferrato C; Gill K; Khan F; Koleva P; Li V; Lloyd C; Lynch T; Ma R; Murphy S; Pabbaraju K; Shokoples S; Thayer J; Tipples G; Whitehouse M; Wong A; Yu C; Zelyas N |
| EPI_ISL_8593220 | Fulgent Genetics | Centers for Disease Control and Prevention Division of Viral Diseases, Pathogen Discovery | Becky Tsai; Benafsh Sapra; Benjamin Rambo-Martin; Christopher Gulvick; Clinton Paden; Dakota Howard; Dhwani Batra; Doreen Ng; Duncan MacCannell; Erisa Sula; Harry Gao; James Xie; Jason Caravas; John Gao; Joseph Fierro; Kristine Lacek; Matthew Schmerer; Mickey Li; Peter Cook; Scott Sammons; Shatavia Morrison; Tymeckia Kendall; Victoria Caban Figueroa; Yan Meng; Yvette Unoarumhi |
| EPI_ISL_8589447 | Jessa | Jessa | Severine Berden et al. on behalf of the Jessa_cmdLab |
| EPI_ISL_8593606, EPI_ISL_8593607, EPI_ISL_8593608 | Kansas Health and Environmental Lab | Kansas Health and Environmental Lab | Amanda Bradley; Carrie Welch; Gary Buruss; Jonathan Barnell; Mike Grose; Victor Anderson; and Phil Adam |
| EPI_ISL_8591441, EPI_ISL_8591446, EPI_ISL_8591447 | Michigan Department of Health and Human Services, Bureau of Laboratories | Michigan Department of Health and Human Services, Bureau of Laboratories | Blankenship HM; Riner D; Soehnlen MK |
| EPI_ISL_8584134 | Mon Health Medical Center | WVU and Marshall University Combined Genomics Core Facilities | James Denvir; Peter Perrotta; Peter Stoilov; Ryan Percifield; Wesley Kimble |
| EPI_ISL_8593976 | Pandemic Response Lab - NYC | Pandemic Response Lab, R&D | Alex Carpio; Cybill del Castillo; Dylan Law; Haiping Hao; Henry Lee; Isabel Fernandez Escapa; Jon Laurent; Melissa Hopkins; Michael Hammerling; Pradeep Bugga; Shinyoung Clair Kang; Sol Rey; William Ward |
| EPI_ISL_8594403 | Regional Medical Sciences Center 7 Khon Kaen | National Institute of Health, Department of Medical Sciences, Ministry of Public Health, Thailand | Archawin Rojanawiwat; Natchaya Khiadsang; Nuttida Thongpramul; Pakorn Piromtong; Pilailuk Okada; Sirikanda Wimol; Siripaporn Phuygun; Sunthareeya Waicharoen; Suratchana Mitrat; Thanutsapa Thanadachakul |
| EPI_ISL_8594402 | World Medical Hospital | National Institute of Health, Department of Medical Sciences, Ministry of Public Health, Thailand | Archawin Rojanawiwat; Natchaya Khiadsang; Nuttida Thongpramul; Pakorn Piromtong; Pilailuk Okada; Sirikanda Wimol; Siripaporn Phuygun; Sunthareeya Waicharoen; Suratchana Mitrat; Thanutsapa Thanadachakul |

We gratefully acknowledge the following Authors from the Originating laboratories responsible for obtaining the specimens, as well as the Submitting laboratories where the genome data were generated and shared via GISAID, on which this research is based.

All Submitters of data may be contacted directly via www.gisaid.org

Authors are sorted alphabetically.

Acknowledgement EPI_SET Identifier: EPI_SET_20220527en

| Accession ID | Originating Laboratory | Submitting Laboratory | Authors |
|---|---|---|---|
| EPI_ISL_8583325, EPI_ISL_8583331, EPI_ISL_8583345, EPI_ISL_8583348, EPI_ISL_8583349, EPI_ISL_8583351, EPI_ISL_8583359, EPI_ISL_8583360, EPI_ISL_8583363, EPI_ISL_8583376, EPI_ISL_8583378, EPI_ISL_8583385, EPI_ISL_8583395, EPI_ISL_8583398, EPI_ISL_8583416, EPI_ISL_8583423, EPI_ISL_8583434, EPI_ISL_8583435, EPI_ISL_8583436, EPI_ISL_8583450, EPI_ISL_8583460, EPI_ISL_8583468, EPI_ISL_8583476, EPI_ISL_8583483, EPI_ISL_8583487, EPI_ISL_8583488, EPI_ISL_8583500, EPI_ISL_8583509, EPI_ISL_8583516, EPI_ISL_8583518, EPI_ISL_8583520, EPI_ISL_8583527, EPI_ISL_8583544, EPI_ISL_8583547, EPI_ISL_8583553, EPI_ISL_8583555, EPI_ISL_8583562, EPI_ISL_8583565, EPI_ISL_8583571, EPI_ISL_8583576, EPI_ISL_8583580, EPI_ISL_8583583, EPI_ISL_8583584, EPI_ISL_8583588, EPI_ISL_8583592, EPI_ISL_8583595, EPI_ISL_8583596, EPI_ISL_8583597, EPI_ISL_8583598, EPI_ISL_8583604 | | | |
| see above | Department of Bacteria, Parasites and Fungi, Statens Serum Institut, Copenhagen, Denmark | Statens Serum Institut Bioinformatics and Microbial Genomics | Danish Covid-19 Genome Consortium |

We gratefully acknowledge the following Authors from the Originating laboratories responsible for obtaining the specimens, as well as the Submitting laboratories where the genome data were generated and shared via GISAID, on which this research is based.

All Submitters of data may be contacted directly via www.gisaid.org

Authors are sorted alphabetically.

| Accession ID | Originating Laboratory | Submitting Laboratory | Authors |
|---|---|---|---|
| EPI_ISL_8582960, EPI_ISL_8582962, EPI_ISL_8582976, EPI_ISL_8582981, EPI_ISL_8582987, EPI_ISL_8582990, EPI_ISL_8582991, EPI_ISL_8582992, EPI_ISL_8582998, EPI_ISL_8583009, EPI_ISL_8583012, EPI_ISL_8583018, EPI_ISL_8583022, EPI_ISL_8583036, EPI_ISL_8583038, EPI_ISL_8583043, EPI_ISL_8583047, EPI_ISL_8583061, EPI_ISL_8583064, EPI_ISL_8583067, EPI_ISL_8583081, EPI_ISL_8583103, EPI_ISL_8583110, EPI_ISL_8583116, EPI_ISL_8583130, EPI_ISL_8583131, EPI_ISL_8583138, EPI_ISL_8583139, EPI_ISL_8583145, EPI_ISL_8583146, EPI_ISL_8583158, EPI_ISL_8583161, EPI_ISL_8583162, EPI_ISL_8583163, EPI_ISL_8583169, EPI_ISL_8583171, EPI_ISL_8583173, EPI_ISL_8583175, EPI_ISL_8583192, EPI_ISL_8583194, EPI_ISL_8583203, EPI_ISL_8583213, EPI_ISL_8583222, EPI_ISL_8583229, EPI_ISL_8583230, EPI_ISL_8583240, EPI_ISL_8583293, EPI_ISL_8583296, EPI_ISL_8583297, EPI_ISL_8583315 | | | |
| see above | Department of Bacteria, Parasites and Fungi, Statens Serum Institut, Copenhagen, Denmark | Statens Serum Institut Bioinformatics and Microbial Genomics | Danish Covid-19 Genome Consortium |

We gratefully acknowledge the following Authors from the Originating laboratories responsible for obtaining the specimens, as well as the Submitting laboratories where the genome data were generated and shared via GISAID, on which this research is based.

All Submitters of data may be contacted directly via www.gisaid.org

Authors are sorted alphabetically.

| Accession ID | Originating Laboratory | Submitting Laboratory | Authors |
|---|---|---|---|
| EPI_ISL_8582673, EPI_ISL_8582674, EPI_ISL_8582684, EPI_ISL_8582693, EPI_ISL_8582698, EPI_ISL_8582700, EPI_ISL_8582701, EPI_ISL_8582706, EPI_ISL_8582720, EPI_ISL_8582723, EPI_ISL_8582725, EPI_ISL_8582727, EPI_ISL_8582733, EPI_ISL_8582744, EPI_ISL_8582751, EPI_ISL_8582753, EPI_ISL_8582765, EPI_ISL_8582766, EPI_ISL_8582768, EPI_ISL_8582772, EPI_ISL_8582775, EPI_ISL_8582777, EPI_ISL_8582782, EPI_ISL_8582785, EPI_ISL_8582786, EPI_ISL_8582788, EPI_ISL_8582795, EPI_ISL_8582801, EPI_ISL_8582805, EPI_ISL_8582818, EPI_ISL_8582821, EPI_ISL_8582828, EPI_ISL_8582853, EPI_ISL_8582855, EPI_ISL_8582859, EPI_ISL_8582863, EPI_ISL_8582869, EPI_ISL_8582877, EPI_ISL_8582881, EPI_ISL_8582891, EPI_ISL_8582893, EPI_ISL_8582914, EPI_ISL_8582918, EPI_ISL_8582919, EPI_ISL_8582921, EPI_ISL_8582926, EPI_ISL_8582932, EPI_ISL_8582934, EPI_ISL_8582943, EPI_ISL_8582944 | | | |
| see above | Department of Bacteria, Parasites and Fungi, Statens Serum Institut, Copenhagen, Denmark | Statens Serum Institut Bioinformatics and Microbial Genomics | Danish Covid-19 Genome Consortium |

We gratefully acknowledge the following Authors from the Originating laboratories responsible for obtaining the specimens, as well as the Submitting laboratories where the genome data were generated and shared via GISAID, on which this research is based.

All Submitters of data may be contacted directly via www.gisaid.org

Authors are sorted alphabetically.

| Accession ID | Originating Laboratory | Submitting Laboratory | Authors |
|---|---|---|---|
| EPI_ISL_8582400, EPI_ISL_8582405, EPI_ISL_8582414, EPI_ISL_8582415, EPI_ISL_8582435, EPI_ISL_8582440, EPI_ISL_8582443, EPI_ISL_8582444, EPI_ISL_8582449, EPI_ISL_8582450, EPI_ISL_8582456, EPI_ISL_8582458, EPI_ISL_8582469, EPI_ISL_8582482, EPI_ISL_8582484, EPI_ISL_8582487, EPI_ISL_8582497, EPI_ISL_8582503, EPI_ISL_8582504, EPI_ISL_8582510, EPI_ISL_8582516, EPI_ISL_8582518, EPI_ISL_8582527, EPI_ISL_8582530, EPI_ISL_8582534, EPI_ISL_8582536, EPI_ISL_8582541, EPI_ISL_8582547, EPI_ISL_8582550, EPI_ISL_8582551, EPI_ISL_8582557, EPI_ISL_8582564, EPI_ISL_8582567, EPI_ISL_8582577, EPI_ISL_8582579, EPI_ISL_8582583, EPI_ISL_8582594, EPI_ISL_8582603, EPI_ISL_8582605, EPI_ISL_8582606, EPI_ISL_8582612, EPI_ISL_8582622, EPI_ISL_8582626, EPI_ISL_8582633, EPI_ISL_8582637, EPI_ISL_8582639, EPI_ISL_8582645, EPI_ISL_8582650, EPI_ISL_8582657, EPI_ISL_8582668 | | | |
| see above | Department of Bacteria, Parasites and Fungi, Statens Serum Institut, Copenhagen, Denmark | Statens Serum Institut Bioinformatics and Microbial Genomics | Danish Covid-19 Genome Consortium |

We gratefully acknowledge the following Authors from the Originating laboratories responsible for obtaining the specimens, as well as the Submitting laboratories where the genome data were generated and shared via GISAID, on which this research is based.

All Submitters of data may be contacted directly via www.gisaid.org

Authors are sorted alphabetically.

Acknowledgement EPI_SET Identifier: EPI_SET_20220527sq

| Accession ID | Originating Laboratory | Submitting Laboratory | Authors |
| --- | --- | --- | --- |
| EPI_ISL_8582142, EPI_ISL_8582172, EPI_ISL_8582178, EPI_ISL_8582187, EPI_ISL_8582188, EPI_ISL_8582200, EPI_ISL_8582202, EPI_ISL_8582204, EPI_ISL_8582214, EPI_ISL_8582217, EPI_ISL_8582218, EPI_ISL_8582219, EPI_ISL_8582224, EPI_ISL_8582238, EPI_ISL_8582240, EPI_ISL_8582244, EPI_ISL_8582245, EPI_ISL_8582247, EPI_ISL_8582248, EPI_ISL_8582249, EPI_ISL_8582252, EPI_ISL_8582258, EPI_ISL_8582259, EPI_ISL_8582265, EPI_ISL_8582271, EPI_ISL_8582272, EPI_ISL_8582273, EPI_ISL_8582274, EPI_ISL_8582275, EPI_ISL_8582279, EPI_ISL_8582290, EPI_ISL_8582296, EPI_ISL_8582304, EPI_ISL_8582307, EPI_ISL_8582308, EPI_ISL_8582318, EPI_ISL_8582321, EPI_ISL_8582327, EPI_ISL_8582328, EPI_ISL_8582329, EPI_ISL_8582330, EPI_ISL_8582337, EPI_ISL_8582344, EPI_ISL_8582347, EPI_ISL_8582351, EPI_ISL_8582357, EPI_ISL_8582358, EPI_ISL_8582362, EPI_ISL_8582378, EPI_ISL_8582393 | | | |
| see above | Department of Bacteria, Parasites and Fungi, Statens Serum Institut, Copenhagen, Denmark | Statens Serum Institut Bioinformatics and Microbial Genomics | Danish Covid-19 Genome Consortium |

We gratefully acknowledge the following Authors from the Originating laboratories responsible for obtaining the specimens, as well as the Submitting laboratories where the genome data were generated and shared via GISAID, on which this research is based.

All Submitters of data may be contacted directly via www.gisaid.org

Authors are sorted alphabetically.

| Accession ID | Originating Laboratory | Submitting Laboratory | Authors |
|---|---|---|---|
| EPI_ISL_8582031, EPI_ISL_8582034, EPI_ISL_8582057, EPI_ISL_8582091, EPI_ISL_8582107, EPI_ISL_8582113, EPI_ISL_8582136, EPI_ISL_8582139 | | | |
| see above | Department of Bacteria, Parasites and Fungi, Statens Serum Institut, Copenhagen, Denmark | Statens Serum Institut Bioinformatics and Microbial Genomics | Danish Covid-19 Genome Consortium |

We gratefully acknowledge the following Authors from the Originating laboratories responsible for obtaining the specimens, as well as the Submitting laboratories where the genome data were generated and shared via GISAID, on which this research is based.

All Submitters of data may be contacted directly via www.gisaid.org

Authors are sorted alphabetically.

| Accession ID | Originating Laboratory | Submitting Laboratory | Authors |
|---|---|---|---|
| EPI_ISL_8604897, EPI_ISL_8604898, EPI_ISL_8604899, EPI_ISL_8604900, EPI_ISL_8604905, EPI_ISL_8604908, EPI_ISL_8604909, EPI_ISL_8604913, EPI_ISL_8604914, EPI_ISL_8604915, EPI_ISL_8604917, EPI_ISL_8604918, EPI_ISL_8604919, EPI_ISL_8604920, EPI_ISL_8604922, EPI_ISL_8604926, EPI_ISL_8604927, EPI_ISL_8604928, EPI_ISL_8604929, EPI_ISL_8604930, EPI_ISL_8604931, EPI_ISL_8604932, EPI_ISL_8604933, EPI_ISL_8604934, EPI_ISL_8604936 | | | |
| see above | AHRI | CERI, Centre for Epidemic Response and Innvoation, Stellenbosch University and KRISP, KZN Research Innovation and Sequencing Platform, UKZN. | Bernstein M; Cele S; Giandhari J; Karim F; Khan K; Moir M; Naidoo Y; Pillay S; Ramphal U; Ramphal Y; San JE; Sigal A; Tegally H; Tshiabuila D; Wilkinson E; de Oliveira T; van Wyk S |
| EPI_ISL_8607974, EPI_ISL_8608035, EPI_ISL_8608121 | Botswana Harvard HIV Reference Laboratory | Botswana Harvard HIV Reference Laboratory | Boitumelo Zuze; Botshelo Radibe; Dorcas Maruapula; Joseph Makhema; Keoratile Ntshambiwa; Kgomotso Moruisi; Legodile Kooepile; Mosepele Mosepele; Mphaphi B. Mbulawa; Ontlametse T. Bareng; Pamela Smith-Lawrence; Roger Shapiro; Sefetogi Ramaologa; Shahin Lockman; Sikhulile Moyo; Simani Gaseitsiwe; Thongbotho Mphoyakgosi; Wonderful T. Choga |
| EPI_ISL_8610825, EPI_ISL_8610838, EPI_ISL_8610866, EPI_ISL_8610875, EPI_ISL_8610879, EPI_ISL_8610887, EPI_ISL_8610904, EPI_ISL_8610948, EPI_ISL_8610955, EPI_ISL_8610958, EPI_ISL_8610972, EPI_ISL_8610981, EPI_ISL_8610986, EPI_ISL_8611012, EPI_ISL_8611027, EPI_ISL_8611046, EPI_ISL_8611058, EPI_ISL_8611061, EPI_ISL_8611069, EPI_ISL_8611085, EPI_ISL_8611124, EPI_ISL_8611140, EPI_ISL_8611164, EPI_ISL_8611173, EPI_ISL_8611208, EPI_ISL_8611258, EPI_ISL_8611275, EPI_ISL_8611293, EPI_ISL_8611321, EPI_ISL_8611339, EPI_ISL_8611349, EPI_ISL_8611361, EPI_ISL_8611365, EPI_ISL_8611377, EPI_ISL_8611379, EPI_ISL_8611394, EPI_ISL_8611453, EPI_ISL_8611493, EPI_ISL_8611500, EPI_ISL_8611528, EPI_ISL_8611532, EPI_ISL_8611540, EPI_ISL_8611547, EPI_ISL_8611550, EPI_ISL_8611569, EPI_ISL_8611599, EPI_ISL_8611605, EPI_ISL_8611616, EPI_ISL_8611629, EPI_ISL_8611641, EPI_ISL_8611655, EPI_ISL_8611658, EPI_ISL_8611667, EPI_ISL_8611670, EPI_ISL_8611685, EPI_ISL_8611690, EPI_ISL_8611698, EPI_ISL_8611735, EPI_ISL_8611759, EPI_ISL_8611770, EPI_ISL_8611775, EPI_ISL_8611778, EPI_ISL_8611783, EPI_ISL_8611787, EPI_ISL_8611796, EPI_ISL_8611805, EPI_ISL_8611810, EPI_ISL_8611816, EPI_ISL_8611818, EPI_ISL_8611819, EPI_ISL_8611827, EPI_ISL_8611840, EPI_ISL_8611857, EPI_ISL_8611864, EPI_ISL_8611889, EPI_ISL_8611895, EPI_ISL_8611898, EPI_ISL_8611904, EPI_ISL_8611913, EPI_ISL_8611917, EPI_ISL_8612031, EPI_ISL_8612035, EPI_ISL_8612043, EPI_ISL_8612056, EPI_ISL_8612070, EPI_ISL_8612074, EPI_ISL_8612078, EPI_ISL_8612089, EPI_ISL_8612100, EPI_ISL_8612111, EPI_ISL_8612159, EPI_ISL_8612161, EPI_ISL_8612167, EPI_ISL_8612171, EPI_ISL_8612183, EPI_ISL_8612200, EPI_ISL_8612203, EPI_ISL_8612216, EPI_ISL_8612224, EPI_ISL_8612271, EPI_ISL_8612275, EPI_ISL_8612284, EPI_ISL_8612301, EPI_ISL_8612306, EPI_ISL_8612323, EPI_ISL_8612325, EPI_ISL_8612341, EPI_ISL_8612346, EPI_ISL_8612371, EPI_ISL_8612379, EPI_ISL_8612410, EPI_ISL_8612412, EPI_ISL_8612430, EPI_ISL_8612450, EPI_ISL_8612465, EPI_ISL_8612472, EPI_ISL_8612491, EPI_ISL_8612500, EPI_ISL_8612529, EPI_ISL_8612531, EPI_ISL_8612533, EPI_ISL_8612540, EPI_ISL_8612546, EPI_ISL_8612547, EPI_ISL_8612573, EPI_ISL_8612577, EPI_ISL_8612580, EPI_ISL_8612583, EPI_ISL_8612588, EPI_ISL_8612591, EPI_ISL_8612607, EPI_ISL_8612617, EPI_ISL_8612649, EPI_ISL_8612662, EPI_ISL_8612704, EPI_ISL_8612726, EPI_ISL_8612730, EPI_ISL_8612744, EPI_ISL_8612754, EPI_ISL_8612776, EPI_ISL_8612778, EPI_ISL_8612795, EPI_ISL_8612806, EPI_ISL_8613830, EPI_ISL_8613832, EPI_ISL_8613872, EPI_ISL_8613888, EPI_ISL_8613902, EPI_ISL_8613930, EPI_ISL_8613938, EPI_ISL_8613939, EPI_ISL_8613959, EPI_ISL_8613969, EPI_ISL_8613979, EPI_ISL_8613987, EPI_ISL_8614023, EPI_ISL_8614027, EPI_ISL_8614034, EPI_ISL_8614042, EPI_ISL_8614054, EPI_ISL_8614071, EPI_ISL_8614088, EPI_ISL_8614106, EPI_ISL_8614126, EPI_ISL_8614131, EPI_ISL_8614150, EPI_ISL_8614183, EPI_ISL_8614234, EPI_ISL_8614245, EPI_ISL_8614288, EPI_ISL_8614339, EPI_ISL_8614365, EPI_ISL_8614369, EPI_ISL_8614409, EPI_ISL_8614444, EPI_ISL_8614448, EPI_ISL_8614452, EPI_ISL_8614472, EPI_ISL_8614495, EPI_ISL_8614501, EPI_ISL_8614508, EPI_ISL_8614518, EPI_ISL_8614537, EPI_ISL_8614545, EPI_ISL_8614560, EPI_ISL_8614590, EPI_ISL_8614615, EPI_ISL_8614619, EPI_ISL_8614634, EPI_ISL_8614658, EPI_ISL_8614664, EPI_ISL_8614668, EPI_ISL_8614710, EPI_ISL_8614749, EPI_ISL_8614753, EPI_ISL_8614763, EPI_ISL_8614780, EPI_ISL_8614788, EPI_ISL_8614791, EPI_ISL_8614808, EPI_ISL_8614812, EPI_ISL_8614885 | | | |
| see above | Department of Bacteria, Parasites and Fungi, Statens Serum Institut, Copenhagen, Denmark | Statens Serum Institut Bioinformatics and Microbial Genomics | Danish Covid-19 Genome Consortium |
| EPI_ISL_8623424 | Department of Clinical Microbiology | GIGA Medical Genomics | Bouchra Boujemla; Claire Gourzonès; Cécile Meex; Keith Durkin; Laurent Gillet; Maria Artesi; Marie-Pierre Hayette; Nadine Cambisano; Nathalie Renotte; Olivier Ek; Sébastien Bontems; Vincent Bours |
| EPI_ISL_8607570, EPI_ISL_8607573, EPI_ISL_8607605, EPI_ISL_8607606, EPI_ISL_8607607, EPI_ISL_8607608, EPI_ISL_8607609, EPI_ISL_8608254, EPI_ISL_8608268, EPI_ISL_8625485, EPI_ISL_8625586, EPI_ISL_8629990, EPI_ISL_8631251 | | | |
| see above | Fulgent Genetics | Centers for Disease Control and Prevention Division of Viral Diseases, Pathogen Discovery | Becky Tsai; Benafsh Sapra; Benjamin Rambo-Martin; Christopher Gulvick; Clinton Paden; Dakota Howard; Dhwani Batra; Doreen Ng; Duncan MacCannell; Erisa Sula; Harry Gao; James Xie; Jason Caravas; John Gao; Joseph Fierro; Kristine Lacek; Matthew Schmerer; Mickey Li; Peter Cook; Scott Sammons; Shatavia Morrison; Tymeckia Kendall; Victoria Caban Figueroa; Yan Meng; Yvette Unoarumhi |
| EPI_ISL_8605571 | GENOME CENTER, Jashore University of Science and Technology | GENOME CENTER, Jashore University of Science and Technology | Hassan M. Al-Emran; Iqbal Kabir Jahid; Md. Ali Ahasan Setu; Md. Anwar Hossain; Md. Rasel Parvez; Md. Shaminur Rahman Prosanto Kumar Das; Md. Shazid Hasan; Shovon Lal Sarkar; Toukir Ahammed |
| EPI_ISL_8605841 | GENOME CENTER, Jashore University of Science and Technology. | GENOME CENTER, Jashore University of Science and Technology. | Hassan M. Al-Emran; Iqbal Kabir Jahid; Md. Ali Ahasan Setu; Md. Anwar Hossain; Md. Rasel Parvez; Md. Shaminur Rahman Prosanto Kumar Das; Md. Shazid Hasan; Shovon Lal Sarkar; Toukir Ahammed |
| EPI_ISL_8619211, EPI_ISL_8619216, EPI_ISL_8619222, EPI_ISL_8619224, EPI_ISL_8619225, EPI_ISL_8619233, EPI_ISL_8619234 | | | |
| see above | Hannover Medical School, Institute of Virology | Hannover Medical School, Institute of Virology | Lars Steinbrück |
| EPI_ISL_8598733 | Hospital Carmen Y S. Ochoa | Laboratorio de Virología HUCA | ; Alba L; Alvarez-Arguelles ME; Boga JA; Costales I; Coto E; González-Alba JM; Gómez de Oña J; Martín-Rodríguez G; Melón S; Perez-Martínez Z; Rojo S; Sandoval M |
| EPI_ISL_8598725, EPI_ISL_8598726, EPI_ISL_8598728, EPI_ISL_8598729, EPI_ISL_8598730, EPI_ISL_8598731, EPI_ISL_8598732, EPI_ISL_8598735, EPI_ISL_8598739, EPI_ISL_8598743 | | | |
| see above | Hospital Universitario Central de Asturias | Laboratorio de Virología HUCA | ; Alba L; Alvarez-Arguelles ME; Boga JA; Costales I; Coto E; González-Alba JM; Gómez de Oña J; Martín-Rodríguez G; Melón S; Perez-Martínez Z; Rojo S; Sandoval M |
| EPI_ISL_8629108, EPI_ISL_8629109, EPI_ISL_8629110, EPI_ISL_8629111, EPI_ISL_8629112, EPI_ISL_8629113, EPI_ISL_8629114, EPI_ISL_8629115, EPI_ISL_8629116, EPI_ISL_8629117, EPI_ISL_8629118, EPI_ISL_8629119, EPI_ISL_8629120, EPI_ISL_8629121, EPI_ISL_8629123, EPI_ISL_8629124, EPI_ISL_8629125, EPI_ISL_8629126, EPI_ISL_8629127, EPI_ISL_8629128, EPI_ISL_8629129, EPI_ISL_8629130, EPI_ISL_8629131, EPI_ISL_8629132, EPI_ISL_8629133, EPI_ISL_8629134, EPI_ISL_8629135, EPI_ISL_8629136, EPI_ISL_8629137, EPI_ISL_8629139, EPI_ISL_8629140, EPI_ISL_8629141, EPI_ISL_8629142, EPI_ISL_8629143, EPI_ISL_8629144, EPI_ISL_8629145, EPI_ISL_8629146, EPI_ISL_8629147, EPI_ISL_8629148, EPI_ISL_8629149, EPI_ISL_8629152, EPI_ISL_8629153, EPI_ISL_8629154, EPI_ISL_8629155, EPI_ISL_8629156, EPI_ISL_8629157, EPI_ISL_8629158, EPI_ISL_8629159, EPI_ISL_8629160, EPI_ISL_8629161, EPI_ISL_8629162, EPI_ISL_8629163, EPI_ISL_8629164, EPI_ISL_8629165, EPI_ISL_8629166, EPI_ISL_8629167, EPI_ISL_8629168, EPI_ISL_8629169, EPI_ISL_8629170, EPI_ISL_8629171, EPI_ISL_8629172, EPI_ISL_8629173, EPI_ISL_8629228, EPI_ISL_8629229, EPI_ISL_8629230, EPI_ISL_8629232, EPI_ISL_8629233, EPI_ISL_8629234, EPI_ISL_8629236, EPI_ISL_8629237, EPI_ISL_8629238, EPI_ISL_8629248, EPI_ISL_8629249, EPI_ISL_8629250, EPI_ISL_8629251, EPI_ISL_8629254, EPI_ISL_8629256, EPI_ISL_8629260, EPI_ISL_8629261, EPI_ISL_8629262, EPI_ISL_8629263, EPI_ISL_8629264, EPI_ISL_8629265, EPI_ISL_8629266, EPI_ISL_8629267, EPI_ISL_8629268, EPI_ISL_8629269, EPI_ISL_8629270, EPI_ISL_8629272, EPI_ISL_8629273 | | | |
| see above | Idaho Bureau of Laboratories | Idaho Bureau of Laboratories | "R. Beukelman; Aimee Ceniseros; Christian Loera; Christopher Ball"; Matthew Charles Burns; Robert L. Voermans |
| EPI_ISL_8601617, EPI_ISL_8602634, EPI_ISL_8603144, EPI_ISL_8603202, EPI_ISL_8603211 | Laboratory Corporation of America | Centers for Disease Control and Prevention Division of Viral Diseases, Pathogen Discovery | Amanda Douglas; Amanda Suchanek; Andrea Throop; Ayla Burns; Benjamin Rambo-Martin; Bobbi Croy; Brian Krueger; Brian Norvell; Christopher Gulvick; Christos Petropoulos; Clinton Paden; Craig Lukasik; Dakota Howard; Debbie Boles; Dhwani Batra; Duncan MacCannell; Eyad Almasri; Goran Stevovic; Howard Engler; Hrushikesh Deshmukh; Jake Humphrey; Jana Schroth; Jason Caravas; Joe Voshell; John Pruitt; Jonathan Meltzer; Jonathan Williams; Kimberly Wagner; Kristine Lacek; Lax Iyer; Lisa Pfefferle; Lyndon Tilson; Manoj Jain; Marcia Eisenberg; Mary Cristobal; Mary Williamson; Matthew Robinson; Matthew Schmerer; Michael Levandoski; Mike Sapeta; Mindy Nye; Minoo Agarwal; Mohan Kolli; Nuthawin Charoensri; Oren Cohen; Peter Cook; Prashant Gupta; Qian Zeng; Rama Ghatti; Scott Parker; Scott Ryan; Scott Sammons; Shatavia Morrison; Stanley Letovsky; Steven Ragan; Suresh Selvaraju; Susan Countryman; Susan Hicks; Suzanne Dale; Thomas Urban; Tim Kuphal; Tricia Zwiefelhofer; Tymeckia Kendall; Victoria Caban Figueroa; Vincent Drouillon; Yvette Unoarumhi |
| EPI_ISL_8623347 | National Platform bis COVID ULB-IBC | National Platform bis COVID ULB-IBC | Arnaud Marchant; Benoit Haerlingen; Coralie Henin; Marie-Luce Delforge; Ricardo De Mendonça |
| EPI_ISL_8597860 | New South Wales Health Pathology Royal Prince Alfred Hospital | Microbiology RPAH | Au, J.; Bull, R.; Deveson, I.; Foster, C.; Rawlinson, W.; Ruiz Silva, M.; Van Hal, S. |
| EPI_ISL_8598240, EPI_ISL_8598245, EPI_ISL_8598248, EPI_ISL_8598252, EPI_ISL_8598254, EPI_ISL_8598265, EPI_ISL_8598266, EPI_ISL_8598275, EPI_ISL_8598278, EPI_ISL_8598279, EPI_ISL_8598281, EPI_ISL_8598284, EPI_ISL_8598288, EPI_ISL_8598289, EPI_ISL_8598305, EPI_ISL_8598309, EPI_ISL_8598310, EPI_ISL_8598312, EPI_ISL_8598331, EPI_ISL_8598332, EPI_ISL_8598338, EPI_ISL_8598353, EPI_ISL_8598360, EPI_ISL_8598361, EPI_ISL_8598363, EPI_ISL_8598364, EPI_ISL_8598365, EPI_ISL_8598366, EPI_ISL_8598374, EPI_ISL_8598379, EPI_ISL_8598383, EPI_ISL_8598388, EPI_ISL_8598391, EPI_ISL_8598394, EPI_ISL_8598396, EPI_ISL_8598397, EPI_ISL_8598405 | | | |
| see above | PHV-FSS | PHV-FSS | Chenwei Wang on behalf of Q-PHIRE Genomics |
| EPI_ISL_8607795 | PO AVEZZANO - U.O.C. Laboratorio Analisi | Istituto Zooprofilattico Sperimentale dell'Abruzzo e Molise "G. Caporale" | Ancora M; Calistri P; Cammà C; Caporale M; Curini V; Delli Compagni E; Di Domenico M; Di Lollo Valeria; Di Pasquale A; Lorusso A; Mangone I; Marcacci M; Puglia I; Rinaldi A; Savini G; Scialabba S |
| EPI_ISL_8603748 | Pejabat Kesihatan KLIA | iPROMISE, UiTM | Ariza Adnan; Fadzilah Mohd Nor; Lim Wai Feng; Mohd Asif Mohd Sukri; Mohd Nur Fakhruzzaman Noorizhab; Mohd Zaki Salleh; Sazzli Shahlan Kassim; Siti Farah Alwani Mohd Nawi; Siti Hamimah Sheikh Abdul Kadir; Teh Lay Kek; Wang Seok Mui |
| EPI_ISL_8625876, EPI_ISL_8625877, EPI_ISL_8625878, EPI_ISL_8625879, EPI_ISL_8625881 | Regional Hospital Liberec | Regional Hospital Liberec | Iva Dolinova; Katerina Arientova; Katerina Stillerova; Martin Kracik; Tomas Zajic |
| EPI_ISL_8616009 | SK-Roy Romanow Provincial Laboratory | Saskatchewan - Roy Romanow Provincial Laboratory (RRPL) | Alanna Senecal; Amanda Lang; Jessica Minion; Kara Loos; Keith MacKenzie; Meredith Faires; Rachel DePaulo; Roy Romanow Provincial Laboratory - Molecular Diagnostics; Ryan McDonald |
| EPI_ISL_8605618, EPI_ISL_8621950, EPI_ISL_8622036 | UW Virology Lab | UW Virology Lab | Alexander Greninger; Hong Xie; Isabel Arnould; Keith R Jerome; Meei-Li Huang; Nathan Breit; Patrick Mathias; Pavitra Roychoudhury; Pooneh Hajian; Ricardo Perez; Robert J. Livingston; Sean Ellis; Seffir T. Wendm; Shah Mohamed Bakhash |
| EPI_ISL_8620724 | Utah Public Health Laboratory | Utah Public Health Laboratory | Erin L. Young; John Arnn; Kelly F. Oakeson; Olinto Linares-Perdomo; Pooja Gupta; Tom Iverson |
| EPI_ISL_8636462, EPI_ISL_8636482 | ZARV/NHLS, Department Medical Virology, University of Pretoria | CERI, Centre for Epidemic Response and Innvoation, Stellenbosch University and KRISP, KZN Research Innovation and Sequencing Platform, UKZN. | Adriano Mendes; Amy Strydom; Giandhari J; Micheala Davids; Moir M; Naidoo Y; Pillay S; Ramphal U; Ramphal Y; San JE; Sim Mayaphi and Marietjie Venter; Tegally H; Tshiabuila D; Van Wyk S; Wilkinson E; de Oliveira T |

We gratefully acknowledge the following Authors from the Originating laboratories responsible for obtaining the specimens, as well as the Submitting laboratories where the genome data were generated and shared via GISAID, on which this research is based.

All Submitters of data may be contacted directly via www.gisaid.org

Authors are sorted alphabetically.

Acknowledgement EPI_SET Identifier: EPI_SET_20220527ga

| Accession ID | Originating Laboratory | Submitting Laboratory | Authors |
|---|---|---|---|
| EPI_ISL_8637800, EPI_ISL_8637814, EPI_ISL_8637817 | AP SSO | CSIR-Centre for Cellular and Molecular Biology-INSACOG | Amareshwar Vodapalli; Ara Sreenivas; Archana Bharadwaj Siva; B Himasri; Divya Tej Sowpati; Jandhyala Sai Krishna; Karthik Bharadwaj Tallapaka; Lamuk Zaveri; Malini Nemalikanti; Priya Nurkurthy; Rakesh K Mishra; Shreekant Verma; Sreelekshmi MS; Sumedha Avadhanula; Surabhi Srivastava; Tulasi Nagabandi; Valli Nagalakshmi Undamatla; Vidhyadhari Methuku |
| EPI_ISL_8637640 | CSIR-National Environmental Engineering Research Institute | CSIR-Centre for Cellular and Molecular Biology-INSACOG | Amareshwar Vodapalli; Ara Sreenivas; Archana Bharadwaj Siva; B Himasri; Divya Tej Sowpati; Karthik Bharadwaj Tallapaka; Krishna Khairnar; Lamuk Zaveri; Malini Nemalikanti; Priya Nurkurthy; Rakesh K Mishra; Shreekant Verma; Sreelekshmi MS; Sumedha Avadhanula; Surabhi Srivastava; Tulasi Nagabandi; Valli Nagalakshmi Undamatla; Vidhyadhari Methuku |
| EPI_ISL_8653120, EPI_ISL_8653178, EPI_ISL_8653181, EPI_ISL_8653190, EPI_ISL_8653193, EPI_ISL_8653196, EPI_ISL_8653208, EPI_ISL_8653211, EPI_ISL_8653213, EPI_ISL_8653254, EPI_ISL_8653257, EPI_ISL_8653284, EPI_ISL_8653288, EPI_ISL_8653311, EPI_ISL_8653328, EPI_ISL_8653331, EPI_ISL_8653345, EPI_ISL_8653352, EPI_ISL_8653363, EPI_ISL_8653367, EPI_ISL_8653382, EPI_ISL_8653384, EPI_ISL_8653399, EPI_ISL_8653409, EPI_ISL_8653437, EPI_ISL_8653478, EPI_ISL_8653499, EPI_ISL_8653508, EPI_ISL_8653519, EPI_ISL_8653522, EPI_ISL_8653525, EPI_ISL_8653543, EPI_ISL_8653555, EPI_ISL_8653591, EPI_ISL_8653600, EPI_ISL_8653622, EPI_ISL_8653630, EPI_ISL_8653633, EPI_ISL_8653642, EPI_ISL_8653645, EPI_ISL_8653653, EPI_ISL_8653661, EPI_ISL_8653674, EPI_ISL_8653696, EPI_ISL_8653699, EPI_ISL_8653701, EPI_ISL_8653711, EPI_ISL_8653717, EPI_ISL_8653723, EPI_ISL_8653737, EPI_ISL_8653743, EPI_ISL_8653760, EPI_ISL_8653763, EPI_ISL_8653792, EPI_ISL_8653808, EPI_ISL_8653814, EPI_ISL_8653834, EPI_ISL_8653843, EPI_ISL_8653846, EPI_ISL_8653849, EPI_ISL_8653852, EPI_ISL_8653858, EPI_ISL_8653881, EPI_ISL_8653899, EPI_ISL_8653905, EPI_ISL_8653908, EPI_ISL_8653942, EPI_ISL_8653961, EPI_ISL_8653980, EPI_ISL_8653983, EPI_ISL_8653986, EPI_ISL_8653991, EPI_ISL_8653996, EPI_ISL_8654012, EPI_ISL_8654027, EPI_ISL_8654029, EPI_ISL_8654047, EPI_ISL_8654052, EPI_ISL_8654055, EPI_ISL_8654063, EPI_ISL_8654091, EPI_ISL_8654093, EPI_ISL_8654100, EPI_ISL_8654109, EPI_ISL_8654120, EPI_ISL_8654134, EPI_ISL_8654149, EPI_ISL_8654152, EPI_ISL_8654153, EPI_ISL_8654185, EPI_ISL_8654191, EPI_ISL_8654198, EPI_ISL_8654206, EPI_ISL_8654221, EPI_ISL_8654230, EPI_ISL_8654233, EPI_ISL_8654246, EPI_ISL_8654261, EPI_ISL_8654272, EPI_ISL_8654281, EPI_ISL_8654295, EPI_ISL_8654335, EPI_ISL_8654340, EPI_ISL_8654352, EPI_ISL_8654358, EPI_ISL_8654361, EPI_ISL_8654379, EPI_ISL_8654395, EPI_ISL_8654404, EPI_ISL_8654407, EPI_ISL_8654410, EPI_ISL_8654432, EPI_ISL_8654460, EPI_ISL_8654466, EPI_ISL_8654472, EPI_ISL_8654474, EPI_ISL_8654478, EPI_ISL_8654493, EPI_ISL_8654498, EPI_ISL_8654544, EPI_ISL_8654562, EPI_ISL_8654565, EPI_ISL_8654574, EPI_ISL_8654584, EPI_ISL_8654599, EPI_ISL_8654609, EPI_ISL_8654630, EPI_ISL_8654643, EPI_ISL_8654680, EPI_ISL_8654713, EPI_ISL_8654725, EPI_ISL_8654733, EPI_ISL_8654750, EPI_ISL_8654764, EPI_ISL_8654767, EPI_ISL_8654773, EPI_ISL_8654795, EPI_ISL_8654802, EPI_ISL_8654804, EPI_ISL_8654807, EPI_ISL_8654813, EPI_ISL_8654814, EPI_ISL_8654815, EPI_ISL_8654819, EPI_ISL_8654821, EPI_ISL_8654832, EPI_ISL_8654851, EPI_ISL_8654859, EPI_ISL_8654873, EPI_ISL_8654882, EPI_ISL_8654897, EPI_ISL_8654907, EPI_ISL_8654914, EPI_ISL_8654956, EPI_ISL_8654994, EPI_ISL_8655008, EPI_ISL_8655013, EPI_ISL_8655031, EPI_ISL_8655033, EPI_ISL_8655060, EPI_ISL_8655069, EPI_ISL_8655077, EPI_ISL_8655083, EPI_ISL_8655098, EPI_ISL_8655109, EPI_ISL_8655117, EPI_ISL_8655173, EPI_ISL_8655189, EPI_ISL_8655192, EPI_ISL_8655204, EPI_ISL_8655217, EPI_ISL_8655260, EPI_ISL_8655266, EPI_ISL_8655275, EPI_ISL_8655283, EPI_ISL_8655295, EPI_ISL_8655305, EPI_ISL_8655307, EPI_ISL_8655312, EPI_ISL_8655324, EPI_ISL_8655385, EPI_ISL_8655391, EPI_ISL_8655403, EPI_ISL_8655418, EPI_ISL_8655421, EPI_ISL_8655447, EPI_ISL_8655455, EPI_ISL_8655470, EPI_ISL_8655482, EPI_ISL_8655508, EPI_ISL_8655529, EPI_ISL_8655547, EPI_ISL_8655562, EPI_ISL_8655574, EPI_ISL_8655581, EPI_ISL_8655593, EPI_ISL_8655595, EPI_ISL_8655604, EPI_ISL_8655605, EPI_ISL_8655611, EPI_ISL_8655617, EPI_ISL_8655622, EPI_ISL_8655623, EPI_ISL_8655628, EPI_ISL_8655635, EPI_ISL_8655641, EPI_ISL_8655643, EPI_ISL_8655646, EPI_ISL_8655661, EPI_ISL_8655664, EPI_ISL_8655667, EPI_ISL_8655732, EPI_ISL_8655735, EPI_ISL_8655752, EPI_ISL_8655770, EPI_ISL_8655773, EPI_ISL_8655794, EPI_ISL_8655803, EPI_ISL_8655806, EPI_ISL_8655818, EPI_ISL_8655836, EPI_ISL_8655851, EPI_ISL_8655871, EPI_ISL_8655876, EPI_ISL_8655901, EPI_ISL_8655909, EPI_ISL_8655921, EPI_ISL_8655932, EPI_ISL_8655944, EPI_ISL_8655956, EPI_ISL_8655962, EPI_ISL_8655966, EPI_ISL_8655974, EPI_ISL_8656009, EPI_ISL_8656012, EPI_ISL_8656017, EPI_ISL_8656027, EPI_ISL_8656033, EPI_ISL_8656046, EPI_ISL_8656074, EPI_ISL_8656094, EPI_ISL_8656103, EPI_ISL_8656131, EPI_ISL_8656156, EPI_ISL_8656163, EPI_ISL_8656166, EPI_ISL_8656172, EPI_ISL_8656184, EPI_ISL_8656188, EPI_ISL_8656198, EPI_ISL_8656201, EPI_ISL_8656217, EPI_ISL_8656218, EPI_ISL_8656232, EPI_ISL_8656240, EPI_ISL_8656248, EPI_ISL_8656289, EPI_ISL_8656293, EPI_ISL_8656308, EPI_ISL_8656328, EPI_ISL_8656336, EPI_ISL_8656345, EPI_ISL_8656351, EPI_ISL_8656354, EPI_ISL_8656365, EPI_ISL_8656372, EPI_ISL_8656400, EPI_ISL_8656406, EPI_ISL_8656407, EPI_ISL_8656592, EPI_ISL_8656598, EPI_ISL_8656602, EPI_ISL_8656609, EPI_ISL_8656613, EPI_ISL_8656616, EPI_ISL_8656621, EPI_ISL_8656627, EPI_ISL_8656630, EPI_ISL_8656632, EPI_ISL_8656637, EPI_ISL_8656654, EPI_ISL_8656679, EPI_ISL_8656682, EPI_ISL_8656685, EPI_ISL_8656693, EPI_ISL_8656708, EPI_ISL_8656738, EPI_ISL_8656767, EPI_ISL_8656777, EPI_ISL_8656812, EPI_ISL_8656849, EPI_ISL_8656852, EPI_ISL_8656885, EPI_ISL_8656894, EPI_ISL_8656900, EPI_ISL_8656904, EPI_ISL_8656937, EPI_ISL_8656963, EPI_ISL_8656965, EPI_ISL_8656969, EPI_ISL_8656972, EPI_ISL_8656978, EPI_ISL_8657001, EPI_ISL_8657042, EPI_ISL_8657068, EPI_ISL_8657070, EPI_ISL_8657082, EPI_ISL_8657110, EPI_ISL_8657142, EPI_ISL_8657148, EPI_ISL_8657154, EPI_ISL_8657156, EPI_ISL_8657167, EPI_ISL_8657199, EPI_ISL_8657208, EPI_ISL_8657226, EPI_ISL_8657232, EPI_ISL_8657239, EPI_ISL_8657241, EPI_ISL_8657244, EPI_ISL_8657247, EPI_ISL_8657269, EPI_ISL_8657280, EPI_ISL_8657287, EPI_ISL_8657307, EPI_ISL_8657315, EPI_ISL_8657339, EPI_ISL_8657367, EPI_ISL_8657378, EPI_ISL_8657392, EPI_ISL_8657394, EPI_ISL_8657399, EPI_ISL_8657400, EPI_ISL_8657401, EPI_ISL_8657402, EPI_ISL_8657587, EPI_ISL_8657594, EPI_ISL_8657608, EPI_ISL_8657623, EPI_ISL_8657640, EPI_ISL_8657648, EPI_ISL_8657654, EPI_ISL_8657660, EPI_ISL_8657699, EPI_ISL_8657710, EPI_ISL_8657716, EPI_ISL_8657728, EPI_ISL_8657736, EPI_ISL_8657746, EPI_ISL_8657774, EPI_ISL_8657780, EPI_ISL_8657797, EPI_ISL_8657805, EPI_ISL_8657811, EPI_ISL_8657817, EPI_ISL_8657820, EPI_ISL_8657826, EPI_ISL_8657831, EPI_ISL_8657838, EPI_ISL_8657852, EPI_ISL_8657866, EPI_ISL_8657903, EPI_ISL_8657909, EPI_ISL_8657915, EPI_ISL_8657923, EPI_ISL_8657925, EPI_ISL_8657928, EPI_ISL_8657942, EPI_ISL_8657951, EPI_ISL_8657958, EPI_ISL_8657963, EPI_ISL_8657973, EPI_ISL_8657978, EPI_ISL_8657987, EPI_ISL_8657993, EPI_ISL_8657999, EPI_ISL_8658021, EPI_ISL_8658024, EPI_ISL_8658028, EPI_ISL_8658033, EPI_ISL_8658057, EPI_ISL_8658090, EPI_ISL_8658093, EPI_ISL_8658102, EPI_ISL_8658108, EPI_ISL_8658125, EPI_ISL_8658144, EPI_ISL_8658148, EPI_ISL_8658154, EPI_ISL_8658159, EPI_ISL_8658167, EPI_ISL_8658173, EPI_ISL_8658200, EPI_ISL_8658205, EPI_ISL_8658227, EPI_ISL_8658267, EPI_ISL_8658272, EPI_ISL_8658283, EPI_ISL_8658299, EPI_ISL_8658313, EPI_ISL_8658316, EPI_ISL_8658323, EPI_ISL_8658326, EPI_ISL_8658354, EPI_ISL_8658363, EPI_ISL_8658644, EPI_ISL_8658647, EPI_ISL_8658649, EPI_ISL_8658652, EPI_ISL_8658660, EPI_ISL_8658666, EPI_ISL_8658668, EPI_ISL_8658671, EPI_ISL_8658681, EPI_ISL_8658696, EPI_ISL_8658707, EPI_ISL_8658719, EPI_ISL_8658737, EPI_ISL_8658755, EPI_ISL_8658766, EPI_ISL_8658769, EPI_ISL_8658816, EPI_ISL_8658830, EPI_ISL_8658833, EPI_ISL_8658850, EPI_ISL_8658853, EPI_ISL_8658866, EPI_ISL_8658880, EPI_ISL_8658889, EPI_ISL_8658895, EPI_ISL_8658897, EPI_ISL_8658906, EPI_ISL_8658911, EPI_ISL_8658917, EPI_ISL_8658926, EPI_ISL_8658941, EPI_ISL_8658947, EPI_ISL_8658961, EPI_ISL_8658974, EPI_ISL_8658980, EPI_ISL_8658983, EPI_ISL_8658991, EPI_ISL_8659004, EPI_ISL_8659009, EPI_ISL_8659014, EPI_ISL_8659041, EPI_ISL_8659063, EPI_ISL_8659081, EPI_ISL_8659085, EPI_ISL_8659088, EPI_ISL_8659093, EPI_ISL_8659096, EPI_ISL_8659102, EPI_ISL_8659107, EPI_ISL_8659126 | | | |
| see above | Department of Bacteria, Parasites and Fungi, Statens Serum Institut, Copenhagen, Denmark | Statens Serum Institut Bioinformatics and Microbial Genomics | Danish Covid-19 Genome Consortium |
| EPI_ISL_8648923 | Dept. of Microbiology and Infection Control, Akershus University Hospital HF | Dept. of Microbiology and Infection Control, Akershus University Hospital HF | Alexander Hesselberg Løvestad; Hanne Berggreen; Hege Vangstein Aamot |
| EPI_ISL_8648853, EPI_ISL_8648854 | HOSPITAL DE CABUEÑES | Laboratorio de Virología HUCA | ; Alba L; Alvarez-Arguelles ME; Boga JA; Costales I; Coto E; González-Alba JM; Gómez de Oña J; Martín-Rodríguez G; Melón S; Perez-Martínez Z; Rojo S; Sandoval M |
| EPI_ISL_8648851, EPI_ISL_8648859 | HOSPITAL UNIVERSITARIO CENTRAL DE ASTURIAS | Laboratorio de Virología HUCA | ; Alba L; Alvarez-Arguelles ME; Boga JA; Costales I; Coto E; González-Alba JM; Gómez de Oña J; Martín-Rodríguez G; Melón S; Perez-Martínez Z; Rojo S; Sandoval M |
| EPI_ISL_8668943 | Klinikum Stuttgart | Robert Koch Institute | |
| EPI_ISL_8651228 | Lighthouse Lab in Alderley Park | Wellcome Sanger Institute for the COVID-19 Genomics UK (COG-UK) Consortium | Cordelia Langford; David K. Jackson; Dominic Kwiatkowski; Ewan Harrison; Ian Johnston; Jacquelyn Wynn; Jeffrey Barrett; John Sillitoe on behalf of the Wellcome Sanger Institute COVID-19 Surveillance Team; Mairead Hyland; Roberto Amato; Sonia Goncalves; The Lighthouse Lab in Alderley Park and Alex Alderton |
| EPI_ISL_8651229 | Lighthouse Lab in Milton Keynes | Wellcome Sanger Institute for the COVID-19 Genomics UK (COG-UK) Consortium | Cordelia Langford; David K. Jackson; Dominic Kwiatkowski; Ewan Harrison; Ian Johnston; Jeffrey Barrett; John Sillitoe on behalf of the Wellcome Sanger Institute COVID-19 Surveillance Team; Roberto Amato; Sonia Goncalves; The Lighthouse Lab in Milton Keynes and Alex Alderton |
| EPI_ISL_8637669, EPI_ISL_8637811, EPI_ISL_8637838 | MapMyGenome | CSIR-Centre for Cellular and Molecular Biology-INSACOG | Amareshwar Vodapalli; Anuradha Acharya; Ara Sreenivas; Archana Bharadwaj Siva; B Himasri; Divya Tej Sowpati; Jandhyala Sai Krishna; Karthik Bharadwaj Tallapaka; Lamuk Zaveri; M. Aravind Kumar; Malini Nemalikanti; Priya Nurkurthy; Rakesh K Mishra; S.P.S. Satish; Sandhya Kiran; Shreekant Verma; Sreelekshmi MS; Sumedha Avadhanula; Surabhi Srivastava; Tulasi Nagabandi; Valli Nagalakshmi Undamatla; Vidhyadhari Methuku |
| EPI_ISL_8650134 | Normah Medical Specialist Centre, Kuching | Institute of Health and Community Medicine | Chan Chia Jui; Chua Hock Hin; David Perera; Ikwannuddin Y; Ooi Mong How; Tonnii Sia Loong Loong; Wong Jyn Shan |
| EPI_ISL_8650409, EPI_ISL_8650410, EPI_ISL_8650411, EPI_ISL_8650412, EPI_ISL_8650414 | SARS-CoV-2 Sequencing Castilla y Leon-Spain Consortium | SARS-CoV-2 Sequencing Castilla y Leon-Spain Consortium | Antonio Orduña-Domingo; Carlos Fuster Foz; Carmen Aldea-Mansilla; Carmen Gimeno Crespo; David Abad; Gabriel March Rosello; Gregoria Megías Lobón; Jose María Eiros Bouza; M. Isabel Fernandez-Natal; Marta Dominguez-Gil; Marta Hernandez; María Antonia García Castro; Mª Fe Brezmes-Valdivieso; Noelia Arenal Andrés; Silvia Rojo; Sonsoles Garcinuño Pérez |
| EPI_ISL_8650133, EPI_ISL_8650135, EPI_ISL_8650136, EPI_ISL_8650137 | Sarawak General Hospital (Kuching) | Institute of Health and Community Medicine | Chan Chia Jui; Chua Hock Hin; David Perera; Ooi Mong How; Tonnii Sia Loong Loong; Wong Jyn Shan |

We gratefully acknowledge the following Authors from the Originating laboratories responsible for obtaining the specimens, as well as the Submitting laboratories where the genome data were generated and shared via GISAID, on which this research is based.

All Submitters of data may be contacted directly via www.gisaid.org

Authors are sorted alphabetically.

Acknowledgement EPI_SET Identifier: EPI_SET_20220527nm

| Accession ID | Originating Laboratory | Submitting Laboratory | Authors |
|---|---|---|---|
| EPI_ISL_8685691, EPI_ISL_8685692, EPI_ISL_8685693, EPI_ISL_8685697, EPI_ISL_8686761, EPI_ISL_8686819 | Aegis Sciences Corporation | Centers for Disease Control and Prevention Division of Viral Diseases, Pathogen Discovery | Alec Vest; Benjamin Rambo-Martin; Christopher Gulvick; Clinton Paden; Cyndi Clark; Dakota Howard; Dhwani Batra; Dillon Nall; Duncan MacCannell; Erisa Sula; Ethan Sanders; Holly Houdeshell; Jason Caravas; Kristine Lacek; Matthew Hardison; Matthew Schmerer; Ola Kvalvaag; Patrick Campbell; Peter Cook; Rob Case; Scott Sammons; Shatavia Morrison; Shaun Westlund; Tymeckia Kendall; Victoria Caban Figueroa; Vikramsinha Ghorpade; Yvette Unoarumhi |
| EPI_ISL_8685512, EPI_ISL_8685513 | LADR Zentrallabor DR. Kramer & Kollegen Geesthacht | Robert Koch Institute | |
| EPI_ISL_8687362, EPI_ISL_8687363, EPI_ISL_8687364, EPI_ISL_8687365, EPI_ISL_8687366, EPI_ISL_8687367, EPI_ISL_8687370, EPI_ISL_8687372, EPI_ISL_8687373, EPI_ISL_8687374, EPI_ISL_8687375, EPI_ISL_8687376, EPI_ISL_8687378, EPI_ISL_8687379, EPI_ISL_8687380, EPI_ISL_8687381, EPI_ISL_8687383, EPI_ISL_8687384, EPI_ISL_8687385, EPI_ISL_8687386, EPI_ISL_8687387, EPI_ISL_8687388, EPI_ISL_8687389, EPI_ISL_8687390, EPI_ISL_8687392, EPI_ISL_8687393, EPI_ISL_8687394, EPI_ISL_8687395, EPI_ISL_8687396, EPI_ISL_8687397, EPI_ISL_8687398, EPI_ISL_8687400, EPI_ISL_8687403, EPI_ISL_8687404, EPI_ISL_8687408, EPI_ISL_8687409, EPI_ISL_8687410, EPI_ISL_8687412, EPI_ISL_8687416, EPI_ISL_8687418, EPI_ISL_8687419, EPI_ISL_8687420 | | | |
| see above | Ochsner Health | BioInfoExperts | Amy Feehan; Ben Lain; Chris Huston; David J. Nolan; Judy Crabtree; Julia-Garcia-Diaz; Lucio Miele; Rebecca Rose; Samual Moot; Susanna L. Lamers; Tessa LaFleur |

We gratefully acknowledge the following Authors from the Originating laboratories responsible for obtaining the specimens, as well as the Submitting laboratories where the genome data were generated and shared via GISAID, on which this research is based.

All Submitters of data may be contacted directly via www.gisaid.org

Authors are sorted alphabetically.

| Accession ID | Originating Laboratory | Submitting Laboratory | Authors |
|---|---|---|---|
| EPI_ISL_8684305, EPI_ISL_8684311 | Invenimus AG | Institute of Medical Virology, University of Zurich | Alexandra Trkola; Annette Audigé; Catharine Aquino; Cyril Shah; Daniel Ehrsam; Gabriela Ziltener; Guido Bloemberg; Hubert Rehrauer; Isabel Stürmer; Joel Wirz; Jon Huder; Jürg Böni; Kevin Steiner; Maria Grünberg; Maryam Zaheri; Michael Huber; Riccarda Capaul; Stefan Schmutz; Verena Kufner; Weihong Qi |
| EPI_ISL_8684213, EPI_ISL_8684225, EPI_ISL_8684231, EPI_ISL_8684238, EPI_ISL_8684240, EPI_ISL_8684243, EPI_ISL_8684244, EPI_ISL_8684245 | | | |
| see above | Kantonsspital Aarau | Institute of Medical Virology, University of Zurich | Alexandra Trkola; Annette Audigé; Catharine Aquino; Cyril Shah; Daniel Ehrsam; Gabriela Ziltener; Guido Bloemberg; Hubert Rehrauer; Isabel Stürmer; Joel Wirz; Jon Huder; Jürg Böni; Kevin Steiner; Maria Grünberg; Maryam Zaheri; Michael Huber; Riccarda Capaul; Stefan Schmutz; Verena Kufner; Weihong Qi |
| EPI_ISL_8684370, EPI_ISL_8684386, EPI_ISL_8684387, EPI_ISL_8684392, EPI_ISL_8684398 | Kantonsspital Winterthur | Institute of Medical Virology, University of Zurich | Alexandra Trkola; Annette Audigé; Catharine Aquino; Cyril Shah; Daniel Ehrsam; Gabriela Ziltener; Guido Bloemberg; Hubert Rehrauer; Isabel Stürmer; Joel Wirz; Jon Huder; Jürg Böni; Kevin Steiner; Maria Grünberg; Maryam Zaheri; Michael Huber; Riccarda Capaul; Stefan Schmutz; Verena Kufner; Weihong Qi |
| EPI_ISL_8685507, EPI_ISL_8685509 | LADR Zentrallabor DR. Kramer & Kollegen Geesthacht | Robert Koch Institute | |
| EPI_ISL_8684246 | Stadtspital Waid | Institute of Medical Virology, University of Zurich | Alexandra Trkola; Annette Audigé; Catharine Aquino; Cyril Shah; Daniel Ehrsam; Gabriela Ziltener; Guido Bloemberg; Hubert Rehrauer; Isabel Stürmer; Joel Wirz; Jon Huder; Jürg Böni; Kevin Steiner; Maria Grünberg; Maryam Zaheri; Michael Huber; Riccarda Capaul; Stefan Schmutz; Verena Kufner; Weihong Qi |
| EPI_ISL_8684362, EPI_ISL_8684363, EPI_ISL_8685314, EPI_ISL_8685323, EPI_ISL_8685330 | Universität Zürich | Institute of Medical Virology, University of Zurich | Alexandra Trkola; Annette Audigé; Catharine Aquino; Cyril Shah; Daniel Ehrsam; Gabriela Ziltener; Guido Bloemberg; Hubert Rehrauer; Isabel Stürmer; Joel Wirz; Jon Huder; Jürg Böni; Kevin Steiner; Maria Grünberg; Maryam Zaheri; Michael Huber; Riccarda Capaul; Stefan Schmutz; Verena Kufner; Weihong Qi |
| EPI_ISL_8684260, EPI_ISL_8684328, EPI_ISL_8684340, EPI_ISL_8684346, EPI_ISL_8684364, EPI_ISL_8684433, EPI_ISL_8685227, EPI_ISL_8685245 | | | |
| see above | Universitäts-Kinderspital Zürich | Institute of Medical Virology, University of Zurich | Alexandra Trkola; Annette Audigé; Catharine Aquino; Cyril Shah; Daniel Ehrsam; Gabriela Ziltener; Guido Bloemberg; Hubert Rehrauer; Isabel Stürmer; Joel Wirz; Jon Huder; Jürg Böni; Kevin Steiner; Maria Grünberg; Maryam Zaheri; Michael Huber; Riccarda Capaul; Stefan Schmutz; Verena Kufner; Weihong Qi |
| EPI_ISL_8684264, EPI_ISL_8684270, EPI_ISL_8684275, EPI_ISL_8684278, EPI_ISL_8684281, EPI_ISL_8684287, EPI_ISL_8684293, EPI_ISL_8684421, EPI_ISL_8684425, EPI_ISL_8684428, EPI_ISL_8685251, EPI_ISL_8685257, EPI_ISL_8685263, EPI_ISL_8685375, EPI_ISL_8685376, EPI_ISL_8685377, EPI_ISL_8685378, EPI_ISL_8685379 | | | |
| see above | UniversitätsSpital Zürich | Institute of Medical Virology, University of Zurich | Alexandra Trkola; Annette Audigé; Catharine Aquino; Cyril Shah; Daniel Ehrsam; Gabriela Ziltener; Guido Bloemberg; Hubert Rehrauer; Isabel Stürmer; Joel Wirz; Jon Huder; Jürg Böni; Kevin Steiner; Maria Grünberg; Maryam Zaheri; Michael Huber; Riccarda Capaul; Stefan Schmutz; Verena Kufner; Weihong Qi |
| EPI_ISL_8684429 | Zentrallabor Zürich | Institute of Medical Virology, University of Zurich | Alexandra Trkola; Annette Audigé; Catharine Aquino; Cyril Shah; Daniel Ehrsam; Gabriela Ziltener; Guido Bloemberg; Hubert Rehrauer; Isabel Stürmer; Joel Wirz; Jon Huder; Jürg Böni; Kevin Steiner; Maria Grünberg; Maryam Zaheri; Michael Huber; Riccarda Capaul; Stefan Schmutz; Verena Kufner; Weihong Qi |

We gratefully acknowledge the following Authors from the Originating laboratories responsible for obtaining the specimens, as well as the Submitting laboratories where the genome data were generated and shared via GISAID, on which this research is based.

All Submitters of data may be contacted directly via www.gisaid.org

Authors are sorted alphabetically.

| Accession ID | Originating Laboratory | Submitting Laboratory | Authors |
|---|---|---|---|
| EPI_ISL_8684196 | Invenimus AG | Institute of Medical Virology, University of Zurich | Alexandra Trkola; Annette Audigé; Catharine Aquino; Cyril Shah; Daniel Ehrsam; Gabriela Ziltener; Guido Bloemberg; Hubert Rehrauer; Isabel Stürmer; Joel Wirz; Jon Huder; Jürg Böni; Kevin Steiner; Maria Grünberg; Maryam Zaheri; Michael Huber; Riccarda Capaul; Stefan Schmutz; Verena Kufner; Weihong Qi |
| EPI_ISL_8682545, EPI_ISL_8682555 | Kantonsspital Baden AG | Institute of Medical Virology, University of Zurich | Alexandra Trkola; Annette Audigé; Catharine Aquino; Cyril Shah; Daniel Ehrsam; Gabriela Ziltener; Guido Bloemberg; Hubert Rehrauer; Isabel Stürmer; Joel Wirz; Jon Huder; Jürg Böni; Kevin Steiner; Maria Grünberg; Maryam Zaheri; Michael Huber; Riccarda Capaul; Stefan Schmutz; Verena Kufner; Weihong Qi |
| EPI_ISL_8682198, EPI_ISL_8682443, EPI_ISL_8682473, EPI_ISL_8682477, EPI_ISL_8682482, EPI_ISL_8682483, EPI_ISL_8682487, EPI_ISL_8682495, EPI_ISL_8682519, EPI_ISL_8682524 | | | |
| see above | Kantonsspital Winterthur | Institute of Medical Virology, University of Zurich | Alexandra Trkola; Annette Audigé; Catharine Aquino; Cyril Shah; Daniel Ehrsam; Gabriela Ziltener; Guido Bloemberg; Hubert Rehrauer; Isabel Stürmer; Joel Wirz; Jon Huder; Jürg Böni; Kevin Steiner; Maria Grünberg; Maryam Zaheri; Michael Huber; Riccarda Capaul; Stefan Schmutz; Verena Kufner; Weihong Qi |
| EPI_ISL_8682669, EPI_ISL_8683655 | Stadtspital Waid | Institute of Medical Virology, University of Zurich | Alexandra Trkola; Annette Audigé; Catharine Aquino; Cyril Shah; Daniel Ehrsam; Gabriela Ziltener; Guido Bloemberg; Hubert Rehrauer; Isabel Stürmer; Joel Wirz; Jon Huder; Jürg Böni; Kevin Steiner; Maria Grünberg; Maryam Zaheri; Michael Huber; Riccarda Capaul; Stefan Schmutz; Verena Kufner; Weihong Qi |
| EPI_ISL_8682563, EPI_ISL_8682573, EPI_ISL_8683696, EPI_ISL_8683701 | Universität Zürich | Institute of Medical Virology, University of Zurich | Alexandra Trkola; Annette Audigé; Catharine Aquino; Cyril Shah; Daniel Ehrsam; Gabriela Ziltener; Guido Bloemberg; Hubert Rehrauer; Isabel Stürmer; Joel Wirz; Jon Huder; Jürg Böni; Kevin Steiner; Maria Grünberg; Maryam Zaheri; Michael Huber; Riccarda Capaul; Stefan Schmutz; Verena Kufner; Weihong Qi |
| EPI_ISL_8682327, EPI_ISL_8682373, EPI_ISL_8682379, EPI_ISL_8682387, EPI_ISL_8682396, EPI_ISL_8682403, EPI_ISL_8682420, EPI_ISL_8682429, EPI_ISL_8682534, EPI_ISL_8682612, EPI_ISL_8682619, EPI_ISL_8682643, EPI_ISL_8682667, EPI_ISL_8682697, EPI_ISL_8684122, EPI_ISL_8684133 | | | |
| see above | Universitäts-Kinderspital Zürich | Institute of Medical Virology, University of Zurich | Alexandra Trkola; Annette Audigé; Catharine Aquino; Cyril Shah; Daniel Ehrsam; Gabriela Ziltener; Guido Bloemberg; Hubert Rehrauer; Isabel Stürmer; Joel Wirz; Jon Huder; Jürg Böni; Kevin Steiner; Maria Grünberg; Maryam Zaheri; Michael Huber; Riccarda Capaul; Stefan Schmutz; Verena Kufner; Weihong Qi |
| EPI_ISL_8682216, EPI_ISL_8682262, EPI_ISL_8682276, EPI_ISL_8682285, EPI_ISL_8682294, EPI_ISL_8682303, EPI_ISL_8682311, EPI_ISL_8682319, EPI_ISL_8682324, EPI_ISL_8682723, EPI_ISL_8682729, EPI_ISL_8683712, EPI_ISL_8684155 | | | |
| see above | UniversitätsSpital Zürich | Institute of Medical Virology, University of Zurich | Alexandra Trkola; Annette Audigé; Catharine Aquino; Cyril Shah; Daniel Ehrsam; Gabriela Ziltener; Guido Bloemberg; Hubert Rehrauer; Isabel Stürmer; Joel Wirz; Jon Huder; Jürg Böni; Kevin Steiner; Maria Grünberg; Maryam Zaheri; Michael Huber; Riccarda Capaul; Stefan Schmutz; Verena Kufner; Weihong Qi |
| EPI_ISL_8683806 | Valais Hospital, Central Institute | Valais Hospital, Central Institute | Alexis Dumoulin; Cedric Howald; Deborah Penet; Henri Pegeot; Ioannis Xenarios; Keith Harshman; Lorenzo Cerutti; Melyssa Elies |
| EPI_ISL_8684184 | Zentrallabor Zürich | Institute of Medical Virology, University of Zurich | Alexandra Trkola; Annette Audigé; Catharine Aquino; Cyril Shah; Daniel Ehrsam; Gabriela Ziltener; Guido Bloemberg; Hubert Rehrauer; Isabel Stürmer; Joel Wirz; Jon Huder; Jürg Böni; Kevin Steiner; Maria Grünberg; Maryam Zaheri; Michael Huber; Riccarda Capaul; Stefan Schmutz; Verena Kufner; Weihong Qi |

We gratefully acknowledge the following Authors from the Originating laboratories responsible for obtaining the specimens, as well as the Submitting laboratories where the genome data were generated and shared via GISAID, on which this research is based.

All Submitters of data may be contacted directly via www.gisaid.org

Authors are sorted alphabetically.

Acknowledgement EPI_SET Identifier: EPI_SET_20220527uq

| Accession ID | Originating Laboratory | Submitting Laboratory | Authors |
|---|---|---|---|
| EPI_ISL_8672912 | Infinity Biologix | Centers for Disease Control and Prevention Division of Viral Diseases, Pathogen Discovery | Benjamin Rambo-Martin; Chirayu Goswami; Christian Bixby; Christopher Gulvick; Clinton Paden; Dakota Howard; Dhwani Batra; Duncan MacCannell; Erisa Sula; Jason Caravas; Jonathan Schultz; Kristine Lacek; Matthew Schmerer; Peter Cook; Robin Grimwood; Russ Hager; Scott Sammons; Shatavia Morrison; Tymeckia Kendall; Victoria Caban Figueroa; Yihe Wang; Yvette Unoarumhi |
| EPI_ISL_8668943, EPI_ISL_8668945, EPI_ISL_8668946, EPI_ISL_8668947, EPI_ISL_8668948, EPI_ISL_8668950, EPI_ISL_8668951, EPI_ISL_8668953, EPI_ISL_8668954, EPI_ISL_8668955, EPI_ISL_8668958, EPI_ISL_8668959, EPI_ISL_8668960, EPI_ISL_8668961, EPI_ISL_8668963, EPI_ISL_8668964, EPI_ISL_8668965, EPI_ISL_8668966, EPI_ISL_8668967, EPI_ISL_8668968, EPI_ISL_8668969, EPI_ISL_8668970 | | | |
| see above | Klinikum Stuttgart | Robert Koch Institute | |
| EPI_ISL_8670505, EPI_ISL_8671355, EPI_ISL_8672018, EPI_ISL_8672560 | LABOPAT | LABOPAT | Cynthia Penaloza; Luis Mendoza; Silvia Montilla |
| EPI_ISL_8680564 | Laborarztpraxis Dres. med. Walther Weindel & Kollegen | Robert Koch Institute | |
| EPI_ISL_8674176, EPI_ISL_8674177, EPI_ISL_8674178 | MVZ Labor Krone GbR | Robert Koch Institute | |
| EPI_ISL_8679094 | Sonic - Bioscientia - MVZ Labor Saar GmbH | Robert Koch Institute | |
| EPI_ISL_8681459, EPI_ISL_8681559 | Stadtspital Waid | Institute of Medical Virology, University of Zurich | Alexandra Trkola; Annette Audigé; Catharine Aquino; Cyril Shah; Daniel Ehrsam; Gabriela Ziltener; Guido Bloemberg; Hubert Rehrauer; Isabel Stürmer; Joel Wirz; Jon Huder; Jürg Böni; Kevin Steiner; Maria Grünberg; Maryam Zaheri; Michael Huber; Riccarda Capaul; Stefan Schmutz; Verena Kufner; Weihong Qi |
| EPI_ISL_8681454, EPI_ISL_8681466, EPI_ISL_8681480, EPI_ISL_8681505, EPI_ISL_8681553 | Universität Zürich | Institute of Medical Virology, University of Zurich | Alexandra Trkola; Annette Audigé; Catharine Aquino; Cyril Shah; Daniel Ehrsam; Gabriela Ziltener; Guido Bloemberg; Hubert Rehrauer; Isabel Stürmer; Joel Wirz; Jon Huder; Jürg Böni; Kevin Steiner; Maria Grünberg; Maryam Zaheri; Michael Huber; Riccarda Capaul; Stefan Schmutz; Verena Kufner; Weihong Qi |
| EPI_ISL_8681448, EPI_ISL_8681566, EPI_ISL_8681575, EPI_ISL_8681580 | Universitäts-Kinderspital Zürich | Institute of Medical Virology, University of Zurich | Alexandra Trkola; Annette Audigé; Catharine Aquino; Cyril Shah; Daniel Ehrsam; Gabriela Ziltener; Guido Bloemberg; Hubert Rehrauer; Isabel Stürmer; Joel Wirz; Jon Huder; Jürg Böni; Kevin Steiner; Maria Grünberg; Maryam Zaheri; Michael Huber; Riccarda Capaul; Stefan Schmutz; Verena Kufner; Weihong Qi |
| EPI_ISL_8681497, EPI_ISL_8681519, EPI_ISL_8681526, EPI_ISL_8681538, EPI_ISL_8681621, EPI_ISL_8682183, EPI_ISL_8682189 | | | |
| see above | UniversitätsSpital Zürich | Institute of Medical Virology, University of Zurich | Alexandra Trkola; Annette Audigé; Catharine Aquino; Cyril Shah; Daniel Ehrsam; Gabriela Ziltener; Guido Bloemberg; Hubert Rehrauer; Isabel Stürmer; Joel Wirz; Jon Huder; Jürg Böni; Kevin Steiner; Maria Grünberg; Maryam Zaheri; Michael Huber; Riccarda Capaul; Stefan Schmutz; Verena Kufner; Weihong Qi |

We gratefully acknowledge the following Authors from the Originating laboratories responsible for obtaining the specimens, as well as the Submitting laboratories where the genome data were generated and shared via GISAID, on which this research is based.

All Submitters of data may be contacted directly via www.gisaid.org

Authors are sorted alphabetically.

| Accession ID | Originating Laboratory | Submitting Laboratory | Authors |
|---|---|---|---|
| EPI_ISL_8660509 | Clinique Saint-Pierre Ottignies | UCLouvain/IREC/MBLG-CTMA | Benoit Kabamba Mukadi; Bertrand Bearzatto; Jean-Luc Gala; Nicolas Pinte; Paul Blanpain; Simon Ophélie; Valentin Coste |
| EPI_ISL_8660507, EPI_ISL_8660508 | Cliniques universitaires Saint-Luc | UCLouvain/IREC/MBLG-CTMA | Benoit Kabamba Mukadi; Bertrand Bearzatto; Jean-Luc Gala; Nicolas Pinte; Paul Blanpain; Simon Ophélie; Valentin Coste |
| EPI_ISL_8659865, EPI_ISL_8659893, EPI_ISL_8659930, EPI_ISL_8659936, EPI_ISL_8659955, EPI_ISL_8660005, EPI_ISL_8660010, EPI_ISL_8660023, EPI_ISL_8660055, EPI_ISL_8660084, EPI_ISL_8660102, EPI_ISL_8660142, EPI_ISL_8660145, EPI_ISL_8660162, EPI_ISL_8660171, EPI_ISL_8660190, EPI_ISL_8660193, EPI_ISL_8660199, EPI_ISL_8660203, EPI_ISL_8660205, EPI_ISL_8660206, EPI_ISL_8660210, EPI_ISL_8660213, EPI_ISL_8660226, EPI_ISL_8660231, EPI_ISL_8660247, EPI_ISL_8660253, EPI_ISL_8660279, EPI_ISL_8660286 | | | |
| see above | Department of Bacteria, Parasites and Fungi, Statens Serum Institut, Copenhagen, Denmark | Statens Serum Institut Bioinformatics and Microbial Genomics | Danish Covid-19 Genome Consortium |
| EPI_ISL_8668942 | Klinikum Stuttgart | Robert Koch Institute | |
| EPI_ISL_8665271, EPI_ISL_8665273, EPI_ISL_8665274, EPI_ISL_8665275, EPI_ISL_8665277, EPI_ISL_8665278, EPI_ISL_8665280, EPI_ISL_8665281 | | | |
| see above | Laboratorio de Pesquisa em Virologia, FAMERP, SJRP | Laboratorio de Pesquisa em Virologia, FAMERP, SJRP | Beatriz de Carvalho Marques; Cecília Artico Banho; Cíntia Bittar; Fábio Sossai Possebon; Guilherme Campos; Helena Lage Ferreira; Jorge A. Petroli Marchesi; João Pessoa Araújo Jr.; Leila Sabrina Ullmann; Lívia Sacchetto; Maisa C. Pereira Parra; Marília Moraes; Maurício L. Nogueira.; Paula Rahal; Paulo Inacio da Costa |
| EPI_ISL_8665230, EPI_ISL_8665242, EPI_ISL_8665300, EPI_ISL_8665319, EPI_ISL_8665344, EPI_ISL_8665617, EPI_ISL_8665682 | | | |
| see above | Medizinisch-Diagnostisches Labor Kempten allgäulab | Robert Koch Institute | |
| EPI_ISL_8668093 | SYNLAB Jena Oncoscreen | Robert Koch Institute | |
| EPI_ISL_8662043 | SYNLAB MVZ Hamburg | Robert Koch Institute | |

We gratefully acknowledge the following Authors from the Originating laboratories responsible for obtaining the specimens, as well as the Submitting laboratories where the genome data were generated and shared via GISAID, on which this research is based.

All Submitters of data may be contacted directly via www.gisaid.org

Authors are sorted alphabetically.

EPI_ISL_8659132, EPI_ISL_8659142, EPI_ISL_8659151, EPI_ISL_8659154, EPI_ISL_8659163, EPI_ISL_8659166, EPI_ISL_8659195, EPI_ISL_8659203, EPI_ISL_8659221, EPI_ISL_8659223, EPI_ISL_8659239, EPI_ISL_8659247, EPI_ISL_8659253, EPI_ISL_8659285, EPI_ISL_8659363, EPI_ISL_8659382, EPI_ISL_8659385, EPI_ISL_8659393, EPI_ISL_8659399, EPI_ISL_8659422, EPI_ISL_8659425, EPI_ISL_8659430, EPI_ISL_8659432, EPI_ISL_8659442, EPI_ISL_8659446, EPI_ISL_8659457, EPI_ISL_8659489, EPI_ISL_8659491, EPI_ISL_8659497, EPI_ISL_8659558, EPI_ISL_8659563, EPI_ISL_8659565, EPI_ISL_8659576, EPI_ISL_8659583, EPI_ISL_8659632, EPI_ISL_8659688, EPI_ISL_8659689, EPI_ISL_8659693, EPI_ISL_8659710, EPI_ISL_8659722, EPI_ISL_8659734, EPI_ISL_8659737, EPI_ISL_8659762, EPI_ISL_8659778, EPI_ISL_8659793, EPI_ISL_8659796, EPI_ISL_8659802, EPI_ISL_8659813, EPI_ISL_8659822, EPI_ISL_8659841

| Accession ID | Originating Laboratory | Submitting Laboratory | Authors |
|---|---|---|---|
| see above | Department of Bacteria, Parasites and Fungi, Statens Serum Institut, Copenhagen, Denmark | Statens Serum Institut Bioinformatics and Microbial Genomics | Danish Covid-19 Genome Consortium |

We gratefully acknowledge the following Authors from the Originating laboratories responsible for obtaining the specimens, as well as the Submitting laboratories where the genome data were generated and shared via GISAID, on which this research is based.

All Submitters of data may be contacted directly via www.gisaid.org

Authors are sorted alphabetically.

Acknowledgement EPI_SET Identifier: EPI_SET_20220527tx

| Accession ID | Originating Laboratory | Submitting Laboratory | Authors |
|---|---|---|---|
| EPI_ISL_8694511 | Center for Laboratory Medicine | Center for Laboratory Medicine, ZLMSG | Yannick Gerth |
| EPI_ISL_8700219, EPI_ISL_8700241, EPI_ISL_8700285, EPI_ISL_8700291, EPI_ISL_8700294, EPI_ISL_8700297, EPI_ISL_8700300, EPI_ISL_8700302, EPI_ISL_8700303, EPI_ISL_8700320, EPI_ISL_8700326, EPI_ISL_8700329, EPI_ISL_8700344, EPI_ISL_8700348, EPI_ISL_8700355, EPI_ISL_8700362, EPI_ISL_8700380, EPI_ISL_8700384, EPI_ISL_8700395, EPI_ISL_8700413, EPI_ISL_8700417, EPI_ISL_8700420, EPI_ISL_8700441, EPI_ISL_8700450, EPI_ISL_8700454, EPI_ISL_8700456, EPI_ISL_8700457, EPI_ISL_8700462, EPI_ISL_8700466, EPI_ISL_8700468, EPI_ISL_8700482, EPI_ISL_8700507, EPI_ISL_8700569, EPI_ISL_8700573, EPI_ISL_8700580, EPI_ISL_8701342, EPI_ISL_8701370, EPI_ISL_8701408, EPI_ISL_8701421, EPI_ISL_8701435, EPI_ISL_8701468, EPI_ISL_8701478, EPI_ISL_8701488, EPI_ISL_8701509, EPI_ISL_8701532, EPI_ISL_8701535, EPI_ISL_8701552, EPI_ISL_8701555, EPI_ISL_8701571, EPI_ISL_8701591, EPI_ISL_8701593, EPI_ISL_8701596, EPI_ISL_8701601, EPI_ISL_8701606, EPI_ISL_8701709, EPI_ISL_8701731, EPI_ISL_8701748, EPI_ISL_8701755, EPI_ISL_8701776, EPI_ISL_8701787, EPI_ISL_8701793, EPI_ISL_8701796, EPI_ISL_8701801, EPI_ISL_8701807, EPI_ISL_8701818, EPI_ISL_8701845, EPI_ISL_8701849, EPI_ISL_8701856, EPI_ISL_8701868, EPI_ISL_8701870, EPI_ISL_8701898, EPI_ISL_8701902, EPI_ISL_8701912, EPI_ISL_8701918, EPI_ISL_8701921, EPI_ISL_8701941, EPI_ISL_8701971, EPI_ISL_8701979, EPI_ISL_8701988, EPI_ISL_8702019, EPI_ISL_8702027, EPI_ISL_8702044, EPI_ISL_8702059, EPI_ISL_8702084, EPI_ISL_8702107, EPI_ISL_8702119, EPI_ISL_8702122, EPI_ISL_8702157, EPI_ISL_8702164, EPI_ISL_8702197, EPI_ISL_8702200, EPI_ISL_8702219, EPI_ISL_8702222, EPI_ISL_8702231, EPI_ISL_8702241, EPI_ISL_8702251, EPI_ISL_8702257, EPI_ISL_8702260, EPI_ISL_8702267, EPI_ISL_8702303, EPI_ISL_8702327, EPI_ISL_8702335, EPI_ISL_8702363, EPI_ISL_8702372, EPI_ISL_8702375, EPI_ISL_8702383, EPI_ISL_8702385, EPI_ISL_8702389, EPI_ISL_8702394, EPI_ISL_8702403, EPI_ISL_8702425, EPI_ISL_8702428, EPI_ISL_8702430, EPI_ISL_8702432, EPI_ISL_8702445, EPI_ISL_8702447, EPI_ISL_8702464, EPI_ISL_8702470, EPI_ISL_8702473, EPI_ISL_8702495, EPI_ISL_8702504, EPI_ISL_8702519, EPI_ISL_8702527, EPI_ISL_8702528, EPI_ISL_8702549, EPI_ISL_8702559, EPI_ISL_8702573, EPI_ISL_8702579, EPI_ISL_8702582, EPI_ISL_8702593, EPI_ISL_8702596, EPI_ISL_8702605, EPI_ISL_8702608, EPI_ISL_8702617, EPI_ISL_8702620, EPI_ISL_8702631, EPI_ISL_8702642, EPI_ISL_8702651, EPI_ISL_8702671, EPI_ISL_8702691, EPI_ISL_8702697, EPI_ISL_8702703, EPI_ISL_8702710, EPI_ISL_8702720, EPI_ISL_8702722, EPI_ISL_8702735, EPI_ISL_8702737, EPI_ISL_8702747, EPI_ISL_8702767, EPI_ISL_8702854, EPI_ISL_8702857, EPI_ISL_8702859, EPI_ISL_8702870, EPI_ISL_8702873, EPI_ISL_8702886, EPI_ISL_8702908, EPI_ISL_8702963, EPI_ISL_8702996, EPI_ISL_8703001, EPI_ISL_8703012, EPI_ISL_8703021, EPI_ISL_8703048, EPI_ISL_8703059, EPI_ISL_8703140, EPI_ISL_8703144, EPI_ISL_8703161, EPI_ISL_8703164, EPI_ISL_8703168, EPI_ISL_8703172, EPI_ISL_8703207, EPI_ISL_8703231, EPI_ISL_8703241, EPI_ISL_8703257, EPI_ISL_8703263, EPI_ISL_8703269, EPI_ISL_8703276, EPI_ISL_8703294, EPI_ISL_8703310, EPI_ISL_8703314, EPI_ISL_8703329, EPI_ISL_8703340, EPI_ISL_8703354, EPI_ISL_8703367, EPI_ISL_8703376, EPI_ISL_8703380, EPI_ISL_8703381, EPI_ISL_8703386, EPI_ISL_8703390, EPI_ISL_8703394, EPI_ISL_8703437, EPI_ISL_8703444, EPI_ISL_8703449, EPI_ISL_8703465, EPI_ISL_8703473, EPI_ISL_8703495, EPI_ISL_8703522, EPI_ISL_8703524, EPI_ISL_8703535, EPI_ISL_8703542, EPI_ISL_8703561, EPI_ISL_8703596, EPI_ISL_8703612, EPI_ISL_8703624, EPI_ISL_8703645, EPI_ISL_8703656, EPI_ISL_8703672, EPI_ISL_8703801, EPI_ISL_8703827, EPI_ISL_8704004, EPI_ISL_8704007, EPI_ISL_8704014, EPI_ISL_8704018, EPI_ISL_8704046, EPI_ISL_8704072, EPI_ISL_8704122, EPI_ISL_8704126, EPI_ISL_8704142, EPI_ISL_8704146, EPI_ISL_8704148, EPI_ISL_8704155, EPI_ISL_8704159, EPI_ISL_8704175, EPI_ISL_8704190, EPI_ISL_8704227, EPI_ISL_8704243, EPI_ISL_8704245, EPI_ISL_8704273, EPI_ISL_8704277, EPI_ISL_8704291, EPI_ISL_8704295, EPI_ISL_8704333, EPI_ISL_8704356, EPI_ISL_8704368, EPI_ISL_8704416, EPI_ISL_8704425, EPI_ISL_8704429, EPI_ISL_8704437, EPI_ISL_8704449, EPI_ISL_8704474, EPI_ISL_8704487, EPI_ISL_8704497, EPI_ISL_8704500, EPI_ISL_8704505, EPI_ISL_8704519, EPI_ISL_8704526, EPI_ISL_8704550, EPI_ISL_8704553, EPI_ISL_8704568, EPI_ISL_8704606, EPI_ISL_8704625, EPI_ISL_8704633, EPI_ISL_8704639, EPI_ISL_8704642, EPI_ISL_8704656, EPI_ISL_8704664, EPI_ISL_8704668, EPI_ISL_8704673, EPI_ISL_8704676, EPI_ISL_8704708, EPI_ISL_8704730, EPI_ISL_8704775, EPI_ISL_8704788, EPI_ISL_8704792, EPI_ISL_8704804, EPI_ISL_8704807, EPI_ISL_8704821, EPI_ISL_8704825, EPI_ISL_8704845, EPI_ISL_8704883, EPI_ISL_8704903, EPI_ISL_8704913, EPI_ISL_8704917, EPI_ISL_8704924, EPI_ISL_8704938, EPI_ISL_8704959, EPI_ISL_8705001, EPI_ISL_8705023, EPI_ISL_8705046, EPI_ISL_8705051, EPI_ISL_8705055, EPI_ISL_8705058, EPI_ISL_8705062, EPI_ISL_8705079, EPI_ISL_8705127, EPI_ISL_8705131, EPI_ISL_8705150, EPI_ISL_8705175, EPI_ISL_8705180, EPI_ISL_8705191, EPI_ISL_8705198, EPI_ISL_8705236, EPI_ISL_8705238, EPI_ISL_8705241, EPI_ISL_8705243, EPI_ISL_8705250, EPI_ISL_8705252, EPI_ISL_8705253, EPI_ISL_8705257, EPI_ISL_8705258, EPI_ISL_8705281, EPI_ISL_8705283, EPI_ISL_8705307, EPI_ISL_8705325, EPI_ISL_8705344, EPI_ISL_8705348, EPI_ISL_8705355, EPI_ISL_8705378, EPI_ISL_8705386, EPI_ISL_8705396, EPI_ISL_8705424, EPI_ISL_8705427, EPI_ISL_8705441, EPI_ISL_8705463, EPI_ISL_8705469, EPI_ISL_8705477, EPI_ISL_8705488, EPI_ISL_8705500, EPI_ISL_8705503, EPI_ISL_8705507, EPI_ISL_8705511, EPI_ISL_8705521, EPI_ISL_8705571, EPI_ISL_8705581, EPI_ISL_8705594, EPI_ISL_8705596, EPI_ISL_8705680, EPI_ISL_8705697, EPI_ISL_8705704, EPI_ISL_8705708, EPI_ISL_8705712, EPI_ISL_8705719, EPI_ISL_8705722, EPI_ISL_8705733, EPI_ISL_8705746, EPI_ISL_8705758, EPI_ISL_8705761, EPI_ISL_8705765, EPI_ISL_8705769, EPI_ISL_8705783, EPI_ISL_8705792, EPI_ISL_8705797, EPI_ISL_8705800, EPI_ISL_8705809, EPI_ISL_8705829, EPI_ISL_8705831, EPI_ISL_8705842, EPI_ISL_8705876, EPI_ISL_8705892, EPI_ISL_8705896, EPI_ISL_8705914, EPI_ISL_8705918, EPI_ISL_8705922, EPI_ISL_8705946, EPI_ISL_8705962, EPI_ISL_8705967, EPI_ISL_8705969, EPI_ISL_8705973, EPI_ISL_8705979, EPI_ISL_8706004, EPI_ISL_8706012, EPI_ISL_8706023, EPI_ISL_8706037, EPI_ISL_8706043, EPI_ISL_8706053, EPI_ISL_8706072, EPI_ISL_8706076, EPI_ISL_8706097, EPI_ISL_8706100, EPI_ISL_8706104, EPI_ISL_8706108, EPI_ISL_8706135, EPI_ISL_8706139, EPI_ISL_8706147, EPI_ISL_8706181, EPI_ISL_8706188, EPI_ISL_8706192 | | | |
| see above | Department of Bacteria, Parasites and Fungi, Statens Serum Institut, Copenhagen, Denmark | Statens Serum Institut Bioinformatics and Microbial Genomics | Danish Covid-19 Genome Consortium |
| EPI_ISL_8700415 | Karolinska University Hospital Huddinge | Karolinska University Hospital | Annika Tiveljung Lindell; Henning Onsbring; Jan Albert; Karina Hentrich; Lynda Eneh; Maria Ropat; Martin Ekman; Natalija Gerasimcik; Robert Dyrdak; Sandra Broddesson; Shambhu Ganeshappa Aralaguppe; Tanja Normark; Tobias Allander; Valtteri Wirta; Zhibing Yun |
| EPI_ISL_8695191, EPI_ISL_8695335, EPI_ISL_8695913, EPI_ISL_8695958, EPI_ISL_8695967, EPI_ISL_8697860, EPI_ISL_8699738, EPI_ISL_8699837, EPI_ISL_8701499, EPI_ISL_8701569, EPI_ISL_8701595 | | | |
| see above | Lighthouse Lab in Alderley Park | Wellcome Sanger Institute for the COVID-19 Genomics UK (COG-UK) Consortium | Cordelia Langford; David K. Jackson; Dominic Kwiatkowski; Ewan Harrison; Ian Johnston; Jacquelyn Wynn; Jeffrey Barrett; John Sillitoe on behalf of the Wellcome Sanger Institute COVID-19 Surveillance Team; Mairead Hyland; Roberto Amato; Sonia Goncalves; The Lighthouse Lab in Alderley Park and Alex Alderton |
| EPI_ISL_8695142, EPI_ISL_8695678, EPI_ISL_8695844, EPI_ISL_8695873, EPI_ISL_8695927, EPI_ISL_8696349, EPI_ISL_8697885, EPI_ISL_8697903, EPI_ISL_8697930, EPI_ISL_8699721, EPI_ISL_8703315, EPI_ISL_8704326, EPI_ISL_8704422, EPI_ISL_8704863, EPI_ISL_8705845, EPI_ISL_8706197 | | | |
| see above | Lighthouse Lab in Glasgow | Wellcome Sanger Institute for the COVID-19 Genomics UK (COG-UK) Consortium | Anna Dominiczak and Alex Alderton; Carol Clugston; Cordelia Langford; David Gray; David K. Jackson; Dominic Kwiatkowski; Ewan Harrison; Harper VanSteenhouse; Ian Johnston; Jeffrey Barrett; John Sillitoe on behalf of the Wellcome Sanger Institute COVID-19 Surveillance Team; Roberto Amato; Sonia Goncalves; Yumi Kasai |
| EPI_ISL_8695718, EPI_ISL_8695740, EPI_ISL_8695877, EPI_ISL_8695910, EPI_ISL_8696735, EPI_ISL_8696857, EPI_ISL_8697593, EPI_ISL_8697689, EPI_ISL_8697756, EPI_ISL_8698826, EPI_ISL_8699265, EPI_ISL_8699445, EPI_ISL_8701597, EPI_ISL_8704563 | | | |
| see above | Lighthouse Lab in Milton Keynes | Wellcome Sanger Institute for the COVID-19 Genomics UK (COG-UK) Consortium | Cordelia Langford; David K. Jackson; Dominic Kwiatkowski; Ewan Harrison; Ian Johnston; Jeffrey Barrett; John Sillitoe on behalf of the Wellcome Sanger Institute COVID-19 Surveillance Team; Roberto Amato; Sonia Goncalves; The Lighthouse Lab in Milton Keynes and Alex Alderton |
| EPI_ISL_8703863, EPI_ISL_8703864, EPI_ISL_8703865, EPI_ISL_8703866, EPI_ISL_8703867, EPI_ISL_8703868, EPI_ISL_8703869, EPI_ISL_8703870, EPI_ISL_8703871, EPI_ISL_8703872, EPI_ISL_8703873, EPI_ISL_8703874, EPI_ISL_8703875, EPI_ISL_8703876, EPI_ISL_8703877, EPI_ISL_8703878, EPI_ISL_8703879, EPI_ISL_8703880, EPI_ISL_8703881, EPI_ISL_8703882, EPI_ISL_8703883, EPI_ISL_8703884, EPI_ISL_8703885, EPI_ISL_8703886, EPI_ISL_8703887, EPI_ISL_8703888, EPI_ISL_8703889, EPI_ISL_8703890, EPI_ISL_8703891, EPI_ISL_8703892, EPI_ISL_8703893, EPI_ISL_8703895, EPI_ISL_8703896, EPI_ISL_8703897, EPI_ISL_8703898, EPI_ISL_8703899, EPI_ISL_8703900, EPI_ISL_8703901, EPI_ISL_8703902, EPI_ISL_8703903, EPI_ISL_8703904 | | | |
| see above | National Public Health Laboratory, National Centre for Infectious Diseases | National Public Health Laboratory, National Centre for Infectious Diseases | Benny Yeo; Grace Ngan; Lin Cui; Raymond Tzer Pin Lin; Royce Ang; Samuel Loo; Zhenyang Zhou |
| EPI_ISL_8696665 | Rosalind Franklin Laboratory | Wellcome Sanger Institute for the COVID-19 Genomics UK (COG-UK) Consortium | Cordelia Langford; David K. Jackson; Dominic Kwiatkowski; Donald Fraser; Ewan Harrison; Ian Johnston; Jeffrey Barrett; John Sillitoe on behalf of the Wellcome Sanger Institute COVID-19 Surveillance Team; Rob Howes; Roberto Amato; Sonia Goncalves; Suki Lee; The Rosalind Franklin Laboratory and Alex Alderton |
| EPI_ISL_8691409 | State Laboratories Division, Hawaii State Department of Health | State Laboratories Division, Hawaii State Department of Health | Ayana Garnet; Briana Ofilas; Cheryl-lynn Daquip; Cheyenne Barela; Daniel Strange; Drew Kuwazaki; Edward Desmond; Jeffrey Au; Mark Nagata; Pamela O'Brien; Razvan Sultana; Remedios Gose; Samantha Cotter; Samantha Sruba |
| EPI_ISL_8699489, EPI_ISL_8699490, EPI_ISL_8699491, EPI_ISL_8699492, EPI_ISL_8699493, EPI_ISL_8699494, EPI_ISL_8699495, EPI_ISL_8699496, EPI_ISL_8699497, EPI_ISL_8699500, EPI_ISL_8699501, EPI_ISL_8699502, EPI_ISL_8699503, EPI_ISL_8699504, EPI_ISL_8699505, EPI_ISL_8699506, EPI_ISL_8699507, EPI_ISL_8699508 | | | |
| see above | Synlab Eesti OÜ | 1. Laboratory of Communicable Diseases (Estonia); 2. Eurofins Genomics Europe Sequencing GmbH | Abroi A.; Avi R.; Dotsenko L.; Epštein J.; Hoidmets D.; Huik K.; Härma M-A.; Jaaniso E.; Kaarna K.; Kallas E.; Koppel I.; Kuzmin I.; Lahesaare A.; Lutsar I.; Metspalu M.; Milani L.; Naaber P.; Niglas H.; Oopkaup O.E.; Pauskar M.; Peterson H.; Päll T.; Ratnik K.; Raudvere U.; Reisberg T.; Sadikova O.; Sepp H.; Shablinskaja A.; Suija H.; Talas U.G.; Truusalu K. |

We gratefully acknowledge the following Authors from the Originating laboratories responsible for obtaining the specimens, as well as the Submitting laboratories where the genome data were generated and shared via GISAID, on which this research is based.

All Submitters of data may be contacted directly via www.gisaid.org

Authors are sorted alphabetically.

| Accession ID | Originating Laboratory | Submitting Laboratory | Authors |
|---|---|---|---|
| EPI_ISL_8723794 | Department of Clinical Microbiology | GIGA Medical Genomics | Bouchra Boujemla; Claire Gourzonès; Cécile Meex; Keith Durkin; Laurent Gillet; Maria Artesi; Marie-Pierre Hayette; Nadine Cambisano; Nathalie Renotte; Olivier Ek; Sébastien Bontems; Vincent Bours |
| EPI_ISL_8713546, EPI_ISL_8714397, EPI_ISL_8714398, EPI_ISL_8714399 | Hrvatski zavod za javno zdravstvo | Hrvatski zavod za javno zdravstvo | Anita Jurić; Dragan Jurić; Irena Tabain; Ivana Ferenčak; Josipa Kuzle |
| EPI_ISL_8717998, EPI_ISL_8718000, EPI_ISL_8718008, EPI_ISL_8718009, EPI_ISL_8718012, EPI_ISL_8718016, EPI_ISL_8718017, EPI_ISL_8718018, EPI_ISL_8718025, EPI_ISL_8718036, EPI_ISL_8718037, EPI_ISL_8718038, EPI_ISL_8718049, EPI_ISL_8718051 | | | |
| see above | Idaho Bureau of Laboratories | Idaho Bureau of Laboratories | "R. Beukelman; Aimee Ceniseros; Christian Loera; Christopher Ball"; Matthew Charles Burns; Robert L. Voermans |
| EPI_ISL_8717063 | Jessa | Jessa | Severine Berden et al. on behalf of the Jessa_cmdLab |
| EPI_ISL_8725069 | KKP Kelas 1 Soekarno-Hatta (iLab) | National Institute of Health Research and Development | Arie Ardiansyah Nugraha; Fajar Nur Sulistiyohadi; Hana Apsari Pawestri; Hartanti Dian Ikawati; Kartika Dewi Puspa; Nelly Puspandari; Nur Ika Hariastuti; Putri Widia; Subangkit; Vivi Setiawaty |
| EPI_ISL_8726190, EPI_ISL_8726191, EPI_ISL_8726192 | Kansas Health and Environmental Lab | Kansas Health and Environmental Lab | Amanda Bradley; Carrie Welch; Gary Buruss; Jonathan Barnell; Mike Grose; Victor Anderson; and Phil Adam |
| EPI_ISL_8713004, EPI_ISL_8713006, EPI_ISL_8713008, EPI_ISL_8713015, EPI_ISL_8713016, EPI_ISL_8713017, EPI_ISL_8713020, EPI_ISL_8713022, EPI_ISL_8713025, EPI_ISL_8713032 | | | |
| see above | Kasturba Hospital Molecular Lab - INSACOG | Kasturba hospital WGS | Jayanthi Shastri; Vidushi Chitalia |
| EPI_ISL_8713269, EPI_ISL_8713270, EPI_ISL_8713271 | Klinika za infektivne bolesti "Dr. Fran Mihaljević" | Hrvatski zavod za javno zdravstvo | Anita Jurić; Dragan Jurić; Irena Tabain; Ivana Ferenčak; Josipa Kuzle |
| EPI_ISL_8718383, EPI_ISL_8718435, EPI_ISL_8718448, EPI_ISL_8718556, EPI_ISL_8718557, EPI_ISL_8718558, EPI_ISL_8718559 | | | |
| see above | Laboratory Corporation of America | Centers for Disease Control and Prevention Division of Viral Diseases, Pathogen Discovery | Amanda Douglas; Amanda Suchanek; Andrea Throop; Ayla Burns; Benjamin Rambo-Martin; Bobbi Croy; Brian Krueger; Brian Norvell; Christopher Gulvick; Christos Petropoulos; Clinton Paden; Craig Lukasik; Dakota Howard; Debbie Boles; Dhwani Batra; Duncan MacCannell; Eyad Almasri; Goran Stevovic; Howard Engler; Hrushikesh Deshmukh; Jake Humphrey; Jana Schroth; Jason Caravas; Joe Voshell; John Pruitt; Jonathan Meltzer; Jonathan Williams; Kimberly Wagner; Kristine Lacek; Lax Iyer; Lisa Pfefferle; Lyndon Tilson; Manoj Jain; Marcia Eisenberg; Mary Cristobal; Mary Williamson; Matthew Robinson; Matthew Schmerer; Michael Levandoski; Mike Sapeta; Mindy Nye; Minoo Agarwal; Mohan Kolli; Nuthawin Charoensri; Oren Cohen; Peter Cook; Prashant Gupta; Qian Zeng; Rama Ghatti; Scott Parker; Scott Ryan; Scott Sammons; Shatavia Morrison; Stanley Letovsky; Steven Ragan; Suresh Selvaraju; Susan Countryman; Susan Hicks; Suzanne Dale; Thomas Urban; Tim Kuphal; Tricia Zwiefelhofer; Tymeckia Kendall; Victoria Caban Figueroa; Vincent Drouillon; Yvette Unoarumhi |
| EPI_ISL_8713545 | Nastavni zavod za javno zdravstvo Primorsko-Goranske županije | Hrvatski zavod za javno zdravstvo | Anita Jurić; Dragan Jurić; Irena Tabain; Ivana Ferenčak; Josipa Kuzle |
| EPI_ISL_8713549 | Opća bolnica Bjelovar | Hrvatski zavod za javno zdravstvo | Anita Jurić; Dragan Jurić; Irena Tabain; Ivana Ferenčak; Josipa Kuzle |
| EPI_ISL_8713778 | Tulare County Public Health Laboratory | Tulare County Public Health Laboratory | Denise L Lopez; Joshua Kootstra |
| EPI_ISL_8713361, EPI_ISL_8714683 | Zavod za javno zdravstvo Dubrovačko-Neretvanske županije | Hrvatski zavod za javno zdravstvo | Anita Jurić; Dragan Jurić; Irena Tabain; Ivana Ferenčak; Josipa Kuzle |
| EPI_ISL_8713547 | Zavod za javno zdravstvo Sisačko-Moslavačke županije | Hrvatski zavod za javno zdravstvo | Anita Jurić; Dragan Jurić; Irena Tabain; Ivana Ferenčak; Josipa Kuzle |

We gratefully acknowledge the following Authors from the Originating laboratories responsible for obtaining the specimens, as well as the Submitting laboratories where the genome data were generated and shared via GISAID, on which this research is based.

All Submitters of data may be contacted directly via www.gisaid.org

Authors are sorted alphabetically.

| Accession ID | Originating Laboratory | Submitting Laboratory | Authors |
|---|---|---|---|
| EPI_ISL_8707664, EPI_ISL_8707673, EPI_ISL_8707679, EPI_ISL_8707683, EPI_ISL_8707688, EPI_ISL_8707690, EPI_ISL_8707769 | | | |
| see above | Department of Bacteria, Parasites and Fungi, Statens Serum Institut, Copenhagen, Denmark | Statens Serum Institut Bioinformatics and Microbial Genomics | Danish Covid-19 Genome Consortium |
| EPI_ISL_8712911, EPI_ISL_8712912, EPI_ISL_8712915, EPI_ISL_8712919, EPI_ISL_8712926, EPI_ISL_8712927, EPI_ISL_8712934, EPI_ISL_8712935, EPI_ISL_8712939, EPI_ISL_8712940, EPI_ISL_8712943, EPI_ISL_8712944, EPI_ISL_8712945, EPI_ISL_8712946, EPI_ISL_8712947, EPI_ISL_8712948, EPI_ISL_8712950, EPI_ISL_8712954, EPI_ISL_8712957, EPI_ISL_8712958, EPI_ISL_8712959, EPI_ISL_8712960, EPI_ISL_8712961, EPI_ISL_8712962, EPI_ISL_8712963, EPI_ISL_8712964, EPI_ISL_8712966, EPI_ISL_8712967, EPI_ISL_8712968, EPI_ISL_8712970, EPI_ISL_8712972, EPI_ISL_8712973, EPI_ISL_8712974, EPI_ISL_8712975, EPI_ISL_8712981, EPI_ISL_8712984, EPI_ISL_8712992, EPI_ISL_8712994, EPI_ISL_8712995, EPI_ISL_8712997, EPI_ISL_8713001, EPI_ISL_8713002, EPI_ISL_8713003 | | | |
| see above | Kasturba Hospital Molecular Lab - INSACOG | Kasturba hospital WGS | Jayanthi Shastri; Vidushi Chitalia |

We gratefully acknowledge the following Authors from the Originating laboratories responsible for obtaining the specimens, as well as the Submitting laboratories where the genome data were generated and shared via GISAID, on which this research is based.

All Submitters of data may be contacted directly via www.gisaid.org

Authors are sorted alphabetically.

Acknowledgement EPI_SET Identifier: EPI_SET_20220527ba

| Accession ID | Originating Laboratory | Submitting Laboratory | Authors |
|---|---|---|---|
| EPI_ISL_8707337, EPI_ISL_8707340, EPI_ISL_8707343, EPI_ISL_8707355, EPI_ISL_8707358, EPI_ISL_8707360, EPI_ISL_8707361, EPI_ISL_8707363, EPI_ISL_8707365, EPI_ISL_8707369, EPI_ISL_8707373, EPI_ISL_8707375, EPI_ISL_8707379, EPI_ISL_8707382, EPI_ISL_8707383, EPI_ISL_8707388, EPI_ISL_8707391, EPI_ISL_8707395, EPI_ISL_8707396, EPI_ISL_8707397, EPI_ISL_8707400, EPI_ISL_8707423, EPI_ISL_8707432, EPI_ISL_8707440, EPI_ISL_8707447, EPI_ISL_8707455, EPI_ISL_8707456, EPI_ISL_8707459, EPI_ISL_8707480, EPI_ISL_8707483, EPI_ISL_8707488, EPI_ISL_8707504, EPI_ISL_8707506, EPI_ISL_8707512, EPI_ISL_8707541, EPI_ISL_8707544, EPI_ISL_8707589, EPI_ISL_8707592, EPI_ISL_8707595, EPI_ISL_8707598, EPI_ISL_8707627, EPI_ISL_8707630, EPI_ISL_8707632, EPI_ISL_8707642, EPI_ISL_8707643, EPI_ISL_8707644, EPI_ISL_8707648, EPI_ISL_8707653, EPI_ISL_8707663 | | | |
| see above | Department of Bacteria, Parasites and Fungi, Statens Serum Institut, Copenhagen, Denmark | Statens Serum Institut Bioinformatics and Microbial Genomics | Danish Covid-19 Genome Consortium |
| EPI_ISL_8707535 | Lighthouse Lab in Glasgow | Wellcome Sanger Institute for the COVID-19 Genomics UK (COG-UK) Consortium | Anna Dominiczak and Alex Alderton; Carol Clugston; Cordelia Langford; David Gray; David K. Jackson; Dominic Kwiatkowski; Ewan Harrison; Harper VanSteenhouse; Ian Johnston; Jeffrey Barrett; John Sillitoe on behalf of the Wellcome Sanger Institute COVID-19 Surveillance Team; Roberto Amato; Sonia Goncalves; Yumi Kasai |

We gratefully acknowledge the following Authors from the Originating laboratories responsible for obtaining the specimens, as well as the Submitting laboratories where the genome data were generated and shared via GISAID, on which this research is based.

All Submitters of data may be contacted directly via www.gisaid.org

Authors are sorted alphabetically.

| Accession ID | Originating Laboratory | Submitting Laboratory | Authors |
|---|---|---|---|
| EPI_ISL_8706808, EPI_ISL_8706819, EPI_ISL_8706828, EPI_ISL_8706841, EPI_ISL_8706874, EPI_ISL_8706885, EPI_ISL_8706893, EPI_ISL_8706900, EPI_ISL_8706907, EPI_ISL_8706910, EPI_ISL_8706918, EPI_ISL_8706936, EPI_ISL_8706940, EPI_ISL_8706974, EPI_ISL_8706981, EPI_ISL_8707007, EPI_ISL_8707011, EPI_ISL_8707013, EPI_ISL_8707016, EPI_ISL_8707024, EPI_ISL_8707032, EPI_ISL_8707037, EPI_ISL_8707054, EPI_ISL_8707068, EPI_ISL_8707076, EPI_ISL_8707085, EPI_ISL_8707088, EPI_ISL_8707089, EPI_ISL_8707117, EPI_ISL_8707120, EPI_ISL_8707146, EPI_ISL_8707170, EPI_ISL_8707173, EPI_ISL_8707179, EPI_ISL_8707185, EPI_ISL_8707188, EPI_ISL_8707193, EPI_ISL_8707212, EPI_ISL_8707223, EPI_ISL_8707226, EPI_ISL_8707229, EPI_ISL_8707233, EPI_ISL_8707256, EPI_ISL_8707265, EPI_ISL_8707277, EPI_ISL_8707293, EPI_ISL_8707298, EPI_ISL_8707300, EPI_ISL_8707306, EPI_ISL_8707313 | | | |
| see above | Department of Bacteria, Parasites and Fungi, Statens Serum Institut, Copenhagen, Denmark | Statens Serum Institut Bioinformatics and Microbial Genomics | Danish Covid-19 Genome Consortium |

We gratefully acknowledge the following Authors from the Originating laboratories responsible for obtaining the specimens, as well as the Submitting laboratories where the genome data were generated and shared via GISAID, on which this research is based.

All Submitters of data may be contacted directly via www.gisaid.org

Authors are sorted alphabetically.

| Accession ID | Originating Laboratory | Submitting Laboratory | Authors |
| --- | --- | --- | --- |
| EPI_ISL_8706216, EPI_ISL_8706227, EPI_ISL_8706231, EPI_ISL_8706239, EPI_ISL_8706243, EPI_ISL_8706256, EPI_ISL_8706275, EPI_ISL_8706276, EPI_ISL_8706281, EPI_ISL_8706286, EPI_ISL_8706295, EPI_ISL_8706305, EPI_ISL_8706309, EPI_ISL_8706320, EPI_ISL_8706339, EPI_ISL_8706348, EPI_ISL_8706366, EPI_ISL_8706370, EPI_ISL_8706433, EPI_ISL_8706477, EPI_ISL_8706483, EPI_ISL_8706492, EPI_ISL_8706507, EPI_ISL_8706510, EPI_ISL_8706523, EPI_ISL_8706525, EPI_ISL_8706548, EPI_ISL_8706554, EPI_ISL_8706557, EPI_ISL_8706561, EPI_ISL_8706616, EPI_ISL_8706627, EPI_ISL_8706630, EPI_ISL_8706634, EPI_ISL_8706656, EPI_ISL_8706660, EPI_ISL_8706686, EPI_ISL_8706708, EPI_ISL_8706710, EPI_ISL_8706721, EPI_ISL_8706726, EPI_ISL_8706730, EPI_ISL_8706743, EPI_ISL_8706748, EPI_ISL_8706765, EPI_ISL_8706783, EPI_ISL_8706790, EPI_ISL_8706794, EPI_ISL_8706801 | | | |
| see above | Department of Bacteria, Parasites and Fungi, Statens Serum Institut, Copenhagen, Denmark | Statens Serum Institut Bioinformatics and Microbial Genomics | Danish Covid-19 Genome Consortium |
| EPI_ISL_8706738 | Lighthouse Laboratory Plymouth | Wellcome Sanger Institute for the COVID-19 Genomics UK (COG-UK) Consortium | Cordelia Langford; David K. Jackson; Dominic Kwiatkowski; Ewan Harrison; Ian Johnston; Jeffrey Barrett; John Sillitoe on behalf of the Wellcome Sanger Institute COVID-19 Surveillance Team; Lighthouse Laboratory Plymouth and Alex Alderton; Roberto Amato; Sonia Goncalves |

We gratefully acknowledge the following Authors from the Originating laboratories responsible for obtaining the specimens, as well as the Submitting laboratories where the genome data were generated and shared via GISAID, on which this research is based.

All Submitters of data may be contacted directly via www.gisaid.org

Authors are sorted alphabetically.

| Accession ID | Originating Laboratory | Submitting Laboratory | Authors |
|---|---|---|---|
| EPI_ISL_8731067, EPI_ISL_8731069, EPI_ISL_8731074 | CDPH VBL | California Department of Public Health | CDPH COVIDNet |
| EPI_ISL_8750817, EPI_ISL_8750827, EPI_ISL_8750828, EPI_ISL_8750829, EPI_ISL_8750830, EPI_ISL_8750833, EPI_ISL_8750834, EPI_ISL_8750835, EPI_ISL_8750836, EPI_ISL_8750837, EPI_ISL_8750838, EPI_ISL_8750839, EPI_ISL_8750840, EPI_ISL_8750841, EPI_ISL_8750843, EPI_ISL_8750845, EPI_ISL_8750846, EPI_ISL_8750847, EPI_ISL_8750848, EPI_ISL_8750849, EPI_ISL_8750850, EPI_ISL_8750851, EPI_ISL_8750852, EPI_ISL_8750853, EPI_ISL_8750854, EPI_ISL_8750855, EPI_ISL_8750856, EPI_ISL_8750857, EPI_ISL_8750859, EPI_ISL_8750861, EPI_ISL_8750865 | | | |
| see above | Idaho Bureau of Laboratories | Idaho Bureau of Laboratories | Aimee Ceniseros; Christian Loera; Christopher Ball; Matthew Charles Burns; R. Beukelman; Robert L. Voermans |
| EPI_ISL_8741961, EPI_ISL_8742089 | Lighthouse Lab in Alderley Park | Wellcome Sanger Institute for the COVID-19 Genomics UK (COG-UK) Consortium | Cordelia Langford; David K. Jackson; Dominic Kwiatkowski; Ewan Harrison; Ian Johnston; Jacquelyn Wynn; Jeffrey Barrett; John Sillitoe on behalf of the Wellcome Sanger Institute COVID-19 Surveillance Team; Mairead Hyland; Roberto Amato; Sonia Goncalves; The Lighthouse Lab in Alderley Park and Alex Alderton |
| EPI_ISL_8739949, EPI_ISL_8741098, EPI_ISL_8741193, EPI_ISL_8741905, EPI_ISL_8742094, EPI_ISL_8742139, EPI_ISL_8743347, EPI_ISL_8743488 | | | |
| see above | Lighthouse Lab in Glasgow | Wellcome Sanger Institute for the COVID-19 Genomics UK (COG-UK) Consortium | Anna Dominiczak and Alex Alderton; Carol Clugston; Cordelia Langford; David Gray; David K. Jackson; Dominic Kwiatkowski; Ewan Harrison; Harper VanSteenhouse; Ian Johnston; Jeffrey Barrett; John Sillitoe on behalf of the Wellcome Sanger Institute COVID-19 Surveillance Team; Roberto Amato; Sonia Goncalves; Yumi Kasai |
| EPI_ISL_8743240, EPI_ISL_8743465, EPI_ISL_8743484, EPI_ISL_8743485 | Lighthouse Lab in Milton Keynes | Wellcome Sanger Institute for the COVID-19 Genomics UK (COG-UK) Consortium | Cordelia Langford; David K. Jackson; Dominic Kwiatkowski; Ewan Harrison; Ian Johnston; Jeffrey Barrett; John Sillitoe on behalf of the Wellcome Sanger Institute COVID-19 Surveillance Team; Roberto Amato; Sonia Goncalves; The Lighthouse Lab in Milton Keynes and Alex Alderton |
| EPI_ISL_8739590, EPI_ISL_8739603, EPI_ISL_8739639, EPI_ISL_8739848, EPI_ISL_8739904 | Lighthouse Laboratory Plymouth | Wellcome Sanger Institute for the COVID-19 Genomics UK (COG-UK) Consortium | Cordelia Langford; David K. Jackson; Dominic Kwiatkowski; Ewan Harrison; Ian Johnston; Jeffrey Barrett; John Sillitoe on behalf of the Wellcome Sanger Institute COVID-19 Surveillance Team; Lighthouse Laboratory Plymouth and Alex Alderton; Roberto Amato; Sonia Goncalves |
| EPI_ISL_8730816, EPI_ISL_8730819, EPI_ISL_8730822, EPI_ISL_8730826, EPI_ISL_8730828, EPI_ISL_8730832, EPI_ISL_8730835 | | | |
| see above | Premier Laboratory Services | BioInfoExperts | Amy Feehan; Ben Lain; Chris Huston; David J. Nolan; Judy Crabtree; Julia-Garcia-Diaz; Lucio Miele; Rebecca Rose; Samual Moot; Susanna L. Lamers; Tessa LaFleur |
| EPI_ISL_8751571 | PrimBio Research Institute LLC | PrimBio Research Institute LLC | Diana Z. Ye; Yang Liu |
| EPI_ISL_8739601, EPI_ISL_8743432 | Rosalind Franklin Laboratory | Wellcome Sanger Institute for the COVID-19 Genomics UK (COG-UK) Consortium | Cordelia Langford; David K. Jackson; Dominic Kwiatkowski; Donald Fraser; Ewan Harrison; Ian Johnston; Jeffrey Barrett; John Sillitoe on behalf of the Wellcome Sanger Institute COVID-19 Surveillance Team; Rob Howes; Roberto Amato; Sonia Goncalves; Suki Lee; The Rosalind Franklin Laboratory and Alex Alderton |
| EPI_ISL_8744182 | Scientific Diagnostic Centre Pvt. Ltd., Ahmedabad | Gujarat Biotechnology Research Centre | Akhilesh Modi; Apurvasinh Puvar; Bhadreshsinh Gohil; Chaitanya Joshi; Disha Vora; Janvi Raval; Jaykumar Rangani; Madhvi Joshi; Moksha Narechania; Nimesh Patel; Nitin Savaliya; NitinShukla; Priyank Chavda; Ramesh Pandit; Roshani Mishra; Sonal Sharma; Tasnim Trivedi; Zarna Patel |
| EPI_ISL_8727742 | State Laboratories Division, Hawaii State Department of Health | State Laboratories Division, Hawaii State Department of Health | Ayana Garnet; Briana Ofilas; Cheryl-lynn Daquip; Cheyenne Barela; Daniel Strange; Drew Kuwazaki; Edward Desmond; Jeffrey Au; Mark Nagata; Pamela O'Brien; Razvan Sultana; Remedios Gose; Samantha Cotter; Samantha Sruba |
| EPI_ISL_8744253, EPI_ISL_8744274, EPI_ISL_8750077, EPI_ISL_8750095, EPI_ISL_8750103, EPI_ISL_8750240, EPI_ISL_8750270, EPI_ISL_8750308, EPI_ISL_8750356 | | | |
| see above | UW Virology Lab | UW Virology Lab | Alexander Greninger; Hong Xie; Isabel Arnould; Keith R Jerome; Meei-Li Huang; Nathan Breit; Patrick Mathias; Pavitra Roychoudhury; Pooneh Hajian; Ricardo Perez; Robert J. Livingston; Saraswathi Sathees; Sean Ellis; Seffir T. Wendm; Shah Mohamed Bakhash; Tien V. Nguyen |
| EPI_ISL_8733062, EPI_ISL_8733065, EPI_ISL_8733067, EPI_ISL_8733070, EPI_ISL_8733071, EPI_ISL_8733076, EPI_ISL_8733077, EPI_ISL_8733079, EPI_ISL_8733080, EPI_ISL_8733081, EPI_ISL_8733086, EPI_ISL_8733089, EPI_ISL_8733091, EPI_ISL_8733093, EPI_ISL_8733099, EPI_ISL_8733108, EPI_ISL_8733115, EPI_ISL_8733122, EPI_ISL_8733123, EPI_ISL_8733126, EPI_ISL_8733128, EPI_ISL_8733130, EPI_ISL_8733131, EPI_ISL_8733132, EPI_ISL_8733135, EPI_ISL_8733140, EPI_ISL_8733144, EPI_ISL_8733145, EPI_ISL_8733146, EPI_ISL_8733148, EPI_ISL_8733150, EPI_ISL_8733151, EPI_ISL_8733160, EPI_ISL_8733162, EPI_ISL_8733163, EPI_ISL_8733165, EPI_ISL_8733173, EPI_ISL_8733183, EPI_ISL_8733185, EPI_ISL_8733186, EPI_ISL_8733187, EPI_ISL_8733188, EPI_ISL_8733190, EPI_ISL_8733191, EPI_ISL_8733192, EPI_ISL_8733193, EPI_ISL_8733194, EPI_ISL_8733195, EPI_ISL_8733196, EPI_ISL_8733197, EPI_ISL_8733198, EPI_ISL_8733201, EPI_ISL_8733202, EPI_ISL_8733203, EPI_ISL_8733205, EPI_ISL_8733206, EPI_ISL_8733207, EPI_ISL_8733208, EPI_ISL_8733210, EPI_ISL_8733211, EPI_ISL_8733212, EPI_ISL_8733213, EPI_ISL_8733214, EPI_ISL_8733215, EPI_ISL_8733217, EPI_ISL_8733218, EPI_ISL_8733221, EPI_ISL_8733222, EPI_ISL_8733223, EPI_ISL_8733225, EPI_ISL_8733227, EPI_ISL_8733228, EPI_ISL_8733232, EPI_ISL_8733234, EPI_ISL_8733240, EPI_ISL_8733243, EPI_ISL_8733244, EPI_ISL_8733247, EPI_ISL_8733251, EPI_ISL_8733254, EPI_ISL_8733255, EPI_ISL_8733256, EPI_ISL_8733259, EPI_ISL_8733260, EPI_ISL_8733261, EPI_ISL_8733262, EPI_ISL_8733263, EPI_ISL_8733264, EPI_ISL_8733267, EPI_ISL_8733271, EPI_ISL_8733276, EPI_ISL_8733277, EPI_ISL_8733284, EPI_ISL_8733285, EPI_ISL_8733302, EPI_ISL_8733303, EPI_ISL_8733305, EPI_ISL_8733307, EPI_ISL_8733308, EPI_ISL_8733309, EPI_ISL_8733310, EPI_ISL_8733322, EPI_ISL_8733333, EPI_ISL_8733344, EPI_ISL_8733345, EPI_ISL_8733346, EPI_ISL_8733347, EPI_ISL_8733348, EPI_ISL_8733349, EPI_ISL_8733351, EPI_ISL_8733356, EPI_ISL_8733357, EPI_ISL_8733358, EPI_ISL_8733369, EPI_ISL_8733375, EPI_ISL_8733379, EPI_ISL_8733380, EPI_ISL_8733381, EPI_ISL_8733383, EPI_ISL_8733388, EPI_ISL_8733389, EPI_ISL_8733392, EPI_ISL_8733395, EPI_ISL_8733397, EPI_ISL_8733398, EPI_ISL_8733402, EPI_ISL_8733403, EPI_ISL_8733405, EPI_ISL_8733406, EPI_ISL_8733409, EPI_ISL_8733410, EPI_ISL_8733423, EPI_ISL_8733425, EPI_ISL_8733430, EPI_ISL_8733433, EPI_ISL_8733434, EPI_ISL_8733438, EPI_ISL_8733439, EPI_ISL_8733441, EPI_ISL_8733442, EPI_ISL_8733448, EPI_ISL_8733454, EPI_ISL_8733462, EPI_ISL_8733465, EPI_ISL_8733466, EPI_ISL_8733474, EPI_ISL_8733544, EPI_ISL_8733568, EPI_ISL_8733618, EPI_ISL_8733622, EPI_ISL_8733623, EPI_ISL_8733624, EPI_ISL_8733625, EPI_ISL_8733626, EPI_ISL_8733627, EPI_ISL_8733628, EPI_ISL_8733630, EPI_ISL_8733631, EPI_ISL_8733633, EPI_ISL_8733638, EPI_ISL_8733641, EPI_ISL_8733642, EPI_ISL_8733645, EPI_ISL_8733646, EPI_ISL_8733652, EPI_ISL_8733656, EPI_ISL_8733667, EPI_ISL_8733676, EPI_ISL_8733678, EPI_ISL_8733686, EPI_ISL_8733693, EPI_ISL_8733699, EPI_ISL_8733703, EPI_ISL_8733712, EPI_ISL_8733715, EPI_ISL_8733716, EPI_ISL_8733719, EPI_ISL_8733721, EPI_ISL_8733722, EPI_ISL_8733724, EPI_ISL_8733726, EPI_ISL_8733730, EPI_ISL_8733733, EPI_ISL_8733735, EPI_ISL_8733737 | | | |
| see above | University of Oregon COVID-19 MAP Laboratory | University of Oregon Genomics and Cell Characericzation Core Facility (GC3F) | Ariana White; Demi Glidden; Douglas Turnbull; Jason Carriere; Jason Sydes; Jeff Bishop; Megan Criss; Peter Batzel; Tina Arredondo |
| EPI_ISL_8751533 | Utah Public Health Laboratory | Utah Public Health Laboratory | Erin L. Young; John Arnn; Kelly F. Oakeson; Olinto Linares-Perdomo; Pooja Gupta; Tom Iverson |

We gratefully acknowledge the following Authors from the Originating laboratories responsible for obtaining the specimens, as well as the Submitting laboratories where the genome data were generated and shared via GISAID, on which this research is based.

All Submitters of data may be contacted directly via www.gisaid.org

Authors are sorted alphabetically.

Acknowledgement EPI_SET Identifier: EPI_SET_20220527rw

| Accession ID | Originating Laboratory | Submitting Laboratory | Authors |
|---|---|---|---|
| EPI_ISL_8754018 | Amirabad Virology Lab, Detection Unit | National Influenza Center | A Nejati; Asghar Abdoli; J Yavarian; K Sadeghi; NZ Shafiei Jandaghi; Sevrin Zadheidar and T Mokhtari Azad; V Salimi |
| EPI_ISL_8754645, EPI_ISL_8754697 | Berkshire and Surrey Pathology Services Lighthouse Laboratory | Wellcome Sanger Institute for the COVID-19 Genomics UK (COG-UK) Consortium | Berkshire and Surrey Pathology Services Lighthouse Laboratory and Alex Alderton; Cordelia Langford; David K. Jackson; Dominic Kwiatkowski; Ewan Harrison; Ian Johnston; Jeffrey Barrett; John Sillitoe on behalf of the Wellcome Sanger Institute COVID-19 Surveillance Team; Roberto Amato; Sonia Goncalves |
| EPI_ISL_8755609 | Integrated Covid Hub North East | Wellcome Sanger Institute for the COVID-19 Genomics UK (COG-UK) Consortium | Cordelia Langford; David K. Jackson; Dominic Kwiatkowski; Ewan Harrison; Ian Johnston; Integrated Covid Hub North East and Alex Alderton; Jeffrey Barrett; John Sillitoe on behalf of the Wellcome Sanger Institute COVID-19 Surveillance Team; Roberto Amato; Sonia Goncalves |
| EPI_ISL_8754374, EPI_ISL_8764986, EPI_ISL_8764987, EPI_ISL_8764988, EPI_ISL_8764999, EPI_ISL_8765002, EPI_ISL_8765011, EPI_ISL_8765014, EPI_ISL_8765028, EPI_ISL_8765030, EPI_ISL_8765031, EPI_ISL_8765033, EPI_ISL_8765034, EPI_ISL_8765037, EPI_ISL_8765039, EPI_ISL_8765041, EPI_ISL_8765045, EPI_ISL_8765046 | | | |
| see above | Lab. Microbiologia e Virologia Cotugno A.O. dei Colli | TIGEM | Antonio Grimaldi Patrizia Annunziata Francesco Panariello Claudia Tiberio Teresa Giuliano Valentina Bouche Chiara Colantuono Michela Daniele Lucio Di Filippo Anna Manfredi Marcello Salvi Antonio Limone Luigi Atripaldi Andrea Ballabio Davide Cacchiarelli |
| EPI_ISL_8754576 | Lighthouse Lab in Alderley Park | Wellcome Sanger Institute for the COVID-19 Genomics UK (COG-UK) Consortium | Cordelia Langford; David K. Jackson; Dominic Kwiatkowski; Ewan Harrison; Ian Johnston; Jacquelyn Wynn; Jeffrey Barrett; John Sillitoe on behalf of the Wellcome Sanger Institute COVID-19 Surveillance Team; Mairead Hyland; Roberto Amato; Sonia Goncalves; The Lighthouse Lab in Alderley Park and Alex Alderton |
| EPI_ISL_8751688 | NJDOH, Public Health and Environmental Laboratories | NJ_PHEL | Byeong Jeong; Chelsea San Filippo; Dana Woell; Lindsey Bodnar; Maria-Magdalene Pugliese; Mohammad M. Ali; Shiv K. Verma |
| EPI_ISL_8757535, EPI_ISL_8757536, EPI_ISL_8760108, EPI_ISL_8760117, EPI_ISL_8760121, EPI_ISL_8760127, EPI_ISL_8760144, EPI_ISL_8760148, EPI_ISL_8760165, EPI_ISL_8760169, EPI_ISL_8760215, EPI_ISL_8760235, EPI_ISL_8760241, EPI_ISL_8760841, EPI_ISL_8760844, EPI_ISL_8760872, EPI_ISL_8760883, EPI_ISL_8760950, EPI_ISL_8761055 | | | |
| see above | Respiratory Virus Unit, Microbiology Services Colindale, Public Health England | COVID-19 Genomics UK (COG-UK) Consortium | PHE Covid Sequencing Team |

We gratefully acknowledge the following Authors from the Originating laboratories responsible for obtaining the specimens, as well as the Submitting laboratories where the genome data were generated and shared via GISAID, on which this research is based.

All Submitters of data may be contacted directly via www.gisaid.org

Authors are sorted alphabetically.

| Accession ID | Originating Laboratory | Submitting Laboratory | Authors |
|---|---|---|---|
| EPI_ISL_8769739 | CNC PATH LAB | ILBS | Arjun Bhugra; Chhagan Bihari Sharma; Ekta Gupta; Pramod Gautam; Rahul Garg; Reshu Agarwal; Shiv Kumar Sarin; Urvinder Kaur; Varun Suroliya |
| EPI_ISL_8769645 | CORE Diagnostics Private Limited | ILBS | Arjun Bhugra; Chhagan Bihari Sharma; Ekta Gupta; Pramod Gautam; Rahul Garg; Reshu Agarwal; Shiv Kumar Sarin; Urvinder Kaur; Varun Suroliya |
| EPI_ISL_8767333, EPI_ISL_8767339, EPI_ISL_8767340, EPI_ISL_8767342, EPI_ISL_8767349, EPI_ISL_8767352, EPI_ISL_8767358, EPI_ISL_8767359, EPI_ISL_8767361, EPI_ISL_8767364, EPI_ISL_8767368, EPI_ISL_8767369, EPI_ISL_8767372, EPI_ISL_8767374, EPI_ISL_8767378, EPI_ISL_8767397, EPI_ISL_8767399, EPI_ISL_8767401, EPI_ISL_8767405, EPI_ISL_8767406, EPI_ISL_8767407, EPI_ISL_8767410, EPI_ISL_8767413, EPI_ISL_8767414, EPI_ISL_8767417, EPI_ISL_8767425, EPI_ISL_8767427, EPI_ISL_8767430, EPI_ISL_8767432, EPI_ISL_8767434, EPI_ISL_8767439, EPI_ISL_8767440, EPI_ISL_8767442, EPI_ISL_8767448, EPI_ISL_8767449, EPI_ISL_8767451, EPI_ISL_8767452, EPI_ISL_8767453, EPI_ISL_8767455, EPI_ISL_8767456, EPI_ISL_8767460, EPI_ISL_8767467, EPI_ISL_8767471, EPI_ISL_8767473, EPI_ISL_8767474, EPI_ISL_8767476, EPI_ISL_8767479, EPI_ISL_8767482, EPI_ISL_8767483, EPI_ISL_8767486, EPI_ISL_8767502, EPI_ISL_8767505, EPI_ISL_8767508, EPI_ISL_8767516, EPI_ISL_8767518, EPI_ISL_8767519, EPI_ISL_8767521, EPI_ISL_8767522, EPI_ISL_8767528, EPI_ISL_8767530, EPI_ISL_8767533, EPI_ISL_8767535, EPI_ISL_8767540, EPI_ISL_8767542, EPI_ISL_8767546, EPI_ISL_8767548, EPI_ISL_8767549, EPI_ISL_8767553, EPI_ISL_8767554, EPI_ISL_8767557, EPI_ISL_8767559, EPI_ISL_8767560, EPI_ISL_8767561, EPI_ISL_8767563, EPI_ISL_8767567, EPI_ISL_8767568, EPI_ISL_8767570, EPI_ISL_8767576, EPI_ISL_8767577, EPI_ISL_8767579, EPI_ISL_8767586, EPI_ISL_8767587, EPI_ISL_8767588, EPI_ISL_8767593, EPI_ISL_8767594, EPI_ISL_8767595, EPI_ISL_8767600, EPI_ISL_8767603, EPI_ISL_8767604, EPI_ISL_8767605, EPI_ISL_8767608, EPI_ISL_8767614, EPI_ISL_8767615, EPI_ISL_8767616, EPI_ISL_8767617, EPI_ISL_8767625, EPI_ISL_8767627, EPI_ISL_8767633, EPI_ISL_8767634, EPI_ISL_8767637, EPI_ISL_8767647, EPI_ISL_8767652, EPI_ISL_8767656, EPI_ISL_8767657, EPI_ISL_8767660, EPI_ISL_8767662, EPI_ISL_8767664, EPI_ISL_8767669, EPI_ISL_8767671, EPI_ISL_8767675, EPI_ISL_8767677, EPI_ISL_8767679, EPI_ISL_8767681, EPI_ISL_8767684, EPI_ISL_8767685, EPI_ISL_8767686, EPI_ISL_8767689, EPI_ISL_8767694, EPI_ISL_8767695, EPI_ISL_8767708, EPI_ISL_8767712, EPI_ISL_8767713, EPI_ISL_8767718, EPI_ISL_8767719, EPI_ISL_8767721, EPI_ISL_8767722, EPI_ISL_8767723, EPI_ISL_8767726, EPI_ISL_8767728, EPI_ISL_8767731, EPI_ISL_8767735, EPI_ISL_8767740, EPI_ISL_8767748, EPI_ISL_8767752, EPI_ISL_8767753, EPI_ISL_8767755, EPI_ISL_8767756, EPI_ISL_8767757, EPI_ISL_8767760, EPI_ISL_8767765, EPI_ISL_8767768, EPI_ISL_8767771, EPI_ISL_8767777, EPI_ISL_8767778, EPI_ISL_8767783, EPI_ISL_8767784, EPI_ISL_8767785, EPI_ISL_8767786, EPI_ISL_8767788, EPI_ISL_8767792, EPI_ISL_8767799, EPI_ISL_8767803, EPI_ISL_8767804, EPI_ISL_8767808, EPI_ISL_8767811, EPI_ISL_8767815, EPI_ISL_8767819, EPI_ISL_8767820, EPI_ISL_8767825, EPI_ISL_8767833, EPI_ISL_8767835, EPI_ISL_8767836, EPI_ISL_8767841, EPI_ISL_8767862, EPI_ISL_8767865, EPI_ISL_8767866, EPI_ISL_8767873, EPI_ISL_8767884, EPI_ISL_8767891, EPI_ISL_8767894, EPI_ISL_8767895, EPI_ISL_8767896, EPI_ISL_8767900, EPI_ISL_8767908, EPI_ISL_8767912, EPI_ISL_8767913, EPI_ISL_8767914, EPI_ISL_8767915, EPI_ISL_8767917, EPI_ISL_8767922, EPI_ISL_8767924, EPI_ISL_8767929, EPI_ISL_8767930, EPI_ISL_8767932, EPI_ISL_8767938, EPI_ISL_8767939, EPI_ISL_8767942, EPI_ISL_8767949, EPI_ISL_8767957, EPI_ISL_8767958, EPI_ISL_8767965, EPI_ISL_8767967, EPI_ISL_8767977, EPI_ISL_8767978, EPI_ISL_8767981, EPI_ISL_8767986, EPI_ISL_8767992, EPI_ISL_8767993, EPI_ISL_8768003, EPI_ISL_8768007, EPI_ISL_8768009, EPI_ISL_8768012, EPI_ISL_8768015, EPI_ISL_8768018, EPI_ISL_8768023, EPI_ISL_8768024, EPI_ISL_8768028, EPI_ISL_8768031, EPI_ISL_8768032, EPI_ISL_8768033, EPI_ISL_8768036, EPI_ISL_8768037, EPI_ISL_8768038, EPI_ISL_8768043, EPI_ISL_8768059, EPI_ISL_8768062, EPI_ISL_8768064, EPI_ISL_8768065, EPI_ISL_8768066, EPI_ISL_8768068, EPI_ISL_8768073, EPI_ISL_8768077, EPI_ISL_8768078, EPI_ISL_8768088, EPI_ISL_8768089, EPI_ISL_8768092, EPI_ISL_8768101, EPI_ISL_8768103, EPI_ISL_8768104, EPI_ISL_8768106, EPI_ISL_8768107, EPI_ISL_8768116, EPI_ISL_8768128, EPI_ISL_8768137, EPI_ISL_8768139, EPI_ISL_8768143, EPI_ISL_8768146, EPI_ISL_8768152, EPI_ISL_8768158, EPI_ISL_8768163, EPI_ISL_8768166, EPI_ISL_8768170, EPI_ISL_8768174, EPI_ISL_8768182, EPI_ISL_8768183, EPI_ISL_8768195, EPI_ISL_8768196, EPI_ISL_8768199, EPI_ISL_8768209, EPI_ISL_8768212, EPI_ISL_8768215, EPI_ISL_8768217, EPI_ISL_8768219, EPI_ISL_8768221, EPI_ISL_8768227, EPI_ISL_8768230, EPI_ISL_8768238, EPI_ISL_8768245, EPI_ISL_8768247, EPI_ISL_8768249, EPI_ISL_8768252, EPI_ISL_8768253, EPI_ISL_8768255, EPI_ISL_8768256, EPI_ISL_8768259, EPI_ISL_8768265, EPI_ISL_8768267, EPI_ISL_8768273, EPI_ISL_8768275, EPI_ISL_8768276, EPI_ISL_8768278, EPI_ISL_8768283, EPI_ISL_8768284, EPI_ISL_8768285, EPI_ISL_8768290, EPI_ISL_8768298, EPI_ISL_8768299, EPI_ISL_8768301, EPI_ISL_8768303, EPI_ISL_8768305, EPI_ISL_8768307, EPI_ISL_8768308, EPI_ISL_8768309, EPI_ISL_8768310, EPI_ISL_8768312, EPI_ISL_8768315, EPI_ISL_8768316, EPI_ISL_8768318, EPI_ISL_8768321, EPI_ISL_8768326, EPI_ISL_8768330, EPI_ISL_8768331, EPI_ISL_8768332, EPI_ISL_8768337, EPI_ISL_8768341, EPI_ISL_8768351, EPI_ISL_8768365, EPI_ISL_8768371, EPI_ISL_8768378, EPI_ISL_8768382, EPI_ISL_8768383, EPI_ISL_8768386, EPI_ISL_8768393, EPI_ISL_8768395, EPI_ISL_8768398, EPI_ISL_8768400, EPI_ISL_8768403 | | | |
| see above | Department of Bacteria, Parasites and Fungi, Statens Serum Institut, Copenhagen, Denmark | Statens Serum Institut Bioinformatics and Microbial Genomics | Danish Covid-19 Genome Consortium |
| EPI_ISL_8769564 | House of Diagnostics | ILBS | Arjun Bhugra; Chhagan Bihari Sharma; Ekta Gupta; Pramod Gautam; Rahul Garg; Reshu Agarwal; Shiv Kumar Sarin; Urvinder Kaur; Varun Suroliya |
| EPI_ISL_8791239, EPI_ISL_8791244 | Invenimus AG | Institute of Medical Virology, University of Zurich | Alexandra Trkola; Annette Audigé; Catharine Aquino; Cyril Shah; Daniel Ehrsam; Gabriela Ziltener; Guido Bloemberg; Hubert Rehrauer; Isabel Stürmer; Joel Wirz; Jon Huder; Jürg Böni; Kevin Steiner; Maria Grünberg; Maryam Zaheri; Michael Huber; Riccarda Capaul; Stefan Schmutz; Verena Kufner; Weihong Qi |
| EPI_ISL_8791242 | Kantonsspital Baden AG | Institute of Medical Virology, University of Zurich | Alexandra Trkola; Annette Audigé; Catharine Aquino; Cyril Shah; Daniel Ehrsam; Gabriela Ziltener; Guido Bloemberg; Hubert Rehrauer; Isabel Stürmer; Joel Wirz; Jon Huder; Jürg Böni; Kevin Steiner; Maria Grünberg; Maryam Zaheri; Michael Huber; Riccarda Capaul; Stefan Schmutz; Verena Kufner; Weihong Qi |
| EPI_ISL_8787930, EPI_ISL_8787931, EPI_ISL_8787932, EPI_ISL_8787934, EPI_ISL_8787935, EPI_ISL_8787936, EPI_ISL_8787937, EPI_ISL_8787938, EPI_ISL_8787941, EPI_ISL_8787942, EPI_ISL_8787943, EPI_ISL_8787944, EPI_ISL_8787945, EPI_ISL_8787946, EPI_ISL_8788031, EPI_ISL_8788039, EPI_ISL_8791189, EPI_ISL_8791191, EPI_ISL_8791194, EPI_ISL_8791272 | | | |
| see above | Kantonsspital Winterthur | Institute of Medical Virology, University of Zurich | Alexandra Trkola; Annette Audigé; Catharine Aquino; Cyril Shah; Daniel Ehrsam; Gabriela Ziltener; Guido Bloemberg; Hubert Rehrauer; Isabel Stürmer; Joel Wirz; Jon Huder; Jürg Böni; Kevin Steiner; Maria Grünberg; Maryam Zaheri; Michael Huber; Riccarda Capaul; Stefan Schmutz; Verena Kufner; Weihong Qi |
| EPI_ISL_8770363 | LABORATOIRE BIOALLIANCE | CNR Virus des Infections Respiratoires - France SUD | Antonin Bal; Bruno Lina; Bruno Simon; Gregory Destras; Gwendolyne Burfin; Hadrien Regue; Laurence Josset; Martine Valette; Quentin Semanas; ThÃ©ophile Boyer |
| EPI_ISL_8769742 | Medibird Diagnostics | ILBS | Arjun Bhugra; Chhagan Bihari Sharma; Ekta Gupta; Pramod Gautam; Rahul Garg; Reshu Agarwal; Shiv Kumar Sarin; Urvinder Kaur; Varun Suroliya |
| EPI_ISL_8769667, EPI_ISL_8769670 | NRL, Dr Lal Path Lab Ltds | ILBS | Arjun Bhugra; Chhagan Bihari Sharma; Ekta Gupta; Pramod Gautam; Rahul Garg; Reshu Agarwal; Shiv Kumar Sarin; Urvinder Kaur; Varun Suroliya |
| EPI_ISL_8770483, EPI_ISL_8770484, EPI_ISL_8770485, EPI_ISL_8770486, EPI_ISL_8770487, EPI_ISL_8770488, EPI_ISL_8770489, EPI_ISL_8770490, EPI_ISL_8770491, EPI_ISL_8770492, EPI_ISL_8770493, EPI_ISL_8770494, EPI_ISL_8770495, EPI_ISL_8770496, EPI_ISL_8770497, EPI_ISL_8770498, EPI_ISL_8770499, EPI_ISL_8770500, EPI_ISL_8770501, EPI_ISL_8770502, EPI_ISL_8770504, EPI_ISL_8770505, EPI_ISL_8770506, EPI_ISL_8770507, EPI_ISL_8770509, EPI_ISL_8770510, EPI_ISL_8770511, EPI_ISL_8770512, EPI_ISL_8770513, EPI_ISL_8770514, EPI_ISL_8770515, EPI_ISL_8770516, EPI_ISL_8770517, EPI_ISL_8770518, EPI_ISL_8770519, EPI_ISL_8770520, EPI_ISL_8770521, EPI_ISL_8770522, EPI_ISL_8770523, EPI_ISL_8770525, EPI_ISL_8770526, EPI_ISL_8770527, EPI_ISL_8770528, EPI_ISL_8770529, EPI_ISL_8770530, EPI_ISL_8770531, EPI_ISL_8770532, EPI_ISL_8770533, EPI_ISL_8770534, EPI_ISL_8770535, EPI_ISL_8770536, EPI_ISL_8770538, EPI_ISL_8770539, EPI_ISL_8770540, EPI_ISL_8770541, EPI_ISL_8770542, EPI_ISL_8770543 | | | |
| see above | National Public Health Laboratory, National Centre for Infectious Diseases | National Public Health Laboratory, National Centre for Infectious Diseases | Benny Yeo; Lin Cui; Raymond Tzer Pin Lin; Royce Ang; Samuel Loo; Zhenyang Zhou |
| EPI_ISL_8786600, EPI_ISL_8786641, EPI_ISL_8786643 | Nebraska Public Health Laboratory | NPHL COVID-19 Response Team | NPHL COVID-19 Response Team |
| EPI_ISL_8784572, EPI_ISL_8784610 | Originating lab: Wales Specialist Virology Centre Sequencing lab: Pathogen Genomics Unit | Public Health Wales Microbiology Cardiff Wales Specialist Virology Centre | Alec Birchley; Alexander Adams; Amy Gaskin; Angela Marchbank; Bree Gatica-Wilcox; Catherine Moore; Jason Coombes; Joanne Watkins; Joel Southgate; Johnathan Evans; Laura Gifford; Lauren Gilbert; Lee Graham; Malorie Perry; Matthew Bull; Nicole Pacchiarini; Sally Corden; Sara Kumziene-Summerhayes; Sara Rey; Sarah Taylor; Simon Cottrell; Sophie Jones; Tom Connor |
| EPI_ISL_8783172 | Polski Bank Komórek Macierzystych S.A. | WSSE w Warszawie | Dorota Wagrocka - Roczniak |
| EPI_ISL_8782794, EPI_ISL_8782795, EPI_ISL_8782796, EPI_ISL_8782799, EPI_ISL_8782800, EPI_ISL_8782803 | Sarawak General Hospital (Kuching) | Institute of Health and Community Medicine | Chan Chia Jui; Chua Hock Hin; David Perera; Ooi Mong How; Tonnii Sia Loong Loong |
| EPI_ISL_8782793, EPI_ISL_8782797, EPI_ISL_8782798 | Sibu Hospital (Sibu, Sarawak) | Institute of Health and Community Medicine | Chan Chia Jui; Chua Hock Hin; David Perera; Ooi Mong How; Tonnii Sia Loong Loong |
| EPI_ISL_8787953, EPI_ISL_8791204, EPI_ISL_8791240 | Stadtspital Waid | Institute of Medical Virology, University of Zurich | Alexandra Trkola; Annette Audigé; Catharine Aquino; Cyril Shah; Daniel Ehrsam; Gabriela Ziltener; Guido Bloemberg; Hubert Rehrauer; Isabel Stürmer; Joel Wirz; Jon Huder; Jürg Böni; Kevin Steiner; Maria Grünberg; Maryam Zaheri; Michael Huber; Riccarda Capaul; Stefan Schmutz; Verena Kufner; Weihong Qi |
| EPI_ISL_8791232 | USZ Flughafen | Institute of Medical Virology, University of Zurich | Alexandra Trkola; Annette Audigé; Catharine Aquino; Cyril Shah; Daniel Ehrsam; Gabriela Ziltener; Guido Bloemberg; Hubert Rehrauer; Isabel Stürmer; Joel Wirz; Jon Huder; Jürg Böni; Kevin Steiner; Maria Grünberg; Maryam Zaheri; Michael Huber; Riccarda Capaul; Stefan Schmutz; Verena Kufner; Weihong Qi |
| EPI_ISL_8787950, EPI_ISL_8787952, EPI_ISL_8787955, EPI_ISL_8787961, EPI_ISL_8787963, EPI_ISL_8787965, EPI_ISL_8787967, EPI_ISL_8787968, EPI_ISL_8787970, EPI_ISL_8787971, EPI_ISL_8787973, EPI_ISL_8787974, EPI_ISL_8787975, EPI_ISL_8787976, EPI_ISL_8787997, EPI_ISL_8788000, EPI_ISL_8788004, EPI_ISL_8788007, EPI_ISL_8788019, EPI_ISL_8791214, EPI_ISL_8791216, EPI_ISL_8791217, EPI_ISL_8791218, EPI_ISL_8791219, EPI_ISL_8791221, EPI_ISL_8791222, EPI_ISL_8791223, EPI_ISL_8791233, EPI_ISL_8791234, EPI_ISL_8791235, EPI_ISL_8791236, EPI_ISL_8791237, EPI_ISL_8791249, EPI_ISL_8791251, EPI_ISL_8791252, EPI_ISL_8791253 | | | |
| see above | Universität Zürich | Institute of Medical Virology, University of Zurich | Alexandra Trkola; Annette Audigé; Catharine Aquino; Cyril Shah; Daniel Ehrsam; Gabriela Ziltener; Guido Bloemberg; Hubert Rehrauer; Isabel Stürmer; Joel Wirz; Jon Huder; Jürg Böni; Kevin Steiner; Maria Grünberg; Maryam Zaheri; Michael Huber; Riccarda Capaul; Stefan Schmutz; Verena Kufner; Weihong Qi |
| EPI_ISL_8788011, EPI_ISL_8788015, EPI_ISL_8791202 | Universitäts-Kinderspital Zürich | Institute of Medical Virology, University of Zurich | Alexandra Trkola; Annette Audigé; Catharine Aquino; Cyril Shah; Daniel Ehrsam; Gabriela Ziltener; Guido Bloemberg; Hubert Rehrauer; Isabel Stürmer; Joel Wirz; Jon Huder; Jürg Böni; Kevin Steiner; Maria Grünberg; Maryam Zaheri; Michael Huber; Riccarda Capaul; Stefan Schmutz; Verena Kufner; Weihong Qi |
| EPI_ISL_8787954, EPI_ISL_8787978, EPI_ISL_8787980, EPI_ISL_8787983, EPI_ISL_8787990, EPI_ISL_8787991, EPI_ISL_8787992, EPI_ISL_8787994, EPI_ISL_8787995, EPI_ISL_8791195, EPI_ISL_8791197, EPI_ISL_8791199, EPI_ISL_8791200, EPI_ISL_8791205, EPI_ISL_8791206, EPI_ISL_8791208, EPI_ISL_8791209, EPI_ISL_8791211, EPI_ISL_8791243, EPI_ISL_8791245, EPI_ISL_8791246, EPI_ISL_8791247, EPI_ISL_8791254, EPI_ISL_8791256, EPI_ISL_8791260, EPI_ISL_8791261, EPI_ISL_8791264, EPI_ISL_8791265, EPI_ISL_8791266, EPI_ISL_8791267, EPI_ISL_8791270 | | | |
| see above | UniversitätsSpital Zürich | Institute of Medical Virology, University of Zurich | Alexandra Trkola; Annette Audigé; Catharine Aquino; Cyril Shah; Daniel Ehrsam; Gabriela Ziltener; Guido Bloemberg; Hubert Rehrauer; Isabel Stürmer; Joel Wirz; Jon Huder; Jürg Böni; Kevin Steiner; Maria Grünberg; Maryam Zaheri; Michael Huber; Riccarda Capaul; Stefan Schmutz; Verena Kufner; Weihong Qi |
| EPI_ISL_8788617, EPI_ISL_8788640 | Viollier AG | Department of Biosystems Science and Engineering, ETH Zürich | Andrea Patrignani; Andrea Patrizia Salzmann; Andreia Cabral de Gouvea; Catharine Aquino Fournier; Chaoran Chen; Christian Urban; Christiane Beckmann; Christoph Noppen; Daniel Ehrsam; David Dreifuss; Doris Popovic; Franziska Singer; Griffin White; Hai Bui; Henriette Kurth; Isabel Stürmer; Ivan Topolsky; Jay Tracy; Kim Philipp Jablonski; Lara Fuhrmann; Laura Neff; Lennart Opitz; Louis du Plessis; Maria Domenica Moccia; Matteo Carrara; Maurice Redondo; Niko Beerenwinkel; Olivier Kobel; Pelin Icer; Ralph Schlapbach; Sarah Nadeau; Shuqing Yu; Simon Grüter; Tanja Stadler; Timothy Sykes |
| EPI_ISL_8791540 | Virology Laboratory, Scientific Department, Army Medical Center | Virology Laboratory, Scientific Department, Army Medical Center | Anella Monte; Anna Anselmo; Antonella Fortunato; Filippo Molinari; Florigio Lista; Francesco Giordani; Giancarlo Petralito; Giandomenico Cerreto; Giulia Campoli; Lucia Nicosia; Marzia Cavalli; Pietro Marco D'Angelo; Riccardo De Sanctis; Rossella Brandi; Silvia Fillo; Vanessa Vera Fain |

We gratefully acknowledge the following Authors from the Originating laboratories responsible for obtaining the specimens, as well as the Submitting laboratories where the genome data were generated and shared via GISAID, on which this research is based.

All Submitters of data may be contacted directly via www.gisaid.org

Authors are sorted alphabetically.

| Accession ID | Originating Laboratory | Submitting Laboratory | Authors |
|---|---|---|---|
| EPI_ISL_8799962 | Cell-T Sp. z o.o. | 1. Tricity SARS-CoV-2 sequencing consortium: University of Gdansk, Medical University of Gdansk, Vaxican Ltd., Invicta Ltd. 2. National Institute of Public Health - National Institute of Hygiene, Warsaw, Poland | Celina Cybulska; Karolina Gackowska; Katarzyna Groth; Katarzyna Zacharczuk; Krystyna Bienkowska Szewczyk; Lukasz Rabalski; Maciej Grzybek; Maciej Kosinski; Magdalena Nowakowska; Marcin Lubocki; Małgorzata Sadkowska-Todys; Tomasz Wołkowicz |
| EPI_ISL_8795428, EPI_ISL_8795499, EPI_ISL_8795673, EPI_ISL_8795704 | Colorado Department of Public Health and Environment | Colorado Department of Public Health and Environment | Alexandria Rossheim; Arianna Smith; Diana Ir; Emily A. Travanty; Laura Bankers; Mandy Waters; Michael Martin; Molly C. Hetherington-Rauth; Shannon R. Matzinger |
| EPI_ISL_8803947, EPI_ISL_8804058, EPI_ISL_8804066, EPI_ISL_8804072, EPI_ISL_8804078, EPI_ISL_8804080, EPI_ISL_8804089, EPI_ISL_8804091, EPI_ISL_8804097, EPI_ISL_8804098, EPI_ISL_8804107, EPI_ISL_8804109, EPI_ISL_8804111, EPI_ISL_8804118, EPI_ISL_8804119, EPI_ISL_8804120, EPI_ISL_8804130, EPI_ISL_8804137, EPI_ISL_8804141, EPI_ISL_8804147, EPI_ISL_8804159, EPI_ISL_8804162, EPI_ISL_8804170, EPI_ISL_8804176, EPI_ISL_8804179, EPI_ISL_8804181, EPI_ISL_8804183, EPI_ISL_8804184, EPI_ISL_8804212, EPI_ISL_8804214, EPI_ISL_8804220, EPI_ISL_8804238, EPI_ISL_8804242, EPI_ISL_8804244, EPI_ISL_8804265, EPI_ISL_8804268, EPI_ISL_8804272, EPI_ISL_8804280, EPI_ISL_8804286, EPI_ISL_8804288, EPI_ISL_8804290, EPI_ISL_8804297, EPI_ISL_8804299, EPI_ISL_8804303, EPI_ISL_8804305, EPI_ISL_8804317, EPI_ISL_8804325, EPI_ISL_8804330, EPI_ISL_8804332, EPI_ISL_8804351, EPI_ISL_8804361, EPI_ISL_8804363, EPI_ISL_8804371, EPI_ISL_8804373, EPI_ISL_8804383, EPI_ISL_8804392, EPI_ISL_8804408, EPI_ISL_8804412, EPI_ISL_8804434, EPI_ISL_8804530, EPI_ISL_8804537, EPI_ISL_8804606, EPI_ISL_8804608, EPI_ISL_8804613, EPI_ISL_8804615, EPI_ISL_8804616, EPI_ISL_8804621, EPI_ISL_8804629, EPI_ISL_8804640, EPI_ISL_8804650, EPI_ISL_8804655, EPI_ISL_8804658, EPI_ISL_8804662, EPI_ISL_8804665, EPI_ISL_8804675, EPI_ISL_8804677, EPI_ISL_8804679, EPI_ISL_8804685, EPI_ISL_8804703, EPI_ISL_8804707, EPI_ISL_8804709, EPI_ISL_8804715, EPI_ISL_8804718, EPI_ISL_8804722, EPI_ISL_8804728, EPI_ISL_8804746, EPI_ISL_8804754, EPI_ISL_8804841, EPI_ISL_8804855, EPI_ISL_8804857, EPI_ISL_8804861, EPI_ISL_8804866, EPI_ISL_8804885, EPI_ISL_8804897, EPI_ISL_8804902, EPI_ISL_8804910, EPI_ISL_8804917, EPI_ISL_8804921, EPI_ISL_8804937, EPI_ISL_8804944, EPI_ISL_8804950, EPI_ISL_8804952, EPI_ISL_8804954, EPI_ISL_8804958, EPI_ISL_8804962, EPI_ISL_8804970, EPI_ISL_8804972, EPI_ISL_8804982, EPI_ISL_8804990, EPI_ISL_8804998, EPI_ISL_8805003, EPI_ISL_8805006, EPI_ISL_8805022, EPI_ISL_8805025, EPI_ISL_8805034, EPI_ISL_8805038, EPI_ISL_8805051, EPI_ISL_8805065, EPI_ISL_8805066, EPI_ISL_8805070, EPI_ISL_8805080, EPI_ISL_8805082, EPI_ISL_8805087, EPI_ISL_8805089, EPI_ISL_8805093, EPI_ISL_8805096, EPI_ISL_8805105, EPI_ISL_8805110, EPI_ISL_8805125, EPI_ISL_8805131, EPI_ISL_8805137, EPI_ISL_8805141, EPI_ISL_8805143, EPI_ISL_8805147, EPI_ISL_8805149, EPI_ISL_8805153, EPI_ISL_8805154, EPI_ISL_8805160, EPI_ISL_8805166, EPI_ISL_8805170, EPI_ISL_8805173, EPI_ISL_8805181, EPI_ISL_8805186, EPI_ISL_8805190, EPI_ISL_8805224, EPI_ISL_8805230, EPI_ISL_8805239, EPI_ISL_8805245, EPI_ISL_8805265, EPI_ISL_8805280, EPI_ISL_8805284, EPI_ISL_8805355, EPI_ISL_8805365, EPI_ISL_8805367, EPI_ISL_8805372, EPI_ISL_8805374, EPI_ISL_8805379, EPI_ISL_8805381, EPI_ISL_8805387, EPI_ISL_8805391, EPI_ISL_8805409, EPI_ISL_8805411, EPI_ISL_8805413, EPI_ISL_8805422, EPI_ISL_8805423, EPI_ISL_8805425, EPI_ISL_8805429, EPI_ISL_8805431, EPI_ISL_8805433, EPI_ISL_8805442, EPI_ISL_8805459, EPI_ISL_8805466, EPI_ISL_8805470, EPI_ISL_8805480, EPI_ISL_8805484, EPI_ISL_8805489, EPI_ISL_8805504, EPI_ISL_8805519, EPI_ISL_8805521, EPI_ISL_8805524, EPI_ISL_8805528, EPI_ISL_8805546, EPI_ISL_8805548, EPI_ISL_8805551, EPI_ISL_8805578, EPI_ISL_8805579, EPI_ISL_8805588, EPI_ISL_8805594, EPI_ISL_8805604, EPI_ISL_8805619, EPI_ISL_8805638, EPI_ISL_8805735, EPI_ISL_8805736, EPI_ISL_8805738, EPI_ISL_8805742, EPI_ISL_8805744, EPI_ISL_8805763, EPI_ISL_8805775, EPI_ISL_8805782, EPI_ISL_8805787, EPI_ISL_8805801, EPI_ISL_8805816, EPI_ISL_8805818, EPI_ISL_8805822, EPI_ISL_8805826, EPI_ISL_8805830, EPI_ISL_8805832, EPI_ISL_8805836, EPI_ISL_8805838, EPI_ISL_8805839, EPI_ISL_8805863, EPI_ISL_8805869, EPI_ISL_8805887, EPI_ISL_8805896, EPI_ISL_8805898, EPI_ISL_8805906, EPI_ISL_8805908, EPI_ISL_8805911, EPI_ISL_8805916, EPI_ISL_8805926, EPI_ISL_8805929, EPI_ISL_8805931, EPI_ISL_8805938, EPI_ISL_8805939, EPI_ISL_8805941, EPI_ISL_8805944, EPI_ISL_8805948, EPI_ISL_8805949, EPI_ISL_8805960, EPI_ISL_8805962, EPI_ISL_8805971, EPI_ISL_8805972, EPI_ISL_8805977, EPI_ISL_8805978, EPI_ISL_8805979, EPI_ISL_8805982, EPI_ISL_8806033, EPI_ISL_8806036, EPI_ISL_8806046, EPI_ISL_8806047, EPI_ISL_8806048, EPI_ISL_8806053, EPI_ISL_8806057, EPI_ISL_8806064, EPI_ISL_8806066, EPI_ISL_8806067, EPI_ISL_8806091, EPI_ISL_8806099, EPI_ISL_8806107, EPI_ISL_8806116, EPI_ISL_8806119, EPI_ISL_8806129, EPI_ISL_8806132, EPI_ISL_8806138, EPI_ISL_8806140, EPI_ISL_8806141, EPI_ISL_8806143, EPI_ISL_8806147, EPI_ISL_8806148, EPI_ISL_8806151, EPI_ISL_8806155, EPI_ISL_8806156, EPI_ISL_8806161, EPI_ISL_8806163, EPI_ISL_8806175, EPI_ISL_8806177, EPI_ISL_8806183, EPI_ISL_8806219, EPI_ISL_8806220, EPI_ISL_8806228, EPI_ISL_8806230, EPI_ISL_8806233, EPI_ISL_8806234, EPI_ISL_8806235, EPI_ISL_8806239, EPI_ISL_8806240, EPI_ISL_8806241, EPI_ISL_8806244, EPI_ISL_8806245, EPI_ISL_8806285, EPI_ISL_8806288, EPI_ISL_8806290, EPI_ISL_8806299, EPI_ISL_8806300, EPI_ISL_8806306, EPI_ISL_8806307, EPI_ISL_8806310, EPI_ISL_8806311, EPI_ISL_8806316, EPI_ISL_8806320, EPI_ISL_8806324, EPI_ISL_8806327, EPI_ISL_8806330, EPI_ISL_8806331, EPI_ISL_8806332, EPI_ISL_8806334, EPI_ISL_8806342, EPI_ISL_8806344, EPI_ISL_8806346, EPI_ISL_8806347, EPI_ISL_8806353, EPI_ISL_8806358, EPI_ISL_8806363, EPI_ISL_8806365, EPI_ISL_8806375, EPI_ISL_8806377, EPI_ISL_8806382, EPI_ISL_8806390, EPI_ISL_8806391, EPI_ISL_8806394, EPI_ISL_8806396, EPI_ISL_8806398, EPI_ISL_8806400, EPI_ISL_8806401, EPI_ISL_8806406, EPI_ISL_8806408, EPI_ISL_8806409, EPI_ISL_8806413, EPI_ISL_8806414, EPI_ISL_8806520, EPI_ISL_8806522, EPI_ISL_8806524, EPI_ISL_8806529, EPI_ISL_8806534, EPI_ISL_8806536, EPI_ISL_8806542, EPI_ISL_8806545, EPI_ISL_8806549, EPI_ISL_8806555, EPI_ISL_8806574, EPI_ISL_8806576, EPI_ISL_8806592, EPI_ISL_8806600, EPI_ISL_8806617, EPI_ISL_8806621, EPI_ISL_8806627, EPI_ISL_8806636, EPI_ISL_8806639, EPI_ISL_8806684, EPI_ISL_8806689, EPI_ISL_8806693, EPI_ISL_8806697, EPI_ISL_8807698, EPI_ISL_8807700, EPI_ISL_8807702, EPI_ISL_8807703, EPI_ISL_8807710, EPI_ISL_8807711, EPI_ISL_8807714, EPI_ISL_8807715, EPI_ISL_8807720, EPI_ISL_8807721, EPI_ISL_8807722, EPI_ISL_8807723, EPI_ISL_8807728, EPI_ISL_8807736 | | | |
| see above | Department of Bacteria, Parasites and Fungi, Statens Serum Institut, Copenhagen, Denmark | Statens Serum Institut Bioinformatics and Microbial Genomics | Danish Covid-19 Genome Consortium |
| EPI_ISL_8804062, EPI_ISL_8804122, EPI_ISL_8804129, EPI_ISL_8804134, EPI_ISL_8804146, EPI_ISL_8804198, EPI_ISL_8804228, EPI_ISL_8804234, EPI_ISL_8804240, EPI_ISL_8804256, EPI_ISL_8804274, EPI_ISL_8804283, EPI_ISL_8804294, EPI_ISL_8804296, EPI_ISL_8804321, EPI_ISL_8804334, EPI_ISL_8804357, EPI_ISL_8804367, EPI_ISL_8804531, EPI_ISL_8804544, EPI_ISL_8804610, EPI_ISL_8804646, EPI_ISL_8804649, EPI_ISL_8804681, EPI_ISL_8804683, EPI_ISL_8804687, EPI_ISL_8804694, EPI_ISL_8804726, EPI_ISL_8804756, EPI_ISL_8804764, EPI_ISL_8804865, EPI_ISL_8804872, EPI_ISL_8804893, EPI_ISL_8804974, EPI_ISL_8805058, EPI_ISL_8805121, EPI_ISL_8805127, EPI_ISL_8805232, EPI_ISL_8805247, EPI_ISL_8805258, EPI_ISL_8805282, EPI_ISL_8805351, EPI_ISL_8805386, EPI_ISL_8805397, EPI_ISL_8805443, EPI_ISL_8805455, EPI_ISL_8805496, EPI_ISL_8805498, EPI_ISL_8805536, EPI_ISL_8805582, EPI_ISL_8805592, EPI_ISL_8805608, EPI_ISL_8805612, EPI_ISL_8805627, EPI_ISL_8805631, EPI_ISL_8805753, EPI_ISL_8805773, EPI_ISL_8805795, EPI_ISL_8805820, EPI_ISL_8805851, EPI_ISL_8805892, EPI_ISL_8805915, EPI_ISL_8805925, EPI_ISL_8805959, EPI_ISL_8805981, EPI_ISL_8806037, EPI_ISL_8806041, EPI_ISL_8806045, EPI_ISL_8806056, EPI_ISL_8806059, EPI_ISL_8806062, EPI_ISL_8806087, EPI_ISL_8806088, EPI_ISL_8806104, EPI_ISL_8806123, EPI_ISL_8806157, EPI_ISL_8806174, EPI_ISL_8806218, EPI_ISL_8806222, EPI_ISL_8806229, EPI_ISL_8806232, EPI_ISL_8806284, EPI_ISL_8806388, EPI_ISL_8806525, EPI_ISL_8806597 | | | |
| see above | Department of Clinical Microbiology and Center for Genomic Medicine, Rigshospitalet, Copenhagen, Denmark | Statens Serum Institut Bioinformatics and Microbial Genomics | Danish Covid-19 Genome Consortium |
| EPI_ISL_8800370 | Department of Public Health Iasi | National Institute of Infectious Diseases-Prof. Dr. Matei Bals Molecular Diagnostics Laboratory | Corina Casangiu; Dan Otelea; Leontina Banica; Marius Surleac; Ovidiu Vlaicu; Petre Milu; Robert Hohan; Simona Paraschiv |
| EPI_ISL_8800328 | Department of Public Health Mures | National Institute of Infectious Diseases-Prof. Dr. Matei Bals Molecular Diagnostics Laboratory | Corina Casangiu; Dan Otelea; Leontina Banica; Marius Surleac; Ovidiu Vlaicu; Petre Milu; Robert Hohan; Simona Paraschiv |
| EPI_ISL_8801461, EPI_ISL_8801462, EPI_ISL_8801463, EPI_ISL_8801466, EPI_ISL_8801467 | KKP Kelas 1 Soekarno-Hatta (FarmaLab) | National Institute of Health Research and Development | Arie Ardiansyah Nugraha; Fajar Nur Sulistiyohadi; Hana Apsari Pawestri; Hartanti Dian Ikawati; Kartika Dewi Puspa; Nelly Puspandari; Nur Ika Hariastuti; Putri Widia; Subangkit; Vivi Setiawaty |
| EPI_ISL_8799194 | LEBOWAKGOMO LABORATORY | National Institute for Communicable Diseases of the National Health Laboratory Service | Amoako DG; Bhiman JN; Everatt J; Ismail A; Kekana D; Mahlangu B; Mnguni A; Mohale T; Ntuli N; Scheepers C; Wolter N |
| EPI_ISL_8799964 | Laboratoria Medyczne Bruss grupa ALAB sp. z o.o.; Medyczne Laboratorium Diagnostyczne, Pracownia Genetyki | 1. Tricity SARS-CoV-2 sequencing consortium: University of Gdansk, Medical University of Gdansk, Vaxican Ltd., Invicta Ltd. 2. National Institute of Public Health - National Institute of Hygiene, Warsaw, Poland | Celina Cybulska; Karolina Gackowska; Katarzyna Groth; Katarzyna Zacharczuk; Krystyna Bienkowska Szewczyk; Lukasz Rabalski; Maciej Grzybek; Maciej Kosinski; Magdalena Nowakowska; Marcin Lubocki; Małgorzata Sadkowska-Todys; Tomasz Wołkowicz |
| EPI_ISL_8800203 | National Institute of Infectious Diseases-Prof. Dr. Matei Bals Molecular Diagnostics Laboratory | National Institute of Infectious Diseases-Prof. Dr. Matei Bals Molecular Diagnostics Laboratory | Corina Casangiu; Dan Otelea; Leontina Banica; Marius Surleac; Ovidiu Vlaicu; Petre Milu; Robert Hohan; Simona Paraschiv |
| EPI_ISL_8806579 | Respiratory Virus Unit, Microbiology Services Colindale, Public Health England | COVID-19 Genomics UK (COG-UK) Consortium | PHE Covid Sequencing Team |
| EPI_ISL_8796869 | SK-Roy Romanow Provincial Laboratory | Saskatchewan - Roy Romanow Provincial Laboratory (RRPL) | Alanna Senecal; Amanda Lang; Jessica Minion; Kara Loos; Keith MacKenzie; Meredith Faires; Rachel DePaulo; Roy Romanow Provincial Laboratory - Molecular Diagnostics; Ryan McDonald |
| EPI_ISL_8807361 | UniversitätsSpital Zürich | Institute of Medical Virology, University of Zurich | Alexandra Trkola; Annette Audigé; Cyril Shah; Gabriela Ziltener; Guido Bloemberg; Jon Huder; Jürg Böni; Kevin Steiner; Maria Grünberg; Maryam Zaheri; Michael Huber; Riccarda Capaul; Stefan Schmutz; Verena Kufner |
| EPI_ISL_8801350, EPI_ISL_8801351, EPI_ISL_8801352, EPI_ISL_8801353, EPI_ISL_8801354, EPI_ISL_8801355, EPI_ISL_8801356, EPI_ISL_8801357, EPI_ISL_8801358, EPI_ISL_8801359, EPI_ISL_8801360, EPI_ISL_8801361, EPI_ISL_8801365, EPI_ISL_8801366, EPI_ISL_8801367, EPI_ISL_8801368, EPI_ISL_8801369, EPI_ISL_8801370, EPI_ISL_8801371, EPI_ISL_8801372, EPI_ISL_8801374, EPI_ISL_8801375, EPI_ISL_8801377, EPI_ISL_8801378, EPI_ISL_8801380 | | | |
| see above | VRDL,GMC,PTA | NIV Influenza | Dr.Rupinder Bakshi |

We gratefully acknowledge the following Authors from the Originating laboratories responsible for obtaining the specimens, as well as the Submitting laboratories where the genome data were generated and shared via GISAID, on which this research is based.

All Submitters of data may be contacted directly via www.gisaid.org

Authors are sorted alphabetically.

| Accession ID | Originating Laboratory | Submitting Laboratory | Authors |
|---|---|---|---|
| EPI_ISL_8814315, EPI_ISL_8814384, EPI_ISL_8815349 | Aegis Sciences Corporation | Centers for Disease Control and Prevention Division of Viral Diseases, Pathogen Discovery | Alec Vest; Benjamin Rambo-Martin; Christopher Gulvick; Clinton Paden; Cyndi Clark; Dakota Howard; Dhwani Batra; Dillon Nall; Duncan MacCannell; Erisa Sula; Ethan Sanders; Holly Houdeshell; Jason Caravas; Kristine Lacek; Matthew Hardison; Matthew Schmerer; Ola Kvalvaag; Patrick Campbell; Peter Cook; Rob Case; Scott Sammons; Shatavia Morrison; Shaun Westlund; Tymeckia Kendall; Victoria Caban Figueroa; Vikramsinha Ghorpade; Yvette Unoarumhi |
| EPI_ISL_8821241 | CDPH VBL | California Department of Public Health | CDPH COVIDNet |
| EPI_ISL_8807737, EPI_ISL_8807742, EPI_ISL_8807747, EPI_ISL_8807749, EPI_ISL_8807751, EPI_ISL_8807752, EPI_ISL_8807753, EPI_ISL_8807754, EPI_ISL_8807755, EPI_ISL_8807756, EPI_ISL_8807765, EPI_ISL_8807767, EPI_ISL_8807769, EPI_ISL_8807780, EPI_ISL_8807788, EPI_ISL_8807792, EPI_ISL_8807795, EPI_ISL_8807798, EPI_ISL_8807802, EPI_ISL_8807805, EPI_ISL_8807809, EPI_ISL_8807814, EPI_ISL_8807818, EPI_ISL_8807822, EPI_ISL_8807919, EPI_ISL_8807926, EPI_ISL_8807931, EPI_ISL_8807933, EPI_ISL_8807935, EPI_ISL_8807937, EPI_ISL_8807941, EPI_ISL_8807946 | | | |
| see above | Department of Bacteria, Parasites and Fungi, Statens Serum Institut, Copenhagen, Denmark | Statens Serum Institut Bioinformatics and Microbial Genomics | Danish Covid-19 Genome Consortium |
| EPI_ISL_8807741, EPI_ISL_8807775, EPI_ISL_8807793, EPI_ISL_8807819, EPI_ISL_8807832, EPI_ISL_8807921, EPI_ISL_8807924 | | | |
| see above | Department of Clinical Microbiology and Center for Genomic Medicine, Rigshospitalet, Copenhagen, Denmark | Statens Serum Institut Bioinformatics and Microbial Genomics | Danish Covid-19 Genome Consortium |
| EPI_ISL_8809026 | Florida Bureau of Public Health Laboratories | Florida Bureau of Public Health Laboratories | Jason Blanton; Namratha Tarigopula; Sarah Schmedes; Tiffany Splatt |
| EPI_ISL_8808115, EPI_ISL_8808118 | Jessa | Jessa | Severine Berden et al. on behalf of the Jessa_cmdLab |
| EPI_ISL_8809739, EPI_ISL_8809757 | Public Health Laboratory, Minnesota Department of Health | University of Minnesota Genomics Center | Corbin Dirkx; Daryl M. Gohl; Jaquelyn Kuriger-Laber; John Garbe |
| EPI_ISL_8824283 | Tripler Army Medical Center | Tripler Army Medical Center | Jonathan Andrew D'Ambrozio |
| EPI_ISL_8818108 | Wyoming Public Health Laboratory | Wyoming Public Health Laboratory | Ashley Norberg; Brian Dominguez; Cameron Ward; Carina Williams; Channing Weber; Chayse Rowley; Cyrus Rautman; Daniel Bolding; Elliot Thomasson; Emma Snow; Jim Mildenberger; Noah Cheshier; Rob Christensen; Robert Petit; Taylor Fearing; and Joseph Reed |

We gratefully acknowledge the following Authors from the Originating laboratories responsible for obtaining the specimens, as well as the Submitting laboratories where the genome data were generated and shared via GISAID, on which this research is based.

All Submitters of data may be contacted directly via www.gisaid.org

Authors are sorted alphabetically.

Acknowledgement EPI_SET Identifier: EPI_SET_20220527nb

| Accession ID | Originating Laboratory | Submitting Laboratory | Authors |
|---|---|---|---|
| EPI_ISL_8827119, EPI_ISL_8827901, EPI_ISL_8828165, EPI_ISL_8828166, EPI_ISL_8830893, EPI_ISL_8831308 | Aegis Sciences Corporation | Centers for Disease Control and Prevention Division of Viral Diseases, Pathogen Discovery | Alec Vest; Benjamin Rambo-Martin; Christopher Gulvick; Clinton Paden; Cyndi Clark; Dakota Howard; Dhwani Batra; Dillon Nall; Duncan MacCannell; Erisa Sula; Ethan Sanders; Holly Houdeshell; Jason Caravas; Kristine Lacek; Matthew Hardison; Matthew Schmerer; Ola Kvalvaag; Patrick Campbell; Peter Cook; Rob Case; Scott Sammons; Shatavia Morrison; Shaun Westlund; Tymeckia Kendall; Victoria Caban Figueroa; Vikramsinha Ghorpade; Yvette Unoarumhi |
| EPI_ISL_8826519 | Botswana Harvard HIV Reference Laboratory | Botswana Harvard HIV Reference Laboratory | Boitumelo J. L Zuze; Botshelo Radibe; Dorcas Maruapula; Joseph Makhema; Keoratile Ntshambiwa; Kgomotso Moruisi; Legodile Kooepile; Letsibogo Gaoraelwe; Mosepele Mosepele; Mphaphi B. Mbulawa; Ontlametse T. Bareng; Pamela Smith-Lawrence; Roger Shapiro; Sefetogi Ramaologa; Shahin Lockman; Sikhulile Moyo; Simani Gaseitsiwe; Thela Tefelo; Thongbotho Mphoyakgosi; Wonderful T. Choga |
| EPI_ISL_8831657, EPI_ISL_8831659, EPI_ISL_8831669, EPI_ISL_8831671, EPI_ISL_8831686, EPI_ISL_8831691, EPI_ISL_8831695, EPI_ISL_8831697, EPI_ISL_8831703, EPI_ISL_8831705, EPI_ISL_8831721, EPI_ISL_8831735, EPI_ISL_8831736, EPI_ISL_8831738, EPI_ISL_8831743, EPI_ISL_8831750, EPI_ISL_8831758, EPI_ISL_8831764, EPI_ISL_8831769, EPI_ISL_8831774, EPI_ISL_8831782, EPI_ISL_8831789, EPI_ISL_8831801, EPI_ISL_8831807, EPI_ISL_8831811, EPI_ISL_8831816, EPI_ISL_8831833, EPI_ISL_8831836, EPI_ISL_8831847, EPI_ISL_8831849, EPI_ISL_8831856, EPI_ISL_8831857, EPI_ISL_8831863, EPI_ISL_8831870, EPI_ISL_8831871, EPI_ISL_8831876, EPI_ISL_8831880, EPI_ISL_8831893, EPI_ISL_8831908, EPI_ISL_8831910, EPI_ISL_8831916, EPI_ISL_8831926, EPI_ISL_8831929, EPI_ISL_8831932, EPI_ISL_8831935, EPI_ISL_8831936, EPI_ISL_8831938, EPI_ISL_8831944, EPI_ISL_8831947, EPI_ISL_8831949, EPI_ISL_8831962, EPI_ISL_8831966, EPI_ISL_8831975, EPI_ISL_8831984, EPI_ISL_8831985, EPI_ISL_8831989, EPI_ISL_8831994, EPI_ISL_8831996, EPI_ISL_8832011, EPI_ISL_8832012, EPI_ISL_8832021, EPI_ISL_8832024, EPI_ISL_8832027, EPI_ISL_8832029, EPI_ISL_8832035, EPI_ISL_8832037, EPI_ISL_8832038, EPI_ISL_8832040, EPI_ISL_8832054, EPI_ISL_8832060, EPI_ISL_8832071, EPI_ISL_8832077, EPI_ISL_8832089, EPI_ISL_8832092, EPI_ISL_8832099, EPI_ISL_8832111, EPI_ISL_8832118, EPI_ISL_8832143, EPI_ISL_8832148, EPI_ISL_8832153, EPI_ISL_8832161, EPI_ISL_8832165, EPI_ISL_8832169, EPI_ISL_8832182, EPI_ISL_8832191, EPI_ISL_8832193, EPI_ISL_8832198, EPI_ISL_8832210, EPI_ISL_8832215, EPI_ISL_8832226, EPI_ISL_8832235, EPI_ISL_8832239, EPI_ISL_8832241, EPI_ISL_8832249, EPI_ISL_8832259, EPI_ISL_8832265, EPI_ISL_8832269, EPI_ISL_8832275, EPI_ISL_8832276, EPI_ISL_8832285, EPI_ISL_8832298, EPI_ISL_8832313, EPI_ISL_8832315, EPI_ISL_8832354, EPI_ISL_8832356, EPI_ISL_8832358, EPI_ISL_8832359, EPI_ISL_8832361, EPI_ISL_8832365, EPI_ISL_8832379, EPI_ISL_8832381, EPI_ISL_8832385, EPI_ISL_8832387, EPI_ISL_8832389, EPI_ISL_8832398, EPI_ISL_8832415, EPI_ISL_8832419, EPI_ISL_8832421, EPI_ISL_8832423, EPI_ISL_8832429, EPI_ISL_8832432, EPI_ISL_8832434, EPI_ISL_8832444, EPI_ISL_8832451, EPI_ISL_8832452, EPI_ISL_8832453, EPI_ISL_8832459, EPI_ISL_8832473, EPI_ISL_8832480, EPI_ISL_8832487, EPI_ISL_8832488, EPI_ISL_8832492, EPI_ISL_8832504, EPI_ISL_8832510, EPI_ISL_8832517, EPI_ISL_8832525, EPI_ISL_8832528, EPI_ISL_8832539, EPI_ISL_8832541, EPI_ISL_8832545, EPI_ISL_8832551, EPI_ISL_8832552, EPI_ISL_8832554, EPI_ISL_8832557, EPI_ISL_8832566, EPI_ISL_8832568, EPI_ISL_8832576, EPI_ISL_8832579, EPI_ISL_8832581, EPI_ISL_8832583, EPI_ISL_8832587, EPI_ISL_8832599, EPI_ISL_8832605, EPI_ISL_8832607, EPI_ISL_8832618, EPI_ISL_8832619, EPI_ISL_8832621, EPI_ISL_8832632, EPI_ISL_8832638, EPI_ISL_8832640, EPI_ISL_8832651, EPI_ISL_8832661, EPI_ISL_8832665, EPI_ISL_8832666, EPI_ISL_8832676, EPI_ISL_8832681, EPI_ISL_8832682, EPI_ISL_8832685, EPI_ISL_8832686, EPI_ISL_8832700, EPI_ISL_8832708, EPI_ISL_8832710, EPI_ISL_8832716, EPI_ISL_8832720, EPI_ISL_8832722, EPI_ISL_8832724, EPI_ISL_8832730, EPI_ISL_8832731, EPI_ISL_8832735, EPI_ISL_8832741, EPI_ISL_8832750, EPI_ISL_8832754, EPI_ISL_8832756, EPI_ISL_8832758, EPI_ISL_8832766, EPI_ISL_8832768, EPI_ISL_8832769, EPI_ISL_8832771, EPI_ISL_8832775, EPI_ISL_8832781, EPI_ISL_8832783, EPI_ISL_8832791, EPI_ISL_8832793, EPI_ISL_8832795, EPI_ISL_8832797, EPI_ISL_8832800, EPI_ISL_8832802, EPI_ISL_8832808, EPI_ISL_8832810, EPI_ISL_8832811, EPI_ISL_8832826, EPI_ISL_8832846, EPI_ISL_8832851, EPI_ISL_8832854, EPI_ISL_8832864, EPI_ISL_8832871, EPI_ISL_8832875, EPI_ISL_8832877, EPI_ISL_8832881, EPI_ISL_8832886, EPI_ISL_8832898, EPI_ISL_8832903, EPI_ISL_8832909, EPI_ISL_8832927, EPI_ISL_8832940, EPI_ISL_8832944, EPI_ISL_8832950, EPI_ISL_8832957, EPI_ISL_8832961, EPI_ISL_8832975, EPI_ISL_8832981, EPI_ISL_8832995, EPI_ISL_8833015, EPI_ISL_8833017, EPI_ISL_8833021, EPI_ISL_8833022, EPI_ISL_8833030, EPI_ISL_8833031, EPI_ISL_8833044, EPI_ISL_8833053, EPI_ISL_8833055, EPI_ISL_8833067, EPI_ISL_8833069, EPI_ISL_8833077, EPI_ISL_8833095, EPI_ISL_8833101, EPI_ISL_8833109, EPI_ISL_8833130, EPI_ISL_8833132, EPI_ISL_8833135, EPI_ISL_8833142, EPI_ISL_8833150, EPI_ISL_8833154, EPI_ISL_8833156, EPI_ISL_8833164, EPI_ISL_8833180, EPI_ISL_8833192, EPI_ISL_8833198, EPI_ISL_8833204, EPI_ISL_8833212, EPI_ISL_8833214, EPI_ISL_8833217, EPI_ISL_8833218, EPI_ISL_8833221, EPI_ISL_8833222, EPI_ISL_8833237, EPI_ISL_8833326, EPI_ISL_8833335, EPI_ISL_8833342, EPI_ISL_8833346, EPI_ISL_8833357, EPI_ISL_8833366, EPI_ISL_8833368, EPI_ISL_8833375, EPI_ISL_8833379, EPI_ISL_8833382, EPI_ISL_8833384, EPI_ISL_8833385, EPI_ISL_8833387, EPI_ISL_8833393, EPI_ISL_8833395, EPI_ISL_8833415, EPI_ISL_8833421, EPI_ISL_8833424, EPI_ISL_8833441, EPI_ISL_8833445, EPI_ISL_8833451, EPI_ISL_8833458, EPI_ISL_8833461, EPI_ISL_8833471, EPI_ISL_8833482, EPI_ISL_8833488, EPI_ISL_8833489, EPI_ISL_8833491, EPI_ISL_8833519, EPI_ISL_8833522, EPI_ISL_8833531, EPI_ISL_8833534, EPI_ISL_8833538, EPI_ISL_8833546, EPI_ISL_8833551, EPI_ISL_8833558, EPI_ISL_8833566, EPI_ISL_8833568, EPI_ISL_8833571, EPI_ISL_8833574, EPI_ISL_8833582, EPI_ISL_8833583, EPI_ISL_8833594, EPI_ISL_8833602, EPI_ISL_8833605, EPI_ISL_8833607, EPI_ISL_8833613, EPI_ISL_8833615, EPI_ISL_8833618, EPI_ISL_8833619, EPI_ISL_8833623, EPI_ISL_8833625, EPI_ISL_8833645, EPI_ISL_8833650, EPI_ISL_8833664, EPI_ISL_8833675, EPI_ISL_8833679, EPI_ISL_8833680, EPI_ISL_8833681, EPI_ISL_8833684, EPI_ISL_8833693, EPI_ISL_8833701, EPI_ISL_8833705, EPI_ISL_8833730, EPI_ISL_8833737, EPI_ISL_8833758, EPI_ISL_8833760, EPI_ISL_8833765, EPI_ISL_8833766, EPI_ISL_8833770, EPI_ISL_8833772, EPI_ISL_8833774, EPI_ISL_8833779, EPI_ISL_8833784, EPI_ISL_8833814, EPI_ISL_8833819, EPI_ISL_8833821, EPI_ISL_8833823, EPI_ISL_8833840, EPI_ISL_8833846, EPI_ISL_8833848, EPI_ISL_8833852, EPI_ISL_8833857, EPI_ISL_8833858, EPI_ISL_8833859, EPI_ISL_8833865, EPI_ISL_8833892, EPI_ISL_8833894, EPI_ISL_8833898, EPI_ISL_8833900, EPI_ISL_8833913, EPI_ISL_8833917, EPI_ISL_8833924, EPI_ISL_8833937, EPI_ISL_8833941, EPI_ISL_8833949, EPI_ISL_8833951, EPI_ISL_8833954, EPI_ISL_8833978, EPI_ISL_8833981, EPI_ISL_8833983, EPI_ISL_8833988, EPI_ISL_8833989, EPI_ISL_8833992, EPI_ISL_8833999, EPI_ISL_8834001, EPI_ISL_8834002, EPI_ISL_8834003, EPI_ISL_8834005, EPI_ISL_8834017, EPI_ISL_8834019, EPI_ISL_8834022, EPI_ISL_8834033, EPI_ISL_8834039, EPI_ISL_8834042, EPI_ISL_8834049, EPI_ISL_8834058, EPI_ISL_8834065, EPI_ISL_8834070, EPI_ISL_8834080, EPI_ISL_8834081, EPI_ISL_8834083, EPI_ISL_8834084, EPI_ISL_8834101, EPI_ISL_8834102, EPI_ISL_8834125, EPI_ISL_8834156, EPI_ISL_8834158, EPI_ISL_8834170, EPI_ISL_8834174, EPI_ISL_8834179, EPI_ISL_8834185, EPI_ISL_8834196, EPI_ISL_8834201, EPI_ISL_8834206, EPI_ISL_8834228, EPI_ISL_8834239, EPI_ISL_8834241, EPI_ISL_8834243, EPI_ISL_8834245, EPI_ISL_8834247, EPI_ISL_8834248, EPI_ISL_8834250, EPI_ISL_8834262, EPI_ISL_8834269, EPI_ISL_8834273, EPI_ISL_8834277, EPI_ISL_8834280, EPI_ISL_8834281, EPI_ISL_8834283, EPI_ISL_8834292, EPI_ISL_8834293, EPI_ISL_8834297, EPI_ISL_8834299, EPI_ISL_8834302, EPI_ISL_8834306, EPI_ISL_8834314, EPI_ISL_8834321, EPI_ISL_8834323, EPI_ISL_8834327, EPI_ISL_8834335, EPI_ISL_8834339 | | | |
| see above | Department of Bacteria, Parasites and Fungi, Statens Serum Institut, Copenhagen, Denmark | Statens Serum Institut Bioinformatics and Microbial Genomics | Danish Covid-19 Genome Consortium |
| EPI_ISL_8831684, EPI_ISL_8831787, EPI_ISL_8831793, EPI_ISL_8831823, EPI_ISL_8831906, EPI_ISL_8831951, EPI_ISL_8832056, EPI_ISL_8832110, EPI_ISL_8832137, EPI_ISL_8832171, EPI_ISL_8832174, EPI_ISL_8832176, EPI_ISL_8832200, EPI_ISL_8832304, EPI_ISL_8832306, EPI_ISL_8832340, EPI_ISL_8832350, EPI_ISL_8832523, EPI_ISL_8832593, EPI_ISL_8832678, EPI_ISL_8832745, EPI_ISL_8832774, EPI_ISL_8832872, EPI_ISL_8832953, EPI_ISL_8833057, EPI_ISL_8833195, EPI_ISL_8833220, EPI_ISL_8833338, EPI_ISL_8833397, EPI_ISL_8833416, EPI_ISL_8833495, EPI_ISL_8833496, EPI_ISL_8833507, EPI_ISL_8833526, EPI_ISL_8833542, EPI_ISL_8833589, EPI_ISL_8833617, EPI_ISL_8833632, EPI_ISL_8833648, EPI_ISL_8833710, EPI_ISL_8833744, EPI_ISL_8833794, EPI_ISL_8833842, EPI_ISL_8833860, EPI_ISL_8833918, EPI_ISL_8833991, EPI_ISL_8834010, EPI_ISL_8834021, EPI_ISL_8834028, EPI_ISL_8834053, EPI_ISL_8834149, EPI_ISL_8834213, EPI_ISL_8834222, EPI_ISL_8834253, EPI_ISL_8834271 | | | |
| see above | Department of Clinical Microbiology and Center for Genomic Medicine, Rigshospitalet, Copenhagen, Denmark | Statens Serum Institut Bioinformatics and Microbial Genomics | Danish Covid-19 Genome Consortium |
| EPI_ISL_8830460, EPI_ISL_8830479 | GenePath, Pune | RF-IISER Pune | Aurnab Ghose; Joy Merwin Monteiro; Krishanpal Karmodiya; Nikhil Phadke; Shatakshi Ranade |
| EPI_ISL_8833405 | KKP Kelas 1 Soekarno-Hatta (iLab) | National Institute of Health Research and Development | Arie Ardiansyah Nugraha; Fajar Nur Sulistiyohadi; Hana Apsari Pawestri; Hartanti Dian Ikawati; Kartika Dewi Puspa; Nelly Puspandari; Nur Ika Hariastuti; Putri Widia; Subangkit; Vivi Setiawaty |
| EPI_ISL_8826405, EPI_ISL_8826406, EPI_ISL_8826408, EPI_ISL_8826429 | Lancet Laboratories Gaborone | Botswana Harvard HIV Reference Laboratory | Boitumelo Zuze; Botshelo Radibe; Dorcas Maruapula; Ellen Nkitse; Joseph Makhema; Keoratile Ntshambiwa; Kgomotso Moruisi; Legodile Kooepile; Leyi Ncube; Mosana Moyo; Mosepele Mosepele; Mphaphi B. Mbulawa; Ontlametse T. Bareng; Pamela Smith-Lawrence; Roger Shapiro; Sefetogi Ramaologa; Shahin Lockman; Sikhulile Moyo; Simani Gaseitsiwe; Thongbotho Mphoyakgosi; Wonderful T. Choga |
| EPI_ISL_8826412, EPI_ISL_8826449 | National Health Laboratory | Botswana Harvard HIV Reference Laboratory | Boitumelo J. L Zuze; Botshelo Radibe; Dorcas Maruapula; Joseph Makhema; Keoratile Ntshambiwa; Kgomotso Moruisi; Legodile Kooepile; Letsibogo Gaoraelwe; Mosepele Mosepele; Mphaphi B. Mbulawa; Ontlametse T. Bareng; Pamela Smith-Lawrence; Roger Shapiro; Sefetogi Ramaologa; Shahin Lockman; Sikhulile Moyo; Simani Gaseitsiwe; Thela Tefelo; Thongbotho Mphoyakgosi; Wonderful T. Choga |
| EPI_ISL_8828687 | Nevada State Public Health Laboratory | Nevada State Public Health Laboratory | Andrew Gorzalski; Lynette Gumbleton; Mark Pandori |
| EPI_ISL_8833404 | PRVKP FKUI | National Institute of Health Research and Development | Arie Ardiansyah Nugraha; Fajar Nur Sulistiyohadi; Hana Apsari Pawestri; Hartanti Dian Ikawati; Kartika Dewi Puspa; Nelly Puspandari; Subangkit; Triyani Soekarso; Vivi Setiawaty |
| EPI_ISL_8826512, EPI_ISL_8826515, EPI_ISL_8826523 | Palapye Primary Hospital Laboratory | Botswana Harvard HIV Reference Laboratory | Boitumelo J. L Zuze; Botshelo Radibe; Dorcas Maruapula; Joseph Makhema; Keoratile Ntshambiwa; Kgomotso Moruisi; Legodile Kooepile; Letsibogo Gaoraelwe; Mosepele Mosepele; Mphaphi B. Mbulawa; Ontlametse T. Bareng; Pamela Smith-Lawrence; Roger Shapiro; Sefetogi Ramaologa; Shahin Lockman; Sikhulile Moyo; Simani Gaseitsiwe; Thela Tefelo; Thongbotho Mphoyakgosi; Wonderful T. Choga |
| EPI_ISL_8829822 | Spital Langenthal | Medizinische Klinik, Spital Region Oberaargau | Alexander Imhof; Cedric Howald; Deborah Penet; Henri Pegeot; Ioannis Xenarios; Keith Harshman; Lorenzo Cerutti; Melyssa Elies |
| EPI_ISL_8828357 | University of New Mexico Hospital | Center for Global Health, University of New Mexico Health Sciences Center | Darrell Dinwiddie; Daryl Domman; Jesse Young; Jon Femling; Justin Bacca; Kurt Schwalm; Valerie Morley |

We gratefully acknowledge the following Authors from the Originating laboratories responsible for obtaining the specimens, as well as the Submitting laboratories where the genome data were generated and shared via GISAID, on which this research is based.

All Submitters of data may be contacted directly via www.gisaid.org

Authors are sorted alphabetically.

| Accession ID | Originating Laboratory | Submitting Laboratory | Authors |
|---|---|---|---|
| EPI_ISL_8870728, EPI_ISL_8870770, EPI_ISL_8871007, EPI_ISL_8871878 | Aegis Sciences Corporation | Centers for Disease Control and Prevention Division of Viral Diseases, Pathogen Discovery | Alec Vest; Benjamin Rambo-Martin; Christopher Gulvick; Clinton Paden; Cyndi Clark; Dakota Howard; Dhwani Batra; Dillon Nall; Duncan MacCannell; Erisa Sula; Ethan Sanders; Holly Houdeshell; Jason Caravas; Kristine Lacek; Matthew Hardison; Matthew Schmerer; Ola Kvalvaag; Patrick Campbell; Peter Cook; Rob Case; Scott Sammons; Shatavia Morrison; Shaun Westlund; Tymeckia Kendall; Victoria Caban Figueroa; Vikramsinha Ghorpade; Yvette Unoarumhi |
| EPI_ISL_8868207 | Dept of Microbiology, Maulana Azad Medical College | GENOME SEQUENCING LABORATARY, Dept of Pediatrics, LOK NAYAK Hospital | Arvind Mohan; Avinash Lomash; INSACOG; Meenakshi Bothra; Mohammed Faruq; Oves Siddiqui; Prashanth N Suravajhala; Seema Kapoor.Sandeep Garg; Somesh Kumar; Sonal Saxena; Sunil Kumar Polipalli; Suresh Kumar; Vikas Manchanda |
| EPI_ISL_8844502, EPI_ISL_8847010, EPI_ISL_8847087 | Edmonton Provincial Lab | Public Health Agency of Canada (PHAC) National Microbiology Laboratory | Buss; Croxen M; Deo A; Dieu P; E; Ferrato C; Gill K; Khan F; Koleva P; Li V; Lloyd C; Lynch T; Ma R; Murphy S; Pabbaraju K; Shokoples S; Thayer J; Tipples G; Whitehouse M; Wong A; Yu C; Zelyas N |
| EPI_ISL_8876222 | FYR Diagnostics | FYR Diagnostics | Blohm; Ghai; Tre; Vikas |
| EPI_ISL_8848580, EPI_ISL_8848581, EPI_ISL_8848582, EPI_ISL_8848584, EPI_ISL_8848797, EPI_ISL_8854084 | Lighthouse Lab in Alderley Park | Wellcome Sanger Institute for the COVID-19 Genomics UK (COG-UK) Consortium | Cordelia Langford; David K. Jackson; Dominic Kwiatkowski; Ewan Harrison; Ian Johnston; Jacquelyn Wynn; Jeffrey Barrett; John Sillitoe on behalf of the Wellcome Sanger Institute COVID-19 Surveillance Team; Mairead Hyland; Roberto Amato; Sonia Goncalves; The Lighthouse Lab in Alderley Park and Alex Alderton |
| EPI_ISL_8864505 | Lighthouse Lab in Glasgow | Wellcome Sanger Institute for the COVID-19 Genomics UK (COG-UK) Consortium | Anna Dominiczak and Alex Alderton; Carol Clugston; Cordelia Langford; David Gray; David K. Jackson; Dominic Kwiatkowski; Ewan Harrison; Harper VanSteenhouse; Ian Johnston; Jeffrey Barrett; John Sillitoe on behalf of the Wellcome Sanger Institute COVID-19 Surveillance Team; Roberto Amato; Sonia Goncalves; Yumi Kasai |
| EPI_ISL_8863120, EPI_ISL_8863128 | Michigan Department of Health and Human Services, Bureau of Laboratories | Michigan Department of Health and Human Services, Bureau of Laboratories | Blankenship HM; Riner D; Soehnlen MK |
| EPI_ISL_8843407 | Ministry of Health Turkey | Ministry of Health Turkey | Fatma Bayrakdar; Gülay Korukluoğlu; Gültekin Ünal; Süleyman Yalcin; Yasemin Coşgun |
| EPI_ISL_8839391, EPI_ISL_8839392, EPI_ISL_8839393, EPI_ISL_8839394, EPI_ISL_8839395, EPI_ISL_8839396, EPI_ISL_8839397, EPI_ISL_8839398, EPI_ISL_8839399 | | | |
| see above | Nastavni zavod za javno zdravstvo Splitsko- Dalmatinske županije | Hrvatski zavod za javno zdravstvo | Anita Jurić; Dragan Jurić; Irena Tabain; Ivana Ferenčak; Josipa Kuzle |
| EPI_ISL_8859258, EPI_ISL_8859272 | PathWest Laboratory Medicine WA | PathWest Laboratory Medicine WA Microbial Surveillance Unit | PathWest Laboratory Medicine WA Microbial Surveillance Unit |
| EPI_ISL_8841258 | Platform BIS UZA/UAntwerpen | Labo Klinische Biologie, UZA | Basil Britto Xavier; Christine Lammens; Herman Goossens; Ines Verbesselt; Jasmine Coppens; Kathleen Holemans; Marie Le Mercier; Veerle Matheeussen |
| EPI_ISL_8873222, EPI_ISL_8873234, EPI_ISL_8873236, EPI_ISL_8873241, EPI_ISL_8873243 | Quest Diagnostics Incorporated | Centers for Disease Control and Prevention Division of Viral Diseases, Pathogen Discovery | A. Gerasimova; A. Perez; B. Anderson; Benjamin Rambo-Martin; Christopher Gulvick; Clinton Paden; Dakota Howard; Dhwani Batra; Duncan MacCannell; Erisa Sula; F. Lacbawan; I. Shlyakhter; Jason Caravas; K. Livingston; Kristine Lacek; L. Bernstein; M. Hua; Matthew Schmerer; P. Tanpaiboon; Peter Cook; R. Kagan; R. Owen; R. Rolando; S. Rosenthal; Scott Sammons; Shatavia Morrison; Tymeckia Kendall; Victoria Caban Figueroa; Y. Liu; Yvette Unoarumhi |
| EPI_ISL_8840922, EPI_ISL_8840937, EPI_ISL_8840983, EPI_ISL_8840998 | Respiratory Virus Unit, Microbiology Services Colindale, Public Health England | COVID-19 Genomics UK (COG-UK) Consortium | PHE Covid Sequencing Team |
| EPI_ISL_8841176, EPI_ISL_8841177, EPI_ISL_8841178, EPI_ISL_8841191, EPI_ISL_8841193, EPI_ISL_8841227, EPI_ISL_8841228, EPI_ISL_8841237 | | | |
| see above | Virology Unit, Institut Pasteur du Cambodge | Virology Unit, Institut Pasteur du Cambodge | Cecile Troupin; Chau Darapheak; Chin Savuth; Erik A Karlsson; Jurre Y Siegers; Kraing Sidonn; Leakhena Pum; Ly Sovann; Sophoannadedh Rath; Veasna Duong; Yi Sengdoeurn |
| EPI_ISL_8840071 | Zavod za javno zdravstvo Istarske županije | Hrvatski zavod za javno zdravstvo | Anita Jurić; Dragan Jurić; Irena Tabain; Ivana Ferenčak; Josipa Kuzle |
| EPI_ISL_8843990 | Zavod za javno zdravstvo Zagrebačke županije | Hrvatski zavod za javno zdravstvo | Anita Jurić; Dragan Jurić; Irena Tabain; Ivana Ferenčak; Josipa Kuzle |

We gratefully acknowledge the following Authors from the Originating laboratories responsible for obtaining the specimens, as well as the Submitting laboratories where the genome data were generated and shared via GISAID, on which this research is based.

All Submitters of data may be contacted directly via www.gisaid.org

Authors are sorted alphabetically.

| Accession ID | Originating Laboratory | Submitting Laboratory | Authors |
|---|---|---|---|
| EPI_ISL_8835086, EPI_ISL_8835091, EPI_ISL_8835098, EPI_ISL_8835119, EPI_ISL_8835123, EPI_ISL_8835128, EPI_ISL_8835129, EPI_ISL_8835133, EPI_ISL_8835137, EPI_ISL_8835139, EPI_ISL_8835146, EPI_ISL_8835152, EPI_ISL_8835154, EPI_ISL_8835156, EPI_ISL_8835163, EPI_ISL_8835164, EPI_ISL_8835166, EPI_ISL_8835168, EPI_ISL_8835171, EPI_ISL_8835176, EPI_ISL_8835178, EPI_ISL_8835190, EPI_ISL_8835196, EPI_ISL_8835205, EPI_ISL_8835207, EPI_ISL_8835227, EPI_ISL_8835231, EPI_ISL_8835236, EPI_ISL_8835239, EPI_ISL_8835249, EPI_ISL_8835251, EPI_ISL_8835269, EPI_ISL_8835275 | | | |
| see above | Department of Bacteria, Parasites and Fungi, Statens Serum Institut, Copenhagen, Denmark | Statens Serum Institut Bioinformatics and Microbial Genomics | Danish Covid-19 Genome Consortium |
| EPI_ISL_8835116, EPI_ISL_8835157, EPI_ISL_8835218, EPI_ISL_8835223, EPI_ISL_8835282 | Department of Clinical Microbiology and Center for Genomic Medicine, Rigshospitalet, Copenhagen, Denmark | Statens Serum Institut Bioinformatics and Microbial Genomics | Danish Covid-19 Genome Consortium |
| EPI_ISL_8836880, EPI_ISL_8836881, EPI_ISL_8836882, EPI_ISL_8836883, EPI_ISL_8836884, EPI_ISL_8836885, EPI_ISL_8836886, EPI_ISL_8836887, EPI_ISL_8836888 | | | |
| see above | Division of Emerging Infectious Diseases, Bureau of Infectious Diseases Diagnosis Control, Korea Disease Control and Prevention Agency | Division of Emerging Infectious Diseases, Bureau of Infectious Diseases Diagnosis Control, Korea Disease Control and Prevention Agency | Ae Kyung Park; Chae Young Lee; Eun-Jin Kim; Hyuck Jin Lee; Il-Hwan Kim; Jeong-Ah Kim |
| EPI_ISL_8837764, EPI_ISL_8837795 | Vitomed Sp. z o.o. | 1. Academic Center for Pathomorphological and Genetic-Molecular Diagnostics ltd, Bialystok, Poland 2. National Institute of Public Health - National Institute of Hygiene, Warsaw, Poland | Anetta Sulewska; Jacek Nikliński; Janusz Dzięcioł; Joanna Kiśluk; Katarzyna Zacharczuk; Konrad Raczkowski; Magdalena Nowakowska; Małgorzata Sadkowska-Todys; Piotr Karabowicz; Piotr Majewski; Przemysław Biecek. Joanna Reszeć; Radosław Charkiewicz; Tomasz Wołkowicz |
| EPI_ISL_8837797 | Wojewódzki Szpital Zespolony w Lesznie | 1. Academic Center for Pathomorphological and Genetic-Molecular Diagnostics ltd, Bialystok, Poland 2. National Institute of Public Health - National Institute of Hygiene, Warsaw, Poland | Anetta Sulewska; Jacek Nikliński; Janusz Dzięcioł; Joanna Kiśluk; Katarzyna Zacharczuk; Konrad Raczkowski; Magdalena Nowakowska; Małgorzata Sadkowska-Todys; Piotr Karabowicz; Piotr Majewski; Przemysław Biecek. Joanna Reszeć; Radosław Charkiewicz; Tomasz Wołkowicz |

We gratefully acknowledge the following Authors from the Originating laboratories responsible for obtaining the specimens, as well as the Submitting laboratories where the genome data were generated and shared via GISAID, on which this research is based.

All Submitters of data may be contacted directly via www.gisaid.org

Authors are sorted alphabetically.

Acknowledgement EPI_SET Identifier: EPI_SET_20220527by

| Accession ID | Originating Laboratory | Submitting Laboratory | Authors |
|---|---|---|---|
| EPI_ISL_8834794, EPI_ISL_8834803, EPI_ISL_8834810, EPI_ISL_8834817, EPI_ISL_8834825, EPI_ISL_8834838, EPI_ISL_8834841, EPI_ISL_8834864, EPI_ISL_8834876, EPI_ISL_8834878, EPI_ISL_8834885, EPI_ISL_8834891, EPI_ISL_8834892, EPI_ISL_8834906, EPI_ISL_8834908, EPI_ISL_8834926, EPI_ISL_8834938, EPI_ISL_8834940, EPI_ISL_8834952, EPI_ISL_8834954, EPI_ISL_8834956, EPI_ISL_8834965, EPI_ISL_8834967, EPI_ISL_8834970, EPI_ISL_8834971, EPI_ISL_8834985, EPI_ISL_8834987, EPI_ISL_8834997, EPI_ISL_8835001, EPI_ISL_8835006, EPI_ISL_8835013, EPI_ISL_8835015, EPI_ISL_8835018, EPI_ISL_8835019, EPI_ISL_8835024, EPI_ISL_8835032, EPI_ISL_8835041, EPI_ISL_8835043, EPI_ISL_8835049, EPI_ISL_8835055, EPI_ISL_8835064, EPI_ISL_8835066, EPI_ISL_8835071, EPI_ISL_8835072, EPI_ISL_8835075, EPI_ISL_8835076, EPI_ISL_8835080 | | | |
| see above | Department of Bacteria, Parasites and Fungi, Statens Serum Institut, Copenhagen, Denmark | Statens Serum Institut Bioinformatics and Microbial Genomics | Danish Covid-19 Genome Consortium |
| EPI_ISL_8834857, EPI_ISL_8834934, EPI_ISL_8835068 | Department of Clinical Microbiology and Center for Genomic Medicine, Rigshospitalet, Copenhagen, Denmark | Statens Serum Institut Bioinformatics and Microbial Genomics | Danish Covid-19 Genome Consortium |

We gratefully acknowledge the following Authors from the Originating laboratories responsible for obtaining the specimens, as well as the Submitting laboratories where the genome data were generated and shared via GISAID, on which this research is based.

All Submitters of data may be contacted directly via www.gisaid.org

Authors are sorted alphabetically.

| Accession ID | Originating Laboratory | Submitting Laboratory | Authors |
|---|---|---|---|
| EPI_ISL_8834544, EPI_ISL_8834547, EPI_ISL_8834558, EPI_ISL_8834560, EPI_ISL_8834582, EPI_ISL_8834585, EPI_ISL_8834588, EPI_ISL_8834593, EPI_ISL_8834595, EPI_ISL_8834597, EPI_ISL_8834612, EPI_ISL_8834616, EPI_ISL_8834626, EPI_ISL_8834636, EPI_ISL_8834639, EPI_ISL_8834641, EPI_ISL_8834646, EPI_ISL_8834654, EPI_ISL_8834667, EPI_ISL_8834671, EPI_ISL_8834685, EPI_ISL_8834688, EPI_ISL_8834690, EPI_ISL_8834696, EPI_ISL_8834703, EPI_ISL_8834705, EPI_ISL_8834707, EPI_ISL_8834709, EPI_ISL_8834711, EPI_ISL_8834716, EPI_ISL_8834720, EPI_ISL_8834721, EPI_ISL_8834727, EPI_ISL_8834730, EPI_ISL_8834734, EPI_ISL_8834739, EPI_ISL_8834751, EPI_ISL_8834756, EPI_ISL_8834758, EPI_ISL_8834764, EPI_ISL_8834770, EPI_ISL_8834776, EPI_ISL_8834780, EPI_ISL_8834783, EPI_ISL_8834785, EPI_ISL_8834792 | | | |
| see above | Department of Bacteria, Parasites and Fungi, Statens Serum Institut, Copenhagen, Denmark | Statens Serum Institut Bioinformatics and Microbial Genomics | Danish Covid-19 Genome Consortium |
| EPI_ISL_8834580, EPI_ISL_8834591, EPI_ISL_8834599, EPI_ISL_8834729 | Department of Clinical Microbiology and Center for Genomic Medicine, Rigshospitalet, Copenhagen, Denmark | Statens Serum Institut Bioinformatics and Microbial Genomics | Danish Covid-19 Genome Consortium |

We gratefully acknowledge the following Authors from the Originating laboratories responsible for obtaining the specimens, as well as the Submitting laboratories where the genome data were generated and shared via GISAID, on which this research is based.

All Submitters of data may be contacted directly via www.gisaid.org

Authors are sorted alphabetically.

| Accession ID | Originating Laboratory | Submitting Laboratory | Authors |
| --- | --- | --- | --- |
| EPI_ISL_8834345, EPI_ISL_8834357, EPI_ISL_8834365, EPI_ISL_8834367, EPI_ISL_8834372, EPI_ISL_8834374, EPI_ISL_8834380, EPI_ISL_8834381, EPI_ISL_8834384, EPI_ISL_8834388, EPI_ISL_8834400, EPI_ISL_8834402, EPI_ISL_8834405, EPI_ISL_8834407, EPI_ISL_8834412, EPI_ISL_8834417, EPI_ISL_8834419, EPI_ISL_8834424, EPI_ISL_8834426, EPI_ISL_8834435, EPI_ISL_8834438, EPI_ISL_8834444, EPI_ISL_8834452, EPI_ISL_8834454, EPI_ISL_8834456, EPI_ISL_8834459, EPI_ISL_8834465, EPI_ISL_8834470, EPI_ISL_8834474, EPI_ISL_8834477, EPI_ISL_8834479, EPI_ISL_8834482, EPI_ISL_8834484, EPI_ISL_8834488, EPI_ISL_8834498, EPI_ISL_8834500, EPI_ISL_8834503, EPI_ISL_8834508, EPI_ISL_8834511, EPI_ISL_8834517, EPI_ISL_8834519, EPI_ISL_8834535, EPI_ISL_8834537, EPI_ISL_8834540 | | | |
| see above | Department of Bacteria, Parasites and Fungi, Statens Serum Institut, Copenhagen, Denmark | Statens Serum Institut Bioinformatics and Microbial Genomics | Danish Covid-19 Genome Consortium |
| EPI_ISL_8834344, EPI_ISL_8834354, EPI_ISL_8834391, EPI_ISL_8834396, EPI_ISL_8834449, EPI_ISL_8834496 | Department of Clinical Microbiology and Center for Genomic Medicine, Rigshospitalet, Copenhagen, Denmark | Statens Serum Institut Bioinformatics and Microbial Genomics | Danish Covid-19 Genome Consortium |

We gratefully acknowledge the following Authors from the Originating laboratories responsible for obtaining the specimens, as well as the Submitting laboratories where the genome data were generated and shared via GISAID, on which this research is based.

All Submitters of data may be contacted directly via www.gisaid.org

Authors are sorted alphabetically.

Acknowledgement EPI_SET Identifier: EPI_SET_20220527ck

| Accession ID | Originating Laboratory | Submitting Laboratory | Authors |
|---|---|---|---|
| EPI_ISL_8886634, EPI_ISL_8886635, EPI_ISL_8886637 | Addu Equitorial Hospital | Addu Equitorial Hospital | D. Fathmath Nazla Rafeeq; Dr. Ibrahim Afzal; Mr. Ibrahim Nishan Ahmed; Ms. Aishath Shuhudha; Ms. Aminath Shazleena Abdul Rahman; Ms. Soafy Mohamed |
| EPI_ISL_8878410 | Broad Institute Clinical Research Sequencing Platform | Infectious Disease Program, Broad Institute of Harvard and MIT | Adams, G.; B.L.; B.W.; Bauer, M.; Birren; Blumenstiel, B.; Brown, C.; Carter, A.; Chaluvadi, S.; D.J.; DeFelice, M.; DeRuff, K.; Dodge, S.; Gabriel, S.; Gallagher, G.; Gladden-Young, A.; Granger, B.; J.E.; K.J.; Lagerborg, K.; Larkin, K.; Lee, M.; Lemieux; Lennon, N.; Loreth, C.; Madoff, L.; McGovern, S.; Meldrim, J.; Normandin, E.; P.C.; Park; Pearlman, L.; Reilly, S.; Rudy, M.; Sabeti; Siddle; Smole, S.; Tomkins-Tinch, C.; Vicente, G.; and MacInnis |
| EPI_ISL_8896430, EPI_ISL_8896433, EPI_ISL_8896435, EPI_ISL_8896444 | CAPRISA | CERI, Centre for Epidemic Response and Innvoation, Stellenbosch University and KRISP, KZN Research Innovation and Sequencing Platform, UKZN. | Aida Sivro; Giandhari J; Moir M; Naidoo Y; Natasha Samsunder; Pillay S; Ramphal U; Ramphal Y; San JE; Tegally H; Tshiabuila D; Van Wyk S; Wilkinson E; de Oliveira T |
| EPI_ISL_8885555, EPI_ISL_8885557, EPI_ISL_8885559, EPI_ISL_8885698, EPI_ISL_8885701, EPI_ISL_8885702, EPI_ISL_8885704, EPI_ISL_8885705, EPI_ISL_8885706, EPI_ISL_8885708, EPI_ISL_8885710, EPI_ISL_8885715, EPI_ISL_8885717, EPI_ISL_8885719, EPI_ISL_8885724, EPI_ISL_8885726, EPI_ISL_8885728, EPI_ISL_8885729, EPI_ISL_8885730, EPI_ISL_8885731, EPI_ISL_8885733, EPI_ISL_8885734, EPI_ISL_8885739, EPI_ISL_8885740, EPI_ISL_8885745, EPI_ISL_8885746, EPI_ISL_8885747, EPI_ISL_8885751, EPI_ISL_8885754, EPI_ISL_8885755, EPI_ISL_8885757, EPI_ISL_8885761, EPI_ISL_8885763, EPI_ISL_8885765, EPI_ISL_8885766, EPI_ISL_8885769, EPI_ISL_8885770, EPI_ISL_8885772, EPI_ISL_8885773, EPI_ISL_8885774, EPI_ISL_8885855, EPI_ISL_8885986, EPI_ISL_8885993, EPI_ISL_8885995, EPI_ISL_8885997, EPI_ISL_8886001, EPI_ISL_8886002, EPI_ISL_8886003, EPI_ISL_8886004, EPI_ISL_8886005, EPI_ISL_8886006, EPI_ISL_8886007, EPI_ISL_8886009, EPI_ISL_8886010, EPI_ISL_8886013, EPI_ISL_8886014, EPI_ISL_8886015, EPI_ISL_8886016, EPI_ISL_8886017, EPI_ISL_8886020, EPI_ISL_8886023, EPI_ISL_8886024, EPI_ISL_8886029, EPI_ISL_8886035, EPI_ISL_8886038, EPI_ISL_8886040, EPI_ISL_8886042, EPI_ISL_8886046, EPI_ISL_8886049, EPI_ISL_8886052, EPI_ISL_8886054, EPI_ISL_8886056, EPI_ISL_8886061, EPI_ISL_8886063, EPI_ISL_8886074, EPI_ISL_8886078, EPI_ISL_8886083, EPI_ISL_8886092, EPI_ISL_8886102, EPI_ISL_8886109, EPI_ISL_8886113, EPI_ISL_8886117, EPI_ISL_8886120, EPI_ISL_8886122, EPI_ISL_8886189, EPI_ISL_8886296, EPI_ISL_8886435, EPI_ISL_8886438, EPI_ISL_8886440, EPI_ISL_8886442, EPI_ISL_8886447, EPI_ISL_8886453, EPI_ISL_8886455, EPI_ISL_8886459, EPI_ISL_8886462, EPI_ISL_8886464, EPI_ISL_8886469, EPI_ISL_8886471, EPI_ISL_8886475, EPI_ISL_8886476, EPI_ISL_8886482, EPI_ISL_8886483, EPI_ISL_8886484, EPI_ISL_8886485, EPI_ISL_8886487, EPI_ISL_8886489, EPI_ISL_8886492, EPI_ISL_8886493, EPI_ISL_8886497, EPI_ISL_8886500, EPI_ISL_8886502, EPI_ISL_8886503, EPI_ISL_8886504, EPI_ISL_8886506, EPI_ISL_8886512, EPI_ISL_8886513, EPI_ISL_8886518, EPI_ISL_8886523, EPI_ISL_8886526, EPI_ISL_8886528, EPI_ISL_8886533, EPI_ISL_8886535, EPI_ISL_8886553, EPI_ISL_8886587, EPI_ISL_8886640, EPI_ISL_8886643, EPI_ISL_8886644, EPI_ISL_8886646, EPI_ISL_8886647, EPI_ISL_8886649, EPI_ISL_8886650, EPI_ISL_8886652, EPI_ISL_8886655, EPI_ISL_8886657, EPI_ISL_8886659, EPI_ISL_8886666, EPI_ISL_8886670, EPI_ISL_8886672, EPI_ISL_8886674, EPI_ISL_8886676, EPI_ISL_8886681, EPI_ISL_8886682, EPI_ISL_8886684, EPI_ISL_8886685, EPI_ISL_8886687, EPI_ISL_8886690, EPI_ISL_8886691, EPI_ISL_8886694, EPI_ISL_8886695, EPI_ISL_8886696, EPI_ISL_8886710, EPI_ISL_8886715, EPI_ISL_8886717, EPI_ISL_8886719, EPI_ISL_8886720, EPI_ISL_8886725, EPI_ISL_8886728, EPI_ISL_8886729, EPI_ISL_8886731, EPI_ISL_8886737, EPI_ISL_8886739, EPI_ISL_8886743, EPI_ISL_8886744, EPI_ISL_8886749, EPI_ISL_8886750, EPI_ISL_8886752, EPI_ISL_8886753, EPI_ISL_8886755, EPI_ISL_8886756, EPI_ISL_8886760, EPI_ISL_8886761, EPI_ISL_8886762, EPI_ISL_8886764, EPI_ISL_8886766, EPI_ISL_8886767, EPI_ISL_8886769, EPI_ISL_8886770, EPI_ISL_8886771, EPI_ISL_8886772, EPI_ISL_8886773, EPI_ISL_8886774, EPI_ISL_8886778, EPI_ISL_8886782, EPI_ISL_8886786, EPI_ISL_8886789, EPI_ISL_8886800, EPI_ISL_8886804, EPI_ISL_8886805, EPI_ISL_8886813, EPI_ISL_8886816, EPI_ISL_8886817, EPI_ISL_8886818, EPI_ISL_8886822, EPI_ISL_8886825, EPI_ISL_8886826, EPI_ISL_8886831, EPI_ISL_8886832, EPI_ISL_8886846, EPI_ISL_8886847, EPI_ISL_8886848, EPI_ISL_8886851, EPI_ISL_8886856, EPI_ISL_8886858, EPI_ISL_8886860, EPI_ISL_8886862, EPI_ISL_8886864, EPI_ISL_8886865, EPI_ISL_8886866, EPI_ISL_8886868, EPI_ISL_8886870, EPI_ISL_8886873, EPI_ISL_8886931, EPI_ISL_8886932, EPI_ISL_8886933, EPI_ISL_8886934, EPI_ISL_8886935, EPI_ISL_8886936, EPI_ISL_8886937, EPI_ISL_8886938, EPI_ISL_8886939, EPI_ISL_8886941, EPI_ISL_8886944, EPI_ISL_8886945, EPI_ISL_8886947, EPI_ISL_8886952, EPI_ISL_8886953, EPI_ISL_8886954, EPI_ISL_8886955, EPI_ISL_8886956, EPI_ISL_8886957, EPI_ISL_8886958, EPI_ISL_8886959, EPI_ISL_8886960, EPI_ISL_8886963, EPI_ISL_8886965, EPI_ISL_8886968, EPI_ISL_8886970, EPI_ISL_8886971, EPI_ISL_8886972, EPI_ISL_8886973, EPI_ISL_8886975, EPI_ISL_8886976, EPI_ISL_8886979, EPI_ISL_8886981, EPI_ISL_8886984, EPI_ISL_8886986, EPI_ISL_8886988, EPI_ISL_8886989, EPI_ISL_8886998, EPI_ISL_8886999, EPI_ISL_8887000, EPI_ISL_8887001, EPI_ISL_8887002, EPI_ISL_8887005, EPI_ISL_8887008, EPI_ISL_8887009, EPI_ISL_8887013, EPI_ISL_8887015, EPI_ISL_8887016, EPI_ISL_8887018, EPI_ISL_8887020, EPI_ISL_8887021, EPI_ISL_8887024, EPI_ISL_8887025, EPI_ISL_8887033, EPI_ISL_8887035, EPI_ISL_8887036, EPI_ISL_8887038, EPI_ISL_8887040, EPI_ISL_8887045, EPI_ISL_8887046, EPI_ISL_8887048, EPI_ISL_8887050, EPI_ISL_8887052, EPI_ISL_8887053, EPI_ISL_8887056, EPI_ISL_8887058, EPI_ISL_8887071, EPI_ISL_8887073, EPI_ISL_8887074, EPI_ISL_8887075, EPI_ISL_8887076, EPI_ISL_8887079, EPI_ISL_8887082, EPI_ISL_8887085, EPI_ISL_8887086, EPI_ISL_8887088, EPI_ISL_8887092, EPI_ISL_8887093, EPI_ISL_8887097, EPI_ISL_8887098, EPI_ISL_8887099, EPI_ISL_8887100, EPI_ISL_8887102, EPI_ISL_8887108, EPI_ISL_8887111, EPI_ISL_8887114, EPI_ISL_8887115, EPI_ISL_8887119, EPI_ISL_8887124, EPI_ISL_8887131, EPI_ISL_8887133, EPI_ISL_8887135, EPI_ISL_8887136, EPI_ISL_8887138, EPI_ISL_8887139, EPI_ISL_8887141, EPI_ISL_8887145, EPI_ISL_8887147, EPI_ISL_8887153, EPI_ISL_8887155, EPI_ISL_8887158, EPI_ISL_8887160, EPI_ISL_8887163, EPI_ISL_8887164, EPI_ISL_8887174, EPI_ISL_8887176, EPI_ISL_8887178, EPI_ISL_8887184, EPI_ISL_8887187, EPI_ISL_8887188, EPI_ISL_8887191, EPI_ISL_8887193, EPI_ISL_8887196, EPI_ISL_8887202, EPI_ISL_8887208, EPI_ISL_8887213, EPI_ISL_8887218, EPI_ISL_8887219, EPI_ISL_8887223, EPI_ISL_8887225, EPI_ISL_8887226, EPI_ISL_8887231, EPI_ISL_8887232, EPI_ISL_8887236, EPI_ISL_8887237, EPI_ISL_8887239, EPI_ISL_8887240, EPI_ISL_8887243, EPI_ISL_8887245, EPI_ISL_8887249, EPI_ISL_8887256, EPI_ISL_8887258, EPI_ISL_8887260, EPI_ISL_8887262, EPI_ISL_8887263, EPI_ISL_8887264, EPI_ISL_8887265, EPI_ISL_8887266, EPI_ISL_8887270, EPI_ISL_8887272, EPI_ISL_8887275, EPI_ISL_8887278, EPI_ISL_8887281, EPI_ISL_8887284, EPI_ISL_8887289, EPI_ISL_8887290, EPI_ISL_8887291, EPI_ISL_8887292, EPI_ISL_8887296, EPI_ISL_8887300, EPI_ISL_8887302, EPI_ISL_8887303, EPI_ISL_8887307, EPI_ISL_8887394, EPI_ISL_8887395, EPI_ISL_8887397, EPI_ISL_8887398, EPI_ISL_8887399, EPI_ISL_8887400, EPI_ISL_8887405, EPI_ISL_8887406, EPI_ISL_8887408, EPI_ISL_8887409, EPI_ISL_8887411, EPI_ISL_8887413, EPI_ISL_8887415, EPI_ISL_8887416, EPI_ISL_8887418, EPI_ISL_8887433, EPI_ISL_8887435, EPI_ISL_8887438, EPI_ISL_8887441, EPI_ISL_8887445, EPI_ISL_8887450, EPI_ISL_8887452, EPI_ISL_8887453, EPI_ISL_8887456, EPI_ISL_8887457, EPI_ISL_8887458, EPI_ISL_8887462, EPI_ISL_8887466, EPI_ISL_8887467, EPI_ISL_8887469, EPI_ISL_8887471, EPI_ISL_8887475, EPI_ISL_8887476, EPI_ISL_8887478, EPI_ISL_8887479, EPI_ISL_8887495, EPI_ISL_8887497, EPI_ISL_8887500, EPI_ISL_8887505, EPI_ISL_8887511, EPI_ISL_8887515, EPI_ISL_8887516, EPI_ISL_8887518, EPI_ISL_8887521, EPI_ISL_8887522, EPI_ISL_8887524, EPI_ISL_8887527, EPI_ISL_8887530, EPI_ISL_8887533, EPI_ISL_8887534, EPI_ISL_8887535, EPI_ISL_8887536, EPI_ISL_8887538, EPI_ISL_8887541, EPI_ISL_8887542, EPI_ISL_8887545, EPI_ISL_8887548, EPI_ISL_8887550, EPI_ISL_8887554, EPI_ISL_8887556, EPI_ISL_8887559, EPI_ISL_8887561, EPI_ISL_8887564, EPI_ISL_8887566, EPI_ISL_8887574, EPI_ISL_8887578, EPI_ISL_8887580, EPI_ISL_8887583, EPI_ISL_8887584, EPI_ISL_8887585, EPI_ISL_8887586, EPI_ISL_8887589, EPI_ISL_8887779, EPI_ISL_8887805, EPI_ISL_8887806, EPI_ISL_8887807, EPI_ISL_8887811, EPI_ISL_8887814, EPI_ISL_8887818, EPI_ISL_8887822, EPI_ISL_8887829, EPI_ISL_8887833, EPI_ISL_8887880, EPI_ISL_8887881, EPI_ISL_8887887, EPI_ISL_8887889, EPI_ISL_8887892 | | | |
| see above | Department of Bacteria, Parasites and Fungi, Statens Serum Institut, Copenhagen, Denmark | Statens Serum Institut Bioinformatics and Microbial Genomics | Danish Covid-19 Genome Consortium |
| EPI_ISL_8885401 | Gujarat Pathology laboratory & Diagnostic Center, Paldi, Ahmedabad | Gujarat Biotechnology Research Centre | Akhilesh Modi; Apurvasinh Puvar; Bhadreshsinh Gohil; Chaitanya Joshi; Disha Vora; Janvi Raval; Jaykumar Rangani; Madhvi Joshi; Nimesh Patel; Niraj Kotha; Nitin Savaliya; NitinShukla; Priyank Chavda; Ramesh Pandit; Roshani Mishra; Sonal Sharma; Tasnim Trivedi; Zarna Patel |
| EPI_ISL_8886546, EPI_ISL_8886554, EPI_ISL_8886555, EPI_ISL_8886563, EPI_ISL_8886564, EPI_ISL_8886575, EPI_ISL_8886576, EPI_ISL_8886586, EPI_ISL_8886594, EPI_ISL_8886595, EPI_ISL_8886596, EPI_ISL_8886600 | | | |
| see above | Indira Gandhi Memorial Hospital | Indira Gandhi Memorial Hospital | D. Fathmath Nazla Rafeeq; Dr. Ibrahim Afzal; Mr. Ibrahim Nishan Ahmed; Ms. Aishath Shuhudha; Ms. Aminath Shazleena Abdul Rahman; Ms. Soafy Mohamed |
| EPI_ISL_8897015 | KKP Kelas 1 Soekarno-Hatta (iLab) | National Institute of Health Research and Development | Arie Ardiansyah Nugraha; Fajar Nur Sulistiyohadi; Hana Apsari Pawestri; Hartanti Dian Ikawati; Kartika Dewi Puspa; Nelly Puspandari; Nur Ika Hariastuti; Putri Widia; Subangkit; Vivi Setiawaty |
| EPI_ISL_8886623, EPI_ISL_8886624 | Kudahuvadhoo Atoll Hospital | Kudahuvadhoo Atoll Hospital | D. Fathmath Nazla Rafeeq; Dr. Ibrahim Afzal; Mr. Ibrahim Nishan Ahmed; Ms. Aishath Shuhudha; Ms. Aminath Shazleena Abdul Rahman; Ms. Soafy Mohamed |
| EPI_ISL_8886609, EPI_ISL_8886610 | Kulhudufushi Regional Hospital | Kulhudufushi Regional Hospital | D. Fathmath Nazla Rafeeq; Dr. Ibrahim Afzal; Mr. Ibrahim Nishan Ahmed; Ms. Aishath Shuhudha; Ms. Aminath Shazleena Abdul Rahman; Ms. Soafy Mohamed |
| EPI_ISL_8892176 | Michigan Department of Health and Human Services, Bureau of Laboratories | Michigan Department of Health and Human Services, Bureau of Laboratories | Blankenship HM; Riner D; Soehnlen MK |
| EPI_ISL_8896467, EPI_ISL_8896476, EPI_ISL_8896477 | National Health Laboratory Service, Kwazulu-Natal, South Africa | CERI, Centre for Epidemic Response and Innvoation, Stellenbosch University and KRISP, KZN Research Innovation and Sequencing Platform, UKZN. | Giandhari J; Moir M; Naidoo Y; Nokukhanya Mdlalose; Pillay S; Ramphal U; Ramphal Y; San JE; Tegally H; Tshiabuila D; Van Wyk S; Wilkinson E; de Oliveira T |
| EPI_ISL_8890649, EPI_ISL_8890650, EPI_ISL_8890651, EPI_ISL_8890652, EPI_ISL_8890653, EPI_ISL_8890654, EPI_ISL_8890655, EPI_ISL_8890656, EPI_ISL_8890657, EPI_ISL_8890658, EPI_ISL_8890659, EPI_ISL_8890675 | | | |
| see above | National Microbiology Reference Laboratory, Ministry of Health, Harare, Zimbabwe | CERI, Centre for Epidemic Response and Innvoation, Stellenbosch University and KRISP, KZN Research Innovation and Sequencing Platform, UKZN. | Agnes Juru; Air Comodor Dr J. Chimedza; Charles Nyagupe; Dr Raiva Simbi; Giandhari J; Hlanai Gumbo; Kenneth Maeka; Moir M; Naidoo Y; Pillay S; Ramphal U; Ramphal Y; San JE; Tapfumanei Mashe; Tatenda Takawira; Tegally H; Tshiabuila D; Van Wyk S; Wilkinson E; de Oliveira T |
| EPI_ISL_8879944, EPI_ISL_8879948 | Nevada State Public Health Laboratory | Nevada State Public Health Laboratory | Andrew Gorzalski; Lynette Gumbleton; Mark Pandori |
| EPI_ISL_8900901, EPI_ISL_8901099 | Pathology West - NSW Health Pathology | NSW Health Pathology - Institute of Clinical Pathology and Medical Research; Westmead Hospital; University of Sydney | Arnott A.; Draper J.; Gall M.; Martinez E.; Rockett R.; Sintchenko V.; on behalf of ICPMR |
| EPI_ISL_8884476, EPI_ISL_8884549 | Respiratory Virus Unit, Microbiology Services Colindale, Public Health England | COVID-19 Genomics UK (COG-UK) Consortium | PHE Covid Sequencing Team |
| EPI_ISL_8884441 | Viral Research and Diagnostic Laboratories | NIV Influenza | Dr.Rupinder Bakshi |

We gratefully acknowledge the following Authors from the Originating laboratories responsible for obtaining the specimens, as well as the Submitting laboratories where the genome data were generated and shared via GISAID, on which this research is based.

All Submitters of data may be contacted directly via www.gisaid.org

Authors are sorted alphabetically.

| Accession ID | Originating Laboratory | Submitting Laboratory | Authors |
|---|---|---|---|
| EPI_ISL_8902791, EPI_ISL_8902818, EPI_ISL_8902819 | Arkansas Public Health Laboratory, Arkansas Department of Health | University of Minnesota Genomics Center | Corbin Dirkx; Daryl M. Gohl; Jaquelyn Kuriger-Laber; John Garbe |
| EPI_ISL_8903524, EPI_ISL_8903528, EPI_ISL_8903529 | Jessa | Jessa | Severine Berden et al. on behalf of the Jessa_cmdLab |
| EPI_ISL_8903174, EPI_ISL_8903217, EPI_ISL_8903251, EPI_ISL_8903317, EPI_ISL_8903373, EPI_ISL_8903423 | National Virus Reference Laboratory | National Virus Reference Laboratory | Charlene Bennett; Cillian F De Gascun; Gabriel Gonzalez; Jonathan Dean; Michael Carr; Zoe Yandle |
| EPI_ISL_8901099, EPI_ISL_8901110 | Pathology West - NSW Health Pathology | NSW Health Pathology - Institute of Clinical Pathology and Medical Research; Westmead Hospital; University of Sydney | Arnott A.; Draper J.; Gall M.; Martinez E.; Rockett R.; Sintchenko V.; on behalf of ICPMR |
| EPI_ISL_8907628, EPI_ISL_8909082 | UW Virology Lab | UW Virology Lab | Alexander Greninger; Hong Xie; Isabel Arnould; Keith R Jerome; Meei-Li Huang; Nathan Breit; Patrick Mathias; Pavitra Roychoudhury; Pooneh Hajian; Ricardo Perez; Robert J. Livingston; Sean Ellis; Seffir T. Wendm; Shah Mohamed Bakhash |

We gratefully acknowledge the following Authors from the Originating laboratories responsible for obtaining the specimens, as well as the Submitting laboratories where the genome data were generated and shared via GISAID, on which this research is based.

All Submitters of data may be contacted directly via www.gisaid.org

Authors are sorted alphabetically.

| Accession ID | Originating Laboratory | Submitting Laboratory | Authors |
|---|---|---|---|
| EPI_ISL_9002785, EPI_ISL_9002788 | Botswana Harvard HIV Reference Laboratory | Botswana Harvard HIV Reference Laboratory | Boitumelo Zuze; Botshelo Radibe; Dorcas Maruapula; Joseph Makhema; Keoratile Ntshambiwa; Kgomotso Moruisi; Legodile Kooepile; Mosepele Mosepele; Mphaphi B. Mbulawa; Ontlametse T. Bareng; Pamela Smith-Lawrence; Roger Shapiro; Sefetogi Ramaologa; Shahin Lockman; Sikhulile Moyo; Simani Gaseitsiwe; Thongbotho Mphoyakgosi; Wonderful T. Choga |
| EPI_ISL_9003447 | CHEDV | Instituto Nacional de Saude (INSA) | Borges et al |
| EPI_ISL_9003416 | CHTMAD | Instituto Nacional de Saude (INSA) | Borges et al |
| EPI_ISL_8986579, EPI_ISL_8986650 | Clinical Molecular Microbiology Laboratory, UNC Hospitals | Jeremy Wang | Bravada Hill; Corbin Jones; Jeremy Wang; Katie Martin; Melissa Miller; Sayal Guirales-Medrano; Tara Skelly |
| EPI_ISL_9003527, EPI_ISL_9003556, EPI_ISL_9003563, EPI_ISL_9003581, EPI_ISL_9003583, EPI_ISL_9003594, EPI_ISL_9003597 | | | |
| see above | Colorado Department of Public Health and Environment | Colorado Department of Public Health and Environment | Alexandria Rossheim; Arianna Smith; Diana Ir; Emily A. Travanty; Laura Bankers; Mandy Waters; Michael Martin; Molly C. Hetherington-Rauth; Shannon R. Matzinger |
| EPI_ISL_8976723, EPI_ISL_8976724, EPI_ISL_8976725, EPI_ISL_8976726, EPI_ISL_8976728, EPI_ISL_8976729, EPI_ISL_8976730, EPI_ISL_8976731, EPI_ISL_8976732, EPI_ISL_8976733, EPI_ISL_8976734, EPI_ISL_8976735, EPI_ISL_8976736, EPI_ISL_8976737 | | | |
| see above | Curative Labs | Curative Labs | Elias L. Salfati; Eugenia Khorosheva; George Way; J.Cesar Ignacio-Espinoza; Janet Chen; Mikhail Hanewich-Hollatz; Nabjot Sandhu; Sophia Quasem; Vladimir Slepnev; Zhiyi Xie |
| EPI_ISL_8995768, EPI_ISL_8995769 | DGHP Laboratory | Thailand MOPH - U.S. CDC Collaboration (TUC) | Beth Skaggs; Piroon Jenjaroenpun; Pongpun Sawatwong; Thidathip Wongsurawat |
| EPI_ISL_8985651 | Diagnostyka. Laboratoria Medyczne. | 1. ViroGenetics - BSL3 Laboratory of Virology, Małopolska Centre of Biotechnology, Jagiellonian University; 2. Diagtron Laboratoria Łukasz Rąbalski | Gromowski, T.; Kowalski, M.; Labaj; Maciej Kosinski; Mazur-Panasiuk, N.; P.P.; Pyrc, K.; Rabalski L.; Rogalska-Kupiec M.; Swadzba J.; Sylwia Januszczak; Szulc, P.; Wydmanski, W. |
| EPI_ISL_8998400, EPI_ISL_8998401, EPI_ISL_8998402, EPI_ISL_8998403, EPI_ISL_8998404, EPI_ISL_8998406 | Division of Emerging Infectious Diseases, Bureau of Infectious Diseases Diagnosis Control, Korea Disease Control and Prevention Agency | Division of Emerging Infectious Diseases, Bureau of Infectious Diseases Diagnosis Control, Korea Disease Control and Prevention Agency | Ae Kyung Park; Chae Young Lee; Eun-Jin Kim; Hyuck Jin Lee; Il-Hwan Kim; Jeong-Ah Kim |
| EPI_ISL_8989282 | Fakultní nemocnice u sv. Anny v Brně | Fakultní nemocnice u sv. Anny v Brně | Lenka Zamecnikova; Milada Dvořáčková. Jan Vrbsky |
| EPI_ISL_8955208 | Fondazione IRCCS Ca' Granda Ospedale Maggiore Policlinico | Fondazione IRCCS Ca' Granda Ospedale Maggiore Policlinico | Ferruccio Ceriotti; Sara Uceda Renteria |
| EPI_ISL_8995757, EPI_ISL_8995763, EPI_ISL_8995764, EPI_ISL_8995766, EPI_ISL_8995767 | Health Department, Bangkok Metropolitan Administration | Thailand MOPH - U.S. CDC Collaboration (TUC) | Beth Skaggs; Piroon Jenjaroenpun; Pongpun Sawatwong; Thidathip Wongsurawat |
| EPI_ISL_8995642, EPI_ISL_8995643, EPI_ISL_8995644 | Mae Sot General Hospital | Thailand MOPH - U.S. CDC Collaboration (TUC) | Beth Skaggs; Piroon Jenjaroenpun; Pongpun Sawatwong; Thidathip Wongsurawat |
| EPI_ISL_8982022, EPI_ISL_8983039 | Molecular Virology section, department of infection & immunity, KFSHRC | Molecular Virology section, department of infection & immunity, KFSHRC | Ahmed A. Al-Qahtani; Basma M. Alahideb; Fatimah S. Alhamlan; Feda A. Alsuwairi; Maha Al-Abdulkarim; Maysoon S. Mutabagani; Mohamed S. Asiri; Reem S. Almaghrabi Madain S. Alsanea; Sahar I. Althawadi; Sara A. Altamimi |
| EPI_ISL_9002773, EPI_ISL_9002790, EPI_ISL_9002795, EPI_ISL_9002810, EPI_ISL_9002814, EPI_ISL_9002815, EPI_ISL_9002822, EPI_ISL_9002836, EPI_ISL_9002859 | | | |
| see above | National Health Laboratory | Botswana Harvard HIV Reference Laboratory | Boitumelo Zuze; Botshelo Radibe; Dorcas Maruapula; Joseph Makhema; Keoratile Ntshambiwa; Kgomotso Moruisi; Legodile Kooepile; Mosepele Mosepele; Mphaphi B. Mbulawa; Ontlametse T. Bareng; Pamela Smith-Lawrence; Roger Shapiro; Sefetogi Ramaologa; Shahin Lockman; Sikhulile Moyo; Simani Gaseitsiwe; Thongbotho Mphoyakgosi; Wonderful T. Choga |
| EPI_ISL_8958708 | Northumbria University / South Tees Hospitals NHS Foundation Trust / North Cumbria Integrated Care NHS Foundation Trust / North Tees and Hartlepool NHS Foundation Trust / Newcastle Hospitals NHS Foundation Trust | COVID-19 Genomics UK (COG-UK) Consortium | Andrew Nelson; Brendan Payne; Clive Graham; Darren L Smith; Debra Padgett; Edward Barton; Emma Swindells; Garren Scott; Gary Black; Gary Eltringham; Giles S Holt; Greg R Young; Jane Greenaway; Jennifer Collins; John Allan; Joshua Loh; Lynn Dover; Matthew Bashton; Mohammad A Tariq; Paul Baker; Sarah Essex; Steve Liggett; Wen C Yew; Yusri Taha |
| EPI_ISL_8977384, EPI_ISL_8977385, EPI_ISL_8977386, EPI_ISL_8977387, EPI_ISL_8977388, EPI_ISL_8977392, EPI_ISL_8977394, EPI_ISL_8977397, EPI_ISL_8977400, EPI_ISL_8977401 | | | |
| see above | Ochsner Health | BioInfoExperts | Amy Feehan; Ben Lain; Chris Huston; David J. Nolan; Judy Crabtree; Julia-Garcia-Diaz; Lucio Miele; Rebecca Rose; Samual Moot; Susanna L. Lamers; Tessa LaFleur |
| EPI_ISL_8960507, EPI_ISL_8960509, EPI_ISL_8961047 | Originating lab: Wales Specialist Virology Centre Sequencing lab: Pathogen Genomics Unit | Public Health Wales Microbiology Cardiff Wales Specialist Virology Centre | Alec Birchley; Alexander Adams; Amy Gaskin; Angela Marchbank; Bree Gatica-Wilcox; Catherine Moore; Jason Coombes; Joanne Watkins; Joel Southgate; Johnathan Evans; Laura Gifford; Lauren Gilbert; Lee Graham; Malorie Perry; Matthew Bull; Nicole Pacchiarini; Sally Corden; Sara Kumziene-Summerhayes; Sara Rey; Sarah Taylor; Simon Cottrell; Sophie Jones; Tom Connor |
| EPI_ISL_8959296, EPI_ISL_8959335, EPI_ISL_8959727 | Respiratory Virus Unit, Microbiology Services Colindale, Public Health England | COVID-19 Genomics UK (COG-UK) Consortium | PHE Covid Sequencing Team |
| EPI_ISL_9003415 | SEEBMO | Instituto Nacional de Saude (INSA) | Borges et al |
| EPI_ISL_8993539, EPI_ISL_8993543 | TSGH-CP molecular lab | TSGH-CP molecular lab | Cherng-Lih Perng; Chien-Wen Chen; Chih-Kai Chang; Feng-Yee Chang; Hsing-Yi Chung; Hung-Sheng Shang; Jung-Chung Lin; Kuo-Ming Yeh; Kuo-Sheng Hung; Ming-Jr JIAN; Sheng-Kang Chiu; Shih-Hung Tsai; Tien-Yao Chang |
| EPI_ISL_8986746 | Tokyo Metropolitan Institute of Public Health | Tokyo Metropolitan Institute of Public Health | Akane Negishi; Arisa Amano; Hirofumi Miyake; Jun Suzuki; Kenji Sadamasu; Kenshirou Kuroki; Mami Nagashima; Masaki Hayashi; Maya Isogai; Takako Yamazaki; Takushi Fujiwara |
| EPI_ISL_8988612, EPI_ISL_8988613, EPI_ISL_8988614, EPI_ISL_8988615, EPI_ISL_8988617, EPI_ISL_8988618, EPI_ISL_8988619, EPI_ISL_8988620, EPI_ISL_8988621, EPI_ISL_8988622, EPI_ISL_8988623, EPI_ISL_8988624, EPI_ISL_8988626 | | | |
| see above | UAN-LANIIA | Microbial Genomics Laboratory | ; Alejandra García-Gasca; Alejandra Hernández-Terán; Alejandro Sánchez-Flores; Alfredo Herrera-Estrella; Alicia Ocaña-Mondragón; Andreu Comas-García; Angel Gustavo Salas-Lais; Antonio Loza Román; Bernardo Martínez-Miguel; Blanca Taboada; Brenda Irasema Maldonado-Meza; Bruno Gómez-Gil; Carla Ivón Herrera-Najera; Carlos F. Arias; Celia Boukadida; Clara Esperanza Santacruz-Tinoco; Concepción Grajales-Muñiz; Consorcio Mexicano de Vigilancia Genómica (CoViGen-Mex). Authors (in alphabetical order): Julio Elias Alvarado-Yaah; Cristóbal Cháidez-Quiróz; Célida Duque Molina; Célida Martínez-Rodríguez; Daniel Fregoso-Rueda; Daniel Lira Morales; Eduardo Becerril-Vargas; Fernando Fontove-Herrera; Fidencio Mejía-Nepomuceno; Francisco Pulido; Gloria Elena Espinosa-Ayala; Gloria María Molina-Salinas; Gloria Vazquez; Hector Esteban Paz-Juárez; Hector Montoya-Fuentes; Helen Haydee Fernanda Ramírez-Plascencia; Irvin González-López; Jean Pierre González; Jesús Hernández; Joel Armando Vázquez-Pérez.; Jorge Salas-Hernández; José Antonio Enciso-Moreno; José Arturo Martínez-Orozco; José Esteban Muñoz-Medina; José de Jesús Nuñez-Contreras; Juan Bautista Chale-Dzul; Julissa Enciso-Ibarra; Luis Alberto Ochoa-Carrera; Manuel Iván Girón Pérez; Margarita Matías-Florentino; Mario Mújica-Sánchez; Marissa Perez-Garcia; María Guadalupe Santiago-Mauricio; María Guadalupe de Jesús Mireles-Rivera; Nelly Sélem-Mojica; Pavel Isa; Ricardo Ciria Merce; Ricardo Grande; Rosa María Gutiérrez Rios; Santiago Ávila-Ríos; Selene Zárate; Susana Lopez; Verónica Mata-Haro; Victor Eduardo García-Arias; Víctor Hugo Borja-Aburto |
| EPI_ISL_9003407 | ULS Castelo Branco | Instituto Nacional de Saude (INSA) | Borges et al |
| EPI_ISL_9003417 | UNILABS | Instituto Nacional de Saude (INSA) | Borges et al |
| EPI_ISL_8976191 | US Air Force School of Aerospace Medicine | US Air Force School of Aerospace Medicine | Anthony Fries; Carol Garrett; Clarise Starr; Deanna Muehleman; Elizabeth Macias; Jennifer Meyer; Kelsey Lanter; Sarah Purves; William Gruner |
| EPI_ISL_8986063, EPI_ISL_8986109, EPI_ISL_8986203, EPI_ISL_8986254, EPI_ISL_8986397, EPI_ISL_8986481, EPI_ISL_8986500 | | | |
| see above | UW Virology Lab | UW Virology Lab | Alexander Greninger; Hong Xie; Isabel Arnould; Keith R Jerome; Meei-Li Huang; Nathan Breit; Patrick Mathias; Pavitra Roychoudhury; Pooneh Hajian; Ricardo Perez; Robert J. Livingston; Sean Ellis; Seffir T. Wendm; Shah Mohamed Bakhash |
| EPI_ISL_8981633 | Virology, Universitätsklinikum des Saarlandes | Epigenetics, Saarland University | Barbara Berko-Göttel Sascha Tierling; Jörn Walter; Kathrin Kattler; Nastasia Seiwert; Sigrun Smola; Thorsten Pfuhl |
| EPI_ISL_8975529 | Zakład Diagnostyki Laboratoryjnej Mazowiecki Szpital Bródnowski Sp. z | 1. Academic Center for Pathomorphological and Genetic- | Anetta Sulewska; Jacek Nikliński; Janusz Dzięcioł; Joanna Kiśluk; Katarzyna Zacharczuk; Konrad Raczkowski; Magdalena Nowakowska; Małgorzata Sadkowska-Todys; Piotr Karabowicz; Piotr Majewski; Przemysław Biecek. Joanna Reszeć; Radosław Charkiewicz; Tomasz Wołkowicz |

| | |
|---|---|
| o.o. ul. Kondratowicza 8, o3-242 Warszawa | Molecular Diagnostics ltd, Bialystok, Poland 2. National Institute of Public Health - National Institute of Hygiene, Warsaw, Poland |

We gratefully acknowledge the following Authors from the Originating laboratories responsible for obtaining the specimens, as well as the Submitting laboratories where the genome data were generated and shared via GISAID, on which this research is based.

All Submitters of data may be contacted directly via www.gisaid.org

Authors are sorted alphabetically.

Acknowledgement EPI_SET Identifier: EPI_SET_20220527bd

| Accession ID | Originating Laboratory | Submitting Laboratory | Authors |
|---|---|---|---|
| EPI_ISL_8921333, EPI_ISL_8921336, EPI_ISL_8921337, EPI_ISL_8921342, EPI_ISL_8921344, EPI_ISL_8921346, EPI_ISL_8921350, EPI_ISL_8921358, EPI_ISL_8921363, EPI_ISL_8921368, EPI_ISL_8921377, EPI_ISL_8921387, EPI_ISL_8921388, EPI_ISL_8921390, EPI_ISL_8921392, EPI_ISL_8921396, EPI_ISL_8921399, EPI_ISL_8921400, EPI_ISL_8921402, EPI_ISL_8921406, EPI_ISL_8921412, EPI_ISL_8921426, EPI_ISL_8921430, EPI_ISL_8921431, EPI_ISL_8921438, EPI_ISL_8921442, EPI_ISL_8921445, EPI_ISL_8921449, EPI_ISL_8921451, EPI_ISL_8921453, EPI_ISL_8921455, EPI_ISL_8921462, EPI_ISL_8921465, EPI_ISL_8921466, EPI_ISL_8921467, EPI_ISL_8921472, EPI_ISL_8921477, EPI_ISL_8921486, EPI_ISL_8921490, EPI_ISL_8921492, EPI_ISL_8921494, EPI_ISL_8921496, EPI_ISL_8921498, EPI_ISL_8921505, EPI_ISL_8921509, EPI_ISL_8921515, EPI_ISL_8921517, EPI_ISL_8921519, EPI_ISL_8921521, EPI_ISL_8921527, EPI_ISL_8921534, EPI_ISL_8921536, EPI_ISL_8921539, EPI_ISL_8921543, EPI_ISL_8921551, EPI_ISL_8921557, EPI_ISL_8921560, EPI_ISL_8921564, EPI_ISL_8921570, EPI_ISL_8921577, EPI_ISL_8921579, EPI_ISL_8921585, EPI_ISL_8921594, EPI_ISL_8921599, EPI_ISL_8921601, EPI_ISL_8921605, EPI_ISL_8921607, EPI_ISL_8921611, EPI_ISL_8921613, EPI_ISL_8921620, EPI_ISL_8921624, EPI_ISL_8921628, EPI_ISL_8921630, EPI_ISL_8921634, EPI_ISL_8921638, EPI_ISL_8921645, EPI_ISL_8921648, EPI_ISL_8921650, EPI_ISL_8921655, EPI_ISL_8921657, EPI_ISL_8921662, EPI_ISL_8921666, EPI_ISL_8921674, EPI_ISL_8921678, EPI_ISL_8921680, EPI_ISL_8921682, EPI_ISL_8921692, EPI_ISL_8921696, EPI_ISL_8921698, EPI_ISL_8921699, EPI_ISL_8921701, EPI_ISL_8921705, EPI_ISL_8921713, EPI_ISL_8921727, EPI_ISL_8921730, EPI_ISL_8921736, EPI_ISL_8921738, EPI_ISL_8921740, EPI_ISL_8921747, EPI_ISL_8921749, EPI_ISL_8921757, EPI_ISL_8921762, EPI_ISL_8921764, EPI_ISL_8921770, EPI_ISL_8921772, EPI_ISL_8921777, EPI_ISL_8921781, EPI_ISL_8921783, EPI_ISL_8921787, EPI_ISL_8921791, EPI_ISL_8921798, EPI_ISL_8921800, EPI_ISL_8921802, EPI_ISL_8921820, EPI_ISL_8921821, EPI_ISL_8921823, EPI_ISL_8921825, EPI_ISL_8921829, EPI_ISL_8921831, EPI_ISL_8921834, EPI_ISL_8921836, EPI_ISL_8921839, EPI_ISL_8921844, EPI_ISL_8921849, EPI_ISL_8921863, EPI_ISL_8921878, EPI_ISL_8921880, EPI_ISL_8921884, EPI_ISL_8921885, EPI_ISL_8921887, EPI_ISL_8921889, EPI_ISL_8921891, EPI_ISL_8921893, EPI_ISL_8921895, EPI_ISL_8921901, EPI_ISL_8921907, EPI_ISL_8921909, EPI_ISL_8921913, EPI_ISL_8921916, EPI_ISL_8921918, EPI_ISL_8921920, EPI_ISL_8921922, EPI_ISL_8921923, EPI_ISL_8921925, EPI_ISL_8921927, EPI_ISL_8921939, EPI_ISL_8921941, EPI_ISL_8921949, EPI_ISL_8921950, EPI_ISL_8921954, EPI_ISL_8921957, EPI_ISL_8921958, EPI_ISL_8921960, EPI_ISL_8921964, EPI_ISL_8921966, EPI_ISL_8921970, EPI_ISL_8921974, EPI_ISL_8921981, EPI_ISL_8921988, EPI_ISL_8921990, EPI_ISL_8921992, EPI_ISL_8922001, EPI_ISL_8922011, EPI_ISL_8922015, EPI_ISL_8922017, EPI_ISL_8922027, EPI_ISL_8922030, EPI_ISL_8922032, EPI_ISL_8922034, EPI_ISL_8922036, EPI_ISL_8922038, EPI_ISL_8922043, EPI_ISL_8922047, EPI_ISL_8922049, EPI_ISL_8922051, EPI_ISL_8922059, EPI_ISL_8922060, EPI_ISL_8922064, EPI_ISL_8922070, EPI_ISL_8922076, EPI_ISL_8922087, EPI_ISL_8922089, EPI_ISL_8922091, EPI_ISL_8922093, EPI_ISL_8922096, EPI_ISL_8922098, EPI_ISL_8922102, EPI_ISL_8922104, EPI_ISL_8922106, EPI_ISL_8922113, EPI_ISL_8922115, EPI_ISL_8922117, EPI_ISL_8922121, EPI_ISL_8922123, EPI_ISL_8922126, EPI_ISL_8922130, EPI_ISL_8922134, EPI_ISL_8922143, EPI_ISL_8922147, EPI_ISL_8922149, EPI_ISL_8922151, EPI_ISL_8922156, EPI_ISL_8922170, EPI_ISL_8922177, EPI_ISL_8922182, EPI_ISL_8922185, EPI_ISL_8922190, EPI_ISL_8922195, EPI_ISL_8922199, EPI_ISL_8922202, EPI_ISL_8922206, EPI_ISL_8922221, EPI_ISL_8922223, EPI_ISL_8922225, EPI_ISL_8922233, EPI_ISL_8922237, EPI_ISL_8922239, EPI_ISL_8922243, EPI_ISL_8922245, EPI_ISL_8922247, EPI_ISL_8922250, EPI_ISL_8922254, EPI_ISL_8922258, EPI_ISL_8922265, EPI_ISL_8922267, EPI_ISL_8922269, EPI_ISL_8922286, EPI_ISL_8922290, EPI_ISL_8922292, EPI_ISL_8922302, EPI_ISL_8922304, EPI_ISL_8922314, EPI_ISL_8922316, EPI_ISL_8922318, EPI_ISL_8922319, EPI_ISL_8922321, EPI_ISL_8922327, EPI_ISL_8922329, EPI_ISL_8922331, EPI_ISL_8922336, EPI_ISL_8922338, EPI_ISL_8922344, EPI_ISL_8922350, EPI_ISL_8922352, EPI_ISL_8922354, EPI_ISL_8922356, EPI_ISL_8922357, EPI_ISL_8922369, EPI_ISL_8922372, EPI_ISL_8922377, EPI_ISL_8922379, EPI_ISL_8922384, EPI_ISL_8922387, EPI_ISL_8922389, EPI_ISL_8922391, EPI_ISL_8922396, EPI_ISL_8922397, EPI_ISL_8922399, EPI_ISL_8922403, EPI_ISL_8922405, EPI_ISL_8922412, EPI_ISL_8922414, EPI_ISL_8922418, EPI_ISL_8922419, EPI_ISL_8922421, EPI_ISL_8922423, EPI_ISL_8922435, EPI_ISL_8922439, EPI_ISL_8922445, EPI_ISL_8922447, EPI_ISL_8922450, EPI_ISL_8922458, EPI_ISL_8922464, EPI_ISL_8922469, EPI_ISL_8922471, EPI_ISL_8922475, EPI_ISL_8922481, EPI_ISL_8922482, EPI_ISL_8922488, EPI_ISL_8922489, EPI_ISL_8922491, EPI_ISL_8922507, EPI_ISL_8922516, EPI_ISL_8922523, EPI_ISL_8922533, EPI_ISL_8922701, EPI_ISL_8922703, EPI_ISL_8922705, EPI_ISL_8922711, EPI_ISL_8922713, EPI_ISL_8922717, EPI_ISL_8922721, EPI_ISL_8922726, EPI_ISL_8922728, EPI_ISL_8922734, EPI_ISL_8922737, EPI_ISL_8922741, EPI_ISL_8922743, EPI_ISL_8922745, EPI_ISL_8922749, EPI_ISL_8922751, EPI_ISL_8922767, EPI_ISL_8922770, EPI_ISL_8922772, EPI_ISL_8922777, EPI_ISL_8922783, EPI_ISL_8922785, EPI_ISL_8922787, EPI_ISL_8922790, EPI_ISL_8922798, EPI_ISL_8922799, EPI_ISL_8922801, EPI_ISL_8922805, EPI_ISL_8922808, EPI_ISL_8922810, EPI_ISL_8922816, EPI_ISL_8922817, EPI_ISL_8922822, EPI_ISL_8922830, EPI_ISL_8922847, EPI_ISL_8922851, EPI_ISL_8922853, EPI_ISL_8922855, EPI_ISL_8922856, EPI_ISL_8922860, EPI_ISL_8922862, EPI_ISL_8922864, EPI_ISL_8922866, EPI_ISL_8922868, EPI_ISL_8922877, EPI_ISL_8922885, EPI_ISL_8922888, EPI_ISL_8922890, EPI_ISL_8922892, EPI_ISL_8922895, EPI_ISL_8922897, EPI_ISL_8922918, EPI_ISL_8922922, EPI_ISL_8922923, EPI_ISL_8922927, EPI_ISL_8922928, EPI_ISL_8922930, EPI_ISL_8922934, EPI_ISL_8922949, EPI_ISL_8922954, EPI_ISL_8922956, EPI_ISL_8922961, EPI_ISL_8922963, EPI_ISL_8922968, EPI_ISL_8922970, EPI_ISL_8922972, EPI_ISL_8922976, EPI_ISL_8922980, EPI_ISL_8922982, EPI_ISL_8922987, EPI_ISL_8922991, EPI_ISL_8922992, EPI_ISL_8922997, EPI_ISL_8922998, EPI_ISL_8923000, EPI_ISL_8923006, EPI_ISL_8923008, EPI_ISL_8923012, EPI_ISL_8923014, EPI_ISL_8923016, EPI_ISL_8923018, EPI_ISL_8923023, EPI_ISL_8923027, EPI_ISL_8923033, EPI_ISL_8923042, EPI_ISL_8923048, EPI_ISL_8923085, EPI_ISL_8923087, EPI_ISL_8923089, EPI_ISL_8923091, EPI_ISL_8923093, EPI_ISL_8923101, EPI_ISL_8923104, EPI_ISL_8923109, EPI_ISL_8923116, EPI_ISL_8923125, EPI_ISL_8923127, EPI_ISL_8923133, EPI_ISL_8923155, EPI_ISL_8923165, EPI_ISL_8923169, EPI_ISL_8923170, EPI_ISL_8923174, EPI_ISL_8923403 | | | |
| see above | Department of Bacteria, Parasites and Fungi, Statens Serum Institut, Copenhagen, Denmark | Statens Serum Institut Bioinformatics and Microbial Genomics | Danish Covid-19 Genome Consortium |
| EPI_ISL_8920923 | INSPI-CRN DE INFLUENZA Y OTROS VIRUS RESPIRATORIOS | INSPI-CRN DE INFLUENZA Y OTROS VIRUS RESPIRATORIOS | Alfredo Bruno; Diego Morales; Fernando Llerena; Jimmy Garcés; Maritza Olmedo; Michelle Páez; Ruben Armas |
| EPI_ISL_8919813, EPI_ISL_8919825, EPI_ISL_8919846, EPI_ISL_8919893, EPI_ISL_8919903, EPI_ISL_8919904, EPI_ISL_8919906, EPI_ISL_8919907, EPI_ISL_8919908, EPI_ISL_8919923, EPI_ISL_8919959, EPI_ISL_8919960, EPI_ISL_8919961, EPI_ISL_8919971, EPI_ISL_8919978, EPI_ISL_8920033, EPI_ISL_8920034, EPI_ISL_8920036, EPI_ISL_8920041, EPI_ISL_8920044, EPI_ISL_8920063, EPI_ISL_8920064, EPI_ISL_8920070, EPI_ISL_8920071, EPI_ISL_8920072, EPI_ISL_8920120, EPI_ISL_8920130, EPI_ISL_8920131, EPI_ISL_8920132, EPI_ISL_8920133, EPI_ISL_8920143, EPI_ISL_8920144, EPI_ISL_8920233, EPI_ISL_8920236, EPI_ISL_8920252, EPI_ISL_8920266, EPI_ISL_8920267, EPI_ISL_8920268, EPI_ISL_8920269, EPI_ISL_8920296, EPI_ISL_8920297, EPI_ISL_8920298, EPI_ISL_8920320, EPI_ISL_8920334, EPI_ISL_8920345, EPI_ISL_8920346, EPI_ISL_8920356, EPI_ISL_8920359, EPI_ISL_8920362, EPI_ISL_8920366, EPI_ISL_8920367, EPI_ISL_8920389, EPI_ISL_8920393, EPI_ISL_8920394, EPI_ISL_8920407, EPI_ISL_8920415, EPI_ISL_8920419, EPI_ISL_8920420, EPI_ISL_8920421, EPI_ISL_8920438 | | | |
| see above | SARS-CoV-2 testing team, National Institute of Infectious Diseases | Pathogen Genomics Center, National Institute of Infectious Diseases | Hazuka Y Furihata; Kentaro Itokawa; Makoto Kuroda; Masanori Hashino; Masumichi Saito; Naomi Nojiri; Nozomu Hanaoka; Rina Tanaka; Tsuguto Fujimoto; Tsuyoshi Sekizuka |
| EPI_ISL_8919712, EPI_ISL_8919721, EPI_ISL_8919724 | Tokyo Metropolitan Institute of Public Health | Tokyo Metropolitan Institute of Public Health | Akane Negishi; Arisa Amano; Hirofumi Miyake; Jun Suzuki; Kenji Sadamasu; Kenshirou Kuroki; Mami Nagashima; Masaki Hayashi; Maya Isogai; Takako Yamazaki; Takushi Fujiwara |

We gratefully acknowledge the following Authors from the Originating laboratories responsible for obtaining the specimens, as well as the Submitting laboratories where the genome data were generated and shared via GISAID, on which this research is based.

All Submitters of data may be contacted directly via www.gisaid.org

Authors are sorted alphabetically.

| Accession ID | Originating Laboratory | Submitting Laboratory | Authors |
|---|---|---|---|
| EPI_ISL_8934094 | ASST GOM NIGUARDA | ASST Grande ospedale Metropolitano Niguarda | Alice Nava |
| EPI_ISL_8943096, EPI_ISL_8947554, EPI_ISL_8947588, EPI_ISL_8950543, EPI_ISL_8950554, EPI_ISL_8950562, EPI_ISL_8950655, EPI_ISL_8951105, EPI_ISL_8951290 | | | |
| see above | Aegis Sciences Corporation | Centers for Disease Control and Prevention Division of Viral Diseases, Pathogen Discovery | Alec Vest; Benjamin Rambo-Martin; Christopher Gulvick; Clinton Paden; Cyndi Clark; Dakota Howard; Dhwani Batra; Dillon Nall; Duncan MacCannell; Erisa Sula; Ethan Sanders; Holly Houdeshell; Jason Caravas; Kristine Lacek; Matthew Hardison; Matthew Schmerer; Ola Kvalvaag; Patrick Campbell; Peter Cook; Rob Case; Scott Sammons; Shatavia Morrison; Shaun Westlund; Tymeckia Kendall; Victoria Caban Figueroa; Vikramsinha Ghorpade; Yvette Unoarumhi |
| EPI_ISL_8935629, EPI_ISL_8935630, EPI_ISL_8935631, EPI_ISL_8935632 | CDPH VBL | California Department of Public Health | CDPH-COVIDNet |
| EPI_ISL_8931581, EPI_ISL_8931603, EPI_ISL_8931629 | Cliniques universitaires Saint-Luc | UCLouvain/IREC/MBLG-CTMA | Benoit Kabamba Mukadi; Bertrand Bearzatto; Jean-Luc Gala; Nicolas Pinte; Paul Blanpain; Simon Ophélie; Valentin Coste |
| EPI_ISL_8953795, EPI_ISL_8953796, EPI_ISL_8953801 | Colorado Department of Public Health and Environment | Colorado Department of Public Health and Environment | Alexandria Rossheim; Arianna Smith; Diana Ir; Emily A. Travanty; Laura Bankers; Mandy Waters; Michael Martin; Molly C. Hetherington-Rauth; Shannon R. Matzinger |
| EPI_ISL_8929317, EPI_ISL_8929318, EPI_ISL_8929319, EPI_ISL_8929320, EPI_ISL_8929321, EPI_ISL_8929322 | Division of Emerging Infectious Diseases, Bureau of Infectious Diseases Diagnosis Control, Korea Disease Control and Prevention Agency | Division of Emerging Infectious Diseases, Bureau of Infectious Diseases Diagnosis Control, Korea Disease Control and Prevention Agency | Ae Kyung Park; Chae Young Lee; Eun-Jin Kim; Hyuck Jin Lee; Il-Hwan Kim; Jeong-Ah Kim |
| EPI_ISL_8938767, EPI_ISL_8940304 | Fulgent Genetics | Centers for Disease Control and Prevention Division of Viral Diseases, Pathogen Discovery | Becky Tsai; Benafsh Sapra; Benjamin Rambo-Martin; Christopher Gulvick; Clinton Paden; Dakota Howard; Dhwani Batra; Doreen Ng; Duncan MacCannell; Erisa Sula; Harry Gao; James Xie; Jason Caravas; John Gao; Joseph Fierro; Kristine Lacek; Matthew Schmerer; Mickey Li; Peter Cook; Scott Sammons; Shatavia Morrison; Tymeckia Kendall; Victoria Caban Figueroa; Yan Meng; Yvette Unoarumhi |
| EPI_ISL_8947237 | Infinity Biologix | Centers for Disease Control and Prevention Division of Viral Diseases, Pathogen Discovery | Benjamin Rambo-Martin; Chirayu Goswami; Christian Bixby; Christopher Gulvick; Clinton Paden; Dakota Howard; Dhwani Batra; Duncan MacCannell; Erisa Sula; Jason Caravas; Jonathan Schultz; Kristine Lacek; Matthew Schmerer; Peter Cook; Robin Grimwood; Russ Hager; Scott Sammons; Shatavia Morrison; Tymeckia Kendall; Victoria Caban Figueroa; Yihe Wang; Yvette Unoarumhi |
| EPI_ISL_8938300 | Laboratorio Central, Ministerio de Salud Córdoba | Fundacion para el Progreso de la Medicina and Instituto de Patologia Vegetal (IPAVE-CIAP-INTA) on behalf of 'Proyecto Argentino Interinstitucional de genomica de SARS-CoV-2' (PAIS Consortium) | Barbas, G.; Castro, G.; Debat, H.; F.D.; Fernández; Goya, S.; Lucca, A.; M.B.; Marquez, N.; Pisano; Re, V.; Sicilia, P.; Zeballos, M. |
| EPI_ISL_8953561 | Laboratorio De Patología Molecular Secretaria De Marina Centro Medico Naval | Laboratorio De Patología Molecular Secretaria De Marina Centro Medico Naval | Morales Ramirez Omar. Baltierra Uribe Shantall. Meixueiro Calderon Claudia. Garcia Alegria Irma. Zuno Coronado Teutli. Alvarez Jimenez Violeta Dejanira. Cabrera Gonzalez Moises. Romero Espejel Maria Elena. Mora Heredia Jose Eduardo. Rubio Ortiz Margarita |
| EPI_ISL_8953052 | Mako Medical | Centers for Disease Control and Prevention Division of Viral Diseases, Pathogen Discovery | Benjamin Rambo-Martin; Christopher Gulvick; Clinton Paden; Dakota Howard; Dhwani Batra; Duncan MacCannell; Erisa Sula; Jason Caravas; Kristine Lacek; Lauren Moon; Matthew Schmerer; Matthew Tugwell; Peter Cook; Scott Sammons; Shatavia Morrison; Tymeckia Kendall; Victoria Caban Figueroa; Yvette Unoarumhi |
| EPI_ISL_8929501, EPI_ISL_8929502 | Ministry of Health Turkey | Ministry of Health Turkey | Fatma Bayrakdar; Gülay Korukluoğlu; Gültekin Ünal; Süleyman Yalcin; Yasemin Coşgun |
| EPI_ISL_8937373 | PathWest Laboratory Medicine WA | PathWest Laboratory Medicine WA Microbial Surveillance Unit | PathWest Laboratory Medicine WA Microbial Surveillance Unit |
| EPI_ISL_8931071 | Polski Bank Komórek Macierzystych | WSSE w Warszawie | Dorota Wagrocka - Roczniak |
| EPI_ISL_8946339 | Rosalind Franklin Laboratory | Wellcome Sanger Institute for the COVID-19 Genomics UK (COG-UK) Consortium | Cordelia Langford; David K. Jackson; Dominic Kwiatkowski; Donald Fraser; Ewan Harrison; Ian Johnston; Jeffrey Barrett; John Sillitoe on behalf of the Wellcome Sanger Institute COVID-19 Surveillance Team; Rob Howes; Roberto Amato; Sonia Goncalves; Suki Lee; The Rosalind Franklin Laboratory and Alex Alderton |
| EPI_ISL_8953837, EPI_ISL_8953838 | Stanford Health Care | Stanford University School of Medicine, Clinical Virology Laboratory | Daniel Solis; Fumiko Yamamoto; James Zehnder; Malaya K. Sahoo; Mamdouh Sibai; Selamawit Bihon; Suzette Santos; and Benjamin A. Pinsky |
| EPI_ISL_8936681 | Tokyo Metropolitan Institute of Public Health | Tokyo Metropolitan Institute of Public Health | Akane Negishi; Arisa Amano; Hirofumi Miyake; Jun Suzuki; Kenji Sadamasu; Kenshirou Kuroki; Mami Nagashima; Masaki Hayashi; Maya Isogai; Takako Yamazaki; Takushi Fujiwara |
| EPI_ISL_8937981, EPI_ISL_8937989, EPI_ISL_8938000, EPI_ISL_8938010, EPI_ISL_8938015 | URMC LABS | URMC LABS | Andrew Cameron; Dwight Hardy; Kelly DeLary; Stepan Voljehn |
| EPI_ISL_8950784, EPI_ISL_8952751 | Utah Public Health Laboratory | Utah Public Health Laboratory | Erin L. Young; John Arnn; Kelly F. Oakeson; Olinto Linares-Perdomo; Pooja Gupta; Tom Iverson |
| EPI_ISL_8933676 | Vibhavadi Hospital | Center for Medical Genomics Ramathibodi Hospital | Anek mungaomklang; Insee Sensorn; Inthuorn Ratchatasetthakul; Kanyarat Janthima; Parsakorn Tapaopong; Poramate Jiaranai; Vichapon Tiacharoen; Wasun Chantratita |
| EPI_ISL_8938156, EPI_ISL_8938170 | Wyoming Public Health Laboratory | Wyoming Public Health Laboratory | Ashley Norberg; Brian Dominguez; Cari Sloma; Channing Weber; Chayse Rowley; Elliot Thomasson; Jim Mildenberger; Marley Goetz; Robert Petit; Sam Britz; Taylor Fearing; and Rob Christensen |

We gratefully acknowledge the following Authors from the Originating laboratories responsible for obtaining the specimens, as well as the Submitting laboratories where the genome data were generated and shared via GISAID, on which this research is based.

All Submitters of data may be contacted directly via www.gisaid.org

Authors are sorted alphabetically.

Acknowledgement EPI_SET Identifier: EPI_SET_20220527nw

| Accession ID | Originating Laboratory | Submitting Laboratory | Authors |
|---|---|---|---|
| EPI_ISL_8928232, EPI_ISL_8928234, EPI_ISL_8928236, EPI_ISL_8928240, EPI_ISL_8928248, EPI_ISL_8928250, EPI_ISL_8928255, EPI_ISL_8928257, EPI_ISL_8928262, EPI_ISL_8928264, EPI_ISL_8928266, EPI_ISL_8928276, EPI_ISL_8928282, EPI_ISL_8928284, EPI_ISL_8928285, EPI_ISL_8928287, EPI_ISL_8928289, EPI_ISL_8928293, EPI_ISL_8928296, EPI_ISL_8928304, EPI_ISL_8928307, EPI_ISL_8928309 | | | |
| see above | Department of Bacteria, Parasites and Fungi, Statens Serum Institut, Copenhagen, Denmark | Statens Serum Institut Bioinformatics and Microbial Genomics | Danish Covid-19 Genome Consortium |
| EPI_ISL_8929304, EPI_ISL_8929305, EPI_ISL_8929306, EPI_ISL_8929307, EPI_ISL_8929308, EPI_ISL_8929309, EPI_ISL_8929310, EPI_ISL_8929311, EPI_ISL_8929312, EPI_ISL_8929313, EPI_ISL_8929314, EPI_ISL_8929315, EPI_ISL_8929316 | | | |
| see above | Division of Emerging Infectious Diseases, Bureau of Infectious Diseases Diagnosis Control, Korea Disease Control and Prevention Agency | Division of Emerging Infectious Diseases, Bureau of Infectious Diseases Diagnosis Control, Korea Disease Control and Prevention Agency | Ae Kyung Park; Chae Young Lee; Eun-Jin Kim; Hyuck Jin Lee; Il-Hwan Kim; Jeong-Ah Kim |
| EPI_ISL_8929209, EPI_ISL_8929225 | Dutch COVID-19 response team | Medical Microbiology, Maastricht University Medical Centre | Brian van der Veer*; Carmen Reumkens; Christian Hoebe; Erik Beuken; Jozef Dingemans*; Lieke van Alphen; Paul Savelkoul |
| EPI_ISL_8928516, EPI_ISL_8928517, EPI_ISL_8928518, EPI_ISL_8928519, EPI_ISL_8928529, EPI_ISL_8928530 | Furst Medical Laboratory | Norwegian Institute of Public Health, Department of Virology | Atiya R Ali; Debech Nadia; Engebretsen Serina Beate; Garcia Llorente Ignacio; Hilde Elshaug; Hilde Vollan; Jon Bråte; Kamilla Heddeland Instefjord; Karoline Bragstad; Kathrine Stene-Johansen; Line Victoria Moen; Marie Paulsen Madsen; Olav Hungnes; Pedersen Benedikte Nevjen; Rasmus Riis Kopperud |
| EPI_ISL_8928326, EPI_ISL_8928388, EPI_ISL_8928431, EPI_ISL_8928434, EPI_ISL_8928436, EPI_ISL_8928460 | Oslo University Hospital, Department of Microbiology | Norwegian Institute of Public Health, Department of Virology | Arvind Yegambaram Meenakshi Sundaram; Cathrine Fladeby; Garcia Llorente Ignacio; Gregor D. Gilfillan; Hilde Elshaug; Hilde Vollan; Jon Bråte; Kamilla Heddeland Instefjord; Karoline Bragstad; Kathrine Stene-Johansen; Line Victoria Moen; Lise Andresen; Mariann Nilsen; Mona Holberg-Petersen; Olav Hungnes; Pedersen Benedikte Nevjen; Pål Marius Bjørnstad; Rasmus Riis Kopperud; Teodora Plamenova Ribarska |
| EPI_ISL_8928596 | Polski Bank Komórek Macierzystych | WSSE w Warszawie | Dorota Wagrocka - Roczniak |

We gratefully acknowledge the following Authors from the Originating laboratories responsible for obtaining the specimens, as well as the Submitting laboratories where the genome data were generated and shared via GISAID, on which this research is based.

All Submitters of data may be contacted directly via www.gisaid.org

Authors are sorted alphabetically.

| Accession ID | Originating Laboratory | Submitting Laboratory | Authors |
| --- | --- | --- | --- |
| EPI_ISL_8927645, EPI_ISL_8927647, EPI_ISL_8927649, EPI_ISL_8927652, EPI_ISL_8927756, EPI_ISL_8927765, EPI_ISL_8927769, EPI_ISL_8927778, EPI_ISL_8927780, EPI_ISL_8927787, EPI_ISL_8927789, EPI_ISL_8927801, EPI_ISL_8927803, EPI_ISL_8927806, EPI_ISL_8927812, EPI_ISL_8927823, EPI_ISL_8927935, EPI_ISL_8928098, EPI_ISL_8928101, EPI_ISL_8928107, EPI_ISL_8928113, EPI_ISL_8928119, EPI_ISL_8928126, EPI_ISL_8928130, EPI_ISL_8928132, EPI_ISL_8928136, EPI_ISL_8928142, EPI_ISL_8928148, EPI_ISL_8928155, EPI_ISL_8928163, EPI_ISL_8928165, EPI_ISL_8928172, EPI_ISL_8928174, EPI_ISL_8928176, EPI_ISL_8928177, EPI_ISL_8928179, EPI_ISL_8928183, EPI_ISL_8928185, EPI_ISL_8928188, EPI_ISL_8928190, EPI_ISL_8928192, EPI_ISL_8928200, EPI_ISL_8928202, EPI_ISL_8928204, EPI_ISL_8928208, EPI_ISL_8928212, EPI_ISL_8928227, EPI_ISL_8928230 | | | |
| see above | Department of Bacteria, Parasites and Fungi, Statens Serum Institut, Copenhagen, Denmark | Statens Serum Institut Bioinformatics and Microbial Genomics | Danish Covid-19 Genome Consortium |
| EPI_ISL_8928084 | Medical Microbiology Unit, Department for Laboratory Medicine, Drammen Hospital, Vestre Viken Health Trust | Norwegian Institute of Public Health, Department of Virology | Atiya R Ali; Debech Nadia; Engebretsen Serina Beate; Garcia Llorente Ignacio; Hilde Elshaug; Hilde Vollan; Jon Bråte; Kamilla Heddeland Instefjord; Karoline Bragstad; Kathrine Stene-Johansen; Line Victoria Moen; Marie Paulsen Madsen; Olav Hungnes; Pedersen Benedikte Nevjen; Rasmus Riis Kopperud |
| EPI_ISL_8928077 | Ostfold Hospital Trust - Kalnes, Centre for Laboratory Medicine, Section for gene technology and infection serology | Norwegian Institute of Public Health, Department of Virology | Atiya R Ali; Debech Nadia; Engebretsen Serina Beate; Garcia Llorente Ignacio; Hilde Elshaug; Hilde Vollan; Jon Bråte; Kamilla Heddeland Instefjord; Karoline Bragstad; Kathrine Stene-Johansen; Line Victoria Moen; Marie Paulsen Madsen; Olav Hungnes; Pedersen Benedikte Nevjen; Rasmus Riis Kopperud |

We gratefully acknowledge the following Authors from the Originating laboratories responsible for obtaining the specimens, as well as the Submitting laboratories where the genome data were generated and shared via GISAID, on which this research is based.

All Submitters of data may be contacted directly via www.gisaid.org

Authors are sorted alphabetically.

| Accession ID | Originating Laboratory | Submitting Laboratory | Authors |
|---|---|---|---|
| EPI_ISL_8927387, EPI_ISL_8927390, EPI_ISL_8927405, EPI_ISL_8927423, EPI_ISL_8927431, EPI_ISL_8927435, EPI_ISL_8927438, EPI_ISL_8927442, EPI_ISL_8927445, EPI_ISL_8927454, EPI_ISL_8927456, EPI_ISL_8927465, EPI_ISL_8927467, EPI_ISL_8927473, EPI_ISL_8927478, EPI_ISL_8927491, EPI_ISL_8927498, EPI_ISL_8927505, EPI_ISL_8927507, EPI_ISL_8927515, EPI_ISL_8927517, EPI_ISL_8927519, EPI_ISL_8927521, EPI_ISL_8927523, EPI_ISL_8927527, EPI_ISL_8927528, EPI_ISL_8927537, EPI_ISL_8927542, EPI_ISL_8927544, EPI_ISL_8927545, EPI_ISL_8927550, EPI_ISL_8927553, EPI_ISL_8927555, EPI_ISL_8927557, EPI_ISL_8927562, EPI_ISL_8927564, EPI_ISL_8927567, EPI_ISL_8927575, EPI_ISL_8927578, EPI_ISL_8927583, EPI_ISL_8927584, EPI_ISL_8927590, EPI_ISL_8927597, EPI_ISL_8927605, EPI_ISL_8927627, EPI_ISL_8927629, EPI_ISL_8927631, EPI_ISL_8927632, EPI_ISL_8927636, EPI_ISL_8927640 | | | |
| see above | Department of Bacteria, Parasites and Fungi, Statens Serum Institut, Copenhagen, Denmark | Statens Serum Institut Bioinformatics and Microbial Genomics | Danish Covid-19 Genome Consortium |

We gratefully acknowledge the following Authors from the Originating laboratories responsible for obtaining the specimens, as well as the Submitting laboratories where the genome data were generated and shared via GISAID, on which this research is based.

All Submitters of data may be contacted directly via www.gisaid.org

Authors are sorted alphabetically.

Acknowledgement EPI_SET Identifier: EPI_SET_20220527yz

| Accession ID | Originating Laboratory | Submitting Laboratory | Authors |
|---|---|---|---|
| EPI_ISL_8927302, EPI_ISL_8927322, EPI_ISL_8927325, EPI_ISL_8927334, EPI_ISL_8927362, EPI_ISL_8927364, EPI_ISL_8927373 | | | |
| see above | Department of Bacteria, Parasites and Fungi, Statens Serum Institut, Copenhagen, Denmark | Statens Serum Institut Bioinformatics and Microbial Genomics | Danish Covid-19 Genome Consortium |
| EPI_ISL_8927251, EPI_ISL_8927252, EPI_ISL_8927253, EPI_ISL_8927254, EPI_ISL_8927255, EPI_ISL_8927256, EPI_ISL_8927257, EPI_ISL_8927258, EPI_ISL_8927259, EPI_ISL_8927261, EPI_ISL_8927262, EPI_ISL_8927263, EPI_ISL_8927264, EPI_ISL_8927265, EPI_ISL_8927266, EPI_ISL_8927267, EPI_ISL_8927268, EPI_ISL_8927269, EPI_ISL_8927270, EPI_ISL_8927271, EPI_ISL_8927272, EPI_ISL_8927273, EPI_ISL_8927274, EPI_ISL_8927275, EPI_ISL_8927276, EPI_ISL_8927277, EPI_ISL_8927278, EPI_ISL_8927279, EPI_ISL_8927280, EPI_ISL_8927281, EPI_ISL_8927282, EPI_ISL_8927283, EPI_ISL_8927284, EPI_ISL_8927285, EPI_ISL_8927286, EPI_ISL_8927287, EPI_ISL_8927288, EPI_ISL_8927289, EPI_ISL_8927290, EPI_ISL_8927291, EPI_ISL_8927292, EPI_ISL_8927293, EPI_ISL_8927295 | | | |
| see above | National Public Health Laboratory, National Centre for Infectious Diseases | National Public Health Laboratory, National Centre for Infectious Diseases | Benny Yeo; Grace Ngan; Lin Cui; Raymond Tzer Pin Lin; Royce Ang; Samuel Loo; Yichen Ding; Zhenyang Zhou |

We gratefully acknowledge the following Authors from the Originating laboratories responsible for obtaining the specimens, as well as the Submitting laboratories where the genome data were generated and shared via GISAID, on which this research is based.

All Submitters of data may be contacted directly via www.gisaid.org

Authors are sorted alphabetically.

| Accession ID | Originating Laboratory | Submitting Laboratory | Authors |
|---|---|---|---|
| EPI_ISL_8927026, EPI_ISL_8927027, EPI_ISL_8927030, EPI_ISL_8927033, EPI_ISL_8927036, EPI_ISL_8927040, EPI_ISL_8927069, EPI_ISL_8927072, EPI_ISL_8927075, EPI_ISL_8927082, EPI_ISL_8927086, EPI_ISL_8927096, EPI_ISL_8927103, EPI_ISL_8927107, EPI_ISL_8927119, EPI_ISL_8927122, EPI_ISL_8927125, EPI_ISL_8927209 | | | |
| see above | Department of Bacteria, Parasites and Fungi, Statens Serum Institut, Copenhagen, Denmark | Statens Serum Institut Bioinformatics and Microbial Genomics | Danish Covid-19 Genome Consortium |
| EPI_ISL_8927219, EPI_ISL_8927220, EPI_ISL_8927221, EPI_ISL_8927222, EPI_ISL_8927223, EPI_ISL_8927224, EPI_ISL_8927225, EPI_ISL_8927226, EPI_ISL_8927227, EPI_ISL_8927228, EPI_ISL_8927229, EPI_ISL_8927230, EPI_ISL_8927231, EPI_ISL_8927232, EPI_ISL_8927233, EPI_ISL_8927234, EPI_ISL_8927235, EPI_ISL_8927236, EPI_ISL_8927237, EPI_ISL_8927238, EPI_ISL_8927239, EPI_ISL_8927240, EPI_ISL_8927241, EPI_ISL_8927242, EPI_ISL_8927243, EPI_ISL_8927244, EPI_ISL_8927245, EPI_ISL_8927246, EPI_ISL_8927247, EPI_ISL_8927248, EPI_ISL_8927249, EPI_ISL_8927250 | | | |
| see above | National Public Health Laboratory, National Centre for Infectious Diseases | National Public Health Laboratory, National Centre for Infectious Diseases | Benny Yeo; Grace Ngan; Lin Cui; Raymond Tzer Pin Lin; Royce Ang; Samuel Loo; Yichen Ding; Zhenyang Zhou |

We gratefully acknowledge the following Authors from the Originating laboratories responsible for obtaining the specimens, as well as the Submitting laboratories where the genome data were generated and shared via GISAID, on which this research is based.

All Submitters of data may be contacted directly via www.gisaid.org

Authors are sorted alphabetically.

| Accession ID | Originating Laboratory | Submitting Laboratory | Authors |
|---|---|---|---|
| EPI_ISL_8926743, EPI_ISL_8926745, EPI_ISL_8926747, EPI_ISL_8926749, EPI_ISL_8926751, EPI_ISL_8926759, EPI_ISL_8926764, EPI_ISL_8926773, EPI_ISL_8926776, EPI_ISL_8926780, EPI_ISL_8926783, EPI_ISL_8926786, EPI_ISL_8926793, EPI_ISL_8926797, EPI_ISL_8926802, EPI_ISL_8926805, EPI_ISL_8926808, EPI_ISL_8926811, EPI_ISL_8926813, EPI_ISL_8926816, EPI_ISL_8926818, EPI_ISL_8926821, EPI_ISL_8926828, EPI_ISL_8926838, EPI_ISL_8926842, EPI_ISL_8926843, EPI_ISL_8926849, EPI_ISL_8926857, EPI_ISL_8926861, EPI_ISL_8926868, EPI_ISL_8926877, EPI_ISL_8926879, EPI_ISL_8926881, EPI_ISL_8926888, EPI_ISL_8926890, EPI_ISL_8926893, EPI_ISL_8926896, EPI_ISL_8926904, EPI_ISL_8926908, EPI_ISL_8926910, EPI_ISL_8926914, EPI_ISL_8926923, EPI_ISL_8926931, EPI_ISL_8926945, EPI_ISL_8926993, EPI_ISL_8927003, EPI_ISL_8927010, EPI_ISL_8927014, EPI_ISL_8927018, EPI_ISL_8927025 | | | |
| see above | Department of Bacteria, Parasites and Fungi, Statens Serum Institut, Copenhagen, Denmark | Statens Serum Institut Bioinformatics and Microbial Genomics | Danish Covid-19 Genome Consortium |

We gratefully acknowledge the following Authors from the Originating laboratories responsible for obtaining the specimens, as well as the Submitting laboratories where the genome data were generated and shared via GISAID, on which this research is based.

All Submitters of data may be contacted directly via www.gisaid.org

Authors are sorted alphabetically.

Acknowledgement EPI_SET Identifier: EPI_SET_20220527mz

| Accession ID | Originating Laboratory | Submitting Laboratory | Authors |
|---|---|---|---|
| EPI_ISL_8926265, EPI_ISL_8926269, EPI_ISL_8926277, EPI_ISL_8926295, EPI_ISL_8926307, EPI_ISL_8926316, EPI_ISL_8926318, EPI_ISL_8926326, EPI_ISL_8926328, EPI_ISL_8926329, EPI_ISL_8926330, EPI_ISL_8926332, EPI_ISL_8926336, EPI_ISL_8926340, EPI_ISL_8926345, EPI_ISL_8926351, EPI_ISL_8926360, EPI_ISL_8926362, EPI_ISL_8926367, EPI_ISL_8926369, EPI_ISL_8926373, EPI_ISL_8926390, EPI_ISL_8926391, EPI_ISL_8926394, EPI_ISL_8926401, EPI_ISL_8926403, EPI_ISL_8926408, EPI_ISL_8926410, EPI_ISL_8926415, EPI_ISL_8926416, EPI_ISL_8926576, EPI_ISL_8926579, EPI_ISL_8926585, EPI_ISL_8926596, EPI_ISL_8926598, EPI_ISL_8926601, EPI_ISL_8926604, EPI_ISL_8926605, EPI_ISL_8926608, EPI_ISL_8926611, EPI_ISL_8926613, EPI_ISL_8926615, EPI_ISL_8926617, EPI_ISL_8926621, EPI_ISL_8926627, EPI_ISL_8926633, EPI_ISL_8926634, EPI_ISL_8926637, EPI_ISL_8926640, EPI_ISL_8926740 | | | |
| see above | Department of Bacteria, Parasites and Fungi, Statens Serum Institut, Copenhagen, Denmark | Statens Serum Institut Bioinformatics and Microbial Genomics | Danish Covid-19 Genome Consortium |

We gratefully acknowledge the following Authors from the Originating laboratories responsible for obtaining the specimens, as well as the Submitting laboratories where the genome data were generated and shared via GISAID, on which this research is based.

All Submitters of data may be contacted directly via www.gisaid.org

Authors are sorted alphabetically.

| Accession ID | Originating Laboratory | Submitting Laboratory | Authors |
| --- | --- | --- | --- |
| EPI_ISL_8925531, EPI_ISL_8925532, EPI_ISL_8925533, EPI_ISL_8925534, EPI_ISL_8925541, EPI_ISL_8925542, EPI_ISL_8925547, EPI_ISL_8925553, EPI_ISL_8925566, EPI_ISL_8925568, EPI_ISL_8925570, EPI_ISL_8925574, EPI_ISL_8925581, EPI_ISL_8925585, EPI_ISL_8925587, EPI_ISL_8925589, EPI_ISL_8925592, EPI_ISL_8925594, EPI_ISL_8925596, EPI_ISL_8925600, EPI_ISL_8925602, EPI_ISL_8925611, EPI_ISL_8925637, EPI_ISL_8925639, EPI_ISL_8925648, EPI_ISL_8925650, EPI_ISL_8925651, EPI_ISL_8925652, EPI_ISL_8925653, EPI_ISL_8925654, EPI_ISL_8925655, EPI_ISL_8925656, EPI_ISL_8925657, EPI_ISL_8925658, EPI_ISL_8925659, EPI_ISL_8925660, EPI_ISL_8925661, EPI_ISL_8925662, EPI_ISL_8925670, EPI_ISL_8925681, EPI_ISL_8925685, EPI_ISL_8925686, EPI_ISL_8926037, EPI_ISL_8926246, EPI_ISL_8926249, EPI_ISL_8926251, EPI_ISL_8926255, EPI_ISL_8926259, EPI_ISL_8926263 | | | |
| see above | Department of Bacteria, Parasites and Fungi, Statens Serum Institut, Copenhagen, Denmark | Statens Serum Institut Bioinformatics and Microbial Genomics | Danish Covid-19 Genome Consortium |
| EPI_ISL_8925674 | Unipath Speciality Laboratory Limited, Ahmedabad | Gujarat Biotechnology Research Centre | Akhilesh Modi; Apurvasinh Puvar; Bhadreshsinh Gohil; Chaitanya Joshi; Disha Vora; Janvi Raval; Jaykumar Rangani; Jwalant Shah; Madhvi Joshi; Nimesh Patel; Nitin Savaliya; NitinShukla; Priyank Chavda; Ramesh Pandit; Roshani Mishra; Sonal Sharma; Tasnim Trivedi; Zarna Patel |

We gratefully acknowledge the following Authors from the Originating laboratories responsible for obtaining the specimens, as well as the Submitting laboratories where the genome data were generated and shared via GISAID, on which this research is based.

All Submitters of data may be contacted directly via www.gisaid.org

Authors are sorted alphabetically.

Acknowledgement EPI_SET Identifier: EPI_SET_20220527wc

| Accession ID | Originating Laboratory | Submitting Laboratory | Authors |
|---|---|---|---|
| EPI_ISL_8925264, EPI_ISL_8925266, EPI_ISL_8925267, EPI_ISL_8925272, EPI_ISL_8925273, EPI_ISL_8925276, EPI_ISL_8925277, EPI_ISL_8925295, EPI_ISL_8925298, EPI_ISL_8925299, EPI_ISL_8925301, EPI_ISL_8925302, EPI_ISL_8925303, EPI_ISL_8925304, EPI_ISL_8925307, EPI_ISL_8925312, EPI_ISL_8925316, EPI_ISL_8925318, EPI_ISL_8925320, EPI_ISL_8925322, EPI_ISL_8925324, EPI_ISL_8925327, EPI_ISL_8925331, EPI_ISL_8925366, EPI_ISL_8925429, EPI_ISL_8925431, EPI_ISL_8925434, EPI_ISL_8925436, EPI_ISL_8925440, EPI_ISL_8925446, EPI_ISL_8925448, EPI_ISL_8925458, EPI_ISL_8925460, EPI_ISL_8925462, EPI_ISL_8925471, EPI_ISL_8925478, EPI_ISL_8925483, EPI_ISL_8925485, EPI_ISL_8925486, EPI_ISL_8925488, EPI_ISL_8925494, EPI_ISL_8925500, EPI_ISL_8925501, EPI_ISL_8925503, EPI_ISL_8925507, EPI_ISL_8925513, EPI_ISL_8925519, EPI_ISL_8925523, EPI_ISL_8925527, EPI_ISL_8925529 | | | |
| see above | Department of Bacteria, Parasites and Fungi, Statens Serum Institut, Copenhagen, Denmark | Statens Serum Institut Bioinformatics and Microbial Genomics | Danish Covid-19 Genome Consortium |

We gratefully acknowledge the following Authors from the Originating laboratories responsible for obtaining the specimens, as well as the Submitting laboratories where the genome data were generated and shared via GISAID, on which this research is based.

All Submitters of data may be contacted directly via www.gisaid.org

Authors are sorted alphabetically.

Acknowledgement EPI_SET Identifier: EPI_SET_20220527kz

| Accession ID | Originating Laboratory | Submitting Laboratory | Authors |
|---|---|---|---|
| EPI_ISL_8925102, EPI_ISL_8925104, EPI_ISL_8925106, EPI_ISL_8925107, EPI_ISL_8925109, EPI_ISL_8925117, EPI_ISL_8925118, EPI_ISL_8925120, EPI_ISL_8925126, EPI_ISL_8925127, EPI_ISL_8925131, EPI_ISL_8925133, EPI_ISL_8925136, EPI_ISL_8925144, EPI_ISL_8925146, EPI_ISL_8925155, EPI_ISL_8925163, EPI_ISL_8925167, EPI_ISL_8925169, EPI_ISL_8925172, EPI_ISL_8925174, EPI_ISL_8925176, EPI_ISL_8925178, EPI_ISL_8925180, EPI_ISL_8925181, EPI_ISL_8925185, EPI_ISL_8925186, EPI_ISL_8925222, EPI_ISL_8925223, EPI_ISL_8925224, EPI_ISL_8925225, EPI_ISL_8925227, EPI_ISL_8925230, EPI_ISL_8925232, EPI_ISL_8925235, EPI_ISL_8925236, EPI_ISL_8925239, EPI_ISL_8925240, EPI_ISL_8925243, EPI_ISL_8925244, EPI_ISL_8925247, EPI_ISL_8925248, EPI_ISL_8925249, EPI_ISL_8925250, EPI_ISL_8925251, EPI_ISL_8925252, EPI_ISL_8925259, EPI_ISL_8925260, EPI_ISL_8925261, EPI_ISL_8925262 | | | |
| see above | Department of Bacteria, Parasites and Fungi, Statens Serum Institut, Copenhagen, Denmark | Statens Serum Institut Bioinformatics and Microbial Genomics | Danish Covid-19 Genome Consortium |

We gratefully acknowledge the following Authors from the Originating laboratories responsible for obtaining the specimens, as well as the Submitting laboratories where the genome data were generated and shared via GISAID, on which this research is based.

All Submitters of data may be contacted directly via www.gisaid.org

Authors are sorted alphabetically.

| Accession ID | Originating Laboratory | Submitting Laboratory | Authors |
|---|---|---|---|
| EPI_ISL_8924860, EPI_ISL_8924861, EPI_ISL_8924867, EPI_ISL_8924868, EPI_ISL_8924869, EPI_ISL_8924870, EPI_ISL_8924872, EPI_ISL_8924873, EPI_ISL_8924876, EPI_ISL_8924880, EPI_ISL_8924881, EPI_ISL_8924883, EPI_ISL_8924884, EPI_ISL_8924890, EPI_ISL_8924891, EPI_ISL_8924899, EPI_ISL_8924905, EPI_ISL_8924911, EPI_ISL_8924921, EPI_ISL_8924924, EPI_ISL_8924926, EPI_ISL_8924973, EPI_ISL_8924975, EPI_ISL_8924978, EPI_ISL_8924982, EPI_ISL_8924986, EPI_ISL_8924991, EPI_ISL_8924993, EPI_ISL_8924994, EPI_ISL_8924996, EPI_ISL_8924999, EPI_ISL_8925007, EPI_ISL_8925012, EPI_ISL_8925016, EPI_ISL_8925018, EPI_ISL_8925022, EPI_ISL_8925026, EPI_ISL_8925027, EPI_ISL_8925030, EPI_ISL_8925039, EPI_ISL_8925043, EPI_ISL_8925049, EPI_ISL_8925051, EPI_ISL_8925055, EPI_ISL_8925057, EPI_ISL_8925083, EPI_ISL_8925085, EPI_ISL_8925089, EPI_ISL_8925098, EPI_ISL_8925100 | | | |
| see above | Department of Bacteria, Parasites and Fungi, Statens Serum Institut, Copenhagen, Denmark | Statens Serum Institut Bioinformatics and Microbial Genomics | Danish Covid-19 Genome Consortium |

We gratefully acknowledge the following Authors from the Originating laboratories responsible for obtaining the specimens, as well as the Submitting laboratories where the genome data were generated and shared via GISAID, on which this research is based.

All Submitters of data may be contacted directly via www.gisaid.org

Authors are sorted alphabetically.

Acknowledgement EPI_SET Identifier: EPI_SET_20220527ua

| Accession ID | Originating Laboratory | Submitting Laboratory | Authors |
|---|---|---|---|
| EPI_ISL_8923793, EPI_ISL_8923797, EPI_ISL_8923802, EPI_ISL_8923804, EPI_ISL_8923806, EPI_ISL_8923812, EPI_ISL_8923815, EPI_ISL_8923817, EPI_ISL_8923833, EPI_ISL_8923835, EPI_ISL_8923838, EPI_ISL_8923843, EPI_ISL_8923845, EPI_ISL_8923846, EPI_ISL_8923848, EPI_ISL_8923854, EPI_ISL_8923869, EPI_ISL_8923872, EPI_ISL_8923877, EPI_ISL_8923881, EPI_ISL_8923886, EPI_ISL_8923897, EPI_ISL_8923899, EPI_ISL_8923901, EPI_ISL_8923907, EPI_ISL_8923908, EPI_ISL_8923910, EPI_ISL_8923914, EPI_ISL_8923916, EPI_ISL_8923922, EPI_ISL_8923924, EPI_ISL_8923926, EPI_ISL_8923928, EPI_ISL_8923934, EPI_ISL_8923941, EPI_ISL_8923942, EPI_ISL_8924574, EPI_ISL_8924578, EPI_ISL_8924584, EPI_ISL_8924586, EPI_ISL_8924600, EPI_ISL_8924607, EPI_ISL_8924610, EPI_ISL_8924622, EPI_ISL_8924625, EPI_ISL_8924630, EPI_ISL_8924699, EPI_ISL_8924723, EPI_ISL_8924857, EPI_ISL_8924859 | | | |
| see above | Department of Bacteria, Parasites and Fungi, Statens Serum Institut, Copenhagen, Denmark | Statens Serum Institut Bioinformatics and Microbial Genomics | Danish Covid-19 Genome Consortium |

We gratefully acknowledge the following Authors from the Originating laboratories responsible for obtaining the specimens, as well as the Submitting laboratories where the genome data were generated and shared via GISAID, on which this research is based.

All Submitters of data may be contacted directly via www.gisaid.org

Authors are sorted alphabetically.

| Accession ID | Originating Laboratory | Submitting Laboratory | Authors |
|---|---|---|---|
| EPI_ISL_8923653, EPI_ISL_8923665, EPI_ISL_8923666, EPI_ISL_8923674, EPI_ISL_8923675, EPI_ISL_8923676, EPI_ISL_8923678, EPI_ISL_8923680, EPI_ISL_8923682, EPI_ISL_8923683, EPI_ISL_8923685, EPI_ISL_8923687, EPI_ISL_8923688, EPI_ISL_8923694, EPI_ISL_8923700, EPI_ISL_8923702, EPI_ISL_8923707, EPI_ISL_8923708, EPI_ISL_8923710, EPI_ISL_8923714, EPI_ISL_8923716, EPI_ISL_8923718, EPI_ISL_8923720, EPI_ISL_8923722, EPI_ISL_8923724, EPI_ISL_8923727, EPI_ISL_8923728, EPI_ISL_8923730, EPI_ISL_8923732, EPI_ISL_8923734, EPI_ISL_8923739, EPI_ISL_8923742, EPI_ISL_8923746, EPI_ISL_8923749, EPI_ISL_8923753, EPI_ISL_8923755, EPI_ISL_8923757, EPI_ISL_8923759, EPI_ISL_8923761, EPI_ISL_8923763, EPI_ISL_8923765, EPI_ISL_8923767, EPI_ISL_8923769, EPI_ISL_8923771, EPI_ISL_8923774, EPI_ISL_8923778, EPI_ISL_8923780, EPI_ISL_8923784, EPI_ISL_8923786, EPI_ISL_8923791 | | | |
| see above | Department of Bacteria, Parasites and Fungi, Statens Serum Institut, Copenhagen, Denmark | Statens Serum Institut Bioinformatics and Microbial Genomics | Danish Covid-19 Genome Consortium |

We gratefully acknowledge the following Authors from the Originating laboratories responsible for obtaining the specimens, as well as the Submitting laboratories where the genome data were generated and shared via GISAID, on which this research is based.

All Submitters of data may be contacted directly via www.gisaid.org

Authors are sorted alphabetically.

Acknowledgement EPI_SET Identifier: EPI_SET_20220527nr

EPI_ISL_8923434, EPI_ISL_8923436, EPI_ISL_8923438, EPI_ISL_8923441, EPI_ISL_8923443, EPI_ISL_8923445, EPI_ISL_8923455, EPI_ISL_8923464, EPI_ISL_8923467, EPI_ISL_8923469, EPI_ISL_8923471, EPI_ISL_8923473, EPI_ISL_8923475, EPI_ISL_8923477, EPI_ISL_8923478, EPI_ISL_8923486, EPI_ISL_8923487, EPI_ISL_8923489, EPI_ISL_8923491, EPI_ISL_8923493, EPI_ISL_8923494, EPI_ISL_8923496, EPI_ISL_8923498, EPI_ISL_8923507, EPI_ISL_8923509, EPI_ISL_8923514, EPI_ISL_8923516, EPI_ISL_8923523, EPI_ISL_8923524, EPI_ISL_8923525, EPI_ISL_8923526, EPI_ISL_8923539, EPI_ISL_8923542, EPI_ISL_8923543, EPI_ISL_8923546, EPI_ISL_8923548, EPI_ISL_8923550, EPI_ISL_8923560, EPI_ISL_8923583, EPI_ISL_8923621, EPI_ISL_8923625, EPI_ISL_8923626, EPI_ISL_8923628, EPI_ISL_8923630, EPI_ISL_8923634, EPI_ISL_8923636, EPI_ISL_8923638, EPI_ISL_8923644, EPI_ISL_8923646, EPI_ISL_8923651

| Accession ID | Originating Laboratory | Submitting Laboratory | Authors |
|---|---|---|---|
| see above | Department of Bacteria, Parasites and Fungi, Statens Serum Institut, Copenhagen, Denmark | Statens Serum Institut Bioinformatics and Microbial Genomics | Danish Covid-19 Genome Consortium |

We gratefully acknowledge the following Authors from the Originating laboratories responsible for obtaining the specimens, as well as the Submitting laboratories where the genome data were generated and shared via GISAID, on which this research is based.

All Submitters of data may be contacted directly via www.gisaid.org

Authors are sorted alphabetically.

| Accession ID | Originating Laboratory | Submitting Laboratory | Authors |
|---|---|---|---|
| EPI_ISL_8923177, EPI_ISL_8923181, EPI_ISL_8923186, EPI_ISL_8923190, EPI_ISL_8923195, EPI_ISL_8923203, EPI_ISL_8923204, EPI_ISL_8923211, EPI_ISL_8923212, EPI_ISL_8923214, EPI_ISL_8923216, EPI_ISL_8923251, EPI_ISL_8923255, EPI_ISL_8923259, EPI_ISL_8923260, EPI_ISL_8923263, EPI_ISL_8923264, EPI_ISL_8923266, EPI_ISL_8923271, EPI_ISL_8923279, EPI_ISL_8923283, EPI_ISL_8923285, EPI_ISL_8923294, EPI_ISL_8923296, EPI_ISL_8923298, EPI_ISL_8923303, EPI_ISL_8923305, EPI_ISL_8923307, EPI_ISL_8923310, EPI_ISL_8923314, EPI_ISL_8923322, EPI_ISL_8923329, EPI_ISL_8923331, EPI_ISL_8923333, EPI_ISL_8923335, EPI_ISL_8923345, EPI_ISL_8923348, EPI_ISL_8923352, EPI_ISL_8923360, EPI_ISL_8923361, EPI_ISL_8923363, EPI_ISL_8923365, EPI_ISL_8923375, EPI_ISL_8923384, EPI_ISL_8923388, EPI_ISL_8923390, EPI_ISL_8923393, EPI_ISL_8923399, EPI_ISL_8923403, EPI_ISL_8923412 | | | |
| see above | Department of Bacteria, Parasites and Fungi, Statens Serum Institut, Copenhagen, Denmark | Statens Serum Institut Bioinformatics and Microbial Genomics | Danish Covid-19 Genome Consortium |

We gratefully acknowledge the following Authors from the Originating laboratories responsible for obtaining the specimens, as well as the Submitting laboratories where the genome data were generated and shared via GISAID, on which this research is based.

All Submitters of data may be contacted directly via www.gisaid.org

Authors are sorted alphabetically.

Acknowledgement EPI_SET Identifier: EPI_SET_20220527pn

| Accession ID | Originating Laboratory | Submitting Laboratory | Authors |
|---|---|---|---|
| EPI_ISL_9300753 | ABC Algarve | Instituto Nacional de Saude (INSA) | Borges et al |
| EPI_ISL_9285973 | Allergy, Immunology and Cell Biology Unit (AICBU) | Allergy, Immunology and Cell Biology Unit (AICBU) | Ayesha Wijesinghe; Chandima Jeewandara; Deshni Jayathilaka; Dinuka Ariyaratne; Diyanath Ranasinghe; Dumni Guasinghe; Farha Bary; Gathsaurie Neelika Malavige; Heshan Kuruppu; Tibutius Thanesh |
| EPI_ISL_9300776 | Avelab | Instituto Nacional de Saude (INSA) | Borges et al |
| EPI_ISL_9301587, EPI_ISL_9301588 | Betong hospital | Prince of Songkhla University | Anusara Wongkotsila; Chanon Kongkamon; Pongsakorn Choochuen; Samonrapat Surasombatpattana; Sarunyou Chusri; Surakameth Mahasirimongkol; Surasak Sangkhathat; Thammasin Ingviya; Thanit Sila; Virasakdi Chongsuvivatwong; Wison Laochareonsuk |
| EPI_ISL_9300755 | CHTMAD - Vila Real | Instituto Nacional de Saude (INSA) | Borges et al |
| EPI_ISL_9289788, EPI_ISL_9289789, EPI_ISL_9289790, EPI_ISL_9289795, EPI_ISL_9289798, EPI_ISL_9289806, EPI_ISL_9289823, EPI_ISL_9289824, EPI_ISL_9289829, EPI_ISL_9289831, EPI_ISL_9289836, EPI_ISL_9289841, EPI_ISL_9289842 | | | |
| see above | CSIR-NEERI, Nagpur Covid-19 Testing Lab | CSIR-NEERI, Nagpur Covid-19 Testing Lab | Krishna Khairnar et al. |
| EPI_ISL_9300754 | H Santarem | Instituto Nacional de Saude (INSA) | Borges et al |
| EPI_ISL_9287725, EPI_ISL_9287732 | Hamera Laboratorium | National Institute of Health Research and Development | Arie Ardiansyah Nugraha; Fajar Nur Sulistiyohadi; Hana Apsari Pawestri; Hartanti Dian Ikawati; Kartika Dewi Puspa; Nelly Puspandari; Nur Ika Hariastuti; Putri Widia; Subangkit; Vivi Setiawaty |
| EPI_ISL_9287822, EPI_ISL_9287824, EPI_ISL_9287828 | Kantor Kesehatan Pelabuhan Kelas II Balikpapan | National Institute of Health Research and Development | Arie Ardiansyah Nugraha; Fajar Nur Sulistiyohadi; Hana Apsari Pawestri; Hartanti Dian Ikawati; Kartika Dewi Puspa; Nelly Puspandari; Nur Ika Hariastuti; Puri Widia; Subangkit; Vivi Setiawaty |
| EPI_ISL_9293265 | Lighthouse Lab in Alderley Park | Wellcome Sanger Institute for the COVID-19 Genomics UK (COG-UK) Consortium | Cordelia Langford; David K. Jackson; Dominic Kwiatkowski; Ewan Harrison; Ian Johnston; Jacquelyn Wynn; Jeffrey Barrett; John Sillitoe on behalf of the Wellcome Sanger Institute COVID-19 Surveillance Team; Mairead Hyland; Roberto Amato; Sonia Goncalves; The Lighthouse Lab in Alderley Park and Alex Alderton |
| EPI_ISL_9293266 | Lighthouse Lab in Glasgow | Wellcome Sanger Institute for the COVID-19 Genomics UK (COG-UK) Consortium | Anna Dominiczak and Alex Alderton; Carol Clugston; Cordelia Langford; David Gray; David K. Jackson; Dominic Kwiatkowski; Ewan Harrison; Harper VanSteenhouse; Ian Johnston; Jeffrey Barrett; John Sillitoe on behalf of the Wellcome Sanger Institute COVID-19 Surveillance Team; Roberto Amato; Sonia Goncalves; Yumi Kasai |
| EPI_ISL_9302406, EPI_ISL_9302879, EPI_ISL_9303021, EPI_ISL_9303065 | Microbiological Diagnostic Unit - Public Health Laboratory (MDU-PHL) | Microbiological Diagnostic Unit - Public Health Laboratory (MDU-PHL) | Horan, K.; N.L.; Seemann, T.; Sherry |
| EPI_ISL_9301675 | N Health Hat Yai | Prince of Songkhla University | Anusara Wongkotsila; Chanon Kongkamon; Pongsakorn Choochuen; Samonrapat Surasombatpattana; Sarunyou Chusri; Surakameth Mahasirimongkol; Surasak Sangkhathat; Thammasin Ingviya; Thanit Sila; Virasakdi Chongsuvivatwong; Wison Laochareonsuk |
| EPI_ISL_9301625, EPI_ISL_9301626, EPI_ISL_9301627, EPI_ISL_9301629, EPI_ISL_9301630 | PR Medical Laboratory. | Prince of Songkhla University | Anusara Wongkotsila; Chanon Kongkamon; Pongsakorn Choochuen; Samonrapat Surasombatpattana; Sarunyou Chusri; Surakameth Mahasirimongkol; Surasak Sangkhathat; Thammasin Ingviya; Thanit Sila; Virasakdi Chongsuvivatwong; Wison Laochareonsuk |
| EPI_ISL_9301585, EPI_ISL_9301586 | Pattani Hospital | Prince of Songkhla University | Anusara Wongkotsila; Chanon Kongkamon; Pongsakorn Choochuen; Samonrapat Surasombatpattana; Sarunyou Chusri; Surakameth Mahasirimongkol; Surasak Sangkhathat; Thammasin Ingviya; Thanit Sila; Virasakdi Chongsuvivatwong; Wison Laochareonsuk |
| EPI_ISL_9301589, EPI_ISL_9301590, EPI_ISL_9301591, EPI_ISL_9301593, EPI_ISL_9301595, EPI_ISL_9301631, EPI_ISL_9301632 | | | |
| see above | RAJYINDEE Hospital | Prince of Songkhla University | Anusara Wongkotsila; Chanon Kongkamon; Pongsakorn Choochuen; Samonrapat Surasombatpattana; Sarunyou Chusri; Surakameth Mahasirimongkol; Surasak Sangkhathat; Thammasin Ingviya; Thanit Sila; Virasakdi Chongsuvivatwong; Wison Laochareonsuk |
| EPI_ISL_9287781 | RS Kartika Pulomas | National Institute of Health Research and Development | Arie Ardiansyah Nugraha; Fajar Nur Sulistiyohadi; Hana Apsari Pawestri; Hartanti Dian Ikawati; Kartika Dewi Puspa; Nelly Puspandari; Nur Ika Hariastuti; Putri Widia; Subangkit; Vivi Setiawaty |
| EPI_ISL_9287817 | RSUP Persahabatan | National Institute of Health Research and Development | Arie Ardiansyah Nugraha; Fajar Nur Sulistiyohadi; Hana Apsari Pawestri; Hartanti Dian Ikawati; Kartika Dewi Puspa; Nelly Puspandari; Nur Ika Hariastuti; Putri Widia; Subangkit; Vivi Setiawaty |
| EPI_ISL_9301655, EPI_ISL_9301656 | Rattaphum Hospital | Prince of Songkhla University | Anusara Wongkotsila; Chanon Kongkamon; Pongsakorn Choochuen; Samonrapat Surasombatpattana; Sarunyou Chusri; Surakameth Mahasirimongkol; Surasak Sangkhathat; Thammasin Ingviya; Thanit Sila; Virasakdi Chongsuvivatwong; Wison Laochareonsuk |
| EPI_ISL_9290286, EPI_ISL_9290322, EPI_ISL_9293221 | Respiratory Virus Unit, Microbiology Services Colindale, Public Health England | COVID-19 Genomics UK (COG-UK) Consortium | PHE Covid Sequencing Team |
| EPI_ISL_9298655 | Rumah Sakit Royal Taruma | Genomik Solidaritas Indonesia Laboratorium | Annisa Muthiah Sukirman; Anuraj Shankar; Ariel Pradipta; Carissa Sintca Wijaya; Gracia Felias Enos Korompis; Liman; Louisa Markus; Meutia Ayuputeri Kumaheri; Reinhart Gabriel; Riana; Stefanus Bernard |
| EPI_ISL_9301652, EPI_ISL_9301653, EPI_ISL_9301654 | SOMDEJPRABOROMRACHINEENART NATAWEE HOSPITAL. | Prince of Songkhla University | Anusara Wongkotsila; Chanon Kongkamon; Pongsakorn Choochuen; Samonrapat Surasombatpattana; Sarunyou Chusri; Surakameth Mahasirimongkol; Surasak Sangkhathat; Thammasin Ingviya; Thanit Sila; Virasakdi Chongsuvivatwong; Wison Laochareonsuk |
| EPI_ISL_9301597 | Sikarin Hospital | Prince of Songkhla University | Anusara Wongkotsila; Chanon Kongkamon; Pongsakorn Choochuen; Samonrapat Surasombatpattana; Sarunyou Chusri; Surakameth Mahasirimongkol; Surasak Sangkhathat; Thammasin Ingviya; Thanit Sila; Virasakdi Chongsuvivatwong; Wison Laochareonsuk |
| EPI_ISL_9301640, EPI_ISL_9301641, EPI_ISL_9301642, EPI_ISL_9301643, EPI_ISL_9301644 | Siroros Hospital | Prince of Songkhla University | Anusara Wongkotsila; Chanon Kongkamon; Pongsakorn Choochuen; Samonrapat Surasombatpattana; Sarunyou Chusri; Surakameth Mahasirimongkol; Surasak Sangkhathat; Thammasin Ingviya; Thanit Sila; Virasakdi Chongsuvivatwong; Wison Laochareonsuk |
| EPI_ISL_9301610, EPI_ISL_9301611, EPI_ISL_9301612, EPI_ISL_9301613, EPI_ISL_9301614, EPI_ISL_9301615, EPI_ISL_9301618, EPI_ISL_9301620, EPI_ISL_9301622, EPI_ISL_9301623 | | | |
| see above | Songkhla Hospital | Prince of Songkhla University | Anusara Wongkotsila; Chanon Kongkamon; Pongsakorn Choochuen; Samonrapat Surasombatpattana; Sarunyou Chusri; Surakameth Mahasirimongkol; Surasak Sangkhathat; Thammasin Ingviya; Thanit Sila; Virasakdi Chongsuvivatwong; Wison Laochareonsuk |
| EPI_ISL_9301687, EPI_ISL_9301688, EPI_ISL_9301689, EPI_ISL_9301690, EPI_ISL_9301694 | Songklanagarind hospital | Prince of Songkhla University | Anusara Wongkotsila; Chanon Kongkamon; Pongsakorn Choochuen; Samonrapat Surasombatpattana; Sarunyou Chusri; Surakameth Mahasirimongkol; Surasak Sangkhathat; Thammasin Ingviya; Thanit Sila; Virasakdi Chongsuvivatwong; Wison Laochareonsuk |
| EPI_ISL_9301598, EPI_ISL_9301599, EPI_ISL_9301600, EPI_ISL_9301601, EPI_ISL_9301602, EPI_ISL_9301603, EPI_ISL_9301604, EPI_ISL_9301605, EPI_ISL_9301607, EPI_ISL_9301658, EPI_ISL_9301659, EPI_ISL_9301661, EPI_ISL_9301662, EPI_ISL_9301663 | | | |
| see above | Su-ngai Kolok Hospital | Prince of Songkhla University | Anusara Wongkotsila; Chanon Kongkamon; Pongsakorn Choochuen; Samonrapat Surasombatpattana; Sarunyou Chusri; Surakameth Mahasirimongkol; Surasak Sangkhathat; Thammasin Ingviya; Thanit Sila; Virasakdi Chongsuvivatwong; Wison Laochareonsuk |
| EPI_ISL_9300770 | ULSNE - Braganca | Instituto Nacional de Saude (INSA) | Borges et al |
| EPI_ISL_9301868, EPI_ISL_9301875, EPI_ISL_9301881, EPI_ISL_9301903 | UW Virology Lab | UW Virology Lab | Alexander Greninger; Hong Xie; Isabel Arnould; Keith R Jerome; Meei-Li Huang; Pavitra Roychoudhury; Pooneh Hajian; Ricardo Perez; Saraswathi Sathees; Sean Ellis; Seffir T. Wendm; Shah Mohamed Bakhash |

We gratefully acknowledge the following Authors from the Originating laboratories responsible for obtaining the specimens, as well as the Submitting laboratories where the genome data were generated and shared via GISAID, on which this research is based.

All Submitters of data may be contacted directly via www.gisaid.org

Authors are sorted alphabetically.

| Accession ID | Originating Laboratory | Submitting Laboratory | Authors |
|---|---|---|---|
| EPI_ISL_9324346, EPI_ISL_9324347, EPI_ISL_9324348 | AIRPORT CLINIC | inStem, NCBS-INSACOG | Aswin SaiNarain; Dasaradhi Palakodeti; Uma Ramakrishnan |
| EPI_ISL_9324342, EPI_ISL_9324344 | ASTER LABS SARJAPUR ROAD | inStem, NCBS-INSACOG | Aswin SaiNarain; Dasaradhi Palakodeti; Uma Ramakrishnan |
| EPI_ISL_9324340 | ASTER R V | inStem, NCBS-INSACOG | Aswin SaiNarain; Dasaradhi Palakodeti; Uma Ramakrishnan |
| EPI_ISL_9331874, EPI_ISL_9332007 | Aegis Sciences Corporation | Centers for Disease Control and Prevention Division of Viral Diseases, Pathogen Discovery | Alec Vest; Benjamin Rambo-Martin; Christopher Gulvick; Clinton Paden; Cyndi Clark; Dakota Howard; Dhwani Batra; Dillon Nall; Duncan MacCannell; Erisa Sula; Ethan Sanders; Holly Houdeshell; Jason Caravas; Kristine Lacek; Matthew Hardison; Matthew Schmerer; Ola Kvalvaag; Patrick Campbell; Peter Cook; Rob Case; Scott Sammons; Shatavia Morrison; Shaun Westlund; Tymeckia Kendall; Victoria Caban Figueroa; Vikramsinha Ghorpade; Yvette Unoarumhi |
| EPI_ISL_9329641, EPI_ISL_9329651, EPI_ISL_9329672, EPI_ISL_9329675, EPI_ISL_9329790, EPI_ISL_9329812, EPI_ISL_9329821, EPI_ISL_9329827, EPI_ISL_9329839, EPI_ISL_9329850, EPI_ISL_9329858, EPI_ISL_9329862, EPI_ISL_9329863, EPI_ISL_9329864, EPI_ISL_9329865, EPI_ISL_9329870, EPI_ISL_9329874, EPI_ISL_9329880 | | | |
| see above | BANGALORE BAPTIST HOSPITAL | National Center for Biological Sciences, TIFR - Rockefeller Foundation | Awadhesh Pandit; Bhagyashree Madhav Shelar; Darshan Sreenivas; Dimple Notani; Kathirvel Kandasamy; Lakshminarayanan CP; Satyajit Mayor; Uma Ramakrishnan |
| EPI_ISL_9336257, EPI_ISL_9336265, EPI_ISL_9336266, EPI_ISL_9336279, EPI_ISL_9336284, EPI_ISL_9336285, EPI_ISL_9336286, EPI_ISL_9336297, EPI_ISL_9336298, EPI_ISL_9336302, EPI_ISL_9336307, EPI_ISL_9336311, EPI_ISL_9336323, EPI_ISL_9336324, EPI_ISL_9336325, EPI_ISL_9336326, EPI_ISL_9336327, EPI_ISL_9336332, EPI_ISL_9336333, EPI_ISL_9336335, EPI_ISL_9336337 | | | |
| see above | CSIR-NEERI, Nagpur Covid-19 Testing Lab | CSIR-NEERI, Nagpur Covid-19 Testing Lab | Krishna Khairnar et al. |
| EPI_ISL_9324334 | DASARHALLI FPEC ASTER | inStem, NCBS-INSACOG | Aswin SaiNarain; Dasaradhi Palakodeti; Uma Ramakrishnan |
| EPI_ISL_9325605 | Department of Microbiology, University Hospital in Pilsen | Department of Microbiology, University Hospital in Pilsen | Jaroslav Hrabak; Katerina Chudejova; Katerina Vlkova; Petra Kocova; Vendula Heringova |
| EPI_ISL_9332008 | Fulgent Genetics | Centers for Disease Control and Prevention Division of Viral Diseases, Pathogen Discovery | Becky Tsai; Benafsh Sapra; Benjamin Rambo-Martin; Christopher Gulvick; Clinton Paden; Dakota Howard; Dhwani Batra; Doreen Ng; Duncan MacCannell; Erisa Sula; Harry Gao; James Xie; Jason Caravas; John Gao; Joseph Fierro; Kristine Lacek; Matthew Schmerer; Mickey Li; Peter Cook; Scott Sammons; Shatavia Morrison; Tymeckia Kendall; Victoria Caban Figueroa; Yan Meng; Yvette Unoarumhi |
| EPI_ISL_9324317, EPI_ISL_9324321 | HOME COLLECTION | inStem, NCBS-INSACOG | Aswin SaiNarain; Dasaradhi Palakodeti; Uma Ramakrishnan |
| EPI_ISL_9325921 | Hospital Santa Bárbara | Hospital General Universitario de Ciudad Real | Cristina Colmenarejo; José Martínez-Alarcón; Lidia García-Agudo; Marta Torres-Narbona; Soledad Illescas Fernández-Bermejo |
| EPI_ISL_9347062 | Houston Methodist Hospital | Houston Methodist Hospital | Akanksha Batajoo; Ilya J. Finkelstein; James J. Davis; Jessica Cambric; Jimmy Gollihar; Kristina Reppond; Madison N. Shyer; Marcus Nguyen; Matthew Ojeda Saavedra; Paul A. Christensen; Randall J. Olsen; Rashi Thakur; Regan Mangham; Richard Snehall; Robert Olson; Ryan Gadd; S. Wesley Long; Sindy Pena; and James M. Musser |
| EPI_ISL_9322394 | Hrvatski zavod za javno zdravstvo | Hrvatski zavod za javno zdravstvo | Anita Jurić; Dragan Jurić; Irena Tabain; Ivana Ferenčak; Josipa Kuzle |
| EPI_ISL_9325315 | Invenimus AG | Institute of Medical Virology, University of Zurich | Alexandra Trkola; Annette Audigé; Catharine Aquino; Cyril Shah; Daniel Ehrsam; Gabriela Ziltener; Guido Bloemberg; Hubert Rehrauer; Isabel Stürmer; Joel Wirz; Jon Huder; Jürg Böni; Kevin Steiner; Maria Grünberg; Maryam Zaheri; Michael Huber; Riccarda Capaul; Stefan Schmutz; Verena Kufner; Weihong Qi |
| EPI_ISL_9306076 | Israel Central Virology laboratory | Israel Central Virology laboratory | Danit Sofer; Efrat Dahan Bucris; Efrat Glick-Saar; Ella Mendelson; Gideon Rechavi; Hagar Morad; Michal Mandelboim; Miranda Geva; Neta Zuckerman; Or Zilbertzan; Oran Erster; Orna Mor |
| EPI_ISL_9306388 | Jerusalem Central Laboratories | Israel Central Virology laboratory | Danit Sofer; Efrat Dahan Bucris; Ella Mendelson; Evan Nachum; Hagar Morad; Julia Vainer; Maya Davidovich; Michal Mandelboim; Michal Zak; Miranda Geva; Neta Zuckerman; Or Zilbertzan; Oran Erster; Orna Mor; Rona Grossman |
| EPI_ISL_9325288 | Kantonsspital Aarau | Institute of Medical Virology, University of Zurich | Alexandra Trkola; Annette Audigé; Catharine Aquino; Cyril Shah; Daniel Ehrsam; Gabriela Ziltener; Guido Bloemberg; Hubert Rehrauer; Isabel Stürmer; Joel Wirz; Jon Huder; Jürg Böni; Kevin Steiner; Maria Grünberg; Maryam Zaheri; Michael Huber; Riccarda Capaul; Stefan Schmutz; Verena Kufner; Weihong Qi |
| EPI_ISL_9324292, EPI_ISL_9324293 | Kims Hubli | inStem, NCBS-INSACOG | Aswin SaiNarain; Dasaradhi Palakodeti; Uma Ramakrishnan |
| EPI_ISL_9344681 | LABORATORIO DE FRANCA - FRANCA - 2064154 | Instituto Butantan | Antonio Jorge Martins; Claudia Renata dos Santos Barros; David Schlesinger; Debora Botequio Moretti; Dimas Tadeu Covas; Elaine Cristina Marqueze; Elaine Vieira Santos; Evandra Strazza Rodrigues; Heidge Fukumasu; Jayme Augusto de Souza-Neto; José Salvatore Leister Patané; Luiz Alcantara; Luiz Lehmann Coutinho; Maria Carolina Elias; Maurício Lacerda Nogueira; Rafael dos Santos Bezerra; Raul Machado Neto; Rejane Maria Tommasini Grotto; Ricardo Haddad; Sandra Coccuzzo Sampaio Vessoni; Simone Kashima; Svetoslav Nanev Slavov; Vincent Louis Viala |
| EPI_ISL_9321624, EPI_ISL_9321625, EPI_ISL_9321627, EPI_ISL_9321630, EPI_ISL_9321632, EPI_ISL_9321633, EPI_ISL_9321635, EPI_ISL_9321637, EPI_ISL_9321639, EPI_ISL_9321643, EPI_ISL_9321647, EPI_ISL_9321648, EPI_ISL_9321651, EPI_ISL_9321653, EPI_ISL_9321659, EPI_ISL_9321661, EPI_ISL_9321662, EPI_ISL_9321664, EPI_ISL_9321672, EPI_ISL_9321673, EPI_ISL_9321674, EPI_ISL_9321675, EPI_ISL_9321678, EPI_ISL_9321683, EPI_ISL_9321685, EPI_ISL_9321686, EPI_ISL_9321689, EPI_ISL_9321691, EPI_ISL_9321692, EPI_ISL_9321693, EPI_ISL_9321696, EPI_ISL_9321698, EPI_ISL_9321699, EPI_ISL_9321700, EPI_ISL_9321701, EPI_ISL_9321704, EPI_ISL_9321705, EPI_ISL_9321709, EPI_ISL_9321713 | | | |
| see above | Lifebrain Covid Labor GmbH | Lifebrain Covid Labor GmbH | Alexandra Wagner; Anna Edermayr; Filip Sima |
| EPI_ISL_9315131, EPI_ISL_9315229 | Lighthouse Lab in Milton Keynes | Wellcome Sanger Institute for the COVID-19 Genomics UK (COG-UK) Consortium | Cordelia Langford; David K. Jackson; Dominic Kwiatkowski; Ewan Harrison; Ian Johnston; Jeffrey Barrett; John Sillitoe on behalf of the Wellcome Sanger Institute COVID-19 Surveillance Team; Roberto Amato; Sonia Goncalves; The Lighthouse Lab in Milton Keynes and Alex Alderton |
| EPI_ISL_9314579 | Lighthouse Laboratory Plymouth | Wellcome Sanger Institute for the COVID-19 Genomics UK (COG-UK) Consortium | Cordelia Langford; David K. Jackson; Dominic Kwiatkowski; Ewan Harrison; Ian Johnston; Jeffrey Barrett; John Sillitoe on behalf of the Wellcome Sanger Institute COVID-19 Surveillance Team; Lighthouse Laboratory Plymouth and Alex Alderton; Roberto Amato; Sonia Goncalves |
| EPI_ISL_9324289 | MAHALAXMI LAYOUT | inStem, NCBS-INSACOG | Aswin SaiNarain; Dasaradhi Palakodeti; Uma Ramakrishnan |
| EPI_ISL_9322383, EPI_ISL_9322384, EPI_ISL_9322385, EPI_ISL_9322386, EPI_ISL_9322387 | Nastavni zavod za javno zdravstvo "Andrija Štampar" | Hrvatski zavod za javno zdravstvo | Anita Jurić; Dragan Jurić; Irena Tabain; Ivana Ferenčak; Josipa Kuzle |
| EPI_ISL_9322388, EPI_ISL_9322389 | Opća bolnica Bjelovar | Hrvatski zavod za javno zdravstvo | Anita Jurić; Dragan Jurić; Irena Tabain; Ivana Ferenčak; Josipa Kuzle |
| EPI_ISL_9303471, EPI_ISL_9303482, EPI_ISL_9303495, EPI_ISL_9303506, EPI_ISL_9303522, EPI_ISL_9303527, EPI_ISL_9303534, EPI_ISL_9303572, EPI_ISL_9303574, EPI_ISL_9303575, EPI_ISL_9303576, EPI_ISL_9303586, EPI_ISL_9303587, EPI_ISL_9303588, EPI_ISL_9303589, EPI_ISL_9303592, EPI_ISL_9303595, EPI_ISL_9303596, EPI_ISL_9303597, EPI_ISL_9303598, EPI_ISL_9303601, EPI_ISL_9303602, EPI_ISL_9303603, EPI_ISL_9303606, EPI_ISL_9303607, EPI_ISL_9303608, EPI_ISL_9303616, EPI_ISL_9303622, EPI_ISL_9303633, EPI_ISL_9303634, EPI_ISL_9303645, EPI_ISL_9303648, EPI_ISL_9303649, EPI_ISL_9303650, EPI_ISL_9303651, EPI_ISL_9303653, EPI_ISL_9303654, EPI_ISL_9303661, EPI_ISL_9303666 | | | |
| see above | SARS-CoV-2 testing team, National Institute of Infectious Diseases | Pathogen Genomics Center, National Institute of Infectious Diseases | Hazuka Y Furihata; Kentaro Itokawa; Makoto Kuroda; Masanori Hashino; Masumichi Saito; Naomi Nojiri; Nozomu Hanaoka; Rina Tanaka; Tsuguto Fujimoto; Tsuyoshi Sekizuka |
| EPI_ISL_9324272 | SYSTEMATIC LABS KOTHUNOOR FPEC | inStem, NCBS-INSACOG | Aswin SaiNarain; Dasaradhi Palakodeti; Uma Ramakrishnan |
| EPI_ISL_9346803, EPI_ISL_9346818 | Seattle Flu Study | Seattle Flu Study | Amanda Adler; Barry R. Lutz; Benjamin Pelle; Caitlin R. Wolf; Chris D. Frazar; Deborah A. Nickerson; Elisabeth Brandstetter; Erica Ryke; Helen Y. Chu; Janet A. Englund; Jay Shendure; Jover Lee; Kairsten Fay; Kirsten Lacombe; Lea M. Starita; Mark J. Rieder; Matthew Richardson; Matthew Thompson; Melissa Truong; Michael Boeckh; Michael Famulare; Misja Ilcisin; Peter D. Han; Thomas R. Sibley; Trevor Bedford |
| EPI_ISL_9324984, EPI_ISL_9324985, EPI_ISL_9325099, EPI_ISL_9325220, EPI_ISL_9325222, EPI_ISL_9325232, EPI_ISL_9325304 | | | |
| see above | Stadtspital Waid | Institute of Medical Virology, University of Zurich | Alexandra Trkola; Annette Audigé; Catharine Aquino; Cyril Shah; Daniel Ehrsam; Gabriela Ziltener; Guido Bloemberg; Hubert Rehrauer; Isabel Stürmer; Joel Wirz; Jon Huder; Jürg Böni; Kevin Steiner; Maria Grünberg; Maryam Zaheri; Michael Huber; Riccarda Capaul; Stefan Schmutz; Verena Kufner; Weihong Qi |
| EPI_ISL_9332967 | The Ohio State University Applied Microbiology Services Laboratory | The Ohio State University Applied Microbiology Services Laboratory | Seth A. Faith PhD |
| EPI_ISL_9303742 | Tokyo Medical and Dental University | Tokyo Medical and Dental University | Akinori Kimura; Hiroaki Takeuchi; Kousuke Tanimoto; Yoko Nukui; Yoshiaki Gu; Yukie Tanaka |
| EPI_ISL_9324824, EPI_ISL_9324825, EPI_ISL_9324826, EPI_ISL_9324871, EPI_ISL_9324962, EPI_ISL_9324963, EPI_ISL_9324968, EPI_ISL_9324969, EPI_ISL_9324970, EPI_ISL_9324971, EPI_ISL_9325091, EPI_ISL_9325092, EPI_ISL_9325093, EPI_ISL_9325094, EPI_ISL_9325096, EPI_ISL_9325097, EPI_ISL_9325098, EPI_ISL_9325100, EPI_ISL_9325101, EPI_ISL_9325102, EPI_ISL_9325103, EPI_ISL_9325104, EPI_ISL_9325105, EPI_ISL_9325106, EPI_ISL_9325107, EPI_ISL_9325109, EPI_ISL_9325201, EPI_ISL_9325202, EPI_ISL_9325203, EPI_ISL_9325204, EPI_ISL_9325205, EPI_ISL_9325206, EPI_ISL_9325210, EPI_ISL_9325211, EPI_ISL_9325212, EPI_ISL_9325214, EPI_ISL_9325215, EPI_ISL_9325216, EPI_ISL_9325218, EPI_ISL_9325219, EPI_ISL_9325512, EPI_ISL_9325513, EPI_ISL_9325515, EPI_ISL_9325516, EPI_ISL_9325517, EPI_ISL_9325518, EPI_ISL_9325519, EPI_ISL_9325520, EPI_ISL_9325521, EPI_ISL_9325525 | | | |
| see above | Universität Zürich | Institute of Medical Virology, University of Zurich | Alexandra Trkola; Annette Audigé; Catharine Aquino; Cyril Shah; Daniel Ehrsam; Gabriela Ziltener; Guido Bloemberg; Hubert Rehrauer; Isabel Stürmer; Joel Wirz; Jon Huder; Jürg Böni; Kevin Steiner; Maria Grünberg; Maryam Zaheri; Michael Huber; Riccarda Capaul; Stefan Schmutz; Verena Kufner; Weihong Qi |
| EPI_ISL_9324996, EPI_ISL_9324998, EPI_ISL_9324999, EPI_ISL_9325228, EPI_ISL_9325229, EPI_ISL_9325230, EPI_ISL_9325234, EPI_ISL_9325237, EPI_ISL_9325305, EPI_ISL_9325309, EPI_ISL_9325310, EPI_ISL_9325311, EPI_ISL_9325312, EPI_ISL_9325313, EPI_ISL_9325505, EPI_ISL_9325506, EPI_ISL_9325507 | | | |
| see above | Universitäts-Kinderspital Zürich | Institute of Medical Virology, University of Zurich | Alexandra Trkola; Annette Audigé; Catharine Aquino; Cyril Shah; Daniel Ehrsam; Gabriela Ziltener; Guido Bloemberg; Hubert Rehrauer; Isabel Stürmer; Joel Wirz; Jon Huder; Jürg Böni; Kevin Steiner; Maria Grünberg; Maryam Zaheri; Michael Huber; Riccarda Capaul; Stefan Schmutz; Verena Kufner; Weihong Qi |
| EPI_ISL_9324974, EPI_ISL_9324975, EPI_ISL_9324976, EPI_ISL_9324977, EPI_ISL_9324978, EPI_ISL_9324979, EPI_ISL_9324986, EPI_ISL_9324987, EPI_ISL_9324988, EPI_ISL_9324989, EPI_ISL_9324990, EPI_ISL_9324991, EPI_ISL_9324992, EPI_ISL_9324993, EPI_ISL_9324994, EPI_ISL_9324995, EPI_ISL_9325002, EPI_ISL_9325003, EPI_ISL_9325088, EPI_ISL_9325089, EPI_ISL_9325090, EPI_ISL_9325111, EPI_ISL_9325113, EPI_ISL_9325114, EPI_ISL_9325116, EPI_ISL_9325117, EPI_ISL_9325207, EPI_ISL_9325209, EPI_ISL_9325223, EPI_ISL_9325226, EPI_ISL_9325227, EPI_ISL_9325231, EPI_ISL_9325233, EPI_ISL_9325238, EPI_ISL_9325240, EPI_ISL_9325241, EPI_ISL_9325242, EPI_ISL_9325243, EPI_ISL_9325244, EPI_ISL_9325279, EPI_ISL_9325280, EPI_ISL_9325281, EPI_ISL_9325282, EPI_ISL_9325283, EPI_ISL_9325285, EPI_ISL_9325286, EPI_ISL_9325287, EPI_ISL_9325289, EPI_ISL_9325290, EPI_ISL_9325291, EPI_ISL_9325292, EPI_ISL_9325293, EPI_ISL_9325294, EPI_ISL_9325295, EPI_ISL_9325296, EPI_ISL_9325298, EPI_ISL_9325299, EPI_ISL_9325300, EPI_ISL_9325314, EPI_ISL_9325503, EPI_ISL_9325509, EPI_ISL_9325510, EPI_ISL_9325511 | | | |
| see above | UniversitätsSpital Zürich | Institute of Medical Virology, University of Zurich | Alexandra Trkola; Annette Audigé; Catharine Aquino; Cyril Shah; Daniel Ehrsam; Gabriela Ziltener; Guido Bloemberg; Hubert Rehrauer; Isabel Stürmer; Joel Wirz; Jon Huder; Jürg Böni; Kevin Steiner; Maria Grünberg; Maryam Zaheri; Michael Huber; Riccarda Capaul; Stefan Schmutz; Verena Kufner; Weihong Qi |
| EPI_ISL_9322465, EPI_ISL_9322466, | Zavod za javno zdravstvo | Hrvatski zavod za javno zdravstvo | Anita Jurić; Dragan Jurić; Irena Tabain; Ivana Ferenčak; Josipa Kuzle |

| | |
|---|---|
| EPI_ISL_9322566, EPI_ISL_9322568,<br>EPI_ISL_9322569, EPI_ISL_9322570 | Šibensko- Kninske županije |

We gratefully acknowledge the following Authors from the Originating laboratories responsible for obtaining the specimens, as well as the Submitting laboratories where the genome data were generated and shared via GISAID, on which this research is based.

All Submitters of data may be contacted directly via www.gisaid.org

Authors are sorted alphabetically.

| Accession ID | Originating Laboratory | Submitting Laboratory | Authors |
| --- | --- | --- | --- |
| EPI_ISL_8439646 | ROB FERREIRA LABORATORY | National Institute for Communicable Diseases of the National Health Laboratory Service | Amoako DG; Bhiman JN; Everatt J; Ismail A; Kekana D; Mahlangu B; Mnguni A; Mohale T; Ntuli N; Scheepers C; Wolter N |

We gratefully acknowledge the following Authors from the Originating laboratories responsible for obtaining the specimens, as well as the Submitting laboratories where the genome data were generated and shared via GISAID, on which this research is based.

All Submitters of data may be contacted directly via www.gisaid.org

Authors are sorted alphabetically.

Acknowledgement EPI_SET Identifier: EPI_SET_20220527kq

| Accession ID | Originating Laboratory | Submitting Laboratory | Authors |
|---|---|---|---|
| EPI_ISL_8476913 | Child Health Research Foundation | Child Health Research Foundation | CHRF Bangladesh Genomics Team |
| EPI_ISL_8452449, EPI_ISL_8452453, EPI_ISL_8452455, EPI_ISL_8452456, EPI_ISL_8452458, EPI_ISL_8452459, EPI_ISL_8452464, EPI_ISL_8452465, EPI_ISL_8452466, EPI_ISL_8452467, EPI_ISL_8452468, EPI_ISL_8452469, EPI_ISL_8452470, EPI_ISL_8452471, EPI_ISL_8452472, EPI_ISL_8452473, EPI_ISL_8452478, EPI_ISL_8452480, EPI_ISL_8452492, EPI_ISL_8452493, EPI_ISL_8452494, EPI_ISL_8452496, EPI_ISL_8452497, EPI_ISL_8452498, EPI_ISL_8452500, EPI_ISL_8452501, EPI_ISL_8452502, EPI_ISL_8452506, EPI_ISL_8452507, EPI_ISL_8452508 | | | |
| see above | Gandhi Medical College and Hospital (GMCH), Secunderabad | NIV Influenza | D.R.Manisha Rani; Dr.G.Sushma Rajya Lakshmi; Dr.K.Nagamani; Dr.Sunitha Pakalapaty |
| EPI_ISL_8445176, EPI_ISL_8445224, EPI_ISL_8445225, EPI_ISL_8445227, EPI_ISL_8445229, EPI_ISL_8445230, EPI_ISL_8445234, EPI_ISL_8445236, EPI_ISL_8445237, EPI_ISL_8445238, EPI_ISL_8445239, EPI_ISL_8445240, EPI_ISL_8445241, EPI_ISL_8445242, EPI_ISL_8445248 | | | |
| see above | ICMR-National Institute of Virology - INSACOG | NIV Influenza | Dr. Varsha Potdar and NIC Team |
| EPI_ISL_8435250, EPI_ISL_8435282, EPI_ISL_8449933, EPI_ISL_8450019 | National Public Health Laboratory, National Centre for Infectious Diseases | National Public Health Laboratory, National Centre for Infectious Diseases | Benny Yeo; Grace Ngan; Lin Cui; Raymond Tzer Pin Lin; Royce Ang; Samuel Loo; Zhenyang Zhou |

We gratefully acknowledge the following Authors from the Originating laboratories responsible for obtaining the specimens, as well as the Submitting laboratories where the genome data were generated and shared via GISAID, on which this research is based.

All Submitters of data may be contacted directly via www.gisaid.org

Authors are sorted alphabetically.

| Accession ID | Originating Laboratory | Submitting Laboratory | Authors |
| --- | --- | --- | --- |
| EPI_ISL_8457787, EPI_ISL_8457848, EPI_ISL_8457866, EPI_ISL_8457869, EPI_ISL_8457900, EPI_ISL_8457968, EPI_ISL_8458005, EPI_ISL_8458066, EPI_ISL_8458091, EPI_ISL_8458107, EPI_ISL_8458112, EPI_ISL_8458115, EPI_ISL_8458202, EPI_ISL_8458234, EPI_ISL_8458251, EPI_ISL_8458287, EPI_ISL_8458295, EPI_ISL_8458315, EPI_ISL_8458335, EPI_ISL_8458358, EPI_ISL_8458365, EPI_ISL_8458395, EPI_ISL_8458401, EPI_ISL_8458441, EPI_ISL_8458499, EPI_ISL_8458519, EPI_ISL_8458533, EPI_ISL_8458549, EPI_ISL_8458581, EPI_ISL_8458697, EPI_ISL_8458709, EPI_ISL_8458712, EPI_ISL_8458719, EPI_ISL_8458720, EPI_ISL_8458725, EPI_ISL_8458805, EPI_ISL_8458827, EPI_ISL_8458882, EPI_ISL_8458893, EPI_ISL_8458949, EPI_ISL_8458987, EPI_ISL_8459001, EPI_ISL_8459015, EPI_ISL_8459054, EPI_ISL_8459102, EPI_ISL_8459153, EPI_ISL_8459200, EPI_ISL_8459211, EPI_ISL_8459223, EPI_ISL_8459226, EPI_ISL_8459231, EPI_ISL_8459237, EPI_ISL_8459273, EPI_ISL_8459292, EPI_ISL_8459303, EPI_ISL_8459314, EPI_ISL_8459318, EPI_ISL_8459340, EPI_ISL_8459365, EPI_ISL_8459409, EPI_ISL_8459443, EPI_ISL_8459450, EPI_ISL_8459452, EPI_ISL_8459457, EPI_ISL_8459458, EPI_ISL_8459460, EPI_ISL_8459462, EPI_ISL_8459468, EPI_ISL_8459472, EPI_ISL_8459480, EPI_ISL_8459496, EPI_ISL_8459500, EPI_ISL_8459553, EPI_ISL_8459569, EPI_ISL_8459616, EPI_ISL_8459644, EPI_ISL_8459664, EPI_ISL_8459728, EPI_ISL_8459737, EPI_ISL_8459747, EPI_ISL_8459753, EPI_ISL_8459775, EPI_ISL_8459800, EPI_ISL_8459852, EPI_ISL_8459872, EPI_ISL_8459884, EPI_ISL_8459932, EPI_ISL_8459947, EPI_ISL_8459957, EPI_ISL_8459972, EPI_ISL_8460062, EPI_ISL_8460088, EPI_ISL_8460097, EPI_ISL_8460119, EPI_ISL_8460132, EPI_ISL_8460139, EPI_ISL_8460165, EPI_ISL_8460171, EPI_ISL_8460178, EPI_ISL_8460189, EPI_ISL_8460312, EPI_ISL_8460322, EPI_ISL_8460345, EPI_ISL_8461029, EPI_ISL_8461911, EPI_ISL_8462122, EPI_ISL_8462203, EPI_ISL_8462224, EPI_ISL_8462240, EPI_ISL_8462259, EPI_ISL_8462279, EPI_ISL_8462434, EPI_ISL_8462475, EPI_ISL_8462489, EPI_ISL_8462517, EPI_ISL_8462580, EPI_ISL_8462585, EPI_ISL_8462604, EPI_ISL_8462611, EPI_ISL_8462613, EPI_ISL_8462614, EPI_ISL_8462622, EPI_ISL_8462628, EPI_ISL_8462649, EPI_ISL_8462658, EPI_ISL_8462661, EPI_ISL_8462674, EPI_ISL_8462686, EPI_ISL_8462688, EPI_ISL_8462701, EPI_ISL_8462703, EPI_ISL_8462709, EPI_ISL_8462717, EPI_ISL_8462724, EPI_ISL_8462730, EPI_ISL_8462732, EPI_ISL_8462739, EPI_ISL_8462760, EPI_ISL_8462772, EPI_ISL_8462797, EPI_ISL_8462811, EPI_ISL_8462812, EPI_ISL_8462817, EPI_ISL_8462818, EPI_ISL_8462824, EPI_ISL_8462829, EPI_ISL_8462831, EPI_ISL_8462842, EPI_ISL_8462878, EPI_ISL_8462892, EPI_ISL_8462897, EPI_ISL_8462898, EPI_ISL_8462899, EPI_ISL_8462902, EPI_ISL_8462904, EPI_ISL_8462908, EPI_ISL_8462912, EPI_ISL_8462929, EPI_ISL_8462930, EPI_ISL_8462935, EPI_ISL_8462956, EPI_ISL_8462972, EPI_ISL_8462979, EPI_ISL_8463053, EPI_ISL_8463055, EPI_ISL_8463061, EPI_ISL_8463082, EPI_ISL_8463087, EPI_ISL_8463091, EPI_ISL_8463097, EPI_ISL_8463101, EPI_ISL_8463111, EPI_ISL_8463112, EPI_ISL_8463119, EPI_ISL_8463120, EPI_ISL_8463121, EPI_ISL_8463124, EPI_ISL_8463128, EPI_ISL_8463130, EPI_ISL_8463136, EPI_ISL_8463143, EPI_ISL_8463151, EPI_ISL_8463153, EPI_ISL_8463165, EPI_ISL_8463182, EPI_ISL_8463199, EPI_ISL_8463207, EPI_ISL_8463215, EPI_ISL_8463230, EPI_ISL_8463243, EPI_ISL_8463261, EPI_ISL_8463279, EPI_ISL_8463289, EPI_ISL_8463290, EPI_ISL_8463292, EPI_ISL_8463296, EPI_ISL_8463302, EPI_ISL_8463303, EPI_ISL_8463306, EPI_ISL_8463324, EPI_ISL_8463328, EPI_ISL_8465069, EPI_ISL_8465070, EPI_ISL_8465082, EPI_ISL_8465083, EPI_ISL_8465088, EPI_ISL_8465089, EPI_ISL_8465090, EPI_ISL_8465095, EPI_ISL_8465096, EPI_ISL_8465100, EPI_ISL_8465119, EPI_ISL_8465121, EPI_ISL_8465123, EPI_ISL_8465125, EPI_ISL_8465128, EPI_ISL_8465133, EPI_ISL_8465135, EPI_ISL_8465137, EPI_ISL_8465140, EPI_ISL_8465141, EPI_ISL_8465142, EPI_ISL_8465151, EPI_ISL_8465154, EPI_ISL_8465156, EPI_ISL_8465157, EPI_ISL_8465165, EPI_ISL_8465169, EPI_ISL_8465170, EPI_ISL_8465178, EPI_ISL_8465181, EPI_ISL_8465187, EPI_ISL_8465190, EPI_ISL_8465192, EPI_ISL_8465193, EPI_ISL_8465200, EPI_ISL_8465206, EPI_ISL_8465207, EPI_ISL_8465210, EPI_ISL_8465213, EPI_ISL_8465214, EPI_ISL_8465220, EPI_ISL_8465222, EPI_ISL_8465226, EPI_ISL_8465243, EPI_ISL_8465244, EPI_ISL_8465246, EPI_ISL_8465247, EPI_ISL_8465248, EPI_ISL_8465251, EPI_ISL_8465258, EPI_ISL_8465262, EPI_ISL_8465268, EPI_ISL_8465273, EPI_ISL_8465280, EPI_ISL_8465283, EPI_ISL_8465284, EPI_ISL_8465285, EPI_ISL_8465288, EPI_ISL_8465290, EPI_ISL_8465291, EPI_ISL_8465293, EPI_ISL_8465294, EPI_ISL_8465296, EPI_ISL_8465301, EPI_ISL_8465303, EPI_ISL_8465307, EPI_ISL_8465309, EPI_ISL_8465314, EPI_ISL_8465315, EPI_ISL_8465319, EPI_ISL_8465322, EPI_ISL_8465331, EPI_ISL_8465333, EPI_ISL_8465343, EPI_ISL_8465351, EPI_ISL_8465355, EPI_ISL_8465356, EPI_ISL_8465361, EPI_ISL_8465367, EPI_ISL_8465370, EPI_ISL_8465371, EPI_ISL_8465372, EPI_ISL_8465373, EPI_ISL_8465377, EPI_ISL_8465383, EPI_ISL_8465385, EPI_ISL_8465386, EPI_ISL_8465394, EPI_ISL_8465396, EPI_ISL_8465401, EPI_ISL_8465410, EPI_ISL_8465414, EPI_ISL_8465416, EPI_ISL_8465418, EPI_ISL_8465419, EPI_ISL_8465421, EPI_ISL_8465422, EPI_ISL_8465425, EPI_ISL_8465427, EPI_ISL_8465441, EPI_ISL_8465445, EPI_ISL_8465449, EPI_ISL_8465450, EPI_ISL_8465456, EPI_ISL_8465458, EPI_ISL_8465462, EPI_ISL_8465465, EPI_ISL_8465477, EPI_ISL_8465490, EPI_ISL_8465491, EPI_ISL_8465495, EPI_ISL_8465497, EPI_ISL_8465498, EPI_ISL_8465499, EPI_ISL_8465502, EPI_ISL_8465508, EPI_ISL_8465513, EPI_ISL_8465518, EPI_ISL_8465521, EPI_ISL_8465522, EPI_ISL_8465525, EPI_ISL_8465527, EPI_ISL_8465797, EPI_ISL_8465798, EPI_ISL_8465799, EPI_ISL_8465800, EPI_ISL_8465801, EPI_ISL_8465806, EPI_ISL_8465810, EPI_ISL_8465815, EPI_ISL_8465816, EPI_ISL_8465819, EPI_ISL_8465829, EPI_ISL_8465834, EPI_ISL_8465837, EPI_ISL_8465840, EPI_ISL_8465847, EPI_ISL_8465850, EPI_ISL_8465855, EPI_ISL_8465856, EPI_ISL_8465860, EPI_ISL_8465864, EPI_ISL_8465876, EPI_ISL_8465879, EPI_ISL_8465889, EPI_ISL_8465895, EPI_ISL_8465898, EPI_ISL_8465900, EPI_ISL_8465902, EPI_ISL_8465904, EPI_ISL_8465915, EPI_ISL_8465918, EPI_ISL_8465921, EPI_ISL_8465930, EPI_ISL_8465931, EPI_ISL_8465944, EPI_ISL_8465950, EPI_ISL_8465953, EPI_ISL_8465956, EPI_ISL_8465957, EPI_ISL_8465958, EPI_ISL_8465959, EPI_ISL_8465962, EPI_ISL_8465971, EPI_ISL_8465972, EPI_ISL_8465974, EPI_ISL_8465975, EPI_ISL_8465976, EPI_ISL_8465977, EPI_ISL_8465979, EPI_ISL_8465989, EPI_ISL_8465991, EPI_ISL_8465998, EPI_ISL_8466001, EPI_ISL_8466004, EPI_ISL_8466019, EPI_ISL_8466022, EPI_ISL_8466023, EPI_ISL_8466027, EPI_ISL_8466031, EPI_ISL_8466033 | | | |
| see above | Department of Bacteria, Parasites and Fungi, Statens Serum Institut, Copenhagen, Denmark | Statens Serum Institut Bioinformatics and Microbial Genomics | Danish Covid-19 Genome Consortium |
| EPI_ISL_8461493 | Dutch COVID-19 response team | National Institute for Public Health and the Environment (RIVM) | Adam Meijer; AnneMarie van den Brandt; Annelies Kroneman; Bas van der Veer; Chantal Reusken; Dennis Schmitz; Dirk Eggink; Florian Zwagemaker; Harry Vennema; Ivo van Walle; Jeroen Cremer; Jil Kocken; Jordy de Bakker; Karim Hajji; Kim Freriks; Linda van Someren; Lisa Wijsman; Lynn Aarts; Ryanne Jaarsma; Sanne Bos; Sharon van den Brink; Stijn van Rossum; on behalf of the national COVID-19 response team |
| EPI_ISL_8455623 | Jessa | Jessa | Severine Berden et al. on behalf of the Jessa_cmdLab |
| EPI_ISL_8467775 | PZU “Eurofarm - Centar Poliklinika” | University of Sarajevo, Veterinary Faculty, Laboratory for Molecular Diagnostic and Research | Goletic S; Goletic T; Hodzic A; Huseinbegovic E; Jazic A; Nicevic M.; Prguda-Mujic J; Sabic E; Softic A; Terzic I |
| EPI_ISL_8444273 | Public Health Laboratory, Public Health Service Amsterdam, The Netherlands | Department of Medical Microbiology & Infection prevention, Amsterdam University Medical Centers location AMC | Akke Cornelissen; Fokla Zorgdrager; Janke Schinkel; Jelle Koopsen; Judith den Uil; Marcel Jonges; Matthijs Welkers; Menno de Jong; Robin van Houdt; Sebastien Matamoros; Sjoerd Rebers; Sylvia Bruisten; Tjalling Leenstra and Mariken van der Lubben on behalf of the Amsterdam Regional Genomic epidemiology and Outbreak Surveillance (ARGOS) consortium |
| EPI_ISL_8448885 | Rosalind Franklin Laboratory | Wellcome Sanger Institute for the COVID-19 Genomics UK (COG-UK) Consortium | Cordelia Langford; David K. Jackson; Dominic Kwiatkowski; Donald Fraser; Ewan Harrison; Ian Johnston; Jeffrey Barrett; John Sillitoe on behalf of the Wellcome Sanger Institute COVID-19 Surveillance Team; Rob Howes; Roberto Amato; Sonia Goncalves; Suki Lee; The Rosalind Franklin Laboratory and Alex Alderton |
| EPI_ISL_8467952, EPI_ISL_8468322 | University Clinical Hospital of Mostar, Department of Microbiology and Molecular Diagnostics | University of Sarajevo, Veterinary Faculty, Laboratory for Molecular Diagnostic and Research | Goletic S; Goletic T; Herceg I; Hodzic A; Jazic A; Nicevic M; Ostojic M; Sabic E; Skocibusic S; Softic A; Terzic I |
| EPI_ISL_8466468, EPI_ISL_8466469 | University Hospitals of Geneva, Laboratory of Virology | HUG, Laboratory of Virology and the Health2030 Genome Center | Aline Mamin; Ana Rita Goncalves; Cedric Howald; Deborah Penet; Francisco Perez; Henri Pegeot; Ioannis Xenarios; Keith Harshman; Laurent Kaiser; Lorenzo Cerutti; Melyssa Elies; Samuel Cordey |
| EPI_ISL_8468540 | University of Sarajevo, Veterinary Faculty, Laboratory for Molecular Diagnostic and Research | University of Sarajevo, Veterinary Faculty, Laboratory for Molecular Diagnostic and Research | Goletic S; Goletic T; Hodzic A; Jazic A; Nicevic M; Sabic E; Softic A; Terzic I |

We gratefully acknowledge the following Authors from the Originating laboratories responsible for obtaining the specimens, as well as the Submitting laboratories where the genome data were generated and shared via GISAID, on which this research is based.

All Submitters of data may be contacted directly via www.gisaid.org

Authors are sorted alphabetically.

Acknowledgement EPI_SET Identifier: EPI_SET_20220527qm

| Accession ID | Originating Laboratory | Submitting Laboratory | Authors |
|---|---|---|---|
| EPI_ISL_8471279, EPI_ISL_8472155 | Aegis Sciences Corporation | Centers for Disease Control and Prevention Division of Viral Diseases, Pathogen Discovery | Alec Vest; Benjamin Rambo-Martin; Christopher Gulvick; Clinton Paden; Cyndi Clark; Dakota Howard; Dhwani Batra; Dillon Nall; Duncan MacCannell; Erisa Sula; Ethan Sanders; Holly Houdeshell; Jason Caravas; Kristine Lacek; Matthew Hardison; Matthew Schmerer; Ola Kvalvaag; Patrick Campbell; Peter Cook; Rob Case; Scott Sammons; Shatavia Morrison; Shaun Westlund; Tymeckia Kendall; Victoria Caban Figueroa; Vikramsinha Ghorpade; Yvette Unoarumhi |
| EPI_ISL_8476795, EPI_ISL_8476798, EPI_ISL_8476799, EPI_ISL_8476800 | Colorado Department of Public Health and Environment | Colorado Department of Public Health and Environment | Alexandria Rossheim; Arianna Smith; Diana Ir; Emily A. Travanty; Laura Bankers; Mandy Waters; Michael Martin; Molly C. Hetherington-Rauth; Shannon R. Matzinger |
| EPI_ISL_8475117 | Fulgent Genetics | Centers for Disease Control and Prevention Division of Viral Diseases, Pathogen Discovery | Becky Tsai; Benafsh Sapra; Benjamin Rambo-Martin; Christopher Gulvick; Clinton Paden; Dakota Howard; Dhwani Batra; Doreen Ng; Duncan MacCannell; Erisa Sula; Harry Gao; James Xie; Jason Caravas; John Gao; Joseph Fierro; Kristine Lacek; Matthew Schmerer; Mickey Li; Peter Cook; Scott Sammons; Shatavia Morrison; Tymeckia Kendall; Victoria Caban Figueroa; Yan Meng; Yvette Unoarumhi |
| EPI_ISL_8481779, EPI_ISL_8482526 | Houston Methodist Hospital | Houston Methodist Hospital | Akanksha Batajoo; Ilya J. Finkelstein; James J. Davis; Jessica Cambric; Jimmy Gollihar; Kristina Reppond; Madison N. Shyer; Marcus Nguyen; Matthew Ojeda Saavedra; Paul A. Christensen; Randall J. Olsen; Rashi Thakur; Regan Mangham; Richard Snehall; Robert Olson; Ryan Gadd; S. Wesley Long; Sindy Pena; and James M. Musser |
| EPI_ISL_8473672 | Johns Hopkins Hospital Department of Pathology | Johns Hopkins Hospital Department of Pathology | Amary Fall; C. Paul Morris; David Gaston; Heba H. Mostafa; Julie M. Norton; Matthew Schwartz; Michael Forman; Raghda Eldesouki |
| EPI_ISL_8478365, EPI_ISL_8478587, EPI_ISL_8478658, EPI_ISL_8479317 | Kaiser Permanente Southern California | Helix | Helix; Kaiser Permanente Southern California |
| EPI_ISL_8469485, EPI_ISL_8469703, EPI_ISL_8469732, EPI_ISL_8469868, EPI_ISL_8469869, EPI_ISL_8469990, EPI_ISL_8470301, EPI_ISL_8470302 | | | |
| see above | Laboratory Corporation of America | Centers for Disease Control and Prevention Division of Viral Diseases, Pathogen Discovery | Amanda Douglas; Amanda Suchanek; Andrea Throop; Ayla Burns; Benjamin Rambo-Martin; Bobbi Croy; Brian Krueger; Brian Norvell; Christopher Gulvick; Christos Petropoulos; Clinton Paden; Craig Lukasik; Dakota Howard; Debbie Boles; Dhwani Batra; Duncan MacCannell; Eyad Almasri; Goran Stevovic; Howard Engler; Hrushikesh Deshmukh; Jake Humphrey; Jana Schroth; Jason Caravas; Joe Voshell; John Pruitt; Jonathan Meltzer; Jonathan Williams; Kimberly Wagner; Kristine Lacek; Lax Iyer; Lisa Pfefferle; Lyndon Tilson; Manoj Jain; Marcia Eisenberg; Mary Cristobal; Mary Williamson; Matthew Robinson; Matthew Schmerer; Michael Levandoski; Mike Sapeta; Mindy Nye; Minoo Agarwal; Mohan Kolli; Nuthawin Charoensri; Oren Cohen; Peter Cook; Prashant Gupta; Qian Zeng; Rama Ghatti; Scott Parker; Scott Ryan; Scott Sammons; Shatavia Morrison; Stanley Letovsky; Steven Ragan; Suresh Selvaraju; Susan Countryman; Susan Hicks; Suzanne Dale; Thomas Urban; Tim Kuphal; Tricia Zwiefelhofer; Tymeckia Kendall; Victoria Caban Figueroa; Vincent Drouillon; Yvette Unoarumhi |
| EPI_ISL_8479529, EPI_ISL_8479530, EPI_ISL_8479531, EPI_ISL_8479532, EPI_ISL_8479533, EPI_ISL_8479534, EPI_ISL_8479535, EPI_ISL_8479536, EPI_ISL_8479537, EPI_ISL_8479538, EPI_ISL_8479539, EPI_ISL_8479540, EPI_ISL_8479541, EPI_ISL_8479542, EPI_ISL_8479543, EPI_ISL_8479544, EPI_ISL_8479545, EPI_ISL_8479546, EPI_ISL_8479547, EPI_ISL_8479548, EPI_ISL_8479549, EPI_ISL_8479550, EPI_ISL_8479551, EPI_ISL_8479552, EPI_ISL_8479553, EPI_ISL_8479554, EPI_ISL_8479555, EPI_ISL_8479556, EPI_ISL_8479557, EPI_ISL_8479558, EPI_ISL_8479559, EPI_ISL_8479560, EPI_ISL_8479561, EPI_ISL_8479562, EPI_ISL_8479563, EPI_ISL_8479564, EPI_ISL_8479565, EPI_ISL_8479586 | | | |
| see above | Ochsner Health | BioInfoExperts | Amy Feehan; Ben Lain; Chris Huston; David J. Nolan; Judy Crabtree; Julia-Garcia-Diaz; Lucio Miele; Rebecca Rose; Samual Moot; Susanna L. Lamers; Tessa LaFleur |
| EPI_ISL_8434986 | State Laboratories Division, Hawaii State Department of Health | State Laboratories Division, Hawaii State Department of Health | Ayana Garnet; Briana Ofilas; Cheryl-lynn Daquip; Cheyenne Barela; Daniel Strange; Drew Kuwazaki; Edward Desmond; Jeffrey Au; Mark Nagata; Pamela O'Brien; Razvan Sultana; Remedios Gose; Samantha Cotter; Samantha Sruba |
| EPI_ISL_8479393, EPI_ISL_8480230, EPI_ISL_8480393, EPI_ISL_8480457 | UW Virology Lab | UW Virology Lab | Alexander Greninger; Hong Xie; Isabel Arnould; Keith R Jerome; Meei-Li Huang; Nathan Breit; Patrick Mathias; Pavitra Roychoudhury; Pooneh Hajian; Ricardo Perez; Robert J. Livingston; Saraswathi Sathees; Sean Ellis; Seffir T. Wendm; Shah Mohamed Bakhash; Tien V. Nguyen |
| EPI_ISL_8435628, EPI_ISL_8435634, EPI_ISL_8435636, EPI_ISL_8435640, EPI_ISL_8435641, EPI_ISL_8435643, EPI_ISL_8435644, EPI_ISL_8435645, EPI_ISL_8435646, EPI_ISL_8435651, EPI_ISL_8435655, EPI_ISL_8435656, EPI_ISL_8435661, EPI_ISL_8435664, EPI_ISL_8435666, EPI_ISL_8435672, EPI_ISL_8435680, EPI_ISL_8435684, EPI_ISL_8435693, EPI_ISL_8435695, EPI_ISL_8435698, EPI_ISL_8435701, EPI_ISL_8435702, EPI_ISL_8435703, EPI_ISL_8435704, EPI_ISL_8435705, EPI_ISL_8435716, EPI_ISL_8435723, EPI_ISL_8435727, EPI_ISL_8435728, EPI_ISL_8435731, EPI_ISL_8435743, EPI_ISL_8435744, EPI_ISL_8435748, EPI_ISL_8435750, EPI_ISL_8435751, EPI_ISL_8435752, EPI_ISL_8435757, EPI_ISL_8435765 | | | |
| see above | University of Oregon COVID-19 MAP Laboratory | University of Oregon Genomics and Cell Charactericzation Core Facility (GC3F) | Ariana White; Demi Glidden; Douglas Turnbull; Jason Carriere; Jason Sydes; Jeff Bishop; Megan Criss; Peter Batzel; Tina Arredondo |

We gratefully acknowledge the following Authors from the Originating laboratories responsible for obtaining the specimens, as well as the Submitting laboratories where the genome data were generated and shared via GISAID, on which this research is based.

All Submitters of data may be contacted directly via www.gisaid.org

Authors are sorted alphabetically.

| Accession ID | Originating Laboratory | Submitting Laboratory | Authors |
| --- | --- | --- | --- |
| EPI_ISL_8437525, EPI_ISL_8437527, EPI_ISL_8437528, EPI_ISL_8437529, EPI_ISL_8437530, EPI_ISL_8437532, EPI_ISL_8437536, EPI_ISL_8437537, EPI_ISL_8437539, EPI_ISL_8437540, EPI_ISL_8437542, EPI_ISL_8437543, EPI_ISL_8437544, EPI_ISL_8437547, EPI_ISL_8437548, EPI_ISL_8437549, EPI_ISL_8437550, EPI_ISL_8437551, EPI_ISL_8437552, EPI_ISL_8437553, EPI_ISL_8437555, EPI_ISL_8437556, EPI_ISL_8437557, EPI_ISL_8437558, EPI_ISL_8437560, EPI_ISL_8437561, EPI_ISL_8437562, EPI_ISL_8437563, EPI_ISL_8437564, EPI_ISL_8437566, EPI_ISL_8437567, EPI_ISL_8437570, EPI_ISL_8437571, EPI_ISL_8437572, EPI_ISL_8437573, EPI_ISL_8437575, EPI_ISL_8437576, EPI_ISL_8437577, EPI_ISL_8437580, EPI_ISL_8437581, EPI_ISL_8437582, EPI_ISL_8437583, EPI_ISL_8437584, EPI_ISL_8437585, EPI_ISL_8437586, EPI_ISL_8437587, EPI_ISL_8437588, EPI_ISL_8437590, EPI_ISL_8437592, EPI_ISL_8437594, EPI_ISL_8437595, EPI_ISL_8437596, EPI_ISL_8437597, EPI_ISL_8437598, EPI_ISL_8437602, EPI_ISL_8437604, EPI_ISL_8437608, EPI_ISL_8437609, EPI_ISL_8437614, EPI_ISL_8437615, EPI_ISL_8437616, EPI_ISL_8437623, EPI_ISL_8437624, EPI_ISL_8437625, EPI_ISL_8437627, EPI_ISL_8437628, EPI_ISL_8437629, EPI_ISL_8437633, EPI_ISL_8437634, EPI_ISL_8437637, EPI_ISL_8437639, EPI_ISL_8437641, EPI_ISL_8437642, EPI_ISL_8437644, EPI_ISL_8437645, EPI_ISL_8437649, EPI_ISL_8437651, EPI_ISL_8437652, EPI_ISL_8437653, EPI_ISL_8437655, EPI_ISL_8437657, EPI_ISL_8437658, EPI_ISL_8437660, EPI_ISL_8437661, EPI_ISL_8437662, EPI_ISL_8437663, EPI_ISL_8437668, EPI_ISL_8437669, EPI_ISL_8437670, EPI_ISL_8437672, EPI_ISL_8437673, EPI_ISL_8437674, EPI_ISL_8437678, EPI_ISL_8437680, EPI_ISL_8437681, EPI_ISL_8437683, EPI_ISL_8437684, EPI_ISL_8437685, EPI_ISL_8437686, EPI_ISL_8437689, EPI_ISL_8437690, EPI_ISL_8437691, EPI_ISL_8437692, EPI_ISL_8437693, EPI_ISL_8437697, EPI_ISL_8437700, EPI_ISL_8437701, EPI_ISL_8437706, EPI_ISL_8437707, EPI_ISL_8437710, EPI_ISL_8437711, EPI_ISL_8437714, EPI_ISL_8437715, EPI_ISL_8437717, EPI_ISL_8437718, EPI_ISL_8437720, EPI_ISL_8437721, EPI_ISL_8437723, EPI_ISL_8437724, EPI_ISL_8437728, EPI_ISL_8437730, EPI_ISL_8437731, EPI_ISL_8437732, EPI_ISL_8437733, EPI_ISL_8437734, EPI_ISL_8437738, EPI_ISL_8437740, EPI_ISL_8437741, EPI_ISL_8437742, EPI_ISL_8437744, EPI_ISL_8437745, EPI_ISL_8437746, EPI_ISL_8437747, EPI_ISL_8437749, EPI_ISL_8437751, EPI_ISL_8437753, EPI_ISL_8437757, EPI_ISL_8437759, EPI_ISL_8437761, EPI_ISL_8437765, EPI_ISL_8437767, EPI_ISL_8437771, EPI_ISL_8437773, EPI_ISL_8437774, EPI_ISL_8437776, EPI_ISL_8437778, EPI_ISL_8437779, EPI_ISL_8437780, EPI_ISL_8437781, EPI_ISL_8437782, EPI_ISL_8437783, EPI_ISL_8437784, EPI_ISL_8437786, EPI_ISL_8437788, EPI_ISL_8437789, EPI_ISL_8437791, EPI_ISL_8437792, EPI_ISL_8437793, EPI_ISL_8437794, EPI_ISL_8437796, EPI_ISL_8437797, EPI_ISL_8437798, EPI_ISL_8437799, EPI_ISL_8437801, EPI_ISL_8437803, EPI_ISL_8437804, EPI_ISL_8437807, EPI_ISL_8437809, EPI_ISL_8437810, EPI_ISL_8437813, EPI_ISL_8437814, EPI_ISL_8437815, EPI_ISL_8437820, EPI_ISL_8437821, EPI_ISL_8437822, EPI_ISL_8437823, EPI_ISL_8437824, EPI_ISL_8437825, EPI_ISL_8437828, EPI_ISL_8437832, EPI_ISL_8437833, EPI_ISL_8437836, EPI_ISL_8437838, EPI_ISL_8437841, EPI_ISL_8437847, EPI_ISL_8437849, EPI_ISL_8437850, EPI_ISL_8437851, EPI_ISL_8437858, EPI_ISL_8437867, EPI_ISL_8437871, EPI_ISL_8437875, EPI_ISL_8437878, EPI_ISL_8437880, EPI_ISL_8437881, EPI_ISL_8437884, EPI_ISL_8437888, EPI_ISL_8437896, EPI_ISL_8437904, EPI_ISL_8437909, EPI_ISL_8437910, EPI_ISL_8437914, EPI_ISL_8437915, EPI_ISL_8437919, EPI_ISL_8437920, EPI_ISL_8437926, EPI_ISL_8437932, EPI_ISL_8437933, EPI_ISL_8437934, EPI_ISL_8437937, EPI_ISL_8437952, EPI_ISL_8437954, EPI_ISL_8437958, EPI_ISL_8437960, EPI_ISL_8437965, EPI_ISL_8437970, EPI_ISL_8437973, EPI_ISL_8437980, EPI_ISL_8437981, EPI_ISL_8437982, EPI_ISL_8437985, EPI_ISL_8437986, EPI_ISL_8437990, EPI_ISL_8437991, EPI_ISL_8437992, EPI_ISL_8437993, EPI_ISL_8437994, EPI_ISL_8437996, EPI_ISL_8437999, EPI_ISL_8438002, EPI_ISL_8438003, EPI_ISL_8438004, EPI_ISL_8438005, EPI_ISL_8438006, EPI_ISL_8438007, EPI_ISL_8438008, EPI_ISL_8438010, EPI_ISL_8438011, EPI_ISL_8438012, EPI_ISL_8438013, EPI_ISL_8438015, EPI_ISL_8438017, EPI_ISL_8438018, EPI_ISL_8438019, EPI_ISL_8438020, EPI_ISL_8438022, EPI_ISL_8438024, EPI_ISL_8438027, EPI_ISL_8438028, EPI_ISL_8438030, EPI_ISL_8438034, EPI_ISL_8438035, EPI_ISL_8438036, EPI_ISL_8438038, EPI_ISL_8438039, EPI_ISL_8438040, EPI_ISL_8438041, EPI_ISL_8438044, EPI_ISL_8438045, EPI_ISL_8438046, EPI_ISL_8438048, EPI_ISL_8438049, EPI_ISL_8438050, EPI_ISL_8438053, EPI_ISL_8438054, EPI_ISL_8438056, EPI_ISL_8438058, EPI_ISL_8438059, EPI_ISL_8438060, EPI_ISL_8438061, EPI_ISL_8438063, EPI_ISL_8438064, EPI_ISL_8438065, EPI_ISL_8438066, EPI_ISL_8438069, EPI_ISL_8438070, EPI_ISL_8438072, EPI_ISL_8438073, EPI_ISL_8438076, EPI_ISL_8438077, EPI_ISL_8438078, EPI_ISL_8438080, EPI_ISL_8438082, EPI_ISL_8438083, EPI_ISL_8438084, EPI_ISL_8438089, EPI_ISL_8438091, EPI_ISL_8438093, EPI_ISL_8438094, EPI_ISL_8438097, EPI_ISL_8438098, EPI_ISL_8438100, EPI_ISL_8438102, EPI_ISL_8438103, EPI_ISL_8438104, EPI_ISL_8438105, EPI_ISL_8438108, EPI_ISL_8438110, EPI_ISL_8438112, EPI_ISL_8438113, EPI_ISL_8438116, EPI_ISL_8438118, EPI_ISL_8438119, EPI_ISL_8438124, EPI_ISL_8438126, EPI_ISL_8438127, EPI_ISL_8438128, EPI_ISL_8438130, EPI_ISL_8438132, EPI_ISL_8438133, EPI_ISL_8438135, EPI_ISL_8438137, EPI_ISL_8438138, EPI_ISL_8438140, EPI_ISL_8438141, EPI_ISL_8438143, EPI_ISL_8438144, EPI_ISL_8438147, EPI_ISL_8438148, EPI_ISL_8438149, EPI_ISL_8438152, EPI_ISL_8438155, EPI_ISL_8438156, EPI_ISL_8438157, EPI_ISL_8438158, EPI_ISL_8438159, EPI_ISL_8438160, EPI_ISL_8438161, EPI_ISL_8438164, EPI_ISL_8438169, EPI_ISL_8438172, EPI_ISL_8438173, EPI_ISL_8438174, EPI_ISL_8438175, EPI_ISL_8438177, EPI_ISL_8438178, EPI_ISL_8438179, EPI_ISL_8438183, EPI_ISL_8438184, EPI_ISL_8438185, EPI_ISL_8438186, EPI_ISL_8438188, EPI_ISL_8438189, EPI_ISL_8438190, EPI_ISL_8438193, EPI_ISL_8438194, EPI_ISL_8438198, EPI_ISL_8438207, EPI_ISL_8438210, EPI_ISL_8438211, EPI_ISL_8438216, EPI_ISL_8438219, EPI_ISL_8438220, EPI_ISL_8438222, EPI_ISL_8438224, EPI_ISL_8438227, EPI_ISL_8438229, EPI_ISL_8438231, EPI_ISL_8438234, EPI_ISL_8438235, EPI_ISL_8438236, EPI_ISL_8438240, EPI_ISL_8438241, EPI_ISL_8438246, EPI_ISL_8438248, EPI_ISL_8438249, EPI_ISL_8438251, EPI_ISL_8438254, EPI_ISL_8438257, EPI_ISL_8438258, EPI_ISL_8438262, EPI_ISL_8438263, EPI_ISL_8438264, EPI_ISL_8438270, EPI_ISL_8438272, EPI_ISL_8438274, EPI_ISL_8438276, EPI_ISL_8438277, EPI_ISL_8438278, EPI_ISL_8438279, EPI_ISL_8438281, EPI_ISL_8438283, EPI_ISL_8438284, EPI_ISL_8438286, EPI_ISL_8438287, EPI_ISL_8438288, EPI_ISL_8438289, EPI_ISL_8438290, EPI_ISL_8438292, EPI_ISL_8438293, EPI_ISL_8438294, EPI_ISL_8438295, EPI_ISL_8438296, EPI_ISL_8438299, EPI_ISL_8438301, EPI_ISL_8438302, EPI_ISL_8438303, EPI_ISL_8438304, EPI_ISL_8438305, EPI_ISL_8438306, EPI_ISL_8438307, EPI_ISL_8438308, EPI_ISL_8438309, EPI_ISL_8438310, EPI_ISL_8438311, EPI_ISL_8438312, EPI_ISL_8438313, EPI_ISL_8438314, EPI_ISL_8438315, EPI_ISL_8438316, EPI_ISL_8438317, EPI_ISL_8438318, EPI_ISL_8438319, EPI_ISL_8438321, EPI_ISL_8438322, EPI_ISL_8438323, EPI_ISL_8438324, EPI_ISL_8438325, EPI_ISL_8438326, EPI_ISL_8438327, EPI_ISL_8438328, EPI_ISL_8438329, EPI_ISL_8438330, EPI_ISL_8438331, EPI_ISL_8438333, EPI_ISL_8438334, EPI_ISL_8438335, EPI_ISL_8438336, EPI_ISL_8438428, EPI_ISL_8438429, EPI_ISL_8438430, EPI_ISL_8438432, EPI_ISL_8438436, EPI_ISL_8438438, EPI_ISL_8438439, EPI_ISL_8438441, EPI_ISL_8438443, EPI_ISL_8438445, EPI_ISL_8438449, EPI_ISL_8438451, EPI_ISL_8438454, EPI_ISL_8438458, EPI_ISL_8438464, EPI_ISL_8438465, EPI_ISL_8438469 | | | |
| see above | PHV-FSS | PHV-FSS | Chenwei Wang on behalf of Q-PHIRE Genomics |

We gratefully acknowledge the following Authors from the Originating laboratories responsible for obtaining the specimens, as well as the Submitting laboratories where the genome data were generated and shared via GISAID, on which this research is based.

All Submitters of data may be contacted directly via www.gisaid.org

Authors are sorted alphabetically.

| Accession ID | Originating Laboratory | Submitting Laboratory | Authors |
|---|---|---|---|
| EPI_ISL_9028453, EPI_ISL_9028454, EPI_ISL_9028455, EPI_ISL_9028456, EPI_ISL_9028457, EPI_ISL_9028458, EPI_ISL_9028459, EPI_ISL_9028460, EPI_ISL_9028461, EPI_ISL_9028462, EPI_ISL_9028463, EPI_ISL_9028464, EPI_ISL_9028465, EPI_ISL_9028466, EPI_ISL_9028467, EPI_ISL_9028468, EPI_ISL_9028469, EPI_ISL_9028470, EPI_ISL_9028471, EPI_ISL_9028472, EPI_ISL_9028473, EPI_ISL_9028474, EPI_ISL_9028475, EPI_ISL_9028477, EPI_ISL_9028478, EPI_ISL_9028480, EPI_ISL_9028481, EPI_ISL_9028482, EPI_ISL_9028483, EPI_ISL_9028484, EPI_ISL_9028485, EPI_ISL_9028486, EPI_ISL_9028487, EPI_ISL_9028488, EPI_ISL_9028489, EPI_ISL_9028490, EPI_ISL_9028491, EPI_ISL_9028492, EPI_ISL_9028493, EPI_ISL_9028495 | | | |
| see above | Ministry of Public Health / Hamad Medical Corporation | Biomedical Research Center (BRC), Qatar University / Qatar Genome Project (QGP) | Asmaa A. Al-Thani. MOPH and HMC: Abdullatif Al-Khal; BRC: Heba A. Al-Khatib; Chadi Saad; Dana Al-Batsh; Dina Elgakhlab. QGP: Fatima H. Al-Kuwari; Einas A. E. Al-Kuwari; Fatiha M. Benslimane; Fatma Hassan Ali; Hadeel T. Mohammed; Hadi M. Yassine; Hamad E. Al-Romaihi; Hamda Alromaihi; Maria K. Smatti; Mashael A. Al-Bader; Mohammed Al-Thani; Muna A. S. Al-Maslamani; Ola Al-Jamal; Peter V. Coyle; Reham A. El-Kahlout. QBB: Tasneem Al-Hamad; Roberto Bertollini; Salih Al-Marri; Swapna Thomas |
| EPI_ISL_9019278, EPI_ISL_9019279 | RS Ciputra Citra Garden | Genomik Solidaritas Indonesia Laboratorium | Aisyi Sakina Rifani; Annisa Muthiah Sukirman; Anuraj Shankar; Ariel Pradipta; Carissa Sintca Wijaya; Dian Tri Wahyudi; Esti Rahayuning Tias; Filly Mandalie; Gracia Felias Enos Korompis; Louisa Markus; Meutia Ayuputeri Kumaheri; Stefanus Bernard |

We gratefully acknowledge the following Authors from the Originating laboratories responsible for obtaining the specimens, as well as the Submitting laboratories where the genome data were generated and shared via GISAID, on which this research is based.

All Submitters of data may be contacted directly via www.gisaid.org

Authors are sorted alphabetically.

Acknowledgement EPI_SET Identifier: EPI_SET_20220527rx

| Accession ID | Originating Laboratory | Submitting Laboratory | Authors |
| --- | --- | --- | --- |
| EPI_ISL_9082856, EPI_ISL_9082858, EPI_ISL_9082859 | CAPRISA | CERI, Centre for Epidemic Response and Innvoation, Stellenbosch University and KRISP, KZN Research Innovation and Sequencing Platform, UKZN. | Aida Sivro; Giandhari J; Naidoo Y; Natasha Samsunder; Pillay S; Ramphal U; Ramphal Y; San JE; Tegally H; Tshiabuila D; Wilkinson E; de Oliveira T; van Wyk S |

We gratefully acknowledge the following Authors from the Originating laboratories responsible for obtaining the specimens, as well as the Submitting laboratories where the genome data were generated and shared via GISAID, on which this research is based.

All Submitters of data may be contacted directly via www.gisaid.org

Authors are sorted alphabetically.

| Accession ID | Originating Laboratory | Submitting Laboratory | Authors |
| --- | --- | --- | --- |
| EPI_ISL_9072095 | Allergy, Immunology and Cell Biology Unit (AICBU) | Allergy, Immunology and Cell Biology Unit (AICBU) | Ayesha Wijesinghe; Chandima Jeewandara; Deshni Jayathilaka; Dinuka Ariyaratne; Diyanath Ranasinghe; Dumni Guasinghe; Farha Bary; Gathsaurie Neelika Malavige; Heshan Kuruppu; Tibutius Thanesh |
| EPI_ISL_9076079, EPI_ISL_9076080, EPI_ISL_9076081, EPI_ISL_9076082, EPI_ISL_9076083, EPI_ISL_9076084, EPI_ISL_9076086, EPI_ISL_9076087, EPI_ISL_9076088, EPI_ISL_9076089, EPI_ISL_9076090, EPI_ISL_9076091, EPI_ISL_9076092, EPI_ISL_9076093, EPI_ISL_9076094, EPI_ISL_9076096, EPI_ISL_9076097, EPI_ISL_9076098, EPI_ISL_9076099, EPI_ISL_9076100, EPI_ISL_9076101, EPI_ISL_9076102, EPI_ISL_9076103, EPI_ISL_9076104, EPI_ISL_9076105, EPI_ISL_9076106, EPI_ISL_9076107, EPI_ISL_9076108, EPI_ISL_9076109 | | | |
| see above | National Public Health Laboratory, National Centre for Infectious Diseases | National Public Health Laboratory, National Centre for Infectious Diseases | Benny Yeo; Grace Ngan; Lin Cui; Raymond Tzer Pin Lin; Royce Ang; Samuel Loo; Yichen Ding; Zhenyang Zhou |

We gratefully acknowledge the following Authors from the Originating laboratories responsible for obtaining the specimens, as well as the Submitting laboratories where the genome data were generated and shared via GISAID, on which this research is based.

All Submitters of data may be contacted directly via www.gisaid.org

Authors are sorted alphabetically.

Acknowledgement EPI_SET Identifier: EPI_SET_20220527ed

| Accession ID | Originating Laboratory | Submitting Laboratory | Authors |
|---|---|---|---|
| EPI_ISL_9064941 | CHUV | Laboratory of genomics and metagenomics, Institute of Microbiology, University Hospital Centre and University of Lausanne | Claire Bertelli; Damien Jacot; Gilbert Greub; Sébastien Aeby; Trestan Pillonel |
| EPI_ISL_9063244, EPI_ISL_9063246, EPI_ISL_9063254, EPI_ISL_9063257, EPI_ISL_9063268, EPI_ISL_9063297, EPI_ISL_9063306, EPI_ISL_9063308, EPI_ISL_9063319, EPI_ISL_9063329, EPI_ISL_9063335, EPI_ISL_9063342, EPI_ISL_9063345, EPI_ISL_9063346, EPI_ISL_9063350, EPI_ISL_9063352, EPI_ISL_9063367, EPI_ISL_9063378, EPI_ISL_9063380, EPI_ISL_9063385, EPI_ISL_9063390, EPI_ISL_9063395, EPI_ISL_9063401, EPI_ISL_9063415, EPI_ISL_9063418, EPI_ISL_9063421, EPI_ISL_9063423, EPI_ISL_9063452, EPI_ISL_9063455, EPI_ISL_9063457, EPI_ISL_9063468, EPI_ISL_9063480, EPI_ISL_9063482, EPI_ISL_9063485, EPI_ISL_9063490, EPI_ISL_9063493, EPI_ISL_9063503, EPI_ISL_9063506, EPI_ISL_9063509, EPI_ISL_9063514, EPI_ISL_9063517, EPI_ISL_9063522, EPI_ISL_9063533, EPI_ISL_9063536, EPI_ISL_9063547, EPI_ISL_9063549, EPI_ISL_9063552, EPI_ISL_9063555, EPI_ISL_9063558, EPI_ISL_9063560, EPI_ISL_9063565, EPI_ISL_9063567, EPI_ISL_9063570, EPI_ISL_9063573, EPI_ISL_9063577, EPI_ISL_9063580, EPI_ISL_9063586, EPI_ISL_9063591, EPI_ISL_9063596, EPI_ISL_9063601, EPI_ISL_9063606, EPI_ISL_9063610, EPI_ISL_9063612, EPI_ISL_9063618, EPI_ISL_9063623, EPI_ISL_9063636, EPI_ISL_9063637, EPI_ISL_9063640, EPI_ISL_9063644, EPI_ISL_9063649, EPI_ISL_9063651, EPI_ISL_9063663, EPI_ISL_9063665, EPI_ISL_9063682, EPI_ISL_9063684, EPI_ISL_9063688, EPI_ISL_9063693, EPI_ISL_9063696, EPI_ISL_9063707, EPI_ISL_9063715, EPI_ISL_9063720, EPI_ISL_9063726, EPI_ISL_9063729, EPI_ISL_9063731, EPI_ISL_9063732, EPI_ISL_9063733, EPI_ISL_9063734, EPI_ISL_9063738, EPI_ISL_9063739, EPI_ISL_9063740, EPI_ISL_9063744, EPI_ISL_9063753, EPI_ISL_9063761, EPI_ISL_9063763, EPI_ISL_9063770, EPI_ISL_9063773, EPI_ISL_9063782, EPI_ISL_9063785, EPI_ISL_9063807, EPI_ISL_9063813, EPI_ISL_9063816, EPI_ISL_9063820, EPI_ISL_9063823, EPI_ISL_9063828, EPI_ISL_9063835, EPI_ISL_9063838, EPI_ISL_9063850, EPI_ISL_9063853, EPI_ISL_9063856, EPI_ISL_9063859, EPI_ISL_9063861, EPI_ISL_9063868, EPI_ISL_9063874, EPI_ISL_9063880, EPI_ISL_9063888, EPI_ISL_9063892, EPI_ISL_9063903, EPI_ISL_9063905, EPI_ISL_9063909, EPI_ISL_9063911, EPI_ISL_9063917, EPI_ISL_9063920, EPI_ISL_9063931, EPI_ISL_9063935, EPI_ISL_9063938, EPI_ISL_9063951, EPI_ISL_9063957, EPI_ISL_9063972, EPI_ISL_9063978, EPI_ISL_9063984, EPI_ISL_9063990, EPI_ISL_9063992, EPI_ISL_9063999, EPI_ISL_9064001, EPI_ISL_9064006, EPI_ISL_9064010, EPI_ISL_9064024, EPI_ISL_9064029, EPI_ISL_9064032, EPI_ISL_9064035, EPI_ISL_9064038, EPI_ISL_9064044, EPI_ISL_9064047, EPI_ISL_9064053, EPI_ISL_9064056, EPI_ISL_9064058, EPI_ISL_9064061, EPI_ISL_9064063, EPI_ISL_9064067, EPI_ISL_9064087, EPI_ISL_9064108, EPI_ISL_9064114, EPI_ISL_9064116, EPI_ISL_9064158, EPI_ISL_9064160, EPI_ISL_9064166, EPI_ISL_9064167, EPI_ISL_9064171, EPI_ISL_9064196, EPI_ISL_9064199, EPI_ISL_9064206, EPI_ISL_9064209, EPI_ISL_9064216, EPI_ISL_9064217, EPI_ISL_9064248, EPI_ISL_9064258, EPI_ISL_9064263, EPI_ISL_9064274, EPI_ISL_9064278, EPI_ISL_9064286, EPI_ISL_9064287, EPI_ISL_9064296, EPI_ISL_9064299, EPI_ISL_9064308, EPI_ISL_9064309, EPI_ISL_9064310, EPI_ISL_9064320, EPI_ISL_9064323, EPI_ISL_9064325, EPI_ISL_9064330, EPI_ISL_9064333, EPI_ISL_9064338, EPI_ISL_9064341, EPI_ISL_9064343, EPI_ISL_9064355, EPI_ISL_9064361, EPI_ISL_9064363, EPI_ISL_9064365, EPI_ISL_9064368, EPI_ISL_9064386, EPI_ISL_9064391, EPI_ISL_9064401, EPI_ISL_9064403, EPI_ISL_9064409, EPI_ISL_9064413, EPI_ISL_9064418, EPI_ISL_9064421, EPI_ISL_9064425, EPI_ISL_9064428, EPI_ISL_9064430, EPI_ISL_9064442, EPI_ISL_9064451, EPI_ISL_9064455, EPI_ISL_9064466, EPI_ISL_9064470, EPI_ISL_9064478, EPI_ISL_9064488, EPI_ISL_9064502, EPI_ISL_9064505, EPI_ISL_9064527, EPI_ISL_9064530, EPI_ISL_9064534, EPI_ISL_9064536, EPI_ISL_9064548, EPI_ISL_9064554, EPI_ISL_9064563, EPI_ISL_9064565, EPI_ISL_9064569, EPI_ISL_9064572, EPI_ISL_9064574, EPI_ISL_9064584, EPI_ISL_9064587, EPI_ISL_9064609, EPI_ISL_9064614, EPI_ISL_9064623, EPI_ISL_9064632, EPI_ISL_9064644, EPI_ISL_9064649, EPI_ISL_9064653, EPI_ISL_9064664, EPI_ISL_9064666, EPI_ISL_9064678, EPI_ISL_9064684, EPI_ISL_9064687, EPI_ISL_9064701, EPI_ISL_9064708, EPI_ISL_9064711, EPI_ISL_9064717, EPI_ISL_9064723, EPI_ISL_9064726, EPI_ISL_9064731, EPI_ISL_9064737, EPI_ISL_9064742, EPI_ISL_9064768, EPI_ISL_9064774, EPI_ISL_9064786, EPI_ISL_9064792, EPI_ISL_9064805, EPI_ISL_9064810, EPI_ISL_9064837, EPI_ISL_9064863, EPI_ISL_9064865, EPI_ISL_9064867, EPI_ISL_9064870, EPI_ISL_9064873, EPI_ISL_9064880, EPI_ISL_9064882, EPI_ISL_9064885, EPI_ISL_9064888, EPI_ISL_9064897, EPI_ISL_9064903, EPI_ISL_9064908, EPI_ISL_9064916, EPI_ISL_9064919, EPI_ISL_9064939, EPI_ISL_9064943, EPI_ISL_9064946, EPI_ISL_9064953, EPI_ISL_9064957, EPI_ISL_9064960, EPI_ISL_9064965, EPI_ISL_9064978, EPI_ISL_9064990, EPI_ISL_9064997, EPI_ISL_9065006, EPI_ISL_9065043, EPI_ISL_9065047, EPI_ISL_9065056, EPI_ISL_9065059, EPI_ISL_9065063, EPI_ISL_9065074, EPI_ISL_9065077, EPI_ISL_9065084, EPI_ISL_9065090, EPI_ISL_9065093, EPI_ISL_9065096, EPI_ISL_9065108, EPI_ISL_9065121, EPI_ISL_9065123, EPI_ISL_9065128, EPI_ISL_9065133, EPI_ISL_9065137, EPI_ISL_9065140, EPI_ISL_9065143, EPI_ISL_9065147, EPI_ISL_9065150, EPI_ISL_9065158, EPI_ISL_9065161, EPI_ISL_9065170, EPI_ISL_9065186, EPI_ISL_9065191, EPI_ISL_9065198, EPI_ISL_9065201, EPI_ISL_9065207, EPI_ISL_9065212, EPI_ISL_9065227, EPI_ISL_9065234, EPI_ISL_9065237, EPI_ISL_9065255, EPI_ISL_9065258, EPI_ISL_9065275, EPI_ISL_9065277, EPI_ISL_9065280, EPI_ISL_9065294, EPI_ISL_9065303, EPI_ISL_9065331, EPI_ISL_9065334, EPI_ISL_9065336, EPI_ISL_9065339, EPI_ISL_9065344, EPI_ISL_9065346, EPI_ISL_9065349, EPI_ISL_9065351, EPI_ISL_9065353, EPI_ISL_9065364, EPI_ISL_9065374, EPI_ISL_9065402, EPI_ISL_9065419, EPI_ISL_9065424, EPI_ISL_9065427, EPI_ISL_9065433, EPI_ISL_9065440, EPI_ISL_9065453, EPI_ISL_9065458, EPI_ISL_9065461, EPI_ISL_9065464, EPI_ISL_9065474, EPI_ISL_9065478, EPI_ISL_9065484, EPI_ISL_9065486, EPI_ISL_9065489, EPI_ISL_9065506, EPI_ISL_9065508, EPI_ISL_9065514, EPI_ISL_9065522, EPI_ISL_9065543, EPI_ISL_9065548, EPI_ISL_9065554, EPI_ISL_9065569, EPI_ISL_9065571, EPI_ISL_9065577, EPI_ISL_9065581, EPI_ISL_9065594, EPI_ISL_9065597, EPI_ISL_9065603, EPI_ISL_9065606, EPI_ISL_9065608, EPI_ISL_9065610, EPI_ISL_9065612, EPI_ISL_9065613, EPI_ISL_9065614, EPI_ISL_9065634, EPI_ISL_9065639, EPI_ISL_9065642, EPI_ISL_9065659, EPI_ISL_9065671, EPI_ISL_9065677, EPI_ISL_9065693, EPI_ISL_9065695, EPI_ISL_9065707, EPI_ISL_9065710, EPI_ISL_9065711, EPI_ISL_9065713, EPI_ISL_9065720, EPI_ISL_9065726, EPI_ISL_9065728, EPI_ISL_9065730, EPI_ISL_9065732, EPI_ISL_9065743 | | | |
| see above | Department of Bacteria, Parasites and Fungi, Statens Serum Institut, Copenhagen, Denmark | Statens Serum Institut Bioinformatics and Microbial Genomics | Danish Covid-19 Genome Consortium |
| EPI_ISL_9058354, EPI_ISL_9058356, EPI_ISL_9058361, EPI_ISL_9058364, EPI_ISL_9058370, EPI_ISL_9058376, EPI_ISL_9058380, EPI_ISL_9058389, EPI_ISL_9058390, EPI_ISL_9058394, EPI_ISL_9058399, EPI_ISL_9058402, EPI_ISL_9058407, EPI_ISL_9058408, EPI_ISL_9058410, EPI_ISL_9058411, EPI_ISL_9058414, EPI_ISL_9058417, EPI_ISL_9058418, EPI_ISL_9058427, EPI_ISL_9058428, EPI_ISL_9058429, EPI_ISL_9058431, EPI_ISL_9058433, EPI_ISL_9058436, EPI_ISL_9058462, EPI_ISL_9058464, EPI_ISL_9058465, EPI_ISL_9058466, EPI_ISL_9058467, EPI_ISL_9058468, EPI_ISL_9058470, EPI_ISL_9058471, EPI_ISL_9058473, EPI_ISL_9058475, EPI_ISL_9058476, EPI_ISL_9058477, EPI_ISL_9058478, EPI_ISL_9058479, EPI_ISL_9058480, EPI_ISL_9058481, EPI_ISL_9058483, EPI_ISL_9058486, EPI_ISL_9058488, EPI_ISL_9058490, EPI_ISL_9058497, EPI_ISL_9058506, EPI_ISL_9058513, EPI_ISL_9058515, EPI_ISL_9058516, EPI_ISL_9058530, EPI_ISL_9058540, EPI_ISL_9058543, EPI_ISL_9058544, EPI_ISL_9058546, EPI_ISL_9058547, EPI_ISL_9058548, EPI_ISL_9058549, EPI_ISL_9058550, EPI_ISL_9058551, EPI_ISL_9058554, EPI_ISL_9058555, EPI_ISL_9058557, EPI_ISL_9058558, EPI_ISL_9058559, EPI_ISL_9058561, EPI_ISL_9058562, EPI_ISL_9058563, EPI_ISL_9058564, EPI_ISL_9058606, EPI_ISL_9058607, EPI_ISL_9058608, EPI_ISL_9058609, EPI_ISL_9058610, EPI_ISL_9058613, EPI_ISL_9058614, EPI_ISL_9058617, EPI_ISL_9058618, EPI_ISL_9058622, EPI_ISL_9058627 | | | |
| see above | Lab. Microbiologia e Virologia Cotugno A.O. dei Colli | TIGEM | Antonio Grimaldi Patrizia Annunziata Francesco Panariello Claudia Tiberio Teresa Giuliano Valentina Bouche Chiara Colantuono Michela Daniele Lucio Di Filippo Anna Manfredi Marcello Salvi Antonio Limone Luigi Atripaldi Andrea Ballabio Davide Cacchiarelli |
| EPI_ISL_9061587 | Respiratory Virus Unit, Microbiology Services Colindale, Public Health England | COVID-19 Genomics UK (COG-UK) Consortium | PHE Covid Sequencing Team |

We gratefully acknowledge the following Authors from the Originating laboratories responsible for obtaining the specimens, as well as the Submitting laboratories where the genome data were generated and shared via GISAID, on which this research is based.

All Submitters of data may be contacted directly via www.gisaid.org

Authors are sorted alphabetically.

Acknowledgement EPI_SET Identifier: EPI_SET_20220527sy

| Accession ID | Originating Laboratory | Submitting Laboratory | Authors |
|---|---|---|---|
| EPI_ISL_9065750, EPI_ISL_9065758, EPI_ISL_9065760, EPI_ISL_9065762, EPI_ISL_9065764, EPI_ISL_9065770, EPI_ISL_9065773, EPI_ISL_9065788, EPI_ISL_9065791, EPI_ISL_9065794, EPI_ISL_9065800, EPI_ISL_9065810, EPI_ISL_9065812, EPI_ISL_9065821, EPI_ISL_9065828, EPI_ISL_9065834, EPI_ISL_9065835, EPI_ISL_9065838, EPI_ISL_9065849, EPI_ISL_9065851, EPI_ISL_9065859, EPI_ISL_9065862, EPI_ISL_9065870, EPI_ISL_9065898, EPI_ISL_9065902, EPI_ISL_9065903, EPI_ISL_9065906, EPI_ISL_9065912, EPI_ISL_9065915, EPI_ISL_9065923, EPI_ISL_9065925, EPI_ISL_9065929, EPI_ISL_9065936, EPI_ISL_9065961, EPI_ISL_9065962, EPI_ISL_9065964, EPI_ISL_9065967, EPI_ISL_9065971, EPI_ISL_9065973, EPI_ISL_9065974, EPI_ISL_9065977, EPI_ISL_9065978, EPI_ISL_9065979, EPI_ISL_9065984, EPI_ISL_9065993, EPI_ISL_9065998, EPI_ISL_9066000, EPI_ISL_9066002, EPI_ISL_9066011, EPI_ISL_9066024, EPI_ISL_9066027, EPI_ISL_9066030, EPI_ISL_9066032, EPI_ISL_9066044, EPI_ISL_9066050, EPI_ISL_9066057, EPI_ISL_9066064, EPI_ISL_9066088, EPI_ISL_9066099, EPI_ISL_9066102, EPI_ISL_9066105, EPI_ISL_9066113, EPI_ISL_9066118, EPI_ISL_9066122, EPI_ISL_9066127, EPI_ISL_9066138, EPI_ISL_9066149, EPI_ISL_9066158, EPI_ISL_9066164, EPI_ISL_9066167, EPI_ISL_9066172, EPI_ISL_9066183, EPI_ISL_9066186, EPI_ISL_9066191, EPI_ISL_9066194, EPI_ISL_9066196, EPI_ISL_9066205, EPI_ISL_9066208, EPI_ISL_9066222, EPI_ISL_9066239, EPI_ISL_9066242, EPI_ISL_9066247, EPI_ISL_9066250, EPI_ISL_9066256, EPI_ISL_9066261, EPI_ISL_9066269, EPI_ISL_9066275, EPI_ISL_9066290, EPI_ISL_9066299, EPI_ISL_9066301, EPI_ISL_9066307, EPI_ISL_9066309, EPI_ISL_9066320, EPI_ISL_9066323, EPI_ISL_9066326, EPI_ISL_9066332, EPI_ISL_9066338, EPI_ISL_9066350, EPI_ISL_9066359, EPI_ISL_9066362, EPI_ISL_9066368, EPI_ISL_9066370, EPI_ISL_9066373, EPI_ISL_9066375, EPI_ISL_9066377, EPI_ISL_9066380, EPI_ISL_9066396, EPI_ISL_9066399, EPI_ISL_9066402, EPI_ISL_9066404, EPI_ISL_9066406, EPI_ISL_9066409, EPI_ISL_9066411, EPI_ISL_9066428, EPI_ISL_9066433, EPI_ISL_9066436, EPI_ISL_9066437, EPI_ISL_9066441, EPI_ISL_9066442, EPI_ISL_9066444, EPI_ISL_9066445, EPI_ISL_9066464, EPI_ISL_9066477, EPI_ISL_9066483, EPI_ISL_9066489, EPI_ISL_9066492, EPI_ISL_9066495, EPI_ISL_9066499, EPI_ISL_9066504, EPI_ISL_9066510, EPI_ISL_9066511, EPI_ISL_9066514, EPI_ISL_9066521, EPI_ISL_9066527, EPI_ISL_9066530, EPI_ISL_9066533, EPI_ISL_9066535, EPI_ISL_9066548, EPI_ISL_9066552, EPI_ISL_9066555, EPI_ISL_9066558, EPI_ISL_9066563, EPI_ISL_9066572, EPI_ISL_9066579, EPI_ISL_9066581, EPI_ISL_9066584, EPI_ISL_9066587, EPI_ISL_9066600, EPI_ISL_9066609, EPI_ISL_9066612, EPI_ISL_9066618, EPI_ISL_9066626, EPI_ISL_9066629, EPI_ISL_9066632, EPI_ISL_9066640, EPI_ISL_9066645, EPI_ISL_9066648, EPI_ISL_9066657, EPI_ISL_9066661, EPI_ISL_9066664, EPI_ISL_9066667, EPI_ISL_9066670, EPI_ISL_9066684, EPI_ISL_9066687, EPI_ISL_9066690, EPI_ISL_9066695, EPI_ISL_9066704, EPI_ISL_9066706, EPI_ISL_9066720, EPI_ISL_9066725, EPI_ISL_9066744, EPI_ISL_9066747, EPI_ISL_9066750, EPI_ISL_9066754, EPI_ISL_9066763, EPI_ISL_9066773, EPI_ISL_9066777, EPI_ISL_9066780, EPI_ISL_9066782, EPI_ISL_9066785, EPI_ISL_9066791, EPI_ISL_9066794, EPI_ISL_9066804, EPI_ISL_9066807, EPI_ISL_9066813, EPI_ISL_9066816, EPI_ISL_9066824, EPI_ISL_9066826, EPI_ISL_9066831, EPI_ISL_9066834, EPI_ISL_9066837, EPI_ISL_9066845, EPI_ISL_9066848, EPI_ISL_9066850, EPI_ISL_9066858, EPI_ISL_9066868, EPI_ISL_9066871, EPI_ISL_9066883, EPI_ISL_9066888, EPI_ISL_9066890, EPI_ISL_9066897, EPI_ISL_9066903, EPI_ISL_9066906, EPI_ISL_9066916, EPI_ISL_9066924, EPI_ISL_9066927, EPI_ISL_9066941, EPI_ISL_9066949, EPI_ISL_9066952, EPI_ISL_9066960, EPI_ISL_9066968, EPI_ISL_9066973, EPI_ISL_9066976, EPI_ISL_9066982, EPI_ISL_9066984, EPI_ISL_9066987, EPI_ISL_9066996, EPI_ISL_9067002, EPI_ISL_9067005, EPI_ISL_9067013, EPI_ISL_9067016, EPI_ISL_9067022, EPI_ISL_9067028, EPI_ISL_9067031, EPI_ISL_9067033, EPI_ISL_9067042, EPI_ISL_9067048, EPI_ISL_9067051, EPI_ISL_9067053, EPI_ISL_9067056, EPI_ISL_9067073, EPI_ISL_9067090, EPI_ISL_9067097, EPI_ISL_9067100, EPI_ISL_9067102, EPI_ISL_9067123, EPI_ISL_9067130, EPI_ISL_9067133, EPI_ISL_9067135, EPI_ISL_9067140, EPI_ISL_9067143, EPI_ISL_9067145, EPI_ISL_9067148, EPI_ISL_9067156, EPI_ISL_9067159, EPI_ISL_9067166, EPI_ISL_9067172, EPI_ISL_9067174, EPI_ISL_9067175, EPI_ISL_9067178, EPI_ISL_9067191, EPI_ISL_9067199, EPI_ISL_9067205, EPI_ISL_9067210, EPI_ISL_9067221, EPI_ISL_9067229, EPI_ISL_9067232, EPI_ISL_9067235, EPI_ISL_9067243, EPI_ISL_9067245, EPI_ISL_9067246, EPI_ISL_9067249, EPI_ISL_9067251, EPI_ISL_9067257, EPI_ISL_9067258, EPI_ISL_9067259, EPI_ISL_9067263, EPI_ISL_9067266, EPI_ISL_9067269, EPI_ISL_9067273, EPI_ISL_9067275, EPI_ISL_9067276, EPI_ISL_9067277, EPI_ISL_9067278, EPI_ISL_9067291, EPI_ISL_9067300, EPI_ISL_9067317, EPI_ISL_9067320, EPI_ISL_9067328, EPI_ISL_9067331, EPI_ISL_9067333, EPI_ISL_9067336, EPI_ISL_9067338, EPI_ISL_9067341, EPI_ISL_9067346, EPI_ISL_9067351, EPI_ISL_9067353, EPI_ISL_9067358, EPI_ISL_9067361, EPI_ISL_9067367, EPI_ISL_9067370, EPI_ISL_9067378, EPI_ISL_9067381, EPI_ISL_9067383, EPI_ISL_9067388, EPI_ISL_9067394, EPI_ISL_9067412, EPI_ISL_9067415, EPI_ISL_9067418, EPI_ISL_9067424, EPI_ISL_9067519, EPI_ISL_9067522, EPI_ISL_9067525, EPI_ISL_9067531, EPI_ISL_9067534, EPI_ISL_9067536, EPI_ISL_9067551, EPI_ISL_9067554, EPI_ISL_9067556, EPI_ISL_9067559, EPI_ISL_9067562, EPI_ISL_9067564, EPI_ISL_9067570, EPI_ISL_9067576, EPI_ISL_9067578, EPI_ISL_9067589, EPI_ISL_9067595, EPI_ISL_9067598, EPI_ISL_9067603, EPI_ISL_9067606, EPI_ISL_9067610, EPI_ISL_9067614, EPI_ISL_9067620, EPI_ISL_9067625, EPI_ISL_9067631, EPI_ISL_9067634, EPI_ISL_9067651, EPI_ISL_9067652, EPI_ISL_9067664, EPI_ISL_9067672, EPI_ISL_9067684, EPI_ISL_9067687, EPI_ISL_9067693, EPI_ISL_9067696, EPI_ISL_9067702, EPI_ISL_9067708, EPI_ISL_9067714, EPI_ISL_9067717, EPI_ISL_9067726, EPI_ISL_9067729, EPI_ISL_9067732, EPI_ISL_9067738, EPI_ISL_9067758, EPI_ISL_9067761, EPI_ISL_9067764, EPI_ISL_9067769, EPI_ISL_9067772, EPI_ISL_9067773, EPI_ISL_9067778, EPI_ISL_9067781, EPI_ISL_9067784, EPI_ISL_9067786, EPI_ISL_9067803, EPI_ISL_9067806, EPI_ISL_9067809, EPI_ISL_9067814, EPI_ISL_9067817, EPI_ISL_9067822, EPI_ISL_9067824, EPI_ISL_9067836, EPI_ISL_9067855, EPI_ISL_9067858, EPI_ISL_9067864, EPI_ISL_9067868, EPI_ISL_9067871, EPI_ISL_9067885, EPI_ISL_9067888, EPI_ISL_9067894, EPI_ISL_9067904, EPI_ISL_9067907, EPI_ISL_9067922, EPI_ISL_9067925, EPI_ISL_9067937, EPI_ISL_9067945, EPI_ISL_9067947, EPI_ISL_9067967, EPI_ISL_9067973, EPI_ISL_9067976, EPI_ISL_9067981, EPI_ISL_9067984, EPI_ISL_9067987, EPI_ISL_9067989, EPI_ISL_9067992, EPI_ISL_9068009, EPI_ISL_9068011, EPI_ISL_9068014, EPI_ISL_9068017, EPI_ISL_9068020, EPI_ISL_9068023, EPI_ISL_9068025, EPI_ISL_9068028, EPI_ISL_9068031, EPI_ISL_9068036, EPI_ISL_9068051, EPI_ISL_9068054, EPI_ISL_9068060, EPI_ISL_9068066, EPI_ISL_9068077, EPI_ISL_9068084, EPI_ISL_9068086, EPI_ISL_9068090, EPI_ISL_9068093, EPI_ISL_9068110, EPI_ISL_9068128, EPI_ISL_9068129, EPI_ISL_9068131, EPI_ISL_9068133, EPI_ISL_9068135, EPI_ISL_9068155, EPI_ISL_9068164, EPI_ISL_9068167, EPI_ISL_9068218, EPI_ISL_9068221, EPI_ISL_9068226, EPI_ISL_9068228, EPI_ISL_9068235, EPI_ISL_9068240, EPI_ISL_9068241, EPI_ISL_9068242, EPI_ISL_9068243, EPI_ISL_9068244, EPI_ISL_9068247, EPI_ISL_9068252, EPI_ISL_9068253, EPI_ISL_9068257, EPI_ISL_9068258, EPI_ISL_9068262, EPI_ISL_9068263, EPI_ISL_9068265, EPI_ISL_9068268, EPI_ISL_9068269, EPI_ISL_9068271, EPI_ISL_9068273, EPI_ISL_9068274, EPI_ISL_9068277, EPI_ISL_9068279, EPI_ISL_9068282, EPI_ISL_9068287, EPI_ISL_9068296, EPI_ISL_9068298, EPI_ISL_9068304, EPI_ISL_9068310, EPI_ISL_9068313, EPI_ISL_9068316, EPI_ISL_9068328, EPI_ISL_9068334, EPI_ISL_9068340, EPI_ISL_9068343, EPI_ISL_9068346, EPI_ISL_9068348, EPI_ISL_9068354, EPI_ISL_9068357, EPI_ISL_9068369, EPI_ISL_9068371, EPI_ISL_9068374, EPI_ISL_9068406, EPI_ISL_9068409, EPI_ISL_9068411, EPI_ISL_9068417, EPI_ISL_9068419, EPI_ISL_9068425, EPI_ISL_9068435, EPI_ISL_9068438, EPI_ISL_9068447, EPI_ISL_9068451, EPI_ISL_9068456, EPI_ISL_9068464, EPI_ISL_9068468, EPI_ISL_9068471, EPI_ISL_9068476, EPI_ISL_9068479, EPI_ISL_9068485, EPI_ISL_9068493, EPI_ISL_9068497, EPI_ISL_9068500, EPI_ISL_9068509, EPI_ISL_9068513, EPI_ISL_9068519, EPI_ISL_9068522, EPI_ISL_9068528, EPI_ISL_9068541, EPI_ISL_9068547, EPI_ISL_9068549, EPI_ISL_9068553, EPI_ISL_9068558, EPI_ISL_9068561, EPI_ISL_9068567, EPI_ISL_9068570, EPI_ISL_9068572, EPI_ISL_9068575, EPI_ISL_9068579, EPI_ISL_9068582, EPI_ISL_9068588, EPI_ISL_9068593, EPI_ISL_9068604, EPI_ISL_9068609, EPI_ISL_9068620, EPI_ISL_9068622, EPI_ISL_9068626, EPI_ISL_9068632 | | | |
| see above | Department of Bacteria, Parasites and Fungi, Statens Serum Institut, Copenhagen, Denmark | Statens Serum Institut Bioinformatics and Microbial Genomics | Danish Covid-19 Genome Consortium |

We gratefully acknowledge the following Authors from the Originating laboratories responsible for obtaining the specimens, as well as the Submitting laboratories where the genome data were generated and shared via GISAID, on which this research is based.

All Submitters of data may be contacted directly via www.gisaid.org

Authors are sorted alphabetically.

| Accession ID | Originating Laboratory | Submitting Laboratory | Authors |
|---|---|---|---|
| EPI_ISL_9080634, EPI_ISL_9080635, EPI_ISL_9085223 | Dutch COVID-19 response team | National Institute for Public Health and the Environment (RIVM) | Adam Meijer; Afke Vogelzang; AnneMarie van den Brandt; Annelies Kroneman; Bas van der Veer; Chantal Reusken; Dennis Schmitz; Dirk Eggink; Florian Zwagemaker; Harry Vennema; Ivo van Walle; Jeroen Cremer; Jil Kocken; Jordy de Bakker; Karim Hajji; Kim Freriks; Linda van Someren; Lisa Wijsman; Lynn Aarts; Ryanne Jaarsma; Sanne Bos; Sharon van den Brink; on behalf of the national COVID-19 response team |
| EPI_ISL_9087313, EPI_ISL_9087316, EPI_ISL_9089615 | Hopital | National Reference Center for Viruses of Respiratory Infections, Institut Pasteur, Paris | Angela Brisebarre; Camille Capel; Christophe Malabat; Corinne Maufrais; Etienne Simon-Lorière; Frédéric Lemoine; Julien Fumey; Louise Lefrançois; Mama Imene FODIL PACHA; Marion Barbet; Maud Vanpeene; Méline Bizard; Slim El Khiari; Sylvie Behillil; Sylvie Van der Werf; Vincent ESTEVE; Vincent Enouf |
| EPI_ISL_9089637 | Labo Analyses Med | National Reference Center for Viruses of Respiratory Infections, Institut Pasteur, Paris | Angela Brisebarre; Camille Capel; Christophe Malabat; Corinne Maufrais; Etienne Simon-Lorière; Frédéric Lemoine; Julien Fumey; Louise Lefrançois; Marion Barbet; Maud Vanpeene; Méline Bizard; Slim El Khiari; Sylvie Behillil; Sylvie Van der Werf; Thierry GUFFOND; Vincent Enouf |
| EPI_ISL_9074304, EPI_ISL_9074867 | MVZ Labor Dr. Fenner und Kollegen (Standort Hamburg) | Robert Koch Institute | |
| EPI_ISL_9083481 | Platform BIS UZA/UAntwerpen | Labo Klinische Biologie, UZA | Basil Britto Xavier; Christine Lammens; Herman Goossens; Ines Verbesselt; Jasmine Coppens; Kathleen Holemans; Marie Le Mercier; Veerle Matheeussen |
| EPI_ISL_9084574, EPI_ISL_9084575, EPI_ISL_9084576, EPI_ISL_9084577, EPI_ISL_9084578, EPI_ISL_9084580, EPI_ISL_9084581, EPI_ISL_9084583, EPI_ISL_9084584, EPI_ISL_9084585 | | | |
| see above | SARS-CoV-2 Sequencing Castilla y Leon-Spain Consortium | SARS-CoV-2 Sequencing Castilla y Leon-Spain Consortium | Antonio Orduña-Domingo; Carlos Fuster Foz; Carmen Aldea-Mansilla; Carmen Gimeno Crespo; David Abad; Gabriel March Rosello; Gregoria Megías Lobón; Jose María Eiros Bouza; M. Isabel Fernandez-Natal; Marta Dominguez-Gil; Marta Hernandez; María Antonia García Castro; Mª Fe Brezmes-Valdivieso; Noelia Arenal Andrés; Silvia Rojo; Sonsoles Garcinuño Pérez |
| EPI_ISL_9080382 | SYNLAB MVZ Leverkusen | Robert Koch Institute | |
| EPI_ISL_9075006, EPI_ISL_9075007, EPI_ISL_9075008, EPI_ISL_9075009, EPI_ISL_9075010, EPI_ISL_9075011, EPI_ISL_9075012, EPI_ISL_9075013, EPI_ISL_9075014, EPI_ISL_9075015, EPI_ISL_9075016, EPI_ISL_9075017, EPI_ISL_9075018, EPI_ISL_9075019, EPI_ISL_9075020, EPI_ISL_9075021, EPI_ISL_9075022, EPI_ISL_9075023, EPI_ISL_9075024, EPI_ISL_9075025, EPI_ISL_9075026, EPI_ISL_9075027, EPI_ISL_9075028, EPI_ISL_9075029, EPI_ISL_9075030, EPI_ISL_9075031 | | | |
| see above | University Clinical Hospital of Mostar, Department of Microbiology and Molecular Diagnostics | University of Sarajevo, Veterinary Faculty, Laboratory for Molecular Diagnostic and Research | Goletic S; Goletic T; Herceg I; Hodzic A; Jazic A; Nicevic M; Ostojic M; Sabic E; Skocibusic S; Softic A; Terzic I |
| EPI_ISL_9075003, EPI_ISL_9075004, EPI_ISL_9075005 | University of Sarajevo, Veterinary Faculty, Laboratory for Molecular Diagnostic and Research | University of Sarajevo, Veterinary Faculty, Laboratory for Molecular Diagnostic and Research | Goletic S; Goletic T; Hodzic A; Jazic A; Nicevic M; Sabic E; Softic A; Terzic I |

We gratefully acknowledge the following Authors from the Originating laboratories responsible for obtaining the specimens, as well as the Submitting laboratories where the genome data were generated and shared via GISAID, on which this research is based.

All Submitters of data may be contacted directly via www.gisaid.org

Authors are sorted alphabetically.

| Accession ID | Originating Laboratory | Submitting Laboratory | Authors |
|---|---|---|---|
| EPI_ISL_9071873, EPI_ISL_9071899, EPI_ISL_9071904, EPI_ISL_9071938, EPI_ISL_9071941, EPI_ISL_9071943, EPI_ISL_9071944, EPI_ISL_9071958, EPI_ISL_9071965, EPI_ISL_9071977, EPI_ISL_9071986, EPI_ISL_9071996 | | | |
| see above | Bayerisches Landesamt fuer Gesundheit und Lebensmittelsicherheit (LGL) | Bayerisches Landesamt fuer Gesundheit und Lebensmittelsicherheit (LGL) | Alexandra Dangel; Andreas Sing; Annika Sprenger; Carola Berger; Laura Weise; Nikolaus Ackermann; Sabrina Hepner |
| EPI_ISL_9070769, EPI_ISL_9070771, EPI_ISL_9070778, EPI_ISL_9070785, EPI_ISL_9070787, EPI_ISL_9070788, EPI_ISL_9070801, EPI_ISL_9070869, EPI_ISL_9070870, EPI_ISL_9070874, EPI_ISL_9070877, EPI_ISL_9070879, EPI_ISL_9070882, EPI_ISL_9070892, EPI_ISL_9070894, EPI_ISL_9070903, EPI_ISL_9070906, EPI_ISL_9070912, EPI_ISL_9070915, EPI_ISL_9070917, EPI_ISL_9070937, EPI_ISL_9070940, EPI_ISL_9070943, EPI_ISL_9070945, EPI_ISL_9070948, EPI_ISL_9070951, EPI_ISL_9070957, EPI_ISL_9070967, EPI_ISL_9070970, EPI_ISL_9070989, EPI_ISL_9071008, EPI_ISL_9071015, EPI_ISL_9071018, EPI_ISL_9071021, EPI_ISL_9071022, EPI_ISL_9071023, EPI_ISL_9071024, EPI_ISL_9071025 | | | |
| see above | Department of Bacteria, Parasites and Fungi, Statens Serum Institut, Copenhagen, Denmark | Statens Serum Institut Bioinformatics and Microbial Genomics | Danish Covid-19 Genome Consortium |

We gratefully acknowledge the following Authors from the Originating laboratories responsible for obtaining the specimens, as well as the Submitting laboratories where the genome data were generated and shared via GISAID, on which this research is based.

All Submitters of data may be contacted directly via www.gisaid.org

Authors are sorted alphabetically.

Acknowledgement EPI_SET Identifier: EPI_SET_20220527ae

| Accession ID | Originating Laboratory | Submitting Laboratory | Authors |
|---|---|---|---|
| EPI_ISL_9070416, EPI_ISL_9070422, EPI_ISL_9070423, EPI_ISL_9070434, EPI_ISL_9070437, EPI_ISL_9070439, EPI_ISL_9070451, EPI_ISL_9070459, EPI_ISL_9070464, EPI_ISL_9070469, EPI_ISL_9070480, EPI_ISL_9070482, EPI_ISL_9070488, EPI_ISL_9070493, EPI_ISL_9070495, EPI_ISL_9070517, EPI_ISL_9070540, EPI_ISL_9070542, EPI_ISL_9070543, EPI_ISL_9070596, EPI_ISL_9070601, EPI_ISL_9070606, EPI_ISL_9070612, EPI_ISL_9070618, EPI_ISL_9070629, EPI_ISL_9070634, EPI_ISL_9070644, EPI_ISL_9070647, EPI_ISL_9070650, EPI_ISL_9070653, EPI_ISL_9070663, EPI_ISL_9070666, EPI_ISL_9070675, EPI_ISL_9070676, EPI_ISL_9070677, EPI_ISL_9070678, EPI_ISL_9070685, EPI_ISL_9070690, EPI_ISL_9070695, EPI_ISL_9070698, EPI_ISL_9070704, EPI_ISL_9070712, EPI_ISL_9070722, EPI_ISL_9070737, EPI_ISL_9070740, EPI_ISL_9070748, EPI_ISL_9070751, EPI_ISL_9070753, EPI_ISL_9070759, EPI_ISL_9070760 | | | |
| see above | Department of Bacteria, Parasites and Fungi, Statens Serum Institut, Copenhagen, Denmark | Statens Serum Institut Bioinformatics and Microbial Genomics | Danish Covid-19 Genome Consortium |

We gratefully acknowledge the following Authors from the Originating laboratories responsible for obtaining the specimens, as well as the Submitting laboratories where the genome data were generated and shared via GISAID, on which this research is based.

All Submitters of data may be contacted directly via www.gisaid.org

Authors are sorted alphabetically.

| Accession ID | Originating Laboratory | Submitting Laboratory | Authors |
|---|---|---|---|
| EPI_ISL_9070067, EPI_ISL_9070080, EPI_ISL_9070083, EPI_ISL_9070087, EPI_ISL_9070091, EPI_ISL_9070092, EPI_ISL_9070099, EPI_ISL_9070102, EPI_ISL_9070105, EPI_ISL_9070111, EPI_ISL_9070132, EPI_ISL_9070163, EPI_ISL_9070171, EPI_ISL_9070174, EPI_ISL_9070179, EPI_ISL_9070183, EPI_ISL_9070188, EPI_ISL_9070189, EPI_ISL_9070190, EPI_ISL_9070191, EPI_ISL_9070192, EPI_ISL_9070198, EPI_ISL_9070201, EPI_ISL_9070211, EPI_ISL_9070217, EPI_ISL_9070220, EPI_ISL_9070222, EPI_ISL_9070232, EPI_ISL_9070238, EPI_ISL_9070264, EPI_ISL_9070270, EPI_ISL_9070273, EPI_ISL_9070322, EPI_ISL_9070325, EPI_ISL_9070331, EPI_ISL_9070333, EPI_ISL_9070342, EPI_ISL_9070347, EPI_ISL_9070351, EPI_ISL_9070358, EPI_ISL_9070362, EPI_ISL_9070372, EPI_ISL_9070375, EPI_ISL_9070380, EPI_ISL_9070383, EPI_ISL_9070391, EPI_ISL_9070394, EPI_ISL_9070408, EPI_ISL_9070410, EPI_ISL_9070413 | | | |
| see above | Department of Bacteria, Parasites and Fungi, Statens Serum Institut, Copenhagen, Denmark | Statens Serum Institut Bioinformatics and Microbial Genomics | Danish Covid-19 Genome Consortium |

We gratefully acknowledge the following Authors from the Originating laboratories responsible for obtaining the specimens, as well as the Submitting laboratories where the genome data were generated and shared via GISAID, on which this research is based.

All Submitters of data may be contacted directly via www.gisaid.org

Authors are sorted alphabetically.

| Accession ID | Originating Laboratory | Submitting Laboratory | Authors |
|---|---|---|---|
| EPI_ISL_9069764, EPI_ISL_9069765, EPI_ISL_9069767, EPI_ISL_9069769, EPI_ISL_9069776, EPI_ISL_9069781, EPI_ISL_9069784, EPI_ISL_9069791, EPI_ISL_9069793, EPI_ISL_9069797, EPI_ISL_9069802, EPI_ISL_9069805, EPI_ISL_9069816, EPI_ISL_9069822, EPI_ISL_9069825, EPI_ISL_9069828, EPI_ISL_9069833, EPI_ISL_9069838, EPI_ISL_9069847, EPI_ISL_9069858, EPI_ISL_9069861, EPI_ISL_9069863, EPI_ISL_9069868, EPI_ISL_9069873, EPI_ISL_9069882, EPI_ISL_9069897, EPI_ISL_9069903, EPI_ISL_9069907, EPI_ISL_9069910, EPI_ISL_9069911, EPI_ISL_9069913, EPI_ISL_9069921, EPI_ISL_9069929, EPI_ISL_9069931, EPI_ISL_9069934, EPI_ISL_9069940, EPI_ISL_9069943, EPI_ISL_9069950, EPI_ISL_9069955, EPI_ISL_9069968, EPI_ISL_9069971, EPI_ISL_9069974, EPI_ISL_9069977, EPI_ISL_9069979, EPI_ISL_9069982, EPI_ISL_9069984, EPI_ISL_9069991, EPI_ISL_9069993, EPI_ISL_9070001, EPI_ISL_9070006 | | | |
| see above | Department of Bacteria, Parasites and Fungi, Statens Serum Institut, Copenhagen, Denmark | Statens Serum Institut Bioinformatics and Microbial Genomics | Danish Covid-19 Genome Consortium |

We gratefully acknowledge the following Authors from the Originating laboratories responsible for obtaining the specimens, as well as the Submitting laboratories where the genome data were generated and shared via GISAID, on which this research is based.

All Submitters of data may be contacted directly via www.gisaid.org

Authors are sorted alphabetically.

Acknowledgement EPI_SET Identifier: EPI_SET_20220527pf

| Accession ID | Originating Laboratory | Submitting Laboratory | Authors |
|---|---|---|---|
| EPI_ISL_9069357, EPI_ISL_9069364, EPI_ISL_9069367, EPI_ISL_9069370, EPI_ISL_9069379, EPI_ISL_9069393, EPI_ISL_9069396, EPI_ISL_9069397, EPI_ISL_9069400, EPI_ISL_9069403, EPI_ISL_9069406, EPI_ISL_9069409, EPI_ISL_9069412, EPI_ISL_9069415, EPI_ISL_9069427, EPI_ISL_9069433, EPI_ISL_9069436, EPI_ISL_9069439, EPI_ISL_9069442, EPI_ISL_9069444, EPI_ISL_9069450, EPI_ISL_9069465, EPI_ISL_9069487, EPI_ISL_9069492, EPI_ISL_9069495, EPI_ISL_9069500, EPI_ISL_9069503, EPI_ISL_9069509, EPI_ISL_9069512, EPI_ISL_9069523, EPI_ISL_9069526, EPI_ISL_9069534, EPI_ISL_9069539, EPI_ISL_9069550, EPI_ISL_9069576, EPI_ISL_9069587, EPI_ISL_9069592, EPI_ISL_9069597, EPI_ISL_9069598, EPI_ISL_9069602, EPI_ISL_9069607, EPI_ISL_9069614, EPI_ISL_9069616, EPI_ISL_9069617, EPI_ISL_9069624, EPI_ISL_9069627, EPI_ISL_9069633, EPI_ISL_9069666, EPI_ISL_9069672, EPI_ISL_9069741 | | | |
| see above | Department of Bacteria, Parasites and Fungi, Statens Serum Institut, Copenhagen, Denmark | Statens Serum Institut Bioinformatics and Microbial Genomics | Danish Covid-19 Genome Consortium |

We gratefully acknowledge the following Authors from the Originating laboratories responsible for obtaining the specimens, as well as the Submitting laboratories where the genome data were generated and shared via GISAID, on which this research is based.

All Submitters of data may be contacted directly via www.gisaid.org

Authors are sorted alphabetically.

| Accession ID | Originating Laboratory | Submitting Laboratory | Authors |
| --- | --- | --- | --- |
| EPI_ISL_9068982, EPI_ISL_9068985, EPI_ISL_9069000, EPI_ISL_9069003, EPI_ISL_9069005, EPI_ISL_9069014, EPI_ISL_9069017, EPI_ISL_9069020, EPI_ISL_9069023, EPI_ISL_9069025, EPI_ISL_9069036, EPI_ISL_9069044, EPI_ISL_9069047, EPI_ISL_9069050, EPI_ISL_9069059, EPI_ISL_9069062, EPI_ISL_9069081, EPI_ISL_9069087, EPI_ISL_9069088, EPI_ISL_9069102, EPI_ISL_9069104, EPI_ISL_9069110, EPI_ISL_9069128, EPI_ISL_9069149, EPI_ISL_9069171, EPI_ISL_9069180, EPI_ISL_9069198, EPI_ISL_9069203, EPI_ISL_9069215, EPI_ISL_9069220, EPI_ISL_9069223, EPI_ISL_9069241, EPI_ISL_9069243, EPI_ISL_9069259, EPI_ISL_9069264, EPI_ISL_9069275, EPI_ISL_9069278, EPI_ISL_9069280, EPI_ISL_9069293, EPI_ISL_9069298, EPI_ISL_9069309, EPI_ISL_9069312, EPI_ISL_9069316, EPI_ISL_9069319, EPI_ISL_9069322, EPI_ISL_9069334, EPI_ISL_9069346, EPI_ISL_9069349, EPI_ISL_9069352, EPI_ISL_9069355 | | | |
| see above | Department of Bacteria, Parasites and Fungi, Statens Serum Institut, Copenhagen, Denmark | Statens Serum Institut Bioinformatics and Microbial Genomics | Danish Covid-19 Genome Consortium |

We gratefully acknowledge the following Authors from the Originating laboratories responsible for obtaining the specimens, as well as the Submitting laboratories where the genome data were generated and shared via GISAID, on which this research is based.

All Submitters of data may be contacted directly via www.gisaid.org

Authors are sorted alphabetically.

| Accession ID | Originating Laboratory | Submitting Laboratory | Authors |
|---|---|---|---|
| EPI_ISL_9068641, EPI_ISL_9068656, EPI_ISL_9068658, EPI_ISL_9068660, EPI_ISL_9068661, EPI_ISL_9068662, EPI_ISL_9068663, EPI_ISL_9068675, EPI_ISL_9068691, EPI_ISL_9068724, EPI_ISL_9068727, EPI_ISL_9068747, EPI_ISL_9068749, EPI_ISL_9068752, EPI_ISL_9068755, EPI_ISL_9068764, EPI_ISL_9068766, EPI_ISL_9068773, EPI_ISL_9068786, EPI_ISL_9068789, EPI_ISL_9068798, EPI_ISL_9068801, EPI_ISL_9068804, EPI_ISL_9068807, EPI_ISL_9068814, EPI_ISL_9068819, EPI_ISL_9068824, EPI_ISL_9068833, EPI_ISL_9068839, EPI_ISL_9068842, EPI_ISL_9068856, EPI_ISL_9068859, EPI_ISL_9068869, EPI_ISL_9068872, EPI_ISL_9068875, EPI_ISL_9068887, EPI_ISL_9068890, EPI_ISL_9068900, EPI_ISL_9068918, EPI_ISL_9068927, EPI_ISL_9068935, EPI_ISL_9068944, EPI_ISL_9068947, EPI_ISL_9068950, EPI_ISL_9068959, EPI_ISL_9068962, EPI_ISL_9068968, EPI_ISL_9068974, EPI_ISL_9068979 | | | |
| see above | Department of Bacteria, Parasites and Fungi, Statens Serum Institut, Copenhagen, Denmark | Statens Serum Institut Bioinformatics and Microbial Genomics | Danish Covid-19 Genome Consortium |
| EPI_ISL_9068734 | Lighthouse Lab in Milton Keynes | Wellcome Sanger Institute for the COVID-19 Genomics UK (COG-UK) Consortium | Cordelia Langford; David K. Jackson; Dominic Kwiatkowski; Ewan Harrison; Ian Johnston; Jeffrey Barrett; John Sillitoe on behalf of the Wellcome Sanger Institute COVID-19 Surveillance Team; Roberto Amato; Sonia Goncalves; The Lighthouse Lab in Milton Keynes and Alex Alderton |

We gratefully acknowledge the following Authors from the Originating laboratories responsible for obtaining the specimens, as well as the Submitting laboratories where the genome data were generated and shared via GISAID, on which this research is based.

All Submitters of data may be contacted directly via www.gisaid.org

Authors are sorted alphabetically.

Acknowledgement EPI_SET Identifier: EPI_SET_20220527zt

| Accession ID | Originating Laboratory | Submitting Laboratory | Authors |
|---|---|---|---|
| EPI_ISL_9149786 | National Health Laboratory Services, Virology | National Health Laboratory Services, Virology | Avani Bharuthram; Florette K. Treurnicht; Kathleen Subramoney; Nkhensani Mtileni |

We gratefully acknowledge the following Authors from the Originating laboratories responsible for obtaining the specimens, as well as the Submitting laboratories where the genome data were generated and shared via GISAID, on which this research is based.

All Submitters of data may be contacted directly via www.gisaid.org

Authors are sorted alphabetically.

| Accession ID | Originating Laboratory | Submitting Laboratory | Authors |
|---|---|---|---|
| EPI_ISL_9102258 | Ramathibodi Hospital | Center for Medical Genomics Ramathibodi Hospital | Anek mungaomklang; Insee Sensorn; Inthuorn Ratchatasetthakul; Kanyarat Janthima; Parsakorn Tapaopong; Poramate Jiaranai; Vichapon Tiacharoen; Wasun Chantratita |

We gratefully acknowledge the following Authors from the Originating laboratories responsible for obtaining the specimens, as well as the Submitting laboratories where the genome data were generated and shared via GISAID, on which this research is based.

All Submitters of data may be contacted directly via www.gisaid.org

Authors are sorted alphabetically.

| Accession ID | Originating Laboratory | Submitting Laboratory | Authors |
|---|---|---|---|
| EPI_ISL_9139503 | Aegis Sciences Corporation | Centers for Disease Control and Prevention Division of Viral Diseases, Pathogen Discovery | Alec Vest; Benjamin Rambo-Martin; Christopher Gulvick; Clinton Paden; Cyndi Clark; Dakota Howard; Dhwani Batra; Dillon Nall; Duncan MacCannell; Erisa Sula; Ethan Sanders; Holly Houdeshell; Jason Caravas; Kristine Lacek; Matthew Hardison; Matthew Schmerer; Ola Kvalvaag; Patrick Campbell; Peter Cook; Rob Case; Scott Sammons; Shatavia Morrison; Shaun Westlund; Tymeckia Kendall; Victoria Caban Figueroa; Vikramsinha Ghorpade; Yvette Unoarumhi |
| EPI_ISL_9129287 | Centers for Disease Control and Prevention, Dengue Branch | Centers for Disease Control and Prevention, Dengue Branch | Betzabel Flores; Gabriela Paz-Bailey; Gilberto A. Santiago; Glenda Gonzalez; Jorge L. Munoz-Jordan; Keyla Charriez |
| EPI_ISL_9112401, EPI_ISL_9112402, EPI_ISL_9112403, EPI_ISL_9112404, EPI_ISL_9112406, EPI_ISL_9112408, EPI_ISL_9112409, EPI_ISL_9112410, EPI_ISL_9112413, EPI_ISL_9112414, EPI_ISL_9112417, EPI_ISL_9112418, EPI_ISL_9112419, EPI_ISL_9112420, EPI_ISL_9112423, EPI_ISL_9112424, EPI_ISL_9112427, EPI_ISL_9112428, EPI_ISL_9112429, EPI_ISL_9112430, EPI_ISL_9112431, EPI_ISL_9112433, EPI_ISL_9112434, EPI_ISL_9112435, EPI_ISL_9112437, EPI_ISL_9112438, EPI_ISL_9112439, EPI_ISL_9112441, EPI_ISL_9112443, EPI_ISL_9112445, EPI_ISL_9112447, EPI_ISL_9112449, EPI_ISL_9112450, EPI_ISL_9112451, EPI_ISL_9112452, EPI_ISL_9112453, EPI_ISL_9112454, EPI_ISL_9112455, EPI_ISL_9112456, EPI_ISL_9112457, EPI_ISL_9112460, EPI_ISL_9112461, EPI_ISL_9112462, EPI_ISL_9112463, EPI_ISL_9112469, EPI_ISL_9112471, EPI_ISL_9112473, EPI_ISL_9112474, EPI_ISL_9112475, EPI_ISL_9112476, EPI_ISL_9112477, EPI_ISL_9112478, EPI_ISL_9112479, EPI_ISL_9112480, EPI_ISL_9112481, EPI_ISL_9112482, EPI_ISL_9112484, EPI_ISL_9112485, EPI_ISL_9112486, EPI_ISL_9112487, EPI_ISL_9112488 | | | |
| see above | Idaho Bureau of Laboratories | Idaho Bureau of Laboratories | Aimee Ceniseros; Christian Loera; Christopher Ball; Matthew Charles Burns; R. Beukelman; Robert L. Voermans |
| EPI_ISL_9100723, EPI_ISL_9100728, EPI_ISL_9100744, EPI_ISL_9100745, EPI_ISL_9100748, EPI_ISL_9100751, EPI_ISL_9100752, EPI_ISL_9100753, EPI_ISL_9100756, EPI_ISL_9100757, EPI_ISL_9100759, EPI_ISL_9100760, EPI_ISL_9100761, EPI_ISL_9100763, EPI_ISL_9100769, EPI_ISL_9100774, EPI_ISL_9100776, EPI_ISL_9100778, EPI_ISL_9100783, EPI_ISL_9100784, EPI_ISL_9100785, EPI_ISL_9100786, EPI_ISL_9100787 | | | |
| see above | Premier Laboratory Servic | BioInfoExperts | Cheryl Metzler; David Nolan; Rebecca Rose; Susanna Lamers; Valerie Murray |
| EPI_ISL_9132681, EPI_ISL_9132682, EPI_ISL_9133195 | Quadrant Laboratories, LLC | SUNY Upstate Medical University | Brian Pavlovitz; Frank Middleton |
| EPI_ISL_9114519 | University of Michigan Clinical Microbiology Laboratory | Lauring Lab, University of Michigan, Department of Microbiology and Immunology | Gilbert |
| EPI_ISL_9135223 | University of Wisconsin-Madison AIDS Vaccine Research Laboratories | University of Wisconsin-Madison AIDS Vaccine Research Laboratories | Gage Moreno; Katarina Braun; et al. AIDS Vaccine Research Laboratories |
| EPI_ISL_9103201, EPI_ISL_9105109, EPI_ISL_9105124, EPI_ISL_9150547, EPI_ISL_9150551 | Utah Public Health Laboratory | Utah Public Health Laboratory | Erin L. Young; John Arnn; Kelly F. Oakeson; Olinto Linares-Perdomo; Pooja Gupta; Tom Iverson |

We gratefully acknowledge the following Authors from the Originating laboratories responsible for obtaining the specimens, as well as the Submitting laboratories where the genome data were generated and shared via GISAID, on which this research is based.

All Submitters of data may be contacted directly via www.gisaid.org

Authors are sorted alphabetically.

| Accession ID | Originating Laboratory | Submitting Laboratory | Authors |
| --- | --- | --- | --- |
| EPI_ISL_9119542, EPI_ISL_9119561, EPI_ISL_9119575, EPI_ISL_9119610, EPI_ISL_9119657, EPI_ISL_9119661, EPI_ISL_9119696, EPI_ISL_9119697, EPI_ISL_9119707, EPI_ISL_9119708, EPI_ISL_9119722, EPI_ISL_9119730, EPI_ISL_9119735, EPI_ISL_9119738, EPI_ISL_9119741, EPI_ISL_9119743, EPI_ISL_9119744, EPI_ISL_9119750, EPI_ISL_9119755, EPI_ISL_9119769, EPI_ISL_9119770, EPI_ISL_9119783, EPI_ISL_9119790, EPI_ISL_9119804, EPI_ISL_9119811, EPI_ISL_9119814, EPI_ISL_9119833, EPI_ISL_9119835, EPI_ISL_9119839, EPI_ISL_9119842, EPI_ISL_9119846, EPI_ISL_9119852, EPI_ISL_9119863, EPI_ISL_9119867, EPI_ISL_9119870, EPI_ISL_9119879, EPI_ISL_9119884, EPI_ISL_9119890, EPI_ISL_9119899, EPI_ISL_9119904, EPI_ISL_9119911, EPI_ISL_9119912, EPI_ISL_9119922, EPI_ISL_9119947, EPI_ISL_9119950, EPI_ISL_9119960, EPI_ISL_9119985, EPI_ISL_9119994, EPI_ISL_9120001, EPI_ISL_9120029, EPI_ISL_9120037, EPI_ISL_9120040, EPI_ISL_9120045, EPI_ISL_9120055, EPI_ISL_9120063, EPI_ISL_9120064, EPI_ISL_9120078, EPI_ISL_9120079, EPI_ISL_9120083, EPI_ISL_9120087, EPI_ISL_9120093, EPI_ISL_9120100, EPI_ISL_9120114, EPI_ISL_9120123, EPI_ISL_9120142, EPI_ISL_9120143, EPI_ISL_9120157, EPI_ISL_9120159, EPI_ISL_9120167, EPI_ISL_9120194, EPI_ISL_9120211, EPI_ISL_9120213, EPI_ISL_9120220, EPI_ISL_9120229, EPI_ISL_9120243, EPI_ISL_9120244, EPI_ISL_9120247, EPI_ISL_9120261, EPI_ISL_9120264, EPI_ISL_9120266, EPI_ISL_9120301, EPI_ISL_9120470, EPI_ISL_9120478, EPI_ISL_9120482, EPI_ISL_9120485, EPI_ISL_9120488, EPI_ISL_9120505, EPI_ISL_9120507, EPI_ISL_9120525, EPI_ISL_9120527, EPI_ISL_9120535, EPI_ISL_9120549, EPI_ISL_9120558, EPI_ISL_9120561, EPI_ISL_9120565, EPI_ISL_9120579, EPI_ISL_9120582, EPI_ISL_9120599, EPI_ISL_9120606, EPI_ISL_9120608, EPI_ISL_9120617, EPI_ISL_9120627, EPI_ISL_9120629, EPI_ISL_9120636, EPI_ISL_9120643, EPI_ISL_9120646, EPI_ISL_9120649, EPI_ISL_9120650, EPI_ISL_9121019, EPI_ISL_9121022, EPI_ISL_9121049, EPI_ISL_9121051, EPI_ISL_9121065, EPI_ISL_9121081, EPI_ISL_9121086, EPI_ISL_9121096, EPI_ISL_9121098, EPI_ISL_9121108, EPI_ISL_9121121, EPI_ISL_9121122, EPI_ISL_9121135, EPI_ISL_9121175, EPI_ISL_9121182, EPI_ISL_9121184, EPI_ISL_9121198, EPI_ISL_9121210, EPI_ISL_9121211, EPI_ISL_9121232, EPI_ISL_9121238, EPI_ISL_9121239, EPI_ISL_9121250, EPI_ISL_9121264, EPI_ISL_9121273, EPI_ISL_9121278, EPI_ISL_9121298, EPI_ISL_9121309, EPI_ISL_9121338, EPI_ISL_9121339, EPI_ISL_9121343, EPI_ISL_9121350, EPI_ISL_9121352, EPI_ISL_9121354, EPI_ISL_9121357, EPI_ISL_9121358, EPI_ISL_9121364, EPI_ISL_9121368, EPI_ISL_9121375, EPI_ISL_9121379, EPI_ISL_9121380, EPI_ISL_9121381, EPI_ISL_9121389, EPI_ISL_9121396, EPI_ISL_9121405, EPI_ISL_9121409, EPI_ISL_9121418, EPI_ISL_9121421, EPI_ISL_9121426, EPI_ISL_9121431, EPI_ISL_9121434, EPI_ISL_9121442, EPI_ISL_9121461, EPI_ISL_9121487, EPI_ISL_9121500, EPI_ISL_9121508, EPI_ISL_9121516, EPI_ISL_9121529, EPI_ISL_9121533, EPI_ISL_9121542, EPI_ISL_9121562, EPI_ISL_9121564, EPI_ISL_9121565, EPI_ISL_9121569, EPI_ISL_9121579, EPI_ISL_9121583, EPI_ISL_9121590, EPI_ISL_9121620, EPI_ISL_9121625, EPI_ISL_9121629, EPI_ISL_9121641, EPI_ISL_9121652, EPI_ISL_9121653, EPI_ISL_9121691, EPI_ISL_9121725, EPI_ISL_9121751, EPI_ISL_9121752, EPI_ISL_9121754, EPI_ISL_9121766, EPI_ISL_9121778, EPI_ISL_9121780, EPI_ISL_9121784, EPI_ISL_9121785, EPI_ISL_9121802, EPI_ISL_9121803, EPI_ISL_9121856, EPI_ISL_9121858, EPI_ISL_9121875, EPI_ISL_9121880, EPI_ISL_9121900, EPI_ISL_9121907, EPI_ISL_9121909, EPI_ISL_9121913, EPI_ISL_9121947, EPI_ISL_9122019, EPI_ISL_9122024, EPI_ISL_9122032, EPI_ISL_9122034, EPI_ISL_9122044, EPI_ISL_9122046, EPI_ISL_9122059, EPI_ISL_9122069, EPI_ISL_9122077, EPI_ISL_9122078, EPI_ISL_9122082, EPI_ISL_9122086, EPI_ISL_9122087, EPI_ISL_9122089, EPI_ISL_9122111, EPI_ISL_9122123, EPI_ISL_9122130, EPI_ISL_9122142, EPI_ISL_9122170, EPI_ISL_9122176, EPI_ISL_9122938, EPI_ISL_9122949, EPI_ISL_9122950, EPI_ISL_9122968, EPI_ISL_9122990, EPI_ISL_9122995, EPI_ISL_9122996, EPI_ISL_9123113, EPI_ISL_9123118, EPI_ISL_9123124, EPI_ISL_9123126, EPI_ISL_9123163, EPI_ISL_9123165, EPI_ISL_9123166, EPI_ISL_9123168, EPI_ISL_9123173, EPI_ISL_9123177, EPI_ISL_9123182, EPI_ISL_9123190, EPI_ISL_9123194, EPI_ISL_9123195, EPI_ISL_9123197, EPI_ISL_9123205, EPI_ISL_9123208, EPI_ISL_9123221, EPI_ISL_9123254, EPI_ISL_9123275, EPI_ISL_9123280, EPI_ISL_9123309, EPI_ISL_9123317, EPI_ISL_9123324, EPI_ISL_9123327, EPI_ISL_9123333, EPI_ISL_9123334, EPI_ISL_9123341, EPI_ISL_9123349, EPI_ISL_9123359, EPI_ISL_9123374, EPI_ISL_9123376, EPI_ISL_9123377, EPI_ISL_9123384, EPI_ISL_9123386, EPI_ISL_9123389, EPI_ISL_9123404, EPI_ISL_9123412, EPI_ISL_9123413, EPI_ISL_9123415, EPI_ISL_9123419, EPI_ISL_9123425, EPI_ISL_9123440, EPI_ISL_9123450, EPI_ISL_9123452, EPI_ISL_9123457, EPI_ISL_9123458, EPI_ISL_9123461, EPI_ISL_9123463, EPI_ISL_9123465, EPI_ISL_9123469, EPI_ISL_9123494, EPI_ISL_9123499, EPI_ISL_9123513, EPI_ISL_9123536, EPI_ISL_9123565, EPI_ISL_9123760, EPI_ISL_9123775, EPI_ISL_9123782, EPI_ISL_9123788, EPI_ISL_9123791, EPI_ISL_9124001, EPI_ISL_9124014, EPI_ISL_9124020, EPI_ISL_9124024, EPI_ISL_9124037, EPI_ISL_9124043, EPI_ISL_9124055, EPI_ISL_9124086, EPI_ISL_9124109, EPI_ISL_9124111, EPI_ISL_9124126, EPI_ISL_9124129, EPI_ISL_9124153, EPI_ISL_9124174, EPI_ISL_9124176, EPI_ISL_9124181, EPI_ISL_9124183, EPI_ISL_9124192, EPI_ISL_9124195, EPI_ISL_9124209, EPI_ISL_9124225, EPI_ISL_9124281, EPI_ISL_9124310, EPI_ISL_9124356, EPI_ISL_9124376, EPI_ISL_9124399, EPI_ISL_9124416, EPI_ISL_9124444, EPI_ISL_9124455, EPI_ISL_9124457, EPI_ISL_9124475, EPI_ISL_9124479, EPI_ISL_9124480, EPI_ISL_9124497, EPI_ISL_9124503, EPI_ISL_9124505, EPI_ISL_9124514, EPI_ISL_9124544, EPI_ISL_9124560, EPI_ISL_9124564, EPI_ISL_9124627, EPI_ISL_9124630, EPI_ISL_9124633, EPI_ISL_9124666, EPI_ISL_9124743, EPI_ISL_9124765, EPI_ISL_9124808, EPI_ISL_9124814, EPI_ISL_9124823, EPI_ISL_9124912, EPI_ISL_9124945, EPI_ISL_9124957, EPI_ISL_9124959, EPI_ISL_9124979, EPI_ISL_9124994, EPI_ISL_9124997, EPI_ISL_9125293, EPI_ISL_9125306, EPI_ISL_9125318, EPI_ISL_9125326, EPI_ISL_9125328, EPI_ISL_9125341, EPI_ISL_9125346, EPI_ISL_9125364, EPI_ISL_9125375, EPI_ISL_9125401, EPI_ISL_9125419, EPI_ISL_9125430, EPI_ISL_9125441, EPI_ISL_9125499, EPI_ISL_9125519, EPI_ISL_9125555, EPI_ISL_9125929, EPI_ISL_9125947, EPI_ISL_9126011, EPI_ISL_9126036, EPI_ISL_9126039, EPI_ISL_9126045, EPI_ISL_9126051, EPI_ISL_9126066, EPI_ISL_9126106, EPI_ISL_9126114, EPI_ISL_9126129, EPI_ISL_9126184, EPI_ISL_9126213, EPI_ISL_9126249, EPI_ISL_9126263, EPI_ISL_9126269, EPI_ISL_9126281, EPI_ISL_9126291, EPI_ISL_9126303, EPI_ISL_9126307, EPI_ISL_9126316, EPI_ISL_9126501, EPI_ISL_9126831, EPI_ISL_9126848, EPI_ISL_9126865, EPI_ISL_9126916, EPI_ISL_9126961, EPI_ISL_9126984, EPI_ISL_9126999, EPI_ISL_9127019, EPI_ISL_9127262, EPI_ISL_9127291, EPI_ISL_9127465, EPI_ISL_9127475, EPI_ISL_9127483, EPI_ISL_9127514, EPI_ISL_9127516, EPI_ISL_9127520, EPI_ISL_9127526, EPI_ISL_9127562, EPI_ISL_9127598, EPI_ISL_9127627, EPI_ISL_9127644, EPI_ISL_9127660, EPI_ISL_9127681, EPI_ISL_9127697, EPI_ISL_9127709, EPI_ISL_9127842, EPI_ISL_9127851, EPI_ISL_9127855, EPI_ISL_9127863, EPI_ISL_9127866, EPI_ISL_9127887, EPI_ISL_9127893, EPI_ISL_9127947, EPI_ISL_9127971, EPI_ISL_9127982, EPI_ISL_9127997, EPI_ISL_9128045, EPI_ISL_9128136, EPI_ISL_9128157, EPI_ISL_9128210, EPI_ISL_9128216, EPI_ISL_9128235, EPI_ISL_9128342, EPI_ISL_9128400, EPI_ISL_9128437, EPI_ISL_9128444, EPI_ISL_9128525, EPI_ISL_9128531, EPI_ISL_9128572, EPI_ISL_9128677, EPI_ISL_9128681, EPI_ISL_9128688, EPI_ISL_9128695, EPI_ISL_9128701, EPI_ISL_9128718, EPI_ISL_9128721, EPI_ISL_9128725, EPI_ISL_9128774, EPI_ISL_9128788, EPI_ISL_9128811, EPI_ISL_9128820, EPI_ISL_9128825, EPI_ISL_9128828, EPI_ISL_9128837, EPI_ISL_9128840, EPI_ISL_9128849, EPI_ISL_9128852, EPI_ISL_9128908, EPI_ISL_9128914, EPI_ISL_9128950, EPI_ISL_9128960, EPI_ISL_9128993, EPI_ISL_9129006, EPI_ISL_9129009, EPI_ISL_9129011, EPI_ISL_9129025, EPI_ISL_9129042, EPI_ISL_9129078, EPI_ISL_9129085, EPI_ISL_9129131, EPI_ISL_9131404, EPI_ISL_9131441, EPI_ISL_9131445, EPI_ISL_9131449, EPI_ISL_9131612, EPI_ISL_9131658, EPI_ISL_9131680, EPI_ISL_9131696, EPI_ISL_9131736, EPI_ISL_9131746, EPI_ISL_9131770, EPI_ISL_9131779, EPI_ISL_9131784, EPI_ISL_9134224, EPI_ISL_9134246, EPI_ISL_9134251, EPI_ISL_9134271, EPI_ISL_9134288, EPI_ISL_9134295, EPI_ISL_9134309, EPI_ISL_9134313, EPI_ISL_9134642 | | | |
| see above | Department of Bacteria, Parasites and Fungi, Statens Serum Institut, Copenhagen, Denmark | Statens Serum Institut Bioinformatics and Microbial Genomics | Danish Covid-19 Genome Consortium |

We gratefully acknowledge the following Authors from the Originating laboratories responsible for obtaining the specimens, as well as the Submitting laboratories where the genome data were generated and shared via GISAID, on which this research is based.

All Submitters of data may be contacted directly via www.gisaid.org

Authors are sorted alphabetically.

Acknowledgement EPI_SET Identifier: EPI_SET_20220527no

| Accession ID | Originating Laboratory | Submitting Laboratory | Authors |
|---|---|---|---|
| EPI_ISL_9119563, EPI_ISL_9119618, EPI_ISL_9119689, EPI_ISL_9119693, EPI_ISL_9119701, EPI_ISL_9119712, EPI_ISL_9119720, EPI_ISL_9119732, EPI_ISL_9119756, EPI_ISL_9119764, EPI_ISL_9119779, EPI_ISL_9119817, EPI_ISL_9119829, EPI_ISL_9119847, EPI_ISL_9119855, EPI_ISL_9119883, EPI_ISL_9119894, EPI_ISL_9119898, EPI_ISL_9119905, EPI_ISL_9119929, EPI_ISL_9119968, EPI_ISL_9120015, EPI_ISL_9120048, EPI_ISL_9120060, EPI_ISL_9120073, EPI_ISL_9120085, EPI_ISL_9120090, EPI_ISL_9120096, EPI_ISL_9120128, EPI_ISL_9120172, EPI_ISL_9120174, EPI_ISL_9120215, EPI_ISL_9120224, EPI_ISL_9120233, EPI_ISL_9120245, EPI_ISL_9120258, EPI_ISL_9120364, EPI_ISL_9120472, EPI_ISL_9120476, EPI_ISL_9120479, EPI_ISL_9120494, EPI_ISL_9120511, EPI_ISL_9120516, EPI_ISL_9120530, EPI_ISL_9120557, EPI_ISL_9120574, EPI_ISL_9120581, EPI_ISL_9120609, EPI_ISL_9120614, EPI_ISL_9120620, EPI_ISL_9120624, EPI_ISL_9120626, EPI_ISL_9120632, EPI_ISL_9120639, EPI_ISL_9121029, EPI_ISL_9121030, EPI_ISL_9121040, EPI_ISL_9121042, EPI_ISL_9121043, EPI_ISL_9121068, EPI_ISL_9121100, EPI_ISL_9121102, EPI_ISL_9121134, EPI_ISL_9121173, EPI_ISL_9121192, EPI_ISL_9121193, EPI_ISL_9121204, EPI_ISL_9121214, EPI_ISL_9121221, EPI_ISL_9121222, EPI_ISL_9121226, EPI_ISL_9121261, EPI_ISL_9121269, EPI_ISL_9121290, EPI_ISL_9121299, EPI_ISL_9121300, EPI_ISL_9121303, EPI_ISL_9121304, EPI_ISL_9121308, EPI_ISL_9121331, EPI_ISL_9121382, EPI_ISL_9121399, EPI_ISL_9121406, EPI_ISL_9121424, EPI_ISL_9121471, EPI_ISL_9121475, EPI_ISL_9121496, EPI_ISL_9121517, EPI_ISL_9121527, EPI_ISL_9121546, EPI_ISL_9121549, EPI_ISL_9121551, EPI_ISL_9121557, EPI_ISL_9121576, EPI_ISL_9121606, EPI_ISL_9121611, EPI_ISL_9121631, EPI_ISL_9121635, EPI_ISL_9121637, EPI_ISL_9121643, EPI_ISL_9121704, EPI_ISL_9121736, EPI_ISL_9121763, EPI_ISL_9121787, EPI_ISL_9121797, EPI_ISL_9121844, EPI_ISL_9121848, EPI_ISL_9121853, EPI_ISL_9121899, EPI_ISL_9121902, EPI_ISL_9122023, EPI_ISL_9122047, EPI_ISL_9122054, EPI_ISL_9122057, EPI_ISL_9122064, EPI_ISL_9122068, EPI_ISL_9122075, EPI_ISL_9122145, EPI_ISL_9122146, EPI_ISL_9122147, EPI_ISL_9122174, EPI_ISL_9122918, EPI_ISL_9122925, EPI_ISL_9122939, EPI_ISL_9123119, EPI_ISL_9123127, EPI_ISL_9123141, EPI_ISL_9123147, EPI_ISL_9123154, EPI_ISL_9123199, EPI_ISL_9123224, EPI_ISL_9123250, EPI_ISL_9123274, EPI_ISL_9123288, EPI_ISL_9123297, EPI_ISL_9123311, EPI_ISL_9123329, EPI_ISL_9123361, EPI_ISL_9123435, EPI_ISL_9123446, EPI_ISL_9123467, EPI_ISL_9123484, EPI_ISL_9123485, EPI_ISL_9123486, EPI_ISL_9123516, EPI_ISL_9123538, EPI_ISL_9123540, EPI_ISL_9123544, EPI_ISL_9123548, EPI_ISL_9123550, EPI_ISL_9123551, EPI_ISL_9123764, EPI_ISL_9123776, EPI_ISL_9123790, EPI_ISL_9124010, EPI_ISL_9124025, EPI_ISL_9124039, EPI_ISL_9124040, EPI_ISL_9124143, EPI_ISL_9124162, EPI_ISL_9124167, EPI_ISL_9124182, EPI_ISL_9124201, EPI_ISL_9124223, EPI_ISL_9124228, EPI_ISL_9124243, EPI_ISL_9124258, EPI_ISL_9124302, EPI_ISL_9124373, EPI_ISL_9124494, EPI_ISL_9124517, EPI_ISL_9124587, EPI_ISL_9124610, EPI_ISL_9124802, EPI_ISL_9124835, EPI_ISL_9124896, EPI_ISL_9124942, EPI_ISL_9124956, EPI_ISL_9125016, EPI_ISL_9125060, EPI_ISL_9125289, EPI_ISL_9125304, EPI_ISL_9125361, EPI_ISL_9125372, EPI_ISL_9125394, EPI_ISL_9125445, EPI_ISL_9125531, EPI_ISL_9125919, EPI_ISL_9125988, EPI_ISL_9126000, EPI_ISL_9126083, EPI_ISL_9126229, EPI_ISL_9126845, EPI_ISL_9126858, EPI_ISL_9126873, EPI_ISL_9126925, EPI_ISL_9126966, EPI_ISL_9126978, EPI_ISL_9126994, EPI_ISL_9127050, EPI_ISL_9127054, EPI_ISL_9127077, EPI_ISL_9127273, EPI_ISL_9127336, EPI_ISL_9127340, EPI_ISL_9127373, EPI_ISL_9127409, EPI_ISL_9127455, EPI_ISL_9127522, EPI_ISL_9127559, EPI_ISL_9127610, EPI_ISL_9127657, EPI_ISL_9127668, EPI_ISL_9127671, EPI_ISL_9127736, EPI_ISL_9127739, EPI_ISL_9127753, EPI_ISL_9127848, EPI_ISL_9127873, EPI_ISL_9127910, EPI_ISL_9127955, EPI_ISL_9128010, EPI_ISL_9128060, EPI_ISL_9128177, EPI_ISL_9128194, EPI_ISL_9128365, EPI_ISL_9128377, EPI_ISL_9128456, EPI_ISL_9128474, EPI_ISL_9128480, EPI_ISL_9128483, EPI_ISL_9128501, EPI_ISL_9128665, EPI_ISL_9128685, EPI_ISL_9128739, EPI_ISL_9128763, EPI_ISL_9128872, EPI_ISL_9128917, EPI_ISL_9129056, EPI_ISL_9129066, EPI_ISL_9131419, EPI_ISL_9131434, EPI_ISL_9131443, EPI_ISL_9131534, EPI_ISL_9131651, EPI_ISL_9131740, EPI_ISL_9131742, EPI_ISL_9134221, EPI_ISL_9134267 | | | |
| see above | Department of Bacteria, Parasites and Fungi, Statens Serum Institut, Copenhagen, Denmark | Statens Serum Institut Bioinformatics and Microbial Genomics | Danish Covid-19 Genome Consortium |

We gratefully acknowledge the following Authors from the Originating laboratories responsible for obtaining the specimens, as well as the Submitting laboratories where the genome data were generated and shared via GISAID, on which this research is based.

All Submitters of data may be contacted directly via www.gisaid.org

Authors are sorted alphabetically.

| Accession ID | Originating Laboratory | Submitting Laboratory | Authors |
|---|---|---|---|
| EPI_ISL_9119550, EPI_ISL_9119599, EPI_ISL_9119665, EPI_ISL_9119667, EPI_ISL_9119690, EPI_ISL_9119725, EPI_ISL_9119746, EPI_ISL_9119762, EPI_ISL_9119766, EPI_ISL_9119780, EPI_ISL_9119797, EPI_ISL_9119818, EPI_ISL_9119848, EPI_ISL_9119878, EPI_ISL_9119881, EPI_ISL_9119893, EPI_ISL_9119907, EPI_ISL_9119952, EPI_ISL_9119962, EPI_ISL_9119963, EPI_ISL_9119981, EPI_ISL_9119992, EPI_ISL_9119998, EPI_ISL_9120013, EPI_ISL_9120020, EPI_ISL_9120024, EPI_ISL_9120025, EPI_ISL_9120034, EPI_ISL_9120069, EPI_ISL_9120070, EPI_ISL_9120082, EPI_ISL_9120092, EPI_ISL_9120121, EPI_ISL_9120133, EPI_ISL_9120158, EPI_ISL_9120168, EPI_ISL_9120183, EPI_ISL_9120186, EPI_ISL_9120193, EPI_ISL_9120218, EPI_ISL_9120221, EPI_ISL_9120225, EPI_ISL_9120252, EPI_ISL_9120468, EPI_ISL_9120492, EPI_ISL_9120526, EPI_ISL_9120552, EPI_ISL_9120556, EPI_ISL_9120641, EPI_ISL_9121023, EPI_ISL_9121036, EPI_ISL_9121057, EPI_ISL_9121095, EPI_ISL_9121099, EPI_ISL_9121115, EPI_ISL_9121194, EPI_ISL_9121201, EPI_ISL_9121237, EPI_ISL_9121262, EPI_ISL_9121289, EPI_ISL_9121294, EPI_ISL_9121328, EPI_ISL_9121361, EPI_ISL_9121371, EPI_ISL_9121392, EPI_ISL_9121420, EPI_ISL_9121422, EPI_ISL_9121432, EPI_ISL_9121466, EPI_ISL_9121505, EPI_ISL_9121535, EPI_ISL_9121536, EPI_ISL_9121543, EPI_ISL_9121554, EPI_ISL_9121566, EPI_ISL_9121567, EPI_ISL_9121572, EPI_ISL_9121580, EPI_ISL_9121723, EPI_ISL_9121741, EPI_ISL_9121779, EPI_ISL_9121862, EPI_ISL_9121868, EPI_ISL_9122020, EPI_ISL_9122035, EPI_ISL_9122041, EPI_ISL_9122042, EPI_ISL_9122048, EPI_ISL_9122049, EPI_ISL_9122056, EPI_ISL_9122074, EPI_ISL_9122083, EPI_ISL_9122101, EPI_ISL_9122118, EPI_ISL_9122129, EPI_ISL_9122133, EPI_ISL_9122143, EPI_ISL_9122151, EPI_ISL_9122920, EPI_ISL_9122923, EPI_ISL_9122926, EPI_ISL_9122954, EPI_ISL_9122959, EPI_ISL_9122976, EPI_ISL_9123047, EPI_ISL_9123122, EPI_ISL_9123129, EPI_ISL_9123160, EPI_ISL_9123170, EPI_ISL_9123226, EPI_ISL_9123230, EPI_ISL_9123235, EPI_ISL_9123249, EPI_ISL_9123291, EPI_ISL_9123306, EPI_ISL_9123314, EPI_ISL_9123323, EPI_ISL_9123335, EPI_ISL_9123355, EPI_ISL_9123403, EPI_ISL_9123408, EPI_ISL_9123417, EPI_ISL_9123432, EPI_ISL_9123473, EPI_ISL_9123495, EPI_ISL_9123498, EPI_ISL_9123521, EPI_ISL_9123789, EPI_ISL_9123798, EPI_ISL_9124104, EPI_ISL_9124134, EPI_ISL_9124187, EPI_ISL_9124237, EPI_ISL_9124255, EPI_ISL_9124279, EPI_ISL_9124441, EPI_ISL_9124488, EPI_ISL_9124687, EPI_ISL_9124774, EPI_ISL_9124884, EPI_ISL_9124951, EPI_ISL_9125343, EPI_ISL_9125547, EPI_ISL_9126072, EPI_ISL_9126075, EPI_ISL_9126140, EPI_ISL_9126142, EPI_ISL_9126219, EPI_ISL_9126296, EPI_ISL_9126505, EPI_ISL_9126829, EPI_ISL_9126888, EPI_ISL_9126892, EPI_ISL_9127008, EPI_ISL_9127083, EPI_ISL_9127276, EPI_ISL_9127297, EPI_ISL_9127302, EPI_ISL_9127332, EPI_ISL_9127407, EPI_ISL_9127412, EPI_ISL_9127414, EPI_ISL_9127418, EPI_ISL_9127444, EPI_ISL_9127543, EPI_ISL_9127564, EPI_ISL_9127570, EPI_ISL_9127573, EPI_ISL_9127621, EPI_ISL_9127646, EPI_ISL_9127679, EPI_ISL_9127700, EPI_ISL_9127723, EPI_ISL_9127823, EPI_ISL_9127862, EPI_ISL_9127869, EPI_ISL_9127899, EPI_ISL_9127913, EPI_ISL_9127925, EPI_ISL_9128000, EPI_ISL_9128007, EPI_ISL_9128057, EPI_ISL_9128113, EPI_ISL_9128124, EPI_ISL_9128269, EPI_ISL_9128368, EPI_ISL_9128462, EPI_ISL_9128479, EPI_ISL_9128540, EPI_ISL_9128612, EPI_ISL_9128615, EPI_ISL_9128666, EPI_ISL_9128704, EPI_ISL_9128730, EPI_ISL_9128748, EPI_ISL_9128911, EPI_ISL_9128936, EPI_ISL_9128957, EPI_ISL_9128984, EPI_ISL_9128996, EPI_ISL_9129014, EPI_ISL_9131414, EPI_ISL_9131444, EPI_ISL_9131491, EPI_ISL_9131545, EPI_ISL_9131580, EPI_ISL_9131625, EPI_ISL_9131676, EPI_ISL_9131733 | | | |
| see above | Department of Bacteria, Parasites and Fungi, Statens Serum Institut, Copenhagen, Denmark | Statens Serum Institut Bioinformatics and Microbial Genomics | Danish Covid-19 Genome Consortium |

We gratefully acknowledge the following Authors from the Originating laboratories responsible for obtaining the specimens, as well as the Submitting laboratories where the genome data were generated and shared via GISAID, on which this research is based.

All Submitters of data may be contacted directly via www.gisaid.org

Authors are sorted alphabetically.

| Accession ID | Originating Laboratory | Submitting Laboratory | Authors |
| --- | --- | --- | --- |
| EPI_ISL_9119565, EPI_ISL_9119596, EPI_ISL_9119636, EPI_ISL_9119641, EPI_ISL_9119652, EPI_ISL_9119683, EPI_ISL_9119685, EPI_ISL_9119700, EPI_ISL_9119713, EPI_ISL_9119742, EPI_ISL_9119749, EPI_ISL_9119751, EPI_ISL_9119753, EPI_ISL_9119754, EPI_ISL_9119785, EPI_ISL_9119799, EPI_ISL_9119800, EPI_ISL_9119805, EPI_ISL_9119807, EPI_ISL_9119816, EPI_ISL_9119819, EPI_ISL_9119820, EPI_ISL_9119825, EPI_ISL_9119832, EPI_ISL_9119850, EPI_ISL_9119856, EPI_ISL_9119862, EPI_ISL_9119872, EPI_ISL_9119877, EPI_ISL_9119906, EPI_ISL_9119910, EPI_ISL_9119933, EPI_ISL_9119936, EPI_ISL_9119939, EPI_ISL_9119942, EPI_ISL_9119953, EPI_ISL_9119957, EPI_ISL_9119961, EPI_ISL_9119966, EPI_ISL_9119972, EPI_ISL_9119976, EPI_ISL_9119977, EPI_ISL_9119978, EPI_ISL_9119986, EPI_ISL_9119999, EPI_ISL_9120003, EPI_ISL_9120023, EPI_ISL_9120027, EPI_ISL_9120028, EPI_ISL_9120031, EPI_ISL_9120049, EPI_ISL_9120052, EPI_ISL_9120053, EPI_ISL_9120071, EPI_ISL_9120072, EPI_ISL_9120075, EPI_ISL_9120077, EPI_ISL_9120110, EPI_ISL_9120119, EPI_ISL_9120122, EPI_ISL_9120125, EPI_ISL_9120131, EPI_ISL_9120171, EPI_ISL_9120175, EPI_ISL_9120177, EPI_ISL_9120185, EPI_ISL_9120200, EPI_ISL_9120204, EPI_ISL_9120207, EPI_ISL_9120209, EPI_ISL_9120214, EPI_ISL_9120216, EPI_ISL_9120219, EPI_ISL_9120226, EPI_ISL_9120228, EPI_ISL_9120231, EPI_ISL_9120246, EPI_ISL_9120254, EPI_ISL_9120262, EPI_ISL_9120480, EPI_ISL_9120493, EPI_ISL_9120497, EPI_ISL_9120503, EPI_ISL_9120512, EPI_ISL_9120514, EPI_ISL_9120518, EPI_ISL_9120519, EPI_ISL_9120529, EPI_ISL_9120537, EPI_ISL_9120540, EPI_ISL_9120544, EPI_ISL_9120545, EPI_ISL_9120547, EPI_ISL_9120570, EPI_ISL_9120578, EPI_ISL_9120591, EPI_ISL_9120592, EPI_ISL_9120594, EPI_ISL_9120596, EPI_ISL_9120597, EPI_ISL_9120598, EPI_ISL_9120602, EPI_ISL_9120610, EPI_ISL_9120628, EPI_ISL_9120637, EPI_ISL_9120902, EPI_ISL_9121028, EPI_ISL_9121039, EPI_ISL_9121052, EPI_ISL_9121056, EPI_ISL_9121058, EPI_ISL_9121080, EPI_ISL_9121082, EPI_ISL_9121084, EPI_ISL_9121104, EPI_ISL_9121117, EPI_ISL_9121118, EPI_ISL_9121119, EPI_ISL_9121133, EPI_ISL_9121141, EPI_ISL_9121178, EPI_ISL_9121179, EPI_ISL_9121195, EPI_ISL_9121202, EPI_ISL_9121203, EPI_ISL_9121209, EPI_ISL_9121213, EPI_ISL_9121218, EPI_ISL_9121219, EPI_ISL_9121227, EPI_ISL_9121251, EPI_ISL_9121255, EPI_ISL_9121258, EPI_ISL_9121267, EPI_ISL_9121271, EPI_ISL_9121280, EPI_ISL_9121282, EPI_ISL_9121296, EPI_ISL_9121307, EPI_ISL_9121322, EPI_ISL_9121327, EPI_ISL_9121332, EPI_ISL_9121342, EPI_ISL_9121360, EPI_ISL_9121366, EPI_ISL_9121373, EPI_ISL_9121388, EPI_ISL_9121412, EPI_ISL_9121417, EPI_ISL_9121425, EPI_ISL_9121430, EPI_ISL_9121447, EPI_ISL_9121457, EPI_ISL_9121464, EPI_ISL_9121467, EPI_ISL_9121468, EPI_ISL_9121476, EPI_ISL_9121486, EPI_ISL_9121491, EPI_ISL_9121504, EPI_ISL_9121507, EPI_ISL_9121513, EPI_ISL_9121523, EPI_ISL_9121525, EPI_ISL_9121552, EPI_ISL_9121558, EPI_ISL_9121581, EPI_ISL_9121591, EPI_ISL_9121592, EPI_ISL_9121602, EPI_ISL_9121604, EPI_ISL_9121610, EPI_ISL_9121612, EPI_ISL_9121616, EPI_ISL_9121627, EPI_ISL_9121630, EPI_ISL_9121634, EPI_ISL_9121639, EPI_ISL_9121647, EPI_ISL_9121648, EPI_ISL_9121651, EPI_ISL_9121695, EPI_ISL_9121710, EPI_ISL_9121724, EPI_ISL_9121742, EPI_ISL_9121746, EPI_ISL_9121764, EPI_ISL_9121765, EPI_ISL_9121773, EPI_ISL_9121782, EPI_ISL_9121794, EPI_ISL_9121795, EPI_ISL_9121845, EPI_ISL_9121857, EPI_ISL_9121860, EPI_ISL_9121864, EPI_ISL_9121865, EPI_ISL_9121878, EPI_ISL_9121885, EPI_ISL_9121886, EPI_ISL_9121892, EPI_ISL_9122014, EPI_ISL_9122017, EPI_ISL_9122036, EPI_ISL_9122038, EPI_ISL_9122067, EPI_ISL_9122070, EPI_ISL_9122071, EPI_ISL_9122079, EPI_ISL_9122091, EPI_ISL_9122095, EPI_ISL_9122100, EPI_ISL_9122120, EPI_ISL_9122124, EPI_ISL_9122136, EPI_ISL_9122137, EPI_ISL_9122152, EPI_ISL_9122163, EPI_ISL_9122166, EPI_ISL_9122921, EPI_ISL_9122931, EPI_ISL_9122948, EPI_ISL_9122953, EPI_ISL_9122966, EPI_ISL_9122967, EPI_ISL_9122989, EPI_ISL_9122991, EPI_ISL_9123001, EPI_ISL_9123117, EPI_ISL_9123120, EPI_ISL_9123130, EPI_ISL_9123131, EPI_ISL_9123132, EPI_ISL_9123133, EPI_ISL_9123151, EPI_ISL_9123153, EPI_ISL_9123162, EPI_ISL_9123180, EPI_ISL_9123186, EPI_ISL_9123214, EPI_ISL_9123217, EPI_ISL_9123219, EPI_ISL_9123222, EPI_ISL_9123231, EPI_ISL_9123239, EPI_ISL_9123240, EPI_ISL_9123259, EPI_ISL_9123261, EPI_ISL_9123262, EPI_ISL_9123269, EPI_ISL_9123281, EPI_ISL_9123298, EPI_ISL_9123300, EPI_ISL_9123304, EPI_ISL_9123307, EPI_ISL_9123316, EPI_ISL_9123322, EPI_ISL_9123326, EPI_ISL_9123328, EPI_ISL_9123337, EPI_ISL_9123345, EPI_ISL_9123346, EPI_ISL_9123350, EPI_ISL_9123357, EPI_ISL_9123358, EPI_ISL_9123371, EPI_ISL_9123394, EPI_ISL_9123397, EPI_ISL_9123400, EPI_ISL_9123401, EPI_ISL_9123405, EPI_ISL_9123434, EPI_ISL_9123460, EPI_ISL_9123471, EPI_ISL_9123472, EPI_ISL_9123475, EPI_ISL_9123477, EPI_ISL_9123481, EPI_ISL_9123487, EPI_ISL_9123489, EPI_ISL_9123496, EPI_ISL_9123500, EPI_ISL_9123504, EPI_ISL_9123505, EPI_ISL_9123507, EPI_ISL_9123518, EPI_ISL_9123525, EPI_ISL_9123528, EPI_ISL_9123530, EPI_ISL_9123556, EPI_ISL_9123558, EPI_ISL_9123561, EPI_ISL_9123562, EPI_ISL_9123754, EPI_ISL_9123768, EPI_ISL_9123771, EPI_ISL_9123793, EPI_ISL_9123997, EPI_ISL_9124007, EPI_ISL_9124017, EPI_ISL_9124019, EPI_ISL_9124023, EPI_ISL_9124035, EPI_ISL_9124047, EPI_ISL_9124052, EPI_ISL_9124058, EPI_ISL_9124114, EPI_ISL_9124140, EPI_ISL_9124145, EPI_ISL_9124156, EPI_ISL_9124161, EPI_ISL_9124190, EPI_ISL_9124215, EPI_ISL_9124217, EPI_ISL_9124220, EPI_ISL_9124240, EPI_ISL_9124284, EPI_ISL_9124296, EPI_ISL_9124308, EPI_ISL_9124323, EPI_ISL_9124344, EPI_ISL_9124370, EPI_ISL_9124379, EPI_ISL_9124382, EPI_ISL_9124405, EPI_ISL_9124418, EPI_ISL_9124421, EPI_ISL_9124424, EPI_ISL_9124432, EPI_ISL_9124435, EPI_ISL_9124482, EPI_ISL_9124546, EPI_ISL_9124561, EPI_ISL_9124562, EPI_ISL_9124581, EPI_ISL_9124624, EPI_ISL_9124647, EPI_ISL_9124663, EPI_ISL_9124668, EPI_ISL_9124783, EPI_ISL_9124829, EPI_ISL_9124832, EPI_ISL_9124849, EPI_ISL_9124859, EPI_ISL_9124907, EPI_ISL_9124923, EPI_ISL_9124960, EPI_ISL_9124986, EPI_ISL_9125000, EPI_ISL_9125338, EPI_ISL_9125357, EPI_ISL_9125366, EPI_ISL_9125422, EPI_ISL_9125438, EPI_ISL_9125443, EPI_ISL_9125444, EPI_ISL_9125446, EPI_ISL_9125452, EPI_ISL_9125466, EPI_ISL_9125475, EPI_ISL_9125478, EPI_ISL_9125489, EPI_ISL_9125496, EPI_ISL_9125515, EPI_ISL_9125522, EPI_ISL_9125529, EPI_ISL_9125905, EPI_ISL_9125924, EPI_ISL_9125953, EPI_ISL_9125956, EPI_ISL_9125962, EPI_ISL_9125997, EPI_ISL_9126019, EPI_ISL_9126033, EPI_ISL_9126174, EPI_ISL_9126200, EPI_ISL_9126222, EPI_ISL_9126250, EPI_ISL_9126256, EPI_ISL_9126272, EPI_ISL_9126319, EPI_ISL_9126895, EPI_ISL_9126911, EPI_ISL_9126940, EPI_ISL_9126958, EPI_ISL_9126981, EPI_ISL_9127087, EPI_ISL_9127268, EPI_ISL_9127334, EPI_ISL_9127389, EPI_ISL_9127430, EPI_ISL_9127450, EPI_ISL_9127525, EPI_ISL_9127528, EPI_ISL_9127532, EPI_ISL_9127537, EPI_ISL_9127556, EPI_ISL_9127582, EPI_ISL_9127585, EPI_ISL_9127599, EPI_ISL_9127616, EPI_ISL_9127632, EPI_ISL_9127692, EPI_ISL_9127695, EPI_ISL_9127703, EPI_ISL_9127720, EPI_ISL_9127759, EPI_ISL_9127784, EPI_ISL_9127818, EPI_ISL_9127826, EPI_ISL_9127846, EPI_ISL_9127854, EPI_ISL_9127858, EPI_ISL_9127859, EPI_ISL_9127860, EPI_ISL_9127870, EPI_ISL_9127883, EPI_ISL_9127896, EPI_ISL_9127916, EPI_ISL_9127927, EPI_ISL_9127950, EPI_ISL_9127994, EPI_ISL_9128021, EPI_ISL_9128047, EPI_ISL_9128048, EPI_ISL_9128062, EPI_ISL_9128108, EPI_ISL_9128121, EPI_ISL_9128128, EPI_ISL_9128146, EPI_ISL_9128174, EPI_ISL_9128238, EPI_ISL_9128290, EPI_ISL_9128293, EPI_ISL_9128316, EPI_ISL_9128397, EPI_ISL_9128469, EPI_ISL_9128495, EPI_ISL_9128514, EPI_ISL_9128517, EPI_ISL_9128520, EPI_ISL_9128529, EPI_ISL_9128543, EPI_ISL_9128547, EPI_ISL_9128550, EPI_ISL_9128554, EPI_ISL_9128560, EPI_ISL_9128566, EPI_ISL_9128581, EPI_ISL_9128633, EPI_ISL_9128637, EPI_ISL_9128642, EPI_ISL_9128645, EPI_ISL_9128654, EPI_ISL_9128672, EPI_ISL_9128676, EPI_ISL_9128683, EPI_ISL_9128684, EPI_ISL_9128742, EPI_ISL_9128757, EPI_ISL_9128776, EPI_ISL_9128780, EPI_ISL_9128791, EPI_ISL_9128798, EPI_ISL_9128808, EPI_ISL_9128866, EPI_ISL_9128875, EPI_ISL_9128942, EPI_ISL_9129017, EPI_ISL_9129030, EPI_ISL_9129040, EPI_ISL_9129082, EPI_ISL_9129114, EPI_ISL_9131411, EPI_ISL_9131436, EPI_ISL_9131517, EPI_ISL_9131524, EPI_ISL_9131566, EPI_ISL_9131573, EPI_ISL_9131589, EPI_ISL_9131593, EPI_ISL_9131596, EPI_ISL_9131606, EPI_ISL_9131666, EPI_ISL_9131711, EPI_ISL_9131720, EPI_ISL_9131759, EPI_ISL_9131773, EPI_ISL_9131776, EPI_ISL_9131781, EPI_ISL_9131809, EPI_ISL_9134208, EPI_ISL_9134212, EPI_ISL_9134228, EPI_ISL_9134243, EPI_ISL_9134250, EPI_ISL_9134265, EPI_ISL_9134320, EPI_ISL_9134618, EPI_ISL_9134663 | | | |
| see above | Department of Bacteria, Parasites and Fungi, Statens Serum Institut, Copenhagen, Denmark | Statens Serum Institut Bioinformatics and Microbial Genomics | Danish Covid-19 Genome Consortium |

We gratefully acknowledge the following Authors from the Originating laboratories responsible for obtaining the specimens, as well as the Submitting laboratories where the genome data were generated and shared via GISAID, on which this research is based.

All Submitters of data may be contacted directly via www.gisaid.org

Authors are sorted alphabetically.

Acknowledgement EPI_SET Identifier: EPI_SET_20220527yx

<table>
<tr><th>Accession ID</th><th>Originating Laboratory</th><th>Submitting Laboratory</th><th>Authors</th></tr>
<tr><td colspan="4">EPI_ISL_9119558, EPI_ISL_9119568, EPI_ISL_9119573, EPI_ISL_9119578, EPI_ISL_9119581, EPI_ISL_9119593, EPI_ISL_9119627, EPI_ISL_9119639, EPI_ISL_9119647, EPI_ISL_9119649, EPI_ISL_9119656, EPI_ISL_9119658, EPI_ISL_9119659, EPI_ISL_9119660, EPI_ISL_9119675, EPI_ISL_9119682, EPI_ISL_9119686, EPI_ISL_9119687, EPI_ISL_9119698, EPI_ISL_9119703, EPI_ISL_9119704, EPI_ISL_9119714, EPI_ISL_9119717, EPI_ISL_9119719, EPI_ISL_9119737, EPI_ISL_9119745, EPI_ISL_9119747, EPI_ISL_9119748, EPI_ISL_9119765, EPI_ISL_9119771, EPI_ISL_9119794, EPI_ISL_9119821, EPI_ISL_9119823, EPI_ISL_9119826, EPI_ISL_9119831, EPI_ISL_9119836, EPI_ISL_9119840, EPI_ISL_9119858, EPI_ISL_9119865, EPI_ISL_9119874, EPI_ISL_9119886, EPI_ISL_9119896, EPI_ISL_9119900, EPI_ISL_9119903, EPI_ISL_9119908, EPI_ISL_9119909, EPI_ISL_9119916, EPI_ISL_9119924, EPI_ISL_9119925, EPI_ISL_9119932, EPI_ISL_9119948, EPI_ISL_9119956, EPI_ISL_9119959, EPI_ISL_9119970, EPI_ISL_9119971, EPI_ISL_9119975, EPI_ISL_9119979, EPI_ISL_9119983, EPI_ISL_9119991, EPI_ISL_9119996, EPI_ISL_9120002, EPI_ISL_9120010, EPI_ISL_9120012, EPI_ISL_9120019, EPI_ISL_9120021, EPI_ISL_9120022, EPI_ISL_9120043, EPI_ISL_9120044, EPI_ISL_9120050, EPI_ISL_9120051, EPI_ISL_9120061, EPI_ISL_9120088, EPI_ISL_9120099, EPI_ISL_9120101, EPI_ISL_9120107, EPI_ISL_9120108, EPI_ISL_9120112, EPI_ISL_9120116, EPI_ISL_9120120, EPI_ISL_9120127, EPI_ISL_9120140, EPI_ISL_9120141, EPI_ISL_9120147, EPI_ISL_9120153, EPI_ISL_9120155, EPI_ISL_9120164, EPI_ISL_9120165, EPI_ISL_9120166, EPI_ISL_9120169, EPI_ISL_9120176, EPI_ISL_9120179, EPI_ISL_9120184, EPI_ISL_9120190, EPI_ISL_9120191, EPI_ISL_9120198, EPI_ISL_9120201, EPI_ISL_9120238, EPI_ISL_9120241, EPI_ISL_9120255, EPI_ISL_9120256, EPI_ISL_9120267, EPI_ISL_9120268, EPI_ISL_9120473, EPI_ISL_9120475, EPI_ISL_9120481, EPI_ISL_9120483, EPI_ISL_9120490, EPI_ISL_9120496, EPI_ISL_9120498, EPI_ISL_9120499, EPI_ISL_9120500, EPI_ISL_9120506, EPI_ISL_9120510, EPI_ISL_9120517, EPI_ISL_9120520, EPI_ISL_9120523, EPI_ISL_9120528, EPI_ISL_9120532, EPI_ISL_9120539, EPI_ISL_9120542, EPI_ISL_9120550, EPI_ISL_9120551, EPI_ISL_9120559, EPI_ISL_9120560, EPI_ISL_9120562, EPI_ISL_9120563, EPI_ISL_9120568, EPI_ISL_9120573, EPI_ISL_9120585, EPI_ISL_9120595, EPI_ISL_9120604, EPI_ISL_9120605, EPI_ISL_9120612, EPI_ISL_9120613, EPI_ISL_9120615, EPI_ISL_9120621, EPI_ISL_9120633, EPI_ISL_9120645, EPI_ISL_9120648, EPI_ISL_9121037, EPI_ISL_9121041, EPI_ISL_9121047, EPI_ISL_9121053, EPI_ISL_9121061, EPI_ISL_9121063, EPI_ISL_9121064, EPI_ISL_9121067, EPI_ISL_9121072, EPI_ISL_9121075, EPI_ISL_9121079, EPI_ISL_9121106, EPI_ISL_9121116, EPI_ISL_9121120, EPI_ISL_9121124, EPI_ISL_9121127, EPI_ISL_9121128, EPI_ISL_9121132, EPI_ISL_9121137, EPI_ISL_9121171, EPI_ISL_9121174, EPI_ISL_9121177, EPI_ISL_9121187, EPI_ISL_9121188, EPI_ISL_9121191, EPI_ISL_9121197, EPI_ISL_9121205, EPI_ISL_9121208, EPI_ISL_9121216, EPI_ISL_9121217, EPI_ISL_9121225, EPI_ISL_9121229, EPI_ISL_9121231, EPI_ISL_9121244, EPI_ISL_9121245, EPI_ISL_9121253, EPI_ISL_9121257, EPI_ISL_9121272, EPI_ISL_9121287, EPI_ISL_9121292, EPI_ISL_9121293, EPI_ISL_9121297, EPI_ISL_9121311, EPI_ISL_9121313, EPI_ISL_9121314, EPI_ISL_9121317, EPI_ISL_9121329, EPI_ISL_9121344, EPI_ISL_9121348, EPI_ISL_9121349, EPI_ISL_9121356, EPI_ISL_9121365, EPI_ISL_9121370, EPI_ISL_9121385, EPI_ISL_9121398, EPI_ISL_9121403, EPI_ISL_9121404, EPI_ISL_9121413, EPI_ISL_9121414, EPI_ISL_9121423, EPI_ISL_9121428, EPI_ISL_9121438, EPI_ISL_9121443, EPI_ISL_9121446, EPI_ISL_9121451, EPI_ISL_9121469, EPI_ISL_9121472, EPI_ISL_9121484, EPI_ISL_9121497, EPI_ISL_9121498, EPI_ISL_9121501, EPI_ISL_9121502, EPI_ISL_9121503, EPI_ISL_9121509, EPI_ISL_9121510, EPI_ISL_9121514, EPI_ISL_9121526, EPI_ISL_9121531, EPI_ISL_9121547, EPI_ISL_9121548, EPI_ISL_9121550, EPI_ISL_9121553, EPI_ISL_9121556, EPI_ISL_9121559, EPI_ISL_9121561, EPI_ISL_9121568, EPI_ISL_9121570, EPI_ISL_9121575, EPI_ISL_9121578, EPI_ISL_9121584, EPI_ISL_9121588, EPI_ISL_9121594, EPI_ISL_9121598, EPI_ISL_9121615, EPI_ISL_9121618, EPI_ISL_9121619, EPI_ISL_9121623, EPI_ISL_9121624, EPI_ISL_9121628, EPI_ISL_9121640, EPI_ISL_9121645, EPI_ISL_9121692, EPI_ISL_9121698, EPI_ISL_9121726, EPI_ISL_9121728, EPI_ISL_9121738, EPI_ISL_9121739, EPI_ISL_9121743, EPI_ISL_9121745, EPI_ISL_9121747, EPI_ISL_9121748, EPI_ISL_9121750, EPI_ISL_9121758, EPI_ISL_9121759, EPI_ISL_9121768, EPI_ISL_9121769, EPI_ISL_9121772, EPI_ISL_9121786, EPI_ISL_9121789, EPI_ISL_9121791, EPI_ISL_9121832, EPI_ISL_9121833, EPI_ISL_9121836, EPI_ISL_9121838, EPI_ISL_9121851, EPI_ISL_9121852, EPI_ISL_9121854, EPI_ISL_9121855, EPI_ISL_9121872, EPI_ISL_9121874, EPI_ISL_9121876, EPI_ISL_9121882, EPI_ISL_9121888, EPI_ISL_9121890, EPI_ISL_9121897, EPI_ISL_9121904, EPI_ISL_9122021, EPI_ISL_9122025, EPI_ISL_9122029, EPI_ISL_9122081, EPI_ISL_9122088, EPI_ISL_9122090, EPI_ISL_9122096, EPI_ISL_9122097, EPI_ISL_9122098, EPI_ISL_9122104, EPI_ISL_9122108, EPI_ISL_9122109, EPI_ISL_9122113, EPI_ISL_9122126, EPI_ISL_9122131, EPI_ISL_9122134, EPI_ISL_9122139, EPI_ISL_9122140, EPI_ISL_9122141, EPI_ISL_9122144, EPI_ISL_9122148, EPI_ISL_9122155, EPI_ISL_9122158, EPI_ISL_9122161, EPI_ISL_9122162, EPI_ISL_9122165, EPI_ISL_9122167, EPI_ISL_9122168, EPI_ISL_9122915, EPI_ISL_9122916, EPI_ISL_9122922, EPI_ISL_9122924, EPI_ISL_9122930, EPI_ISL_9122932, EPI_ISL_9122935, EPI_ISL_9122940, EPI_ISL_9122944, EPI_ISL_9122945, EPI_ISL_9122946, EPI_ISL_9122947, EPI_ISL_9122958, EPI_ISL_9122974, EPI_ISL_9122982, EPI_ISL_9122987, EPI_ISL_9122998, EPI_ISL_9123123, EPI_ISL_9123135, EPI_ISL_9123137, EPI_ISL_9123139, EPI_ISL_9123140, EPI_ISL_9123142, EPI_ISL_9123144, EPI_ISL_9123156, EPI_ISL_9123171, EPI_ISL_9123172, EPI_ISL_9123175, EPI_ISL_9123179, EPI_ISL_9123191, EPI_ISL_9123193, EPI_ISL_9123201, EPI_ISL_9123215, EPI_ISL_9123216, EPI_ISL_9123229, EPI_ISL_9123234, EPI_ISL_9123236, EPI_ISL_9123248, EPI_ISL_9123253, EPI_ISL_9123260, EPI_ISL_9123263, EPI_ISL_9123264, EPI_ISL_9123267, EPI_ISL_9123272, EPI_ISL_9123273, EPI_ISL_9123276, EPI_ISL_9123290, EPI_ISL_9123292, EPI_ISL_9123295, EPI_ISL_9123299, EPI_ISL_9123305, EPI_ISL_9123310, EPI_ISL_9123312, EPI_ISL_9123313, EPI_ISL_9123318, EPI_ISL_9123343, EPI_ISL_9123351, EPI_ISL_9123362, EPI_ISL_9123363, EPI_ISL_9123366, EPI_ISL_9123369, EPI_ISL_9123373, EPI_ISL_9123378, EPI_ISL_9123380, EPI_ISL_9123382, EPI_ISL_9123383, EPI_ISL_9123392, EPI_ISL_9123398, EPI_ISL_9123399, EPI_ISL_9123410, EPI_ISL_9123416, EPI_ISL_9123426, EPI_ISL_9123430, EPI_ISL_9123433, EPI_ISL_9123438, EPI_ISL_9123443, EPI_ISL_9123447, EPI_ISL_9123454, EPI_ISL_9123455, EPI_ISL_9123464, EPI_ISL_9123479, EPI_ISL_9123480, EPI_ISL_9123488, EPI_ISL_9123490, EPI_ISL_9123491, EPI_ISL_9123493, EPI_ISL_9123501, EPI_ISL_9123506, EPI_ISL_9123515, EPI_ISL_9123529, EPI_ISL_9123533, EPI_ISL_9123537, EPI_ISL_9123547, EPI_ISL_9123549, EPI_ISL_9123553, EPI_ISL_9123554, EPI_ISL_9123560, EPI_ISL_9123563, EPI_ISL_9123755, EPI_ISL_9123757, EPI_ISL_9123759, EPI_ISL_9123761, EPI_ISL_9123763, EPI_ISL_9123770, EPI_ISL_9123772, EPI_ISL_9123773, EPI_ISL_9123779, EPI_ISL_9123785, EPI_ISL_9123786, EPI_ISL_9123792, EPI_ISL_9123797, EPI_ISL_9123994, EPI_ISL_9124003, EPI_ISL_9124004, EPI_ISL_9124005, EPI_ISL_9124008, EPI_ISL_9124009, EPI_ISL_9124018, EPI_ISL_9124022, EPI_ISL_9124027, EPI_ISL_9124030, EPI_ISL_9124031, EPI_ISL_9124034, EPI_ISL_9124046, EPI_ISL_9124049, EPI_ISL_9124080, EPI_ISL_9124097, EPI_ISL_9124100, EPI_ISL_9124120, EPI_ISL_9124132, EPI_ISL_9124137, EPI_ISL_9124158, EPI_ISL_9124166, EPI_ISL_9124168, EPI_ISL_9124267, EPI_ISL_9124276, EPI_ISL_9124299, EPI_ISL_9124329, EPI_ISL_9124347, EPI_ISL_9124364, EPI_ISL_9124367, EPI_ISL_9124390, EPI_ISL_9124393, EPI_ISL_9124413, EPI_ISL_9124429, EPI_ISL_9124447, EPI_ISL_9124452, EPI_ISL_9124485, EPI_ISL_9124491, EPI_ISL_9124500, EPI_ISL_9124520, EPI_ISL_9124526, EPI_ISL_9124537, EPI_ISL_9124542, EPI_ISL_9124553, EPI_ISL_9124558, EPI_ISL_9124565, EPI_ISL_9124567, EPI_ISL_9124571, EPI_ISL_9124576, EPI_ISL_9124584, EPI_ISL_9124592, EPI_ISL_9124613, EPI_ISL_9124635, EPI_ISL_9124652, EPI_ISL_9124694, EPI_ISL_9124721, EPI_ISL_9124726, EPI_ISL_9124729, EPI_ISL_9124733</td></tr>
<tr><td>see above</td><td>Department of Bacteria, Parasites and Fungi, Statens Serum Institut, Copenhagen, Denmark</td><td>Statens Serum Institut Bioinformatics and Microbial Genomics</td><td>Danish Covid-19 Genome Consortium</td></tr>
</table>

We gratefully acknowledge the following Authors from the Originating laboratories responsible for obtaining the specimens, as well as the Submitting laboratories where the genome data were generated and shared via GISAID, on which this research is based.

All Submitters of data may be contacted directly via www.gisaid.org

Authors are sorted alphabetically.

| Accession ID | Originating Laboratory | Submitting Laboratory | Authors |
|---|---|---|---|
| EPI_ISL_9128636, EPI_ISL_9128638, EPI_ISL_9128664, EPI_ISL_9128670, EPI_ISL_9128679, EPI_ISL_9128751, EPI_ISL_9128771, EPI_ISL_9128786, EPI_ISL_9128793, EPI_ISL_9128800, EPI_ISL_9128817, EPI_ISL_9128835, EPI_ISL_9128869, EPI_ISL_9128878, EPI_ISL_9128919, EPI_ISL_9128934, EPI_ISL_9128970, EPI_ISL_9128973, EPI_ISL_9129020, EPI_ISL_9129036, EPI_ISL_9129072, EPI_ISL_9129100, EPI_ISL_9129109, EPI_ISL_9131406, EPI_ISL_9131409, EPI_ISL_9131422, EPI_ISL_9131438, EPI_ISL_9131446, EPI_ISL_9131453, EPI_ISL_9131484, EPI_ISL_9131520, EPI_ISL_9131559, EPI_ISL_9131576, EPI_ISL_9131599, EPI_ISL_9131617, EPI_ISL_9131623, EPI_ISL_9131632, EPI_ISL_9131647, EPI_ISL_9131684, EPI_ISL_9131727, EPI_ISL_9131786, EPI_ISL_9132000, EPI_ISL_9134248, EPI_ISL_9134301, EPI_ISL_9134316, EPI_ISL_9134323, EPI_ISL_9134326, EPI_ISL_9134332, EPI_ISL_9134627, EPI_ISL_9134644 | | | |
| see above | Department of Bacteria, Parasites and Fungi, Statens Serum Institut, Copenhagen, Denmark | Statens Serum Institut Bioinformatics and Microbial Genomics | Danish Covid-19 Genome Consortium |

We gratefully acknowledge the following Authors from the Originating laboratories responsible for obtaining the specimens, as well as the Submitting laboratories where the genome data were generated and shared via GISAID, on which this research is based.

All Submitters of data may be contacted directly via www.gisaid.org

Authors are sorted alphabetically.

| Accession ID | Originating Laboratory | Submitting Laboratory | Authors |
|---|---|---|---|
| EPI_ISL_9127795, EPI_ISL_9127798, EPI_ISL_9127805, EPI_ISL_9127816, EPI_ISL_9127820, EPI_ISL_9127836, EPI_ISL_9127839, EPI_ISL_9127844, EPI_ISL_9127856, EPI_ISL_9127865, EPI_ISL_9127868, EPI_ISL_9127879, EPI_ISL_9127905, EPI_ISL_9127933, EPI_ISL_9127938, EPI_ISL_9128031, EPI_ISL_9128034, EPI_ISL_9128042, EPI_ISL_9128074, EPI_ISL_9128093, EPI_ISL_9128099, EPI_ISL_9128116, EPI_ISL_9128118, EPI_ISL_9128151, EPI_ISL_9128180, EPI_ISL_9128191, EPI_ISL_9128207, EPI_ISL_9128222, EPI_ISL_9128224, EPI_ISL_9128230, EPI_ISL_9128241, EPI_ISL_9128276, EPI_ISL_9128282, EPI_ISL_9128285, EPI_ISL_9128299, EPI_ISL_9128336, EPI_ISL_9128339, EPI_ISL_9128351, EPI_ISL_9128371, EPI_ISL_9128382, EPI_ISL_9128429, EPI_ISL_9128431, EPI_ISL_9128471, EPI_ISL_9128476, EPI_ISL_9128482, EPI_ISL_9128502, EPI_ISL_9128504, EPI_ISL_9128523, EPI_ISL_9128590, EPI_ISL_9128630 | | | |
| see above | Department of Bacteria, Parasites and Fungi, Statens Serum Institut, Copenhagen, Denmark | Statens Serum Institut Bioinformatics and Microbial Genomics | Danish Covid-19 Genome Consortium |

We gratefully acknowledge the following Authors from the Originating laboratories responsible for obtaining the specimens, as well as the Submitting laboratories where the genome data were generated and shared via GISAID, on which this research is based.

All Submitters of data may be contacted directly via www.gisaid.org

Authors are sorted alphabetically.

Acknowledgement EPI_SET Identifier: EPI_SET_20220527go

| Accession ID | Originating Laboratory | Submitting Laboratory | Authors |
|---|---|---|---|
| EPI_ISL_9126266, EPI_ISL_9126274, EPI_ISL_9126284, EPI_ISL_9126293, EPI_ISL_9126498, EPI_ISL_9126500, EPI_ISL_9126502, EPI_ISL_9126843, EPI_ISL_9126853, EPI_ISL_9126861, EPI_ISL_9126898, EPI_ISL_9126908, EPI_ISL_9126922, EPI_ISL_9126946, EPI_ISL_9126963, EPI_ISL_9126971, EPI_ISL_9127005, EPI_ISL_9127035, EPI_ISL_9127065, EPI_ISL_9127067, EPI_ISL_9127071, EPI_ISL_9127146, EPI_ISL_9127285, EPI_ISL_9127289, EPI_ISL_9127307, EPI_ISL_9127313, EPI_ISL_9127324, EPI_ISL_9127343, EPI_ISL_9127346, EPI_ISL_9127383, EPI_ISL_9127392, EPI_ISL_9127420, EPI_ISL_9127424, EPI_ISL_9127439, EPI_ISL_9127447, EPI_ISL_9127461, EPI_ISL_9127481, EPI_ISL_9127489, EPI_ISL_9127494, EPI_ISL_9127518, EPI_ISL_9127527, EPI_ISL_9127540, EPI_ISL_9127618, EPI_ISL_9127624, EPI_ISL_9127649, EPI_ISL_9127666, EPI_ISL_9127706, EPI_ISL_9127742, EPI_ISL_9127756, EPI_ISL_9127792 | | | |
| see above | Department of Bacteria, Parasites and Fungi, Statens Serum Institut, Copenhagen, Denmark | Statens Serum Institut Bioinformatics and Microbial Genomics | Danish Covid-19 Genome Consortium |

We gratefully acknowledge the following Authors from the Originating laboratories responsible for obtaining the specimens, as well as the Submitting laboratories where the genome data were generated and shared via GISAID, on which this research is based.

All Submitters of data may be contacted directly via www.gisaid.org

Authors are sorted alphabetically.

EPI_ISL_9124736, EPI_ISL_9124790, EPI_ISL_9124796, EPI_ISL_9124818, EPI_ISL_9124821, EPI_ISL_9124826, EPI_ISL_9124841, EPI_ISL_9124844, EPI_ISL_9124878, EPI_ISL_9124926, EPI_ISL_9124929, EPI_ISL_9124932, EPI_ISL_9124937, EPI_ISL_9124950, EPI_ISL_9124958, EPI_ISL_9124977, EPI_ISL_9124982, EPI_ISL_9125291, EPI_ISL_9125295, EPI_ISL_9125388, EPI_ISL_9125410, EPI_ISL_9125424, EPI_ISL_9125427, EPI_ISL_9125449, EPI_ISL_9125457, EPI_ISL_9125472, EPI_ISL_9125481, EPI_ISL_9125483, EPI_ISL_9125526, EPI_ISL_9125534, EPI_ISL_9125881, EPI_ISL_9125920, EPI_ISL_9125934, EPI_ISL_9125937, EPI_ISL_9125955, EPI_ISL_9125964, EPI_ISL_9125994, EPI_ISL_9126002, EPI_ISL_9126005, EPI_ISL_9126009, EPI_ISL_9126025, EPI_ISL_9126042, EPI_ISL_9126086, EPI_ISL_9126100, EPI_ISL_9126117, EPI_ISL_9126131, EPI_ISL_9126143, EPI_ISL_9126145, EPI_ISL_9126225, EPI_ISL_9126247

| Accession ID | Originating Laboratory | Submitting Laboratory | Authors |
|---|---|---|---|
| see above | Department of Bacteria, Parasites and Fungi, Statens Serum Institut, Copenhagen, Denmark | Statens Serum Institut Bioinformatics and Microbial Genomics | Danish Covid-19 Genome Consortium |

We gratefully acknowledge the following Authors from the Originating laboratories responsible for obtaining the specimens, as well as the Submitting laboratories where the genome data were generated and shared via GISAID, on which this research is based.

All Submitters of data may be contacted directly via www.gisaid.org

Authors are sorted alphabetically.

| Accession ID | Originating Laboratory | Submitting Laboratory | Authors |
| --- | --- | --- | --- |
| EPI_ISL_9123002, EPI_ISL_9123003, EPI_ISL_9123004 | Lifebrain Covid Labor GmbH | Lifebrain Covid Labor GmbH | Alexandra Wagner; Anna Edermayr; Filip Sima |

We gratefully acknowledge the following Authors from the Originating laboratories responsible for obtaining the specimens, as well as the Submitting laboratories where the genome data were generated and shared via GISAID, on which this research is based.

All Submitters of data may be contacted directly via www.gisaid.org

Authors are sorted alphabetically.

| Accession ID | Originating Laboratory | Submitting Laboratory | Authors |
|---|---|---|---|
| EPI_ISL_9143636 | Hrvatski zavod za javno zdravstvo | Hrvatski zavod za javno zdravstvo | Anita Jurić; Dragan Jurić; Irena Tabain; Ivana Ferenčak; Josipa Kuzle |

We gratefully acknowledge the following Authors from the Originating laboratories responsible for obtaining the specimens, as well as the Submitting laboratories where the genome data were generated and shared via GISAID, on which this research is based.

All Submitters of data may be contacted directly via www.gisaid.org

Authors are sorted alphabetically.

| Accession ID | Originating Laboratory | Submitting Laboratory | Authors |
|---|---|---|---|
| EPI_ISL_9143643 | Zavod za javno zdravstvo Istarske županije | Hrvatski zavod za javno zdravstvo | Anita Jurić; Dragan Jurić; Irena Tabain; Ivana Ferenčak; Josipa Kuzle |

We gratefully acknowledge the following Authors from the Originating laboratories responsible for obtaining the specimens, as well as the Submitting laboratories where the genome data were generated and shared via GISAID, on which this research is based.

All Submitters of data may be contacted directly via www.gisaid.org

Authors are sorted alphabetically.

Acknowledgement EPI_SET Identifier: EPI_SET_20220527xb

| Accession ID | Originating Laboratory | Submitting Laboratory | Authors |
|---|---|---|---|
| EPI_ISL_9143539, EPI_ISL_9143540 | Zavod za javno zdravstvo Ličko-senjske županije | Hrvatski zavod za javno zdravstvo | Anita Jurić; Dragan Jurić; Irena Tabain; Ivana Ferenčak; Josipa Kuzle |

We gratefully acknowledge the following Authors from the Originating laboratories responsible for obtaining the specimens, as well as the Submitting laboratories where the genome data were generated and shared via GISAID, on which this research is based.

All Submitters of data may be contacted directly via www.gisaid.org

Authors are sorted alphabetically.

| Accession ID | Originating Laboratory | Submitting Laboratory | Authors |
| --- | --- | --- | --- |
| EPI_ISL_9143535, EPI_ISL_9143536, EPI_ISL_9143537, EPI_ISL_9143538 | Zavod za javno zdravstvo Šibensko- Kninske županije | Hrvatski zavod za javno zdravstvo | Anita Jurić; Dragan Jurić; Irena Tabain; Ivana Ferenčak; Josipa Kuzle |

We gratefully acknowledge the following Authors from the Originating laboratories responsible for obtaining the specimens, as well as the Submitting laboratories where the genome data were generated and shared via GISAID, on which this research is based.

All Submitters of data may be contacted directly via www.gisaid.org

Authors are sorted alphabetically.

| Accession ID | Originating Laboratory | Submitting Laboratory | Authors |
| --- | --- | --- | --- |
| EPI_ISL_9143619 | Hrvatski zavod za javno zdravstvo | Hrvatski zavod za javno zdravstvo | Anita Jurić; Dragan Jurić; Irena Tabain; Ivana Ferenčak; Josipa Kuzle |

We gratefully acknowledge the following Authors from the Originating laboratories responsible for obtaining the specimens, as well as the Submitting laboratories where the genome data were generated and shared via GISAID, on which this research is based.

All Submitters of data may be contacted directly via www.gisaid.org

Authors are sorted alphabetically.

| Accession ID | Originating Laboratory | Submitting Laboratory | Authors |
| --- | --- | --- | --- |
| EPI_ISL_9143619 | Hrvatski zavod za javno zdravstvo | Hrvatski zavod za javno zdravstvo | Anita Jurić; Dragan Jurić; Irena Tabain; Ivana Ferenčak; Josipa Kuzle |

We gratefully acknowledge the following Authors from the Originating laboratories responsible for obtaining the specimens, as well as the Submitting laboratories where the genome data were generated and shared via GISAID, on which this research is based.

All Submitters of data may be contacted directly via www.gisaid.org

Authors are sorted alphabetically.

| Accession ID | Originating Laboratory | Submitting Laboratory | Authors |
| --- | --- | --- | --- |
| EPI_ISL_9143611, EPI_ISL_9143612, EPI_ISL_9143614 | Hrvatski zavod za javno zdravstvo | Hrvatski zavod za javno zdravstvo | Anita Jurić; Dragan Jurić; Irena Tabain; Ivana Ferenčak; Josipa Kuzle |

We gratefully acknowledge the following Authors from the Originating laboratories responsible for obtaining the specimens, as well as the Submitting laboratories where the genome data were generated and shared via GISAID, on which this research is based.

All Submitters of data may be contacted directly via www.gisaid.org

Authors are sorted alphabetically.

| Accession ID | Originating Laboratory | Submitting Laboratory | Authors |
|---|---|---|---|
| EPI_ISL_9143613, EPI_ISL_9143615, EPI_ISL_9143616, EPI_ISL_9143617, EPI_ISL_9143618, EPI_ISL_9143620, EPI_ISL_9143621, EPI_ISL_9143622, EPI_ISL_9143623, EPI_ISL_9143624, EPI_ISL_9143625, EPI_ISL_9143626, EPI_ISL_9143627, EPI_ISL_9143628, EPI_ISL_9143629, EPI_ISL_9143630, EPI_ISL_9143631, EPI_ISL_9143632, EPI_ISL_9143633, EPI_ISL_9143634, EPI_ISL_9143635, EPI_ISL_9143637, EPI_ISL_9143638, EPI_ISL_9143639, EPI_ISL_9143640, EPI_ISL_9143641, EPI_ISL_9143642, EPI_ISL_9143644 | | | |
| see above | Hrvatski zavod za javno zdravstvo | Hrvatski zavod za javno zdravstvo | Anita Jurić; Dragan Jurić; Irena Tabain; Ivana Ferenčak; Josipa Kuzle |
| EPI_ISL_9143679, EPI_ISL_9143680, EPI_ISL_9143681, EPI_ISL_9143682, EPI_ISL_9143683, EPI_ISL_9143684, EPI_ISL_9143685, EPI_ISL_9143686, EPI_ISL_9143687, EPI_ISL_9143688, EPI_ISL_9143689, EPI_ISL_9143690, EPI_ISL_9143691, EPI_ISL_9143692, EPI_ISL_9143693, EPI_ISL_9143694, EPI_ISL_9143695, EPI_ISL_9143696, EPI_ISL_9143697, EPI_ISL_9143698, EPI_ISL_9143699, EPI_ISL_9143700, EPI_ISL_9143701, EPI_ISL_9143702, EPI_ISL_9143703, EPI_ISL_9143704, EPI_ISL_9143705, EPI_ISL_9143706, EPI_ISL_9143707, EPI_ISL_9143708, EPI_ISL_9143709, EPI_ISL_9143710, EPI_ISL_9143711, EPI_ISL_9143712, EPI_ISL_9143713, EPI_ISL_9143714, EPI_ISL_9143715, EPI_ISL_9143716, EPI_ISL_9143717, EPI_ISL_9143718, EPI_ISL_9143719, EPI_ISL_9143720, EPI_ISL_9143721, EPI_ISL_9143722, EPI_ISL_9143723, EPI_ISL_9143724, EPI_ISL_9143725, EPI_ISL_9143726, EPI_ISL_9143727, EPI_ISL_9143728, EPI_ISL_9143729, EPI_ISL_9143730, EPI_ISL_9143731, EPI_ISL_9143732, EPI_ISL_9143734, EPI_ISL_9143735, EPI_ISL_9143736, EPI_ISL_9143737, EPI_ISL_9143738, EPI_ISL_9143739 | | | |
| see above | Klinika za infektivne bolesti "Dr. Fran Mihaljević" | Hrvatski zavod za javno zdravstvo | Anita Jurić; Dragan Jurić; Irena Tabain; Ivana Ferenčak; Josipa Kuzle |

We gratefully acknowledge the following Authors from the Originating laboratories responsible for obtaining the specimens, as well as the Submitting laboratories where the genome data were generated and shared via GISAID, on which this research is based.

All Submitters of data may be contacted directly via www.gisaid.org

Authors are sorted alphabetically.

Acknowledgement EPI_SET Identifier: EPI_SET_20220527bt

| Accession ID | Originating Laboratory | Submitting Laboratory | Authors |
|---|---|---|---|
| EPI_ISL_9106321, EPI_ISL_9106341, EPI_ISL_9106347 | Limbach - MVZ Labor Ludwigsburg | Robert Koch Institute | |
| EPI_ISL_9107465, EPI_ISL_9107469, EPI_ISL_9107476, EPI_ISL_9107494, EPI_ISL_9108599 | MVZ Labor Dr. Limbach & Kollegen GbR | Robert Koch Institute | |
| EPI_ISL_9107666, EPI_ISL_9107668 | SYNLAB MVZ Leinfelden-Echterdingen | Robert Koch Institute | |

We gratefully acknowledge the following Authors from the Originating laboratories responsible for obtaining the specimens, as well as the Submitting laboratories where the genome data were generated and shared via GISAID, on which this research is based.

All Submitters of data may be contacted directly via www.gisaid.org

Authors are sorted alphabetically.

| Accession ID | Originating Laboratory | Submitting Laboratory | Authors |
|---|---|---|---|
| EPI_ISL_9110249 | IMD - Institut für Medizinische Diagnostik Berlin-Potsdam | Robert Koch Institute | |
| EPI_ISL_9108810, EPI_ISL_9108829, EPI_ISL_9108835, EPI_ISL_9108852 | MDI Limbach Berlin GmbH; MVZ Labor Berlin | Robert Koch Institute | |

We gratefully acknowledge the following Authors from the Originating laboratories responsible for obtaining the specimens, as well as the Submitting laboratories where the genome data were generated and shared via GISAID, on which this research is based.

All Submitters of data may be contacted directly via www.gisaid.org

Authors are sorted alphabetically.

| Accession ID | Originating Laboratory | Submitting Laboratory | Authors |
| --- | --- | --- | --- |
| EPI_ISL_9108907 | MVZ Medizinisches Labor Hannover GmbH | Robert Koch Institute | |

We gratefully acknowledge the following Authors from the Originating laboratories responsible for obtaining the specimens, as well as the Submitting laboratories where the genome data were generated and shared via GISAID, on which this research is based.

All Submitters of data may be contacted directly via www.gisaid.org

Authors are sorted alphabetically.

| Accession ID | Originating Laboratory | Submitting Laboratory | Authors |
|---|---|---|---|
| EPI_ISL_9107511, EPI_ISL_9107535 | Clemenshospital - Alexianer Clemenshospital Münster | Robert Koch Institute | |
| EPI_ISL_9109828 | MVZ Labor Dr. Quade & Kollegen GmbH | Robert Koch Institute | |

We gratefully acknowledge the following Authors from the Originating laboratories responsible for obtaining the specimens, as well as the Submitting laboratories where the genome data were generated and shared via GISAID, on which this research is based.

All Submitters of data may be contacted directly via www.gisaid.org

Authors are sorted alphabetically.

| Accession ID | Originating Laboratory | Submitting Laboratory | Authors |
|---|---|---|---|
| EPI_ISL_9111052, EPI_ISL_9111116 | A.S.L. ALESSANDRIA | Fondazione del Piemonte per l'Oncologia IRCCS | Antonino Sottile; Giorgio Giardina; Paola Marino; Silvia Brossa |
| EPI_ISL_9110835, EPI_ISL_9110844, EPI_ISL_9110860, EPI_ISL_9110864 | Arcispedale Santa Maria Nuova Autoimmunità Allergologia e Biotecnologie Innovative | Istituto Zooprofilattico Sperimentale della Lombardia e dell'Emilia Romagna (IZSLER), Risk Analysis and Genomic Epidemiology Unit | Alessandro Zerbini; Erika Scaltriti; Ilaria Menozzi; Lucia Belloni; Marina Morganti; Stefania Croci; Stefano Pongolini |
| EPI_ISL_9110686, EPI_ISL_9110695 | Azienda Sanitaria Locale di Piacenza - Presidio Ospedaliero - Laboratorio di Microbiologia | Istituto Zooprofilattico Sperimentale della Lombardia e dell'Emilia Romagna (IZSLER), Risk Analysis and Genomic Epidemiology Unit | Erika Scaltriti; Ilaria Menozzi; Lo Cascio Giuliana; Marina Morganti; Peroni Irene; Schiavo Roberta; Stefano Pongolini |
| EPI_ISL_9110772 | Universita di Parma Laboratorio di Igiene e Sanita Pubblica | Istituto Zooprofilattico Sperimentale della Lombardia e dell'Emilia Romagna (IZSLER), Risk Analysis and Genomic Epidemiology Unit | Erika Scaltriti; Ilaria Menozzi; Marina Morganti; Stefano Pongolini |

We gratefully acknowledge the following Authors from the Originating laboratories responsible for obtaining the specimens, as well as the Submitting laboratories where the genome data were generated and shared via GISAID, on which this research is based.

All Submitters of data may be contacted directly via www.gisaid.org

Authors are sorted alphabetically.

| Accession ID | Originating Laboratory | Submitting Laboratory | Authors |
|---|---|---|---|
| EPI_ISL_9139390 | Microvida | Microvida | Jaco J. Verweij; Joep J. J. M. Stohr; Suzan D. Pas |

We gratefully acknowledge the following Authors from the Originating laboratories responsible for obtaining the specimens, as well as the Submitting laboratories where the genome data were generated and shared via GISAID, on which this research is based.

All Submitters of data may be contacted directly via www.gisaid.org

Authors are sorted alphabetically.

| Accession ID | Originating Laboratory | Submitting Laboratory | Authors |
|---|---|---|---|
| EPI_ISL_9129404 | Lighthouse Lab in Milton Keynes | Wellcome Sanger Institute for the COVID-19 Genomics UK (COG-UK) Consortium | Cordelia Langford; David K. Jackson; Dominic Kwiatkowski; Ewan Harrison; Ian Johnston; Jeffrey Barrett; John Sillitoe on behalf of the Wellcome Sanger Institute COVID-19 Surveillance Team; Roberto Amato; Sonia Goncalves; The Lighthouse Lab in Milton Keynes and Alex Alderton |

We gratefully acknowledge the following Authors from the Originating laboratories responsible for obtaining the specimens, as well as the Submitting laboratories where the genome data were generated and shared via GISAID, on which this research is based.

All Submitters of data may be contacted directly via www.gisaid.org

Authors are sorted alphabetically.

Acknowledgement EPI_SET Identifier: EPI_SET_20220527qv

| Accession ID | Originating Laboratory | Submitting Laboratory | Authors |
|---|---|---|---|
| EPI_ISL_9175910, EPI_ISL_9175911, EPI_ISL_9175996, EPI_ISL_9175998, EPI_ISL_9175999, EPI_ISL_9176000, EPI_ISL_9176001, EPI_ISL_9176002 | | | |
| see above | BBMP- Mahadevapura | inStem, NCBS-INSACOG | Aswin SaiNarain; Dasaradhi Palakodeti; Uma Ramakrishnan |
| EPI_ISL_9175915, EPI_ISL_9175931, EPI_ISL_9175932, EPI_ISL_9175935, EPI_ISL_9175936, EPI_ISL_9175937, EPI_ISL_9175938, EPI_ISL_9175939, EPI_ISL_9175940, EPI_ISL_9176034, EPI_ISL_9176041, EPI_ISL_9176043, EPI_ISL_9176045, EPI_ISL_9176046 | | | |
| see above | BBMP- West | inStem, NCBS-INSACOG | Aswin SaiNarain; Dasaradhi Palakodeti; Uma Ramakrishnan |
| EPI_ISL_9175916, EPI_ISL_9175919, EPI_ISL_9175920, EPI_ISL_9175921, EPI_ISL_9175923, EPI_ISL_9175925, EPI_ISL_9175926, EPI_ISL_9175927 | | | |
| see above | BBMP- Yelahanka | inStem, NCBS-INSACOG | Aswin SaiNarain; Dasaradhi Palakodeti; Uma Ramakrishnan |
| EPI_ISL_9176003, EPI_ISL_9176009, EPI_ISL_9176011, EPI_ISL_9176016, EPI_ISL_9176018, EPI_ISL_9176019, EPI_ISL_9176020, EPI_ISL_9176022, EPI_ISL_9176023 | | | |
| see above | BBMP-Dasarahalli | inStem, NCBS-INSACOG | Aswin SaiNarain; Dasaradhi Palakodeti; Uma Ramakrishnan |
| EPI_ISL_9175909 | BBMP-Mahadevapura | inStem, NCBS-INSACOG | Aswin SaiNarain; Dasaradhi Palakodeti; Uma Ramakrishnan |
| EPI_ISL_9175941, EPI_ISL_9175943, EPI_ISL_9175946, EPI_ISL_9175947, EPI_ISL_9175949, EPI_ISL_9175953, EPI_ISL_9175954, EPI_ISL_9175955, EPI_ISL_9175956, EPI_ISL_9175957, EPI_ISL_9175959, EPI_ISL_9175960, EPI_ISL_9175961, EPI_ISL_9175965, EPI_ISL_9175967, EPI_ISL_9175970, EPI_ISL_9175971, EPI_ISL_9175973, EPI_ISL_9175978, EPI_ISL_9175979, EPI_ISL_9175981, EPI_ISL_9175982, EPI_ISL_9175983, EPI_ISL_9175984, EPI_ISL_9175985, EPI_ISL_9175988, EPI_ISL_9175991, EPI_ISL_9175992, EPI_ISL_9175993 | | | |
| see above | Bangalore Airport | inStem, NCBS-INSACOG | Aswin SaiNarain; Dasaradhi Palakodeti; Uma Ramakrishnan |
| EPI_ISL_9176024, EPI_ISL_9176025, EPI_ISL_9176026, EPI_ISL_9176028, EPI_ISL_9176030, EPI_ISL_9176031, EPI_ISL_9176032 | | | |
| see above | Bengaluru Rural | inStem, NCBS-INSACOG | Aswin SaiNarain; Dasaradhi Palakodeti; Uma Ramakrishnan |
| EPI_ISL_9161266, EPI_ISL_9161312, EPI_ISL_9161326, EPI_ISL_9164578 | Genome Information Research Center, Research Institute for Microbial Diseases, Osaka University | Genome Information Research Center, Research Institute for Microbial Diseases, Osaka University | Daisuke Motooka; Hiroya Oki; Kentaro Tanaka; and Shota Nakamura |
| EPI_ISL_9166255 | MapMyGenome | CSIR-Centre for Cellular and Molecular Biology-INSACOG | Amareshwar Vodapalli; Anuradha Acharya; Ara Sreenivas; Archana Bharadwaj Siva; B Himasri; Divya Tej Sowpati; Jandhyala Sai Krishna; Karthik Bharadwaj Tallapaka; Lamuk Zaveri; M. Aravind Kumar; Malini Nemalikanti; Priya Nurkurthy; Rakesh K Mishra; S.P.S. Satish; Sandhya Kiran; Shreekant Verma; Sreelekshmi MS; Sumedha Avadhanula; Surabhi Srivastava; Tulasi Nagabandi; Valli Nagalakshmi Undamatla; Vidhyadhari Methuku |

We gratefully acknowledge the following Authors from the Originating laboratories responsible for obtaining the specimens, as well as the Submitting laboratories where the genome data were generated and shared via GISAID, on which this research is based.

All Submitters of data may be contacted directly via www.gisaid.org

Authors are sorted alphabetically.

| Accession ID | Originating Laboratory | Submitting Laboratory | Authors |
|---|---|---|---|
| EPI_ISL_9197851 | Edmonton Provincial Lab | Public Health Agency of Canada (PHAC) National Microbiology Laboratory | Buss; Croxen M; Deo A; Dieu P; E; Ferrato C; Gill K; Koleva P; Li V; Lloyd C; Lynch T; Ma R; Murphy S; Pabbaraju K; Shideler S.; Shokoples S; Skitsko T; Thayer J; Tipples G; Wong A; Yu C; Zelyas N |
| EPI_ISL_9196371, EPI_ISL_9197958, EPI_ISL_9197966, EPI_ISL_9197992, EPI_ISL_9198059, EPI_ISL_9198061, EPI_ISL_9198127, EPI_ISL_9198150, EPI_ISL_9198156, EPI_ISL_9198230, EPI_ISL_9198317, EPI_ISL_9198325, EPI_ISL_9198375, EPI_ISL_9198394, EPI_ISL_9198431, EPI_ISL_9198639, EPI_ISL_9198737, EPI_ISL_9198908, EPI_ISL_9199414, EPI_ISL_9199528, EPI_ISL_9199533, EPI_ISL_9199543, EPI_ISL_9199652, EPI_ISL_9199684, EPI_ISL_9199742, EPI_ISL_9199922, EPI_ISL_9199938, EPI_ISL_9200022, EPI_ISL_9200126, EPI_ISL_9200129, EPI_ISL_9200291, EPI_ISL_9200390, EPI_ISL_9200435, EPI_ISL_9200441, EPI_ISL_9200462, EPI_ISL_9200476, EPI_ISL_9200478, EPI_ISL_9200507, EPI_ISL_9200522, EPI_ISL_9200544, EPI_ISL_9200629, EPI_ISL_9200714, EPI_ISL_9200724, EPI_ISL_9200725, EPI_ISL_9200866 | | | |
| see above | Houston Methodist Hospital | Houston Methodist Hospital | Akanksha Batajoo; Ilya J. Finkelstein; James J. Davis; Jessica Cambric; Jimmy Gollihar; Kristina Reppond; Madison N. Shyer; Marcus Nguyen; Matthew Ojeda Saavedra; Paul A. Christensen; Randall J. Olsen; Rashi Thakur; Regan Mangham; Richard Snehall; Robert Olson; Ryan Gadd; S. Wesley Long; Sindy Pena; and James M. Musser |
| EPI_ISL_9177653 | Kansas Health and Environmental Lab | Kansas Health and Environmental Lab | Amanda Bradley; Carrie Welch; Gary Buruss; Jonathan Barnell; Mike Grose; Victor Anderson; and Phil Adam |
| EPI_ISL_9188890, EPI_ISL_9188891, EPI_ISL_9188892, EPI_ISL_9188920 | Laboratory Corporation of America | Centers for Disease Control and Prevention Division of Viral Diseases, Pathogen Discovery | Amanda Douglas; Amanda Suchanek; Andrea Throop; Ayla Burns; Benjamin Rambo-Martin; Bobbi Croy; Brian Krueger; Brian Norvell; Christopher Gulvick; Christos Petropoulos; Clinton Paden; Craig Lukasik; Dakota Howard; Debbie Boles; Dhwani Batra; Duncan MacCannell; Eyad Almasri; Goran Stevovic; Howard Engler; Hrushikesh Deshmukh; Jake Humphrey; Jana Schroth; Jason Caravas; Joe Voshell; John Pruitt; Jonathan Meltzer; Jonathan Williams; Kimberly Wagner; Kristine Lacek; Lax Iyer; Lisa Pfefferle; Lyndon Tilson; Manoj Jain; Marcia Eisenberg; Mary Cristobal; Mary Williamson; Matthew Robinson; Matthew Schmerer; Michael Levandoski; Mike Sapeta; Mindy Nye; Minoo Agarwal; Mohan Kolli; Nuthawin Charoensri; Oren Cohen; Peter Cook; Prashant Gupta; Qian Zeng; Rama Ghatti; Scott Parker; Scott Ryan; Scott Sammons; Shatavia Morrison; Stanley Letovsky; Steven Ragan; Suresh Selvaraju; Susan Countryman; Susan Hicks; Suzanne Dale; Thomas Urban; Tim Kuphal; Tricia Zwiefelhofer; Tymeckia Kendall; Victoria Caban Figueroa; Vincent Drouillon; Yvette Unoarumhi |
| EPI_ISL_9184105, EPI_ISL_9184106, EPI_ISL_9184107, EPI_ISL_9184108 | Ochsner Health | BioInfoExperts | Amy Feehan; Ben Lain; Chris Huston; David J. Nolan; Judy Crabtree; Julia-Garcia-Diaz; Lucio Miele; Rebecca Rose; Samual Moot; Susanna L. Lamers; Tessa LaFleur |
| EPI_ISL_9167820 | SALUD DIGNA | Instituto Nacional de Medicina Genomica | Abraham Campos-Romero; Cedro-Tanda A; Cisneros-Villanueva M; Escobar-Arrazola MA; Garnica-Lopez Dora; Herrera-Montalvo LA.; Hidalgo-Miranda A; Luna-Ruiz Marco; Mendoza-Vargas A; Moreno-Camacho José Luis; Ramirez-Vega O; Rangel-DeLeon D; Reyes-Grajeda JP; Rodriguez-Gallegos Jorge; Yair Alfaro-Mora |
| EPI_ISL_9196278, EPI_ISL_9196321 | State Laboratories Division, Hawaii State Department of Health | State Laboratories Division, Hawaii State Department of Health | Ayana Garnet; Briana Ofilas; Cheryl-lynn Daquip; Cheyenne Barela; Daniel Strange; Drew Kuwazaki; Edward Desmond; Jeffrey Au; Mark Nagata; Pamela O'Brien; Razvan Sultana; Remedios Gose; Samantha Cotter; Samantha Sruba |
| EPI_ISL_9154978 | UW Virology Lab | UW Virology Lab | Alexander Greninger; Hong Xie; Isabel Arnould; Keith R Jerome; Meei-Li Huang; Nathan Breit; Patrick Mathias; Pavitra Roychoudhury; Pooneh Hajian; Ricardo Perez; Robert J. Livingston; Sean Ellis; Seffir T. Wendm; Shah Mohamed Bakhash |

We gratefully acknowledge the following Authors from the Originating laboratories responsible for obtaining the specimens, as well as the Submitting laboratories where the genome data were generated and shared via GISAID, on which this research is based.

All Submitters of data may be contacted directly via www.gisaid.org

Authors are sorted alphabetically.

| Accession ID | Originating Laboratory | Submitting Laboratory | Authors |
|---|---|---|---|
| EPI_ISL_9165857 | Lifebrain Covid Labor GmbH | Lifebrain Covid Labor GmbH | Alexandra Wagner; Anna Edermayr; Filip Sima |

We gratefully acknowledge the following Authors from the Originating laboratories responsible for obtaining the specimens, as well as the Submitting laboratories where the genome data were generated and shared via GISAID, on which this research is based.

All Submitters of data may be contacted directly via www.gisaid.org

Authors are sorted alphabetically.

Acknowledgement EPI_SET Identifier: EPI_SET_20220527dg

| Accession ID | Originating Laboratory | Submitting Laboratory | Authors |
| --- | --- | --- | --- |
| EPI_ISL_9164048, EPI_ISL_9164064, EPI_ISL_9164068, EPI_ISL_9164109 | Cliniques universitaires Saint-Luc | UCLouvain/IREC/MBLG-CTMA | Benoit Kabamba Mukadi; Bertrand Bearzatto; Jean-Luc Gala; Nicolas Pinte; Paul Blanpain; Simon Ophélie; Valentin Coste |

We gratefully acknowledge the following Authors from the Originating laboratories responsible for obtaining the specimens, as well as the Submitting laboratories where the genome data were generated and shared via GISAID, on which this research is based.

All Submitters of data may be contacted directly via www.gisaid.org

Authors are sorted alphabetically.

| Accession ID | Originating Laboratory | Submitting Laboratory | Authors |
|---|---|---|---|
| EPI_ISL_9177472, EPI_ISL_9177474, EPI_ISL_9178070, EPI_ISL_9178071 | CHU UCL Namur, Avenue G. Thérasse 1, 5530 Yvoir, Belgium | Plateforme de testing Namuroise | Degosserie Jonathan; Demars Aurore; Denis Olivier; Drugmand Jonathan; Maschietto Céline; Mullier François; Nobis Chloé; Otto Gaetan |

We gratefully acknowledge the following Authors from the Originating laboratories responsible for obtaining the specimens, as well as the Submitting laboratories where the genome data were generated and shared via GISAID, on which this research is based.

All Submitters of data may be contacted directly via www.gisaid.org

Authors are sorted alphabetically.

| Accession ID | Originating Laboratory | Submitting Laboratory | Authors |
|---|---|---|---|
| EPI_ISL_9162360, EPI_ISL_9162374, EPI_ISL_9162378, EPI_ISL_9162422, EPI_ISL_9162426, EPI_ISL_9162434, EPI_ISL_9162436, EPI_ISL_9162442, EPI_ISL_9162455, EPI_ISL_9162467, EPI_ISL_9162471, EPI_ISL_9162503, EPI_ISL_9162511, EPI_ISL_9162535, EPI_ISL_9162538, EPI_ISL_9162546, EPI_ISL_9162550, EPI_ISL_9162602, EPI_ISL_9162607, EPI_ISL_9162608, EPI_ISL_9162612, EPI_ISL_9162621, EPI_ISL_9162645, EPI_ISL_9162653, EPI_ISL_9162654, EPI_ISL_9162665, EPI_ISL_9162675, EPI_ISL_9162679, EPI_ISL_9162687, EPI_ISL_9162699, EPI_ISL_9162710, EPI_ISL_9162712, EPI_ISL_9162731, EPI_ISL_9162763, EPI_ISL_9162768, EPI_ISL_9162770, EPI_ISL_9162775, EPI_ISL_9162785, EPI_ISL_9162804, EPI_ISL_9162806, EPI_ISL_9162819, EPI_ISL_9162829, EPI_ISL_9162857, EPI_ISL_9162869, EPI_ISL_9162897, EPI_ISL_9162920, EPI_ISL_9162936, EPI_ISL_9162944, EPI_ISL_9162989, EPI_ISL_9163016, EPI_ISL_9163030, EPI_ISL_9163042, EPI_ISL_9163044, EPI_ISL_9163056, EPI_ISL_9163062, EPI_ISL_9163076, EPI_ISL_9163096, EPI_ISL_9163116, EPI_ISL_9163180, EPI_ISL_9163188, EPI_ISL_9163189, EPI_ISL_9163227, EPI_ISL_9163258, EPI_ISL_9163268, EPI_ISL_9163290, EPI_ISL_9163301, EPI_ISL_9163317, EPI_ISL_9163322, EPI_ISL_9163343, EPI_ISL_9163347, EPI_ISL_9163383, EPI_ISL_9163388, EPI_ISL_9163396, EPI_ISL_9163401, EPI_ISL_9163445, EPI_ISL_9163448, EPI_ISL_9163450, EPI_ISL_9163453, EPI_ISL_9163461, EPI_ISL_9163486, EPI_ISL_9163492, EPI_ISL_9163493, EPI_ISL_9163495, EPI_ISL_9163508, EPI_ISL_9163536, EPI_ISL_9163580, EPI_ISL_9163587, EPI_ISL_9163600, EPI_ISL_9163606, EPI_ISL_9163616, EPI_ISL_9163620, EPI_ISL_9163622, EPI_ISL_9163666, EPI_ISL_9163690, EPI_ISL_9163692, EPI_ISL_9163710, EPI_ISL_9163748, EPI_ISL_9163758, EPI_ISL_9163760, EPI_ISL_9163773, EPI_ISL_9163788, EPI_ISL_9163789, EPI_ISL_9163794, EPI_ISL_9163807, EPI_ISL_9163817, EPI_ISL_9163834, EPI_ISL_9163841, EPI_ISL_9163853, EPI_ISL_9163867, EPI_ISL_9163896, EPI_ISL_9163902, EPI_ISL_9163908, EPI_ISL_9163922, EPI_ISL_9163928, EPI_ISL_9163932, EPI_ISL_9163938, EPI_ISL_9163982, EPI_ISL_9163986, EPI_ISL_9163992, EPI_ISL_9164002, EPI_ISL_9164004, EPI_ISL_9164021, EPI_ISL_9164147, EPI_ISL_9164152, EPI_ISL_9164169, EPI_ISL_9164171, EPI_ISL_9164182, EPI_ISL_9164206, EPI_ISL_9164212, EPI_ISL_9164227, EPI_ISL_9164244, EPI_ISL_9164268, EPI_ISL_9164277, EPI_ISL_9164287, EPI_ISL_9164298, EPI_ISL_9164311, EPI_ISL_9164313, EPI_ISL_9164322, EPI_ISL_9164345, EPI_ISL_9164369, EPI_ISL_9164383, EPI_ISL_9164388, EPI_ISL_9164414, EPI_ISL_9164442, EPI_ISL_9164445, EPI_ISL_9164458, EPI_ISL_9164464, EPI_ISL_9164466, EPI_ISL_9164472, EPI_ISL_9164475, EPI_ISL_9164479, EPI_ISL_9164496, EPI_ISL_9164529, EPI_ISL_9164537, EPI_ISL_9164566, EPI_ISL_9164599, EPI_ISL_9164606, EPI_ISL_9164613, EPI_ISL_9164618, EPI_ISL_9164624, EPI_ISL_9164645, EPI_ISL_9164647, EPI_ISL_9164661, EPI_ISL_9164682, EPI_ISL_9164691, EPI_ISL_9164693, EPI_ISL_9164697, EPI_ISL_9164703, EPI_ISL_9164717, EPI_ISL_9164734, EPI_ISL_9164738, EPI_ISL_9164764, EPI_ISL_9164770, EPI_ISL_9164806, EPI_ISL_9164832, EPI_ISL_9164851, EPI_ISL_9164874, EPI_ISL_9164891, EPI_ISL_9164905, EPI_ISL_9164986, EPI_ISL_9164993, EPI_ISL_9165014, EPI_ISL_9165016, EPI_ISL_9165038, EPI_ISL_9165041, EPI_ISL_9165058, EPI_ISL_9165064, EPI_ISL_9165071, EPI_ISL_9165094, EPI_ISL_9165096, EPI_ISL_9165101, EPI_ISL_9165124, EPI_ISL_9165135, EPI_ISL_9165140, EPI_ISL_9165149, EPI_ISL_9165159, EPI_ISL_9165197, EPI_ISL_9165199, EPI_ISL_9165214, EPI_ISL_9165219, EPI_ISL_9165223, EPI_ISL_9165240, EPI_ISL_9165242, EPI_ISL_9165243, EPI_ISL_9165255, EPI_ISL_9165259, EPI_ISL_9165262, EPI_ISL_9165264, EPI_ISL_9165265, EPI_ISL_9165291, EPI_ISL_9165292, EPI_ISL_9165294, EPI_ISL_9165312, EPI_ISL_9165314, EPI_ISL_9165332, EPI_ISL_9165348, EPI_ISL_9165394, EPI_ISL_9165402, EPI_ISL_9165420, EPI_ISL_9165464, EPI_ISL_9165499, EPI_ISL_9165507, EPI_ISL_9165518, EPI_ISL_9165524, EPI_ISL_9165528, EPI_ISL_9165531, EPI_ISL_9165543, EPI_ISL_9165547, EPI_ISL_9165568, EPI_ISL_9165590, EPI_ISL_9165616, EPI_ISL_9165620, EPI_ISL_9165622, EPI_ISL_9165624, EPI_ISL_9165642, EPI_ISL_9165644, EPI_ISL_9165649, EPI_ISL_9165659, EPI_ISL_9165675, EPI_ISL_9165683, EPI_ISL_9165687, EPI_ISL_9165700, EPI_ISL_9165714, EPI_ISL_9165715, EPI_ISL_9165720, EPI_ISL_9165741, EPI_ISL_9165743, EPI_ISL_9165745, EPI_ISL_9165764, EPI_ISL_9165778, EPI_ISL_9165780, EPI_ISL_9165785, EPI_ISL_9165806, EPI_ISL_9165813, EPI_ISL_9165908, EPI_ISL_9165919, EPI_ISL_9165929, EPI_ISL_9165952, EPI_ISL_9165987, EPI_ISL_9165996, EPI_ISL_9166009, EPI_ISL_9166025, EPI_ISL_9166028, EPI_ISL_9166030, EPI_ISL_9166033, EPI_ISL_9166034, EPI_ISL_9166049, EPI_ISL_9166055, EPI_ISL_9166113, EPI_ISL_9166122, EPI_ISL_9166146, EPI_ISL_9166150, EPI_ISL_9166158, EPI_ISL_9166164, EPI_ISL_9166214, EPI_ISL_9166275, EPI_ISL_9166280, EPI_ISL_9166289, EPI_ISL_9166312, EPI_ISL_9166329, EPI_ISL_9166338, EPI_ISL_9166396, EPI_ISL_9166439, EPI_ISL_9166445, EPI_ISL_9166450, EPI_ISL_9166477, EPI_ISL_9166538, EPI_ISL_9166571, EPI_ISL_9166695, EPI_ISL_9166714, EPI_ISL_9166721, EPI_ISL_9166785, EPI_ISL_9166788, EPI_ISL_9166792, EPI_ISL_9166794, EPI_ISL_9166796, EPI_ISL_9166819, EPI_ISL_9166829, EPI_ISL_9166844, EPI_ISL_9166856, EPI_ISL_9166871, EPI_ISL_9166877, EPI_ISL_9166879, EPI_ISL_9166881, EPI_ISL_9166922, EPI_ISL_9166930, EPI_ISL_9166932, EPI_ISL_9166940, EPI_ISL_9166956, EPI_ISL_9166962, EPI_ISL_9166972, EPI_ISL_9167001, EPI_ISL_9167006, EPI_ISL_9167010, EPI_ISL_9167015, EPI_ISL_9167029, EPI_ISL_9167036, EPI_ISL_9167050, EPI_ISL_9167054, EPI_ISL_9167084, EPI_ISL_9167087, EPI_ISL_9167097, EPI_ISL_9167100, EPI_ISL_9167125, EPI_ISL_9167131, EPI_ISL_9167147, EPI_ISL_9167151, EPI_ISL_9167154, EPI_ISL_9167156, EPI_ISL_9167220, EPI_ISL_9167244, EPI_ISL_9167255, EPI_ISL_9167257, EPI_ISL_9167271, EPI_ISL_9167275, EPI_ISL_9167279, EPI_ISL_9167283, EPI_ISL_9167287, EPI_ISL_9167311, EPI_ISL_9167315, EPI_ISL_9167332, EPI_ISL_9167347, EPI_ISL_9167350, EPI_ISL_9167362, EPI_ISL_9167373, EPI_ISL_9167377, EPI_ISL_9167387, EPI_ISL_9167391, EPI_ISL_9167399, EPI_ISL_9167416, EPI_ISL_9167421, EPI_ISL_9167469, EPI_ISL_9167471, EPI_ISL_9167488, EPI_ISL_9167496, EPI_ISL_9167553, EPI_ISL_9167576, EPI_ISL_9167582, EPI_ISL_9167594, EPI_ISL_9167617, EPI_ISL_9167658, EPI_ISL_9167691, EPI_ISL_9167695, EPI_ISL_9167712, EPI_ISL_9167721, EPI_ISL_9167727, EPI_ISL_9167734, EPI_ISL_9167764, EPI_ISL_9167778, EPI_ISL_9167782, EPI_ISL_9167837, EPI_ISL_9167839, EPI_ISL_9167847, EPI_ISL_9167851, EPI_ISL_9167868, EPI_ISL_9167870, EPI_ISL_9167883, EPI_ISL_9167899, EPI_ISL_9167905, EPI_ISL_9167911, EPI_ISL_9167924, EPI_ISL_9167944, EPI_ISL_9167946, EPI_ISL_9167976, EPI_ISL_9168014, EPI_ISL_9168016, EPI_ISL_9168022, EPI_ISL_9168040, EPI_ISL_9168044, EPI_ISL_9168086, EPI_ISL_9168131, EPI_ISL_9168155, EPI_ISL_9168157, EPI_ISL_9168166, EPI_ISL_9168167, EPI_ISL_9168170, EPI_ISL_9168194, EPI_ISL_9168213, EPI_ISL_9168215, EPI_ISL_9168219, EPI_ISL_9168242, EPI_ISL_9168248, EPI_ISL_9168252, EPI_ISL_9168256, EPI_ISL_9168260, EPI_ISL_9168269, EPI_ISL_9168271, EPI_ISL_9168286, EPI_ISL_9168290, EPI_ISL_9168304, EPI_ISL_9168333, EPI_ISL_9168339, EPI_ISL_9168347, EPI_ISL_9168348, EPI_ISL_9168362, EPI_ISL_9168366, EPI_ISL_9168374, EPI_ISL_9168388, EPI_ISL_9168400, EPI_ISL_9168407, EPI_ISL_9168450, EPI_ISL_9168462, EPI_ISL_9168469, EPI_ISL_9168474, EPI_ISL_9168482, EPI_ISL_9168484, EPI_ISL_9168488, EPI_ISL_9168491, EPI_ISL_9168498, EPI_ISL_9168509, EPI_ISL_9168511, EPI_ISL_9168515, EPI_ISL_9168518, EPI_ISL_9168528, EPI_ISL_9168543, EPI_ISL_9168558, EPI_ISL_9168561, EPI_ISL_9168565, EPI_ISL_9168566, EPI_ISL_9168567, EPI_ISL_9168573, EPI_ISL_9168575, EPI_ISL_9168579, EPI_ISL_9168585, EPI_ISL_9168589, EPI_ISL_9168593, EPI_ISL_9168595, EPI_ISL_9168599, EPI_ISL_9168601, EPI_ISL_9168623, EPI_ISL_9168628, EPI_ISL_9168636, EPI_ISL_9168638, EPI_ISL_9168672, EPI_ISL_9168680, EPI_ISL_9168682, EPI_ISL_9168695, EPI_ISL_9168699, EPI_ISL_9168752, EPI_ISL_9168764, EPI_ISL_9168781, EPI_ISL_9168785, EPI_ISL_9168814, EPI_ISL_9168815, EPI_ISL_9168823, EPI_ISL_9168896, EPI_ISL_9168901, EPI_ISL_9168963, EPI_ISL_9168978, EPI_ISL_9168989, EPI_ISL_9168990, EPI_ISL_9168992, EPI_ISL_9169000, EPI_ISL_9169004, EPI_ISL_9169010, EPI_ISL_9169024, EPI_ISL_9169025, EPI_ISL_9169030, EPI_ISL_9169040, EPI_ISL_9169058, EPI_ISL_9169069, EPI_ISL_9169070, EPI_ISL_9169071, EPI_ISL_9169072, EPI_ISL_9169075, EPI_ISL_9169086, EPI_ISL_9169095, EPI_ISL_9169096, EPI_ISL_9169097, EPI_ISL_9169099, EPI_ISL_9169100, EPI_ISL_9169106, EPI_ISL_9169107, EPI_ISL_9169111, EPI_ISL_9169119, EPI_ISL_9169122, EPI_ISL_9169125, EPI_ISL_9169131, EPI_ISL_9169138 | | | |
| see above | Department of Bacteria, Parasites and Fungi, Statens Serum Institut, Copenhagen, Denmark | Statens Serum Institut Bioinformatics and Microbial Genomics | Danish Covid-19 Genome Consortium |

We gratefully acknowledge the following Authors from the Originating laboratories responsible for obtaining the specimens, as well as the Submitting laboratories where the genome data were generated and shared via GISAID, on which this research is based.

All Submitters of data may be contacted directly via www.gisaid.org

Authors are sorted alphabetically.

| Accession ID | Originating Laboratory | Submitting Laboratory | Authors |
|---|---|---|---|
| EPI_ISL_9169142, EPI_ISL_9169197, EPI_ISL_9169200, EPI_ISL_9169210, EPI_ISL_9169224, EPI_ISL_9169235, EPI_ISL_9169236, EPI_ISL_9169242, EPI_ISL_9169249, EPI_ISL_9169254, EPI_ISL_9169255, EPI_ISL_9169263, EPI_ISL_9169270, EPI_ISL_9169276, EPI_ISL_9169281, EPI_ISL_9169291, EPI_ISL_9169302, EPI_ISL_9169310, EPI_ISL_9169322, EPI_ISL_9169324, EPI_ISL_9169326, EPI_ISL_9169327, EPI_ISL_9169333, EPI_ISL_9169336, EPI_ISL_9169358, EPI_ISL_9169366, EPI_ISL_9169370, EPI_ISL_9169388, EPI_ISL_9169391, EPI_ISL_9169397, EPI_ISL_9169504, EPI_ISL_9169505 | | | |
| see above | Department of Bacteria, Parasites and Fungi, Statens Serum Institut, Copenhagen, Denmark | Statens Serum Institut Bioinformatics and Microbial Genomics | Danish Covid-19 Genome Consortium |

We gratefully acknowledge the following Authors from the Originating laboratories responsible for obtaining the specimens, as well as the Submitting laboratories where the genome data were generated and shared via GISAID, on which this research is based.

All Submitters of data may be contacted directly via www.gisaid.org

Authors are sorted alphabetically.

Acknowledgement EPI_SET Identifier: EPI_SET_20220527qy

<table>
<tr><th>Accession ID</th><th>Originating Laboratory</th><th>Submitting Laboratory</th><th>Authors</th></tr>
<tr><td colspan="4">EPI_ISL_9162368, EPI_ISL_9162372, EPI_ISL_9162410, EPI_ISL_9162462, EPI_ISL_9162464, EPI_ISL_9162479, EPI_ISL_9162505, EPI_ISL_9162513, EPI_ISL_9162519, EPI_ISL_9162525, EPI_ISL_9162529, EPI_ISL_9162552, EPI_ISL_9162580, EPI_ISL_9162587, EPI_ISL_9162593, EPI_ISL_9162610, EPI_ISL_9162638, EPI_ISL_9162640, EPI_ISL_9162641, EPI_ISL_9162656, EPI_ISL_9162669, EPI_ISL_9162686, EPI_ISL_9162695, EPI_ISL_9162714, EPI_ISL_9162718, EPI_ISL_9162722, EPI_ISL_9162732, EPI_ISL_9162746, EPI_ISL_9162748, EPI_ISL_9162750, EPI_ISL_9162754, EPI_ISL_9162796, EPI_ISL_9162802, EPI_ISL_9162808, EPI_ISL_9162823, EPI_ISL_9162837, EPI_ISL_9162841, EPI_ISL_9162845, EPI_ISL_9162871, EPI_ISL_9162904, EPI_ISL_9162917, EPI_ISL_9162927, EPI_ISL_9162962, EPI_ISL_9162968, EPI_ISL_9162976, EPI_ISL_9163001, EPI_ISL_9163003, EPI_ISL_9163006, EPI_ISL_9163012, EPI_ISL_9163046, EPI_ISL_9163065, EPI_ISL_9163073, EPI_ISL_9163077, EPI_ISL_9163102, EPI_ISL_9163106, EPI_ISL_9163129, EPI_ISL_9163134, EPI_ISL_9163146, EPI_ISL_9163198, EPI_ISL_9163222, EPI_ISL_9163237, EPI_ISL_9163263, EPI_ISL_9163298, EPI_ISL_9163335, EPI_ISL_9163338, EPI_ISL_9163346, EPI_ISL_9163368, EPI_ISL_9163376, EPI_ISL_9163380, EPI_ISL_9163385, EPI_ISL_9163412, EPI_ISL_9163423, EPI_ISL_9163457, EPI_ISL_9163466, EPI_ISL_9163484, EPI_ISL_9163503, EPI_ISL_9163526, EPI_ISL_9163555, EPI_ISL_9163565, EPI_ISL_9163576, EPI_ISL_9163593, EPI_ISL_9163598, EPI_ISL_9163610, EPI_ISL_9163638, EPI_ISL_9163640, EPI_ISL_9163642, EPI_ISL_9163644, EPI_ISL_9163653, EPI_ISL_9163662, EPI_ISL_9163672, EPI_ISL_9163673, EPI_ISL_9163700, EPI_ISL_9163703, EPI_ISL_9163715, EPI_ISL_9163728, EPI_ISL_9163743, EPI_ISL_9163754, EPI_ISL_9163756, EPI_ISL_9163762, EPI_ISL_9163764, EPI_ISL_9163766, EPI_ISL_9163796, EPI_ISL_9163802, EPI_ISL_9163819, EPI_ISL_9163855, EPI_ISL_9163857, EPI_ISL_9163859, EPI_ISL_9163862, EPI_ISL_9163880, EPI_ISL_9163889, EPI_ISL_9163894, EPI_ISL_9163900, EPI_ISL_9163912, EPI_ISL_9163914, EPI_ISL_9163936, EPI_ISL_9163940, EPI_ISL_9163948, EPI_ISL_9163984, EPI_ISL_9164015, EPI_ISL_9164017, EPI_ISL_9164023, EPI_ISL_9164141, EPI_ISL_9164157, EPI_ISL_9164163, EPI_ISL_9164194, EPI_ISL_9164196, EPI_ISL_9164204, EPI_ISL_9164210, EPI_ISL_9164228, EPI_ISL_9164283, EPI_ISL_9164300, EPI_ISL_9164329, EPI_ISL_9164357, EPI_ISL_9164358, EPI_ISL_9164363, EPI_ISL_9164365, EPI_ISL_9164367, EPI_ISL_9164387, EPI_ISL_9164390, EPI_ISL_9164397, EPI_ISL_9164402, EPI_ISL_9164405, EPI_ISL_9164418, EPI_ISL_9164428, EPI_ISL_9164439, EPI_ISL_9164444, EPI_ISL_9164447, EPI_ISL_9164452, EPI_ISL_9164470, EPI_ISL_9164502, EPI_ISL_9164512, EPI_ISL_9164523, EPI_ISL_9164604, EPI_ISL_9164611, EPI_ISL_9164614, EPI_ISL_9164615, EPI_ISL_9164629, EPI_ISL_9164665, EPI_ISL_9164667, EPI_ISL_9164678, EPI_ISL_9164684, EPI_ISL_9164686, EPI_ISL_9164695, EPI_ISL_9164780, EPI_ISL_9164787, EPI_ISL_9164800, EPI_ISL_9164802, EPI_ISL_9164807, EPI_ISL_9164814, EPI_ISL_9164816, EPI_ISL_9164866, EPI_ISL_9164876, EPI_ISL_9164910, EPI_ISL_9164948, EPI_ISL_9164953, EPI_ISL_9164955, EPI_ISL_9164968, EPI_ISL_9165019, EPI_ISL_9165048, EPI_ISL_9165061, EPI_ISL_9165069, EPI_ISL_9165074, EPI_ISL_9165079, EPI_ISL_9165080, EPI_ISL_9165095, EPI_ISL_9165119, EPI_ISL_9165145, EPI_ISL_9165147, EPI_ISL_9165161, EPI_ISL_9165179, EPI_ISL_9165184, EPI_ISL_9165203, EPI_ISL_9165205, EPI_ISL_9165221, EPI_ISL_9165271, EPI_ISL_9165286, EPI_ISL_9165289, EPI_ISL_9165299, EPI_ISL_9165324, EPI_ISL_9165346, EPI_ISL_9165398, EPI_ISL_9165400, EPI_ISL_9165414, EPI_ISL_9165422, EPI_ISL_9165456, EPI_ISL_9165458, EPI_ISL_9165466, EPI_ISL_9165470, EPI_ISL_9165475, EPI_ISL_9165476, EPI_ISL_9165491, EPI_ISL_9165502, EPI_ISL_9165504, EPI_ISL_9165512, EPI_ISL_9165520, EPI_ISL_9165530, EPI_ISL_9165541, EPI_ISL_9165581, EPI_ISL_9165610, EPI_ISL_9165640, EPI_ISL_9165646, EPI_ISL_9165657, EPI_ISL_9165689, EPI_ISL_9165698, EPI_ISL_9165704, EPI_ISL_9165710, EPI_ISL_9165721, EPI_ISL_9165740, EPI_ISL_9165762, EPI_ISL_9165782, EPI_ISL_9165804, EPI_ISL_9165814, EPI_ISL_9165911, EPI_ISL_9165920, EPI_ISL_9165935, EPI_ISL_9165937, EPI_ISL_9165944, EPI_ISL_9165954, EPI_ISL_9165989, EPI_ISL_9165993, EPI_ISL_9165998, EPI_ISL_9166002, EPI_ISL_9166027, EPI_ISL_9166036, EPI_ISL_9166041, EPI_ISL_9166047, EPI_ISL_9166072, EPI_ISL_9166078, EPI_ISL_9166093, EPI_ISL_9166095, EPI_ISL_9166097, EPI_ISL_9166116, EPI_ISL_9166135, EPI_ISL_9166139, EPI_ISL_9166159, EPI_ISL_9166166, EPI_ISL_9166188, EPI_ISL_9166191, EPI_ISL_9166209, EPI_ISL_9166270, EPI_ISL_9166284, EPI_ISL_9166309, EPI_ISL_9166343, EPI_ISL_9166353, EPI_ISL_9166355, EPI_ISL_9166365, EPI_ISL_9166375, EPI_ISL_9166419, EPI_ISL_9166421, EPI_ISL_9166427, EPI_ISL_9166448, EPI_ISL_9166454, EPI_ISL_9166466, EPI_ISL_9166548, EPI_ISL_9166554, EPI_ISL_9166556, EPI_ISL_9166567, EPI_ISL_9166579, EPI_ISL_9166693, EPI_ISL_9166701, EPI_ISL_9166708, EPI_ISL_9166710, EPI_ISL_9166738, EPI_ISL_9166739, EPI_ISL_9166750, EPI_ISL_9166774, EPI_ISL_9166780, EPI_ISL_9166782, EPI_ISL_9166806, EPI_ISL_9166815, EPI_ISL_9166852, EPI_ISL_9166861, EPI_ISL_9166867, EPI_ISL_9166885, EPI_ISL_9166890, EPI_ISL_9166896, EPI_ISL_9166936, EPI_ISL_9166976, EPI_ISL_9166997, EPI_ISL_9167008, EPI_ISL_9167017, EPI_ISL_9167022, EPI_ISL_9167031, EPI_ISL_9167034, EPI_ISL_9167040, EPI_ISL_9167042, EPI_ISL_9167044, EPI_ISL_9167068, EPI_ISL_9167103, EPI_ISL_9167108, EPI_ISL_9167123, EPI_ISL_9167132, EPI_ISL_9167141, EPI_ISL_9167158, EPI_ISL_9167175, EPI_ISL_9167185, EPI_ISL_9167189, EPI_ISL_9167194, EPI_ISL_9167196, EPI_ISL_9167209, EPI_ISL_9167218, EPI_ISL_9167226, EPI_ISL_9167232, EPI_ISL_9167273, EPI_ISL_9167285, EPI_ISL_9167297, EPI_ISL_9167308, EPI_ISL_9167319, EPI_ISL_9167324, EPI_ISL_9167340, EPI_ISL_9167342, EPI_ISL_9167410, EPI_ISL_9167449, EPI_ISL_9167467, EPI_ISL_9167473, EPI_ISL_9167541, EPI_ISL_9167555, EPI_ISL_9167563, EPI_ISL_9167611, EPI_ISL_9167621, EPI_ISL_9167653, EPI_ISL_9167677, EPI_ISL_9167681, EPI_ISL_9167707, EPI_ISL_9167723, EPI_ISL_9167750, EPI_ISL_9167754, EPI_ISL_9167760, EPI_ISL_9167784, EPI_ISL_9167787, EPI_ISL_9167793, EPI_ISL_9167795, EPI_ISL_9167801, EPI_ISL_9167811, EPI_ISL_9167825, EPI_ISL_9167827, EPI_ISL_9167862, EPI_ISL_9167872, EPI_ISL_9167893, EPI_ISL_9167913, EPI_ISL_9167919, EPI_ISL_9167932, EPI_ISL_9167936, EPI_ISL_9167950, EPI_ISL_9167964, EPI_ISL_9167972, EPI_ISL_9168006, EPI_ISL_9168008, EPI_ISL_9168024, EPI_ISL_9168026, EPI_ISL_9168048, EPI_ISL_9168052, EPI_ISL_9168057, EPI_ISL_9168061, EPI_ISL_9168071, EPI_ISL_9168073, EPI_ISL_9168077, EPI_ISL_9168092, EPI_ISL_9168106, EPI_ISL_9168108, EPI_ISL_9168149, EPI_ISL_9168159, EPI_ISL_9168208, EPI_ISL_9168240, EPI_ISL_9168265, EPI_ISL_9168267, EPI_ISL_9168283, EPI_ISL_9168317, EPI_ISL_9168326, EPI_ISL_9168327, EPI_ISL_9168334, EPI_ISL_9168340, EPI_ISL_9168341, EPI_ISL_9168346, EPI_ISL_9168356, EPI_ISL_9168369, EPI_ISL_9168370, EPI_ISL_9168373, EPI_ISL_9168381, EPI_ISL_9168386, EPI_ISL_9168390, EPI_ISL_9168394, EPI_ISL_9168398, EPI_ISL_9168403, EPI_ISL_9168405, EPI_ISL_9168411, EPI_ISL_9168413, EPI_ISL_9168414, EPI_ISL_9168453, EPI_ISL_9168457, EPI_ISL_9168463, EPI_ISL_9168466, EPI_ISL_9168476, EPI_ISL_9168481, EPI_ISL_9168487, EPI_ISL_9168513, EPI_ISL_9168517, EPI_ISL_9168522, EPI_ISL_9168525, EPI_ISL_9168537, EPI_ISL_9168544, EPI_ISL_9168546, EPI_ISL_9168551, EPI_ISL_9168554, EPI_ISL_9168563, EPI_ISL_9168568, EPI_ISL_9168576, EPI_ISL_9168577, EPI_ISL_9168578, EPI_ISL_9168584, EPI_ISL_9168586, EPI_ISL_9168591, EPI_ISL_9168604, EPI_ISL_9168609, EPI_ISL_9168614, EPI_ISL_9168617, EPI_ISL_9168634, EPI_ISL_9168635, EPI_ISL_9168641, EPI_ISL_9168644, EPI_ISL_9168647, EPI_ISL_9168648, EPI_ISL_9168660, EPI_ISL_9168665, EPI_ISL_9168676, EPI_ISL_9168684, EPI_ISL_9168686, EPI_ISL_9168687, EPI_ISL_9168692, EPI_ISL_9168698, EPI_ISL_9168710, EPI_ISL_9168712, EPI_ISL_9168756, EPI_ISL_9168757, EPI_ISL_9168761, EPI_ISL_9168765, EPI_ISL_9168768, EPI_ISL_9168783, EPI_ISL_9168784, EPI_ISL_9168790, EPI_ISL_9168793, EPI_ISL_9168799, EPI_ISL_9168808, EPI_ISL_9168903, EPI_ISL_9168907, EPI_ISL_9168969, EPI_ISL_9168975, EPI_ISL_9168984, EPI_ISL_9168999, EPI_ISL_9169002, EPI_ISL_9169007, EPI_ISL_9169013, EPI_ISL_9169014, EPI_ISL_9169015, EPI_ISL_9169016, EPI_ISL_9169018, EPI_ISL_9169022, EPI_ISL_9169023, EPI_ISL_9169031, EPI_ISL_9169046, EPI_ISL_9169048, EPI_ISL_9169052, EPI_ISL_9169057, EPI_ISL_9169059, EPI_ISL_9169063, EPI_ISL_9169064, EPI_ISL_9169068, EPI_ISL_9169074</td></tr>
<tr><td>see above</td><td>Department of Bacteria, Parasites and Fungi, Statens Serum Institut, Copenhagen, Denmark</td><td>Statens Serum Institut Bioinformatics and Microbial Genomics</td><td>Danish Covid-19 Genome Consortium</td></tr>
</table>

We gratefully acknowledge the following Authors from the Originating laboratories responsible for obtaining the specimens, as well as the Submitting laboratories where the genome data were generated and shared via GISAID, on which this research is based.

All Submitters of data may be contacted directly via www.gisaid.org

Authors are sorted alphabetically.

| Accession ID | Originating Laboratory | Submitting Laboratory | Authors |
|---|---|---|---|
| EPI_ISL_9169080, EPI_ISL_9169091, EPI_ISL_9169094, EPI_ISL_9169103, EPI_ISL_9169105, EPI_ISL_9169108, EPI_ISL_9169113, EPI_ISL_9169123, EPI_ISL_9169136, EPI_ISL_9169146, EPI_ISL_9169194, EPI_ISL_9169202, EPI_ISL_9169204, EPI_ISL_9169207, EPI_ISL_9169211, EPI_ISL_9169212, EPI_ISL_9169218, EPI_ISL_9169220, EPI_ISL_9169229, EPI_ISL_9169243, EPI_ISL_9169248, EPI_ISL_9169252, EPI_ISL_9169253, EPI_ISL_9169256, EPI_ISL_9169259, EPI_ISL_9169260, EPI_ISL_9169269, EPI_ISL_9169273, EPI_ISL_9169278, EPI_ISL_9169299, EPI_ISL_9169303, EPI_ISL_9169307, EPI_ISL_9169328, EPI_ISL_9169331, EPI_ISL_9169335, EPI_ISL_9169338, EPI_ISL_9169341, EPI_ISL_9169342, EPI_ISL_9169343, EPI_ISL_9169345, EPI_ISL_9169365, EPI_ISL_9169372, EPI_ISL_9169376, EPI_ISL_9169378, EPI_ISL_9169383, EPI_ISL_9169386, EPI_ISL_9169394, EPI_ISL_9169396, EPI_ISL_9169400, EPI_ISL_9169503 | | | |
| see above | Department of Bacteria, Parasites and Fungi, Statens Serum Institut, Copenhagen, Denmark | Statens Serum Institut Bioinformatics and Microbial Genomics | Danish Covid-19 Genome Consortium |

We gratefully acknowledge the following Authors from the Originating laboratories responsible for obtaining the specimens, as well as the Submitting laboratories where the genome data were generated and shared via GISAID, on which this research is based.

All Submitters of data may be contacted directly via www.gisaid.org

Authors are sorted alphabetically.

| Accession ID | Originating Laboratory | Submitting Laboratory | Authors |
| --- | --- | --- | --- |
| EPI_ISL_9162354, EPI_ISL_9162458, EPI_ISL_9162465, EPI_ISL_9162477, EPI_ISL_9162496, EPI_ISL_9162537, EPI_ISL_9162542, EPI_ISL_9162556, EPI_ISL_9162568, EPI_ISL_9162651, EPI_ISL_9162667, EPI_ISL_9162673, EPI_ISL_9162744, EPI_ISL_9162798, EPI_ISL_9162825, EPI_ISL_9162880, EPI_ISL_9162938, EPI_ISL_9162952, EPI_ISL_9162981, EPI_ISL_9163052, EPI_ISL_9163113, EPI_ISL_9163137, EPI_ISL_9163256, EPI_ISL_9163261, EPI_ISL_9163310, EPI_ISL_9163332, EPI_ISL_9163352, EPI_ISL_9163358, EPI_ISL_9163452, EPI_ISL_9163488, EPI_ISL_9163518, EPI_ISL_9163549, EPI_ISL_9163557, EPI_ISL_9163634, EPI_ISL_9163667, EPI_ISL_9163733, EPI_ISL_9163770, EPI_ISL_9163801, EPI_ISL_9163823, EPI_ISL_9163904, EPI_ISL_9164175, EPI_ISL_9164180, EPI_ISL_9164198, EPI_ISL_9164270, EPI_ISL_9164501, EPI_ISL_9164506, EPI_ISL_9164518, EPI_ISL_9164653, EPI_ISL_9164655, EPI_ISL_9164723, EPI_ISL_9164731, EPI_ISL_9164751, EPI_ISL_9164753, EPI_ISL_9164782, EPI_ISL_9164822, EPI_ISL_9164826, EPI_ISL_9164895, EPI_ISL_9164958, EPI_ISL_9164982, EPI_ISL_9165187, EPI_ISL_9165207, EPI_ISL_9165249, EPI_ISL_9165408, EPI_ISL_9165442, EPI_ISL_9165522, EPI_ISL_9165601, EPI_ISL_9165605, EPI_ISL_9165614, EPI_ISL_9165628, EPI_ISL_9165708, EPI_ISL_9165733, EPI_ISL_9165735, EPI_ISL_9165738, EPI_ISL_9165739, EPI_ISL_9165796, EPI_ISL_9165905, EPI_ISL_9165942, EPI_ISL_9165991, EPI_ISL_9166137, EPI_ISL_9166141, EPI_ISL_9166154, EPI_ISL_9166172, EPI_ISL_9166324, EPI_ISL_9166358, EPI_ISL_9166459, EPI_ISL_9166505, EPI_ISL_9166510, EPI_ISL_9166516, EPI_ISL_9166520, EPI_ISL_9166522, EPI_ISL_9166523, EPI_ISL_9166544, EPI_ISL_9166552, EPI_ISL_9166752, EPI_ISL_9166800, EPI_ISL_9166823, EPI_ISL_9166874, EPI_ISL_9166901, EPI_ISL_9166979, EPI_ISL_9167020, EPI_ISL_9167027, EPI_ISL_9167038, EPI_ISL_9167065, EPI_ISL_9167095, EPI_ISL_9167138, EPI_ISL_9167212, EPI_ISL_9167238, EPI_ISL_9167259, EPI_ISL_9167295, EPI_ISL_9167302, EPI_ISL_9167323, EPI_ISL_9167354, EPI_ISL_9167371, EPI_ISL_9167407, EPI_ISL_9167419, EPI_ISL_9167493, EPI_ISL_9167675, EPI_ISL_9167679, EPI_ISL_9167758, EPI_ISL_9167803, EPI_ISL_9167845, EPI_ISL_9167917, EPI_ISL_9168042, EPI_ISL_9168046, EPI_ISL_9168119, EPI_ISL_9168124, EPI_ISL_9168180, EPI_ISL_9168221, EPI_ISL_9168225, EPI_ISL_9168236, EPI_ISL_9168238, EPI_ISL_9168246, EPI_ISL_9168279, EPI_ISL_9168302, EPI_ISL_9168344, EPI_ISL_9168350, EPI_ISL_9168352, EPI_ISL_9168378, EPI_ISL_9168395, EPI_ISL_9168454, EPI_ISL_9168529, EPI_ISL_9168536, EPI_ISL_9168547, EPI_ISL_9168612, EPI_ISL_9168646, EPI_ISL_9168651, EPI_ISL_9168654, EPI_ISL_9168678, EPI_ISL_9168694, EPI_ISL_9168751, EPI_ISL_9168914, EPI_ISL_9168918, EPI_ISL_9168970, EPI_ISL_9169006, EPI_ISL_9169021, EPI_ISL_9169027, EPI_ISL_9169028, EPI_ISL_9169045, EPI_ISL_9169054, EPI_ISL_9169120, EPI_ISL_9169143, EPI_ISL_9169144, EPI_ISL_9169201, EPI_ISL_9169223, EPI_ISL_9169237, EPI_ISL_9169251, EPI_ISL_9169257, EPI_ISL_9169265, EPI_ISL_9169272, EPI_ISL_9169294, EPI_ISL_9169298, EPI_ISL_9169309, EPI_ISL_9169330, EPI_ISL_9169360, EPI_ISL_9169389, EPI_ISL_9169392 | | | |
| see above | Department of Bacteria, Parasites and Fungi, Statens Serum Institut, Copenhagen, Denmark | Statens Serum Institut Bioinformatics and Microbial Genomics | Danish Covid-19 Genome Consortium |

We gratefully acknowledge the following Authors from the Originating laboratories responsible for obtaining the specimens, as well as the Submitting laboratories where the genome data were generated and shared via GISAID, on which this research is based.

All Submitters of data may be contacted directly via www.gisaid.org

Authors are sorted alphabetically.

| Accession ID | Originating Laboratory | Submitting Laboratory | Authors |
|---|---|---|---|
| EPI_ISL_9162416, EPI_ISL_9162430, EPI_ISL_9162489, EPI_ISL_9162521, EPI_ISL_9162544, EPI_ISL_9162596, EPI_ISL_9162705, EPI_ISL_9162709, EPI_ISL_9162742, EPI_ISL_9162839, EPI_ISL_9162851, EPI_ISL_9162863, EPI_ISL_9162867, EPI_ISL_9162923, EPI_ISL_9162960, EPI_ISL_9162964, EPI_ISL_9162983, EPI_ISL_9162985, EPI_ISL_9163050, EPI_ISL_9163081, EPI_ISL_9163173, EPI_ISL_9163243, EPI_ISL_9163313, EPI_ISL_9163435, EPI_ISL_9163459, EPI_ISL_9163542, EPI_ISL_9163582, EPI_ISL_9163614, EPI_ISL_9163629, EPI_ISL_9163648, EPI_ISL_9163650, EPI_ISL_9163670, EPI_ISL_9163705, EPI_ISL_9163717, EPI_ISL_9163719, EPI_ISL_9163752, EPI_ISL_9163777, EPI_ISL_9163783, EPI_ISL_9163827, EPI_ISL_9163878, EPI_ISL_9163950, EPI_ISL_9164006, EPI_ISL_9164254, EPI_ISL_9164285, EPI_ISL_9164301, EPI_ISL_9164305, EPI_ISL_9164392, EPI_ISL_9164398, EPI_ISL_9164434, EPI_ISL_9164637, EPI_ISL_9164659, EPI_ISL_9164706, EPI_ISL_9164709, EPI_ISL_9164715, EPI_ISL_9164719, EPI_ISL_9164737, EPI_ISL_9164745, EPI_ISL_9164786, EPI_ISL_9164789, EPI_ISL_9164864, EPI_ISL_9164901, EPI_ISL_9164922, EPI_ISL_9164963, EPI_ISL_9164991, EPI_ISL_9165001, EPI_ISL_9165009, EPI_ISL_9165129, EPI_ISL_9165130, EPI_ISL_9165166, EPI_ISL_9165193, EPI_ISL_9165201, EPI_ISL_9165236, EPI_ISL_9165267, EPI_ISL_9165278, EPI_ISL_9165303, EPI_ISL_9165308, EPI_ISL_9165406, EPI_ISL_9165430, EPI_ISL_9165432, EPI_ISL_9165434, EPI_ISL_9165446, EPI_ISL_9165473, EPI_ISL_9165562, EPI_ISL_9165572, EPI_ISL_9165681, EPI_ISL_9165694, EPI_ISL_9165723, EPI_ISL_9165853, EPI_ISL_9165903, EPI_ISL_9165913, EPI_ISL_9165917, EPI_ISL_9166007, EPI_ISL_9166010, EPI_ISL_9166143, EPI_ISL_9166178, EPI_ISL_9166231, EPI_ISL_9166282, EPI_ISL_9166370, EPI_ISL_9166389, EPI_ISL_9166495, EPI_ISL_9166535, EPI_ISL_9166616, EPI_ISL_9166746, EPI_ISL_9166803, EPI_ISL_9166808, EPI_ISL_9166846, EPI_ISL_9166863, EPI_ISL_9166869, EPI_ISL_9166883, EPI_ISL_9166887, EPI_ISL_9166902, EPI_ISL_9166963, EPI_ISL_9166989, EPI_ISL_9167048, EPI_ISL_9167086, EPI_ISL_9167090, EPI_ISL_9167152, EPI_ISL_9167205, EPI_ISL_9167214, EPI_ISL_9167281, EPI_ISL_9167396, EPI_ISL_9167461, EPI_ISL_9167504, EPI_ISL_9167567, EPI_ISL_9167578, EPI_ISL_9167629, EPI_ISL_9167641, EPI_ISL_9167647, EPI_ISL_9167663, EPI_ISL_9167725, EPI_ISL_9167736, EPI_ISL_9167774, EPI_ISL_9167829, EPI_ISL_9167833, EPI_ISL_9167942, EPI_ISL_9167958, EPI_ISL_9167968, EPI_ISL_9167993, EPI_ISL_9167997, EPI_ISL_9168035, EPI_ISL_9168038, EPI_ISL_9168055, EPI_ISL_9168081, EPI_ISL_9168200, EPI_ISL_9168365, EPI_ISL_9168478, EPI_ISL_9168538, EPI_ISL_9168611, EPI_ISL_9168655, EPI_ISL_9168658, EPI_ISL_9168664, EPI_ISL_9168666, EPI_ISL_9168673, EPI_ISL_9168685, EPI_ISL_9168704, EPI_ISL_9168748, EPI_ISL_9168753, EPI_ISL_9168758, EPI_ISL_9168792, EPI_ISL_9168797, EPI_ISL_9168805, EPI_ISL_9168813, EPI_ISL_9168894, EPI_ISL_9168902, EPI_ISL_9168915, EPI_ISL_9168973, EPI_ISL_9168977, EPI_ISL_9168993, EPI_ISL_9168995, EPI_ISL_9169008, EPI_ISL_9169041, EPI_ISL_9169042, EPI_ISL_9169090, EPI_ISL_9169117, EPI_ISL_9169118, EPI_ISL_9169137, EPI_ISL_9169147, EPI_ISL_9169208, EPI_ISL_9169215, EPI_ISL_9169250, EPI_ISL_9169261, EPI_ISL_9169267, EPI_ISL_9169293, EPI_ISL_9169325, EPI_ISL_9169329, EPI_ISL_9169347, EPI_ISL_9169355, EPI_ISL_9169380, EPI_ISL_9169382, EPI_ISL_9169387 | | | |
| see above | Department of Bacteria, Parasites and Fungi, Statens Serum Institut, Copenhagen, Denmark | Statens Serum Institut Bioinformatics and Microbial Genomics | Danish Covid-19 Genome Consortium |

We gratefully acknowledge the following Authors from the Originating laboratories responsible for obtaining the specimens, as well as the Submitting laboratories where the genome data were generated and shared via GISAID, on which this research is based.

All Submitters of data may be contacted directly via www.gisaid.org

Authors are sorted alphabetically.

| Accession ID | Originating Laboratory | Submitting Laboratory | Authors |
|---|---|---|---|
| EPI_ISL_9162358, EPI_ISL_9162413, EPI_ISL_9162415, EPI_ISL_9162427, EPI_ISL_9162432, EPI_ISL_9162440, EPI_ISL_9162449, EPI_ISL_9162452, EPI_ISL_9162453, EPI_ISL_9162473, EPI_ISL_9162491, EPI_ISL_9162501, EPI_ISL_9162507, EPI_ISL_9162531, EPI_ISL_9162540, EPI_ISL_9162548, EPI_ISL_9162558, EPI_ISL_9162564, EPI_ISL_9162570, EPI_ISL_9162582, EPI_ISL_9162658, EPI_ISL_9162671, EPI_ISL_9162682, EPI_ISL_9162688, EPI_ISL_9162690, EPI_ISL_9162752, EPI_ISL_9162766, EPI_ISL_9162777, EPI_ISL_9162811, EPI_ISL_9162817, EPI_ISL_9162835, EPI_ISL_9162853, EPI_ISL_9162865, EPI_ISL_9162906, EPI_ISL_9162913, EPI_ISL_9162929, EPI_ISL_9162942, EPI_ISL_9162946, EPI_ISL_9162966, EPI_ISL_9162978, EPI_ISL_9163020, EPI_ISL_9163022, EPI_ISL_9163028, EPI_ISL_9163036, EPI_ISL_9163067, EPI_ISL_9163090, EPI_ISL_9163108, EPI_ISL_9163111, EPI_ISL_9163139, EPI_ISL_9163152, EPI_ISL_9163166, EPI_ISL_9163184, EPI_ISL_9163192, EPI_ISL_9163248, EPI_ISL_9163273, EPI_ISL_9163281, EPI_ISL_9163283, EPI_ISL_9163287, EPI_ISL_9163303, EPI_ISL_9163319, EPI_ISL_9163340, EPI_ISL_9163404, EPI_ISL_9163408, EPI_ISL_9163413, EPI_ISL_9163440, EPI_ISL_9163441, EPI_ISL_9163446, EPI_ISL_9163455, EPI_ISL_9163478, EPI_ISL_9163499, EPI_ISL_9163511, EPI_ISL_9163515, EPI_ISL_9163540, EPI_ISL_9163547, EPI_ISL_9163573, EPI_ISL_9163574, EPI_ISL_9163585, EPI_ISL_9163618, EPI_ISL_9163659, EPI_ISL_9163675, EPI_ISL_9163682, EPI_ISL_9163684, EPI_ISL_9163694, EPI_ISL_9163725, EPI_ISL_9163732, EPI_ISL_9163735, EPI_ISL_9163799, EPI_ISL_9163847, EPI_ISL_9163849, EPI_ISL_9163869, EPI_ISL_9163874, EPI_ISL_9163881, EPI_ISL_9163888, EPI_ISL_9163906, EPI_ISL_9163910, EPI_ISL_9163920, EPI_ISL_9163930, EPI_ISL_9163946, EPI_ISL_9163958, EPI_ISL_9163966, EPI_ISL_9164000, EPI_ISL_9164027, EPI_ISL_9164029, EPI_ISL_9164031, EPI_ISL_9164033, EPI_ISL_9164132, EPI_ISL_9164188, EPI_ISL_9164241, EPI_ISL_9164246, EPI_ISL_9164266, EPI_ISL_9164309, EPI_ISL_9164316, EPI_ISL_9164347, EPI_ISL_9164353, EPI_ISL_9164360, EPI_ISL_9164408, EPI_ISL_9164420, EPI_ISL_9164449, EPI_ISL_9164454, EPI_ISL_9164456, EPI_ISL_9164462, EPI_ISL_9164510, EPI_ISL_9164521, EPI_ISL_9164609, EPI_ISL_9164625, EPI_ISL_9164649, EPI_ISL_9164651, EPI_ISL_9164673, EPI_ISL_9164733, EPI_ISL_9164747, EPI_ISL_9164776, EPI_ISL_9164830, EPI_ISL_9164847, EPI_ISL_9164852, EPI_ISL_9164862, EPI_ISL_9164868, EPI_ISL_9164884, EPI_ISL_9164887, EPI_ISL_9164897, EPI_ISL_9164913, EPI_ISL_9164929, EPI_ISL_9164932, EPI_ISL_9164945, EPI_ISL_9164961, EPI_ISL_9164989, EPI_ISL_9164998, EPI_ISL_9165005, EPI_ISL_9165028, EPI_ISL_9165076, EPI_ISL_9165116, EPI_ISL_9165133, EPI_ISL_9165143, EPI_ISL_9165171, EPI_ISL_9165209, EPI_ISL_9165216, EPI_ISL_9165253, EPI_ISL_9165257, EPI_ISL_9165284, EPI_ISL_9165305, EPI_ISL_9165307, EPI_ISL_9165340, EPI_ISL_9165436, EPI_ISL_9165452, EPI_ISL_9165468, EPI_ISL_9165481, EPI_ISL_9165484, EPI_ISL_9165486, EPI_ISL_9165495, EPI_ISL_9165535, EPI_ISL_9165536, EPI_ISL_9165549, EPI_ISL_9165564, EPI_ISL_9165574, EPI_ISL_9165579, EPI_ISL_9165582, EPI_ISL_9165626, EPI_ISL_9165636, EPI_ISL_9165654, EPI_ISL_9165670, EPI_ISL_9165702, EPI_ISL_9165719, EPI_ISL_9165747, EPI_ISL_9165758, EPI_ISL_9165766, EPI_ISL_9165802, EPI_ISL_9165925, EPI_ISL_9165979, EPI_ISL_9166016, EPI_ISL_9166045, EPI_ISL_9166057, EPI_ISL_9166061, EPI_ISL_9166131, EPI_ISL_9166148, EPI_ISL_9166176, EPI_ISL_9166190, EPI_ISL_9166193, EPI_ISL_9166212, EPI_ISL_9166277, EPI_ISL_9166315, EPI_ISL_9166363, EPI_ISL_9166373, EPI_ISL_9166394, EPI_ISL_9166416, EPI_ISL_9166429, EPI_ISL_9166444, EPI_ISL_9166464, EPI_ISL_9166469, EPI_ISL_9166515, EPI_ISL_9166519, EPI_ISL_9166569, EPI_ISL_9166585, EPI_ISL_9166587, EPI_ISL_9166591, EPI_ISL_9166703, EPI_ISL_9166706, EPI_ISL_9166716, EPI_ISL_9166729, EPI_ISL_9166735, EPI_ISL_9166741, EPI_ISL_9166760, EPI_ISL_9166821, EPI_ISL_9166850, EPI_ISL_9166892, EPI_ISL_9166899, EPI_ISL_9166908, EPI_ISL_9166924, EPI_ISL_9166942, EPI_ISL_9166944, EPI_ISL_9166958, EPI_ISL_9166960, EPI_ISL_9166965, EPI_ISL_9166967, EPI_ISL_9166974, EPI_ISL_9166981, EPI_ISL_9166985, EPI_ISL_9166987, EPI_ISL_9166993, EPI_ISL_9166999, EPI_ISL_9167046, EPI_ISL_9167092, EPI_ISL_9167094, EPI_ISL_9167127, EPI_ISL_9167134, EPI_ISL_9167149, EPI_ISL_9167191, EPI_ISL_9167199, EPI_ISL_9167216, EPI_ISL_9167222, EPI_ISL_9167224, EPI_ISL_9167228, EPI_ISL_9167240, EPI_ISL_9167242, EPI_ISL_9167267, EPI_ISL_9167269, EPI_ISL_9167299, EPI_ISL_9167309, EPI_ISL_9167317, EPI_ISL_9167326, EPI_ISL_9167328, EPI_ISL_9167336, EPI_ISL_9167358, EPI_ISL_9167360, EPI_ISL_9167369, EPI_ISL_9167383, EPI_ISL_9167385, EPI_ISL_9167401, EPI_ISL_9167451, EPI_ISL_9167463, EPI_ISL_9167481, EPI_ISL_9167498, EPI_ISL_9167500, EPI_ISL_9167539, EPI_ISL_9167580, EPI_ISL_9167590, EPI_ISL_9167615, EPI_ISL_9167625, EPI_ISL_9167635, EPI_ISL_9167637, EPI_ISL_9167673, EPI_ISL_9167689, EPI_ISL_9167693, EPI_ISL_9167697, EPI_ISL_9167740, EPI_ISL_9167744, EPI_ISL_9167756, EPI_ISL_9167766, EPI_ISL_9167770, EPI_ISL_9167780, EPI_ISL_9167791, EPI_ISL_9167805, EPI_ISL_9167854, EPI_ISL_9167858, EPI_ISL_9167860, EPI_ISL_9167887, EPI_ISL_9167889, EPI_ISL_9167907, EPI_ISL_9167909, EPI_ISL_9167926, EPI_ISL_9167995, EPI_ISL_9168029, EPI_ISL_9168063, EPI_ISL_9168078, EPI_ISL_9168084, EPI_ISL_9168112, EPI_ISL_9168114, EPI_ISL_9168118, EPI_ISL_9168122, EPI_ISL_9168176, EPI_ISL_9168184, EPI_ISL_9168226, EPI_ISL_9168244, EPI_ISL_9168273, EPI_ISL_9168275, EPI_ISL_9168294, EPI_ISL_9168329, EPI_ISL_9168335, EPI_ISL_9168354, EPI_ISL_9168359, EPI_ISL_9168361, EPI_ISL_9168364, EPI_ISL_9168367, EPI_ISL_9168372, EPI_ISL_9168376, EPI_ISL_9168377, EPI_ISL_9168379, EPI_ISL_9168385, EPI_ISL_9168387, EPI_ISL_9168391, EPI_ISL_9168402, EPI_ISL_9168409, EPI_ISL_9168412, EPI_ISL_9168438, EPI_ISL_9168448, EPI_ISL_9168464, EPI_ISL_9168470, EPI_ISL_9168475, EPI_ISL_9168479, EPI_ISL_9168486, EPI_ISL_9168492, EPI_ISL_9168494, EPI_ISL_9168497, EPI_ISL_9168499, EPI_ISL_9168501, EPI_ISL_9168502, EPI_ISL_9168508, EPI_ISL_9168520, EPI_ISL_9168527, EPI_ISL_9168542, EPI_ISL_9168545, EPI_ISL_9168548, EPI_ISL_9168552, EPI_ISL_9168553, EPI_ISL_9168569, EPI_ISL_9168570, EPI_ISL_9168582, EPI_ISL_9168590, EPI_ISL_9168594, EPI_ISL_9168603, EPI_ISL_9168610, EPI_ISL_9168615, EPI_ISL_9168630, EPI_ISL_9168631, EPI_ISL_9168639, EPI_ISL_9168650, EPI_ISL_9168656, EPI_ISL_9168661, EPI_ISL_9168677, EPI_ISL_9168681, EPI_ISL_9168683, EPI_ISL_9168691, EPI_ISL_9168696, EPI_ISL_9168697, EPI_ISL_9168702, EPI_ISL_9168705, EPI_ISL_9168749, EPI_ISL_9168766, EPI_ISL_9168770, EPI_ISL_9168775, EPI_ISL_9168791, EPI_ISL_9168795, EPI_ISL_9168900, EPI_ISL_9168909, EPI_ISL_9168910, EPI_ISL_9168911, EPI_ISL_9168916, EPI_ISL_9168917, EPI_ISL_9168986, EPI_ISL_9168987, EPI_ISL_9168994, EPI_ISL_9168996, EPI_ISL_9169003, EPI_ISL_9169019, EPI_ISL_9169020, EPI_ISL_9169026, EPI_ISL_9169034, EPI_ISL_9169035, EPI_ISL_9169039, EPI_ISL_9169044, EPI_ISL_9169050, EPI_ISL_9169051, EPI_ISL_9169053, EPI_ISL_9169055, EPI_ISL_9169060, EPI_ISL_9169065, EPI_ISL_9169085, EPI_ISL_9169087, EPI_ISL_9169092, EPI_ISL_9169102, EPI_ISL_9169104, EPI_ISL_9169139, EPI_ISL_9169149, EPI_ISL_9169196, EPI_ISL_9169203, EPI_ISL_9169213, EPI_ISL_9169219, EPI_ISL_9169227, EPI_ISL_9169228, EPI_ISL_9169234, EPI_ISL_9169240, EPI_ISL_9169241, EPI_ISL_9169258, EPI_ISL_9169280, EPI_ISL_9169283, EPI_ISL_9169285, EPI_ISL_9169286, EPI_ISL_9169287, EPI_ISL_9169289, EPI_ISL_9169292, EPI_ISL_9169296, EPI_ISL_9169312, EPI_ISL_9169315, EPI_ISL_9169319, EPI_ISL_9169320, EPI_ISL_9169349, EPI_ISL_9169351, EPI_ISL_9169361, EPI_ISL_9169362, EPI_ISL_9169368, EPI_ISL_9169374, EPI_ISL_9169375, EPI_ISL_9169385, EPI_ISL_9169399, EPI_ISL_9169500 | | | |
| see above | Department of Bacteria, Parasites and Fungi, Statens Serum Institut, Copenhagen, Denmark | Statens Serum Institut Bioinformatics and Microbial Genomics | Danish Covid-19 Genome Consortium |

We gratefully acknowledge the following Authors from the Originating laboratories responsible for obtaining the specimens, as well as the Submitting laboratories where the genome data were generated and shared via GISAID, on which this research is based.

All Submitters of data may be contacted directly via www.gisaid.org

Authors are sorted alphabetically.

Acknowledgement EPI_SET Identifier: EPI_SET_20220527ts

| Accession ID | Originating Laboratory | Submitting Laboratory | Authors |
|---|---|---|---|
| EPI_ISL_9186382, EPI_ISL_9186384, EPI_ISL_9186385, EPI_ISL_9186412, EPI_ISL_9186447 | A. Krumbholz, Labor Dr. Krause und Kollegen MVZ GmbH, Kiel | Charité Universitätsmedizin Berlin, Institut für Virologie | Andi Krumbholz; Barbara Mühlemann; Christian Drosten; Julia Schneider; Julia Tesch; Jörn Beheim-Schwarzbach; Talitha Veith; Terry Jones; Tobias Bleicker; Victor M Corman |

We gratefully acknowledge the following Authors from the Originating laboratories responsible for obtaining the specimens, as well as the Submitting laboratories where the genome data were generated and shared via GISAID, on which this research is based.

All Submitters of data may be contacted directly via www.gisaid.org

Authors are sorted alphabetically.

| Accession ID | Originating Laboratory | Submitting Laboratory | Authors |
| --- | --- | --- | --- |
| EPI_ISL_9171918, EPI_ISL_9171922 | Azienda Sanitaria Locale di Piacenza - Presidio Ospedaliero - Laboratorio di Microbiologia | Istituto Zooprofilattico Sperimentale della Lombardia e dell'Emilia Romagna (IZSLER), Risk Analysis and Genomic Epidemiology Unit | Erika Scaltriti; Ilaria Menozzi; Lo Cascio Giuliana; Marina Morganti; Peroni Irene; Schiavo Roberta; Stefano Pongolini |

We gratefully acknowledge the following Authors from the Originating laboratories responsible for obtaining the specimens, as well as the Submitting laboratories where the genome data were generated and shared via GISAID, on which this research is based.

All Submitters of data may be contacted directly via www.gisaid.org

Authors are sorted alphabetically.

| Accession ID | Originating Laboratory | Submitting Laboratory | Authors |
|---|---|---|---|
| EPI_ISL_9181711, EPI_ISL_9181713 | Laboratoire national de sante, Microbiology, Virology | Laboratoire national de sante, Microbiology, Microbial Genomics Platform | Anke Wienecke-Baldacchino; Catherine Ragimbeau; Elodie Solarino; Fatu Djabi; Jessica Tapp; Lise Pignon; Raoul Salmon; Sibel Berger; Tamir Abdelrahman; Trung Nguyen Nguyen; Virginie Jover |

We gratefully acknowledge the following Authors from the Originating laboratories responsible for obtaining the specimens, as well as the Submitting laboratories where the genome data were generated and shared via GISAID, on which this research is based.

All Submitters of data may be contacted directly via www.gisaid.org

Authors are sorted alphabetically.

| Accession ID | Originating Laboratory | Submitting Laboratory | Authors |
|---|---|---|---|
| EPI_ISL_9154325 | Medyczne Laboratorium Diagnostyczne Medrex | 1. Academic Center for Pathomorphological and Genetic-Molecular Diagnostics ltd, Bialystok, Poland 2. National Institute of Public Health - National Institute of Hygiene, Warsaw, Poland | Anetta Sulewska; Jacek Nikliński; Janusz Dzięcioł; Joanna Kiśluk; Katarzyna Zacharczuk; Konrad Raczkowski; Magdalena Nowakowska; Małgorzata Sadkowska-Todys; Piotr Karabowicz; Piotr Majewski; Przemysław Biecek. Joanna Reszeć; Radosław Charkiewicz; Tomasz Wołkowicz |
| EPI_ISL_9154334, EPI_ISL_9154349 | Szpital Wojewódzki im. Kardynała Stefana Wyszyńskiego w Łomży | 1. Academic Center for Pathomorphological and Genetic-Molecular Diagnostics ltd, Bialystok, Poland 2. National Institute of Public Health - National Institute of Hygiene, Warsaw, Poland | Anetta Sulewska; Jacek Nikliński; Janusz Dzięcioł; Joanna Kiśluk; Katarzyna Zacharczuk; Konrad Raczkowski; Magdalena Nowakowska; Małgorzata Sadkowska-Todys; Piotr Karabowicz; Piotr Majewski; Przemysław Biecek. Joanna Reszeć; Radosław Charkiewicz; Tomasz Wołkowicz |
| EPI_ISL_9154644 | Vitomed Sp. z o.o. | 1. Academic Center for Pathomorphological and Genetic-Molecular Diagnostics ltd, Bialystok, Poland 2. National Institute of Public Health - National Institute of Hygiene, Warsaw, Poland | Anetta Sulewska; Jacek Nikliński; Janusz Dzięcioł; Joanna Kiśluk; Katarzyna Zacharczuk; Konrad Raczkowski; Magdalena Nowakowska; Małgorzata Sadkowska-Todys; Piotr Karabowicz; Piotr Majewski; Przemysław Biecek. Joanna Reszeć; Radosław Charkiewicz; Tomasz Wołkowicz |

We gratefully acknowledge the following Authors from the Originating laboratories responsible for obtaining the specimens, as well as the Submitting laboratories where the genome data were generated and shared via GISAID, on which this research is based.

All Submitters of data may be contacted directly via www.gisaid.org

Authors are sorted alphabetically.

| Accession ID | Originating Laboratory | Submitting Laboratory | Authors |
|---|---|---|---|
| EPI_ISL_9156927 | Center for Laboratory Medicine | Center for Laboratory Medicine, ZLMSG | Yannick Gerth |

We gratefully acknowledge the following Authors from the Originating laboratories responsible for obtaining the specimens, as well as the Submitting laboratories where the genome data were generated and shared via GISAID, on which this research is based.

All Submitters of data may be contacted directly via www.gisaid.org

Authors are sorted alphabetically.

| Accession ID | Originating Laboratory | Submitting Laboratory | Authors |
| --- | --- | --- | --- |
| EPI_ISL_9163539 | Originating lab: Wales Specialist Virology Centre Sequencing lab: Pathogen Genomics Unit | Public Health Wales Microbiology Cardiff Wales Specialist Virology Centre | Alec Birchley; Alexander Adams; Amy Gaskin; Angela Marchbank; Bree Gatica-Wilcox; Catherine Moore; Jason Coombes; Joanne Watkins; Joel Southgate; Johnathan Evans; Laura Gifford; Lauren Gilbert; Lee Graham; Malorie Perry; Matthew Bull; Nicole Pacchiarini; Sally Corden; Sara Kumziene-Summerhayes; Sara Rey; Sarah Taylor; Simon Cottrell; Sophie Jones; Tom Connor |
| EPI_ISL_9159155, EPI_ISL_9159156, EPI_ISL_9159159, EPI_ISL_9159160, EPI_ISL_9159164, EPI_ISL_9159166, EPI_ISL_9159170, EPI_ISL_9159171, EPI_ISL_9159177, EPI_ISL_9159178, EPI_ISL_9159184, EPI_ISL_9159190, EPI_ISL_9159195, EPI_ISL_9159197, EPI_ISL_9159199, EPI_ISL_9159200, EPI_ISL_9159203, EPI_ISL_9159204, EPI_ISL_9159207, EPI_ISL_9159208, EPI_ISL_9159210, EPI_ISL_9159216, EPI_ISL_9159238, EPI_ISL_9159240, EPI_ISL_9159243, EPI_ISL_9159246, EPI_ISL_9159249, EPI_ISL_9159251, EPI_ISL_9159252, EPI_ISL_9159253, EPI_ISL_9159256, EPI_ISL_9159260, EPI_ISL_9159261, EPI_ISL_9159263, EPI_ISL_9159264, EPI_ISL_9159270, EPI_ISL_9159271, EPI_ISL_9159272, EPI_ISL_9159297, EPI_ISL_9159299, EPI_ISL_9159304, EPI_ISL_9159336, EPI_ISL_9159341, EPI_ISL_9159344, EPI_ISL_9159346, EPI_ISL_9159351, EPI_ISL_9159409, EPI_ISL_9159418, EPI_ISL_9159423, EPI_ISL_9159432, EPI_ISL_9159439, EPI_ISL_9159466, EPI_ISL_9159467, EPI_ISL_9159468, EPI_ISL_9159471, EPI_ISL_9159473, EPI_ISL_9159475, EPI_ISL_9159478, EPI_ISL_9159479, EPI_ISL_9159481, EPI_ISL_9159484, EPI_ISL_9159518, EPI_ISL_9159519, EPI_ISL_9159538, EPI_ISL_9159570, EPI_ISL_9160198, EPI_ISL_9160202, EPI_ISL_9160204, EPI_ISL_9160207, EPI_ISL_9160261, EPI_ISL_9160265, EPI_ISL_9160266, EPI_ISL_9160268, EPI_ISL_9160273, EPI_ISL_9160274, EPI_ISL_9160278, EPI_ISL_9160281, EPI_ISL_9160282, EPI_ISL_9160283, EPI_ISL_9160313, EPI_ISL_9160316, EPI_ISL_9160322, EPI_ISL_9160323, EPI_ISL_9160326, EPI_ISL_9160329, EPI_ISL_9160330, EPI_ISL_9160333, EPI_ISL_9160337, EPI_ISL_9160564, EPI_ISL_9160637, EPI_ISL_9160638, EPI_ISL_9160639, EPI_ISL_9160649, EPI_ISL_9160697 | | | |
| see above | Respiratory Virus Unit, Microbiology Services Colindale, Public Health England | COVID-19 Genomics UK (COG-UK) Consortium | PHE Covid Sequencing Team |

We gratefully acknowledge the following Authors from the Originating laboratories responsible for obtaining the specimens, as well as the Submitting laboratories where the genome data were generated and shared via GISAID, on which this research is based.

All Submitters of data may be contacted directly via www.gisaid.org

Authors are sorted alphabetically.

Acknowledgement EPI_SET Identifier: EPI_SET_20220527my

| Accession ID | Originating Laboratory | Submitting Laboratory | Authors |
|---|---|---|---|
| EPI_ISL_9201580 | VERMAAK | National Institute for Communicable Diseases of the National Health Laboratory Service | Amoako DG; Bhiman JN; Everatt J; Ismail A; Kekana D; Mahlangu B; Mnguni A; Mohale T; Ntuli N; Scheepers C; Wolter N |

We gratefully acknowledge the following Authors from the Originating laboratories responsible for obtaining the specimens, as well as the Submitting laboratories where the genome data were generated and shared via GISAID, on which this research is based.

All Submitters of data may be contacted directly via www.gisaid.org

Authors are sorted alphabetically.

| Accession ID | Originating Laboratory | Submitting Laboratory | Authors |
|---|---|---|---|
| EPI_ISL_9232037, EPI_ISL_9232038, EPI_ISL_9232039, EPI_ISL_9232044, EPI_ISL_9232045, EPI_ISL_9232046, EPI_ISL_9232048, EPI_ISL_9232060, EPI_ISL_9232086, EPI_ISL_9232088, EPI_ISL_9232089, EPI_ISL_9232090, EPI_ISL_9232091, EPI_ISL_9232097, EPI_ISL_9232102, EPI_ISL_9232108, EPI_ISL_9232130, EPI_ISL_9232136, EPI_ISL_9232146, EPI_ISL_9232154, EPI_ISL_9232155, EPI_ISL_9232156, EPI_ISL_9232160, EPI_ISL_9232161, EPI_ISL_9232163, EPI_ISL_9232168, EPI_ISL_9232178, EPI_ISL_9232179, EPI_ISL_9232256, EPI_ISL_9232266, EPI_ISL_9232278, EPI_ISL_9232279, EPI_ISL_9232308 | | | |
| see above | AP SSO | CSIR-Centre for Cellular and Molecular Biology-INSACOG | Amareshwar Vodapalli; Ara Sreenivas; Archana Bharadwaj Siva; B Himasri; Divya Tej Sowpati; Jandhyala Sai Krishna; Karthik Bharadwaj Tallapaka; Lamuk Zaveri; Malini Nemalikanti; Priya Nurkurthy; Rakesh K Mishra; Shreekant Verma; Sreelekshmi MS; Sumedha Avadhanula; Surabhi Srivastava; Tulasi Nagabandi; Valli Nagalakshmi Undamatla; Vidhyadhari Methuku |
| EPI_ISL_9236277 | BJ Medical College, Ahmedabad | Gujarat Biotechnology Research Centre | Akhilesh Modi; Apurvasinh Puvar; Bhadreshsinh Gohil; Chaitanya Joshi; Disha Vora; Janvi Raval; Jaykumar Rangani; Madhvi Joshi; Nimesh Patel; Nitin Savaliya; NitinShukla; Priyank Chavda; Ramesh Pandit; Roshani Mishra; Sonal Sharma; Sumita Soni; Tasnim Trivedi; Zarna Patel |
| EPI_ISL_9232398 | Continental Hospital | CSIR-Centre for Cellular and Molecular Biology | Amareshwar Vodapalli; Ara Sreenivas; Archana Bharadwaj Siva; B Himasri; Divya Tej Sowpati; Jandhyala Sai Krishna; Karthik Bharadwaj Tallapaka; Lamuk Zaveri; Malini Nemalikanti; Priya Nurkurthy; Rakesh K Mishra; Shreekant Verma; Sreelekshmi MS; Sumedha Avadhanula; Surabhi Srivastava; Tulasi Nagabandi; Valli Nagalakshmi Undamatla; Vidhyadhari Methuku |
| EPI_ISL_9233793 | Fukushima Prefectural Institute of Public Health | Fukushima Prefectural Institute of Public Health | Kadzuhiro Kitakawa; Kazunori Suzuki; Kentaro Itokawa; Makoto Kuroda; Masanori Hashino; Naoko Kashiwabara; Rie Suzuki; Rina Tanaka; Shohei Fujita; Tsuyoshi Sekizuka |
| EPI_ISL_9234081, EPI_ISL_9234082 | GMERS Medical College, Valsad | Gujarat Biotechnology Research Centre | Akhilesh Modi; Apurvasinh Puvar; Bhadreshsinh Gohil; Chaitanya Joshi; Disha Vora; Janvi Raval; Jaykumar Rangani; Madhvi Joshi; Nimesh Patel; Nitin Savaliya; NitinShukla; Priyank Chavda; Ramesh Pandit; Roshani Mishra; Sonal Sharma; Tasnim Trivedi; Vicky Gandhi; Zarna Patel |
| EPI_ISL_9236268 | General Hospital, Lunavada, Mahisagar | Gujarat Biotechnology Research Centre | Akhilesh Modi; Apurvasinh Puvar; Bhadreshsinh Gohil; C.K.Solanki; Chaitanya Joshi; Disha Vora; Janvi Raval; Jaykumar Rangani; Madhvi Joshi; Nimesh Patel; Nitin Savaliya; NitinShukla; Priyank Chavda; Ramesh Pandit; Roshani Mishra; Sonal Sharma; Tasnim Trivedi; Zarna Patel |
| EPI_ISL_9234083 | Green Cross Genetics Lab Pvt Ltd, Ahmedabad | Gujarat Biotechnology Research Centre | Akhilesh Modi; Apurvasinh Puvar; Bhadreshsinh Gohil; Chaitanya Joshi; Disha Vora; Janvi Raval; Jaykumar Rangani; Madhvi Joshi; Nimesh Patel; Nitin Savaliya; NitinShukla; Priyank Chavda; Ramesh Pandit; Roshani Mishra; Sonal Sharma; Tasnim Trivedi; Umang Patel; Zarna Patel |
| EPI_ISL_9236266 | Manglam In Vitro Laboratory, Rajkot | Gujarat Biotechnology Research Centre | Akhilesh Modi; Apurvasinh Puvar; Bhadreshsinh Gohil; Chaitanya Joshi; Disha Vora; Janvi Raval; Jaykumar Rangani; Madhvi Joshi; Nimesh Patel; Nitin Savaliya; NitinShukla; Priyank Chavda; Rahul Thumbar; Ramesh Pandit; Roshani Mishra; Sonal Sharma; Tasnim Trivedi; Zarna Patel |
| EPI_ISL_9232081, EPI_ISL_9232100, EPI_ISL_9232101, EPI_ISL_9232105, EPI_ISL_9232106, EPI_ISL_9232126, EPI_ISL_9232128, EPI_ISL_9232141, EPI_ISL_9232184, EPI_ISL_9232185, EPI_ISL_9232186, EPI_ISL_9232231, EPI_ISL_9232232, EPI_ISL_9232285 | | | |
| see above | MapMyGenome | CSIR-Centre for Cellular and Molecular Biology-INSACOG | Amareshwar Vodapalli; Anuradha Acharya; Ara Sreenivas; Archana Bharadwaj Siva; B Himasri; Divya Tej Sowpati; Jandhyala Sai Krishna; Karthik Bharadwaj Tallapaka; Lamuk Zaveri; M. Aravind Kumar; Malini Nemalikanti; Priya Nurkurthy; Rakesh K Mishra; S.P.S. Satish; Sandhya Kiran; Shreekant Verma; Sreelekshmi MS; Sumedha Avadhanula; Surabhi Srivastava; Tulasi Nagabandi; Valli Nagalakshmi Undamatla; Vidhyadhari Methuku |
| EPI_ISL_9219357 | Mie Prefecture Health and Environment Research Institute | Mie Prefecture Health and Environment Research Institute | Kentaro Itokawa; Makoto Kuroda; Masanori Hashino; Rina Tanaka; Takuya Yano; Tsuyoshi Sekizuka |
| EPI_ISL_9234084, EPI_ISL_9234085 | Molecular lab, Shree Krishna Hospital, Karamsad, Anand | Gujarat Biotechnology Research Centre | Akhilesh Modi; Apurvasinh Puvar; Bhadreshsinh Gohil; Chaitanya Joshi; Chirag; Disha Vora; Janvi Raval; Jaykumar Rangani; Madhvi Joshi; Mustafa; Nimesh Patel; Nitin Savaliya; NitinShukla; Priyank Chavda; Ramesh Pandit; Roshani Mishra; Sonal Sharma; Tasnim Trivedi; Zarna Patel |
| EPI_ISL_9248707 | Molecular unit, Clinical Pathology Laboratory, Medical correctional Institution | Center for Medical Genomics Ramathibodi Hospital | Anek mungaomklang; Insee Sensorn; Inthuorn Ratchatasetthakul; Kanyarat Janthima; Parsakorn Tapaopong; Poramate Jiaranai; Vichapon Tiacharoen; Wasun Chantratita |
| EPI_ISL_9233081, EPI_ISL_9233082, EPI_ISL_9233083, EPI_ISL_9233084, EPI_ISL_9233085, EPI_ISL_9233086, EPI_ISL_9233087, EPI_ISL_9233088, EPI_ISL_9233090, EPI_ISL_9233091, EPI_ISL_9233092, EPI_ISL_9233093, EPI_ISL_9233094, EPI_ISL_9233095, EPI_ISL_9233096, EPI_ISL_9233097, EPI_ISL_9233098, EPI_ISL_9233099, EPI_ISL_9233100, EPI_ISL_9233101, EPI_ISL_9233102, EPI_ISL_9233103, EPI_ISL_9233104, EPI_ISL_9233105, EPI_ISL_9233106, EPI_ISL_9233107, EPI_ISL_9233108, EPI_ISL_9233109, EPI_ISL_9233110, EPI_ISL_9233111, EPI_ISL_9233113, EPI_ISL_9233114, EPI_ISL_9233115, EPI_ISL_9233116, EPI_ISL_9233117, EPI_ISL_9233118, EPI_ISL_9233119, EPI_ISL_9233120, EPI_ISL_9233121, EPI_ISL_9233122, EPI_ISL_9233123, EPI_ISL_9233124, EPI_ISL_9233125, EPI_ISL_9233126, EPI_ISL_9233127, EPI_ISL_9233128, EPI_ISL_9233129, EPI_ISL_9233130, EPI_ISL_9233131, EPI_ISL_9233132, EPI_ISL_9233133, EPI_ISL_9233134, EPI_ISL_9233135, EPI_ISL_9233136, EPI_ISL_9233137, EPI_ISL_9233138, EPI_ISL_9233139, EPI_ISL_9233140, EPI_ISL_9233141, EPI_ISL_9233142, EPI_ISL_9233143, EPI_ISL_9233144, EPI_ISL_9233145, EPI_ISL_9233146, EPI_ISL_9233147, EPI_ISL_9233148, EPI_ISL_9233149, EPI_ISL_9233150, EPI_ISL_9233151, EPI_ISL_9233152, EPI_ISL_9233153, EPI_ISL_9233154, EPI_ISL_9233156, EPI_ISL_9233157, EPI_ISL_9233158, EPI_ISL_9233159, EPI_ISL_9233160, EPI_ISL_9233161, EPI_ISL_9233162, EPI_ISL_9233163, EPI_ISL_9233164, EPI_ISL_9233165, EPI_ISL_9233166, EPI_ISL_9233167, EPI_ISL_9233168, EPI_ISL_9233169, EPI_ISL_9233170, EPI_ISL_9233171, EPI_ISL_9233172, EPI_ISL_9233173, EPI_ISL_9233174, EPI_ISL_9233175, EPI_ISL_9233176 | | | |
| see above | National Public Health Laboratory, National Centre for Infectious Diseases | National Public Health Laboratory, National Centre for Infectious Diseases | Benny Yeo; Grace Ngan; Lin Cui; Raymond Tzer Pin Lin; Royce Ang; Samuel Loo; Yichen Ding; Zhenyang Zhou |
| EPI_ISL_9236270 | Nidan Path Lab, Gandhinagar | Gujarat Biotechnology Research Centre | Akhilesh Modi; Apurvasinh Puvar; Bhadreshsinh Gohil; Chaitanya Joshi; Disha Vora; Himani Patel; Janvi Raval; Jaykumar Rangani; Madhvi Joshi; Nimesh Patel; Nitin Savaliya; NitinShukla; Priyank Chavda; Ramesh Pandit; Roshani Mishra; Sonal Sharma; Tasnim Trivedi; Zarna Patel |
| EPI_ISL_9236271 | Northstar Pathology Laboratory, Ahmedabad | Gujarat Biotechnology Research Centre | Akhilesh Modi; Apurvasinh Puvar; Bhadreshsinh Gohil; Chaitanya Joshi; Disha Vora; Janvi Raval; Jaykumar Rangani; Kanupriya; Madhvi Joshi; Nimesh Patel; Nitin Savaliya; NitinShukla; Priyank Chavda; Ramesh Pandit; Roshani Mishra; Sonal Sharma; Tasnim Trivedi; Zarna Patel |
| EPI_ISL_9236267 | Pathocare Pathology Laboratory, Vadodara | Gujarat Biotechnology Research Centre | Akhilesh Modi; Apurvasinh Puvar; Bhadreshsinh Gohil; Chaitanya Joshi; Disha Vora; Janvi Raval; Jaykumar Rangani; Madhvi Joshi; Nimesh Patel; Nitin Savaliya; NitinShukla; Priyank Chavda; Ramesh Pandit; Roshani Mishra; Sonal Sharma; Tasnim Trivedi; Viral Patel; Zarna Patel |
| EPI_ISL_9232416, EPI_ISL_9232419, EPI_ISL_9232422, EPI_ISL_9232426, EPI_ISL_9232428, EPI_ISL_9232429, EPI_ISL_9232435, EPI_ISL_9232437, EPI_ISL_9232447, EPI_ISL_9232448, EPI_ISL_9232449, EPI_ISL_9232454, EPI_ISL_9232455, EPI_ISL_9232457, EPI_ISL_9232459, EPI_ISL_9232464 | | | |
| see above | Rainbow Hospital | CSIR-Centre for Cellular and Molecular Biology | Amareshwar Vodapalli; Ara Sreenivas; Archana Bharadwaj Siva; B Himasri; Divya Tej Sowpati; Jandhyala Sai Krishna; Kalyan Ram Uppaluri; Karthik Bharadwaj Tallapaka; Lamuk Zaveri; Malini Nemalikanti; Priya Nurkurthy; Rakesh K Mishra; Shreekant Verma; Smita Juvvadi; Sreelekshmi MS; Sricharan Devineni; Sumedha Avadhanula; Surabhi Srivastava; Tulasi Nagabandi; Valli Nagalakshmi Undamatla; Vidhyadhari Methuku |
| EPI_ISL_9248700, EPI_ISL_9248701 | Ramathibodi Hospital | Center for Medical Genomics Ramathibodi Hospital | Anek mungaomklang; Insee Sensorn; Inthuorn Ratchatasetthakul; Kanyarat Janthima; Parsakorn Tapaopong; Poramate Jiaranai; Vichapon Tiacharoen; Wasun Chantratita |
| EPI_ISL_9236274 | SSG Hospital Vadodara | Gujarat Biotechnology Research Centre | Akhilesh Modi; Apurvasinh Puvar; Bhadreshsinh Gohil; Chaitanya Joshi; Disha Vora; Janvi Raval; Jaykumar Rangani; Jigna Karia; Madhvi Joshi; Nimesh Patel; Nitin Savaliya; NitinShukla; Priyank Chavda; Ramesh Pandit; Roshani Mishra; Sonal Sharma; Tasnim Trivedi; Zarna Patel |
| EPI_ISL_9234086 | Supratech Micropath Diagnostics & Research laboratory Ahmedabad | Gujarat Biotechnology Research Centre | Akhilesh Modi; Amee Shukla; Apurvasinh Puvar; Bhadreshsinh Gohil; Chaitanya Joshi; Disha Vora; Janvi Raval; Jaykumar Rangani; Madhvi Joshi; Nimesh Patel; Nitin Savaliya; NitinShukla; Priyank Chavda; Ramesh Pandit; Roshani Mishra; Sonal Sharma; Tasnim Trivedi; Zarna Patel |
| EPI_ISL_9236272, EPI_ISL_9236273, EPI_ISL_9236276 | Symmers Pathocare Laboratory -Ahmedabad | Gujarat Biotechnology Research Centre | Akhilesh Modi; Apurvasinh Puvar; Bhadreshsinh Gohil; Chaitanya Joshi; Disha Vora; Janvi Raval; Jaykumar Rangani; Madhvi Joshi; Nimesh Patel; Nitin Savaliya; NitinShukla; Priyank Chavda; Ramesh Pandit; Roshani Mishra; Sonal Sharma; Tasnim Trivedi; Umang Gandhi; Zarna Patel |
| EPI_ISL_9234080 | Universal Pathology & Molocular Lab, New Ranip, Ahmedabad | Gujarat Biotechnology Research Centre | Akhilesh Modi; Apurvasinh Puvar; Bhadreshsinh Gohil; Bipin Patel; Chaitanya Joshi; Disha Vora; Janvi Raval; Jaykumar Rangani; Madhvi Joshi; Nimesh Patel; Nitin Savaliya; NitinShukla; Priyank Chavda; Ramesh Pandit; Roshani Mishra; Sonal Sharma; Tasnim Trivedi; Zarna Patel |
| EPI_ISL_9236269 | Zydus Hospital & Healthcare Research Pvt. Ltd. | Gujarat Biotechnology Research Centre | Akhilesh Modi; Apurvasinh Puvar; Bhadreshsinh Gohil; Chaitanya Joshi; Disha Vora; Janvi Raval; Jaykumar Rangani; Jaymin Patel; Madhvi Joshi; Nimesh Patel; Nitin Savaliya; NitinShukla; Priyank Chavda; Ramesh Pandit; Roshani Mishra; Sonal Sharma; Tasnim Trivedi; Zarna Patel |

We gratefully acknowledge the following Authors from the Originating laboratories responsible for obtaining the specimens, as well as the Submitting laboratories where the genome data were generated and shared via GISAID, on which this research is based.

All Submitters of data may be contacted directly via www.gisaid.org

Authors are sorted alphabetically.

| Accession ID | Originating Laboratory | Submitting Laboratory | Authors |
| --- | --- | --- | --- |
| EPI_ISL_9252474 | CHU UCL Namur, Avenue G. Thérasse 1, 5530 Yvoir, Belgium | Plateforme de testing Namuroise | Degosserie Jonathan; Demars Aurore; Denis Olivier; Drugmand Jonathan; Maschietto Céline; Mullier François; Nobis Chloé; Otto Gaetan |
| EPI_ISL_9246983 | Department of Clinical Microbiology | GIGA Medical Genomics | Claire Gourzonès; Cécile Meex; Keith Durkin; Laurent Gillet; Maria Artesi; Marie-Pierre Hayette; Nadine Cambisano; Nathalie Renotte; Olivier Ek; Sébastien Bontems; Vincent Bours |

We gratefully acknowledge the following Authors from the Originating laboratories responsible for obtaining the specimens, as well as the Submitting laboratories where the genome data were generated and shared via GISAID, on which this research is based.

All Submitters of data may be contacted directly via www.gisaid.org

Authors are sorted alphabetically.

| Accession ID | Originating Laboratory | Submitting Laboratory | Authors |
| --- | --- | --- | --- |
| EPI_ISL_9235998, EPI_ISL_9236007, EPI_ISL_9236015, EPI_ISL_9236022, EPI_ISL_9236043, EPI_ISL_9236060, EPI_ISL_9236062, EPI_ISL_9236079, EPI_ISL_9236112, EPI_ISL_9236118, EPI_ISL_9236127, EPI_ISL_9236292, EPI_ISL_9236297, EPI_ISL_9236302, EPI_ISL_9236316, EPI_ISL_9236335, EPI_ISL_9236505, EPI_ISL_9236541, EPI_ISL_9236582, EPI_ISL_9236592, EPI_ISL_9236624, EPI_ISL_9236646, EPI_ISL_9236667, EPI_ISL_9236674, EPI_ISL_9236696, EPI_ISL_9236699, EPI_ISL_9236707, EPI_ISL_9236720, EPI_ISL_9236722, EPI_ISL_9236728, EPI_ISL_9236730, EPI_ISL_9236737, EPI_ISL_9236744, EPI_ISL_9236751, EPI_ISL_9236755, EPI_ISL_9236772, EPI_ISL_9236774, EPI_ISL_9236775, EPI_ISL_9236788, EPI_ISL_9236800, EPI_ISL_9236840, EPI_ISL_9236841, EPI_ISL_9236847, EPI_ISL_9236855, EPI_ISL_9236858, EPI_ISL_9236867, EPI_ISL_9236880, EPI_ISL_9236887, EPI_ISL_9236889, EPI_ISL_9236929, EPI_ISL_9236942, EPI_ISL_9236947, EPI_ISL_9236958, EPI_ISL_9236981, EPI_ISL_9237008, EPI_ISL_9237021, EPI_ISL_9237029, EPI_ISL_9237043, EPI_ISL_9237047, EPI_ISL_9237052, EPI_ISL_9237059, EPI_ISL_9237065, EPI_ISL_9237088, EPI_ISL_9237096, EPI_ISL_9237100, EPI_ISL_9237119, EPI_ISL_9237153, EPI_ISL_9237157, EPI_ISL_9237213, EPI_ISL_9237227, EPI_ISL_9237234, EPI_ISL_9237245, EPI_ISL_9237305, EPI_ISL_9237308, EPI_ISL_9237327, EPI_ISL_9237329, EPI_ISL_9237397, EPI_ISL_9237416, EPI_ISL_9237421, EPI_ISL_9237429, EPI_ISL_9237435, EPI_ISL_9237539, EPI_ISL_9237628, EPI_ISL_9237657, EPI_ISL_9237688, EPI_ISL_9237702, EPI_ISL_9237734, EPI_ISL_9237765, EPI_ISL_9237797, EPI_ISL_9237807, EPI_ISL_9237816, EPI_ISL_9237835, EPI_ISL_9237872, EPI_ISL_9237925, EPI_ISL_9238185, EPI_ISL_9238218, EPI_ISL_9238241, EPI_ISL_9238254, EPI_ISL_9238289, EPI_ISL_9238327, EPI_ISL_9238383, EPI_ISL_9238410, EPI_ISL_9238450, EPI_ISL_9238476, EPI_ISL_9238708, EPI_ISL_9238743, EPI_ISL_9238751, EPI_ISL_9238910, EPI_ISL_9238916, EPI_ISL_9238937, EPI_ISL_9238946, EPI_ISL_9238965, EPI_ISL_9239031, EPI_ISL_9239091, EPI_ISL_9239101, EPI_ISL_9239133, EPI_ISL_9239157, EPI_ISL_9239245, EPI_ISL_9239277, EPI_ISL_9239285, EPI_ISL_9239325, EPI_ISL_9239331, EPI_ISL_9239389, EPI_ISL_9239396, EPI_ISL_9239431, EPI_ISL_9239449, EPI_ISL_9239490, EPI_ISL_9239503, EPI_ISL_9239621, EPI_ISL_9239704, EPI_ISL_9239755, EPI_ISL_9239789, EPI_ISL_9239808, EPI_ISL_9239836, EPI_ISL_9239852, EPI_ISL_9239922, EPI_ISL_9239934, EPI_ISL_9239946, EPI_ISL_9240080, EPI_ISL_9240121, EPI_ISL_9240161, EPI_ISL_9240173, EPI_ISL_9240221, EPI_ISL_9240256, EPI_ISL_9240260, EPI_ISL_9240265, EPI_ISL_9240270, EPI_ISL_9240342, EPI_ISL_9240365, EPI_ISL_9240391, EPI_ISL_9240418, EPI_ISL_9240483, EPI_ISL_9240512, EPI_ISL_9240562, EPI_ISL_9240593, EPI_ISL_9240608, EPI_ISL_9240614, EPI_ISL_9240630, EPI_ISL_9240676, EPI_ISL_9240695, EPI_ISL_9240797, EPI_ISL_9240807, EPI_ISL_9240814, EPI_ISL_9240907, EPI_ISL_9240948, EPI_ISL_9240975, EPI_ISL_9241011, EPI_ISL_9241017, EPI_ISL_9241039, EPI_ISL_9241051, EPI_ISL_9241091, EPI_ISL_9241139, EPI_ISL_9241189, EPI_ISL_9241247, EPI_ISL_9241251, EPI_ISL_9241290, EPI_ISL_9241312, EPI_ISL_9241323, EPI_ISL_9241355, EPI_ISL_9241442, EPI_ISL_9241453, EPI_ISL_9241474, EPI_ISL_9241490, EPI_ISL_9241515, EPI_ISL_9241604, EPI_ISL_9241735, EPI_ISL_9241745, EPI_ISL_9241778, EPI_ISL_9241789, EPI_ISL_9241798, EPI_ISL_9241819, EPI_ISL_9241832, EPI_ISL_9241871, EPI_ISL_9241873, EPI_ISL_9241892, EPI_ISL_9241899, EPI_ISL_9241921, EPI_ISL_9241929, EPI_ISL_9241930, EPI_ISL_9241984, EPI_ISL_9241990, EPI_ISL_9242004, EPI_ISL_9242006, EPI_ISL_9242017, EPI_ISL_9242085, EPI_ISL_9242106, EPI_ISL_9242110, EPI_ISL_9242145, EPI_ISL_9242160, EPI_ISL_9242167, EPI_ISL_9242183, EPI_ISL_9242185, EPI_ISL_9242255, EPI_ISL_9242305, EPI_ISL_9242341, EPI_ISL_9242349, EPI_ISL_9242356, EPI_ISL_9242359, EPI_ISL_9242370, EPI_ISL_9242389, EPI_ISL_9242392, EPI_ISL_9242396, EPI_ISL_9242401, EPI_ISL_9242407, EPI_ISL_9242600, EPI_ISL_9242605, EPI_ISL_9242614, EPI_ISL_9242618, EPI_ISL_9242621, EPI_ISL_9242623, EPI_ISL_9242648, EPI_ISL_9242659, EPI_ISL_9242669, EPI_ISL_9242681, EPI_ISL_9242687, EPI_ISL_9242691, EPI_ISL_9242715, EPI_ISL_9242736, EPI_ISL_9242742, EPI_ISL_9242765, EPI_ISL_9242777, EPI_ISL_9242795, EPI_ISL_9242818, EPI_ISL_9242823, EPI_ISL_9242825, EPI_ISL_9242833, EPI_ISL_9242837, EPI_ISL_9242840, EPI_ISL_9242851, EPI_ISL_9242874, EPI_ISL_9242900, EPI_ISL_9242902, EPI_ISL_9242910, EPI_ISL_9242940, EPI_ISL_9242954, EPI_ISL_9242959, EPI_ISL_9242965, EPI_ISL_9242974, EPI_ISL_9242977, EPI_ISL_9243022, EPI_ISL_9243025, EPI_ISL_9243046, EPI_ISL_9243050, EPI_ISL_9243075, EPI_ISL_9243082, EPI_ISL_9243112, EPI_ISL_9243132, EPI_ISL_9243146, EPI_ISL_9243148, EPI_ISL_9243161, EPI_ISL_9243165, EPI_ISL_9243169, EPI_ISL_9243175, EPI_ISL_9243187, EPI_ISL_9243192, EPI_ISL_9243215, EPI_ISL_9243233, EPI_ISL_9243239, EPI_ISL_9243241, EPI_ISL_9243245 | | | |
| see above | Department of Bacteria, Parasites and Fungi, Statens Serum Institut, Copenhagen, Denmark | Statens Serum Institut Bioinformatics and Microbial Genomics | Danish Covid-19 Genome Consortium |
| EPI_ISL_9236011, EPI_ISL_9236037, EPI_ISL_9236085, EPI_ISL_9236114, EPI_ISL_9236123, EPI_ISL_9236161, EPI_ISL_9236300, EPI_ISL_9236310, EPI_ISL_9236453, EPI_ISL_9236527, EPI_ISL_9236529, EPI_ISL_9236537, EPI_ISL_9236555, EPI_ISL_9236578, EPI_ISL_9236614, EPI_ISL_9236644, EPI_ISL_9236663, EPI_ISL_9236677, EPI_ISL_9236705, EPI_ISL_9236709, EPI_ISL_9236798, EPI_ISL_9236813, EPI_ISL_9236852, EPI_ISL_9236860, EPI_ISL_9236864, EPI_ISL_9236878, EPI_ISL_9236903, EPI_ISL_9236935, EPI_ISL_9236968, EPI_ISL_9236975, EPI_ISL_9237000, EPI_ISL_9237002, EPI_ISL_9237207, EPI_ISL_9237260, EPI_ISL_9237372, EPI_ISL_9237558, EPI_ISL_9237643, EPI_ISL_9237670, EPI_ISL_9237751, EPI_ISL_9237782, EPI_ISL_9237909, EPI_ISL_9237935, EPI_ISL_9238027, EPI_ISL_9238102, EPI_ISL_9238141, EPI_ISL_9238322, EPI_ISL_9238333, EPI_ISL_9238470, EPI_ISL_9238604, EPI_ISL_9238662, EPI_ISL_9238823, EPI_ISL_9239058, EPI_ISL_9239129, EPI_ISL_9239166, EPI_ISL_9239170, EPI_ISL_9239197, EPI_ISL_9239225, EPI_ISL_9239268, EPI_ISL_9239559, EPI_ISL_9239760, EPI_ISL_9239762, EPI_ISL_9239771, EPI_ISL_9239823, EPI_ISL_9239826, EPI_ISL_9239918, EPI_ISL_9239936, EPI_ISL_9240310, EPI_ISL_9240459, EPI_ISL_9240554, EPI_ISL_9240640, EPI_ISL_9240702, EPI_ISL_9240718, EPI_ISL_9240782, EPI_ISL_9240841, EPI_ISL_9240863, EPI_ISL_9241066, EPI_ISL_9241068, EPI_ISL_9241151, EPI_ISL_9241277, EPI_ISL_9241344, EPI_ISL_9241372, EPI_ISL_9241411, EPI_ISL_9241437, EPI_ISL_9241457, EPI_ISL_9241491, EPI_ISL_9241597, EPI_ISL_9241612, EPI_ISL_9241702, EPI_ISL_9241720, EPI_ISL_9241810, EPI_ISL_9241876, EPI_ISL_9241894, EPI_ISL_9241919, EPI_ISL_9241974, EPI_ISL_9241999, EPI_ISL_9242026, EPI_ISL_9242043, EPI_ISL_9242058, EPI_ISL_9242080, EPI_ISL_9242093, EPI_ISL_9242122, EPI_ISL_9242124, EPI_ISL_9242129, EPI_ISL_9242140, EPI_ISL_9242163, EPI_ISL_9242178, EPI_ISL_9242240, EPI_ISL_9242326, EPI_ISL_9242330, EPI_ISL_9242351, EPI_ISL_9242366, EPI_ISL_9242671, EPI_ISL_9242747, EPI_ISL_9242749, EPI_ISL_9242754, EPI_ISL_9242782, EPI_ISL_9242853, EPI_ISL_9242860, EPI_ISL_9242945, EPI_ISL_9242956, EPI_ISL_9242970, EPI_ISL_9242978, EPI_ISL_9242988, EPI_ISL_9243031, EPI_ISL_9243084, EPI_ISL_9243087, EPI_ISL_9243130, EPI_ISL_9243136, EPI_ISL_9243205, EPI_ISL_9243206, EPI_ISL_9243270 | | | |
| see above | Department of Clinical Microbiology and Center for Genomic Medicine, Rigshospitalet, Copenhagen, Denmark | Statens Serum Institut Bioinformatics and Microbial Genomics | Danish Covid-19 Genome Consortium |

We gratefully acknowledge the following Authors from the Originating laboratories responsible for obtaining the specimens, as well as the Submitting laboratories where the genome data were generated and shared via GISAID, on which this research is based.

All Submitters of data may be contacted directly via www.gisaid.org

Authors are sorted alphabetically.

| Accession ID | Originating Laboratory | Submitting Laboratory | Authors |
|---|---|---|---|
| EPI_ISL_9235984, EPI_ISL_9236000, EPI_ISL_9236002, EPI_ISL_9236017, EPI_ISL_9236028, EPI_ISL_9236033, EPI_ISL_9236047, EPI_ISL_9236049, EPI_ISL_9236103, EPI_ISL_9236110, EPI_ISL_9236152, EPI_ISL_9236286, EPI_ISL_9236306, EPI_ISL_9236314, EPI_ISL_9236508, EPI_ISL_9236544, EPI_ISL_9236552, EPI_ISL_9236564, EPI_ISL_9236567, EPI_ISL_9236590, EPI_ISL_9236618, EPI_ISL_9236626, EPI_ISL_9236628, EPI_ISL_9236633, EPI_ISL_9236636, EPI_ISL_9236638, EPI_ISL_9236653, EPI_ISL_9236686, EPI_ISL_9236731, EPI_ISL_9236781, EPI_ISL_9236784, EPI_ISL_9236790, EPI_ISL_9236820, EPI_ISL_9236835, EPI_ISL_9236856, EPI_ISL_9236873, EPI_ISL_9236949, EPI_ISL_9236961, EPI_ISL_9236966, EPI_ISL_9236969, EPI_ISL_9236971, EPI_ISL_9237015, EPI_ISL_9237019, EPI_ISL_9237034, EPI_ISL_9237085, EPI_ISL_9237092, EPI_ISL_9237108, EPI_ISL_9237111, EPI_ISL_9237130, EPI_ISL_9237159, EPI_ISL_9237171, EPI_ISL_9237187, EPI_ISL_9237204, EPI_ISL_9237221, EPI_ISL_9237273, EPI_ISL_9237547, EPI_ISL_9237585, EPI_ISL_9237604, EPI_ISL_9237824, EPI_ISL_9237876, EPI_ISL_9238046, EPI_ISL_9238082, EPI_ISL_9238088, EPI_ISL_9238093, EPI_ISL_9238118, EPI_ISL_9238126, EPI_ISL_9238164, EPI_ISL_9238265, EPI_ISL_9238309, EPI_ISL_9238358, EPI_ISL_9238557, EPI_ISL_9238641, EPI_ISL_9238661, EPI_ISL_9238672, EPI_ISL_9238719, EPI_ISL_9238757, EPI_ISL_9238790, EPI_ISL_9238818, EPI_ISL_9238855, EPI_ISL_9238943, EPI_ISL_9239231, EPI_ISL_9239235, EPI_ISL_9239320, EPI_ISL_9239441, EPI_ISL_9239476, EPI_ISL_9239496, EPI_ISL_9239545, EPI_ISL_9239606, EPI_ISL_9239627, EPI_ISL_9239730, EPI_ISL_9239884, EPI_ISL_9239893, EPI_ISL_9239950, EPI_ISL_9239955, EPI_ISL_9239957, EPI_ISL_9239973, EPI_ISL_9240067, EPI_ISL_9240133, EPI_ISL_9240170, EPI_ISL_9240235, EPI_ISL_9240240, EPI_ISL_9240244, EPI_ISL_9240278, EPI_ISL_9240287, EPI_ISL_9240344, EPI_ISL_9240350, EPI_ISL_9240357, EPI_ISL_9240368, EPI_ISL_9240408, EPI_ISL_9240487, EPI_ISL_9240530, EPI_ISL_9240588, EPI_ISL_9240619, EPI_ISL_9240665, EPI_ISL_9240932, EPI_ISL_9240999, EPI_ISL_9241080, EPI_ISL_9241205, EPI_ISL_9241213, EPI_ISL_9241341, EPI_ISL_9241416, EPI_ISL_9241601, EPI_ISL_9241705, EPI_ISL_9241707, EPI_ISL_9241729, EPI_ISL_9241773, EPI_ISL_9241881, EPI_ISL_9241916, EPI_ISL_9241986, EPI_ISL_9242046, EPI_ISL_9242055, EPI_ISL_9242060, EPI_ISL_9242068, EPI_ISL_9242088, EPI_ISL_9242111, EPI_ISL_9242118, EPI_ISL_9242132, EPI_ISL_9242199, EPI_ISL_9242209, EPI_ISL_9242230, EPI_ISL_9242322, EPI_ISL_9242337, EPI_ISL_9242346, EPI_ISL_9242602, EPI_ISL_9242617, EPI_ISL_9242636, EPI_ISL_9242657, EPI_ISL_9242678, EPI_ISL_9242693, EPI_ISL_9242702, EPI_ISL_9242728, EPI_ISL_9242731, EPI_ISL_9242788, EPI_ISL_9242791, EPI_ISL_9242845, EPI_ISL_9242911, EPI_ISL_9242915, EPI_ISL_9242918, EPI_ISL_9242923, EPI_ISL_9242929, EPI_ISL_9242972, EPI_ISL_9242985, EPI_ISL_9243006, EPI_ISL_9243010, EPI_ISL_9243029, EPI_ISL_9243062, EPI_ISL_9243079, EPI_ISL_9243094, EPI_ISL_9243107, EPI_ISL_9243120, EPI_ISL_9243126, EPI_ISL_9243128, EPI_ISL_9243138, EPI_ISL_9243159, EPI_ISL_9243163, EPI_ISL_9243172, EPI_ISL_9243184, EPI_ISL_9243190, EPI_ISL_9243230, EPI_ISL_9243235 | | | |
| see above | Department of Bacteria, Parasites and Fungi, Statens Serum Institut, Copenhagen, Denmark | Statens Serum Institut Bioinformatics and Microbial Genomics | Danish Covid-19 Genome Consortium |

We gratefully acknowledge the following Authors from the Originating laboratories responsible for obtaining the specimens, as well as the Submitting laboratories where the genome data were generated and shared via GISAID, on which this research is based.

All Submitters of data may be contacted directly via www.gisaid.org

Authors are sorted alphabetically.

| Accession ID | Originating Laboratory | Submitting Laboratory | Authors |
| --- | --- | --- | --- |
| EPI_ISL_9236051, EPI_ISL_9236141, EPI_ISL_9236285, EPI_ISL_9236317, EPI_ISL_9236616, EPI_ISL_9236726, EPI_ISL_9236747, EPI_ISL_9236771, EPI_ISL_9236808, EPI_ISL_9236833, EPI_ISL_9236888, EPI_ISL_9236897, EPI_ISL_9236912, EPI_ISL_9236979, EPI_ISL_9236989, EPI_ISL_9237005, EPI_ISL_9237082, EPI_ISL_9237125, EPI_ISL_9237216, EPI_ISL_9237268, EPI_ISL_9237335, EPI_ISL_9237674, EPI_ISL_9237810, EPI_ISL_9237820, EPI_ISL_9237914, EPI_ISL_9237938, EPI_ISL_9237971, EPI_ISL_9238069, EPI_ISL_9238260, EPI_ISL_9238724, EPI_ISL_9238951, EPI_ISL_9238971, EPI_ISL_9239219, EPI_ISL_9239272, EPI_ISL_9239380, EPI_ISL_9239437, EPI_ISL_9239487, EPI_ISL_9239511, EPI_ISL_9239554, EPI_ISL_9239593, EPI_ISL_9239650, EPI_ISL_9240148, EPI_ISL_9240186, EPI_ISL_9241109, EPI_ISL_9241261, EPI_ISL_9241318, EPI_ISL_9241507, EPI_ISL_9241713, EPI_ISL_9241787, EPI_ISL_9241853, EPI_ISL_9241972, EPI_ISL_9242083, EPI_ISL_9242503, EPI_ISL_9242639, EPI_ISL_9242641, EPI_ISL_9242674, EPI_ISL_9242849, EPI_ISL_9242896, EPI_ISL_9242920, EPI_ISL_9243048, EPI_ISL_9243064, EPI_ISL_9243262 | | | |
| see above | Department of Bacteria, Parasites and Fungi, Statens Serum Institut, Copenhagen, Denmark | Statens Serum Institut Bioinformatics and Microbial Genomics | Danish Covid-19 Genome Consortium |

We gratefully acknowledge the following Authors from the Originating laboratories responsible for obtaining the specimens, as well as the Submitting laboratories where the genome data were generated and shared via GISAID, on which this research is based.

All Submitters of data may be contacted directly via www.gisaid.org

Authors are sorted alphabetically.

| Accession ID | Originating Laboratory | Submitting Laboratory | Authors |
|---|---|---|---|
| EPI_ISL_9235987, EPI_ISL_9236053, EPI_ISL_9236107, EPI_ISL_9236136, EPI_ISL_9236284, EPI_ISL_9236296, EPI_ISL_9236561, EPI_ISL_9236640, EPI_ISL_9236671, EPI_ISL_9236684, EPI_ISL_9236694, EPI_ISL_9236735, EPI_ISL_9236739, EPI_ISL_9236760, EPI_ISL_9236764, EPI_ISL_9236794, EPI_ISL_9236802, EPI_ISL_9236883, EPI_ISL_9236923, EPI_ISL_9236973, EPI_ISL_9236983, EPI_ISL_9236991, EPI_ISL_9237012, EPI_ISL_9237017, EPI_ISL_9237056, EPI_ISL_9237074, EPI_ISL_9237250, EPI_ISL_9237263, EPI_ISL_9237286, EPI_ISL_9237300, EPI_ISL_9237348, EPI_ISL_9237443, EPI_ISL_9237475, EPI_ISL_9237543, EPI_ISL_9237553, EPI_ISL_9237593, EPI_ISL_9237693, EPI_ISL_9237722, EPI_ISL_9237918, EPI_ISL_9238098, EPI_ISL_9238190, EPI_ISL_9238273, EPI_ISL_9238338, EPI_ISL_9238353, EPI_ISL_9238424, EPI_ISL_9238462, EPI_ISL_9238531, EPI_ISL_9238534, EPI_ISL_9238955, EPI_ISL_9239062, EPI_ISL_9239072, EPI_ISL_9239086, EPI_ISL_9239115, EPI_ISL_9239143, EPI_ISL_9239454, EPI_ISL_9239474, EPI_ISL_9239500, EPI_ISL_9239538, EPI_ISL_9239598, EPI_ISL_9239741, EPI_ISL_9239779, EPI_ISL_9239909, EPI_ISL_9240031, EPI_ISL_9240058, EPI_ISL_9240062, EPI_ISL_9240088, EPI_ISL_9240112, EPI_ISL_9240212, EPI_ISL_9240248, EPI_ISL_9240328, EPI_ISL_9240336, EPI_ISL_9240432, EPI_ISL_9240446, EPI_ISL_9240738, EPI_ISL_9240768, EPI_ISL_9240774, EPI_ISL_9240880, EPI_ISL_9241103, EPI_ISL_9241179, EPI_ISL_9241224, EPI_ISL_9241265, EPI_ISL_9241351, EPI_ISL_9241676, EPI_ISL_9241757, EPI_ISL_9241801, EPI_ISL_9241826, EPI_ISL_9241960, EPI_ISL_9241976, EPI_ISL_9241996, EPI_ISL_9242015, EPI_ISL_9242121, EPI_ISL_9242125, EPI_ISL_9242214, EPI_ISL_9242226, EPI_ISL_9242312, EPI_ISL_9242319, EPI_ISL_9242323, EPI_ISL_9242334, EPI_ISL_9242344, EPI_ISL_9242402, EPI_ISL_9242695, EPI_ISL_9242717, EPI_ISL_9242756, EPI_ISL_9242768, EPI_ISL_9242787, EPI_ISL_9242814, EPI_ISL_9242828, EPI_ISL_9242830, EPI_ISL_9242883, EPI_ISL_9242894, EPI_ISL_9242936, EPI_ISL_9242948, EPI_ISL_9242950, EPI_ISL_9242992, EPI_ISL_9243032, EPI_ISL_9243072, EPI_ISL_9243092, EPI_ISL_9243140, EPI_ISL_9243142, EPI_ISL_9243171, EPI_ISL_9243183, EPI_ISL_9243194, EPI_ISL_9243210, EPI_ISL_9243219, EPI_ISL_9243268, EPI_ISL_9243272, EPI_ISL_9243277 | | | |
| see above | Department of Bacteria, Parasites and Fungi, Statens Serum Institut, Copenhagen, Denmark | Statens Serum Institut Bioinformatics and Microbial Genomics | Danish Covid-19 Genome Consortium |
| EPI_ISL_9237756, EPI_ISL_9241431, EPI_ISL_9241687 | Department of Clinical Microbiology and Center for Genomic Medicine, Rigshospitalet, Copenhagen, Denmark | Statens Serum Institut Bioinformatics and Microbial Genomics | Danish Covid-19 Genome Consortium |

We gratefully acknowledge the following Authors from the Originating laboratories responsible for obtaining the specimens, as well as the Submitting laboratories where the genome data were generated and shared via GISAID, on which this research is based.

All Submitters of data may be contacted directly via www.gisaid.org

Authors are sorted alphabetically.

Acknowledgement EPI_SET Identifier: EPI_SET_20220527tw

| Accession ID | Originating Laboratory | Submitting Laboratory | Authors |
| --- | --- | --- | --- |
| EPI_ISL_9235982, EPI_ISL_9236041, EPI_ISL_9236077, EPI_ISL_9236099, EPI_ISL_9236105, EPI_ISL_9236122, EPI_ISL_9236140, EPI_ISL_9236147, EPI_ISL_9236160, EPI_ISL_9236282, EPI_ISL_9236288, EPI_ISL_9236313, EPI_ISL_9236512, EPI_ISL_9236531, EPI_ISL_9236539, EPI_ISL_9236548, EPI_ISL_9236550, EPI_ISL_9236570, EPI_ISL_9236599, EPI_ISL_9236601, EPI_ISL_9236676, EPI_ISL_9236681, EPI_ISL_9236697, EPI_ISL_9236724, EPI_ISL_9236786, EPI_ISL_9236796, EPI_ISL_9236804, EPI_ISL_9236872, EPI_ISL_9236895, EPI_ISL_9236996, EPI_ISL_9237040, EPI_ISL_9237041, EPI_ISL_9237054, EPI_ISL_9237098, EPI_ISL_9237183, EPI_ISL_9237229, EPI_ISL_9237313, EPI_ISL_9237340, EPI_ISL_9237439, EPI_ISL_9237513, EPI_ISL_9237608, EPI_ISL_9237613, EPI_ISL_9237903, EPI_ISL_9237931, EPI_ISL_9237966, EPI_ISL_9238041, EPI_ISL_9238054, EPI_ISL_9238079, EPI_ISL_9238136, EPI_ISL_9238181, EPI_ISL_9238203, EPI_ISL_9238554, EPI_ISL_9238566, EPI_ISL_9238650, EPI_ISL_9238666, EPI_ISL_9238703, EPI_ISL_9238869, EPI_ISL_9238977, EPI_ISL_9238979, EPI_ISL_9239034, EPI_ISL_9239119, EPI_ISL_9239124, EPI_ISL_9239176, EPI_ISL_9239305, EPI_ISL_9239375, EPI_ISL_9239602, EPI_ISL_9239631, EPI_ISL_9239722, EPI_ISL_9239775, EPI_ISL_9239798, EPI_ISL_9239882, EPI_ISL_9239901, EPI_ISL_9239941, EPI_ISL_9239983, EPI_ISL_9239988, EPI_ISL_9240033, EPI_ISL_9240086, EPI_ISL_9240119, EPI_ISL_9240154, EPI_ISL_9240166, EPI_ISL_9240169, EPI_ISL_9240355, EPI_ISL_9240399, EPI_ISL_9240402, EPI_ISL_9240501, EPI_ISL_9240558, EPI_ISL_9240578, EPI_ISL_9240604, EPI_ISL_9240688, EPI_ISL_9240711, EPI_ISL_9240818, EPI_ISL_9240923, EPI_ISL_9241030, EPI_ISL_9241044, EPI_ISL_9241054, EPI_ISL_9241074, EPI_ISL_9241129, EPI_ISL_9241136, EPI_ISL_9241201, EPI_ISL_9241284, EPI_ISL_9241294, EPI_ISL_9241332, EPI_ISL_9241336, EPI_ISL_9241478, EPI_ISL_9241539, EPI_ISL_9241545, EPI_ISL_9241743, EPI_ISL_9241774, EPI_ISL_9241780, EPI_ISL_9241792, EPI_ISL_9241818, EPI_ISL_9241865, EPI_ISL_9241885, EPI_ISL_9241901, EPI_ISL_9241939, EPI_ISL_9241942, EPI_ISL_9242065, EPI_ISL_9242077, EPI_ISL_9242114, EPI_ISL_9242143, EPI_ISL_9242150, EPI_ISL_9242152, EPI_ISL_9242166, EPI_ISL_9242172, EPI_ISL_9242176, EPI_ISL_9242180, EPI_ISL_9242187, EPI_ISL_9242212, EPI_ISL_9242221, EPI_ISL_9242223, EPI_ISL_9242228, EPI_ISL_9242253, EPI_ISL_9242607, EPI_ISL_9242625, EPI_ISL_9242630, EPI_ISL_9242664, EPI_ISL_9242666, EPI_ISL_9242763, EPI_ISL_9242793, EPI_ISL_9242843, EPI_ISL_9242871, EPI_ISL_9242881, EPI_ISL_9242884, EPI_ISL_9242891, EPI_ISL_9242963, EPI_ISL_9243004, EPI_ISL_9243066, EPI_ISL_9243105, EPI_ISL_9243144, EPI_ISL_9243191, EPI_ISL_9243203, EPI_ISL_9243223, EPI_ISL_9243243, EPI_ISL_9243248, EPI_ISL_9243279 | | | |
| see above | Department of Bacteria, Parasites and Fungi, Statens Serum Institut, Copenhagen, Denmark | Statens Serum Institut Bioinformatics and Microbial Genomics | Danish Covid-19 Genome Consortium |

We gratefully acknowledge the following Authors from the Originating laboratories responsible for obtaining the specimens, as well as the Submitting laboratories where the genome data were generated and shared via GISAID, on which this research is based.

All Submitters of data may be contacted directly via www.gisaid.org

Authors are sorted alphabetically.

| Accession ID | Originating Laboratory | Submitting Laboratory | Authors |
|---|---|---|---|
| EPI_ISL_9256852 | Accolab Sud Ouest | CERBA HealthCare | Bénédicte Roquebert; Laura Verdurme; Mathilde Roussel; Sabine Trombert; Stéphanie Haim-Boukobza |
| EPI_ISL_9257606 | BIO DOMES UNILABS SITE DANZIGER | CERBA HealthCare | Bénédicte Roquebert; Laura Verdurme; Mathilde Roussel; Sabine Trombert; Stéphanie Haim-Boukobza |
| EPI_ISL_9256823, EPI_ISL_9256824 | Bio Domes Unilabs Site Danziger | CERBA HealthCare | Bénédicte Roquebert; Laura Verdurme; Mathilde Roussel; Sabine Trombert; Stéphanie Haim-Boukobza |
| EPI_ISL_9256851 | Bio Lbs - Lbm De Lillebonne | CERBA HealthCare | Bénédicte Roquebert; Laura Verdurme; Mathilde Roussel; Sabine Trombert; Stéphanie Haim-Boukobza |
| EPI_ISL_9256822 | Biolab Unilabs Site Boucicaut | CERBA HealthCare | Bénédicte Roquebert; Laura Verdurme; Mathilde Roussel; Sabine Trombert; Stéphanie Haim-Boukobza |
| EPI_ISL_9257607 | CERBA | CERBA HealthCare | Bénédicte Roquebert; Laura Verdurme; Mathilde Roussel; Sabine Trombert; Stéphanie Haim-Boukobza |
| EPI_ISL_9257727 | CERBALLIANCE ALPES DURANCE | CERBA HealthCare | Bénédicte Roquebert; Laura Verdurme; Mathilde Roussel; Sabine Trombert; Stéphanie Haim-Boukobza |
| EPI_ISL_9257730 | CERBALLIANCE ALPES DURANCE SITE TOKORO | CERBA HealthCare | Bénédicte Roquebert; Laura Verdurme; Mathilde Roussel; Sabine Trombert; Stéphanie Haim-Boukobza |
| EPI_ISL_9257722, EPI_ISL_9257723, EPI_ISL_9257725 | CERBALLIANCE ARTOIS | CERBA HealthCare | Bénédicte Roquebert; Laura Verdurme; Mathilde Roussel; Sabine Trombert; Stéphanie Haim-Boukobza |
| EPI_ISL_9257726 | CERBALLIANCE COTE D'AZUR | CERBA HealthCare | Bénédicte Roquebert; Laura Verdurme; Mathilde Roussel; Sabine Trombert; Stéphanie Haim-Boukobza |
| EPI_ISL_9257728 | CERBALLIANCE COTE D'AZUR SITE ST LAURENT | CERBA HealthCare | Bénédicte Roquebert; Laura Verdurme; Mathilde Roussel; Sabine Trombert; Stéphanie Haim-Boukobza |
| EPI_ISL_9257729 | CERBALLIANCE COTE D'AZUR SITE STRASBOURG | CERBA HealthCare | Bénédicte Roquebert; Laura Verdurme; Mathilde Roussel; Sabine Trombert; Stéphanie Haim-Boukobza |
| EPI_ISL_9257847 | CERBALLIANCE IDF OUEST SITE FOCH | CERBA HealthCare | Bénédicte Roquebert; Laura Verdurme; Mathilde Roussel; Sabine Trombert; Stéphanie Haim-Boukobza |
| EPI_ISL_9257846 | CERBALLIANCE IDF OUEST SITE LES CLAYES | CERBA HealthCare | Bénédicte Roquebert; Laura Verdurme; Mathilde Roussel; Sabine Trombert; Stéphanie Haim-Boukobza |
| EPI_ISL_9257605, EPI_ISL_9257608, EPI_ISL_9257609, EPI_ISL_9257610, EPI_ISL_9257611, EPI_ISL_9257612, EPI_ISL_9257613, EPI_ISL_9257614, EPI_ISL_9257615, EPI_ISL_9257616, EPI_ISL_9257617, EPI_ISL_9257618, EPI_ISL_9257619, EPI_ISL_9257731, EPI_ISL_9257732, EPI_ISL_9257733, EPI_ISL_9257734, EPI_ISL_9257735, EPI_ISL_9257736, EPI_ISL_9257737, EPI_ISL_9257738, EPI_ISL_9257739, EPI_ISL_9257740, EPI_ISL_9257741, EPI_ISL_9257742, EPI_ISL_9257743, EPI_ISL_9257744, EPI_ISL_9257745, EPI_ISL_9257848, EPI_ISL_9257849, EPI_ISL_9257850, EPI_ISL_9257851, EPI_ISL_9257856 | | | |
| see above | CERBALLIANCE PARIS ET IDF EST | CERBA HealthCare | Bénédicte Roquebert; Laura Verdurme; Mathilde Roussel; Sabine Trombert; Stéphanie Haim-Boukobza |
| EPI_ISL_9257852, EPI_ISL_9257853, EPI_ISL_9257855 | CERBALLIANCE PARIS IDF EST SITE PYRENEES | CERBA HealthCare | Bénédicte Roquebert; Laura Verdurme; Mathilde Roussel; Sabine Trombert; Stéphanie Haim-Boukobza |
| EPI_ISL_9257854 | CERBALLIANCE PARIS IDF EST TROCADER0 | CERBA HealthCare | Bénédicte Roquebert; Laura Verdurme; Mathilde Roussel; Sabine Trombert; Stéphanie Haim-Boukobza |
| EPI_ISL_9257604, EPI_ISL_9257620, EPI_ISL_9257621, EPI_ISL_9257622, EPI_ISL_9257623, EPI_ISL_9257624, EPI_ISL_9257625, EPI_ISL_9257626, EPI_ISL_9257627, EPI_ISL_9257628, EPI_ISL_9257629, EPI_ISL_9257630, EPI_ISL_9257631, EPI_ISL_9257632, EPI_ISL_9257633, EPI_ISL_9257634, EPI_ISL_9257635, EPI_ISL_9257636, EPI_ISL_9257640, EPI_ISL_9257641 | | | |
| see above | CERBALLIANCE RHONE ALPES | CERBA HealthCare | Bénédicte Roquebert; Laura Verdurme; Mathilde Roussel; Sabine Trombert; Stéphanie Haim-Boukobza |
| EPI_ISL_9257721, EPI_ISL_9257724 | CERBALLIANCE SOMME | CERBA HealthCare | Bénédicte Roquebert; Laura Verdurme; Mathilde Roussel; Sabine Trombert; Stéphanie Haim-Boukobza |
| EPI_ISL_9246081 | CHRU BREST-Site Carhaix | laboratoire de Virologie CHRU Brest | Adissa Tran Minoui; Christopher Payan; Léa Pilorgé; Pauline Bargain; Simon Rambaut; Sophie Vallet |
| EPI_ISL_9246091 | CHU BREST | laboratoire de Virologie CHRU Brest | Adissa Tran Minoui; Christopher Payan; Léa Pilorgé; Pauline Bargain; Simon Rambaut; Sophie Vallet |
| EPI_ISL_9257018 | Centre Municipal De Sante | CERBA HealthCare | Bénédicte Roquebert; Laura Verdurme; Sabine Trombert; Stéphanie Haim-Boukobza |
| EPI_ISL_9256853 | Cerballiance Cote D'azur | CERBA HealthCare | Bénédicte Roquebert; Laura Verdurme; Mathilde Roussel; Sabine Trombert; Stéphanie Haim-Boukobza |
| EPI_ISL_9256835 | Cerballiance Idf Ouest Site Les Clayes | CERBA HealthCare | Bénédicte Roquebert; Laura Verdurme; Mathilde Roussel; Sabine Trombert; Stéphanie Haim-Boukobza |
| EPI_ISL_9256825, EPI_ISL_9256826, EPI_ISL_9256827, EPI_ISL_9256828, EPI_ISL_9256830, EPI_ISL_9256831, EPI_ISL_9256847, EPI_ISL_9256848, EPI_ISL_9256849, EPI_ISL_9256850, EPI_ISL_9256854, EPI_ISL_9257019 | | | |
| see above | Cerballiance Paris Et Idf Est | CERBA HealthCare | Bénédicte Roquebert; Laura Verdurme; Mathilde Roussel; Sabine Trombert; Stéphanie Haim-Boukobza |
| EPI_ISL_9257637, EPI_ISL_9257638 | EXALAB BISCARROSSE | CERBA HealthCare | Bénédicte Roquebert; Laura Verdurme; Mathilde Roussel; Sabine Trombert; Stéphanie Haim-Boukobza |
| EPI_ISL_9256832 | Hopital Foch | CERBA HealthCare | Bénédicte Roquebert; Laura Verdurme; Mathilde Roussel; Sabine Trombert; Stéphanie Haim-Boukobza |
| EPI_ISL_9257718, EPI_ISL_9257719 | LABM CANDIA MADONUCCIA | CERBA HealthCare | Bénédicte Roquebert; Laura Verdurme; Mathilde Roussel; Sabine Trombert; Stéphanie Haim-Boukobza |
| EPI_ISL_9257639 | LABORATOIRE ACCOLAB | CERBA HealthCare | Bénédicte Roquebert; Laura Verdurme; Mathilde Roussel; Sabine Trombert; Stéphanie Haim-Boukobza |
| EPI_ISL_9257720 | LABORATOIRE AUDOLYS - LONGUENESSE | CERBA HealthCare | Bénédicte Roquebert; Laura Verdurme; Mathilde Roussel; Sabine Trombert; Stéphanie Haim-Boukobza |
| EPI_ISL_9256845, EPI_ISL_9256846 | Labm Bonhoure | CERBA HealthCare | Bénédicte Roquebert; Laura Verdurme; Mathilde Roussel; Sabine Trombert; Stéphanie Haim-Boukobza |
| EPI_ISL_9256821 | Labm Eylau Unilabs Suchet | CERBA HealthCare | Bénédicte Roquebert; Laura Verdurme; Mathilde Roussel; Sabine Trombert; Stéphanie Haim-Boukobza |
| EPI_ISL_9256836, EPI_ISL_9256837, EPI_ISL_9256838, EPI_ISL_9256839, EPI_ISL_9256840, EPI_ISL_9256841, EPI_ISL_9256842, EPI_ISL_9256843, EPI_ISL_9256844 | | | |
| see above | Labm Kuate | CERBA HealthCare | Bénédicte Roquebert; Laura Verdurme; Mathilde Roussel; Sabine Trombert; Stéphanie Haim-Boukobza |
| EPI_ISL_9245672, EPI_ISL_9245684 | Labo Analyses Med | National Reference Center for Viruses of Respiratory Infections, Institut Pasteur, Paris | Angela Brisebarre; Camille Capel; Christophe Malabat; Corinne Maufrais; Etienne Simon-Lorière; Frédéric Lemoine; J.M CORCY; Julien Fumey; Louise Lefrançois; Marion Barbet; Maud Vanpeene; Méline Bizard; Slim El Khiari; Sylvie Behillil; Sylvie Van der Werf; Vincent Enouf |
| EPI_ISL_9256834 | Laboratoire Clinique De Meudon La Foret | CERBA HealthCare | Bénédicte Roquebert; Laura Verdurme; Mathilde Roussel; Sabine Trombert; Stéphanie Haim-Boukobza |
| EPI_ISL_9256829 | Laboratoire De La Cpam De Paris | CERBA HealthCare | Bénédicte Roquebert; Laura Verdurme; Mathilde Roussel; Sabine Trombert; Stéphanie Haim-Boukobza |

We gratefully acknowledge the following Authors from the Originating laboratories responsible for obtaining the specimens, as well as the Submitting laboratories where the genome data were generated and shared via GISAID, on which this research is based.

All Submitters of data may be contacted directly via www.gisaid.org

Authors are sorted alphabetically.

| Accession ID | Originating Laboratory | Submitting Laboratory | Authors |
|---|---|---|---|
| EPI_ISL_9220562, EPI_ISL_9220563, EPI_ISL_9220734 | Bioscientia Labor Wermsdorf | Robert Koch Institute | |
| EPI_ISL_9210610 | CENTOGENE Frankfurt Laboratory: Niederlassung Industriepark Höchst | Robert Koch Institute | |
| EPI_ISL_9214040, EPI_ISL_9214049 | Clemenshospital - Alexianer Clemenshospital Münster | Robert Koch Institute | |
| EPI_ISL_9227746, EPI_ISL_9227774 | Department für Labormedizin Abteilung III Bereich Molekulare Diagnostik Universitätsklinikum Halle(Saale) | Robert Koch Institute | |
| EPI_ISL_9230811 | Helios Universitätsklinikum Wuppertal: Universität Witten/Herdecke: Institut für Medizinische Labordiagnostik | Robert Koch Institute | |
| EPI_ISL_9215680, EPI_ISL_9215705, EPI_ISL_9215706, EPI_ISL_9215737, EPI_ISL_9215856 | IMD Labor Frankfurt | Robert Koch Institute | |
| EPI_ISL_9233956, EPI_ISL_9233967, EPI_ISL_9234559 | Institut für Immunologie und Genetik Kaiserslautern: Medizinisches Labor Dr. med. B. Thiele | Robert Koch Institute | |
| EPI_ISL_9228558 | Kerckhoff-Klinik GmbH | Robert Koch Institute | |
| EPI_ISL_9212611, EPI_ISL_9212614, EPI_ISL_9212617, EPI_ISL_9212624, EPI_ISL_9212625, EPI_ISL_9212627, EPI_ISL_9212634, EPI_ISL_9212635, EPI_ISL_9212637, EPI_ISL_9212641, EPI_ISL_9212643, EPI_ISL_9212644, EPI_ISL_9212647, EPI_ISL_9212649, EPI_ISL_9212652, EPI_ISL_9212655 | | | |
| see above | Klinikum Stuttgart | Robert Koch Institute | |
| EPI_ISL_9211268, EPI_ISL_9216283, EPI_ISL_9216611, EPI_ISL_9216619, EPI_ISL_9216626, EPI_ISL_9216644, EPI_ISL_9216646, EPI_ISL_9216662, EPI_ISL_9221467, EPI_ISL_9221483, EPI_ISL_9223230, EPI_ISL_9223234, EPI_ISL_9228669 | | | |
| see above | Labor Mönchengladbach MVZ Dr. Stein + Kollegen GbR | Robert Koch Institute | |
| EPI_ISL_9215843, EPI_ISL_9215852 | Labormedizin Darmstadt | Robert Koch Institute | |
| EPI_ISL_9230344 | Landesamt für Verbraucherschutz des Landes Sachsen-Anhalt | Robert Koch Institute | |
| EPI_ISL_9223182 | Limbach - MVZ Labor Vorpommern-Rügen Stralsund | Robert Koch Institute | |
| EPI_ISL_9216728, EPI_ISL_9216738, EPI_ISL_9216756, EPI_ISL_9216819, EPI_ISL_9216845, EPI_ISL_9216861, EPI_ISL_9216896, EPI_ISL_9216901, EPI_ISL_9217082, EPI_ISL_9217101, EPI_ISL_9217109, EPI_ISL_9217199, EPI_ISL_9217315, EPI_ISL_9217364, EPI_ISL_9217375, EPI_ISL_9217407, EPI_ISL_9217423 | | | |
| see above | Limbach - MVZ Labor Dr. Volkmann & Kollegen | Robert Koch Institute | |
| EPI_ISL_9216058, EPI_ISL_9216098, EPI_ISL_9216100, EPI_ISL_9216190, EPI_ISL_9216989, EPI_ISL_9221391 | MDI Limbach Berlin GmbH; MVZ Labor Berlin | Robert Koch Institute | |
| EPI_ISL_9211745, EPI_ISL_9222028, EPI_ISL_9222032, EPI_ISL_9222037, EPI_ISL_9222085, EPI_ISL_9222110, EPI_ISL_9222169, EPI_ISL_9222251 | | | |
| see above | MVZ Labor Dr. Fenner und Kollegen (Standort Hamburg) | Robert Koch Institute | |
| EPI_ISL_9217018, EPI_ISL_9221237, EPI_ISL_9221264, EPI_ISL_9221422, EPI_ISL_9223212, EPI_ISL_9227890, EPI_ISL_9227931, EPI_ISL_9228410, EPI_ISL_9228449, EPI_ISL_9228465, EPI_ISL_9228521, EPI_ISL_9228533, EPI_ISL_9228539, EPI_ISL_9228592, EPI_ISL_9228654, EPI_ISL_9228701, EPI_ISL_9230255 | | | |
| see above | MVZ Labor Dr. Limbach & Kollegen GbR | Robert Koch Institute | |
| EPI_ISL_9217306 | MVZ Labor Dr. Reising-Ackermann und Kollegen | Robert Koch Institute | |
| EPI_ISL_9228029, EPI_ISL_9228115, EPI_ISL_9228244 | MVZ Labor Krone GbR | Robert Koch Institute | |
| EPI_ISL_9221270, EPI_ISL_9221389 | MVZ Medizinisches Labor Hannover GmbH | Robert Koch Institute | |
| EPI_ISL_9216903 | Medizinisches Labor Ostsachsen MVZ GbR Nebenbetriebsstätte Dresden | Robert Koch Institute | |
| EPI_ISL_9218276 | SYNLAB MVZ Leinfelden-Echterdingen | Robert Koch Institute | |
| EPI_ISL_9218317 | SYNLAB MVZ Leverkusen | Robert Koch Institute | |
| EPI_ISL_9220560 | Sonic - Labor Dr. von Froreich GmbH | Robert Koch Institute | |
| EPI_ISL_9229725, EPI_ISL_9229810 | Uniklinikum Carl Gustav Carus an der TU Dresden; Institut für Virologie | Robert Koch Institute | |
| EPI_ISL_9225281 | nordlab - Partnerschaftspraxis für Laboratoriumsmedizin | Robert Koch Institute | |

We gratefully acknowledge the following Authors from the Originating laboratories responsible for obtaining the specimens, as well as the Submitting laboratories where the genome data were generated and shared via GISAID, on which this research is based.

All Submitters of data may be contacted directly via www.gisaid.org

Authors are sorted alphabetically.

| Accession ID | Originating Laboratory | Submitting Laboratory | Authors |
|---|---|---|---|
| EPI_ISL_9244732 | INT Fondazione Pascale | INT Fondazione Pascale | N. Normanno INT Fondazione Pascale |

We gratefully acknowledge the following Authors from the Originating laboratories responsible for obtaining the specimens, as well as the Submitting laboratories where the genome data were generated and shared via GISAID, on which this research is based.

All Submitters of data may be contacted directly via www.gisaid.org

Authors are sorted alphabetically.

Acknowledgement EPI_SET Identifier: EPI_SET_20220527wn

| Accession ID | Originating Laboratory | Submitting Laboratory | Authors |
|---|---|---|---|
| EPI_ISL_9247372 | Atrium Health Wake Forest Baptist Microbiology Lab | Wake Forest School of Medicine, Internal Medicine | Gregory Hawkins; Kimberly Reeves |
| EPI_ISL_9260026, EPI_ISL_9260031, EPI_ISL_9260034 | Community Labs, San Antonio | STRL UT Health San Antonio, Greehey Children's Cancer Research Institute | Bethany Landry; Dawn Garcia; Guillermo Nunez; Hongxin Fan; Josefina Stoever; Korri Weldon; Kumari Vadlamudi; Marjorie Parker David; San Antonio Metropolitan Health District; Texas Department of State Health Services; Weijing He; Yidong Chen; Zhao Lai; Zhenqing Ye |
| EPI_ISL_9212993, EPI_ISL_9212999, EPI_ISL_9213000, EPI_ISL_9213040, EPI_ISL_9213148, EPI_ISL_9213151, EPI_ISL_9213170, EPI_ISL_9215135, EPI_ISL_9215138, EPI_ISL_9215151, EPI_ISL_9215156, EPI_ISL_9215175, EPI_ISL_9215197, EPI_ISL_9215205, EPI_ISL_9215214, EPI_ISL_9215232, EPI_ISL_9215307, EPI_ISL_9215319, EPI_ISL_9215323, EPI_ISL_9215324, EPI_ISL_9215336, EPI_ISL_9215338, EPI_ISL_9215352, EPI_ISL_9215402, EPI_ISL_9215459, EPI_ISL_9215463, EPI_ISL_9215466, EPI_ISL_9215472 | | | |
| see above | Curative Labs | Curative Labs | Elias L. Salfati; Eugenia Khorosheva; George Way; J.Cesar Ignacio-Espinoza; Janet Chen; Mikhail Hanewich-Hollatz; Nabjot Sandhu; Sophia Quasem; Vladimir Slepnev; Zhiyi Xie |
| EPI_ISL_9203423, EPI_ISL_9205186, EPI_ISL_9205211, EPI_ISL_9205233, EPI_ISL_9205301, EPI_ISL_9205589, EPI_ISL_9205638 | | | |
| see above | Houston Methodist Hospital | Houston Methodist Hospital | Akanksha Batajoo; Ilya J. Finkelstein; James J. Davis; Jessica Cambric; Jimmy Gollihar; Kristina Reppond; Madison N. Shyer; Marcus Nguyen; Matthew Ojeda Saavedra; Paul A. Christensen; Randall J. Olsen; Rashi Thakur; Regan Mangham; Richard Snehall; Robert Olson; Ryan Gadd; S. Wesley Long; Sindy Pena; and James M. Musser |
| EPI_ISL_9249877 | Kansas Health and Environmental Lab | Kansas Health and Environmental Lab | Amanda Bradley; Carrie Welch; Gary Buruss; Jonathan Barnell; Mike Grose; Victor Anderson; and Phil Adam |
| EPI_ISL_9257017 | LABORATORIO DE PATOLOGÍA MOLECULAR CENTRO MEDICO NAVAL SECRETARIA DE MARINA ARMADA DE MÉXICO | LABORATORIO DE PATOLOGÍA MOLECULAR CENTRO MEDICO NAVAL SECRETARIA DE MARINA ARMADA DE MÉXICO | ALVAREZ JIMENEZ VIOLETA DEJANIRA; BALTIERRA URIBE SHANTAL; GARCIA ALEGRIA IRMA.; MEIXUEIRO CALDERON CLAUDIA; MORALES RAMIREZ OMAR; ZUNO CORONADO TEUTLI |
| EPI_ISL_9257041, EPI_ISL_9257047, EPI_ISL_9257052, EPI_ISL_9257056, EPI_ISL_9257062, EPI_ISL_9257104, EPI_ISL_9257126 | | | |
| see above | Michigan Department of Health and Human Services, Bureau of Laboratories | Michigan Department of Health and Human Services, Bureau of Laboratories | Blankenship HM; Riner D; Soehnlen MK |
| EPI_ISL_9206155, EPI_ISL_9206179 | Stanford Health Care | Stanford University School of Medicine, Clinical Virology Laboratory | Malaya K. Sahoo; Mamdouh Sibai; Selamawit Bihon; Suzette Santos; and Benjamin A. Pinsky |
| EPI_ISL_9242509 | The Jackson Laboratory | The Jackson Laboratory | Adams M; Kelly K; Li L; Long E; Omerza G; Renzette N |

We gratefully acknowledge the following Authors from the Originating laboratories responsible for obtaining the specimens, as well as the Submitting laboratories where the genome data were generated and shared via GISAID, on which this research is based.

All Submitters of data may be contacted directly via www.gisaid.org

Authors are sorted alphabetically.

| Accession ID | Originating Laboratory | Submitting Laboratory | Authors |
| --- | --- | --- | --- |
| EPI_ISL_9209546 | LabPLUS | Institute of Environmental Science and Research (ESR) | Anja Werno; Antje van der Linden; Arlo Upton; Chris Mansell; Clare Gebbie; David Hammer; Dhanisha Patel; Dragana Drinkovic; Erasmus Smit; Gary McAuliffe; Hana Sofia Andersson; Hermes Perez; James Ussher; Jill Sherwood; Jing Wang; Joep de Ligt; Josh Freeman; Julia Howard; Juliet Elvy; Lauren Jelly; Mary DeAlmeida; Matt Blakiston; Matt Storey; Matthew Rogers; Max Bloomfield; Michael Addidle; Michelle Balm; Muhammad Faisal; Nikki Freed; Olin Silander; Olivia Stroeven; Rachel Boyle; Sally Roberts; SallyAnn Harbison; Sarah Cockerton; Sarah Jefferies; Sharmini Muttaiyah; Susan Morpeth; Susan Taylor; Timothy Blackmore; Vani Sathyendran; Veronica Playle; Virginia Hope; Xiaoyun Ren |

We gratefully acknowledge the following Authors from the Originating laboratories responsible for obtaining the specimens, as well as the Submitting laboratories where the genome data were generated and shared via GISAID, on which this research is based.

All Submitters of data may be contacted directly via www.gisaid.org

Authors are sorted alphabetically.

| Accession ID | Originating Laboratory | Submitting Laboratory | Authors |
|---|---|---|---|
| EPI_ISL_9268793, EPI_ISL_9268813, EPI_ISL_9268821, EPI_ISL_9268835, EPI_ISL_9268852, EPI_ISL_9268857, EPI_ISL_9268877, EPI_ISL_9268886, EPI_ISL_9268891, EPI_ISL_9268892, EPI_ISL_9268893, EPI_ISL_9268896, EPI_ISL_9268900, EPI_ISL_9268902, EPI_ISL_9268930, EPI_ISL_9268932, EPI_ISL_9268946, EPI_ISL_9268956, EPI_ISL_9268959, EPI_ISL_9268962, EPI_ISL_9268964, EPI_ISL_9268974, EPI_ISL_9268991, EPI_ISL_9268994, EPI_ISL_9268996, EPI_ISL_9269010, EPI_ISL_9269011, EPI_ISL_9269019, EPI_ISL_9269023, EPI_ISL_9269030, EPI_ISL_9269031, EPI_ISL_9269032, EPI_ISL_9269034, EPI_ISL_9269041, EPI_ISL_9269043, EPI_ISL_9269049, EPI_ISL_9269051, EPI_ISL_9269055, EPI_ISL_9269056, EPI_ISL_9269057, EPI_ISL_9269063, EPI_ISL_9269071, EPI_ISL_9269078, EPI_ISL_9269101, EPI_ISL_9269102, EPI_ISL_9269105, EPI_ISL_9269106, EPI_ISL_9269111, EPI_ISL_9269112, EPI_ISL_9269117, EPI_ISL_9269118, EPI_ISL_9269125, EPI_ISL_9269126, EPI_ISL_9269131, EPI_ISL_9269139, EPI_ISL_9269152, EPI_ISL_9269164, EPI_ISL_9269188, EPI_ISL_9269189, EPI_ISL_9269191, EPI_ISL_9269197, EPI_ISL_9269198, EPI_ISL_9269200, EPI_ISL_9269204, EPI_ISL_9269211, EPI_ISL_9269212, EPI_ISL_9269216, EPI_ISL_9269229, EPI_ISL_9269234, EPI_ISL_9269242, EPI_ISL_9269258, EPI_ISL_9269283, EPI_ISL_9269307, EPI_ISL_9269313, EPI_ISL_9269325, EPI_ISL_9269332, EPI_ISL_9269340, EPI_ISL_9269347, EPI_ISL_9269349, EPI_ISL_9269350, EPI_ISL_9269363, EPI_ISL_9269381, EPI_ISL_9269383, EPI_ISL_9269388, EPI_ISL_9269390, EPI_ISL_9269393, EPI_ISL_9269394, EPI_ISL_9269401, EPI_ISL_9269403, EPI_ISL_9269406, EPI_ISL_9269427, EPI_ISL_9269455, EPI_ISL_9269459, EPI_ISL_9269467, EPI_ISL_9269470, EPI_ISL_9269472, EPI_ISL_9269477, EPI_ISL_9269482, EPI_ISL_9269487, EPI_ISL_9269490, EPI_ISL_9269502, EPI_ISL_9269503, EPI_ISL_9269505, EPI_ISL_9269509, EPI_ISL_9269510, EPI_ISL_9269512, EPI_ISL_9269514, EPI_ISL_9269515, EPI_ISL_9269516, EPI_ISL_9269520, EPI_ISL_9269528, EPI_ISL_9269531, EPI_ISL_9269533, EPI_ISL_9269554, EPI_ISL_9269555, EPI_ISL_9269559, EPI_ISL_9269562, EPI_ISL_9269568, EPI_ISL_9269584, EPI_ISL_9269591, EPI_ISL_9269592, EPI_ISL_9269599, EPI_ISL_9269603, EPI_ISL_9269608, EPI_ISL_9269618, EPI_ISL_9269627, EPI_ISL_9269628, EPI_ISL_9269631, EPI_ISL_9269643, EPI_ISL_9269652, EPI_ISL_9269653, EPI_ISL_9269669, EPI_ISL_9269689, EPI_ISL_9269703, EPI_ISL_9269710, EPI_ISL_9269721, EPI_ISL_9269728, EPI_ISL_9269731, EPI_ISL_9269734, EPI_ISL_9269737, EPI_ISL_9269739, EPI_ISL_9269741, EPI_ISL_9269744, EPI_ISL_9269754, EPI_ISL_9269758, EPI_ISL_9269781, EPI_ISL_9269787, EPI_ISL_9269788, EPI_ISL_9269790, EPI_ISL_9269791, EPI_ISL_9269799, EPI_ISL_9269819, EPI_ISL_9269832, EPI_ISL_9269833, EPI_ISL_9269834, EPI_ISL_9269843, EPI_ISL_9269845, EPI_ISL_9269852, EPI_ISL_9269856, EPI_ISL_9269857, EPI_ISL_9269861, EPI_ISL_9269866, EPI_ISL_9269869, EPI_ISL_9269873, EPI_ISL_9269898, EPI_ISL_9269899, EPI_ISL_9269912, EPI_ISL_9269928, EPI_ISL_9269930, EPI_ISL_9269945, EPI_ISL_9269956, EPI_ISL_9269968, EPI_ISL_9269970, EPI_ISL_9269977, EPI_ISL_9269979, EPI_ISL_9269980, EPI_ISL_9269998, EPI_ISL_9270004, EPI_ISL_9270032, EPI_ISL_9270033, EPI_ISL_9270034, EPI_ISL_9270039, EPI_ISL_9270063, EPI_ISL_9270068, EPI_ISL_9270069, EPI_ISL_9270072, EPI_ISL_9270074, EPI_ISL_9270080, EPI_ISL_9270084, EPI_ISL_9270088, EPI_ISL_9270092, EPI_ISL_9270096, EPI_ISL_9270103, EPI_ISL_9270105, EPI_ISL_9270106, EPI_ISL_9270118, EPI_ISL_9270121, EPI_ISL_9270122, EPI_ISL_9270123, EPI_ISL_9270141, EPI_ISL_9270150, EPI_ISL_9270164, EPI_ISL_9270172, EPI_ISL_9270191, EPI_ISL_9270196, EPI_ISL_9270197, EPI_ISL_9270203, EPI_ISL_9270207, EPI_ISL_9270220, EPI_ISL_9270221, EPI_ISL_9270224, EPI_ISL_9270239, EPI_ISL_9270255, EPI_ISL_9270259, EPI_ISL_9270288, EPI_ISL_9270293, EPI_ISL_9270296, EPI_ISL_9270298, EPI_ISL_9270311, EPI_ISL_9270330, EPI_ISL_9270331, EPI_ISL_9270334, EPI_ISL_9270341, EPI_ISL_9270346, EPI_ISL_9270357, EPI_ISL_9270361, EPI_ISL_9270390, EPI_ISL_9270392, EPI_ISL_9270395, EPI_ISL_9270406, EPI_ISL_9270415, EPI_ISL_9270416, EPI_ISL_9270420, EPI_ISL_9270424, EPI_ISL_9270426, EPI_ISL_9270428, EPI_ISL_9270429, EPI_ISL_9270431, EPI_ISL_9270433, EPI_ISL_9270437, EPI_ISL_9270439, EPI_ISL_9270440, EPI_ISL_9270442, EPI_ISL_9270445, EPI_ISL_9270447, EPI_ISL_9270458, EPI_ISL_9270461, EPI_ISL_9270466, EPI_ISL_9270470, EPI_ISL_9270473, EPI_ISL_9270478, EPI_ISL_9270479, EPI_ISL_9270491, EPI_ISL_9270494, EPI_ISL_9270501, EPI_ISL_9270511, EPI_ISL_9270518, EPI_ISL_9270522, EPI_ISL_9270523, EPI_ISL_9270528, EPI_ISL_9270531, EPI_ISL_9270538, EPI_ISL_9270542, EPI_ISL_9270555, EPI_ISL_9270556, EPI_ISL_9270559, EPI_ISL_9270560, EPI_ISL_9270562, EPI_ISL_9270567, EPI_ISL_9270576, EPI_ISL_9270577, EPI_ISL_9270582, EPI_ISL_9270587, EPI_ISL_9270604, EPI_ISL_9270618, EPI_ISL_9270621, EPI_ISL_9270643, EPI_ISL_9270644, EPI_ISL_9270655, EPI_ISL_9270673, EPI_ISL_9270684, EPI_ISL_9270690, EPI_ISL_9270691, EPI_ISL_9270699, EPI_ISL_9270700, EPI_ISL_9270714, EPI_ISL_9270727, EPI_ISL_9270738, EPI_ISL_9270749, EPI_ISL_9270763, EPI_ISL_9270773, EPI_ISL_9270776, EPI_ISL_9270784, EPI_ISL_9270789, EPI_ISL_9270800, EPI_ISL_9270801, EPI_ISL_9270802, EPI_ISL_9270807, EPI_ISL_9270813, EPI_ISL_9270817, EPI_ISL_9270835, EPI_ISL_9270842, EPI_ISL_9270846, EPI_ISL_9270849, EPI_ISL_9270853, EPI_ISL_9270862, EPI_ISL_9270872, EPI_ISL_9270893, EPI_ISL_9270901, EPI_ISL_9270902, EPI_ISL_9270908, EPI_ISL_9270914, EPI_ISL_9270924, EPI_ISL_9270928, EPI_ISL_9270930, EPI_ISL_9270932, EPI_ISL_9270934, EPI_ISL_9270935, EPI_ISL_9270940, EPI_ISL_9270944, EPI_ISL_9270947, EPI_ISL_9270952, EPI_ISL_9270957, EPI_ISL_9270958, EPI_ISL_9270960, EPI_ISL_9270961, EPI_ISL_9270968, EPI_ISL_9270972, EPI_ISL_9270979, EPI_ISL_9270986, EPI_ISL_9270990, EPI_ISL_9270998, EPI_ISL_9271011, EPI_ISL_9271012, EPI_ISL_9271015, EPI_ISL_9271020, EPI_ISL_9271021, EPI_ISL_9271027, EPI_ISL_9271032, EPI_ISL_9271037, EPI_ISL_9271041, EPI_ISL_9271053, EPI_ISL_9271066, EPI_ISL_9271079, EPI_ISL_9271095, EPI_ISL_9271100, EPI_ISL_9271110, EPI_ISL_9271116, EPI_ISL_9271119, EPI_ISL_9271120, EPI_ISL_9271121, EPI_ISL_9271126, EPI_ISL_9271129, EPI_ISL_9271133, EPI_ISL_9271144, EPI_ISL_9271146, EPI_ISL_9271147, EPI_ISL_9271164, EPI_ISL_9271168, EPI_ISL_9271173, EPI_ISL_9271176, EPI_ISL_9271208, EPI_ISL_9271212, EPI_ISL_9271213, EPI_ISL_9271214, EPI_ISL_9271218, EPI_ISL_9271229, EPI_ISL_9271232, EPI_ISL_9271248, EPI_ISL_9271272, EPI_ISL_9271280, EPI_ISL_9271291, EPI_ISL_9271293, EPI_ISL_9271294, EPI_ISL_9271313, EPI_ISL_9271321, EPI_ISL_9271326, EPI_ISL_9271327, EPI_ISL_9271330, EPI_ISL_9271374, EPI_ISL_9271377, EPI_ISL_9271382, EPI_ISL_9271386, EPI_ISL_9271390, EPI_ISL_9271392, EPI_ISL_9271403, EPI_ISL_9271413, EPI_ISL_9271451, EPI_ISL_9271463, EPI_ISL_9271465, EPI_ISL_9271484, EPI_ISL_9271489, EPI_ISL_9271496, EPI_ISL_9271500, EPI_ISL_9271501, EPI_ISL_9271505, EPI_ISL_9271526, EPI_ISL_9271527, EPI_ISL_9271541, EPI_ISL_9271546, EPI_ISL_9271547, EPI_ISL_9271559, EPI_ISL_9271563, EPI_ISL_9271571, EPI_ISL_9271572, EPI_ISL_9271575, EPI_ISL_9271576, EPI_ISL_9271577, EPI_ISL_9271579, EPI_ISL_9271583, EPI_ISL_9271590, EPI_ISL_9271594, EPI_ISL_9271605, EPI_ISL_9271615, EPI_ISL_9271621 | | | |
| see above | Department of Bacteria, Parasites and Fungi, Statens Serum Institut, Copenhagen, Denmark | Statens Serum Institut Bioinformatics and Microbial Genomics | Danish Covid-19 Genome Consortium |

We gratefully acknowledge the following Authors from the Originating laboratories responsible for obtaining the specimens, as well as the Submitting laboratories where the genome data were generated and shared via GISAID, on which this research is based.

All Submitters of data may be contacted directly via www.gisaid.org

Authors are sorted alphabetically.

<table>
<tr><th>Accession ID</th><th>Originating Laboratory</th><th>Submitting Laboratory</th><th>Authors</th></tr>
<tr><td colspan="4">EPI_ISL_9268790, EPI_ISL_9268791, EPI_ISL_9268798, EPI_ISL_9268802, EPI_ISL_9268803, EPI_ISL_9268824, EPI_ISL_9268825, EPI_ISL_9268830, EPI_ISL_9268831, EPI_ISL_9268832, EPI_ISL_9268844, EPI_ISL_9268849, EPI_ISL_9268858, EPI_ISL_9268862, EPI_ISL_9268863, EPI_ISL_9268865, EPI_ISL_9268876, EPI_ISL_9268885, EPI_ISL_9268894, EPI_ISL_9268907, EPI_ISL_9268909, EPI_ISL_9268917, EPI_ISL_9268923, EPI_ISL_9268925, EPI_ISL_9268936, EPI_ISL_9268952, EPI_ISL_9268960, EPI_ISL_9268966, EPI_ISL_9268967, EPI_ISL_9268981, EPI_ISL_9268983, EPI_ISL_9268985, EPI_ISL_9268989, EPI_ISL_9269000, EPI_ISL_9269003, EPI_ISL_9269004, EPI_ISL_9269005, EPI_ISL_9269016, EPI_ISL_9269018, EPI_ISL_9269028, EPI_ISL_9269037, EPI_ISL_9269042, EPI_ISL_9269050, EPI_ISL_9269052, EPI_ISL_9269061, EPI_ISL_9269062, EPI_ISL_9269064, EPI_ISL_9269076, EPI_ISL_9269083, EPI_ISL_9269086, EPI_ISL_9269096, EPI_ISL_9269113, EPI_ISL_9269120, EPI_ISL_9269121, EPI_ISL_9269134, EPI_ISL_9269150, EPI_ISL_9269161, EPI_ISL_9269171, EPI_ISL_9269176, EPI_ISL_9269178, EPI_ISL_9269184, EPI_ISL_9269186, EPI_ISL_9269206, EPI_ISL_9269210, EPI_ISL_9269223, EPI_ISL_9269228, EPI_ISL_9269231, EPI_ISL_9269232, EPI_ISL_9269260, EPI_ISL_9269261, EPI_ISL_9269269, EPI_ISL_9269281, EPI_ISL_9269296, EPI_ISL_9269298, EPI_ISL_9269303, EPI_ISL_9269315, EPI_ISL_9269316, EPI_ISL_9269319, EPI_ISL_9269326, EPI_ISL_9269338, EPI_ISL_9269356, EPI_ISL_9269360, EPI_ISL_9269361, EPI_ISL_9269365, EPI_ISL_9269366, EPI_ISL_9269376, EPI_ISL_9269378, EPI_ISL_9269398, EPI_ISL_9269404, EPI_ISL_9269405, EPI_ISL_9269414, EPI_ISL_9269417, EPI_ISL_9269422, EPI_ISL_9269424, EPI_ISL_9269438, EPI_ISL_9269449, EPI_ISL_9269457, EPI_ISL_9269460, EPI_ISL_9269463, EPI_ISL_9269466, EPI_ISL_9269506, EPI_ISL_9269523, EPI_ISL_9269538, EPI_ISL_9269544, EPI_ISL_9269545, EPI_ISL_9269546, EPI_ISL_9269551, EPI_ISL_9269557, EPI_ISL_9269564, EPI_ISL_9269570, EPI_ISL_9269571, EPI_ISL_9269588, EPI_ISL_9269594, EPI_ISL_9269595, EPI_ISL_9269596, EPI_ISL_9269597, EPI_ISL_9269598, EPI_ISL_9269600, EPI_ISL_9269610, EPI_ISL_9269611, EPI_ISL_9269614, EPI_ISL_9269616, EPI_ISL_9269619, EPI_ISL_9269642, EPI_ISL_9269648, EPI_ISL_9269651, EPI_ISL_9269666, EPI_ISL_9269671, EPI_ISL_9269680, EPI_ISL_9269686, EPI_ISL_9269688, EPI_ISL_9269692, EPI_ISL_9269696, EPI_ISL_9269697, EPI_ISL_9269698, EPI_ISL_9269700, EPI_ISL_9269705, EPI_ISL_9269726, EPI_ISL_9269733, EPI_ISL_9269753, EPI_ISL_9269755, EPI_ISL_9269756, EPI_ISL_9269762, EPI_ISL_9269769, EPI_ISL_9269775, EPI_ISL_9269784, EPI_ISL_9269803, EPI_ISL_9269820, EPI_ISL_9269821, EPI_ISL_9269825, EPI_ISL_9269840, EPI_ISL_9269849, EPI_ISL_9269854, EPI_ISL_9269859, EPI_ISL_9269862, EPI_ISL_9269864, EPI_ISL_9269874, EPI_ISL_9269885, EPI_ISL_9269886, EPI_ISL_9269890, EPI_ISL_9269910, EPI_ISL_9269911, EPI_ISL_9269957, EPI_ISL_9269966, EPI_ISL_9269974, EPI_ISL_9269985, EPI_ISL_9269987, EPI_ISL_9270026, EPI_ISL_9270027, EPI_ISL_9270031, EPI_ISL_9270046, EPI_ISL_9270048, EPI_ISL_9270054, EPI_ISL_9270061, EPI_ISL_9270062, EPI_ISL_9270075, EPI_ISL_9270093, EPI_ISL_9270100, EPI_ISL_9270104, EPI_ISL_9270126, EPI_ISL_9270128, EPI_ISL_9270132, EPI_ISL_9270134, EPI_ISL_9270137, EPI_ISL_9270148, EPI_ISL_9270149, EPI_ISL_9270155, EPI_ISL_9270157, EPI_ISL_9270168, EPI_ISL_9270169, EPI_ISL_9270174, EPI_ISL_9270176, EPI_ISL_9270185, EPI_ISL_9270194, EPI_ISL_9270216, EPI_ISL_9270225, EPI_ISL_9270226, EPI_ISL_9270237, EPI_ISL_9270241, EPI_ISL_9270245, EPI_ISL_9270246, EPI_ISL_9270249, EPI_ISL_9270271, EPI_ISL_9270275, EPI_ISL_9270278, EPI_ISL_9270282, EPI_ISL_9270284, EPI_ISL_9270290, EPI_ISL_9270297, EPI_ISL_9270299, EPI_ISL_9270302, EPI_ISL_9270305, EPI_ISL_9270310, EPI_ISL_9270316, EPI_ISL_9270348, EPI_ISL_9270350, EPI_ISL_9270353, EPI_ISL_9270359, EPI_ISL_9270368, EPI_ISL_9270379, EPI_ISL_9270397, EPI_ISL_9270418, EPI_ISL_9270438, EPI_ISL_9270441, EPI_ISL_9270444, EPI_ISL_9270452, EPI_ISL_9270471, EPI_ISL_9270474, EPI_ISL_9270476, EPI_ISL_9270480, EPI_ISL_9270495, EPI_ISL_9270496, EPI_ISL_9270500, EPI_ISL_9270502, EPI_ISL_9270507, EPI_ISL_9270515, EPI_ISL_9270516, EPI_ISL_9270519, EPI_ISL_9270525, EPI_ISL_9270529, EPI_ISL_9270533, EPI_ISL_9270535, EPI_ISL_9270539, EPI_ISL_9270554, EPI_ISL_9270563, EPI_ISL_9270568, EPI_ISL_9270571, EPI_ISL_9270574, EPI_ISL_9270607, EPI_ISL_9270611, EPI_ISL_9270614, EPI_ISL_9270622, EPI_ISL_9270625, EPI_ISL_9270628, EPI_ISL_9270650, EPI_ISL_9270652, EPI_ISL_9270653, EPI_ISL_9270654, EPI_ISL_9270669, EPI_ISL_9270679, EPI_ISL_9270683, EPI_ISL_9270692, EPI_ISL_9270694, EPI_ISL_9270697, EPI_ISL_9270701, EPI_ISL_9270702, EPI_ISL_9270726, EPI_ISL_9270734, EPI_ISL_9270737, EPI_ISL_9270741, EPI_ISL_9270744, EPI_ISL_9270777, EPI_ISL_9270779, EPI_ISL_9270781, EPI_ISL_9270787, EPI_ISL_9270794, EPI_ISL_9270795, EPI_ISL_9270799, EPI_ISL_9270810, EPI_ISL_9270812, EPI_ISL_9270815, EPI_ISL_9270818, EPI_ISL_9270820, EPI_ISL_9270824, EPI_ISL_9270864, EPI_ISL_9270868, EPI_ISL_9270881, EPI_ISL_9270888, EPI_ISL_9270892, EPI_ISL_9270899, EPI_ISL_9270910, EPI_ISL_9270919, EPI_ISL_9270920, EPI_ISL_9270936, EPI_ISL_9270941, EPI_ISL_9270946, EPI_ISL_9270955, EPI_ISL_9270966, EPI_ISL_9270983, EPI_ISL_9270985, EPI_ISL_9270996, EPI_ISL_9271005, EPI_ISL_9271009, EPI_ISL_9271023, EPI_ISL_9271034, EPI_ISL_9271056, EPI_ISL_9271068, EPI_ISL_9271070, EPI_ISL_9271073, EPI_ISL_9271074, EPI_ISL_9271084, EPI_ISL_9271085, EPI_ISL_9271092, EPI_ISL_9271103, EPI_ISL_9271112, EPI_ISL_9271127, EPI_ISL_9271137, EPI_ISL_9271150, EPI_ISL_9271154, EPI_ISL_9271155, EPI_ISL_9271156, EPI_ISL_9271159, EPI_ISL_9271160, EPI_ISL_9271175, EPI_ISL_9271182, EPI_ISL_9271192, EPI_ISL_9271202, EPI_ISL_9271205, EPI_ISL_9271209, EPI_ISL_9271215, EPI_ISL_9271223, EPI_ISL_9271225, EPI_ISL_9271227, EPI_ISL_9271238, EPI_ISL_9271239, EPI_ISL_9271241, EPI_ISL_9271244, EPI_ISL_9271247, EPI_ISL_9271250, EPI_ISL_9271261, EPI_ISL_9271287, EPI_ISL_9271295, EPI_ISL_9271299, EPI_ISL_9271300, EPI_ISL_9271301, EPI_ISL_9271304, EPI_ISL_9271312, EPI_ISL_9271314, EPI_ISL_9271325, EPI_ISL_9271331, EPI_ISL_9271337, EPI_ISL_9271339, EPI_ISL_9271340, EPI_ISL_9271341, EPI_ISL_9271349, EPI_ISL_9271359, EPI_ISL_9271375, EPI_ISL_9271383, EPI_ISL_9271387, EPI_ISL_9271391, EPI_ISL_9271405, EPI_ISL_9271411, EPI_ISL_9271429, EPI_ISL_9271431, EPI_ISL_9271435, EPI_ISL_9271441, EPI_ISL_9271444, EPI_ISL_9271448, EPI_ISL_9271450, EPI_ISL_9271453, EPI_ISL_9271459, EPI_ISL_9271468, EPI_ISL_9271476, EPI_ISL_9271481, EPI_ISL_9271492, EPI_ISL_9271493, EPI_ISL_9271494, EPI_ISL_9271502, EPI_ISL_9271514, EPI_ISL_9271521, EPI_ISL_9271528, EPI_ISL_9271530, EPI_ISL_9271534, EPI_ISL_9271540, EPI_ISL_9271555, EPI_ISL_9271566, EPI_ISL_9271567, EPI_ISL_9271578, EPI_ISL_9271585, EPI_ISL_9271591, EPI_ISL_9271595, EPI_ISL_9271604, EPI_ISL_9271618, EPI_ISL_9271627</td></tr>
<tr><td>see above</td><td>Department of Bacteria, Parasites and Fungi, Statens Serum Institut, Copenhagen, Denmark</td><td>Statens Serum Institut Bioinformatics and Microbial Genomics</td><td>Danish Covid-19 Genome Consortium</td></tr>
</table>

We gratefully acknowledge the following Authors from the Originating laboratories responsible for obtaining the specimens, as well as the Submitting laboratories where the genome data were generated and shared via GISAID, on which this research is based.

All Submitters of data may be contacted directly via www.gisaid.org

Authors are sorted alphabetically.

| Accession ID | Originating Laboratory | Submitting Laboratory | Authors |
| --- | --- | --- | --- |
| EPI_ISL_9268856, EPI_ISL_9268908, EPI_ISL_9268915, EPI_ISL_9268927, EPI_ISL_9268931, EPI_ISL_9268943, EPI_ISL_9268949, EPI_ISL_9268971, EPI_ISL_9268972, EPI_ISL_9268980, EPI_ISL_9269014, EPI_ISL_9269053, EPI_ISL_9269080, EPI_ISL_9269103, EPI_ISL_9269108, EPI_ISL_9269133, EPI_ISL_9269158, EPI_ISL_9269259, EPI_ISL_9269282, EPI_ISL_9269317, EPI_ISL_9269330, EPI_ISL_9269343, EPI_ISL_9269345, EPI_ISL_9269375, EPI_ISL_9269387, EPI_ISL_9269407, EPI_ISL_9269411, EPI_ISL_9269419, EPI_ISL_9269431, EPI_ISL_9269444, EPI_ISL_9269462, EPI_ISL_9269491, EPI_ISL_9269507, EPI_ISL_9269582, EPI_ISL_9269630, EPI_ISL_9269636, EPI_ISL_9269656, EPI_ISL_9269658, EPI_ISL_9269714, EPI_ISL_9269715, EPI_ISL_9269718, EPI_ISL_9269729, EPI_ISL_9269730, EPI_ISL_9269800, EPI_ISL_9269835, EPI_ISL_9269860, EPI_ISL_9269915, EPI_ISL_9269922, EPI_ISL_9269923, EPI_ISL_9269949, EPI_ISL_9269955, EPI_ISL_9269958, EPI_ISL_9269960, EPI_ISL_9269973, EPI_ISL_9269993, EPI_ISL_9269999, EPI_ISL_9270009, EPI_ISL_9270014, EPI_ISL_9270023, EPI_ISL_9270035, EPI_ISL_9270041, EPI_ISL_9270055, EPI_ISL_9270060, EPI_ISL_9270085, EPI_ISL_9270089, EPI_ISL_9270133, EPI_ISL_9270138, EPI_ISL_9270175, EPI_ISL_9270181, EPI_ISL_9270183, EPI_ISL_9270192, EPI_ISL_9270200, EPI_ISL_9270219, EPI_ISL_9270242, EPI_ISL_9270257, EPI_ISL_9270295, EPI_ISL_9270324, EPI_ISL_9270333, EPI_ISL_9270335, EPI_ISL_9270358, EPI_ISL_9270377, EPI_ISL_9270382, EPI_ISL_9270399, EPI_ISL_9270421, EPI_ISL_9270435, EPI_ISL_9270455, EPI_ISL_9270486, EPI_ISL_9270508, EPI_ISL_9270550, EPI_ISL_9270566, EPI_ISL_9270570, EPI_ISL_9270586, EPI_ISL_9270589, EPI_ISL_9270602, EPI_ISL_9270631, EPI_ISL_9270708, EPI_ISL_9270709, EPI_ISL_9270710, EPI_ISL_9270712, EPI_ISL_9270713, EPI_ISL_9270722, EPI_ISL_9270728, EPI_ISL_9270730, EPI_ISL_9270758, EPI_ISL_9270783, EPI_ISL_9270804, EPI_ISL_9270827, EPI_ISL_9270830, EPI_ISL_9270844, EPI_ISL_9270850, EPI_ISL_9270852, EPI_ISL_9270858, EPI_ISL_9270876, EPI_ISL_9270880, EPI_ISL_9270906, EPI_ISL_9270911, EPI_ISL_9270943, EPI_ISL_9270963, EPI_ISL_9270964, EPI_ISL_9270969, EPI_ISL_9270971, EPI_ISL_9270982, EPI_ISL_9270987, EPI_ISL_9270989, EPI_ISL_9271064, EPI_ISL_9271077, EPI_ISL_9271178, EPI_ISL_9271188, EPI_ISL_9271245, EPI_ISL_9271273, EPI_ISL_9271278, EPI_ISL_9271286, EPI_ISL_9271289, EPI_ISL_9271307, EPI_ISL_9271351, EPI_ISL_9271428, EPI_ISL_9271432, EPI_ISL_9271437, EPI_ISL_9271462, EPI_ISL_9271466, EPI_ISL_9271532, EPI_ISL_9271544, EPI_ISL_9271545, EPI_ISL_9271548, EPI_ISL_9271550, EPI_ISL_9271561, EPI_ISL_9271562, EPI_ISL_9271564, EPI_ISL_9271588, EPI_ISL_9271592, EPI_ISL_9271600, EPI_ISL_9271628 | | | |
| see above | Department of Bacteria, Parasites and Fungi, Statens Serum Institut, Copenhagen, Denmark | Statens Serum Institut Bioinformatics and Microbial Genomics | Danish Covid-19 Genome Consortium |

We gratefully acknowledge the following Authors from the Originating laboratories responsible for obtaining the specimens, as well as the Submitting laboratories where the genome data were generated and shared via GISAID, on which this research is based.

All Submitters of data may be contacted directly via www.gisaid.org

Authors are sorted alphabetically.

Acknowledgement EPI_SET Identifier: EPI_SET_20220527kt

| Accession ID | Originating Laboratory | Submitting Laboratory | Authors |
|---|---|---|---|
| EPI_ISL_9268794, EPI_ISL_9268827, EPI_ISL_9268834, EPI_ISL_9268846, EPI_ISL_9268860, EPI_ISL_9268864, EPI_ISL_9268901, EPI_ISL_9268919, EPI_ISL_9268934, EPI_ISL_9268997, EPI_ISL_9269007, EPI_ISL_9269012, EPI_ISL_9269073, EPI_ISL_9269095, EPI_ISL_9269155, EPI_ISL_9269156, EPI_ISL_9269168, EPI_ISL_9269175, EPI_ISL_9269215, EPI_ISL_9269221, EPI_ISL_9269224, EPI_ISL_9269227, EPI_ISL_9269233, EPI_ISL_9269275, EPI_ISL_9269290, EPI_ISL_9269305, EPI_ISL_9269323, EPI_ISL_9269333, EPI_ISL_9269359, EPI_ISL_9269389, EPI_ISL_9269402, EPI_ISL_9269430, EPI_ISL_9269443, EPI_ISL_9269445, EPI_ISL_9269468, EPI_ISL_9269478, EPI_ISL_9269499, EPI_ISL_9269513, EPI_ISL_9269547, EPI_ISL_9269563, EPI_ISL_9269625, EPI_ISL_9269641, EPI_ISL_9269660, EPI_ISL_9269665, EPI_ISL_9269674, EPI_ISL_9269675, EPI_ISL_9269702, EPI_ISL_9269717, EPI_ISL_9269724, EPI_ISL_9269740, EPI_ISL_9269765, EPI_ISL_9269768, EPI_ISL_9269795, EPI_ISL_9269814, EPI_ISL_9269815, EPI_ISL_9269818, EPI_ISL_9269884, EPI_ISL_9269887, EPI_ISL_9269892, EPI_ISL_9269903, EPI_ISL_9269916, EPI_ISL_9269963, EPI_ISL_9269976, EPI_ISL_9269982, EPI_ISL_9269992, EPI_ISL_9270008, EPI_ISL_9270022, EPI_ISL_9270044, EPI_ISL_9270091, EPI_ISL_9270107, EPI_ISL_9270110, EPI_ISL_9270111, EPI_ISL_9270144, EPI_ISL_9270166, EPI_ISL_9270190, EPI_ISL_9270211, EPI_ISL_9270222, EPI_ISL_9270228, EPI_ISL_9270232, EPI_ISL_9270240, EPI_ISL_9270258, EPI_ISL_9270268, EPI_ISL_9270320, EPI_ISL_9270322, EPI_ISL_9270360, EPI_ISL_9270383, EPI_ISL_9270391, EPI_ISL_9270403, EPI_ISL_9270432, EPI_ISL_9270449, EPI_ISL_9270460, EPI_ISL_9270475, EPI_ISL_9270482, EPI_ISL_9270504, EPI_ISL_9270506, EPI_ISL_9270572, EPI_ISL_9270573, EPI_ISL_9270590, EPI_ISL_9270597, EPI_ISL_9270599, EPI_ISL_9270605, EPI_ISL_9270613, EPI_ISL_9270639, EPI_ISL_9270648, EPI_ISL_9270674, EPI_ISL_9270682, EPI_ISL_9270716, EPI_ISL_9270759, EPI_ISL_9270762, EPI_ISL_9270765, EPI_ISL_9270768, EPI_ISL_9270786, EPI_ISL_9270825, EPI_ISL_9270831, EPI_ISL_9270851, EPI_ISL_9270866, EPI_ISL_9270886, EPI_ISL_9270889, EPI_ISL_9270916, EPI_ISL_9270933, EPI_ISL_9270945, EPI_ISL_9270949, EPI_ISL_9270995, EPI_ISL_9271008, EPI_ISL_9271018, EPI_ISL_9271042, EPI_ISL_9271048, EPI_ISL_9271062, EPI_ISL_9271114, EPI_ISL_9271158, EPI_ISL_9271163, EPI_ISL_9271167, EPI_ISL_9271186, EPI_ISL_9271187, EPI_ISL_9271194, EPI_ISL_9271206, EPI_ISL_9271220, EPI_ISL_9271240, EPI_ISL_9271259, EPI_ISL_9271275, EPI_ISL_9271281, EPI_ISL_9271308, EPI_ISL_9271317, EPI_ISL_9271345, EPI_ISL_9271346, EPI_ISL_9271348, EPI_ISL_9271356, EPI_ISL_9271357, EPI_ISL_9271364, EPI_ISL_9271365, EPI_ISL_9271366, EPI_ISL_9271367, EPI_ISL_9271379, EPI_ISL_9271388, EPI_ISL_9271393, EPI_ISL_9271410, EPI_ISL_9271423, EPI_ISL_9271430, EPI_ISL_9271438, EPI_ISL_9271449, EPI_ISL_9271454, EPI_ISL_9271479, EPI_ISL_9271490, EPI_ISL_9271497, EPI_ISL_9271522, EPI_ISL_9271539, EPI_ISL_9271549, EPI_ISL_9271586, EPI_ISL_9271598, EPI_ISL_9271607, EPI_ISL_9271609, EPI_ISL_9271611, EPI_ISL_9271612, EPI_ISL_9271614 | | | |
| see above | Department of Bacteria, Parasites and Fungi, Statens Serum Institut, Copenhagen, Denmark | Statens Serum Institut Bioinformatics and Microbial Genomics | Danish Covid-19 Genome Consortium |

We gratefully acknowledge the following Authors from the Originating laboratories responsible for obtaining the specimens, as well as the Submitting laboratories where the genome data were generated and shared via GISAID, on which this research is based.

All Submitters of data may be contacted directly via www.gisaid.org

Authors are sorted alphabetically.

| Accession ID | Originating Laboratory | Submitting Laboratory | Authors |
| --- | --- | --- | --- |
| EPI_ISL_9268792, EPI_ISL_9268795, EPI_ISL_9268797, EPI_ISL_9268799, EPI_ISL_9268804, EPI_ISL_9268806, EPI_ISL_9268811, EPI_ISL_9268812, EPI_ISL_9268814, EPI_ISL_9268815, EPI_ISL_9268816, EPI_ISL_9268819, EPI_ISL_9268826, EPI_ISL_9268839, EPI_ISL_9268843, EPI_ISL_9268867, EPI_ISL_9268868, EPI_ISL_9268874, EPI_ISL_9268884, EPI_ISL_9268887, EPI_ISL_9268889, EPI_ISL_9268904, EPI_ISL_9268920, EPI_ISL_9268954, EPI_ISL_9268965, EPI_ISL_9268968, EPI_ISL_9268970, EPI_ISL_9268995, EPI_ISL_9269008, EPI_ISL_9269024, EPI_ISL_9269025, EPI_ISL_9269035, EPI_ISL_9269059, EPI_ISL_9269065, EPI_ISL_9269066, EPI_ISL_9269067, EPI_ISL_9269091, EPI_ISL_9269107, EPI_ISL_9269116, EPI_ISL_9269135, EPI_ISL_9269146, EPI_ISL_9269148, EPI_ISL_9269149, EPI_ISL_9269159, EPI_ISL_9269166, EPI_ISL_9269170, EPI_ISL_9269174, EPI_ISL_9269177, EPI_ISL_9269181, EPI_ISL_9269199, EPI_ISL_9269201, EPI_ISL_9269202, EPI_ISL_9269205, EPI_ISL_9269208, EPI_ISL_9269209, EPI_ISL_9269217, EPI_ISL_9269226, EPI_ISL_9269230, EPI_ISL_9269235, EPI_ISL_9269237, EPI_ISL_9269241, EPI_ISL_9269244, EPI_ISL_9269245, EPI_ISL_9269248, EPI_ISL_9269251, EPI_ISL_9269253, EPI_ISL_9269263, EPI_ISL_9269268, EPI_ISL_9269274, EPI_ISL_9269280, EPI_ISL_9269284, EPI_ISL_9269287, EPI_ISL_9269289, EPI_ISL_9269293, EPI_ISL_9269304, EPI_ISL_9269314, EPI_ISL_9269321, EPI_ISL_9269324, EPI_ISL_9269335, EPI_ISL_9269336, EPI_ISL_9269337, EPI_ISL_9269341, EPI_ISL_9269344, EPI_ISL_9269371, EPI_ISL_9269386, EPI_ISL_9269392, EPI_ISL_9269397, EPI_ISL_9269408, EPI_ISL_9269413, EPI_ISL_9269451, EPI_ISL_9269452, EPI_ISL_9269453, EPI_ISL_9269454, EPI_ISL_9269469, EPI_ISL_9269480, EPI_ISL_9269483, EPI_ISL_9269486, EPI_ISL_9269494, EPI_ISL_9269497, EPI_ISL_9269498, EPI_ISL_9269508, EPI_ISL_9269511, EPI_ISL_9269522, EPI_ISL_9269525, EPI_ISL_9269530, EPI_ISL_9269532, EPI_ISL_9269536, EPI_ISL_9269537, EPI_ISL_9269539, EPI_ISL_9269540, EPI_ISL_9269541, EPI_ISL_9269548, EPI_ISL_9269550, EPI_ISL_9269566, EPI_ISL_9269572, EPI_ISL_9269580, EPI_ISL_9269583, EPI_ISL_9269586, EPI_ISL_9269587, EPI_ISL_9269612, EPI_ISL_9269620, EPI_ISL_9269629, EPI_ISL_9269633, EPI_ISL_9269637, EPI_ISL_9269640, EPI_ISL_9269657, EPI_ISL_9269663, EPI_ISL_9269664, EPI_ISL_9269679, EPI_ISL_9269681, EPI_ISL_9269693, EPI_ISL_9269695, EPI_ISL_9269711, EPI_ISL_9269713, EPI_ISL_9269719, EPI_ISL_9269720, EPI_ISL_9269722, EPI_ISL_9269725, EPI_ISL_9269736, EPI_ISL_9269749, EPI_ISL_9269763, EPI_ISL_9269773, EPI_ISL_9269785, EPI_ISL_9269789, EPI_ISL_9269796, EPI_ISL_9269801, EPI_ISL_9269802, EPI_ISL_9269811, EPI_ISL_9269812, EPI_ISL_9269823, EPI_ISL_9269827, EPI_ISL_9269831, EPI_ISL_9269844, EPI_ISL_9269846, EPI_ISL_9269870, EPI_ISL_9269875, EPI_ISL_9269878, EPI_ISL_9269879, EPI_ISL_9269881, EPI_ISL_9269889, EPI_ISL_9269900, EPI_ISL_9269904, EPI_ISL_9269907, EPI_ISL_9269913, EPI_ISL_9269921, EPI_ISL_9269926, EPI_ISL_9269927, EPI_ISL_9269932, EPI_ISL_9269940, EPI_ISL_9269942, EPI_ISL_9269944, EPI_ISL_9269946, EPI_ISL_9269962, EPI_ISL_9269965, EPI_ISL_9269975, EPI_ISL_9269990, EPI_ISL_9269997, EPI_ISL_9270013, EPI_ISL_9270017, EPI_ISL_9270019, EPI_ISL_9270020, EPI_ISL_9270021, EPI_ISL_9270028, EPI_ISL_9270030, EPI_ISL_9270036, EPI_ISL_9270047, EPI_ISL_9270049, EPI_ISL_9270071, EPI_ISL_9270077, EPI_ISL_9270081, EPI_ISL_9270083, EPI_ISL_9270095, EPI_ISL_9270101, EPI_ISL_9270109, EPI_ISL_9270113, EPI_ISL_9270115, EPI_ISL_9270124, EPI_ISL_9270139, EPI_ISL_9270140, EPI_ISL_9270145, EPI_ISL_9270147, EPI_ISL_9270154, EPI_ISL_9270156, EPI_ISL_9270158, EPI_ISL_9270160, EPI_ISL_9270162, EPI_ISL_9270163, EPI_ISL_9270165, EPI_ISL_9270167, EPI_ISL_9270171, EPI_ISL_9270179, EPI_ISL_9270187, EPI_ISL_9270188, EPI_ISL_9270189, EPI_ISL_9270199, EPI_ISL_9270209, EPI_ISL_9270210, EPI_ISL_9270214, EPI_ISL_9270217, EPI_ISL_9270218, EPI_ISL_9270229, EPI_ISL_9270230, EPI_ISL_9270234, EPI_ISL_9270235, EPI_ISL_9270248, EPI_ISL_9270250, EPI_ISL_9270262, EPI_ISL_9270265, EPI_ISL_9270272, EPI_ISL_9270273, EPI_ISL_9270277, EPI_ISL_9270292, EPI_ISL_9270300, EPI_ISL_9270307, EPI_ISL_9270313, EPI_ISL_9270315, EPI_ISL_9270323, EPI_ISL_9270325, EPI_ISL_9270332, EPI_ISL_9270351, EPI_ISL_9270352, EPI_ISL_9270355, EPI_ISL_9270364, EPI_ISL_9270367, EPI_ISL_9270380, EPI_ISL_9270381, EPI_ISL_9270400, EPI_ISL_9270409, EPI_ISL_9270411, EPI_ISL_9270417, EPI_ISL_9270425, EPI_ISL_9270430, EPI_ISL_9270436, EPI_ISL_9270443, EPI_ISL_9270446, EPI_ISL_9270450, EPI_ISL_9270464, EPI_ISL_9270481, EPI_ISL_9270483, EPI_ISL_9270485, EPI_ISL_9270487, EPI_ISL_9270490, EPI_ISL_9270505, EPI_ISL_9270512, EPI_ISL_9270513, EPI_ISL_9270517, EPI_ISL_9270520, EPI_ISL_9270530, EPI_ISL_9270541, EPI_ISL_9270543, EPI_ISL_9270548, EPI_ISL_9270557, EPI_ISL_9270564, EPI_ISL_9270575, EPI_ISL_9270583, EPI_ISL_9270585, EPI_ISL_9270588, EPI_ISL_9270592, EPI_ISL_9270595, EPI_ISL_9270596, EPI_ISL_9270600, EPI_ISL_9270601, EPI_ISL_9270603, EPI_ISL_9270610, EPI_ISL_9270630, EPI_ISL_9270640, EPI_ISL_9270641, EPI_ISL_9270651, EPI_ISL_9270667, EPI_ISL_9270678, EPI_ISL_9270685, EPI_ISL_9270686, EPI_ISL_9270689, EPI_ISL_9270705, EPI_ISL_9270718, EPI_ISL_9270723, EPI_ISL_9270724, EPI_ISL_9270731, EPI_ISL_9270735, EPI_ISL_9270739, EPI_ISL_9270743, EPI_ISL_9270745, EPI_ISL_9270752, EPI_ISL_9270757, EPI_ISL_9270772, EPI_ISL_9270793, EPI_ISL_9270796, EPI_ISL_9270805, EPI_ISL_9270811, EPI_ISL_9270816, EPI_ISL_9270829, EPI_ISL_9270832, EPI_ISL_9270833, EPI_ISL_9270854, EPI_ISL_9270855, EPI_ISL_9270857, EPI_ISL_9270861, EPI_ISL_9270863, EPI_ISL_9270874, EPI_ISL_9270877, EPI_ISL_9270878, EPI_ISL_9270879, EPI_ISL_9270890, EPI_ISL_9270891, EPI_ISL_9270897, EPI_ISL_9270907, EPI_ISL_9270913, EPI_ISL_9270917, EPI_ISL_9270921, EPI_ISL_9270923, EPI_ISL_9270931, EPI_ISL_9270938, EPI_ISL_9270956, EPI_ISL_9270959, EPI_ISL_9270977, EPI_ISL_9270984, EPI_ISL_9270988, EPI_ISL_9270993, EPI_ISL_9271001, EPI_ISL_9271002, EPI_ISL_9271004, EPI_ISL_9271006, EPI_ISL_9271010, EPI_ISL_9271013, EPI_ISL_9271024, EPI_ISL_9271028, EPI_ISL_9271031, EPI_ISL_9271036, EPI_ISL_9271038, EPI_ISL_9271044, EPI_ISL_9271051, EPI_ISL_9271052, EPI_ISL_9271054, EPI_ISL_9271057, EPI_ISL_9271058, EPI_ISL_9271083, EPI_ISL_9271088, EPI_ISL_9271089, EPI_ISL_9271096, EPI_ISL_9271101, EPI_ISL_9271102, EPI_ISL_9271113, EPI_ISL_9271115, EPI_ISL_9271122, EPI_ISL_9271131, EPI_ISL_9271132, EPI_ISL_9271135, EPI_ISL_9271143, EPI_ISL_9271174, EPI_ISL_9271177, EPI_ISL_9271179, EPI_ISL_9271185, EPI_ISL_9271190, EPI_ISL_9271198, EPI_ISL_9271200, EPI_ISL_9271210, EPI_ISL_9271224, EPI_ISL_9271228, EPI_ISL_9271231, EPI_ISL_9271235, EPI_ISL_9271254, EPI_ISL_9271258, EPI_ISL_9271263, EPI_ISL_9271268, EPI_ISL_9271269, EPI_ISL_9271270, EPI_ISL_9271277, EPI_ISL_9271297, EPI_ISL_9271302, EPI_ISL_9271303, EPI_ISL_9271305, EPI_ISL_9271315, EPI_ISL_9271318, EPI_ISL_9271320, EPI_ISL_9271324, EPI_ISL_9271328, EPI_ISL_9271344, EPI_ISL_9271352, EPI_ISL_9271354, EPI_ISL_9271368, EPI_ISL_9271376, EPI_ISL_9271398, EPI_ISL_9271400, EPI_ISL_9271401, EPI_ISL_9271404, EPI_ISL_9271408, EPI_ISL_9271412, EPI_ISL_9271414, EPI_ISL_9271417, EPI_ISL_9271418, EPI_ISL_9271422, EPI_ISL_9271426, EPI_ISL_9271427, EPI_ISL_9271439, EPI_ISL_9271446, EPI_ISL_9271460, EPI_ISL_9271473, EPI_ISL_9271474, EPI_ISL_9271478, EPI_ISL_9271495, EPI_ISL_9271499, EPI_ISL_9271510, EPI_ISL_9271513, EPI_ISL_9271517, EPI_ISL_9271531, EPI_ISL_9271535, EPI_ISL_9271537, EPI_ISL_9271553, EPI_ISL_9271565, EPI_ISL_9271574, EPI_ISL_9271580, EPI_ISL_9271602, EPI_ISL_9271608, EPI_ISL_9271613, EPI_ISL_9271619, EPI_ISL_9271623, EPI_ISL_9271625 | | | |
| see above | Department of Bacteria, Parasites and Fungi, Statens Serum Institut, Copenhagen, Denmark | Statens Serum Institut Bioinformatics and Microbial Genomics | Danish Covid-19 Genome Consortium |

We gratefully acknowledge the following Authors from the Originating laboratories responsible for obtaining the specimens, as well as the Submitting laboratories where the genome data were generated and shared via GISAID, on which this research is based.

All Submitters of data may be contacted directly via www.gisaid.org

Authors are sorted alphabetically.

| Accession ID | Originating Laboratory | Submitting Laboratory | Authors |
|---|---|---|---|
| EPI_ISL_9267862 | Respiratory Virus Unit, Microbiology Services Colindale, Public Health England | COVID-19 Genomics UK (COG-UK) Consortium | PHE Covid Sequencing Team |

We gratefully acknowledge the following Authors from the Originating laboratories responsible for obtaining the specimens, as well as the Submitting laboratories where the genome data were generated and shared via GISAID, on which this research is based.

All Submitters of data may be contacted directly via www.gisaid.org

Authors are sorted alphabetically.

| Accession ID | Originating Laboratory | Submitting Laboratory | Authors |
|---|---|---|---|
| EPI_ISL_9264723, EPI_ISL_9264727 | CDPH VBL | California Department of Public Health | CDPH COVIDNet |
| EPI_ISL_9264440, EPI_ISL_9264441, EPI_ISL_9264442, EPI_ISL_9264447, EPI_ISL_9264448, EPI_ISL_9264449, EPI_ISL_9264451 | | | |
| see above | Pathology, UAB | Pathology, UAB | E.J.; Hendrickson; Leal; Lefkowitz; Moates, D.; R.C.; S.M.; Van Der Pol; W.J. |
| EPI_ISL_9263791 | State Laboratories Division, Hawaii State Department of Health | State Laboratories Division, Hawaii State Department of Health | Ayana Garnet; Briana Ofilas; Cheryl-lynn Daquip; Cheyenne Barela; Daniel Strange; Drew Kuwazaki; Edward Desmond; Jeffrey Au; Mark Nagata; Pamela O'Brien; Razvan Sultana; Remedios Gose; Samantha Cotter; Samantha Sruba |

We gratefully acknowledge the following Authors from the Originating laboratories responsible for obtaining the specimens, as well as the Submitting laboratories where the genome data were generated and shared via GISAID, on which this research is based.

All Submitters of data may be contacted directly via www.gisaid.org

Authors are sorted alphabetically.

| Accession ID | Originating Laboratory | Submitting Laboratory | Authors |
|---|---|---|---|
| EPI_ISL_9274900 | Hospital da Baleia | Instituto Rene Rachou / Fiocruz Minas | Alana Oliveira; Anna Salim; Camila Corsini; Daniel Miranda; Gabriel Fernandes; Mozar de Castro; Nathalie Almeida; Pedro Alves; Priscilla Filgueiras; Rafaella Fortini; Raphael Silva; Raquel Vilela; Rubens do Monte Neto; Sarah Gomes; Thais Silva; Wander Jeremias |
| EPI_ISL_9274974, EPI_ISL_9274978, EPI_ISL_9274982, EPI_ISL_9274983, EPI_ISL_9275005, EPI_ISL_9275023, EPI_ISL_9275024, EPI_ISL_9275031, EPI_ISL_9275047, EPI_ISL_9275059, EPI_ISL_9275061 | | | |
| see above | Laboratorio Municipal de Biologia Molecular | Instituto Rene Rachou / Fiocruz Minas | Andre Menezes; Anna Salim; Eneida Oliveira; Gabriel Fernandes; Pedro Alves; Rubens do Monte Neto; Thais Silva |
